# Global burden of musculoskeletal disorders in postmenopausal women across 204 countries and territories: a systematic analysis of the 2021 Global Burden of Disease Study with projections to 2050

**DOI:** 10.1101/2025.04.27.25326322

**Authors:** Gan Li, Lingkang Dong, Jibing He, Qihang Fang, Chao Zhang, Chu’an Gao, Daoyu Zhu, Haoyu Fang, Binyu chen, Xiao Xie, Chenyi Jiang, Peng Ding, Youshui Gao

## Abstract

**Objective:** This study aimed to systematically analyze the epidemiological characteristics of six major musculoskeletal disorders (MSDs) among postmenopausal women globally from 1990 to 2021, with a focus on the spatiotemporal distribution patterns of disease prevalence and associated disability-adjusted life years (DALYs).

**Methods:** Utilizing the 2021 Global Burden of Disease (GBD) database, we examined six MSDs in postmenopausal women: rheumatoid arthritis, osteoarthritis, low back pain, neck pain, gout, and other musculoskeletal disorders. Age-standardized rates (ASRs) were calculated using internationally recognized methods, and estimated annual percentage changes (EAPCs) were employed to assess temporal trends in disease burden. The analysis incorporated considerations of age structure, sociodemographic index (SDI), and various risk factors.

**Results:** From 1990 to 2021, the global burden of MSDs among postmenopausal women showed a persistent increase, with marked heterogeneity across disease subtypes and geographical regions. Osteoarthritis represented the predominant disease burden, while gout exhibited the most rapid growth despite its lower baseline prevalence. Notably, low back pain was the only subtype demonstrating a declining trend. The burden of MSDs shifted from high-SDI regions to developing areas, with middle-SDI countries emerging as new priorities for intervention. Older age groups bore a disproportionately high disease burden, and obesity was identified as the leading risk factor driving disease progression, whereas tobacco control measures showed positive effects. Projections indicated that MSD burden will continue to rise with population aging, necessitating tailored prevention and management strategies to address this major public health challenge.

**Conclusion:** The prevalence of MSDs among postmenopausal women continues to rise, underscoring its profound impact on global health in aging female populations. To address this challenge, a multidimensional intervention approach is required, including: (1) enhanced early screening and standardized management of MSD-related risk factors; (2) personalized lifestyle interventions based on individual BMI levels; and (3) implementation of differentiated public health policies tailored to population-specific characteristics. The comprehensive adoption of these measures will help improve musculoskeletal health in postmenopausal women and reduce the overall disease burden attributable to MSDs.

## Introduction

With the accelerating global aging population, the prevention and control of musculoskeletal disorders (MSDs) have emerged as a critical public health priority.^1–4^ These conditions primarily affect bones, joints, and surrounding soft tissues, characterized by progressive degenerative pathological changes accompanied by chronic pain and deteriorating functional impairment.^5^ Epidemiological studies indicate that MSDs rank among the leading causes of disability and diminished quality of life worldwide, with disease burden escalating alongside demographic aging.^6^ Current clinical management of MSDs presents distinct challenges across disease subtypes. Therapeutically, while effective pharmacological treatments exist for rheumatoid arthritis and gout, osteoarthritis—the most prevalent degenerative joint disease—lacks breakthrough therapies.^7^ From a healthcare delivery perspective, chronic low back and neck pain, despite causing persistent pain, progressive functional limitations, and substantial socioeconomic burdens,^4, 8^ are frequently categorized as “non-specific” conditions and consequently receive inadequate attention. This landscape underscores the urgent need for enhanced early prevention and standardized management protocols, particularly for postmenopausal women as a high-risk population.

Menopause marks a critical transition point for women’s musculoskeletal health. The sharp decline in estrogen levels accelerates bone loss and joint degeneration, which without timely intervention will significantly increase disease burden in later life. Notably, the global average postmenopausal life expectancy exceeds 20 years, with women living 5·1 years longer than men,^9, 10^ giving postmenopausal women’s health management special importance in addressing global aging challenges. Existing evidence shows MSDs are significantly associated with loss of functional independence and reduced quality of life in postmenopausal women.^11^ Therefore, establishing comprehensive MSD prevention and control strategies through early intervention and standardized treatment is essential to improve musculoskeletal health and maintain functional independence.

The disparities in prevalence and disease burden of MSDs among postmenopausal women primarily arise from inequitable distribution of healthcare resources. The perimenopausal period serves as a critical transitional phase for women’s musculoskeletal health, during which the quality of health management is directly impacted by the completeness of healthcare systems. Data demonstrate substantial gaps in medical resource allocation across nations with varying socioeconomic development levels, where high SDI countries achieve a healthcare access and quality index of 83·4 compared to merely 30·7 in low SDI countries.^12^ In terms of universal health coverage, Nordic nations exceed 95% while certain sub-Saharan African regions remain below 25%.^13, 14^ Significant geographical variations exist in healthcare workforce distribution, evidenced by a nearly ten-fold difference in physician-to-population ratios between high-income and low-income countries,^15, 16^ with even more pronounced disparities in per capita health expenditures. This unbalanced resource allocation creates substantial differences in access to essential medical services for perimenopausal women during this physiologically crucial transition period, consequently exerting adverse effects on long-term skeletal health outcomes.

Although the epidemiological data of MSD in postmenopausal women and its impact on global aging have garnered significant attention,^11, 17–21^ there remains a substantial gap in understanding the evolving patterns of disease burden, risk factor distribution, and health disparities among this population at global, regional, and national levels. Against this backdrop, this study aims to address the following key questions: What dynamic changes occurred in the spectrum of MSD burden, risk factor composition, and degree of health inequality among postmenopausal women between 1990 and 2021? How will the MSD burden in this population develop from 2022 to 2050? Do health disparities in MSD burden among postmenopausal women across different countries exhibit correlations with SDI?

## Research in Context

### Evidence Before This Study

The GBD study has long focused on MSDs in middle-aged and older populations but overlooked postmenopausal women as a critical subgroup. The sharp decline in estrogen levels after menopause accelerates bone and joint degeneration, yet systematic assessments of MSDs in this population remain lacking. Existing data indicate that MSDs are a leading cause of disability and reduced quality of life in older women, with postmenopausal women facing over 20 years of survival on average, underscoring the unique importance of their health management. To address this gap, we conducted a systematic evaluation of MSD epidemiology from 1990 to 2021 using GBD 2021 data.

### Added Value of This Study

As the first global study on MSDs in postmenopausal women, our analysis of GBD 2021 data reveals three key trends: (1) By 2050, the burden of MSDs in postmenopausal women will continue to rise, with osteoarthritis remaining predominant while gout shows the fastest growth; (2) Among modifiable risk factors, obesity has become the leading driver of disease progression, with its contribution increasing significantly; (3) The disease burden exhibits a clear geographic shift, with middle-SDI countries replacing high-income regions as the new priority for intervention. This provides important implications for targeted interventions.

### Implications of All Available Evidence

The global burden of MSDs in postmenopausal women is escalating, driven primarily by population aging, the obesity epidemic, and advances in diagnosis and treatment. Importantly, this burden shows marked regional disparities. Our findings provide evidence-based guidance for health policymakers to develop tailored prevention and control strategies, addressing this major public health challenge.

## Methods

### Data Source

The data resources from the Global Burden of Disease Study (GBD) 2021 are accessible via the website of the Institute for Health Metrics and Evaluation (https://www.healthdata.org/). Employing a standardized analytical methodology, this database systematically evaluates the incidence, prevalence trends, and metrics such as disability-adjusted life years (DALYs) for 371 diseases and injuries across 204 countries and territories from 1990 to 2021. Additionally, it examines their attributable risk factors, including smoking, high body mass index (BMI), kidney dysfunction, and occupational ergonomic factors. Detailed research protocols and technical approaches can be found in the GBD thematic series publications.^22–24^ We extracted epidemiological indicators related to musculoskeletal disorders (MSDs), such as prevalence and DALY rates, and further conducted calculations for age-standardized rates (ASRs). The estimated annual percentage change (EAPC) was also computed. Postmenopausal status was approximated using an age cutoff of 55 years and older, consistent with prior epidemiological studies utilizing similar data sources.^25, 26^ This study did not require review by an institutional ethics committee, as the data were obtained from the publicly available GBD 2021 database. All data had undergone anonymization and aggregate processing, posing no potential threat to individual privacy.

### Definitions

The specific definitions of musculoskeletal disorders and their six subtypes, along with their corresponding ICD-10 and ICD-9 codes in the GBD system, are detailed in the Supplementary Materials.

### Predictive Analysis

This study employs the Autoregressive Integrated Moving Average (ARIMA) model to forecast the prevalence and DALYs trends of MSD over the next 29 years. The model integrates Autoregressive (AR), Integrated (I), and Moving Average (MA) components to identify trends and cyclical patterns in the data, thereby enabling the prediction of future changes based on past observations. Specifically, we selected historical data from 1990 to 2021 as the modeling foundation and used the auto. ARIMA() function to determine the optimal ARIMA model based on the Akaike Information Criterion and Bayesian Information Criterion.

### Statistical Analysis

We conducted a comprehensive analysis of the global and regional trends in MSD, prevalence, and DALYs from 1990 to 2021 to assess the overall burden of the disease. These metrics were expressed per 100,000 population, with 95% uncertainty intervals (UI). Additionally, we calculated two key indicators from the GBD study: the age-standardized rate (ASR) and the estimated annual percentage change (EAPC), as well as the age-standardized prevalence rate (ASPR) and the age-standardized DALY rate (ASDR). The ASR was used to adjust for variations in age structure, enabling more accurate comparisons of disease prevalence or mortality across different regions or time periods.^27^ The EAPC measures the average annual percentage change in the metric over time, providing insights into both the rate and direction of trends.^28^ The EAPC was calculated using the following formula:

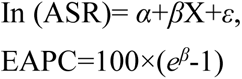

where X represents the calendar year, α denotes the intercept, β indicates the annual change in ln(ASR), and ε is the error term. The 95% confidence interval (CI) of the EAPC was also calculated based on this linear model^29^. A negative EAPC value with an upper 95% CI limit below zero indicates a declining trend in ASR, whereas a positive EAPC value with a lower 95% CI limit above zero suggests an increasing trend. If the 95% CI includes zero, it indicates a stable ASR trend. We further examined the relationship between Socio-demographic Index (SDI) and ASDR across the 21 GBD regions using Pearson’s correlation coefficient and locally weighted scatterplot smoothing. All analyses were performed using R software (version 4·2·3), with visualizations created using the ggplot2 package. Figures were subsequently compiled using Microsoft PowerPoint (version 365; Microsoft Corporation).

## Results

### The Global Burden of Musculoskeletal Disorders in Postmenopausal Women

From 1990 to 2021, the absolute disease burden indicators of MSDs in postmenopausal women, including case numbers and total DALYs, showed significant growth, with increases far exceeding those of age-standardized indicators (age-standardized prevalence rate [ASPR] and age-standardized DALY rate [ASDR]). In 2021, the number of MSD cases among postmenopausal women reached 476 million, with total DALYs of 45·0 million - representing dramatic increases of 126·6% and 122·2% respectively compared to 1990 levels. In contrast, the annualized growth rates (EAPC) of age-standardized rates (ASPR/ASDR) during the same period were only 0·15% and 0·10% (Figure 1A; Table 1).

**Figure 1.**
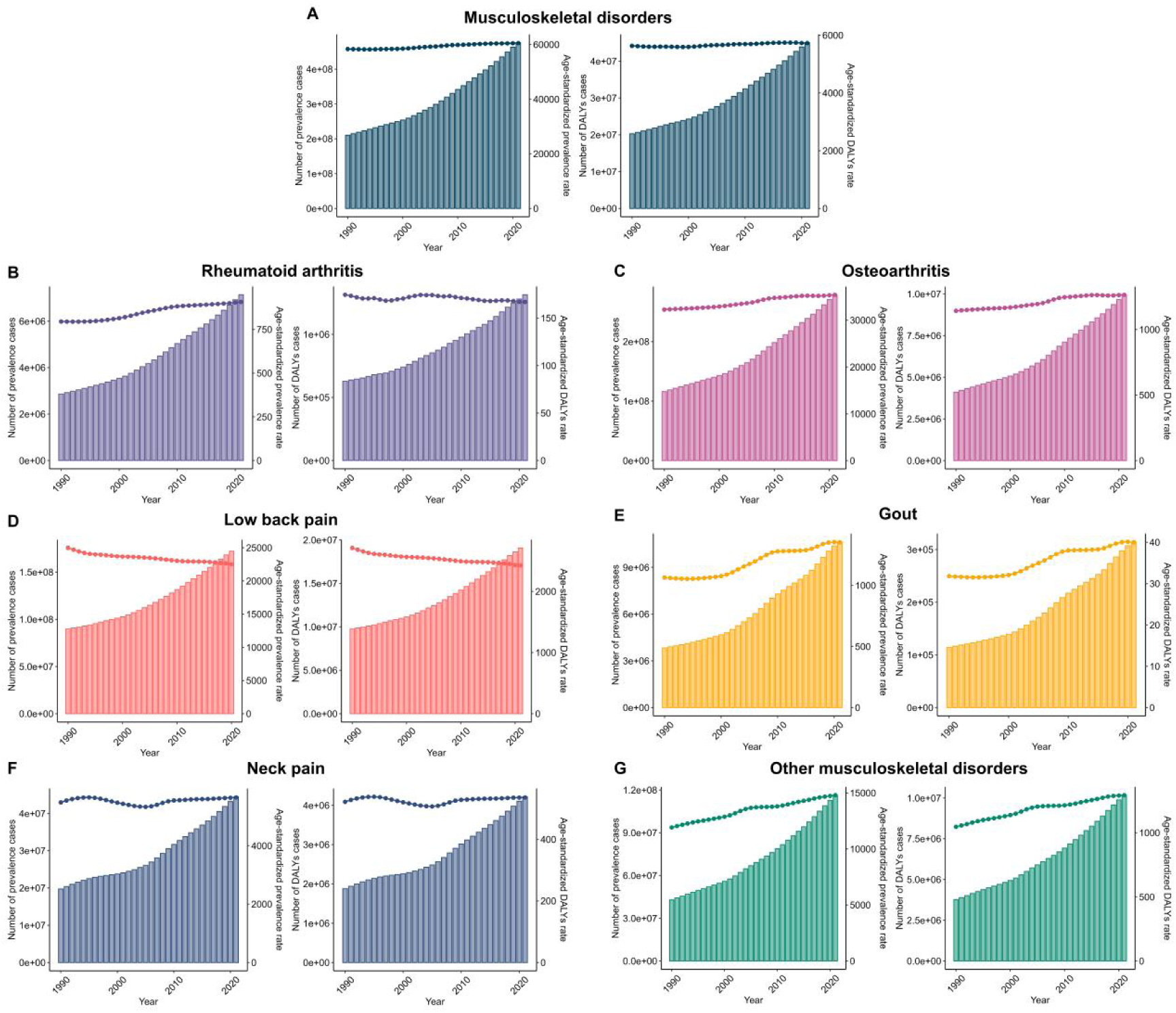
Prevalence and DALYs of musculoskeletal disorders (MSD) among postmenopausal women from 1990 to 2020.A): Musculoskeletal disorders (overall).B): Rheumatoid arthritis.C): Osteoarthritis.D): Low back pain.E): Gout.F): Neck pain.G): Other musculoskeletal disorders.Each panel shows the number of prevalent cases (left) and age-standardized DALYs per 100,000 population (right).

**Table 1.**
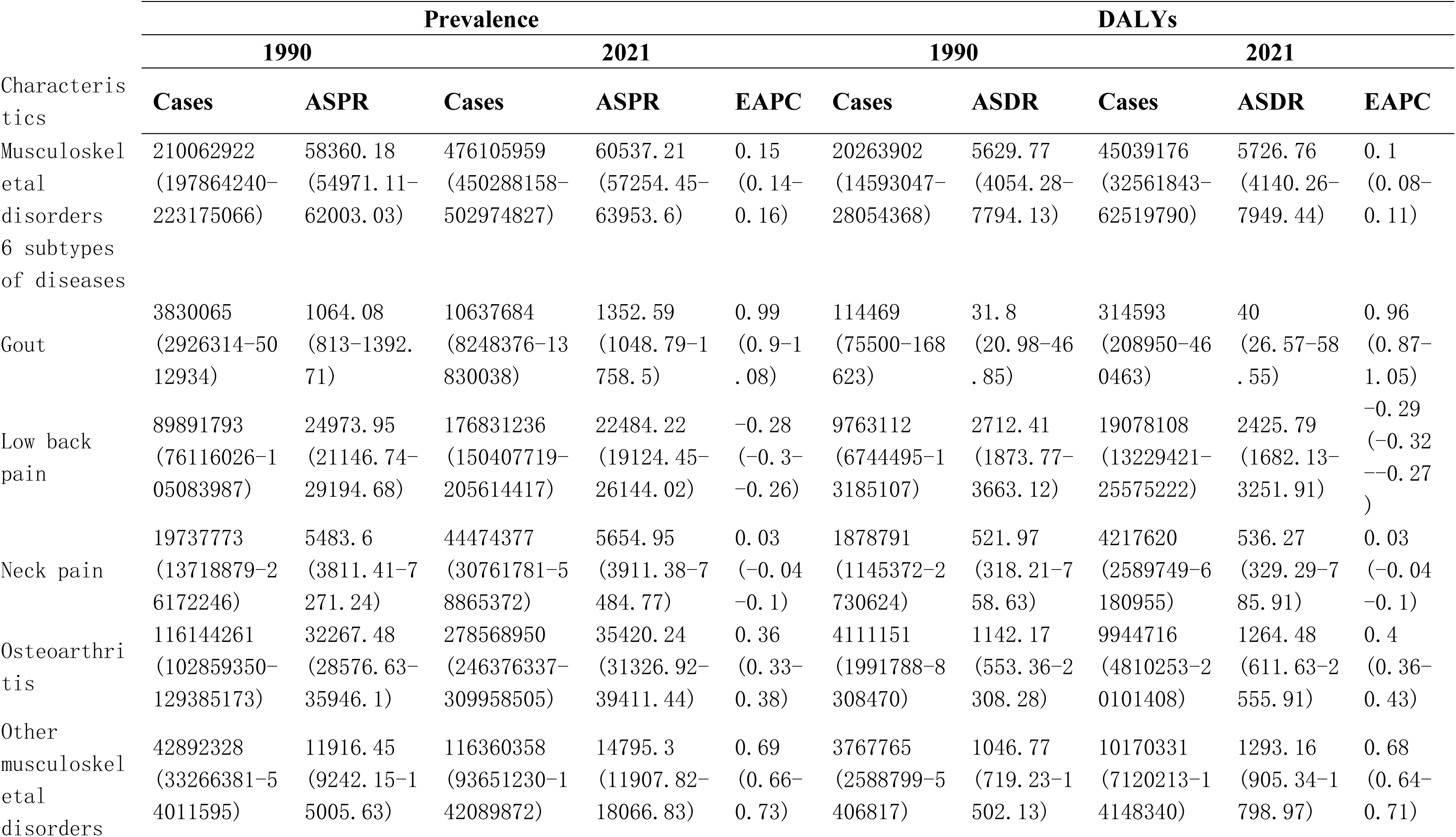
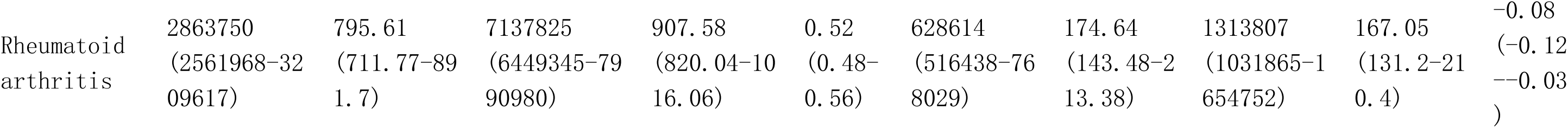
Global burden in Prevalence and DALYs of Musculoskeletal disorders among postmenopausal women from 1990 to 2021.

Analysis of six major MSD subtypes from 1990 to 2021 revealed distinct heterogeneous trends against the backdrop of significantly increased overall MSD burden. Osteoarthritis maintained its predominance, with 279 million cases in 2021, accounting for 58·6% of total MSD cases (point estimate), while demonstrating sustained growth in ASPR (EAPC=0·36) (Figure 1C, Table 1). Notably, although low back pain emerged as the most disabling subtype with 19·08 million DALYs and an ASDR of 2425·79 per 100,000, both its ASPR (EAPC=-0·28) and ASDR (EAPC=-0·29) showed significant declines (Figure 1D, Table 1), resulting in reduced contribution to overall MSD burden. Meanwhile, other musculoskeletal disorders (OMSD) exhibited substantial burden increase, surpassing osteoarthritis to become the second most significant disease subtype after low back pain (Figure 1G, Table 1).

Among the remaining MSD subtypes, rheumatoid arthritis and gout demonstrated relatively low levels across all evaluated indicators. Notably, rheumatoid arthritis exhibited a characteristic pattern of significant increase in ASPR (EAPC=0·52) coupled with a slight decrease in ASDR (EAPC=-0·08) (Figure 1B, Table 1). In contrast, although starting from a smaller baseline, gout showed the most pronounced upward trend (ASPR-EAPC=0·99; ASDR-EAPC=0·96) (Figure 1E, Table 1). The disease burden indicators for neck pain remained relatively stable (ASPR-EAPC=0·03; ASDR-EAPC=0·03) (Figure 1F, Table 1).

### Musculoskeletal Disease Burden in Postmenopausal Women by Age and SDI

The number of cases and DALYs in all age groups showed a significant increase (Figure S1A, Table 2), while the DALYs and case numbers decreased with advancing age groups (Figure S1B, Table 2). Notably, the age group over 95 exhibited the largest increase in both cases and DALYs, whereas the 75-79 age group showed the smallest increase (Table 2). Overall, from 1990 to 2021, the ASPR of MSD in postmenopausal women steadily rose globally across all age groups (Figure S2A, Table 2). The ASPR displayed a characteristic “inverted U-shaped” age distribution pattern (Figure S2B, Table 2), peaking in the 70-74 age group (6327·62). Meanwhile, the estimated annual percentage change (EAPC) of ASPR followed a “positive U-shaped” trend, with the fastest growth observed in the over-95 age group (EAPC=0·19) and the slowest in the 80-84 age group (EAPC=0·04) (Table 2).

**Table 2.**
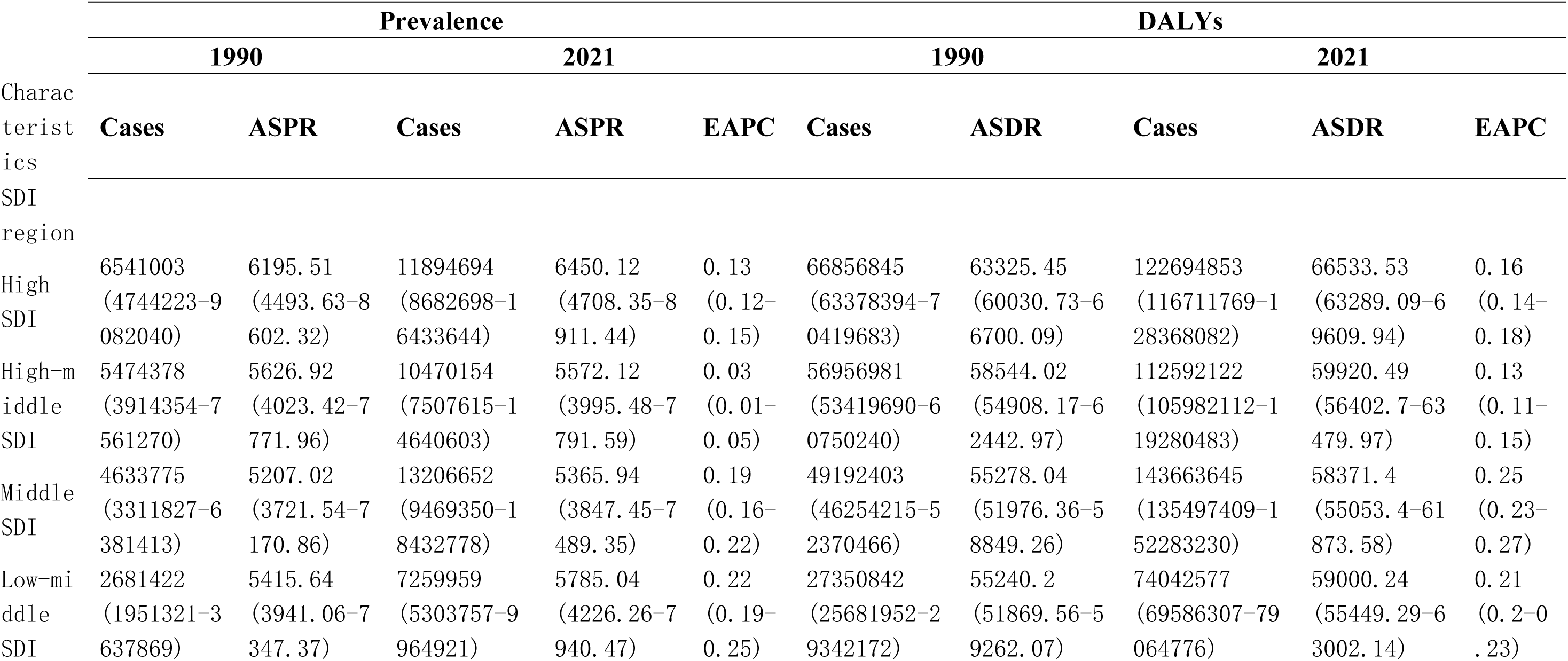
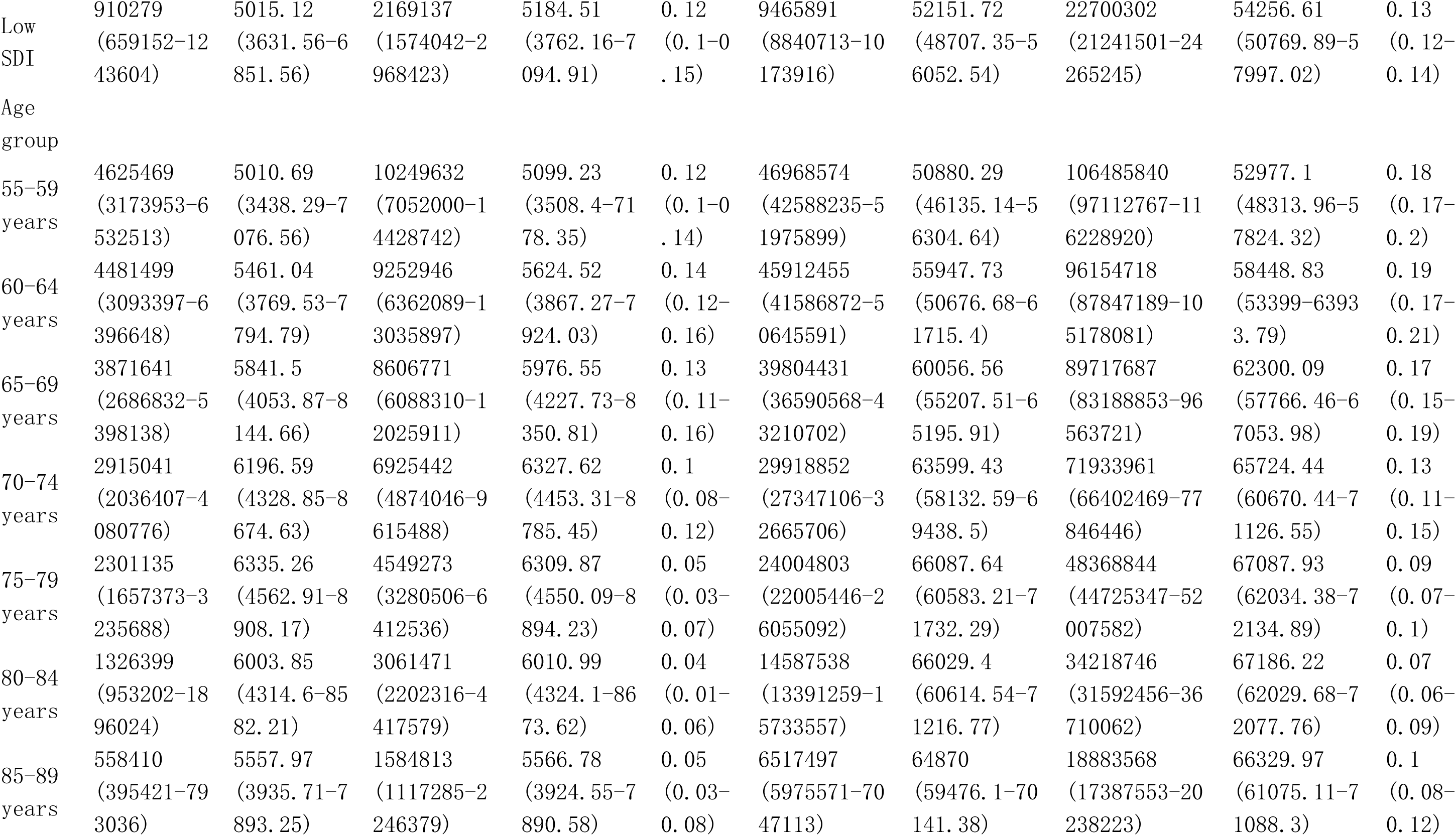
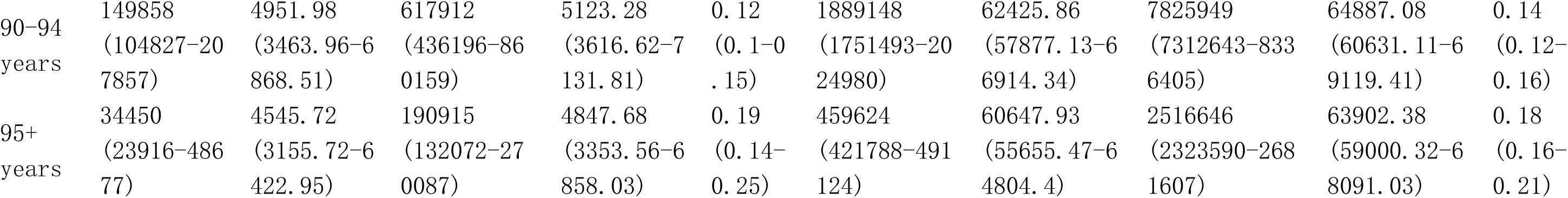
Global burden in Precalence and DALYs of Musculoskeletal disorders among postmenopausal women from 1990 to 2021 by 5 SDI regions and age groups.

In terms of disease severity, the age heterogeneity of ASDR was more pronounced. The “inverted U-shaped” peak of ASDR also appeared in the 75-84 age group, with its absolute value approximately ten times that of ASPR (Figure S2B, Table 2). Notably, the EAPC of ASDR exhibited an approximately “positive U-shaped” distribution, with the fastest growth in the 60-64 age group (EAPC=0·19) and the slowest growth in the 80-84 age group (EAPC=0·07) (Table 2). Further data analysis revealed distinct characteristics across three key age groups: 1) The middle-aged and early elderly group (55–64 years) had a relatively low ASPR, but its DALYs growth rate was significantly higher than the prevalence growth rate; 2) The elderly group (65–84 years) exhibited a typical “bimodal” pattern in both ASPR and ASDR; 3) The oldest-old group (85+ years) showed a “low baseline–high growth rate” trend, with the 95+ age subgroup having the largest increases in case numbers and DALYs, while maintaining consistently high EAPC levels (Table 2).

From 1990 to 2021, both the number of cases and DALYs in all SDI groups showed significant growth (Figure S1C, Table 2). Notably, the middle SDI region exhibited the most pronounced increase, with its growth in cases and DALYs surpassing that of the high SDI region in the past decade, making it the fastest-growing area for disease burden (Figure S1C-D, Table 2). In terms of disease prevalence, the high SDI region consistently maintained the highest ASPR (6450·12), but its estimated annual percentage change (EAPC = 0·13) was relatively moderate (Figure S2C, Table 2). In contrast, the middle SDI region (EAPC = 0·19) and the low-middle SDI region (EAPC = 0·22) showed significant upward trends in ASDR (Figure S2C, Table 2). Regarding disease severity, the spatial distribution of ASDR displayed a more complex gradient pattern. While high-SDI regions maintained the highest absolute ASDR values (66,533.53) (Figure S2D, Table 2), their growth rates remained stable (Figure S2C). Over the past 15 years, ASDR in low- and middle-SDI regions has shown significant acceleration (Figure S2C, Table 2). A deeper analysis of the burden disparity between high and low SDI regions reveals that the ASPR and ASDR in high SDI regions were 1·24 and 1·23 times those of low SDI regions, respectively. Meanwhile, the middle SDI region exhibited a distinct “high-growth, moderate-burden” pattern.

SDI showed a positive correlation with the overall ASDR of MSD (Figure 2A), but the association patterns between SDI and different disease subtypes varied significantly. Specifically, the ASDR of low back pain and neck pain exhibited distinct nonlinear relationships with SDI: the ASDR of low back pain declined slowly in low-to-middle SDI regions, then increased sharply in middle-to-high SDI regions, but decreased again at high SDI levels (Figure 2D); meanwhile, the ASDR of neck pain rose gradually at lower SDI levels before plateauing and declining after reaching middle SDI (Figure 2F). In contrast, the other four disease subtypes (e.g., osteoarthritis and rheumatoid arthritis) all demonstrated nonlinear positive correlations between ASDR and SDI (Figure 2B, C, E, G). As SDI increased, regional disparities in MSD-related ASDR widened, with high-SDI regions showing close alignment between observed ASDR and SDI-based theoretical predictions, indicating a significant association between SDI and ASDR of MSD in postmenopausal women.

**Figure 2.**
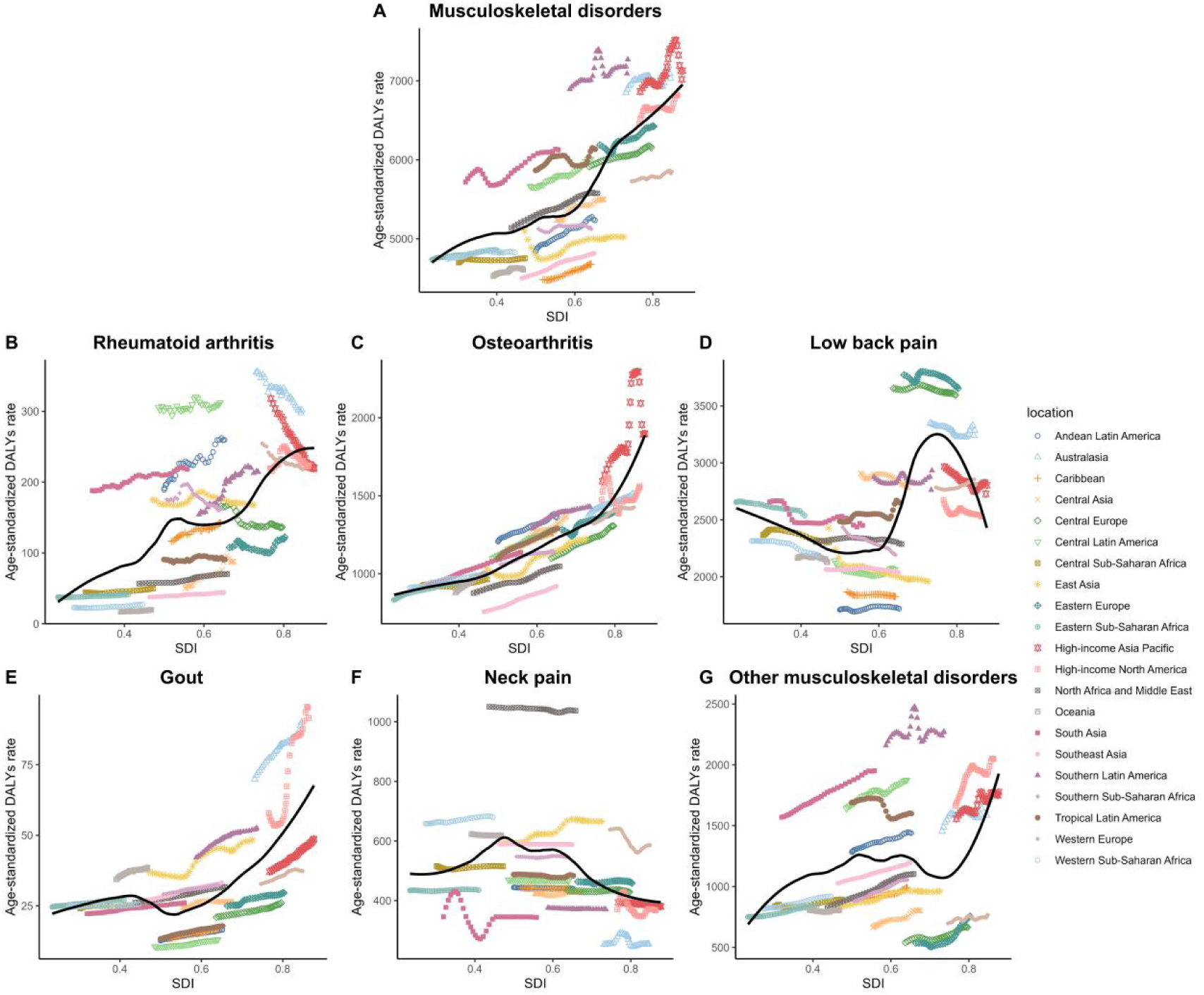
Age-standardized DALYs rates of musculoskeletal disorders (MSD) among postmenopausal women across different SDI regions in 2021. A): Musculoskeletal disorders (overall). B): Rheumatoid arthritis. C): Osteoarthritis. D): Low back pain. E): Gout. F): Neck pain. G): Other musculoskeletal disorders. Each panel shows age-standardized DALYs rates by SDI region, with different colors representing specific geographic locations.

### The burden of musculoskeletal disorders among postmenopausal women at regional and national levels

The GBD study data revealed that the ASPR and ASDR of MSD in postmenopausal women showed an upward trend across all studied regions (Figure 3,Figure S3, Table S2), but with significant regional heterogeneity. Based on disease burden levels and changing trends, these regions can be categorized into four distinct types. Oceania, sub-Saharan Africa, and the Caribbean exhibited a typical low-burden stable pattern. For example, Oceania had an ASPR of 4609·37 with an EAPC of 0·06 (Figure 3C, E; Table S1), and an ASDR of 52160·44 with an EAPC of 0·12 (Figure 3D, F; Table S2). In contrast, high-income regions such as Western Europe, high-income North America, and Australasia displayed a high-burden improving pattern. Although their ASPR (Western Europe: 5849·32) (Figure 3C, Table S2) and ASDR (Western Europe: 62047·32) (Figure 3D, Table S2) remained high, they had effectively controlled disease growth (Western Europe ASPR-EAPC = 0·06 (Figure 3E, Table S2); ASDR-EAPC=0·1 (Figure 3F, Table S2)). Notably, high-income Asia Pacific, North Africa and the Middle East, and Latin America have emerged as prominent high-burden worsening regions, with not only the highest global levels of ASPR (high-income Asia Pacific: 7127·69) (Figure 3C, Table S2) and ASDR (high-income Asia Pacific: 72297·43) (Figure 3D, Table S2) but also significant EAPC growth trends (ASPR-EAPC = 0·22) (Figure 3E; Table S2); ASDR-EAPC=0·32(Figure 3F;Table S2). Additionally, middle-income regions such as East Asia and Southeast Asia exhibited transitional characteristics as moderate-burden transforming regions. Their ASPR (East Asia: 5023·11) (Figure 3C, Table S2) and ASDR (56818·27) (Figure 3D, Table S2) were at intermediate levels, but with persistently rising EAPC trends (ASPR-EAPC = 0·15 (Figure 3E, Table S2); ASDR-EAPC = 0·28 (Figure 3F; Table S2).

**Figure 3.**
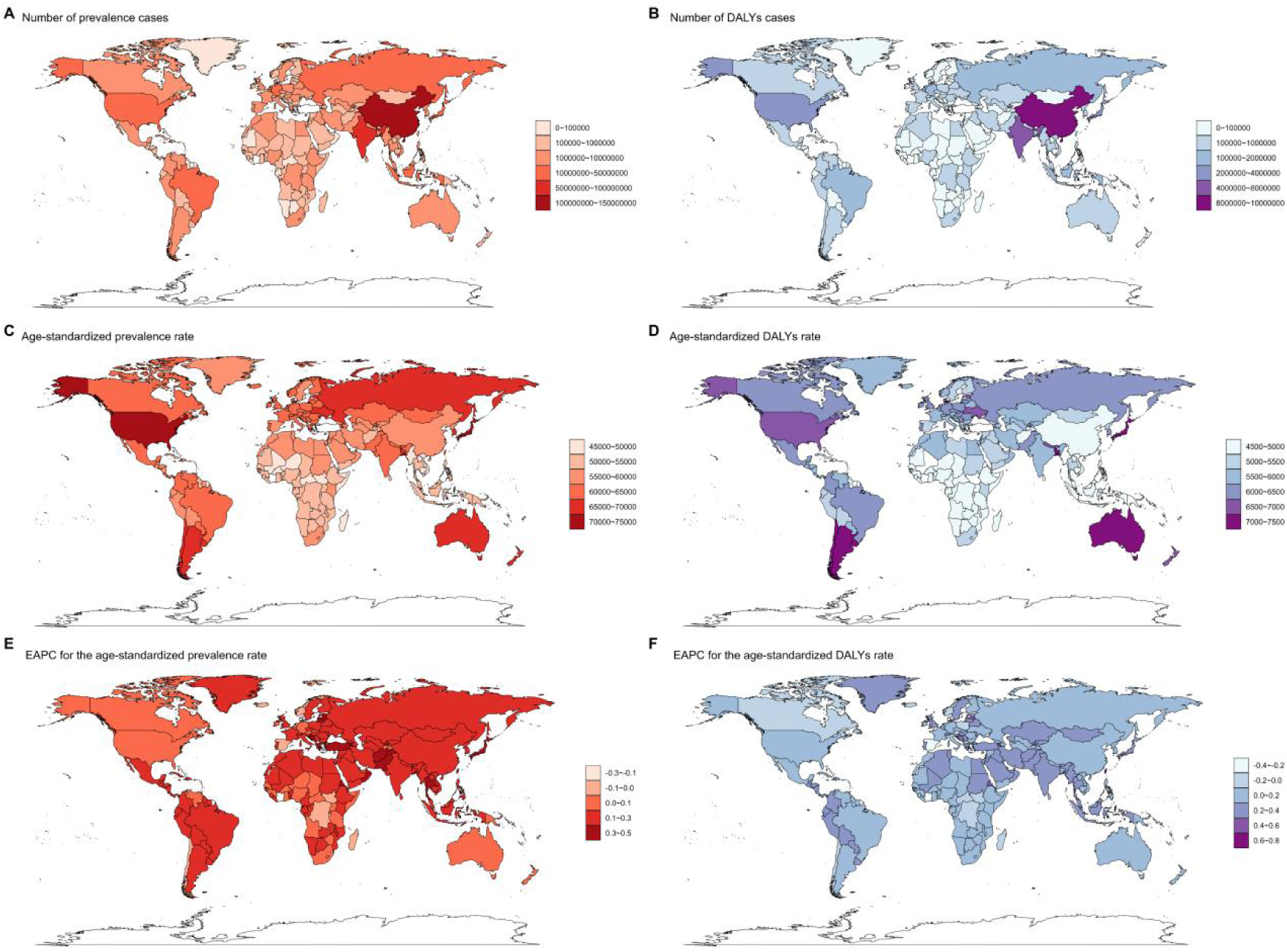
Global epidemiological burden of musculoskeletal disorders (MSD) among postmenopausal women in 2020. A): Number of prevalent cases.B): Number of DALYs cases.C): Age-standardized prevalence rate per 100,000 population.D): Age-standardized DALYs rate per 100,000 population.E): EAPC (annual percentage change) for age-standardized prevalence rate.F): EAPC for age-standardized DALYs rate.Color gradients represent data ranges, with darker shades indicating higher values or rates.

A more granular country-level analysis revealed internal heterogeneity within the regional classification framework. Specifically, Papua New Guinea (ASPR 50099·28; EAPC=0·10) and Haiti (ASPR 48758·02; EAPC=0·08) exhibited slow growth consistent with regional characteristics, while Burundi showed significant negative growth (ASPR-EAPC=−0·21), with its ASDR decline rate (EAPC=−0·30) even exceeding that of ASPR. Meanwhile, Denmark (ASPR 63831·64; EAPC=−0·15) and Spain (ASDR-EAPC=−0·34) displayed negative growth trends, whereas the U.S. continued to experience rising ASPR (70266·28; EAPC=0·08), and Canada’s ASDR growth rate approached zero (EAPC=−0·05). Japan (ASPR 73324·06; EAPC=0·37) and South Korea (ASDR 6441·37; EAPC=0·19) showed accelerated deterioration, while Taiwan, China (ASDR-EAPC=0·62) and Saudi Arabia (EAPC=0·28) exhibited extreme growth. In contrast, medium-burden countries faced particularly notable common challenges during their transitional phase. Thailand (ASPR-EAPC=0·32; ASDR-EAPC=0·27) and North Korea (ASDR 5388·29; EAPC=0·15), while China’s ASPR (56682·18; EAPC=0·28) and ASDR (4992·53; EAPC=0·14) all exhibited the typical characteristic of increasing disease burden alongside economic growth (Table S2).

The geographical distribution differences of six diseases are as follows: rheumatoid arthritis shows stable high-burden regions but rapid increases in some countries, with East Asia, South Asia, and Western Europe having the highest absolute burden, while Australasia and Central Latin America rank highest after age standardization; Ireland and other European countries remain high-incidence, Equatorial Guinea grows fastest (EAPC=2·67), and Norway and Japan show declining trends(Figure S7-8,Table S6). Osteoarthritis is relatively evenly distributed globally but dominated by high-income countries, with Japan and South Korea having the heaviest burden and African nations (e.g., Madagascar, ASPR 22515·25) the lightest; Equatorial Guinea and Armenia show significant growth among middle- and low-income countries, while Israel and the U.S. decline(Figure S9-10,Table S7). Low back pain showed the highest ASDR in Eastern Europe (e.g., Ukraine, ASDR 4001·11). While most regions globally exhibited declining disease burdens, the UK and China’s Taiwan region bucked this trend. Denmark demonstrated the most significant decline (EAPC = -0.81)(Figure S11-12,Table S8). Neck pain was most prevalent in North Africa and the Middle East, while being least common in Australasia. China (EAPC=0·52) showed the fastest growth rate, whereas Spain experienced a sharp decline (EAPC=-2·48)(Figure S13-14,Table S9). Gout burden was heaviest and rising most rapidly in upper-middle-income countries, particularly in North America (ASDR-EAPC 2·3) and East Asia(ASDR-EAPC 1·16), with the United States and China showing marked increases. Sub-Saharan Africa had the lowest burden(Figure S15-16,Table S10). Other musculoskeletal disorders grow fastest in Eastern Europe (ASPR-EAPC=1·05) and North Africa and Middle East (ASPR-EAPC=1·01), while tropical Latin America declines; Bangladesh and Argentina bear the heaviest burden, Eritrea has the sharpest increase (EAPC=3·89), and Burundi shows the steepest decline (EAPC=-1·49)(Figure S17-18,Table S11).

### The impact of different attributions on DALYs of musculoskeletal disorders

From 1990 to 2021, DALYs attributable to various risk factors all showed an increasing trend (Figure S4B), while the trends in ASDR varied (Figure S4A). High body mass index (BMI) has emerged as the primary risk factor affecting musculoskeletal health in postmenopausal women. Regarding the burden of gout, BMI-related DALYs increased by 246·4%, with ASDR rising from 9·47 to 15·02 (EAPC=1·75), while kidney dysfunction-related burden showed relatively slower growth (EAPC=0·72). For low back pain cases, BMI-related DALYs increased by 178·2%, with ASDR reaching 344·44 (EAPC=0·83). Notably, although occupational ergonomic factor-related DALYs increased by 93·3%, ASDR showed a downward trend (EAPC=-0·27). For osteoarthritis, BMI-related DALYs surged by 197·7%, with ASDR reaching 262·38 (EAPC=1·02), highlighting the significant impact of obesity on this condition. In contrast, smoking-related disease burden showed positive changes: ASDR for low back pain decreased significantly (EAPC=-1·13), and rheumatoid arthritis ASDR declined to 6·18 (EAPC=-1·51) (Table S1).

### Projection of musculoskeletal disease burden: prevalence and risk factor-attributable DALYs in postmenopausal women (2022–2050)

The analysis of MSD burden based on the 2022–2050 projection model indicates a continued upward trend with significant epidemiological heterogeneity. Aggregate projections show that the global number of cases will increase from 489 million in 2022 to 835 million by 2050, while the ASPR is expected to rise from 60,600·52 to 62,432·34 per 100,000 (Figure S5, Table S3). Notably, the uncertainty intervals of projections widen substantially over time, reflecting the high sensitivity of long-term forecasts to changes in population structure and disease risk factors. Subtype analysis reveals divergent trends among the six major categories of musculoskeletal disorders: osteoarthritis exhibits the most pronounced growth (case count +70·9%, ASPR +6·7%), while low back pain and neck pain show declining prevalence rates (ASPR -28·5%, ASPR -2·7%) (Table S3).

The burden of musculoskeletal disorders in postmenopausal women exhibits unique epidemiological characteristics. Among postmenopausal women, the number of DALYs related to musculoskeletal disorders is projected to increase from 45·1 million in 2022 to 66·4 million in 2050, while the ASDR remains stable (5726·76 per 100,000). Osteoarthritis (high BMI) and low back pain (high BMI) show the most significant growth in disease burden, with low back pain (high BMI) expected to become the highest-burden subtype by 2050 (5·53 million cases, ASDR 441·56 per 100,000). Meanwhile, the burden of occupational ergonomic factor-related low back pain continues to rise, projected to increase from 2·47 million cases in 2022 to 4·83 million cases in 2050. Notably, smoking-related low back pain and rheumatoid arthritis exhibit declining ASDR trends, with the predicted uncertainty interval for rheumatoid arthritis (smoking) turning negative after 2040. The ASDR for gout (kidney dysfunction) and gout (high BMI) remains relatively stable (Figure S6, Table S4-5).

## Discussion

Postmenopausal women constitute a crucial demographic in aging societies, generally enjoying longer life expectancy than their male counterparts,^9, 10^ yet bearing a disproportionately heavier disease burden.^30, 31^ This study presents the first systematic examination of the epidemiological distribution characteristics of MSD among postmenopausal women across 204 countries and territories from 1990 to 2021. The findings reveal a persistent upward trend in MSD incidence within this population, establishing it as one of the primary disease burdens threatening the health of middle-aged and elderly women - a trend consistent with observations in the general population.^32, 33^ The paradoxical coexistence of rising absolute disease burden metrics (case numbers, DALYs) with declining age-standardized rates (ASPR/ASDR) underscores population aging as the predominant driver of disease burden escalation. Significant epidemiological variations emerge across disease subtypes, with osteoarthritis dominating prevalence trends while low back pain carries the heaviest disease burden, and OMSD demonstrates rapid growth - indicating the necessity for differentiated prevention and control strategies tailored to distinct MSD burden profiles in postmenopausal women. Geospatial analysis identifies four distinct regional distribution patterns (low-stable, high-improving, high-worsening, and medium-transitional), reflecting underlying structural contradictions: the persistent challenge of population aging and chronic disease management in developed regions; asynchronous lifestyle transitions and healthcare system development in middle-income countries; and statistical biases from insufficient disease surveillance sensitivity in low-income areas. More critically, the widening intra-regional disparities (e.g., US EAPC=0·08 vs Canada EAPC=-0·05) demonstrate that economic development alone can no longer explain disease burden variations, necessitating multidimensional interpretation through healthcare system architecture and primary prevention efficacy.

Our findings reveal that the burden of MSD in postmenopausal women exhibits significant dual heterogeneity in terms of age and socioeconomic status. In the age distribution dimension, a characteristic “dual U-shaped” phenomenon was observed: ASPR and ASDR showed an inverted U-shaped curve, peaking in the elderly group aged 65–84, while EAPC displayed a positive U-shaped distribution, with the most pronounced growth rate in the oldest age group over 85. This pattern suggests two critical disease progression pathways: a hidden risk of rapid progression in the elderly (where DALYs growth significantly outpaces ASPR) and an explosive accumulation of disease burden in the very old age group. In terms of socioeconomic development, first, high-SDI regions still maintain the highest absolute values of ASPR (6450·12 per 100,000) and ASDR (66533·53 per 100,000). Second, the study found that middle-SDI regions have become the fastest-growing areas in terms of disease burden.

This distribution pattern may reflect three key issues: First, the paradox between advanced healthcare systems and chronic disease management. Despite abundant medical resources in high-SDI regions, population aging and the cumulative effects of chronic diseases sustain a persistently high disease burden.^34, 35^ Second, rapid socioeconomic transitions may lead to the swift adoption of high-calorie diets and sedentary lifestyles in middle-SDI regions,^36^ with risk factors such as obesity accelerating the epidemiological shift of MSD in postmenopausal women. Third, limited healthcare accessibility in low-SDI regions may result in a systemic underestimation of disease severity. Additionally, as SDI increases, interregional disparities in ASDR show an expanding trend, highlighting the socioeconomic inequalities in the current global MSD prevention and control system for postmenopausal women. More notably, age and socioeconomic factors exhibit significant interaction effects: in high-SDI regions, medical advancements prolong survival but exacerbate disease accumulation, whereas in low- and middle-SDI regions, resource shortages accelerate the growth of burden in the elderly population. This disease evolution pattern under dual pressures poses new challenges to the current global MSD prevention and control framework, urgently calling for the establishment of precision strategies that account for both age-specific and socioeconomic disparities.

BMI is a major risk factor for various MSDs, and our study highlights its significant role in postmenopausal women. Over the past 30 years, MSD conditions attributable to high BMI—including osteoarthritis, gout, and low back pain—have each shown a substantial increase (EAPC>0·8), consistent with trends observed in previous studies.^33, 37, 38^ This rise far exceeds that of other risk factors, aligning with the global surge in obesity rates during the same period. Therefore, it is recommended to prioritize personalized weight management programs for postmenopausal women, including community-based health promotion initiatives or specialized weight management clinics. Regular physical activity should be encouraged to enhance overall bodily function. Early screening should be integrated with health education, with a focus on improving awareness of musculoskeletal health—particularly the importance of proper nutrition, exercise therapy, and preventive medical care. Public health policies must systematically incorporate these interventions to address the ongoing rise in MSD burden and ultimately improve the overall health of this high-risk population.

Looking ahead to the next 30 years, the global disease burden of MSD in postmenopausal women is projected to continue rising. The increase in case numbers will primarily be driven by population aging and the trend of earlier onset of musculoskeletal disorders, while the significant growth in DALYs alongside stable ASDR suggests that the disease burden in this population is heavily influenced by demographic shifts. With increasing global life expectancy, the cumulative burden of MSD will further escalate without effective early intervention measures. Notably, modifiable risk factors such as high BMI and occupational ergonomic factors will become key variables influencing the MSD burden. Projections indicate that osteoarthritis and low back pain associated with high BMI will exhibit the most substantial rise in disease burden in the coming decades, while the number of occupational-related low back pain cases may even double. This trend underscores the need for public health strategies to prioritize interventions targeting obesity and risk-related factors. Furthermore, the MSD burden among postmenopausal women will continue to intensify in the coming years. Although existing clinical interventions can partially mitigate disease impact—such as the use of biologics to control rheumatoid arthritis progression,^39, 40^ urate-lowering therapy to manage gout symptoms,^41^ and cognitive behavioral therapy to alleviate low back pain^42^—these treatments remain insufficient to fundamentally curb the growing disease burden trend.

Several methodological constraints should be acknowledged. First, substantial heterogeneity in data quality across regions was observed, particularly due to incomplete disease surveillance systems in low- and middle-income countries, potentially compromising the accuracy of global MSD burden estimates. Second, while the GBD study employs sophisticated modeling approaches to address data gaps, residual uncertainties in model parameters may propagate and amplify underlying data limitations. Most notably, our use of 55 years as the menopausal age threshold introduces two critical issues: (1) reduced sensitivity in identifying women with premature menopause (<45 years), and (2) insufficient consideration of well-documented racial and geographical variations in natural menopause timing. This classification may systematically exclude menopausal women aged 45-54 years from analyses, leading to underestimation of burden in this subgroup. These limitations collectively impact the generalizability and interpretation of our findings.

## Data Availability

All data produced in the present study are available upon reasonable request to the authors
All data produced in the present work are contained in the manuscript
All data produced are available online at https://ghdx.healthdata.org/gbd-2021

## Contributors

This study was conceived by GL, LD, and JH, who also interpreted the results and contributed to manuscript writing. QF, CZ, and CG were responsible for data analysis, while DZ and HF participated in data collection and verification. BC contributed to literature search and manuscript revision. XX, CJ, PD, and YG provided critical review of the intellectual content. All authors reviewed and approved the final manuscript.

## Acknowledgments

This research was supported by the National Natural Science Foundation of China (No. 82402849, to P.Ding; No. 82272474, to Y.Gao). This research was conducted as part of the Global Burden of Diseases, Injuries, and Risk Factors Study 2021. We thank all individuals who contributed to this study, including the relevant collaborative group at the Institute for Health Metrics and Evaluation.

## 1. Definitions

### Musculoskeletal disorders

These diseases encompass severe functional impairments and fatalities caused by rheumatoid arthritis, osteoarthritis, low back pain, neck pain, gout, and various other musculoskeletal disorders.

### Gout

Gout is a rheumatic disease characterized by the deposition of monosodium urate crystals in joint synovial fluid and other tissues, triggering inflammatory responses. The GBD study adopts the 1977 diagnostic criteria for primary gout established by the American College of Rheumatology.

### Low Back Pain

Low back pain is defined as pain occurring in the posterior region of the body between the lower margin of the twelfth ribs and the inferior gluteal folds (with or without referred pain to one or both lower limbs), lasting for at least one day.

### Neck Pain

Neck pain is defined as pain localized within the anatomical region extending from the occiput to the first thoracic vertebra (with or without referred pain to the upper limbs), persisting for a minimum of one day.

### Rheumatoid Arthritis

Rheumatoid arthritis is a systemic autoimmune disorder that causes joint pain, swelling, and deformity, potentially accompanied by systemic manifestations. The reference case definition for Rheumatoid arthritis is based on the 1987 criteria set by the American College of Rheumatology.

### Osteoarthritis

Osteoarthritis, the most prevalent form of arthritis, involves chronic inflammation, degeneration, and structural changes affecting the entire joint.

### Other Musculoskeletal Disorders

This heterogeneous residual category encompasses a spectrum of muscle, bone, and ligament conditions not covered by the five GBD-defined musculoskeletal disorders (rheumatoid arthritis, osteoarthritis, low back pain, neck pain, and gout).

### The Socio-demographic Index (SDI)

SDI is a comprehensive indicator used to measure the economic and social development status of a country or region, with its value range set between 0 and 1. Here, 0 represents the lowest level of development, while 1 signifies the highest level of development. Based on SDI data, this study categorizes countries into five distinct tiers to analyze disparities in health outcomes, thereby providing crucial insights for academic research and policy-making.

## 2. ICD-10 and ICD-9 Codes

The specific ICD-10 and ICD-9 codes for musculoskeletal disorders and their six subtypes are provided in Table as follows:

| Cause | ICD10 | ICD9 |
| --- | --- | --- |
| Musculoskeletal disorders | G54.2-G54.4, I27.1, L93-L93.2, M00-M10.19, M10.3-M25.9, M28-M32.9, M34-M49, M49.2-M59, M61-M72.4, M72.8-M73, M74-M79.676, M79.8- M87.09, M87.2-M89.59, M89.7-M95.9, M97-M99.9, N09 | 274-274.9, 353.1-353.4, 416.1, 446-446.9, 710-710.2, 710.5, 724.9, 726-727.9, 729-730.39, 730.7-739.9, V13.4-V13.59, V17.7-V17.89, V43.6-V43.8, V58.64-V58.65, V58.78, V77.5, V82.1-V82.2 |
| Rheumatoid arthritis | M05-M05.9, M08-M09.8 | 714-714.9 |
| Osteoarthritis | M16-M18.9 | 715-715.98 |
| Neck pain | G54.2, M47.011-M47.013, M47.11-M47.13, M47.21, M47.23, M47.811-M47.813, M47.892-M47.894, M48.01-M48.03, M48.12-M48.14, M48.21-M48.23, M48.31-M48.33, M48.41-M48.43, | 353.2, 721.0-721.1, 722.0, 722.71, 722.81, 722.91, 723-723.9 |
|  | M48.51-M48.53,<br>M49.81-M49.83, M50-M50.93,<br>M53.0-M53.1, M53.81-M53.83,<br>M54.01-M54.03,<br>M54.11-M54.13, M54.2, M54.81,<br>M99.01, M99.11, M99.21,<br>M99.31, M99.41, M99.51,<br>M99.61, M99.71, M99.81 |  |
| Gout | M10-M10.19, M10.3-M10.9,<br>M1A00X0-M1A9XX1 | 274-274.9, 712.0-712.09 |
| Other<br>musculoskeletal<br>disorders | G54.3, I27.1, L93-L93.2,<br>M00-M03.6, M06-M07.69,<br>M11-M15.9, M19-M19.93,<br>M20-M25.9, M30-M32.9,<br>M34-M36.8, M40-M43.9,<br>M45-M47.01, M47.014,<br>M47.02-M47.10, M47.14,<br>M47.2-M47.20, M47.24,<br>M47.8-M47.81, M47.814,<br>M47.819-M47.891, M47.895,<br>M47.9-M48.00, M48.04, M48.1<br>M48.11, M48.15, M48.2-M48.20,<br>M48.24, M48.3-M48.30,<br>M48.34, M48.4-M48.40, M48.44,<br>M48.5-M48.50, M48.54,<br>M48.8-M49, M49.2-M49.80,<br>M49.84, M49.89, M51-M51.04,<br>M51.1-M51.14, M51.2-M51.24,<br>M51.3-M51.34, M51.4-M51.44,<br>M51.8-M51.84, M51.9, M53,<br>M53.2, M53.8-M53.80, M53.84,<br>M53.9-M54.00, M54.04, M54.1-<br>M54.10, M54.14, M54.6-M54.8,<br>M54.89-M54.9, M61-M63.89,<br>M65-M68.8, M70-M72.4, M72.8- | 353.3, 416.1, 446-446.9,<br>710-710.2, 710.5-712, 712.1-713.8, 716-721, 721.2,<br>721.4-721.41, 721.5-722,<br>722.1, 722.11-722.31,<br>722.39-722.51, 722.6-722.70,<br>722.72, 722.8 722.80, 722.82,<br>722.9-722.90, 722.92,<br>724-724.01, 724.09- 724.1,<br>724.4-724.5, 724.8-724.9,<br>726-727.9, 729-730.39,<br>730.7-739.9 |
|  | M73, M75-M77.9,<br>M79-M79.676, M79.8-M87.09,<br>M87.2-M89.59, M89.7-M95.9,<br>M99-M99.00,<br>M99.02,M99.05-M99.10,M99.12,<br>M99.15-M99.20,<br>M99.22,M99.25-M99.30,M99.32,<br>M99.35-M99.40,<br>M99.42,M99.45-M99.50,M99.52,<br>M99.55-M99.60,<br>M99.62,M99.65-M99.70,M99.72,<br>M99.75-M99.80, M99.82,<br>M99.85-M99.9 |  |

## 3. Supplementary Figure

**Figure S1.**
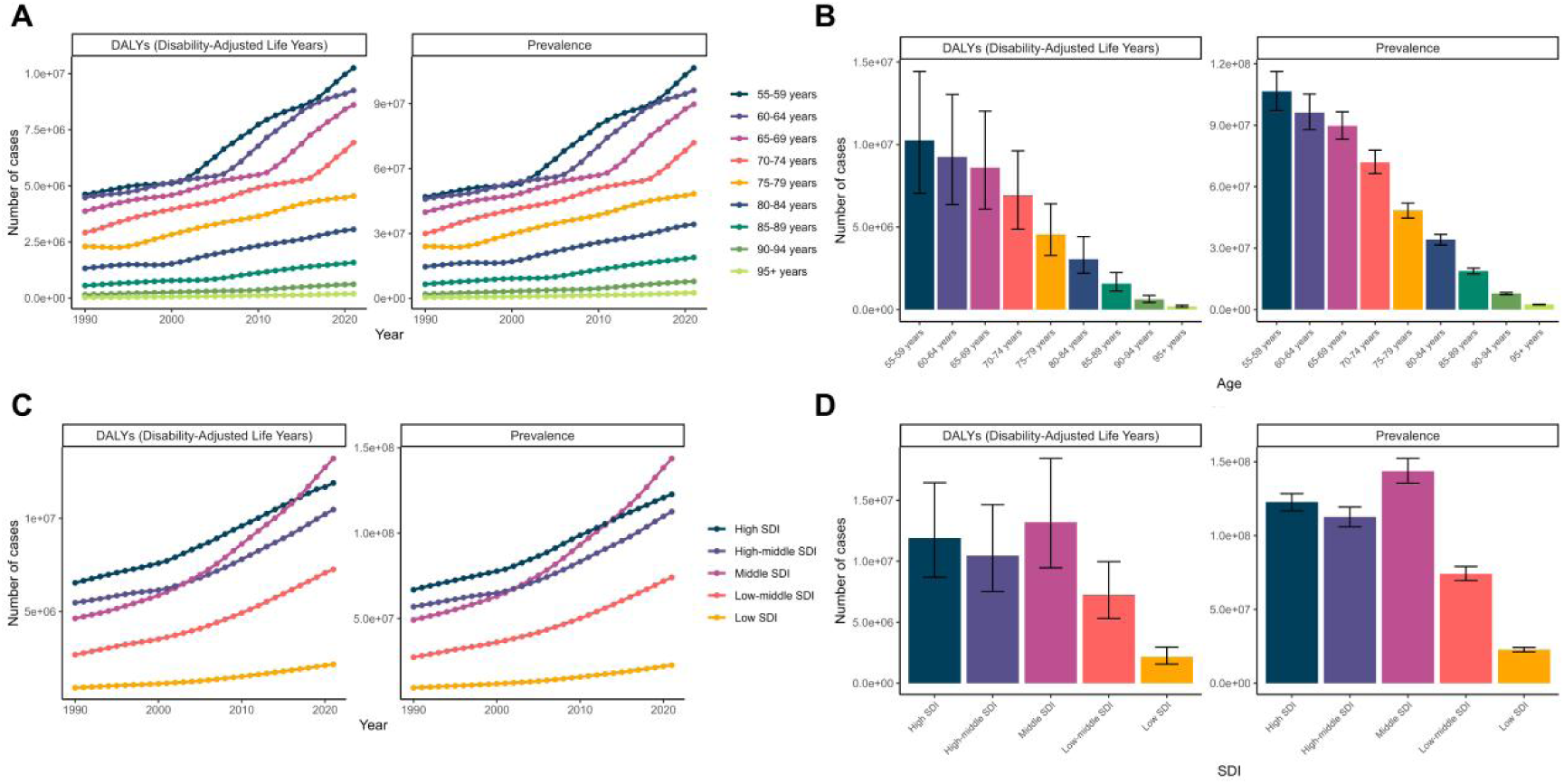
Epidemiological burden of musculoskeletal disorders (MSD) among postmenopausal women globally, by age group and SDI region from 1990 to 2021. A: Age-specific trends in DALYs (left) and prevalence (right) over time (1990–2021). B: Age-specific DALYs (left) and prevalence (right) in 2021, with 95% uncertainty intervals. C: SDI-specific trends in DALYs (left) and prevalence (right) over time (1990–2021). D: SDI-specific DALYs (left) and prevalence (right) in 2021, with 95% uncertainty intervals.

**Figure S2.**
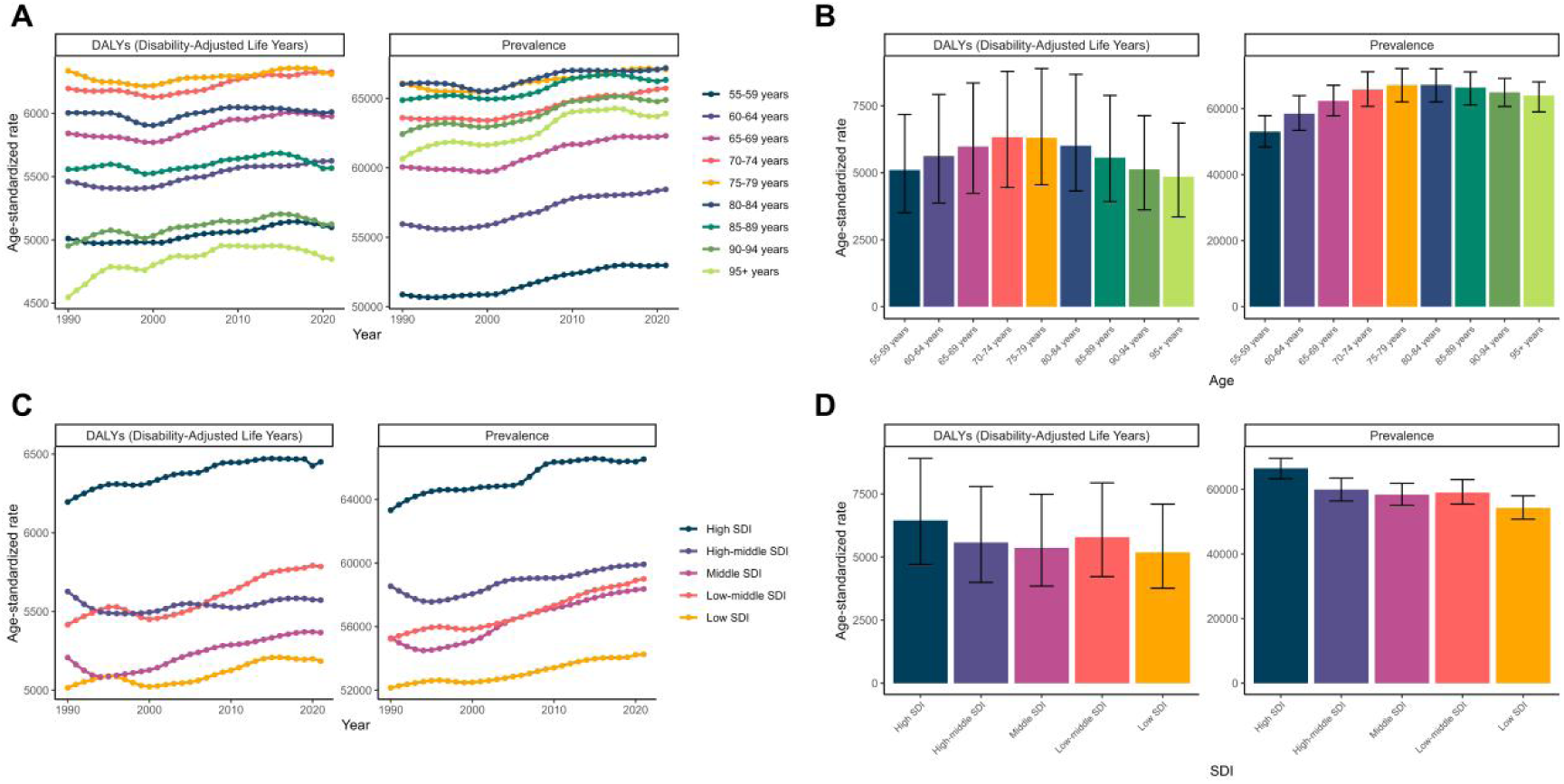
Epidemiological burden of musculoskeletal disorders (MSD) among postmenopausal women globally, by age group and SDI region from 1990 to 2021. A: Age-specific trends in age-standardized DALYs (left) and prevalence (right) over time (1990–2021). B: Age-specific age-standardized DALYs (left) and prevalence (right) in 2021, with 95% uncertainty intervals. C: SDI-specific trends in age-standardized DALYs (left) and prevalence (right) over time (1990–2021). D: SDI-specific age-standardized DALYs (left) and prevalence (right) in 2021, with 95% uncertainty intervals.

**Figure S3.**
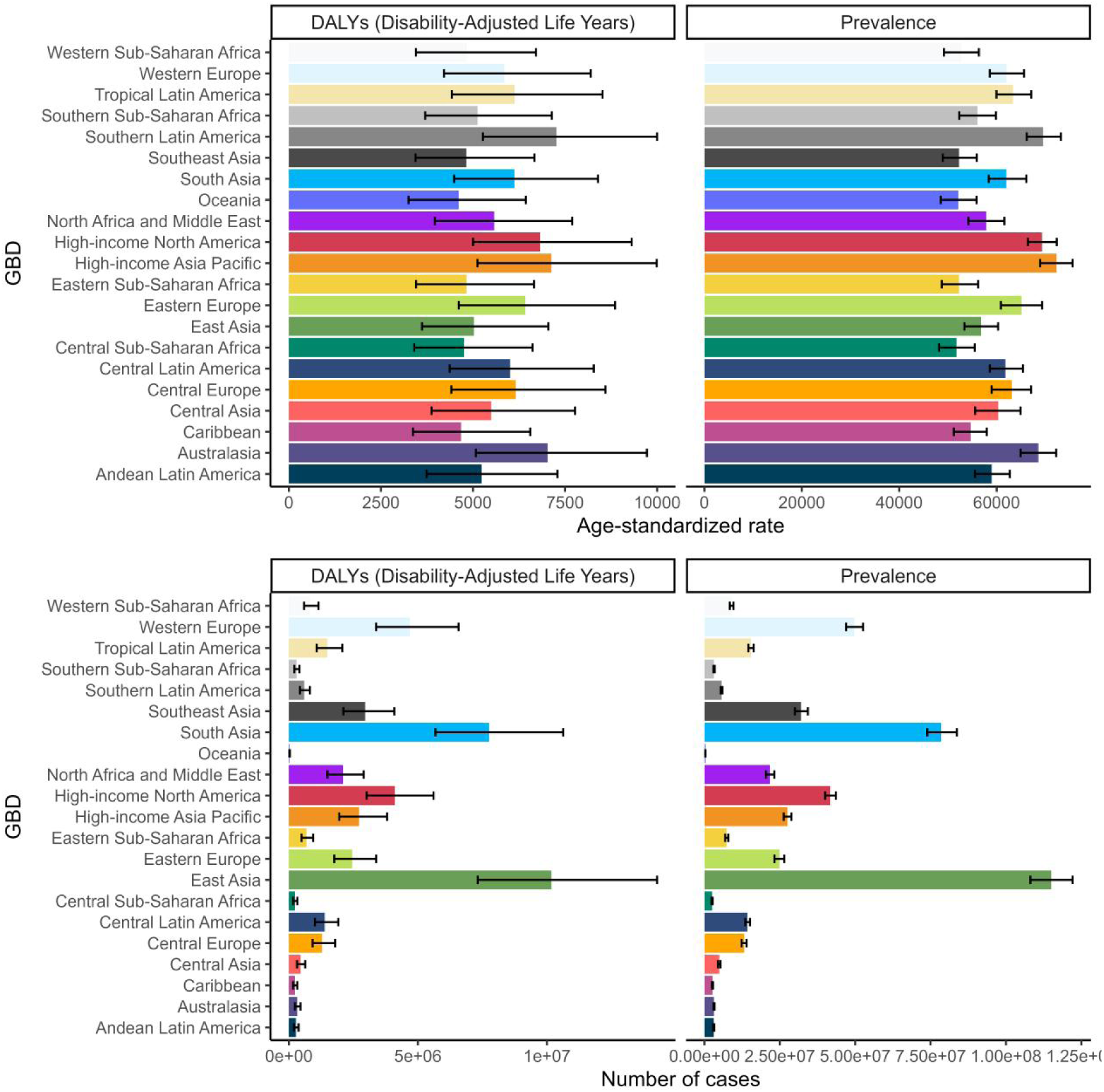
Epidemiological burden of MSD among postmenopausal women across 21 global regions in 2021.

**Figure S4.**
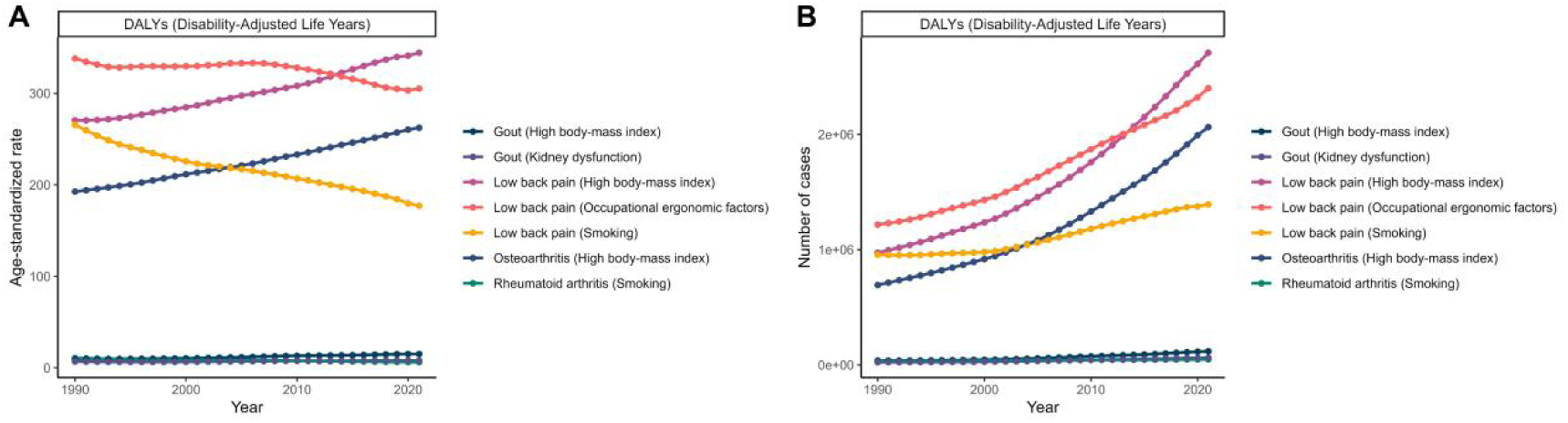
Impact of risk factors on musculoskeletal disorders (MSD) among postmenopausal women from 1990 to 2021.A): Age-standardized DALYs rates attributed to specific risk factors. B): Number of DALYs cases attributed to specific risk factors. Different colors represent DALYs associated with specific risk factors, including high body-mass index, kidney dysfunction, occupational ergonomic factors, and smoking.

**Figure S5.**
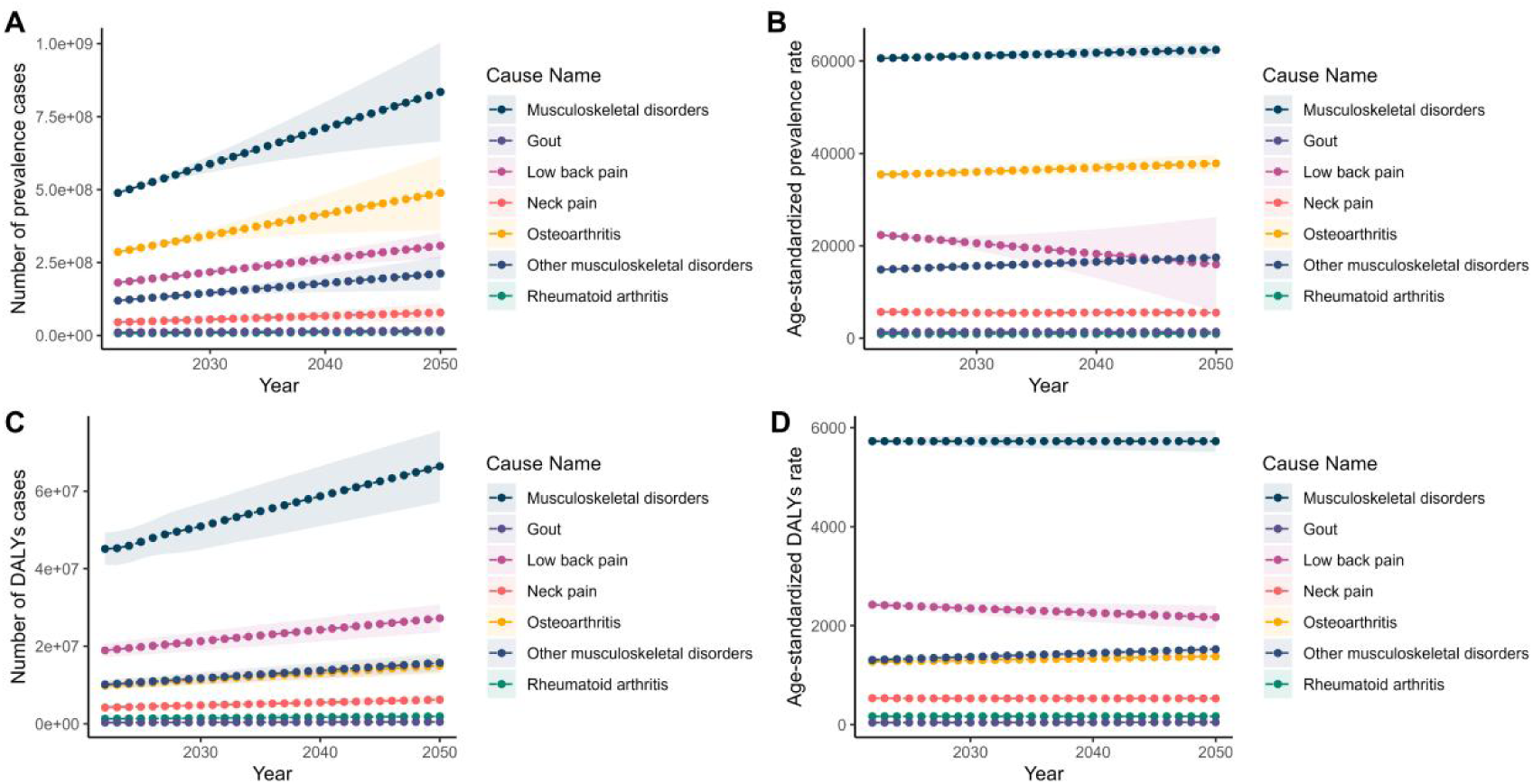
Projected burden of musculoskeletal disorders (MSD) among postmenopausal women from 2022 to 2050. A): Number of prevalent cases. B): Age-standardized prevalence rate. C): Number of DALYs cases. D): Age-standardized DALYs rate. Different colors represent specific MSD categories, including musculoskeletal disorders (overall), gout, low back pain, neck pain, osteoarthritis, other musculoskeletal disorders, and rheumatoid arthritis.

**Figure S6.**
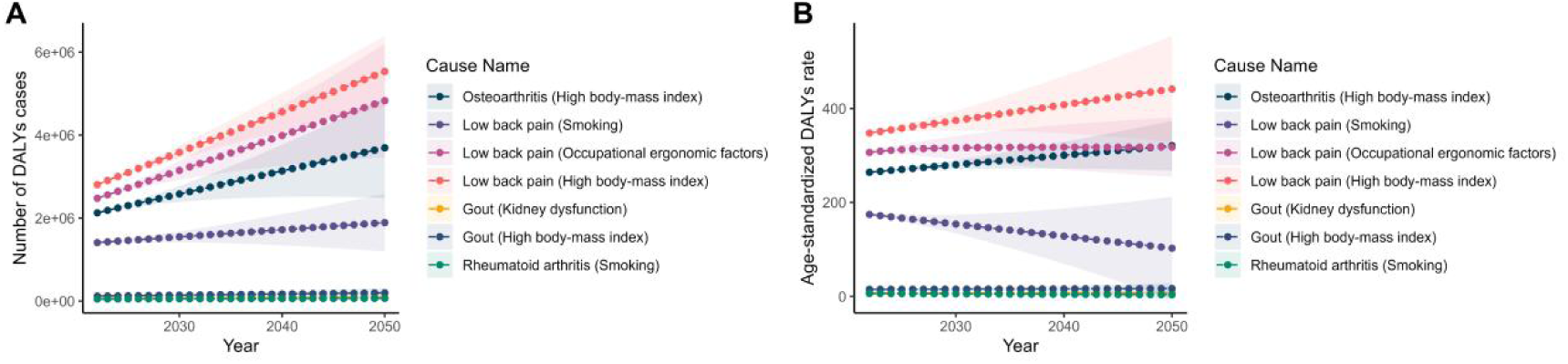
Projected burden of musculoskeletal disorders (MSD) among postmenopausal women from 2022 to 2050, attributed to specific risk factors. A): Number of DALYs cases. B): Age-standardized DALYs rate. Different colors and line styles represent DALYs attributed to specific risk factors, including high body-mass index, smoking, occupational ergonomic factors, and kidney dysfunction.

**Figure S7.**
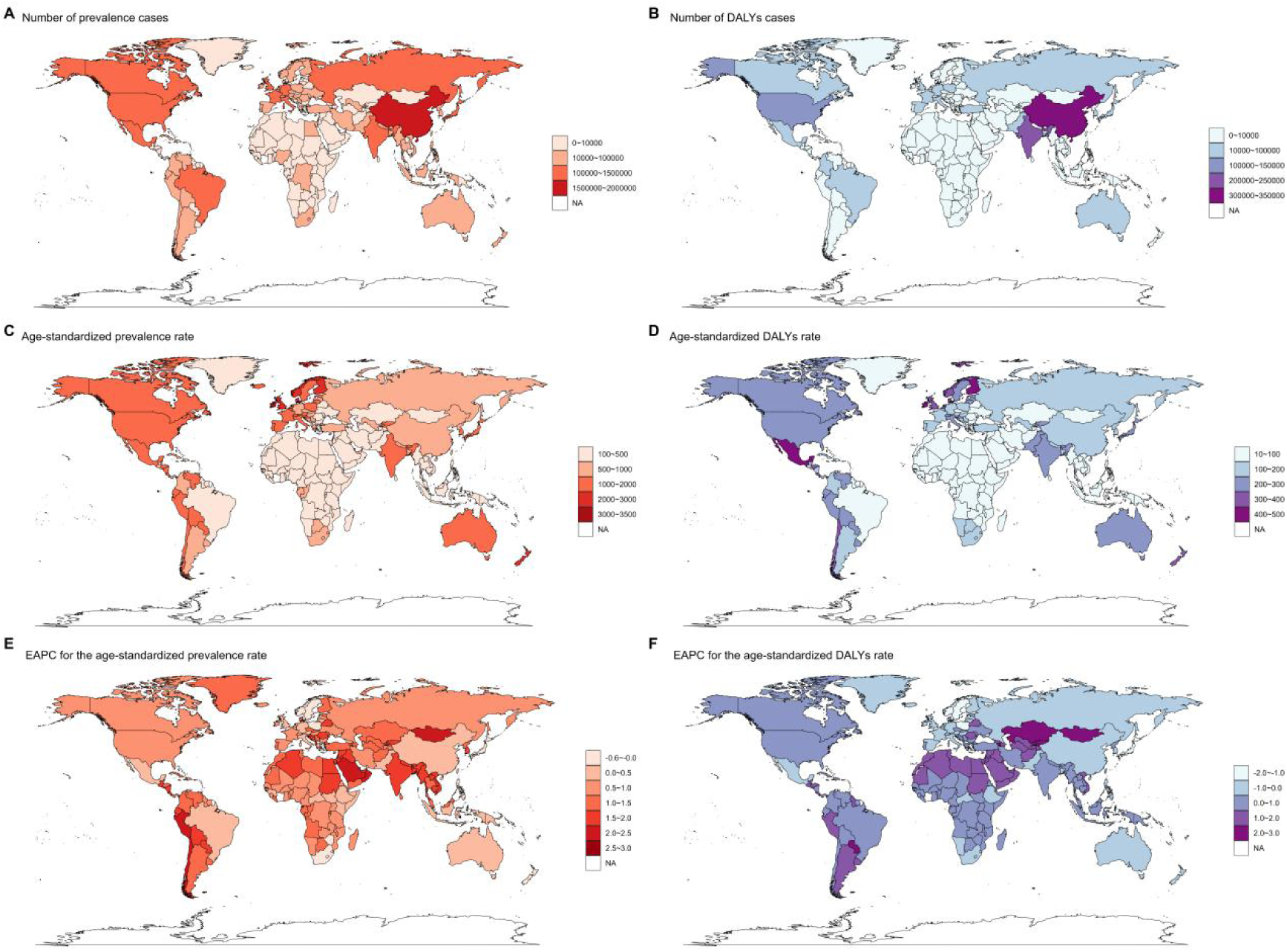
Global geographical distribution characteristics of rheumatoid arthritis. A): Geographical distribution of the number of prevalent cases. B): Geographical distribution of DALYs. C): Geographical distribution of age-standardized prevalence rates. D): Geographical distribution of age-standardized DALY rates. E): Geographical distribution of EAPC in age-standardized prevalence rates. F): Geographical distribution of EAPC in age-standardized DALY rates.

**Figure S8.**
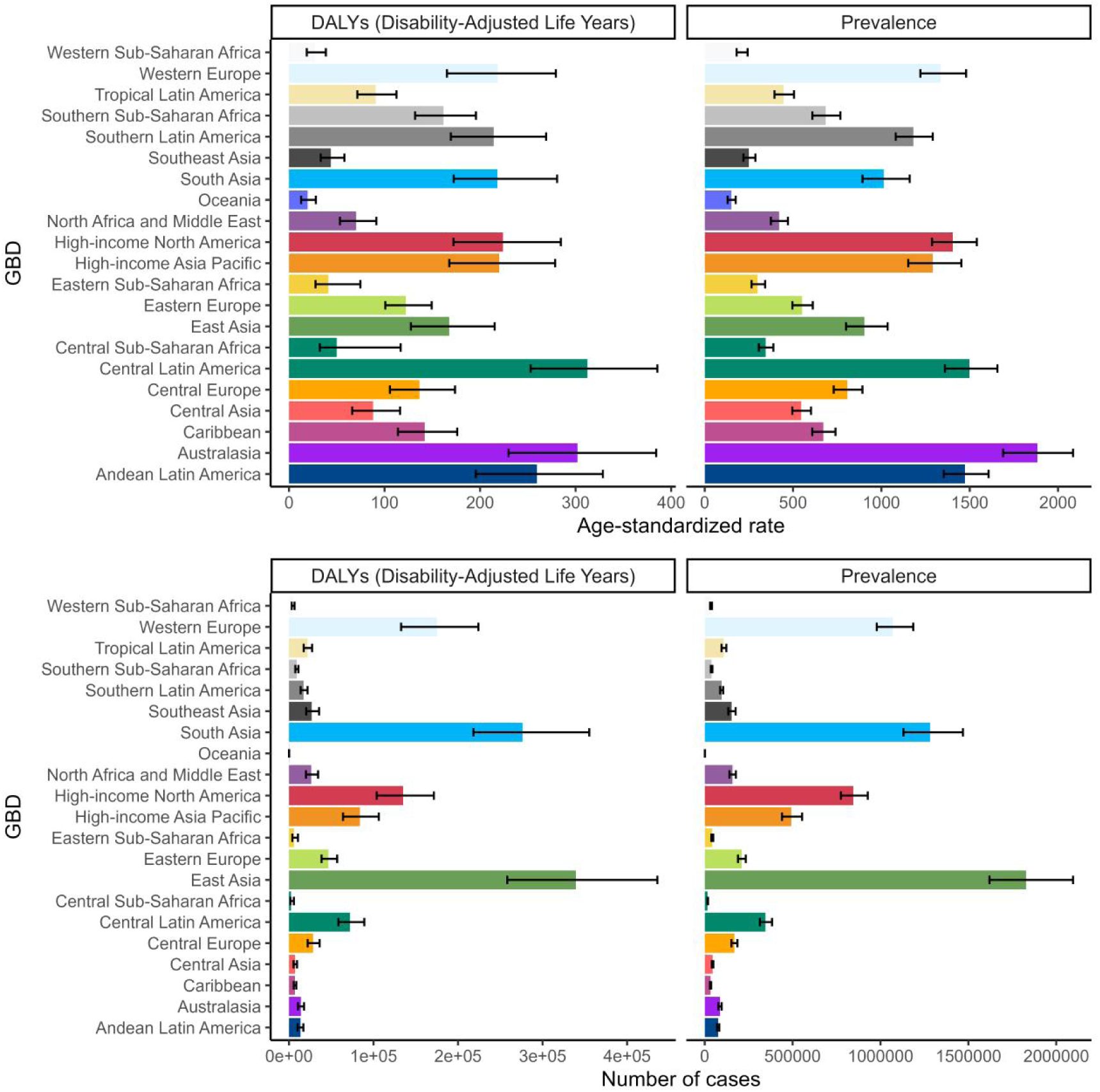
The global epidemiological burden of rheumatoid arthritis across 21 regions in 2021.

**Figure S9.**
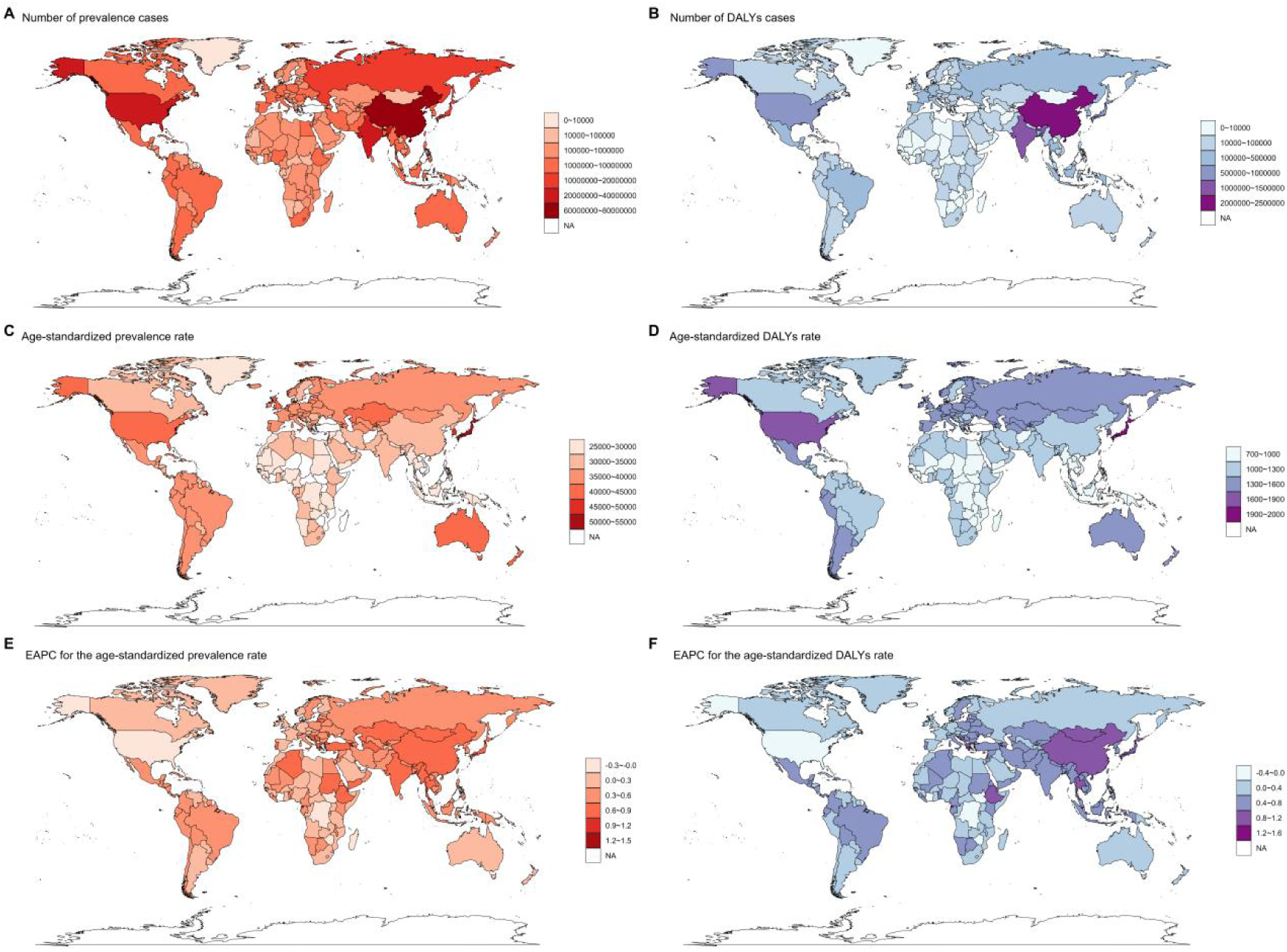
Global geographical distribution characteristics of osteoarthritis. A): Geographical distribution of the number of prevalent cases. B): Geographical distribution of DALYs. C): Geographical distribution of age-standardized prevalence rates. D): Geographical distribution of age-standardized DALY rates. E): Geographical distribution of EAPC in age-standardized prevalence rates. F): Geographical distribution of EAPC in age-standardized DALY rates.

**Figure S10.**
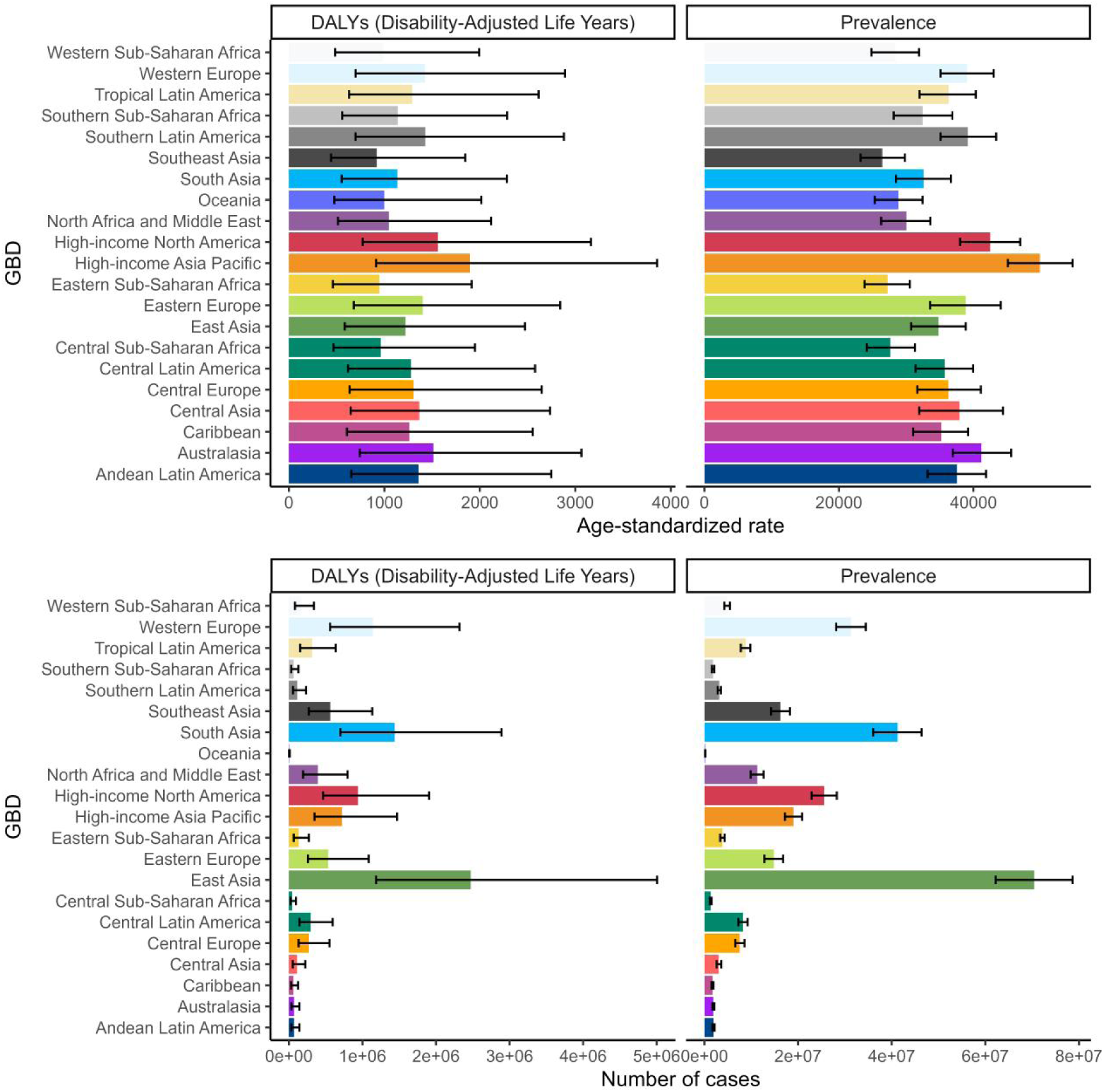
The global epidemiological burden of osteoarthritis across 21 regions in 2021.

**Figure S11.**
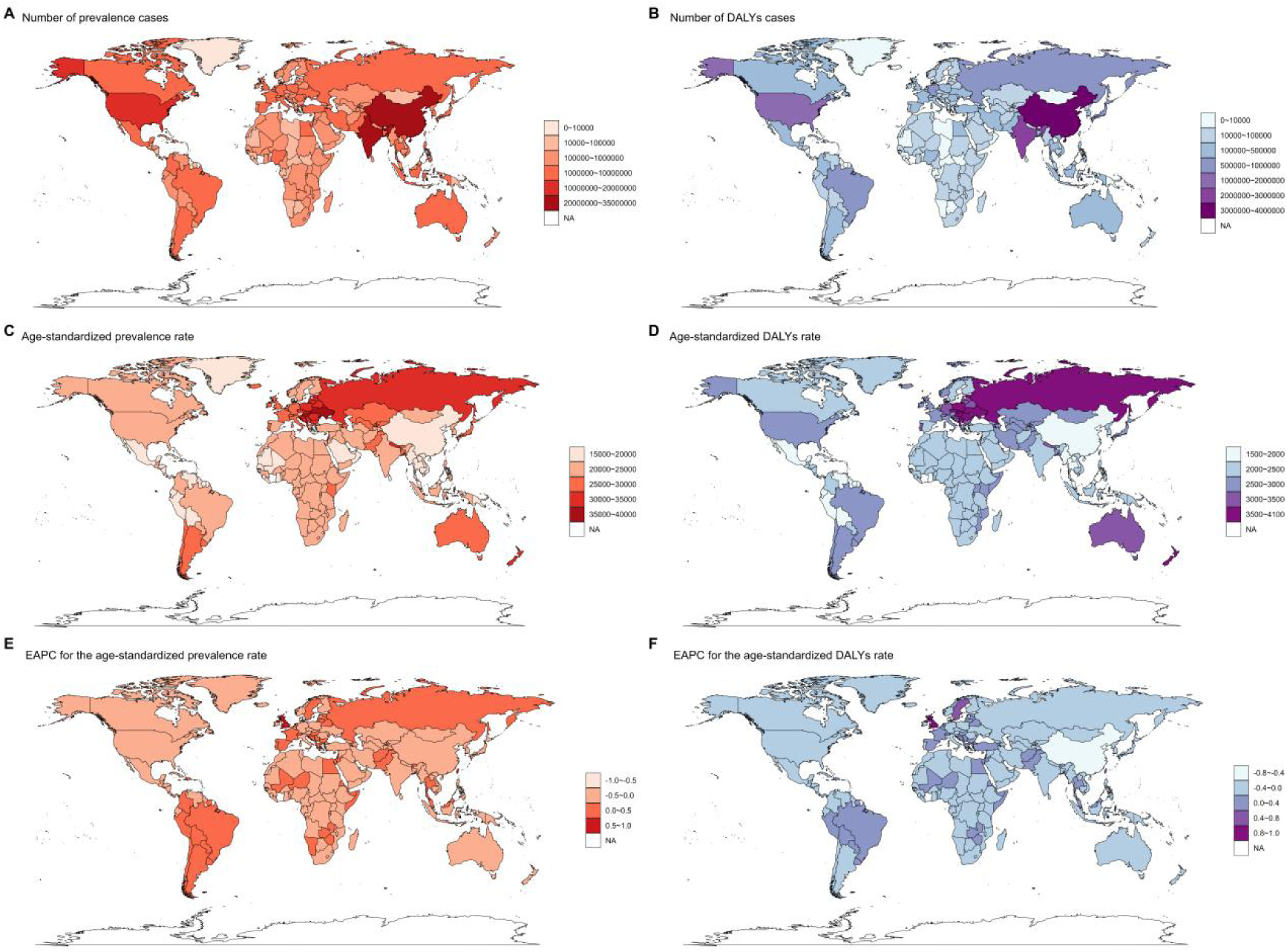
Global geographical distribution characteristics of low back pain. A): Geographical distribution of the number of prevalent cases. B): Geographical distribution of DALYs. C): Geographical distribution of age-standardized prevalence rates. D): Geographical distribution of age-standardized DALY rates. E): Geographical distribution of EAPC in age-standardized prevalence rates. F): Geographical distribution of EAPC in age-standardized DALY rates.

**Figure S12.**
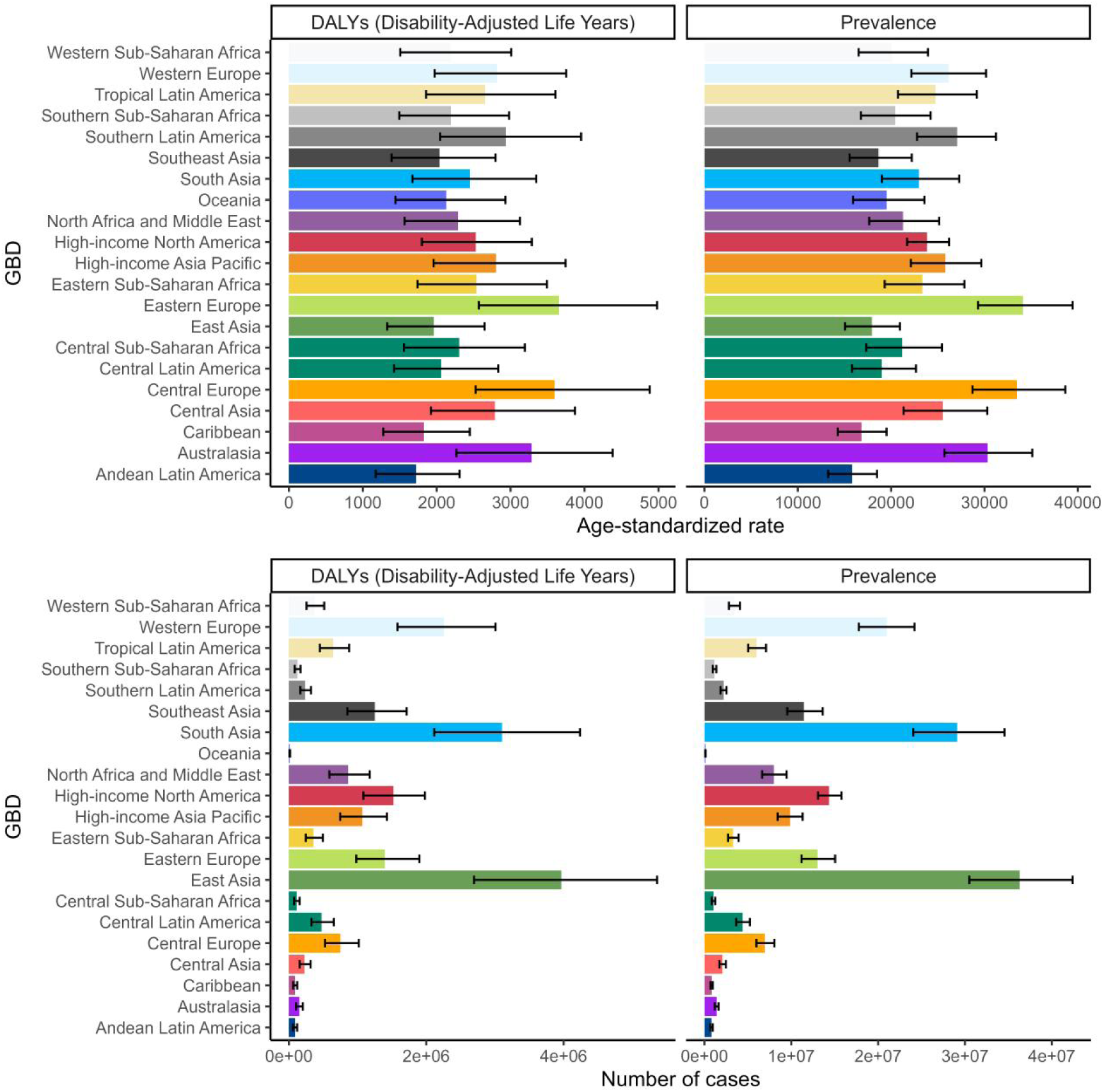
The global epidemiological burden of low back pain across 21 regions in 2021.

**Figure S13.**
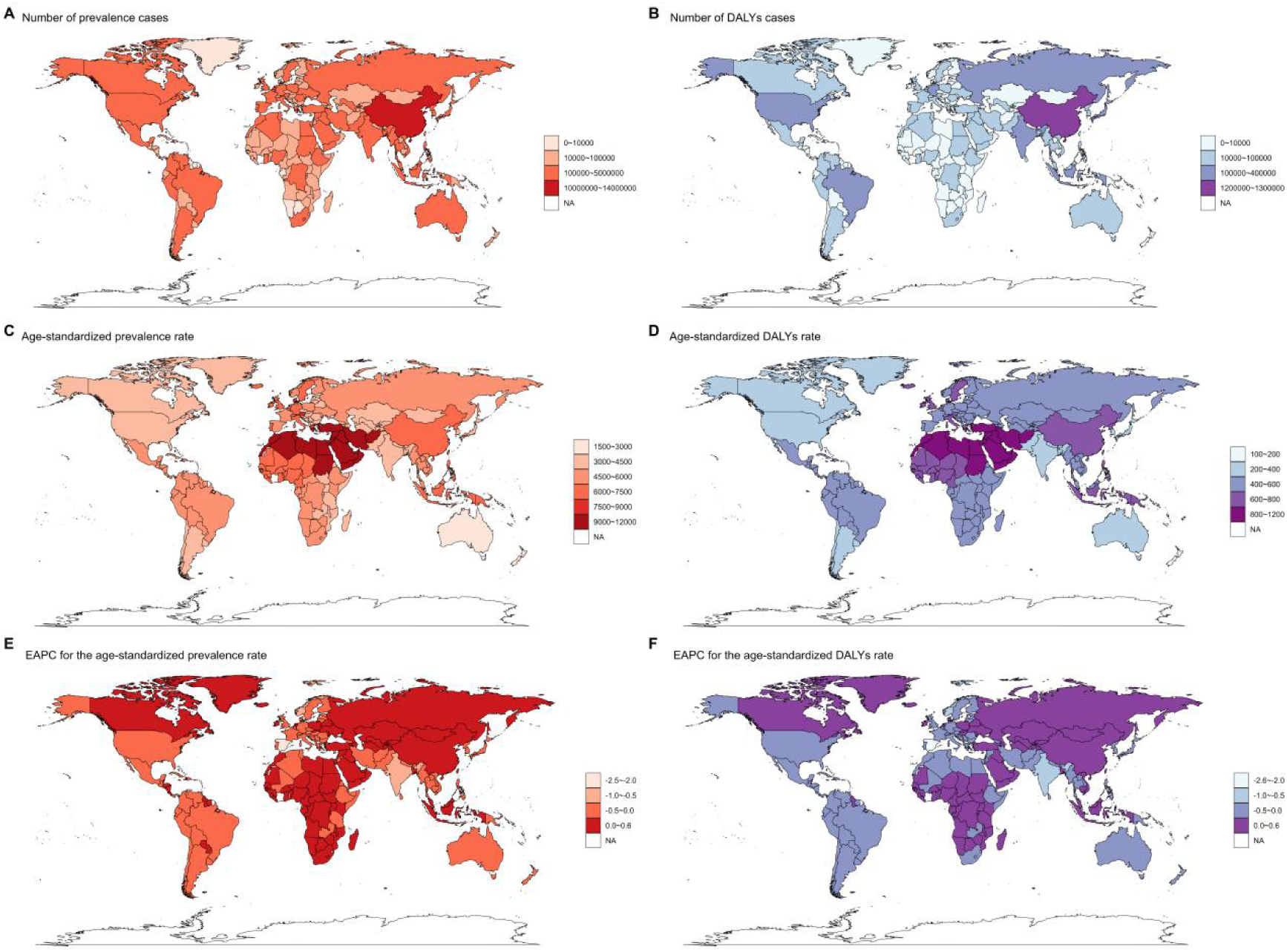
Global geographical distribution characteristics of neck pain. A): Geographical distribution of the number of prevalent cases. B): Geographical distribution of DALYs. C): Geographical distribution of age-standardized prevalence rates. D): Geographical distribution of age-standardized DALY rates. E): Geographical distribution of EAPC in age-standardized prevalence rates. F): Geographical distribution of EAPC in age-standardized DALY rates.

**Figure S14.**
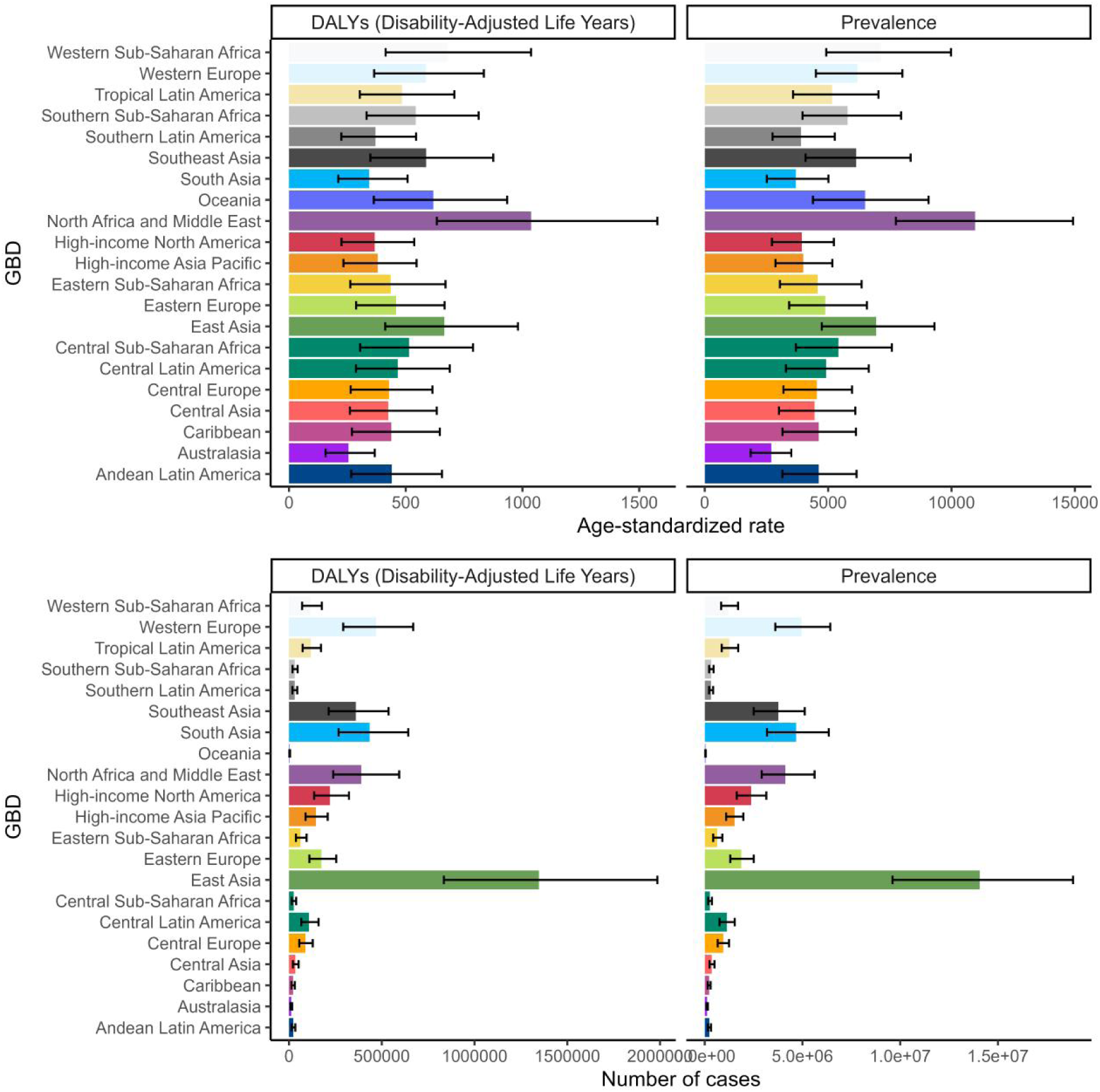
The global epidemiological burden of Neck pain across 21 regions in 2021.

**Figure S15.**
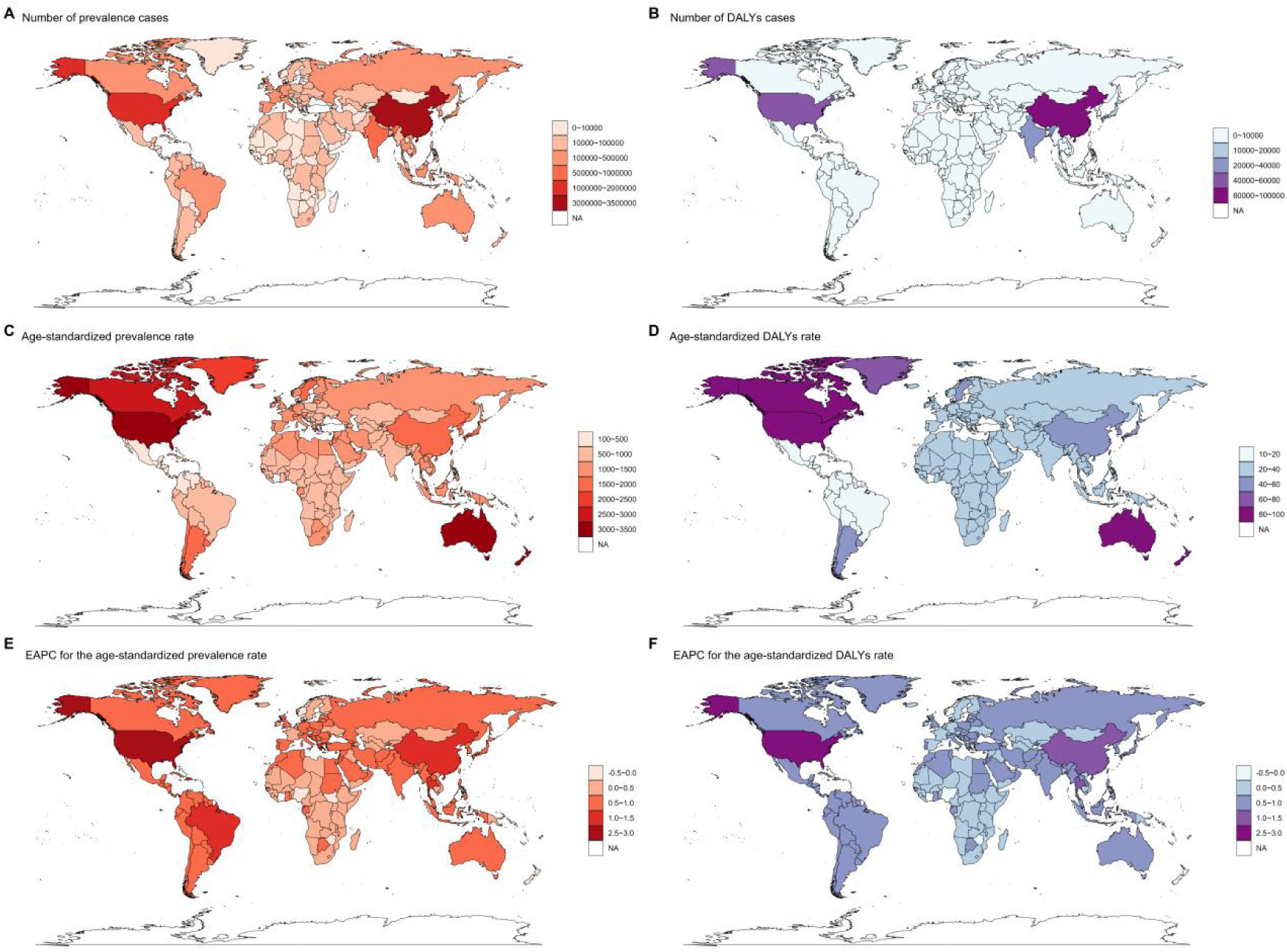
Global geographical distribution characteristics of gout. A): Geographical distribution of the number of prevalent cases. B): Geographical distribution of DALYs. C): Geographical distribution of age-standardized prevalence rates. D): Geographical distribution of age-standardized DALY rates. E): Geographical distribution of EAPC in age-standardized prevalence rates. F): Geographical distribution of EAPC in age-standardized DALY rates.

**Figure S16.**
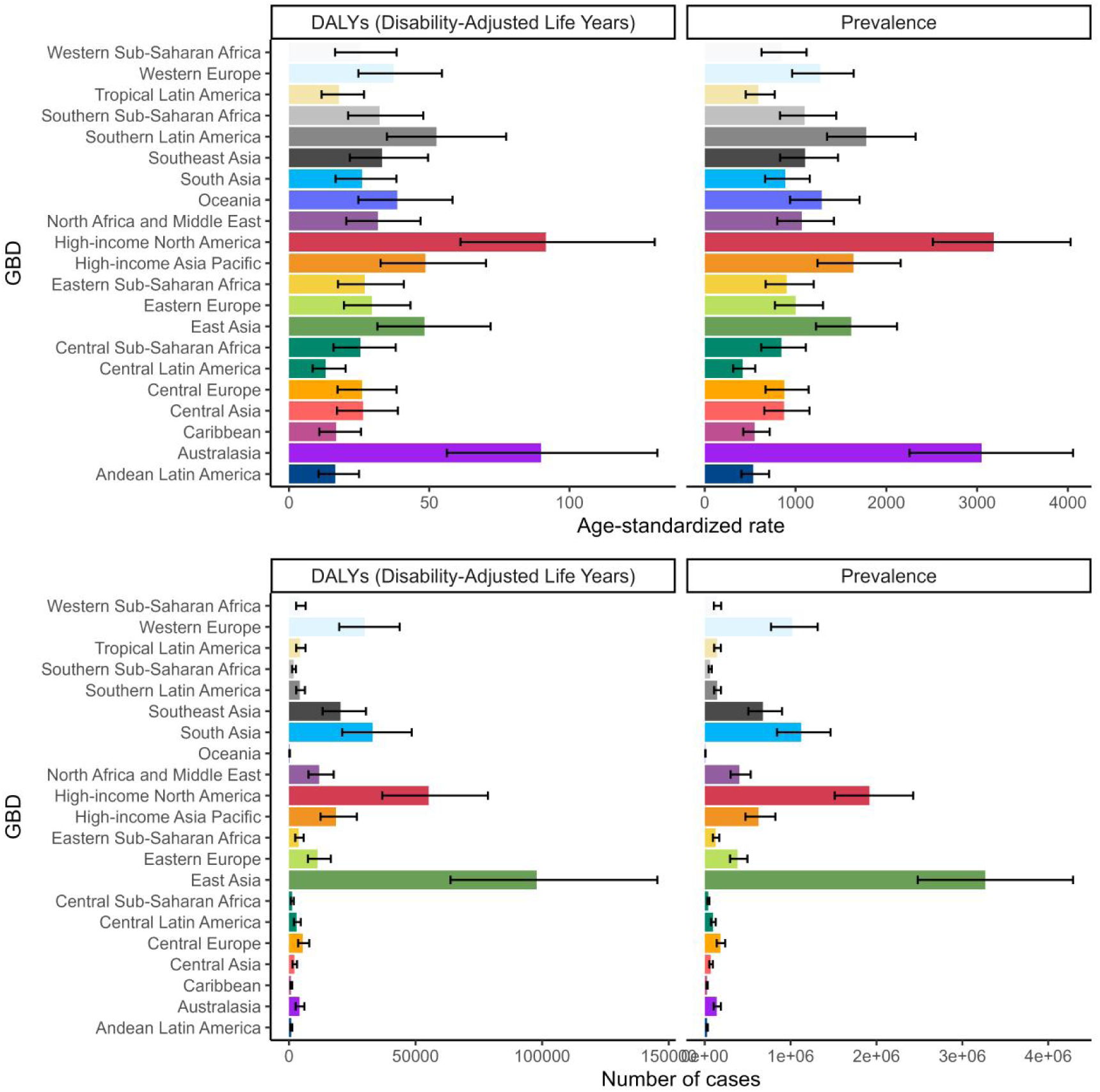
The global epidemiological burden of gout across 21 regions in 2021.

**Figure S17.**
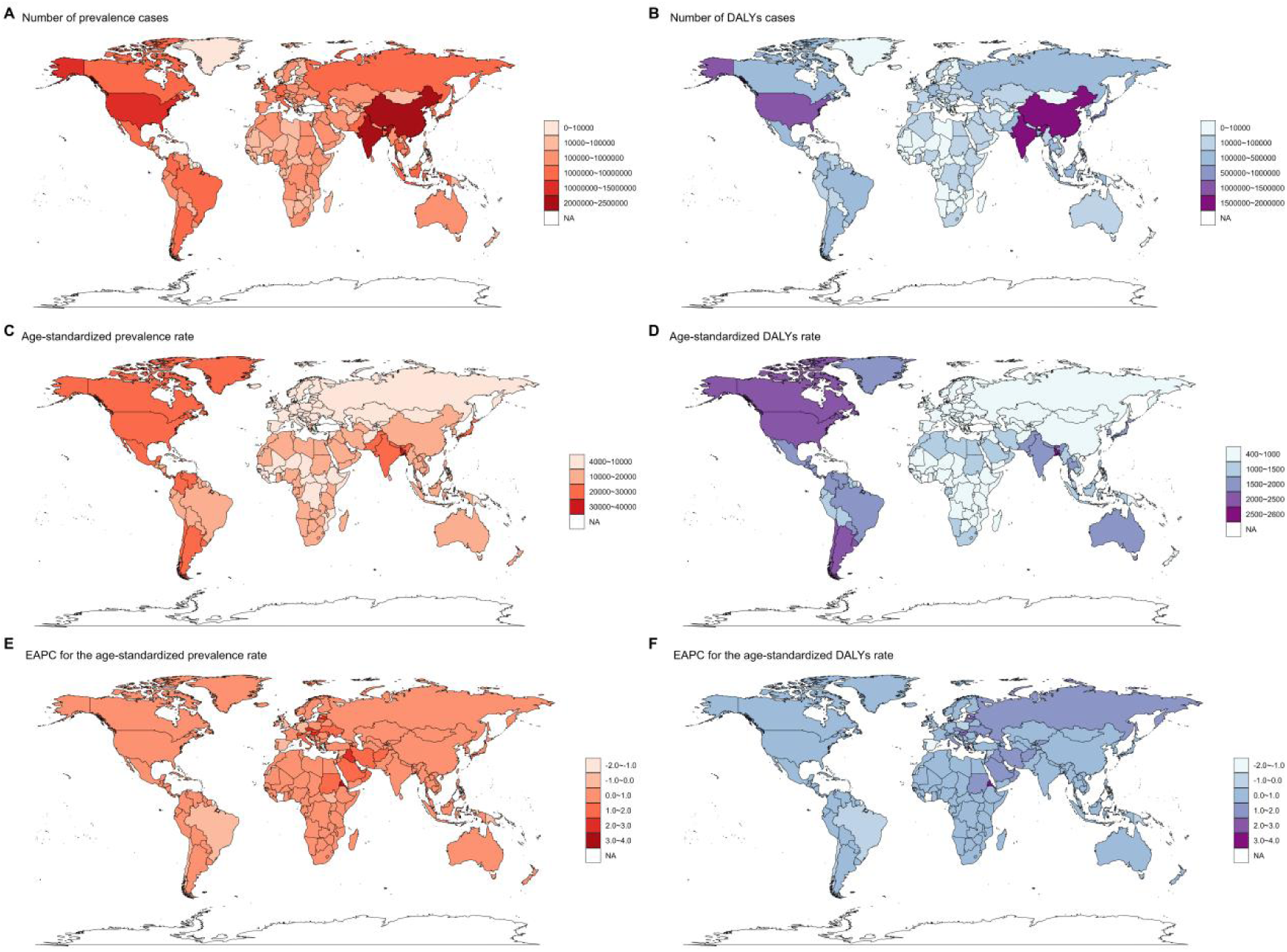
Global geographical distribution characteristics of OMSD. A): Geographical distribution of the number of prevalent cases. B): Geographical distribution of DALYs. C): Geographical distribution of age-standardized prevalence rates. D): Geographical distribution of age-standardized DALY rates. E): Geographical distribution of EAPC in age-standardized prevalence rates. F): Geographical distribution of EAPC in age-standardized DALY rates.

**Figure S18.**
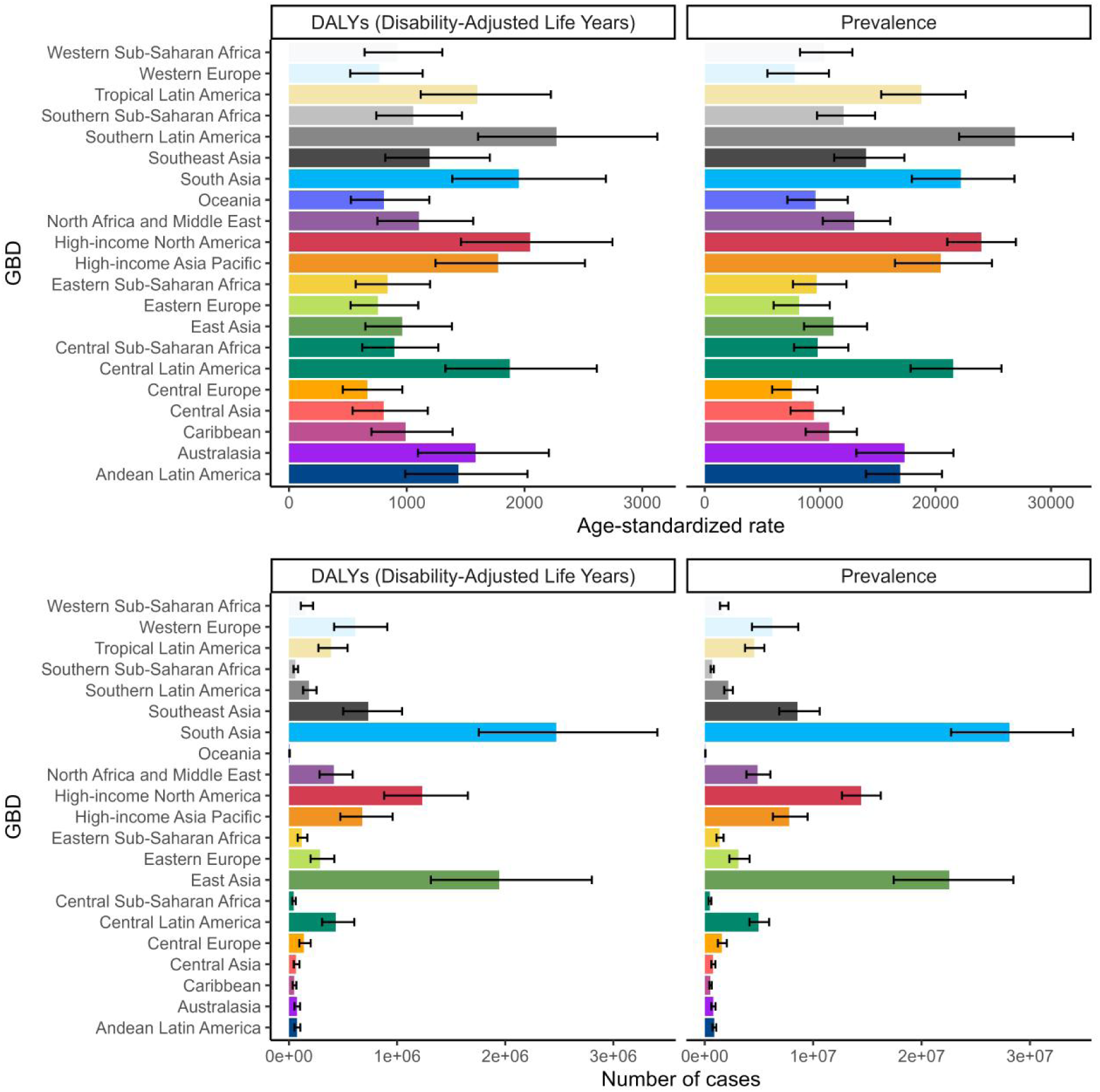
The global epidemiological burden of OMSD across 21 regions in 2021.

**Table S1.** Global Burden in DALYs of Musculoskeletal Disorders Attributable to Associated Risk Factors among postmenopausal women, 1990-2021.

| Disease and Associated Risk Factors | DALYs |  |  |  |  |
| --- | --- | --- | --- | --- | --- |
|  | 1990 |  | 2021 |  |  |
|  | Cases | ASDR | Cases | ASDR | EAPC |
| Gout (High body-mass index) | 34096<br>(20823–53280) | 9.47 (5.79–14.8) | 118101<br>(72608–181205) | 15.02<br>(9.23–23.04) | 1.75<br>(1.65–1.86) |
| Gout (Kidney dysfunction) | 24600<br>(15133–36748) | 6.83 (4.2–10.21) | 60971 (38214–89489) | 7.75<br>(4.86–11.38) | 0.72<br>(0.62–0.83) |
| Low back pain (High body-mass index) | 973690<br>(103147–2026395) | 270.51<br>(28.66–562.98) | 2708944<br>(292322–5543617) | 344.44<br>(37.17–704.87) | 0.83<br>(0.8–0.87) |
| Low back pain (Occupational ergonomic factors) | 1216909<br>(807567–1733865) | 338.08<br>(224.36–481.71) | 2400874<br>(1604144–3425392) | 305.27<br>(203.97–435.54) | –0.27<br>(–0.34––0.2) |
| Low back pain (Smoking) | 956221<br>(540673–1470183) | 265.66<br>(150.21–408.45) | 1392955<br>(772161–2163180) | 177.12<br>(98.18–275.05) | –1.13<br>(–1.18––1.09) |
| Osteoarthritis (High body-mass index) | 693112<br>(–60263–1998715) | 192.56<br>(–16.74–555.29) | 2063539<br>(–193179–5761850) | 262.38<br>(–24.56–732.62) | 1.02 (1–1.03) |
| Rheumatoid arthritis (Smoking) | 37459<br>(27288–50209) | 10.41<br>(7.58–13.95) | 48630 (33502–67323) | 6.18 (4.26–8.56) | –1.51<br>(–1.57––1.46) |

**Table S2.**
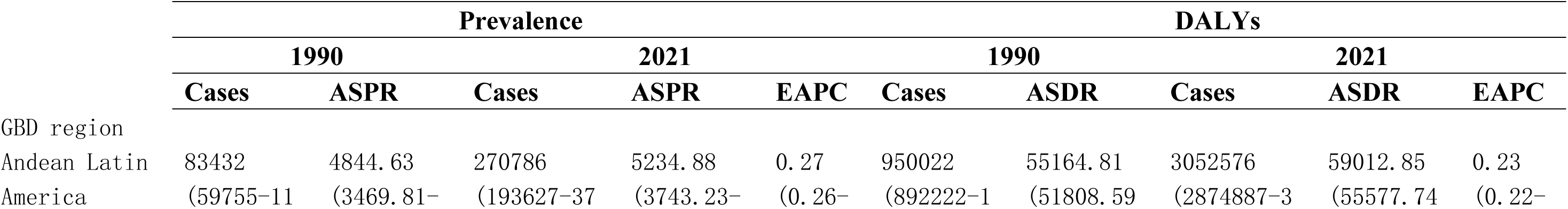
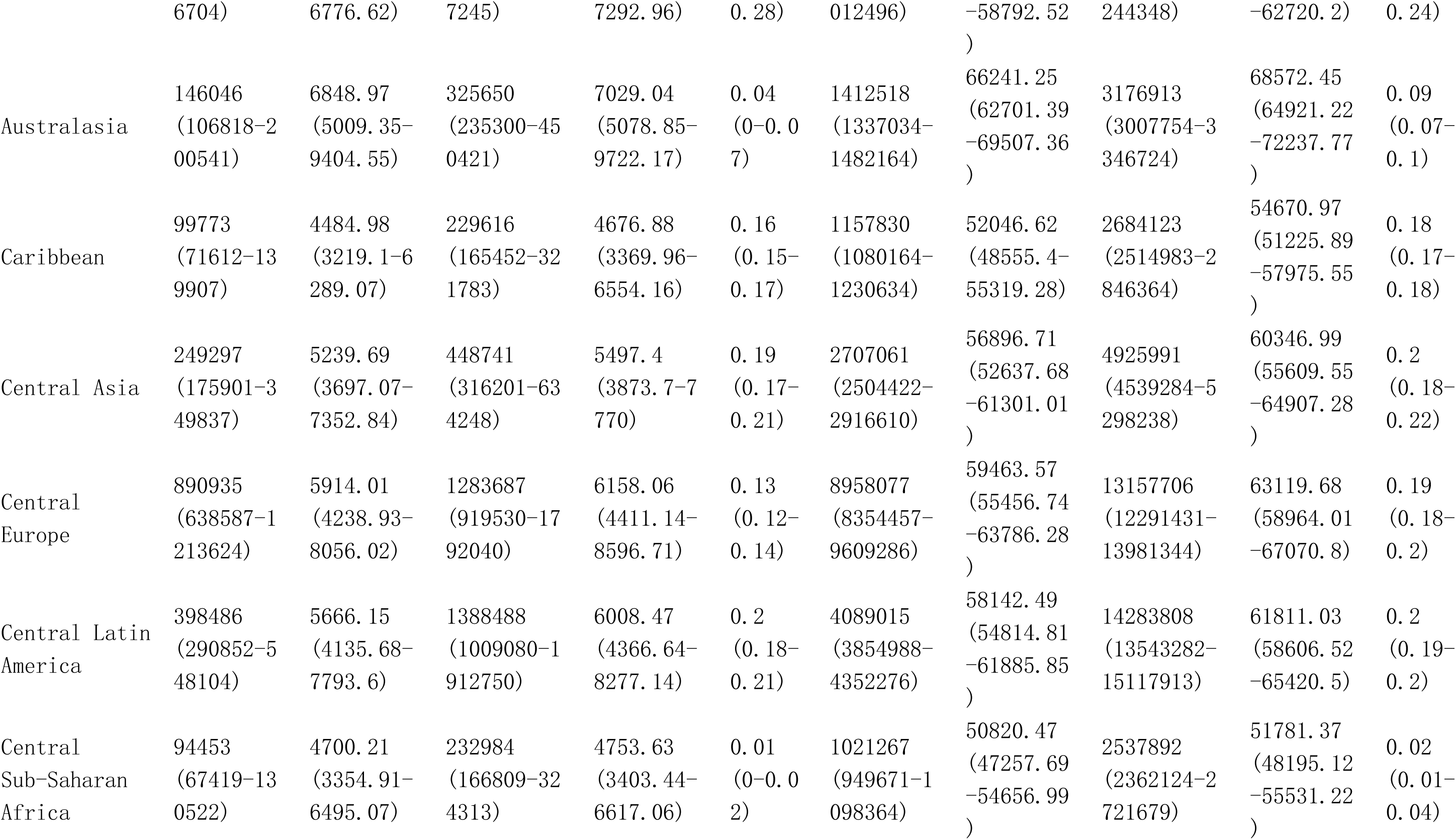
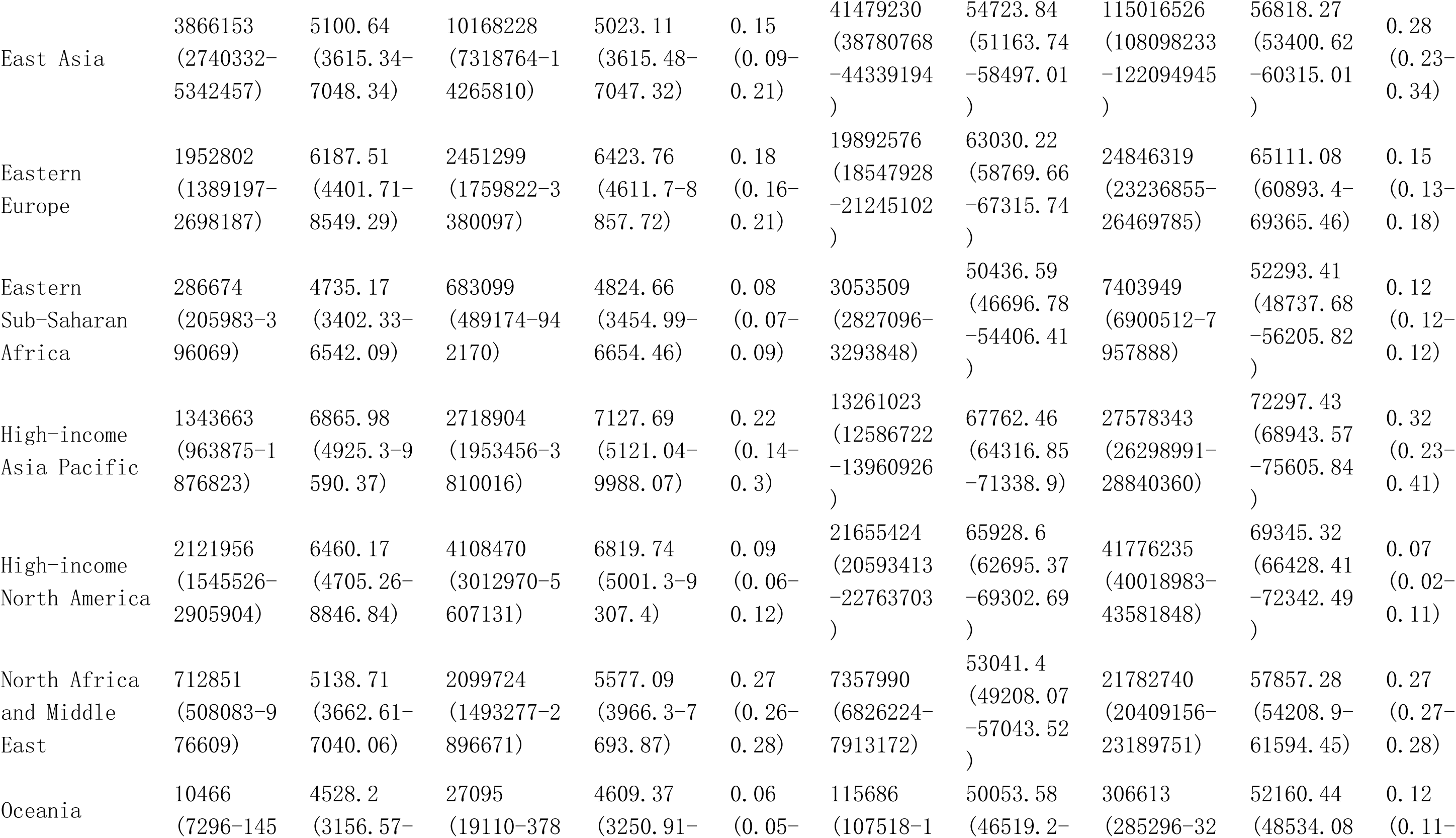
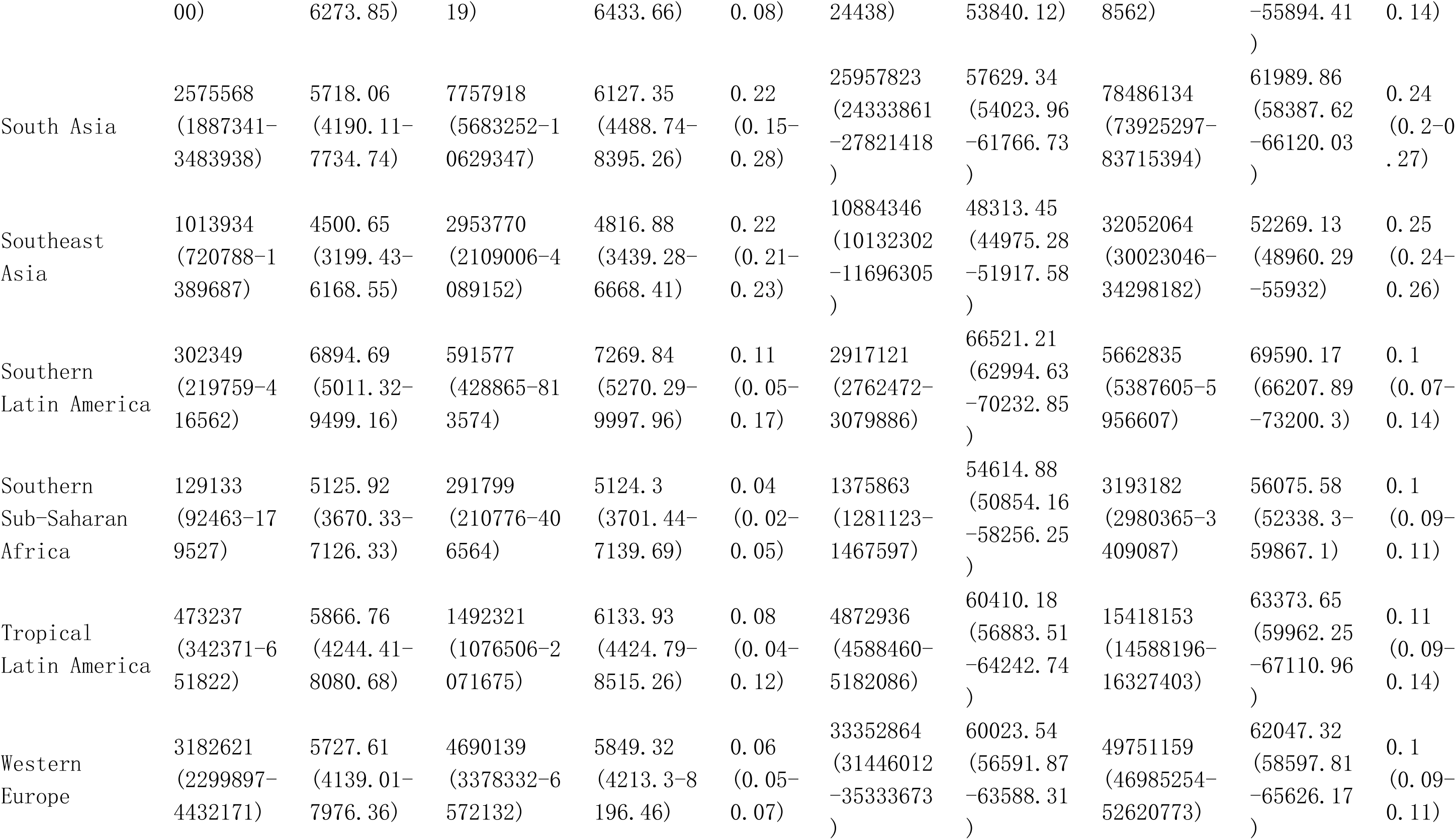
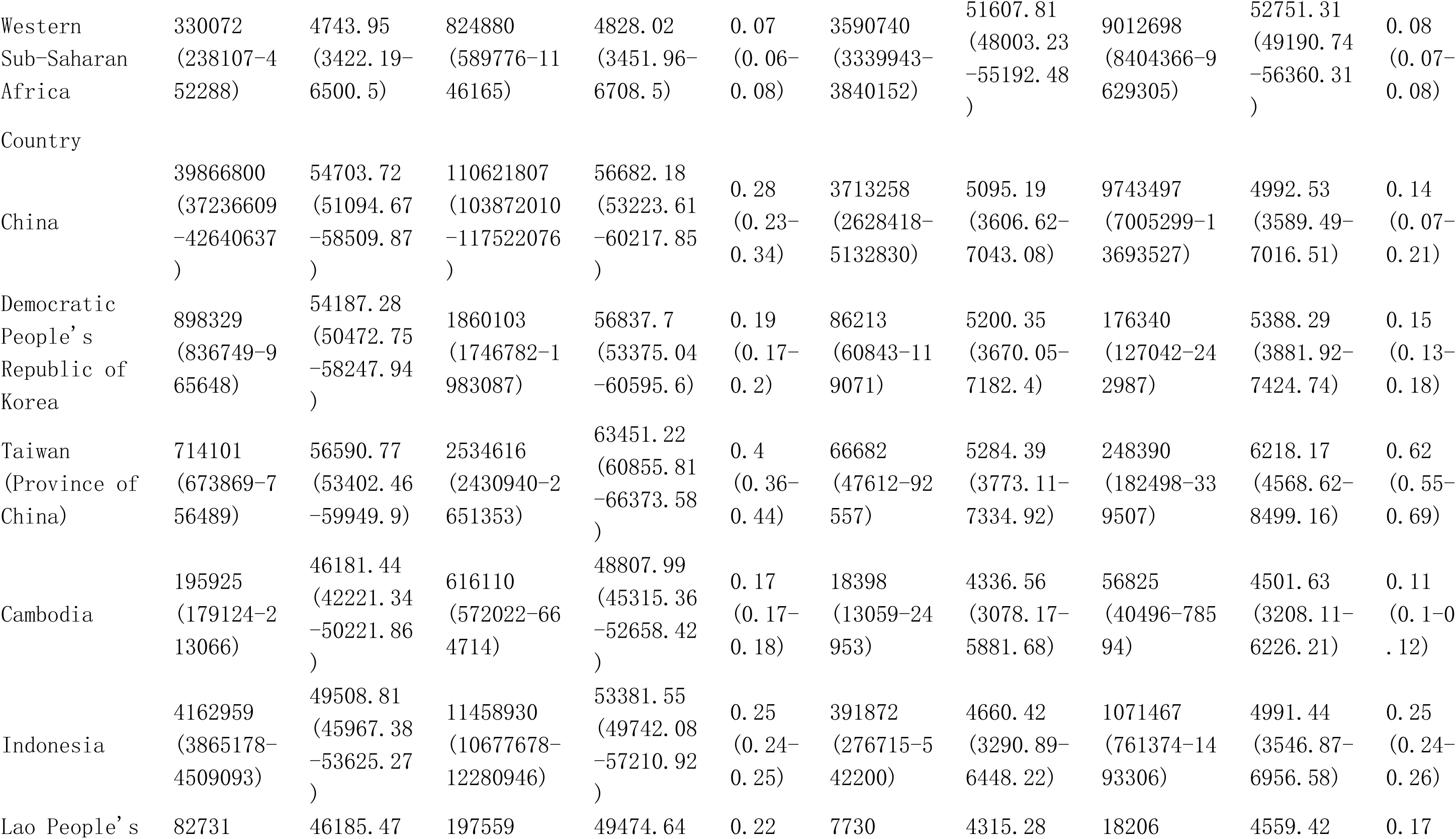
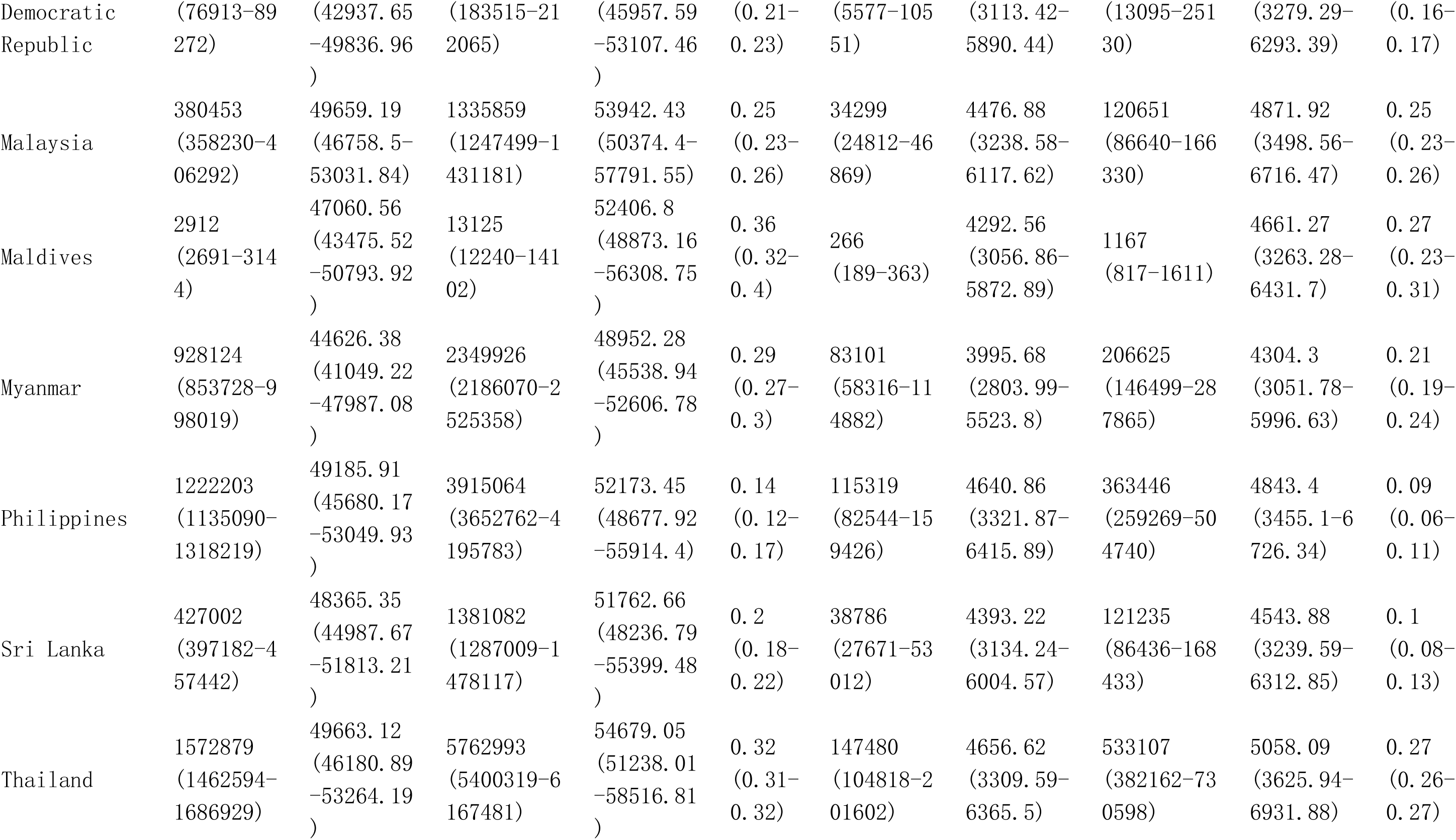
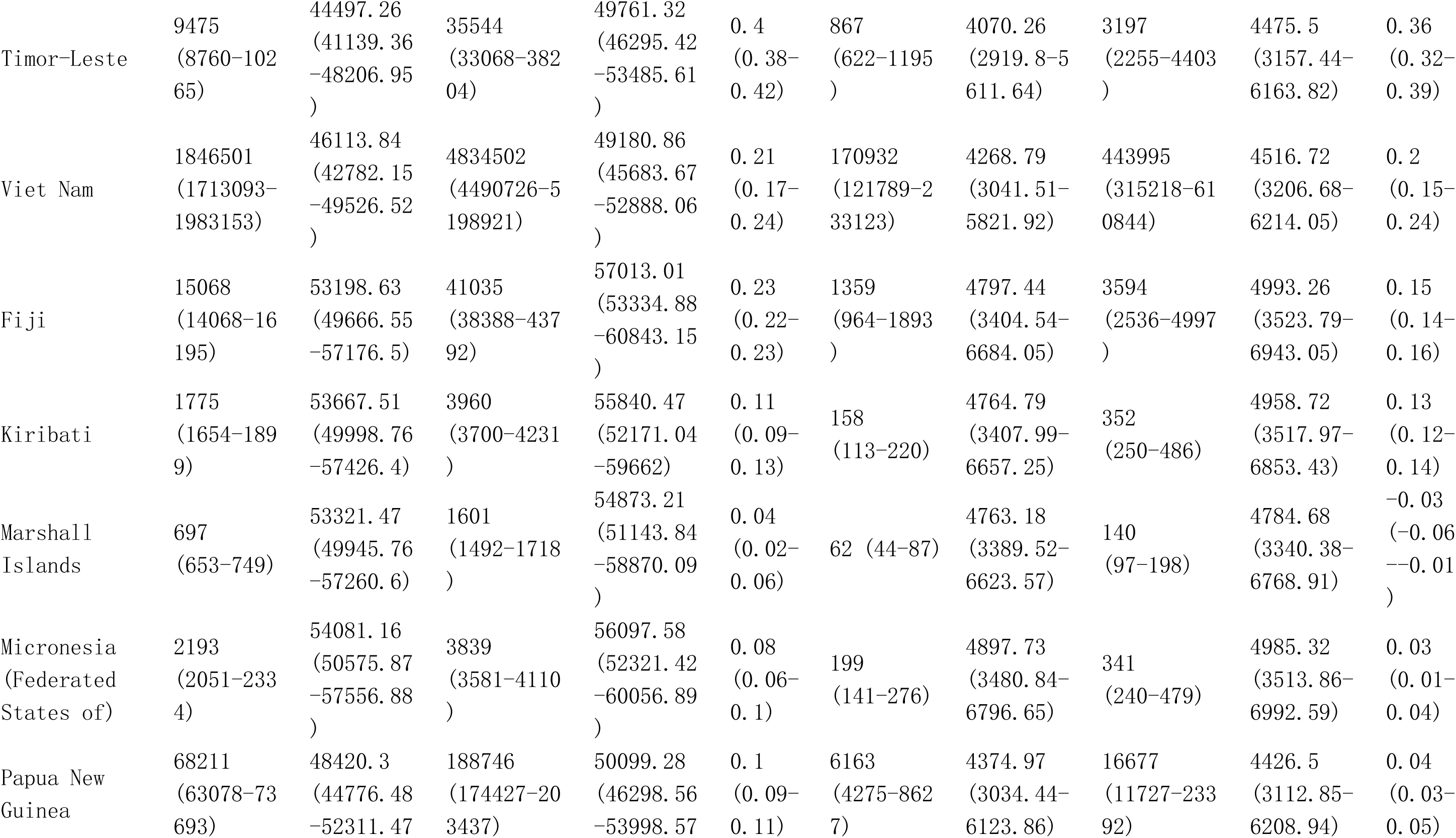
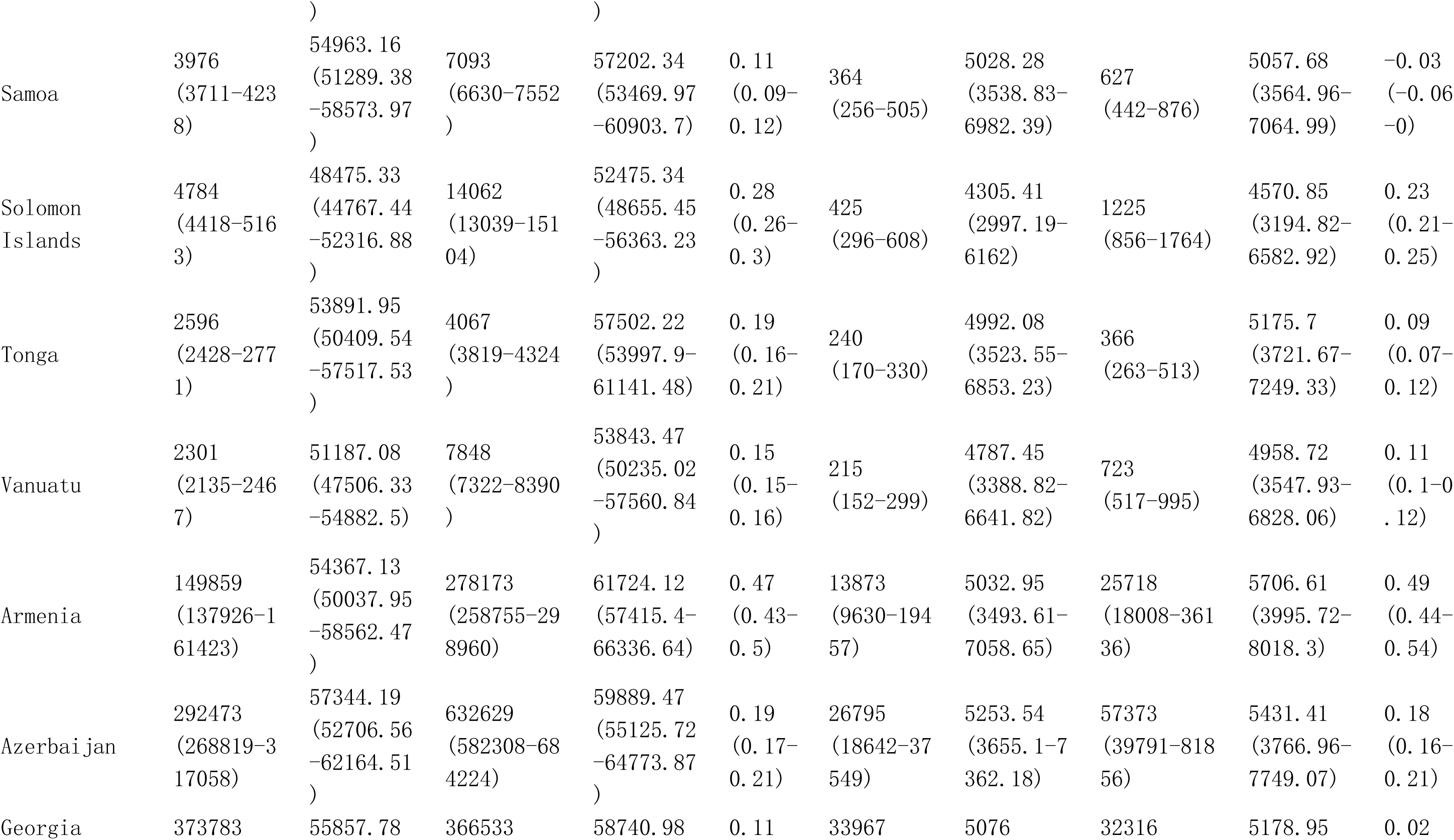
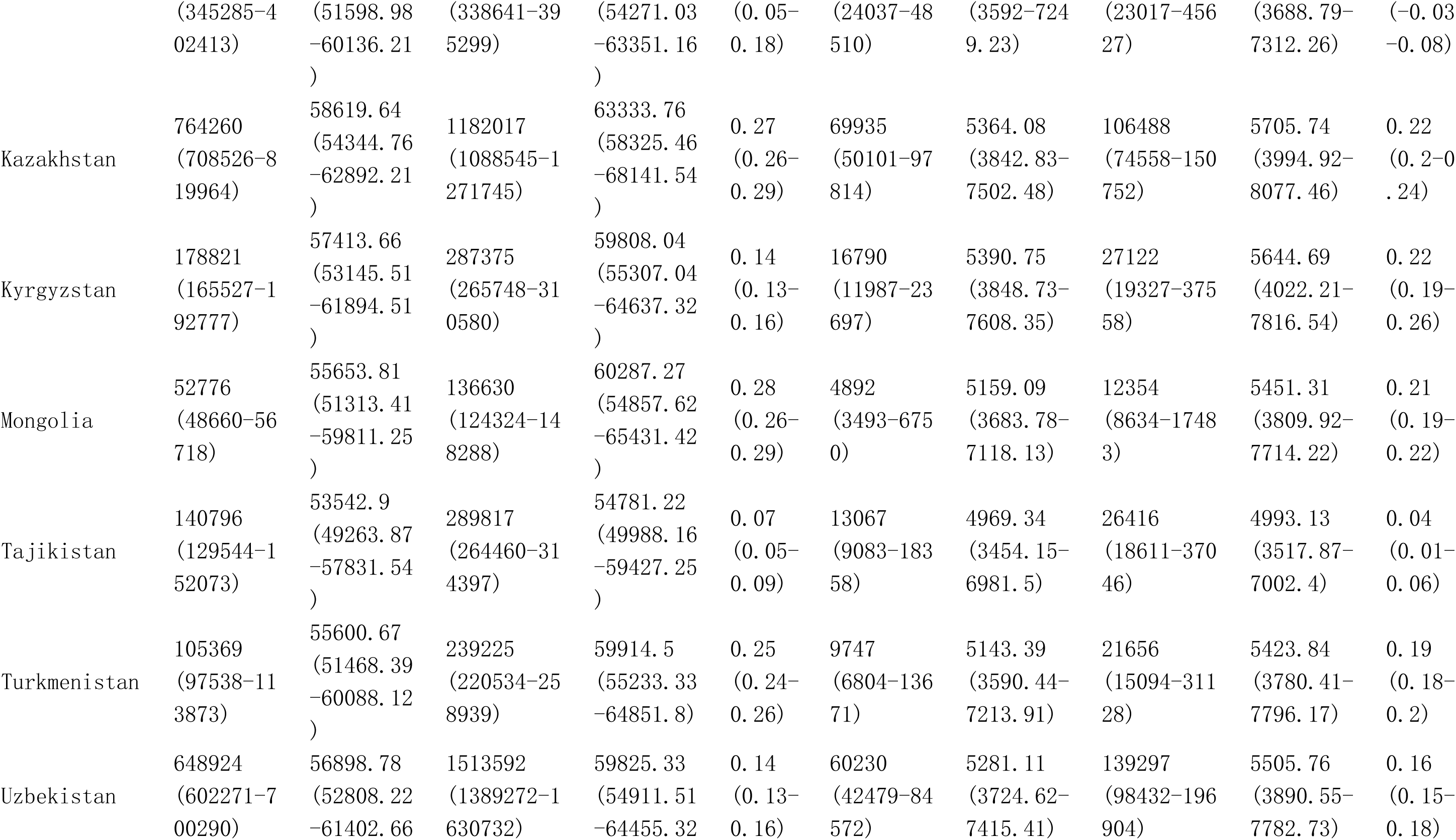
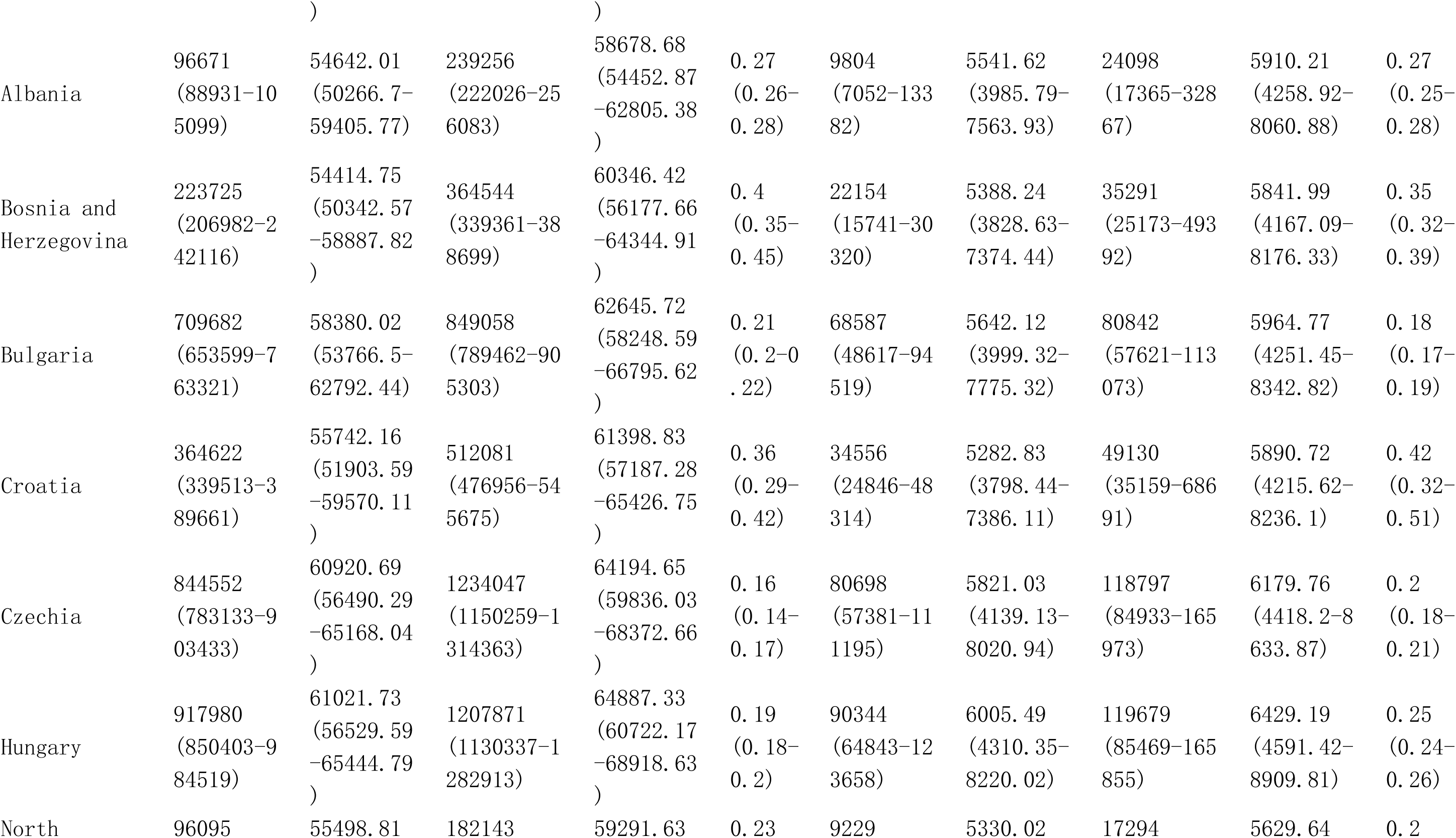
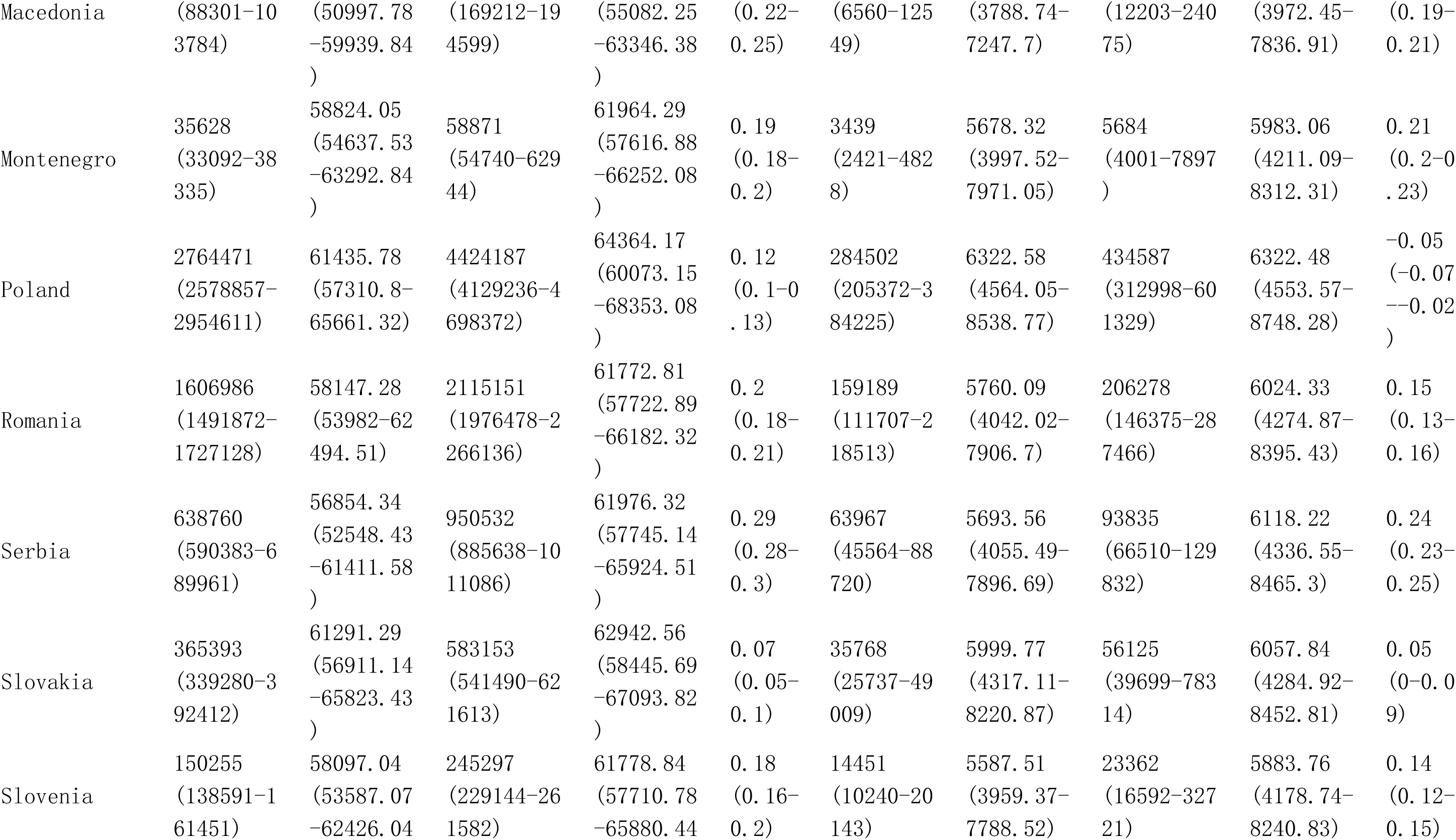
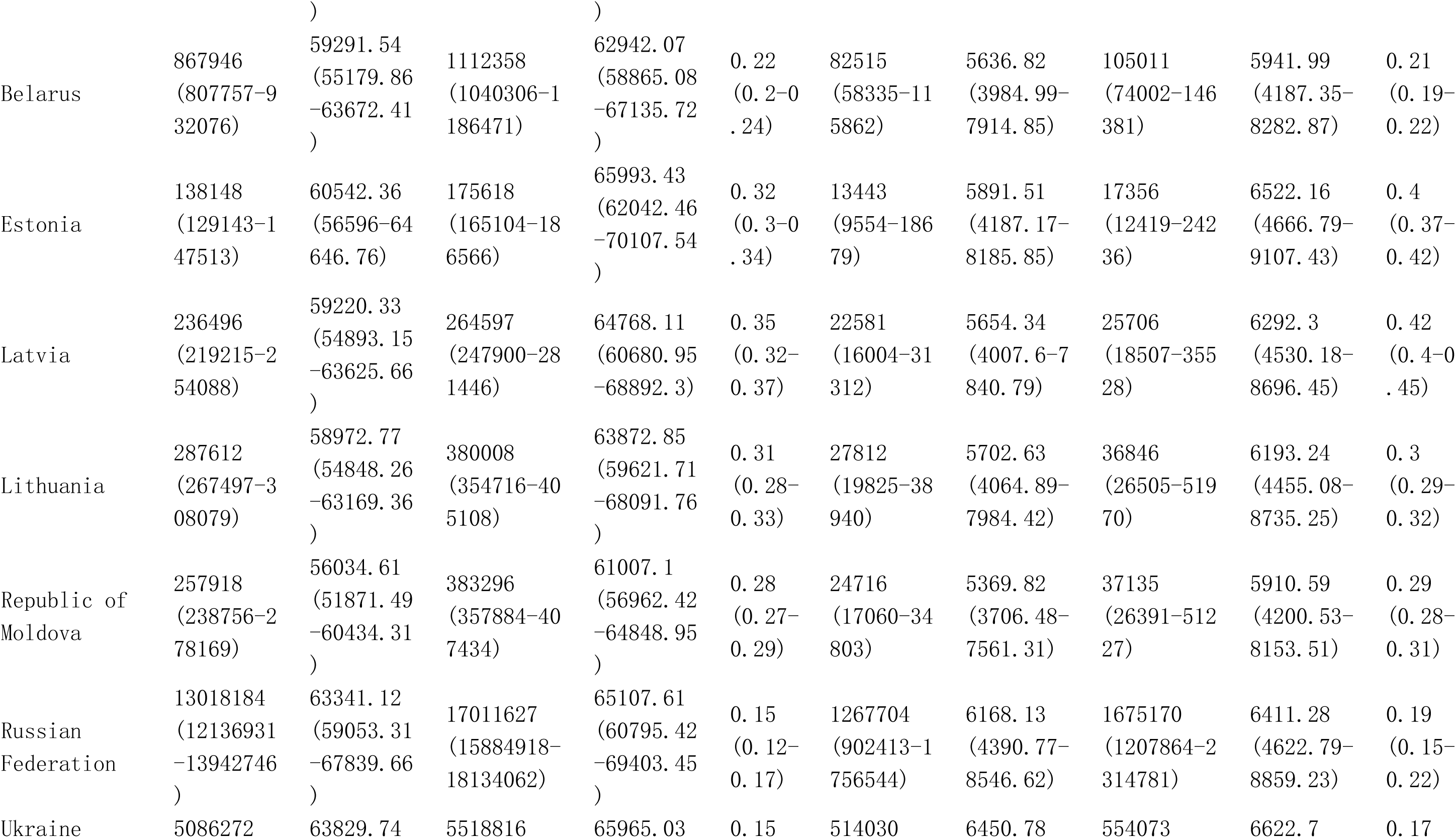
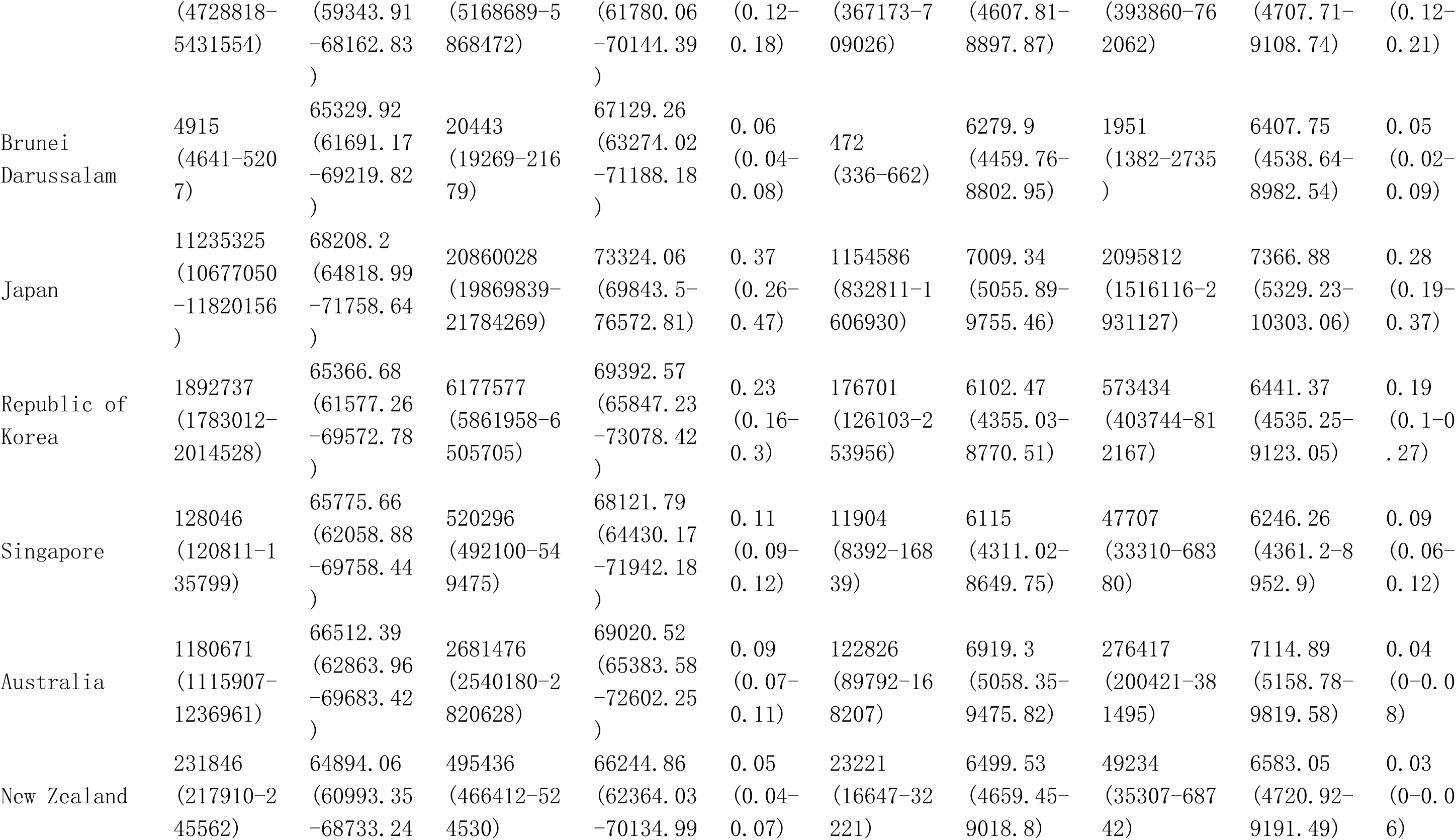
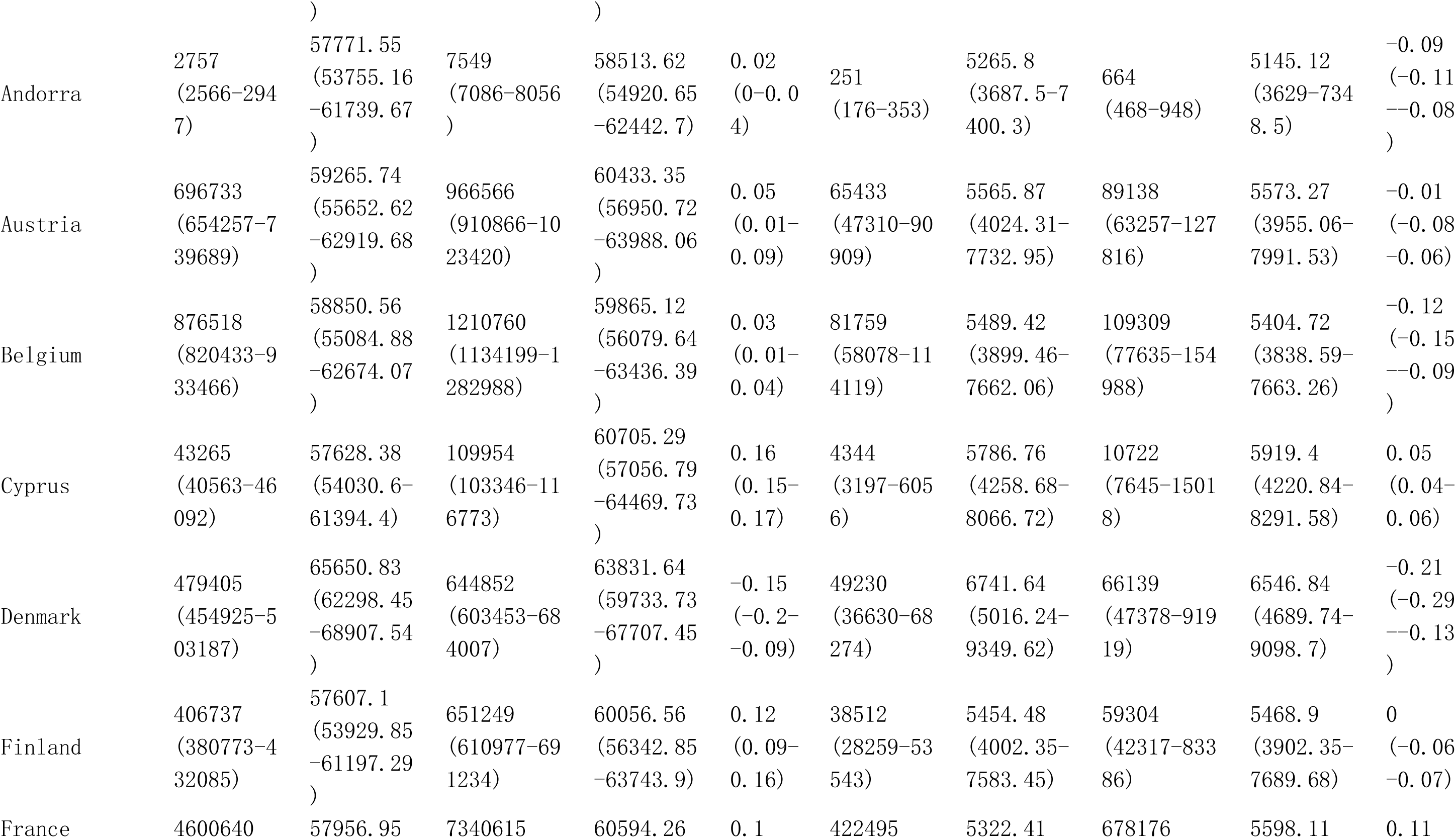
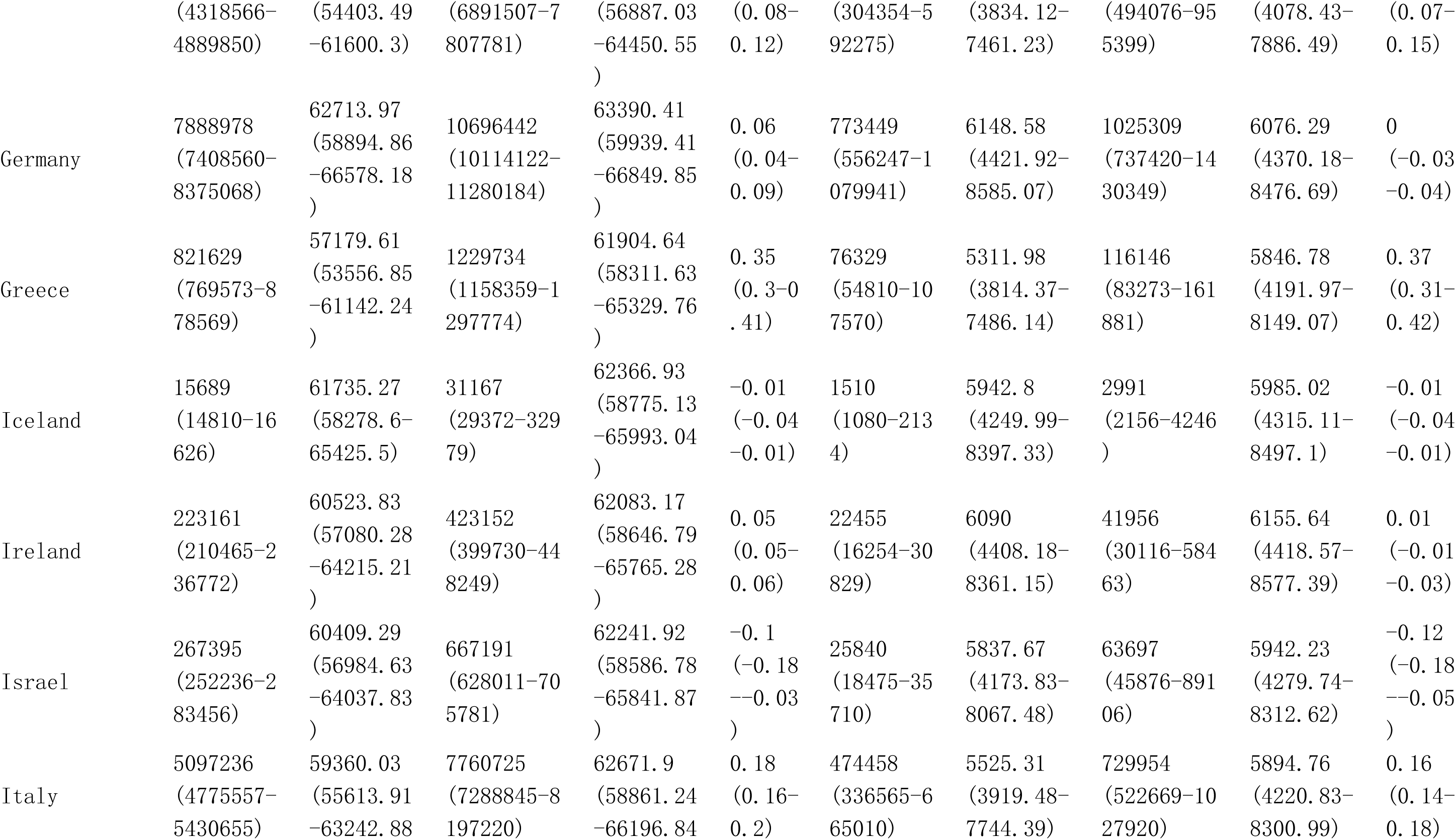
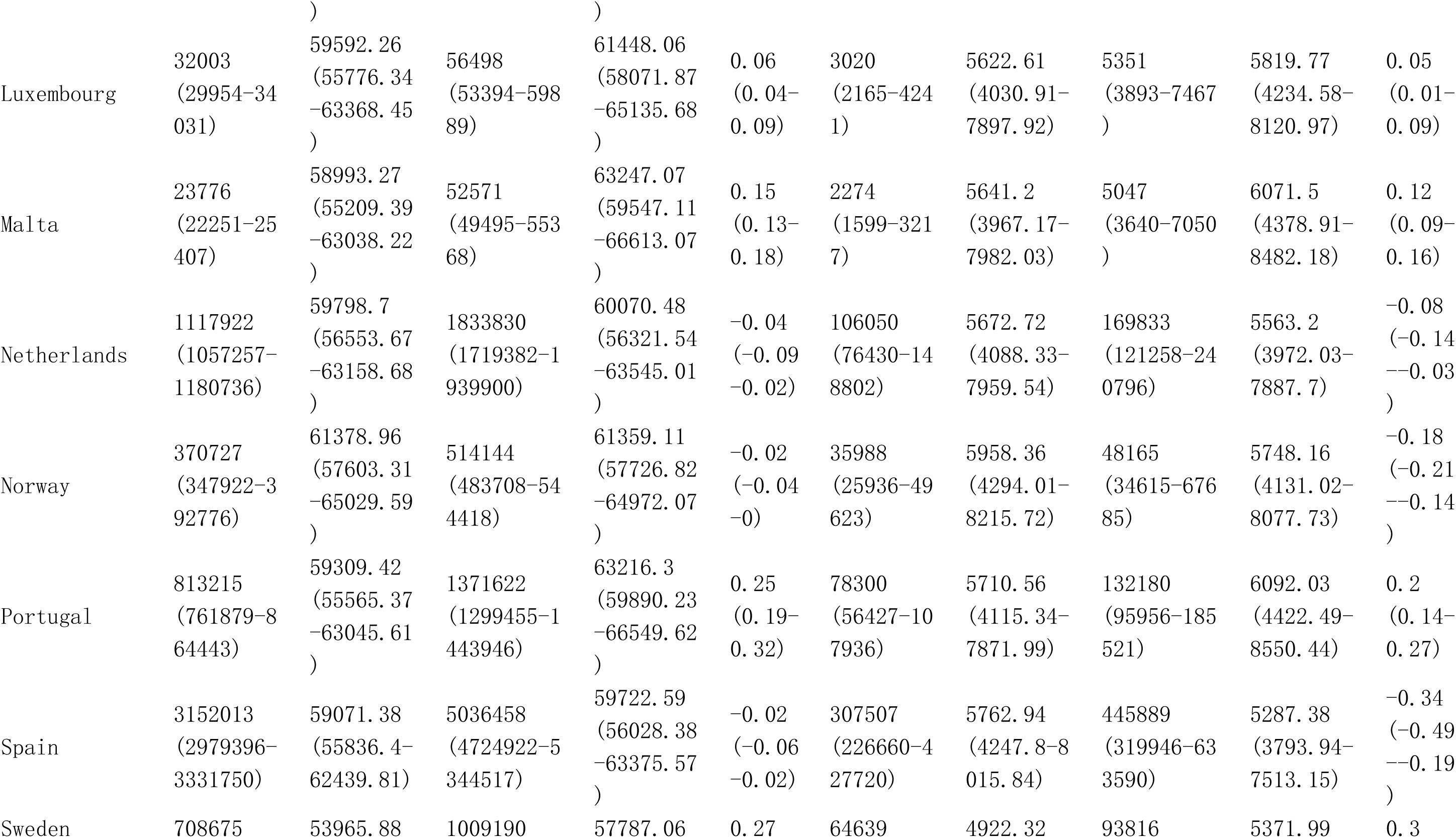
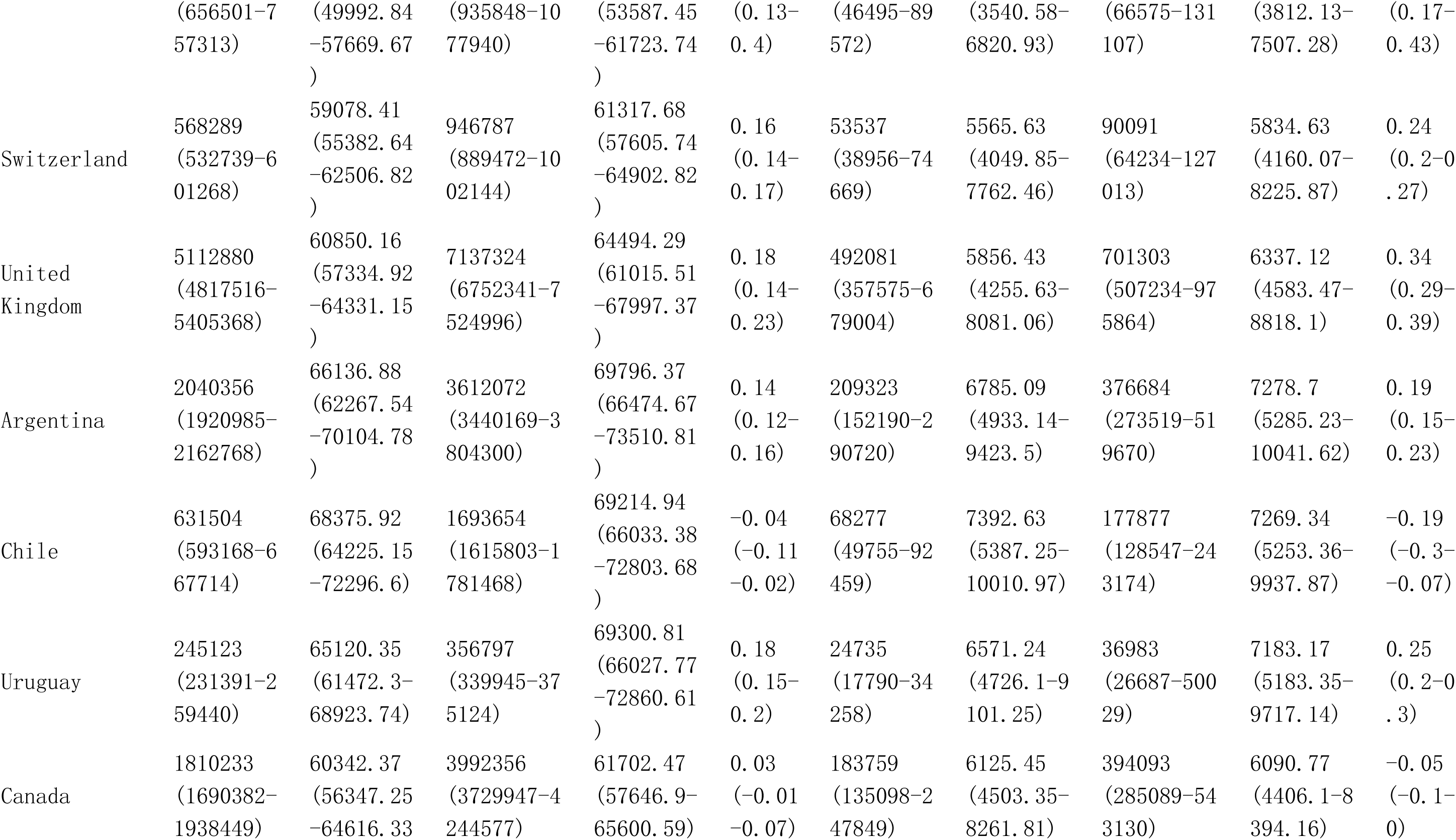
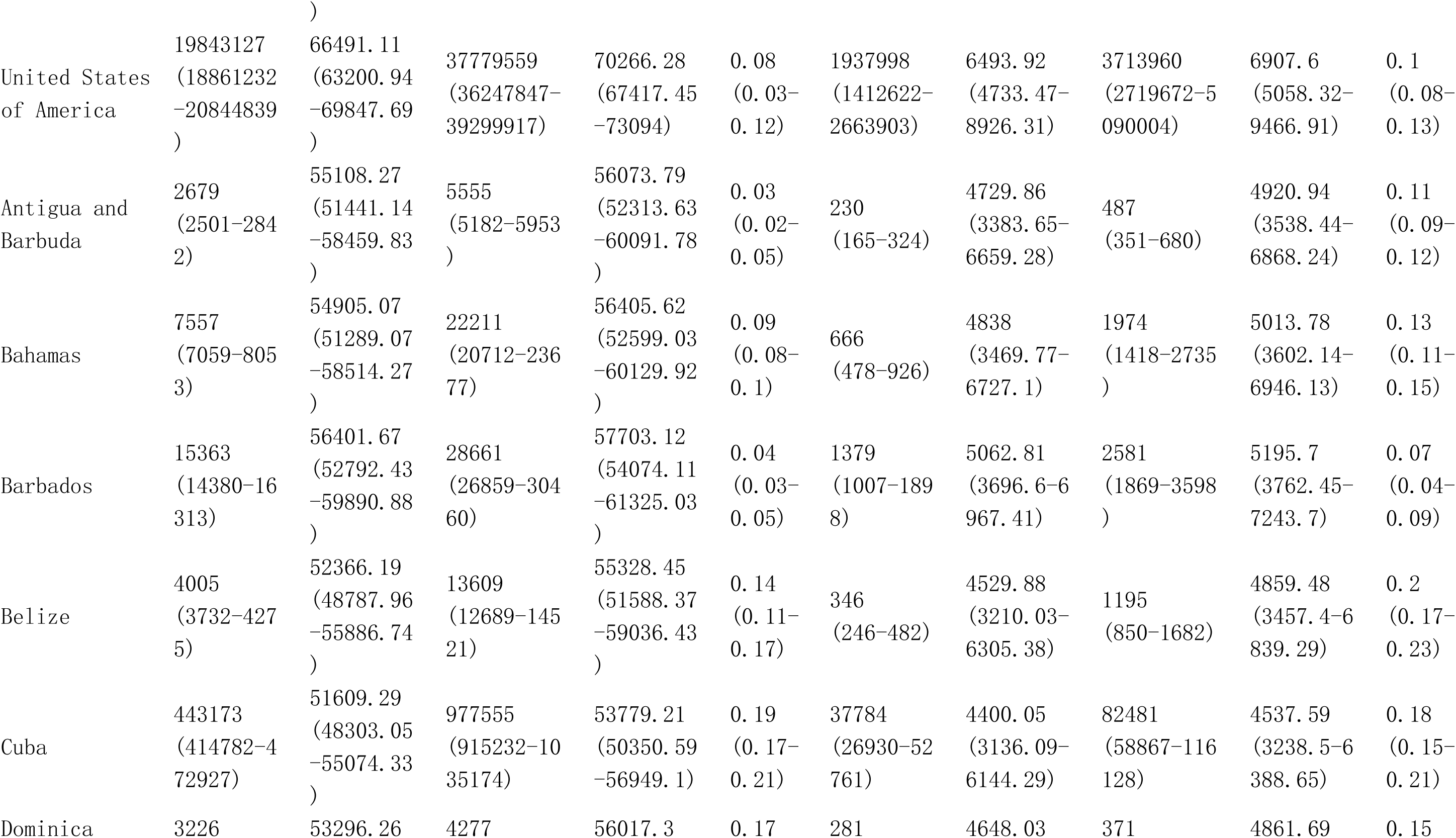
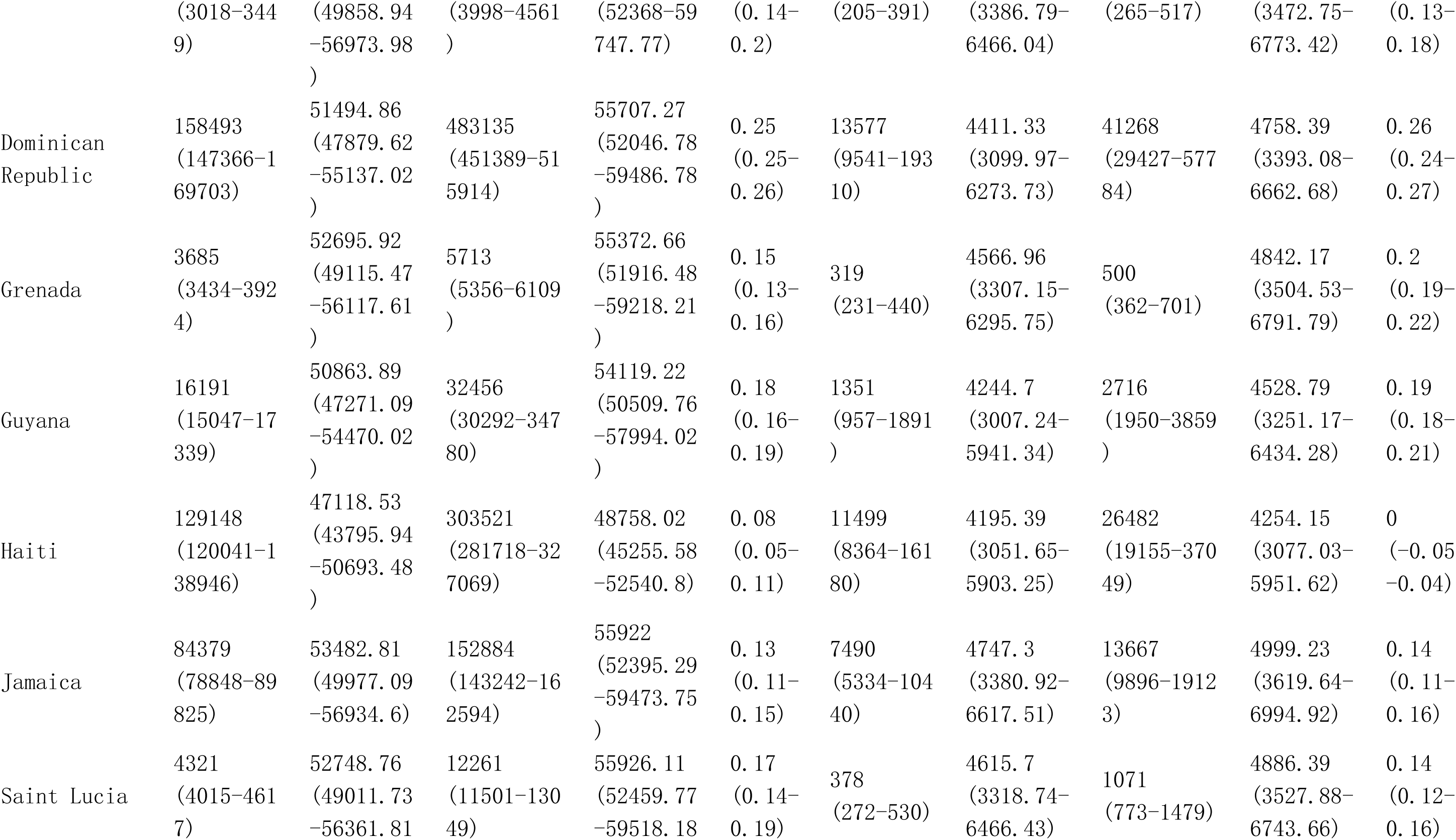
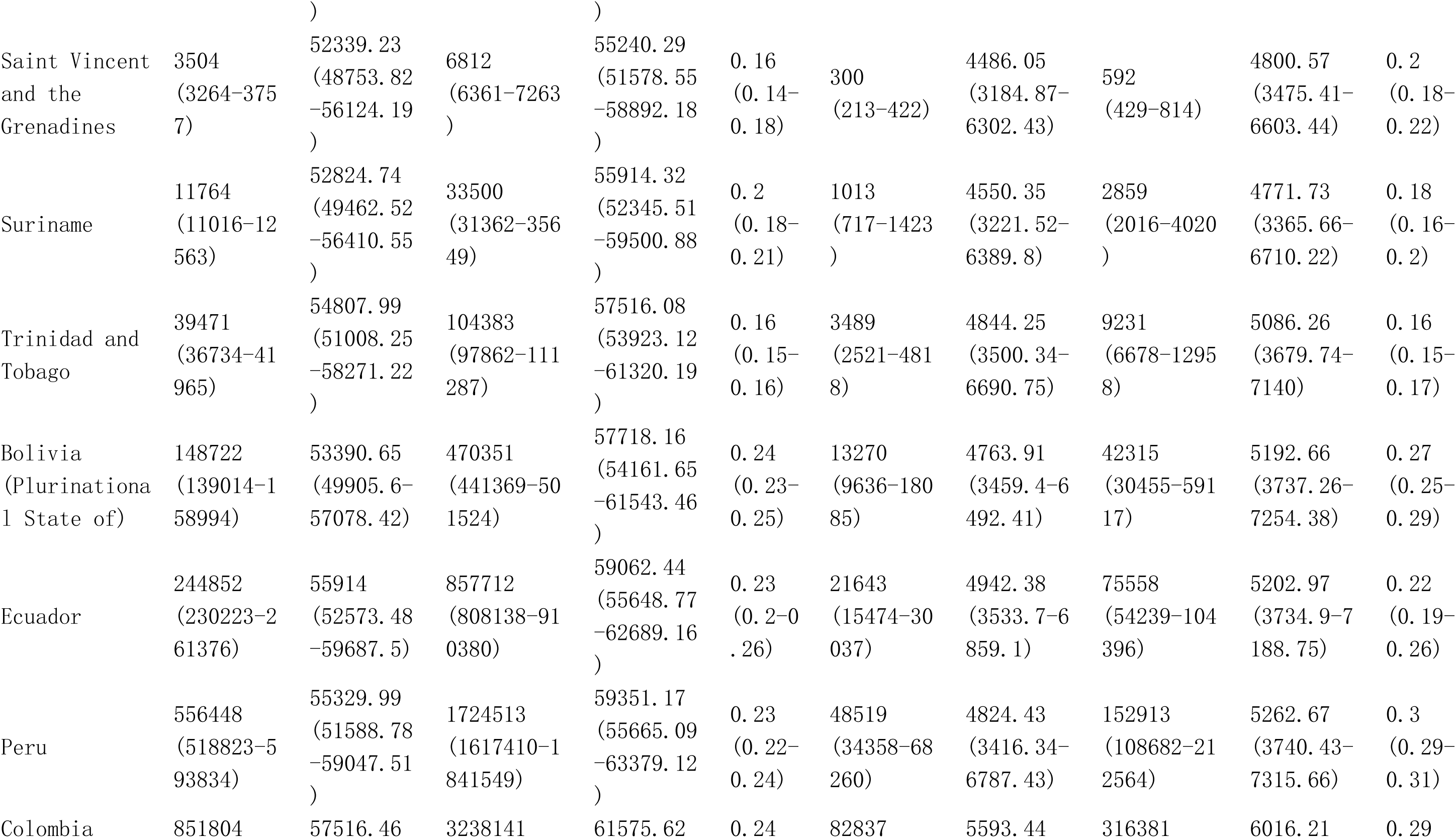
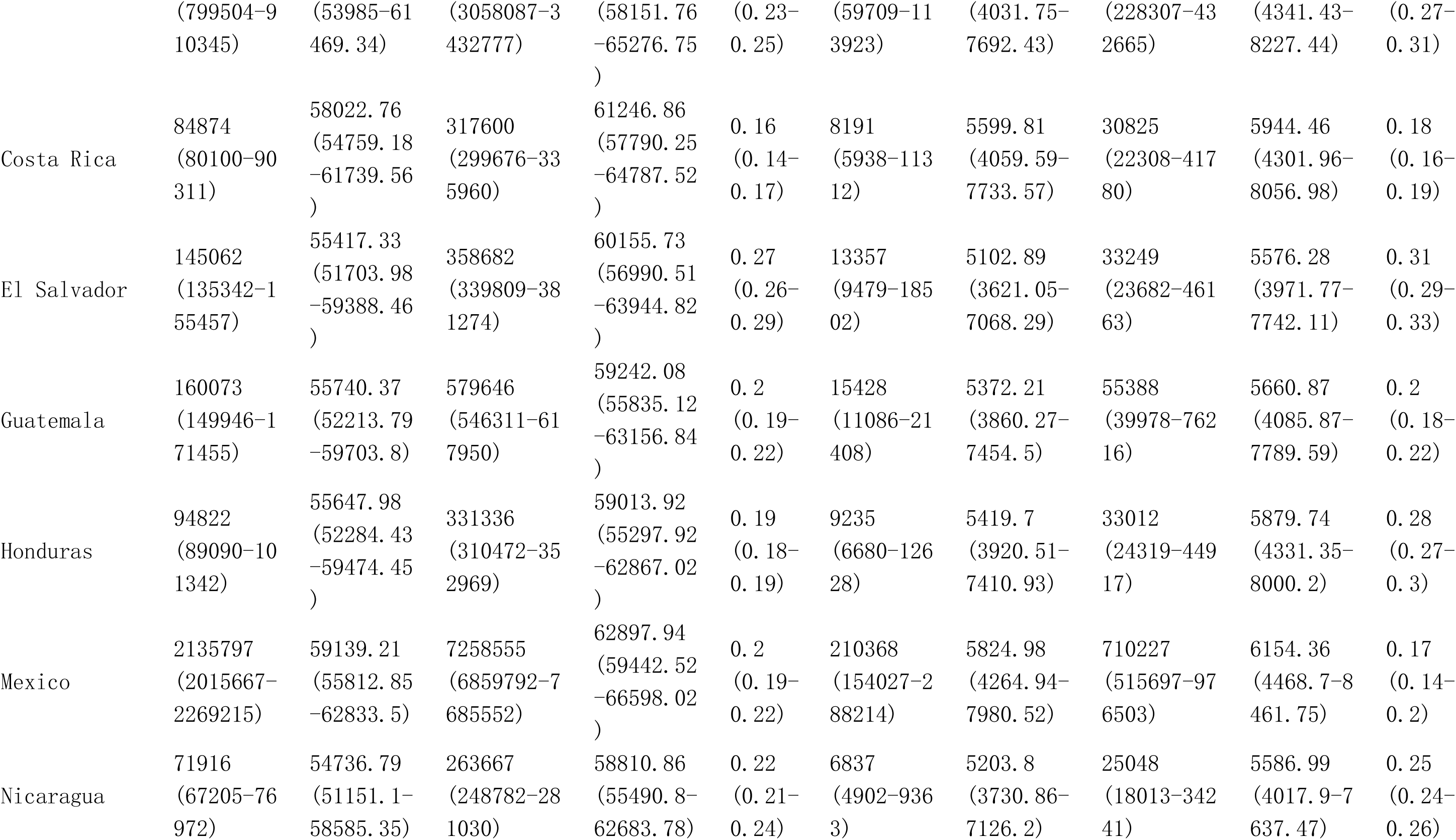
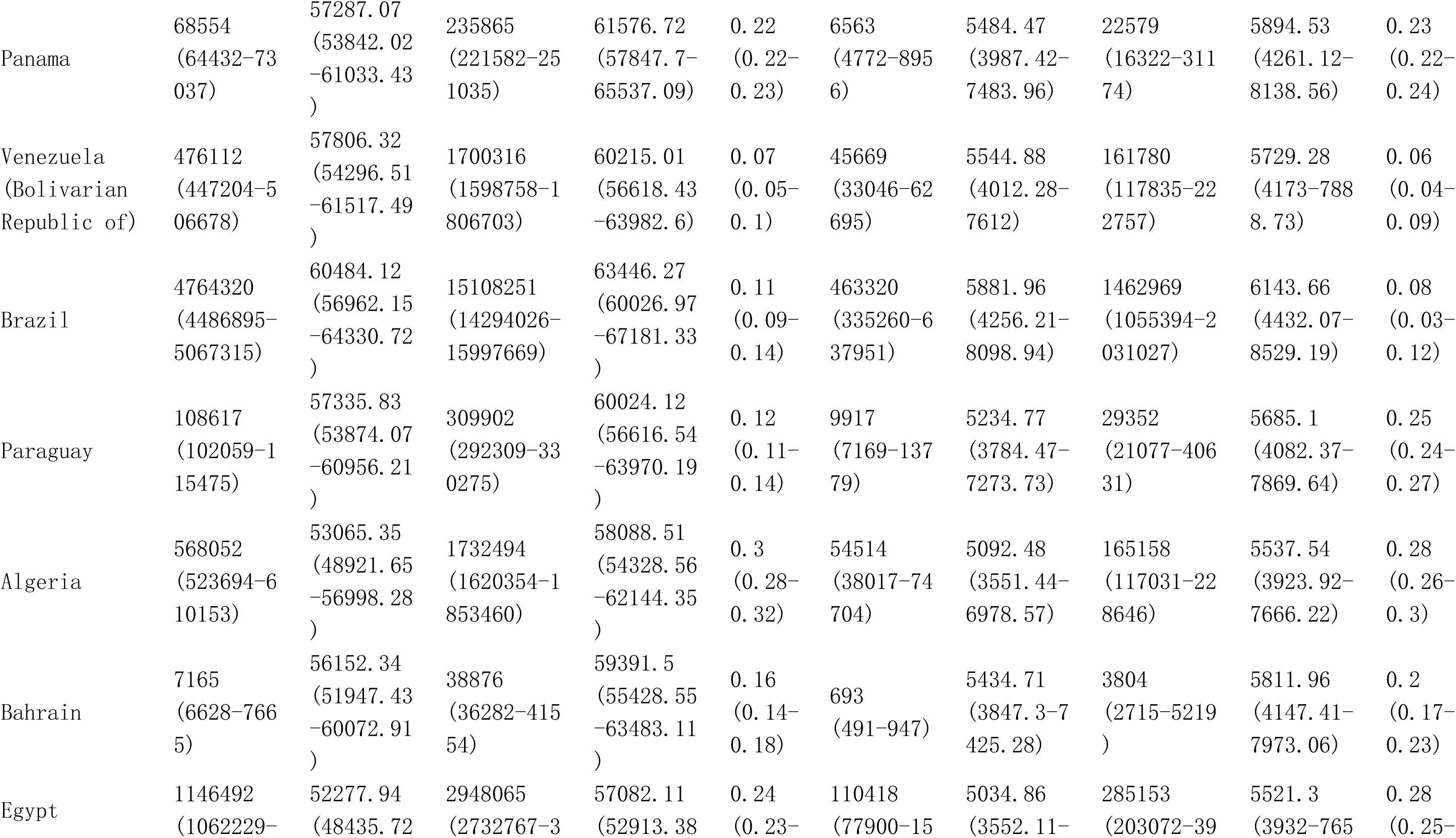
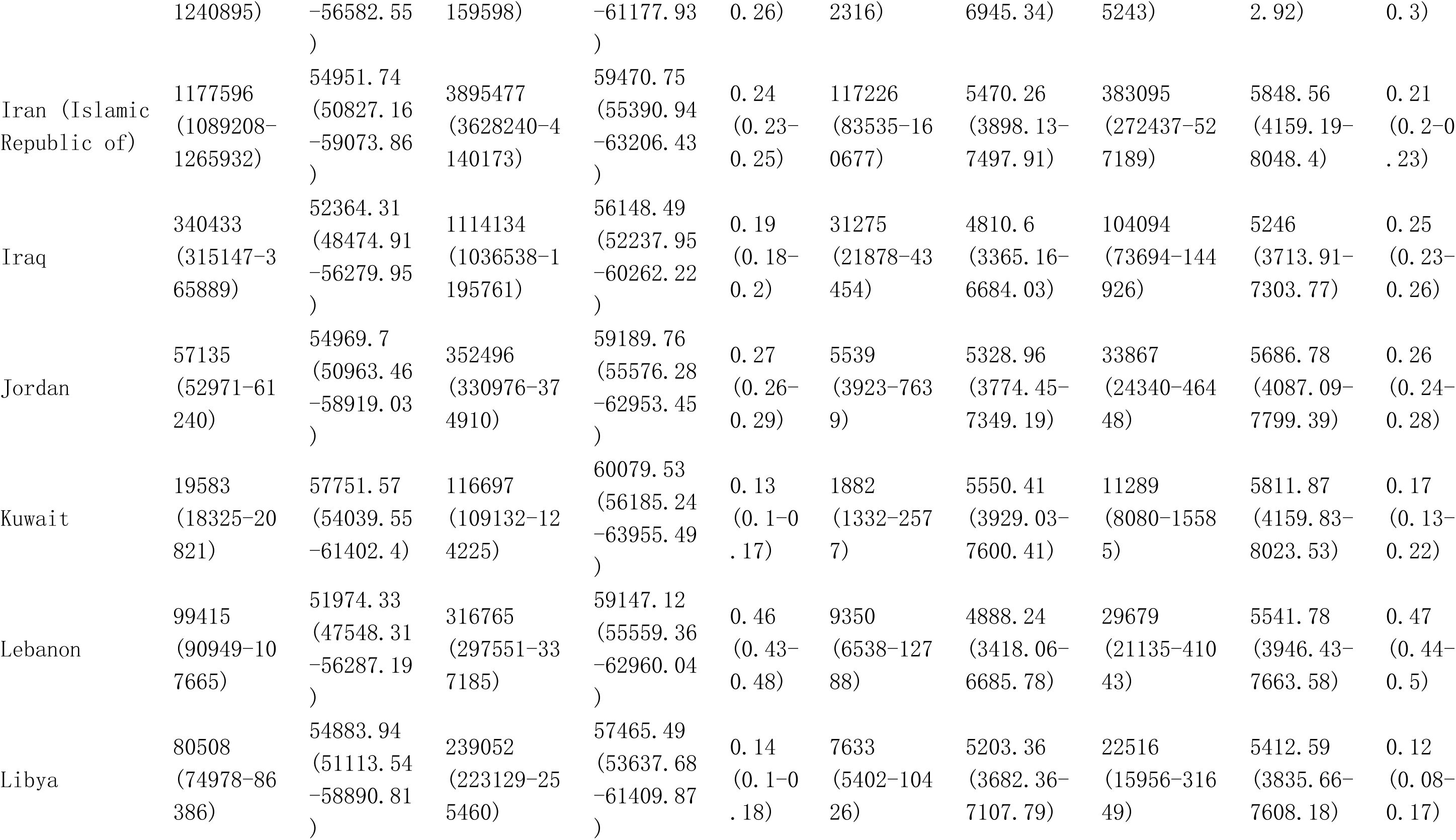
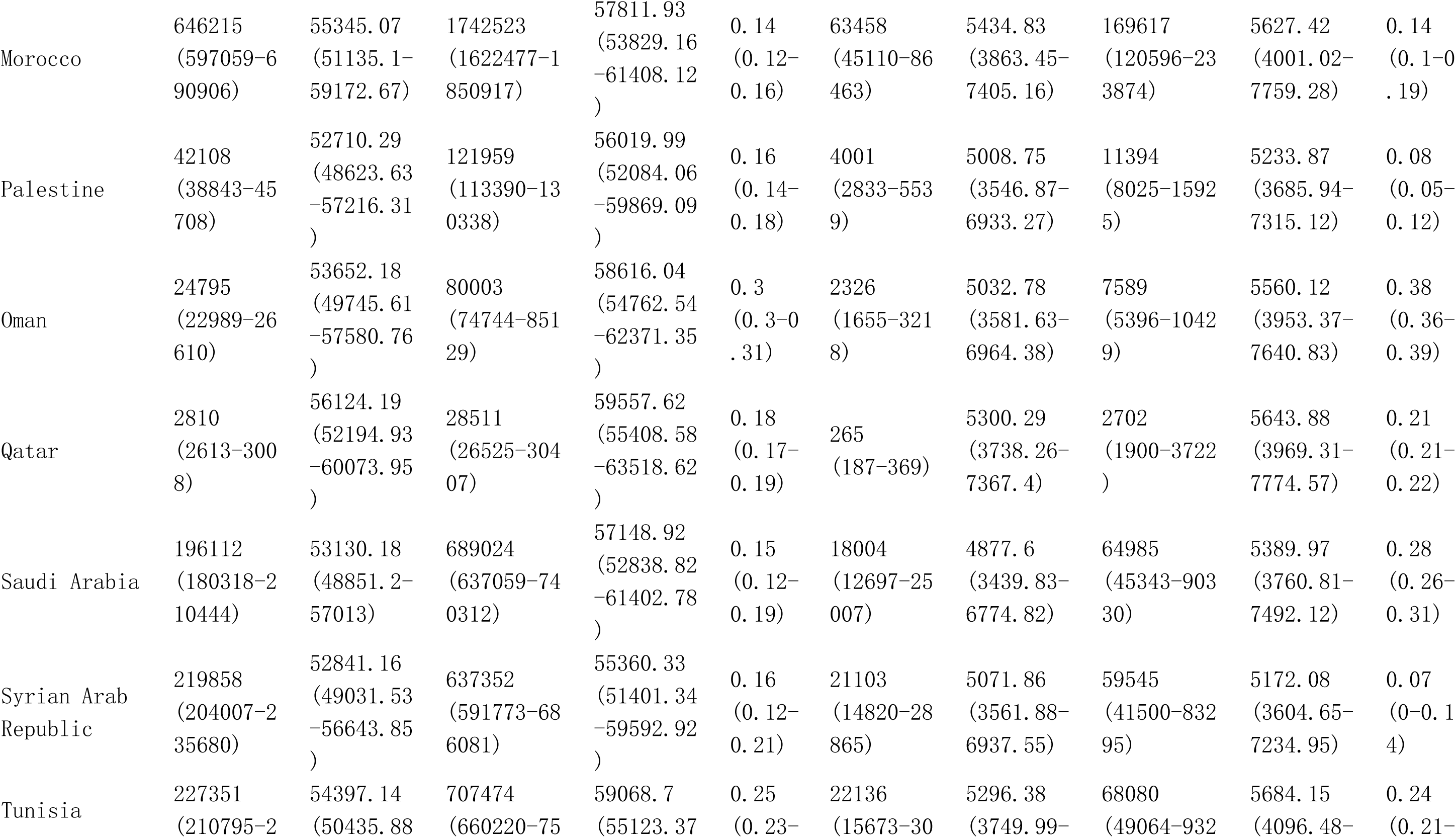
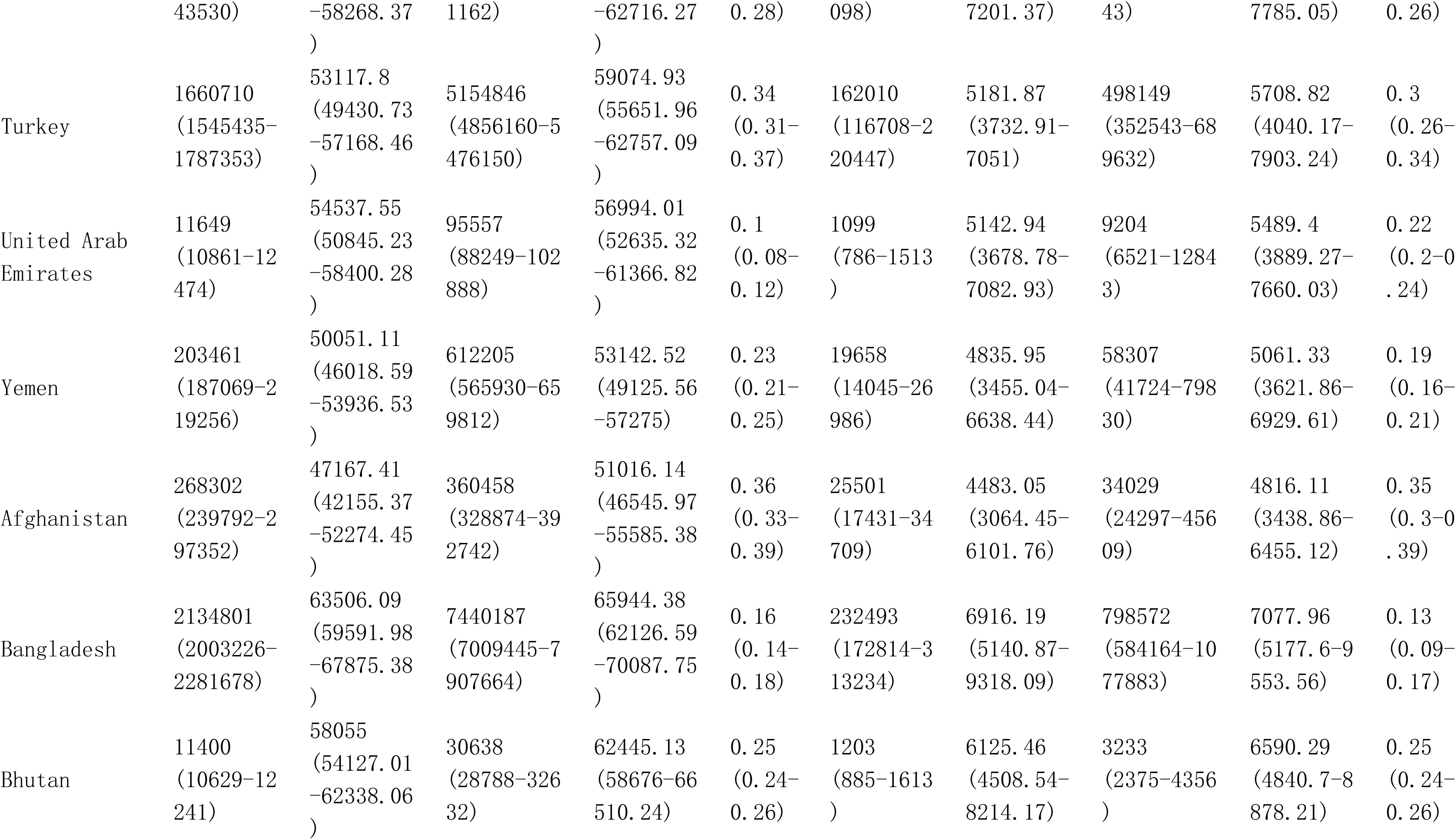
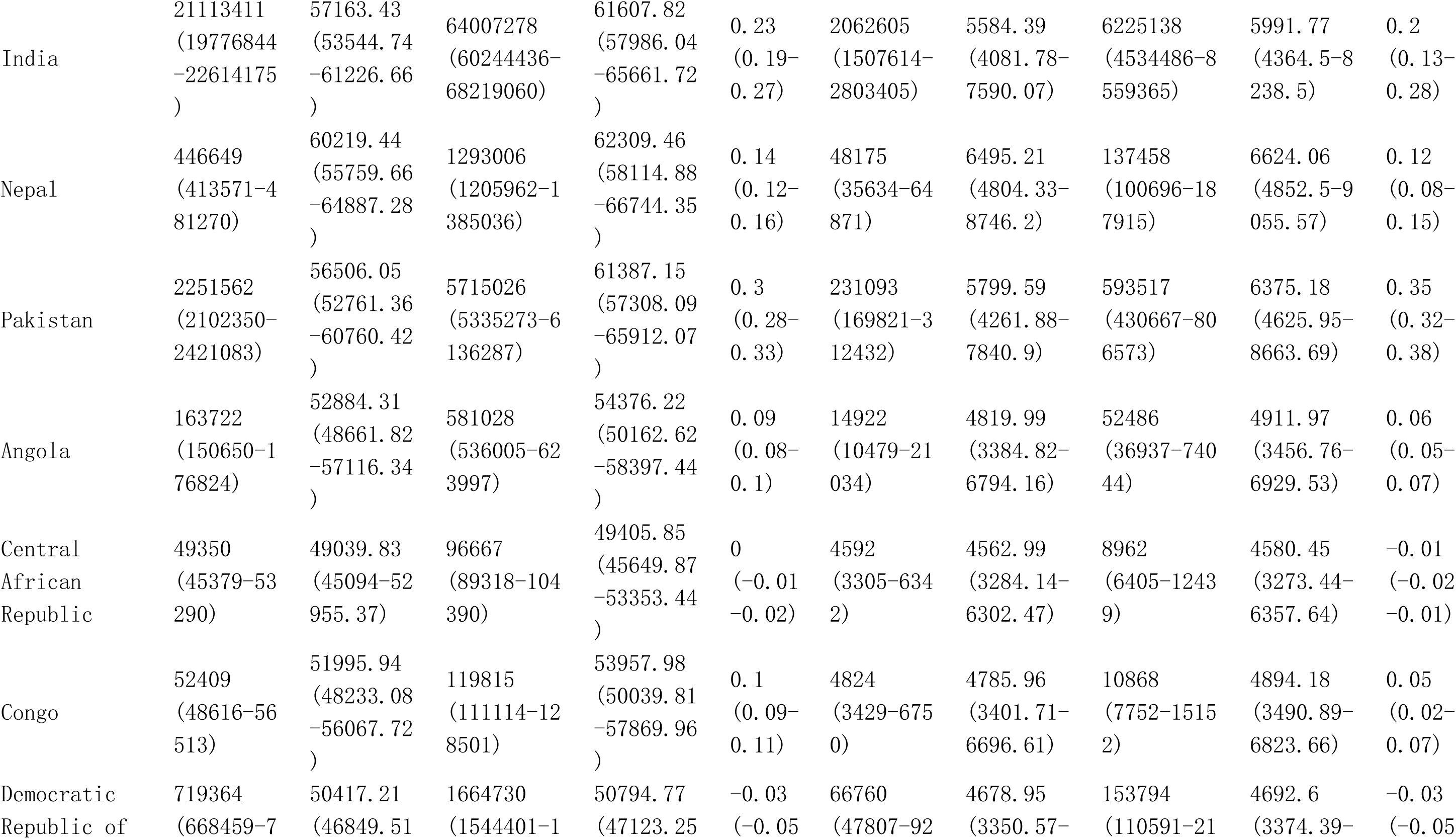
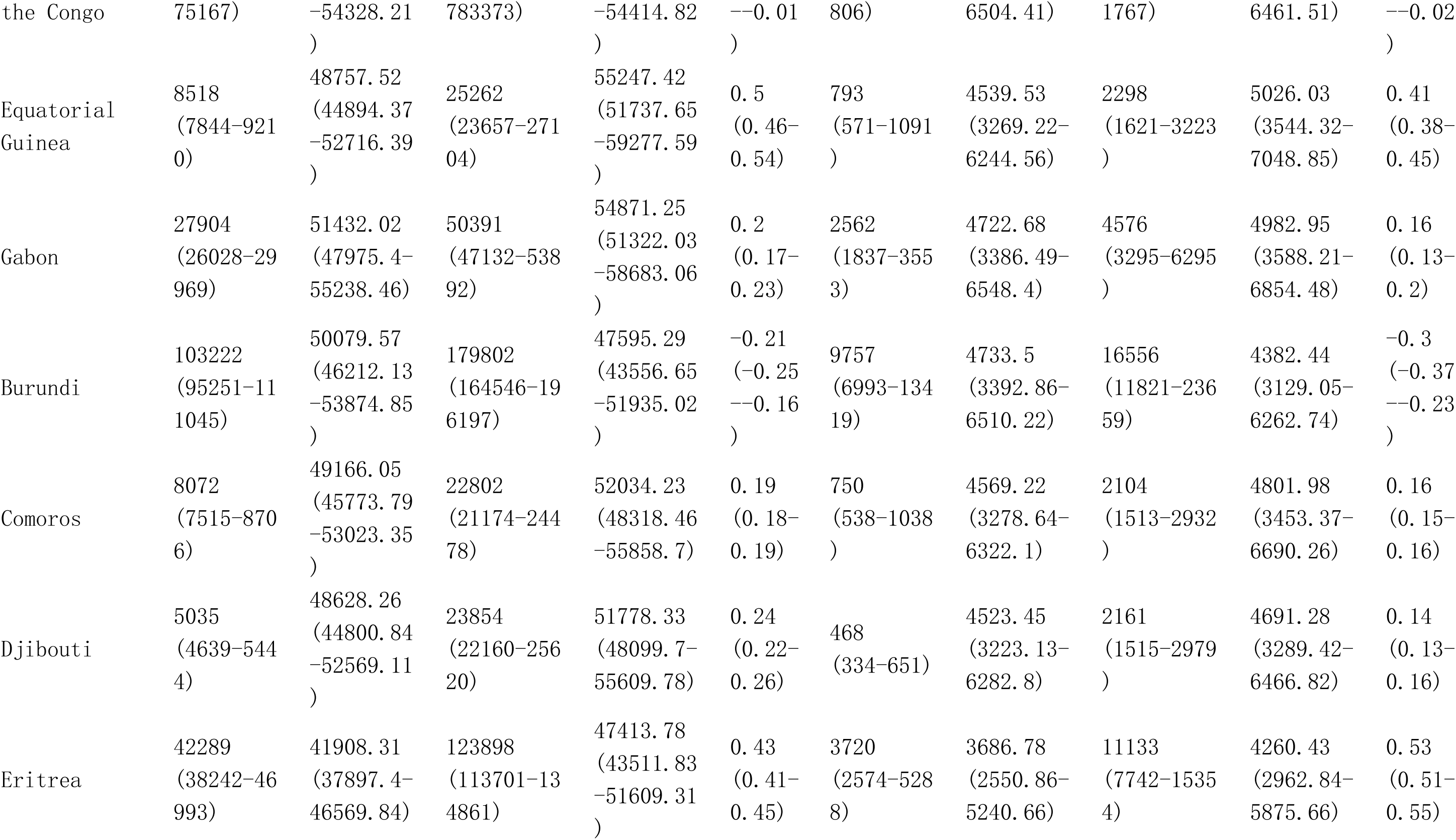
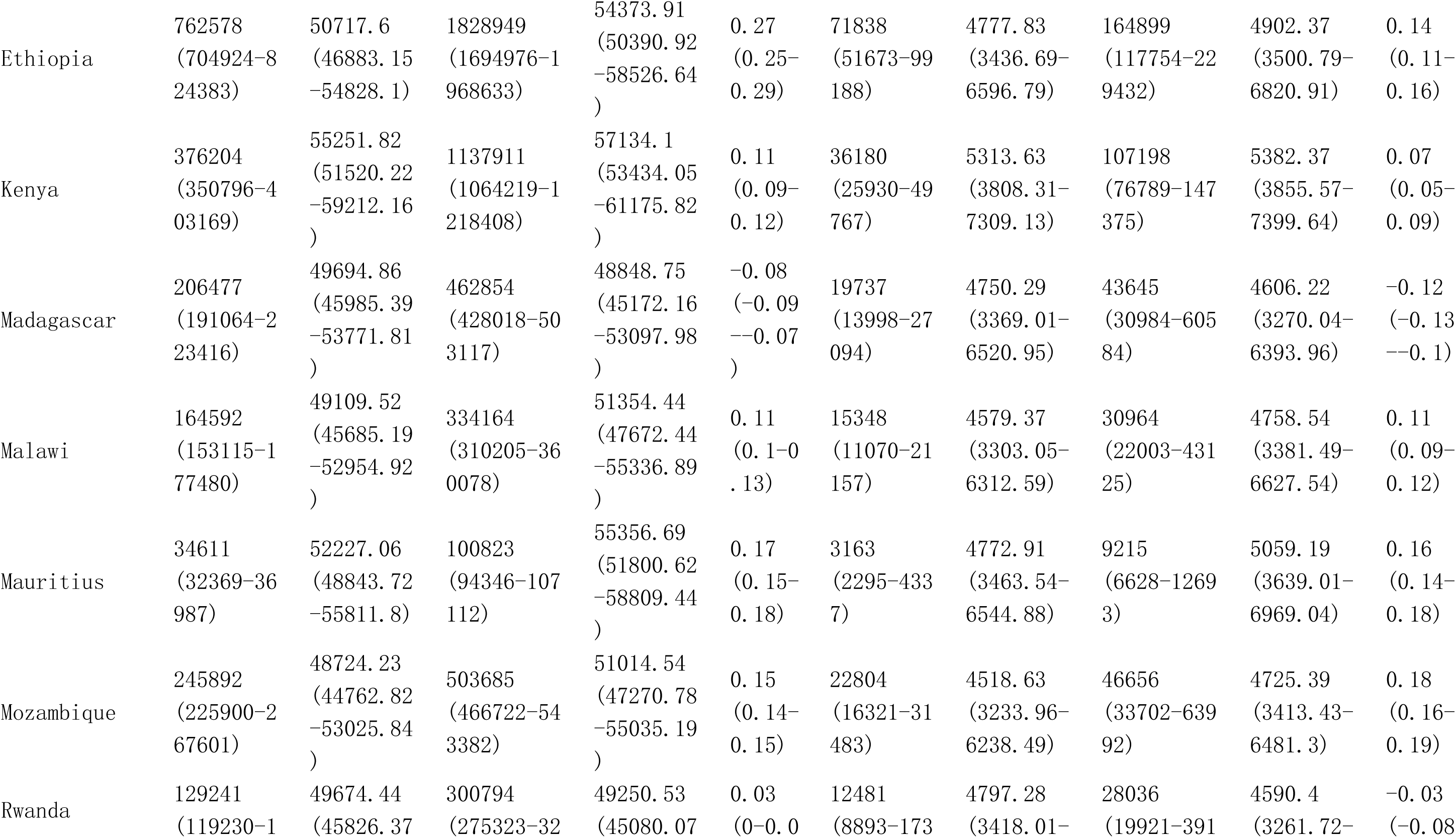
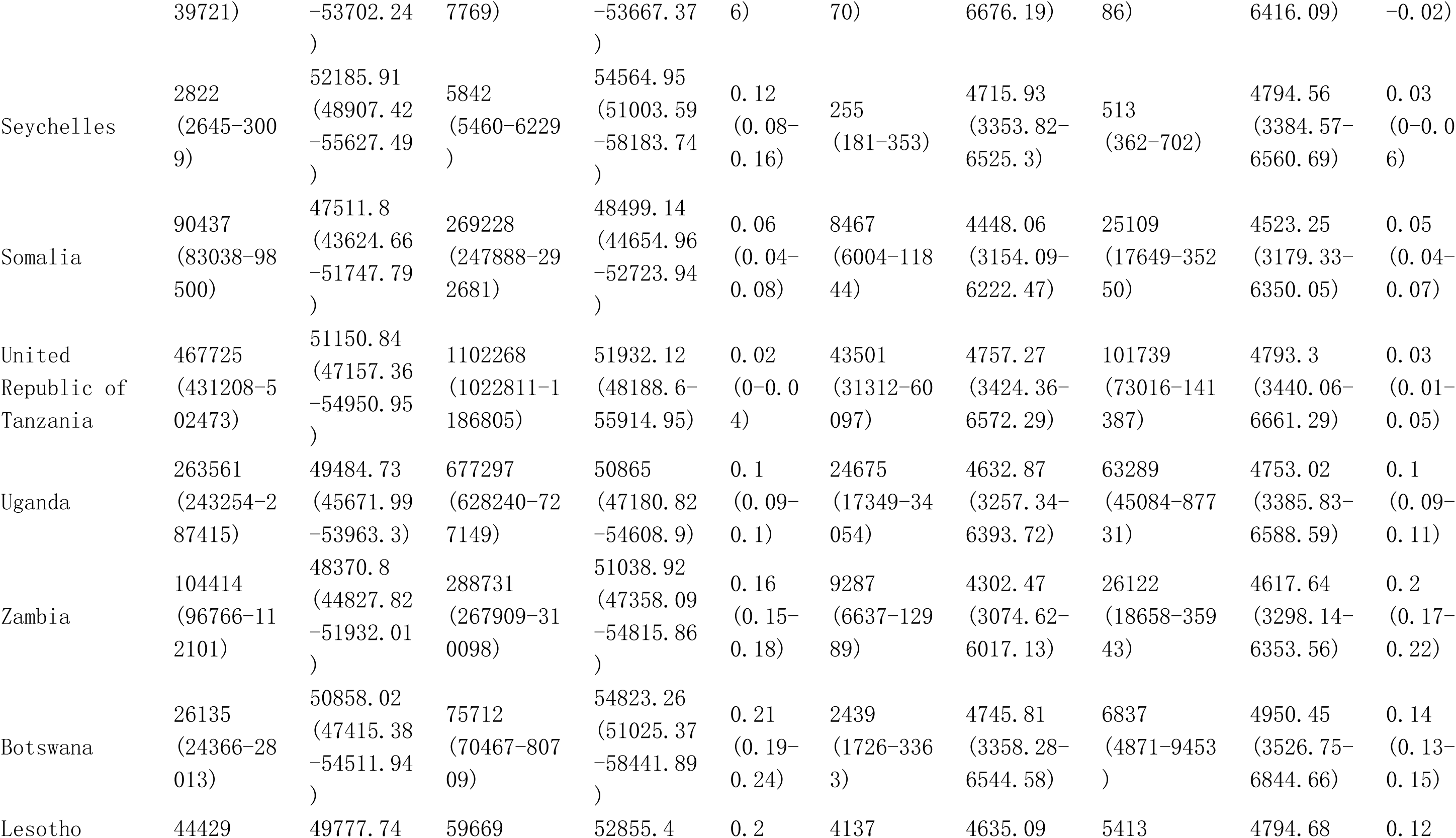
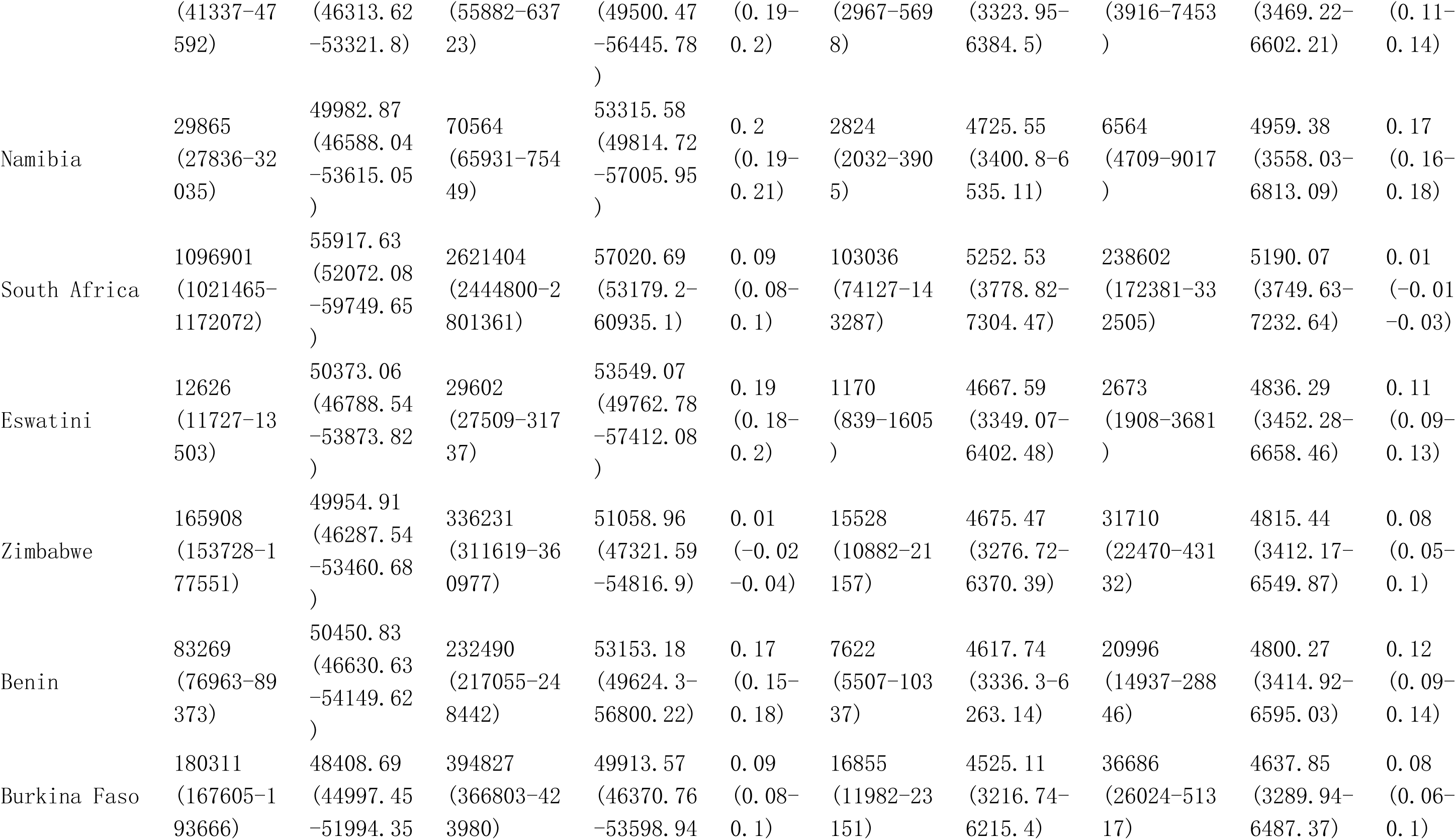
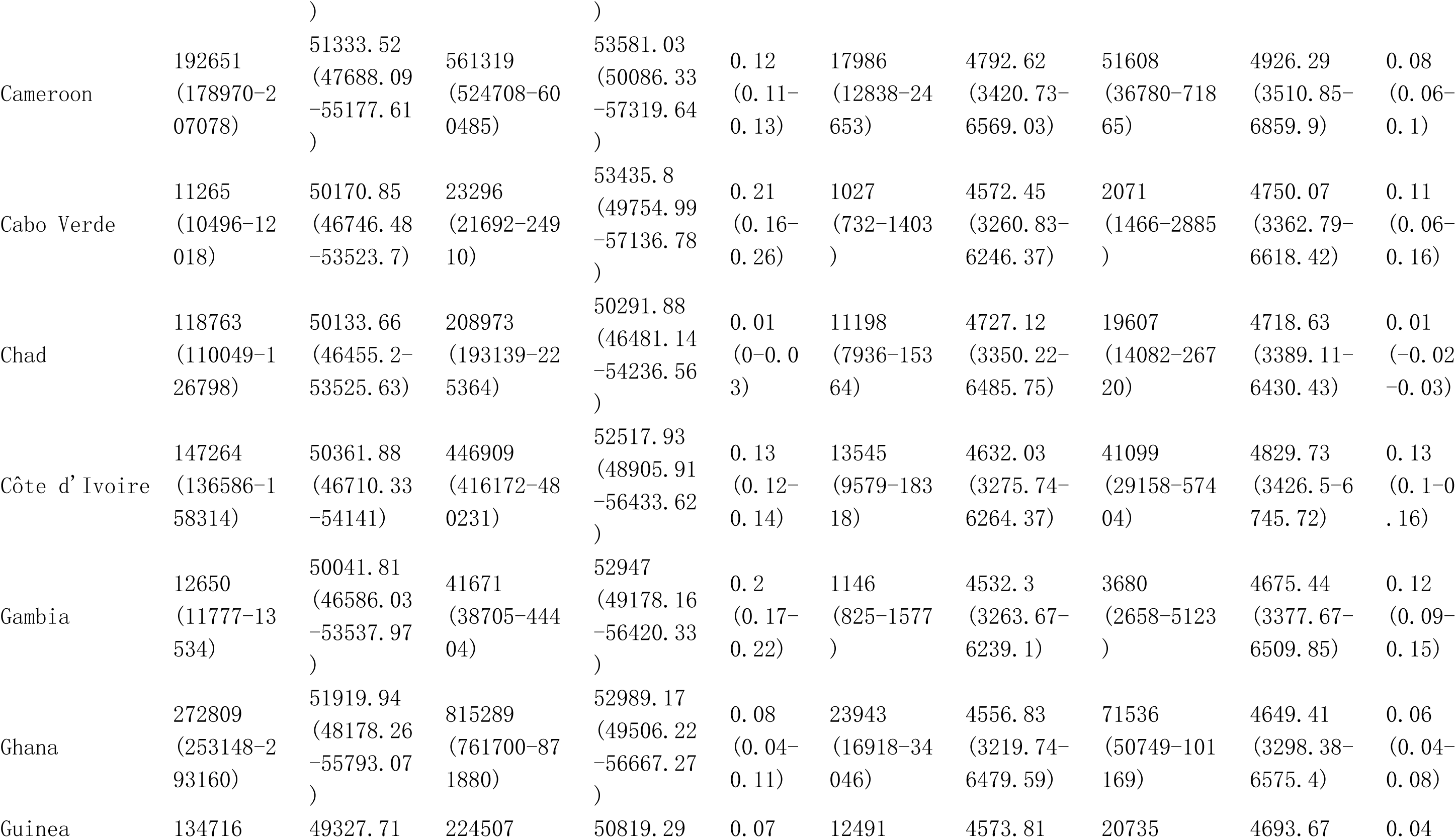
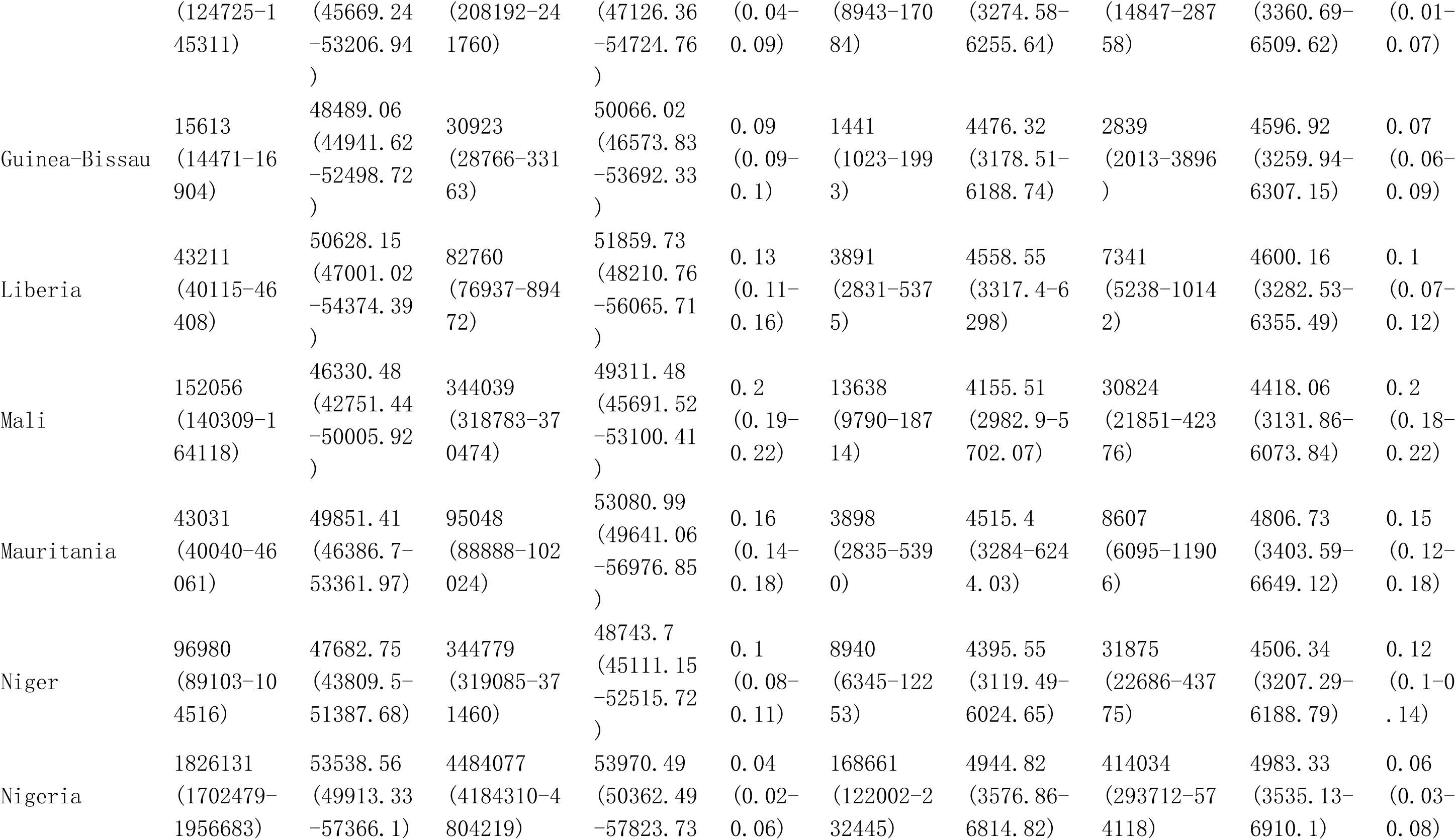
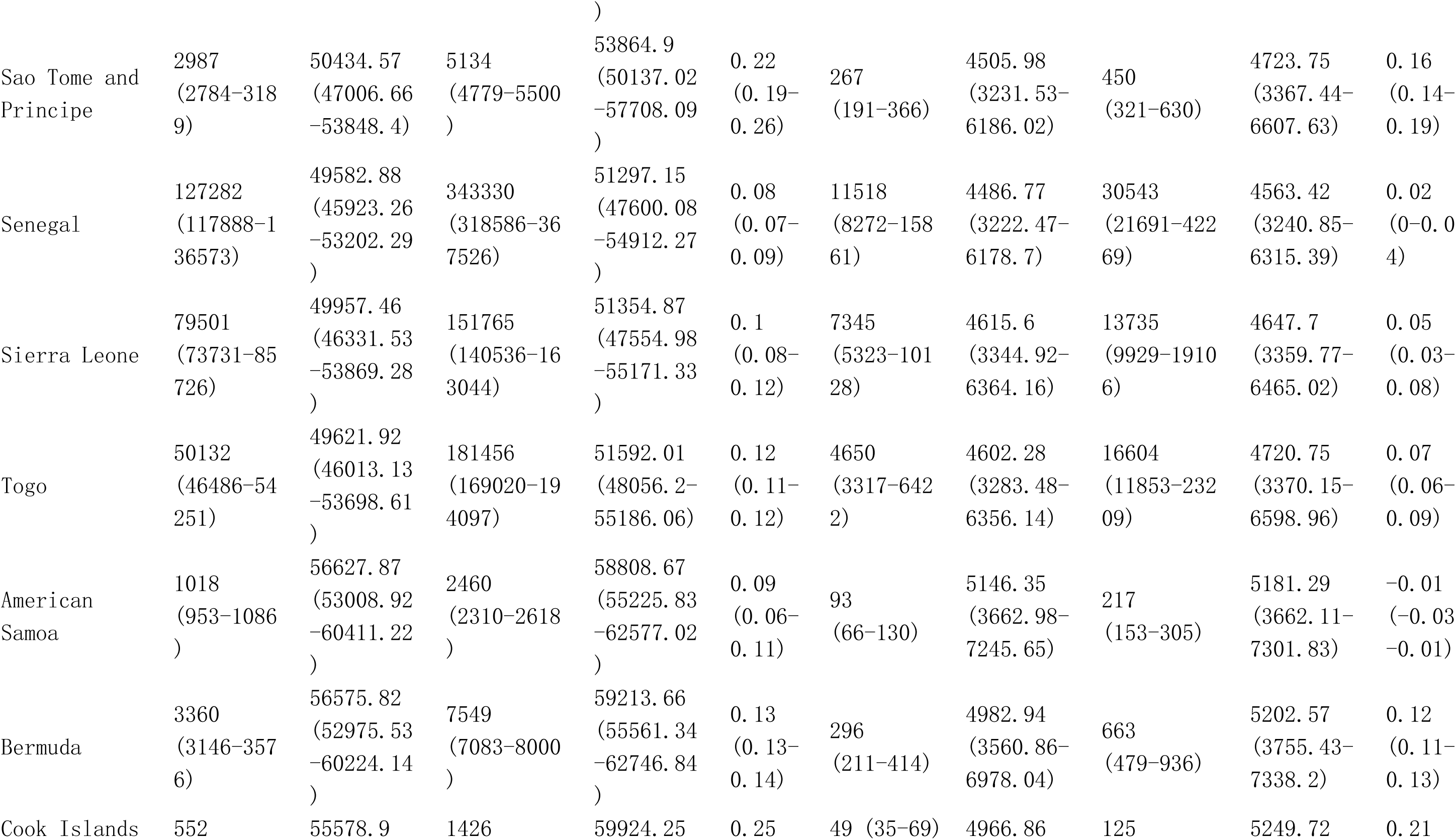
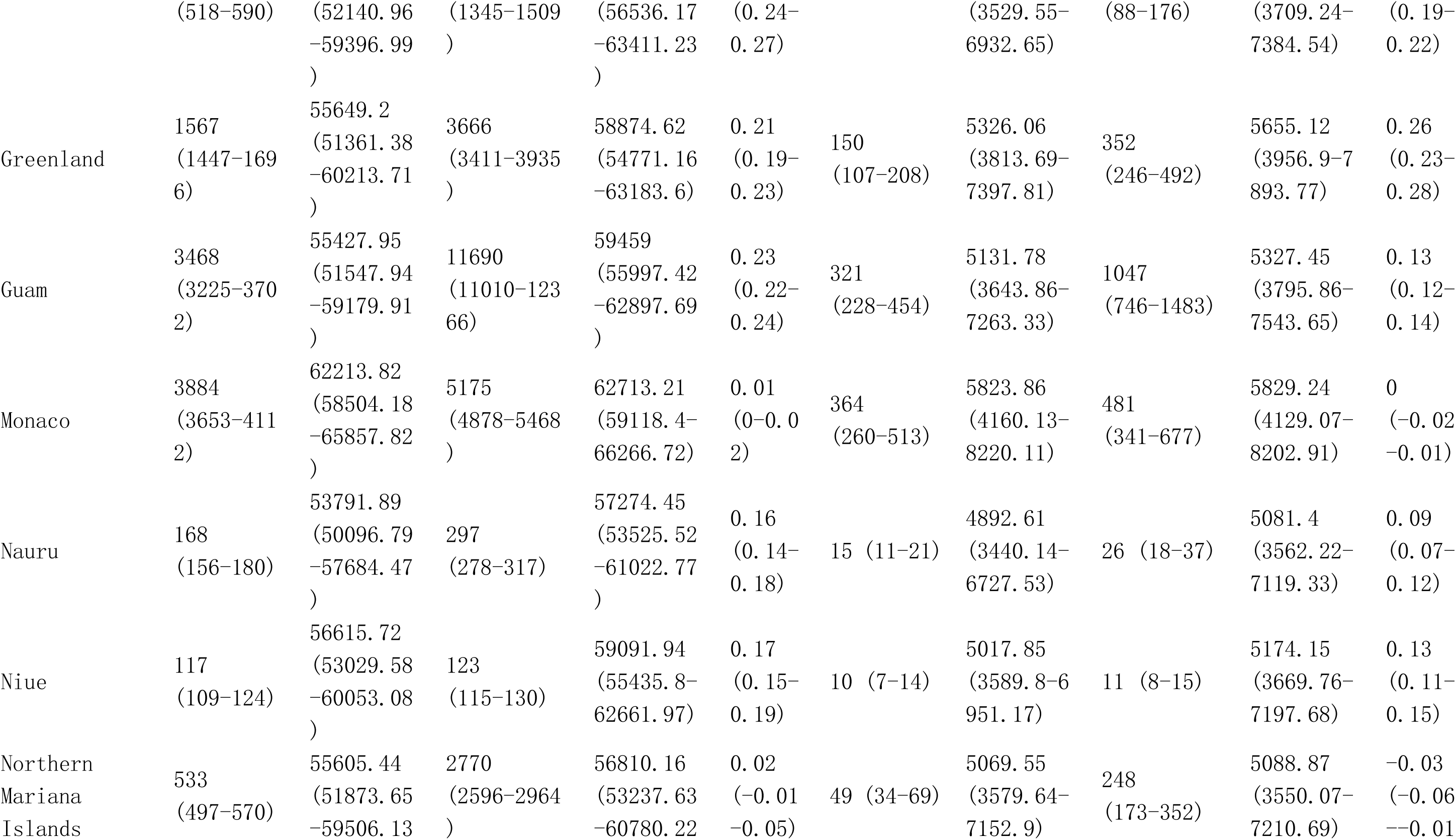
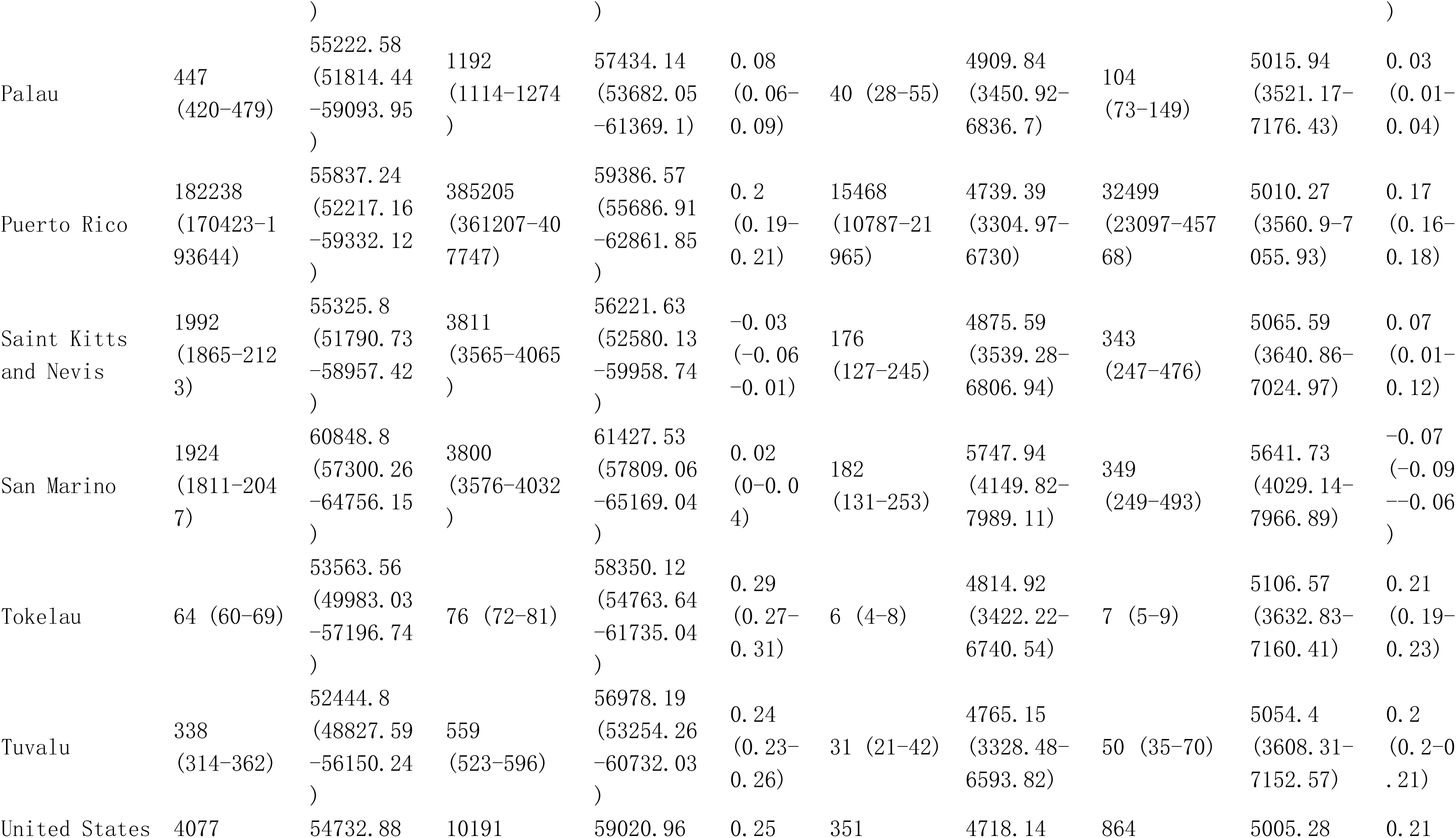
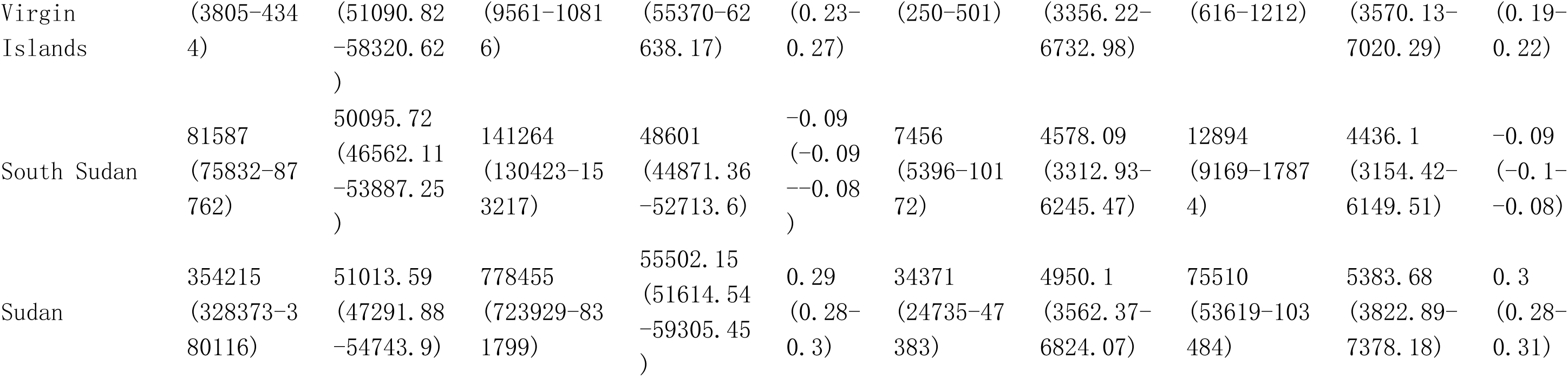
Global burden in Prevalence and DALYs of Musculoskeletal disorders among postmenopausal women from 1990 to 2021 by 21 GBD geographical regions, and 204 countries and territories.

**Table S3.**
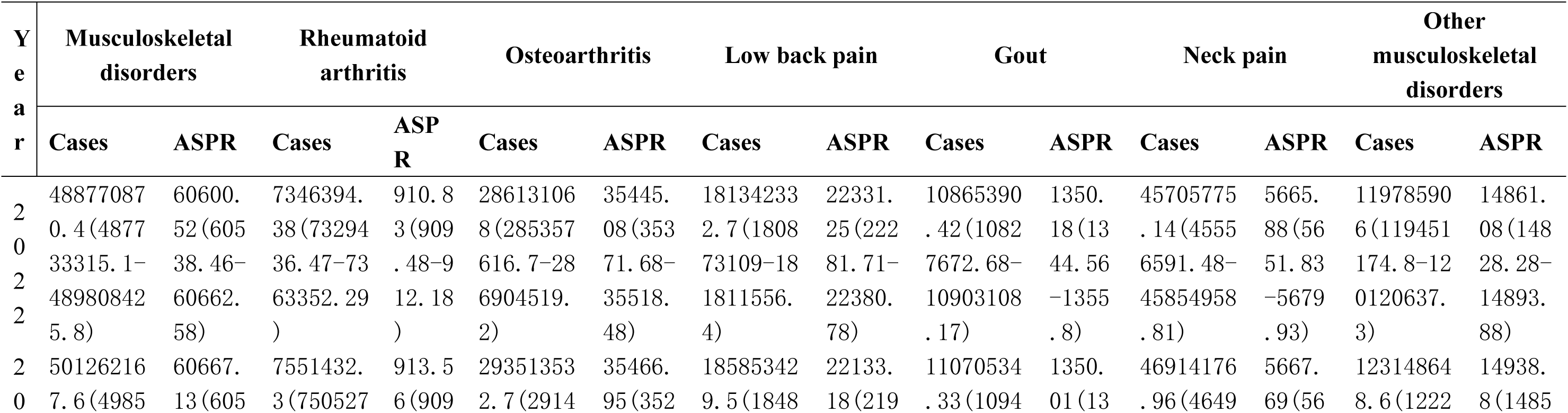
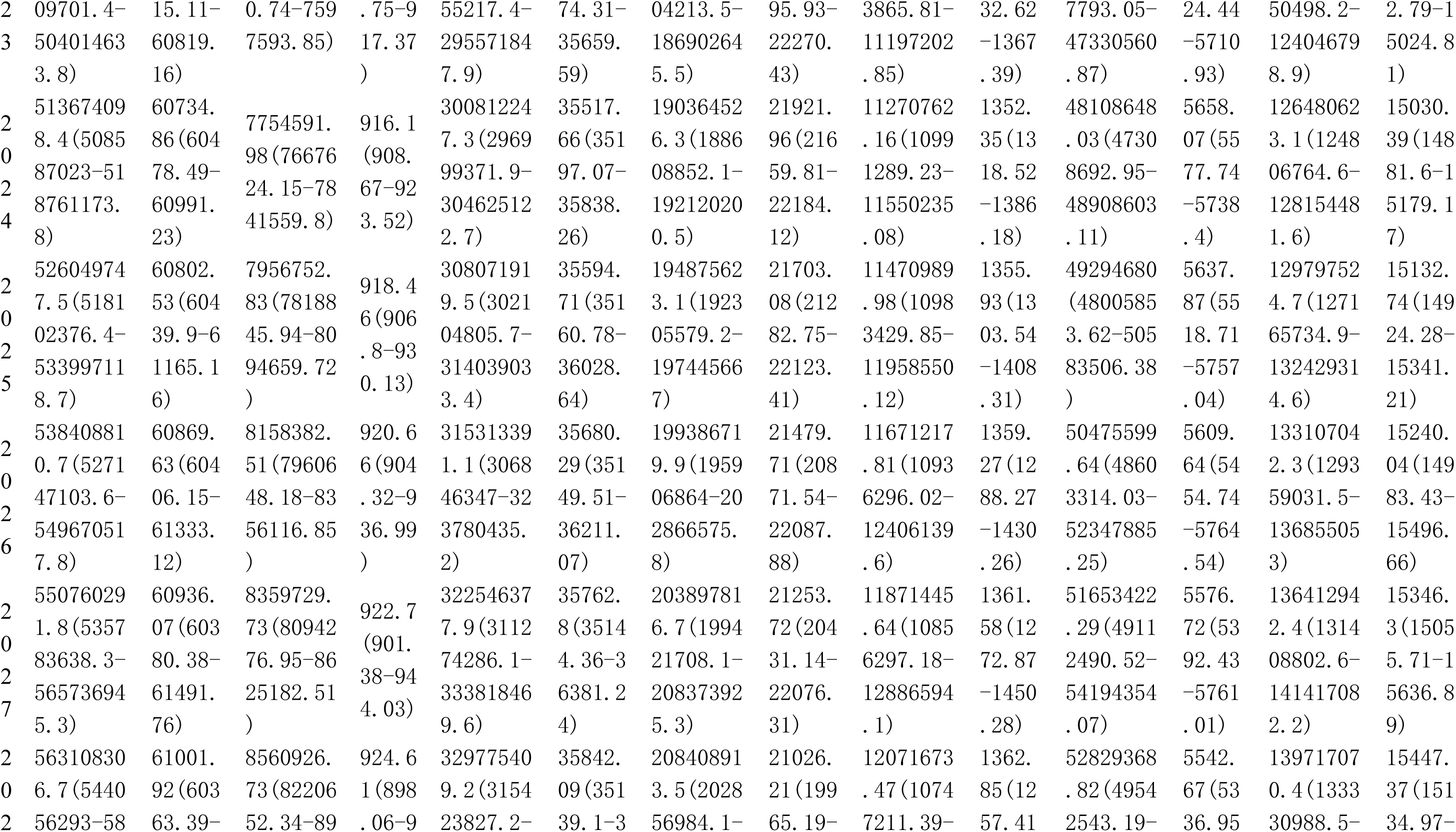
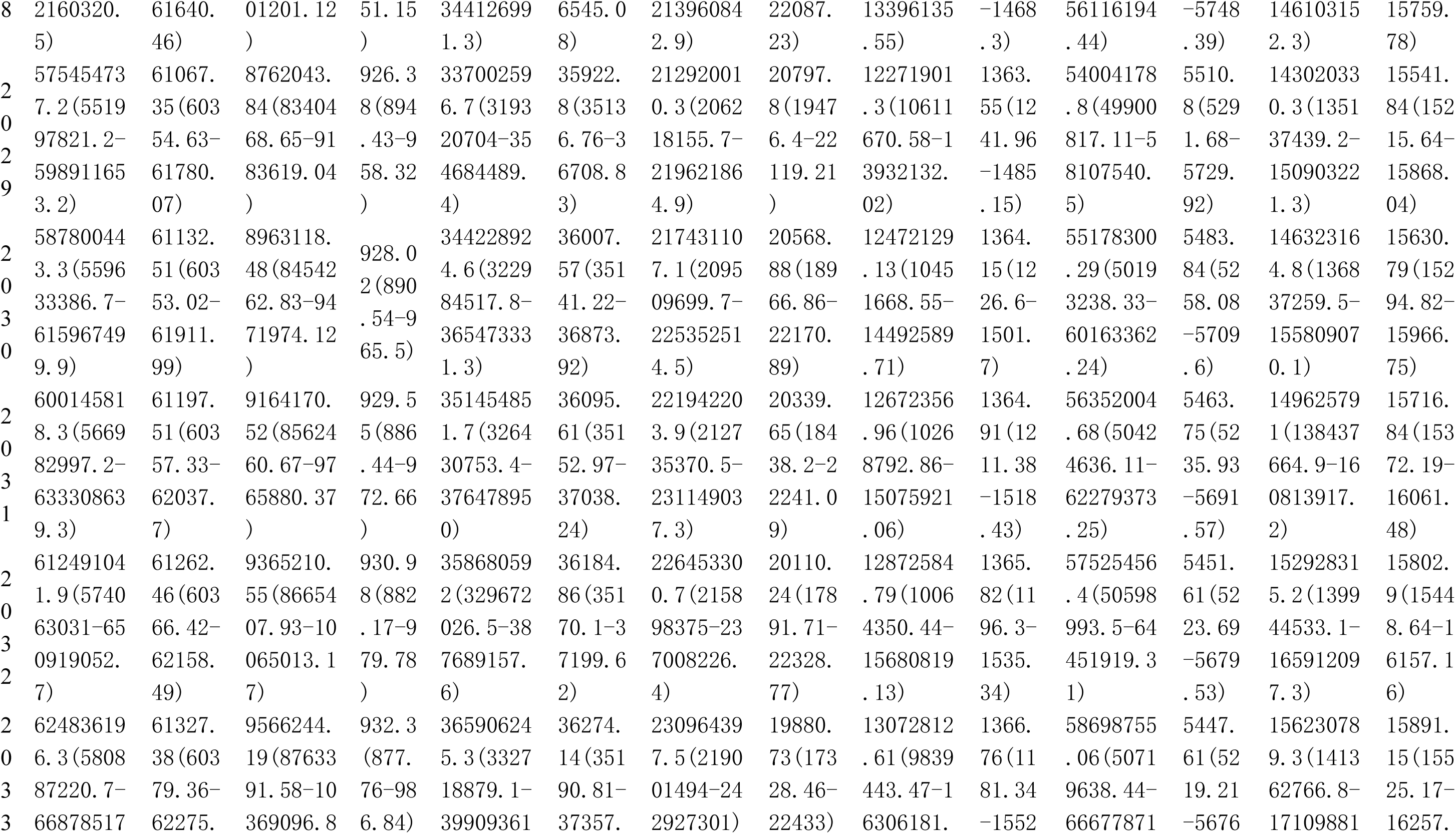
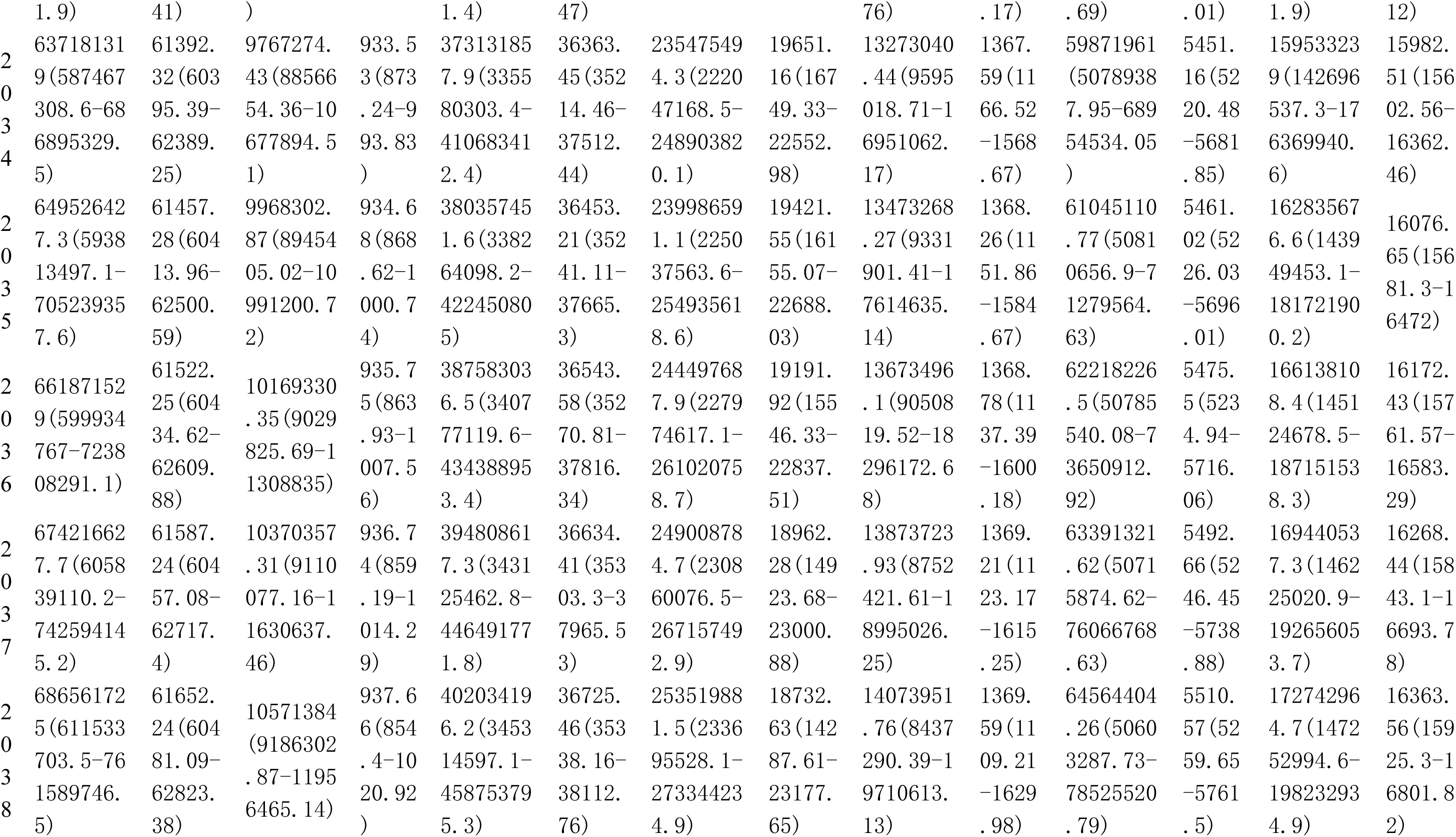
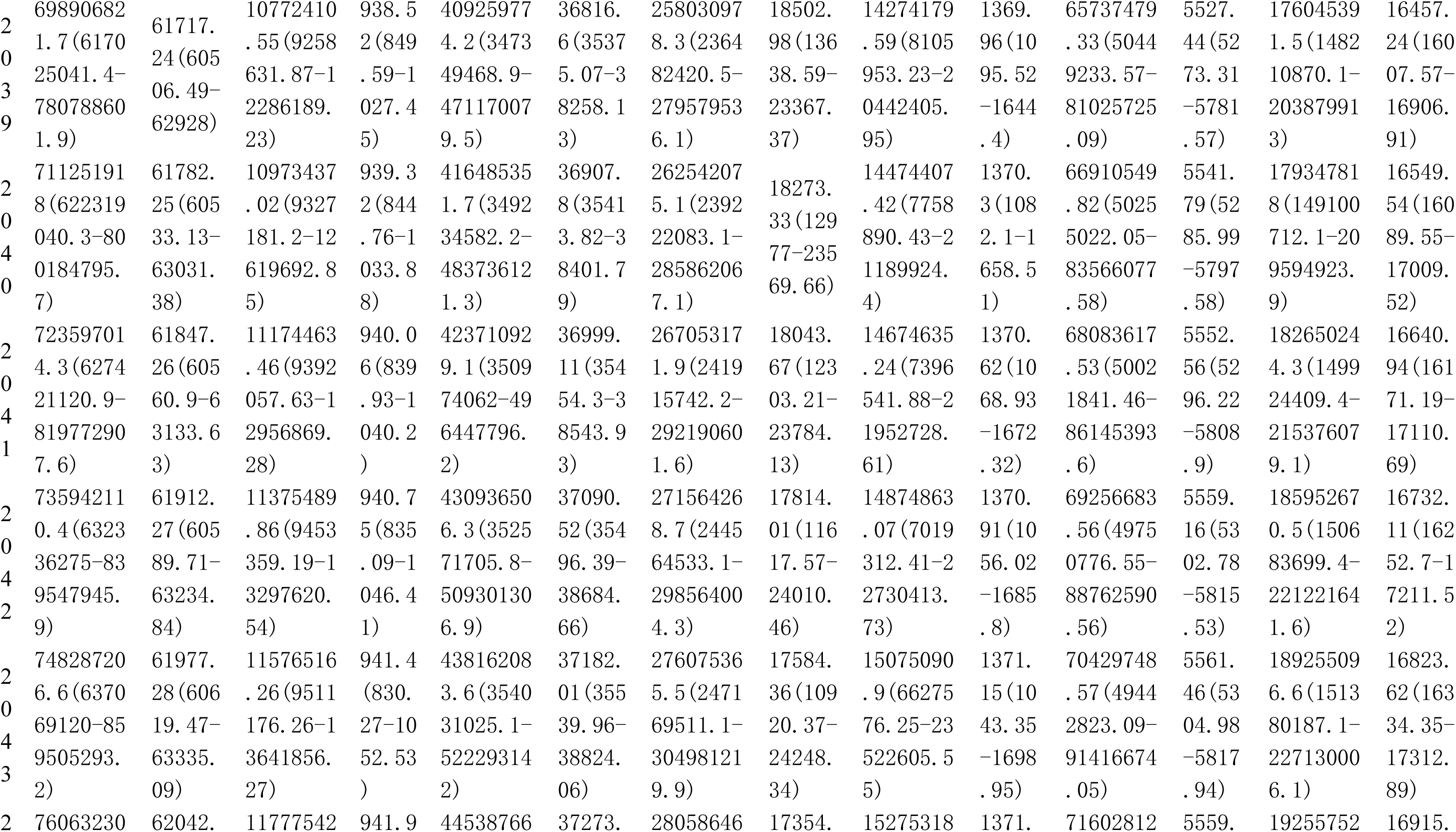
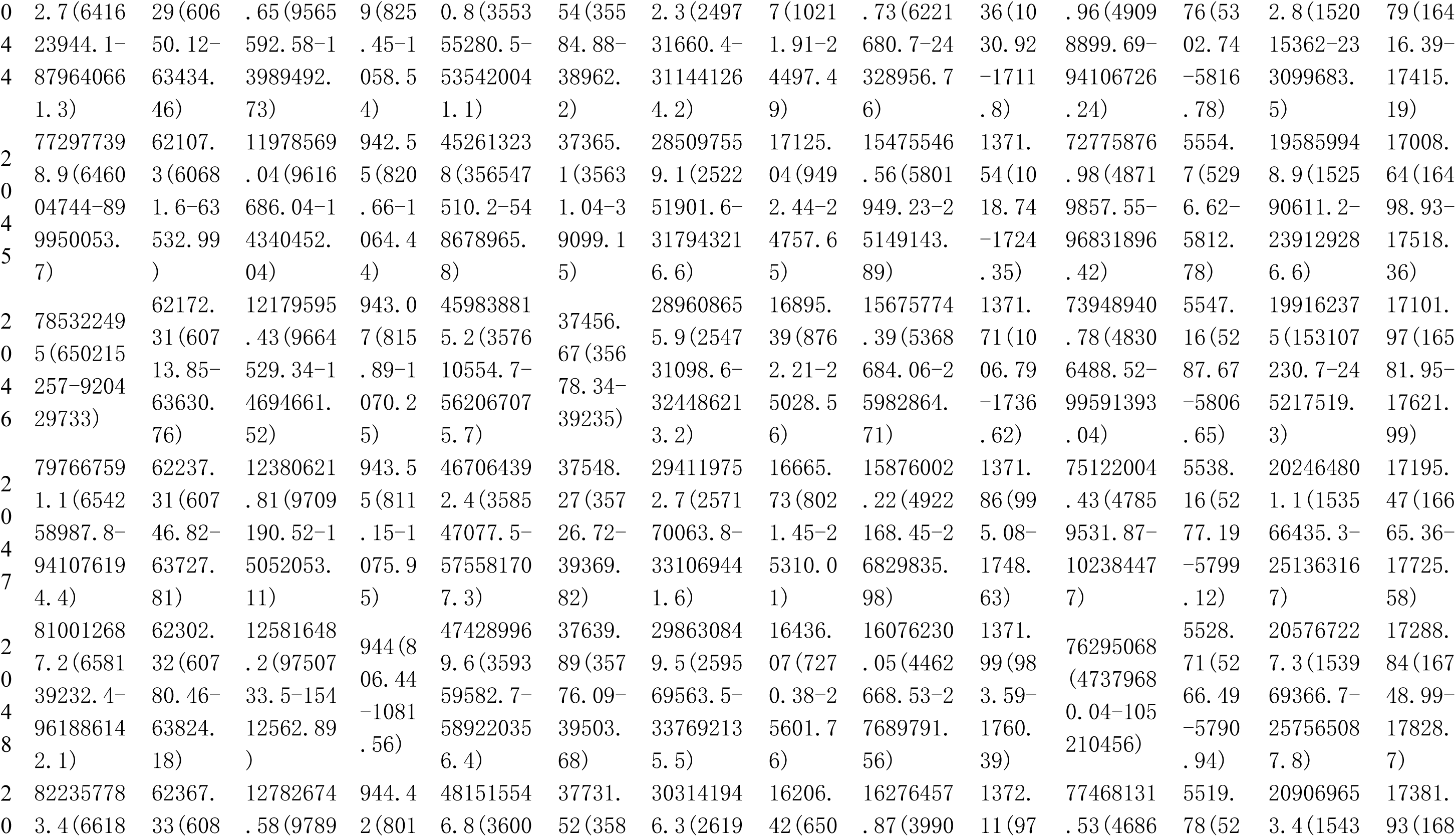
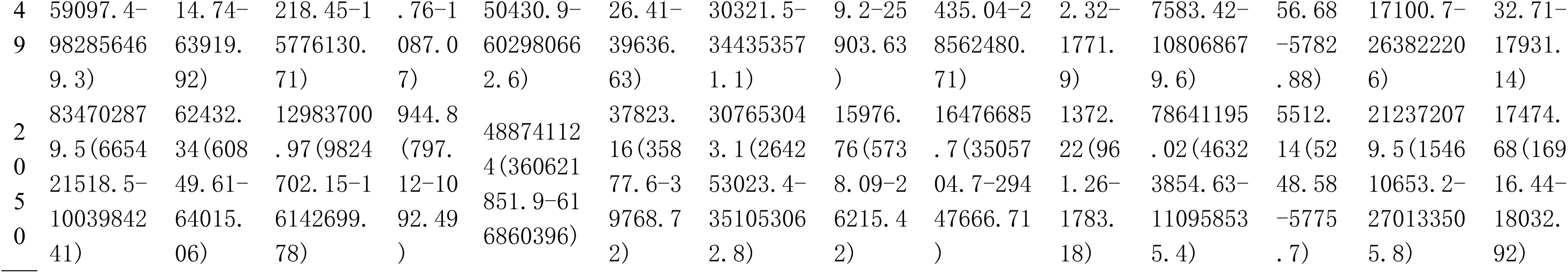
Predictive analysis of Prevalence among postmenopausal women from 2022 to 2050.

**Table S4.**
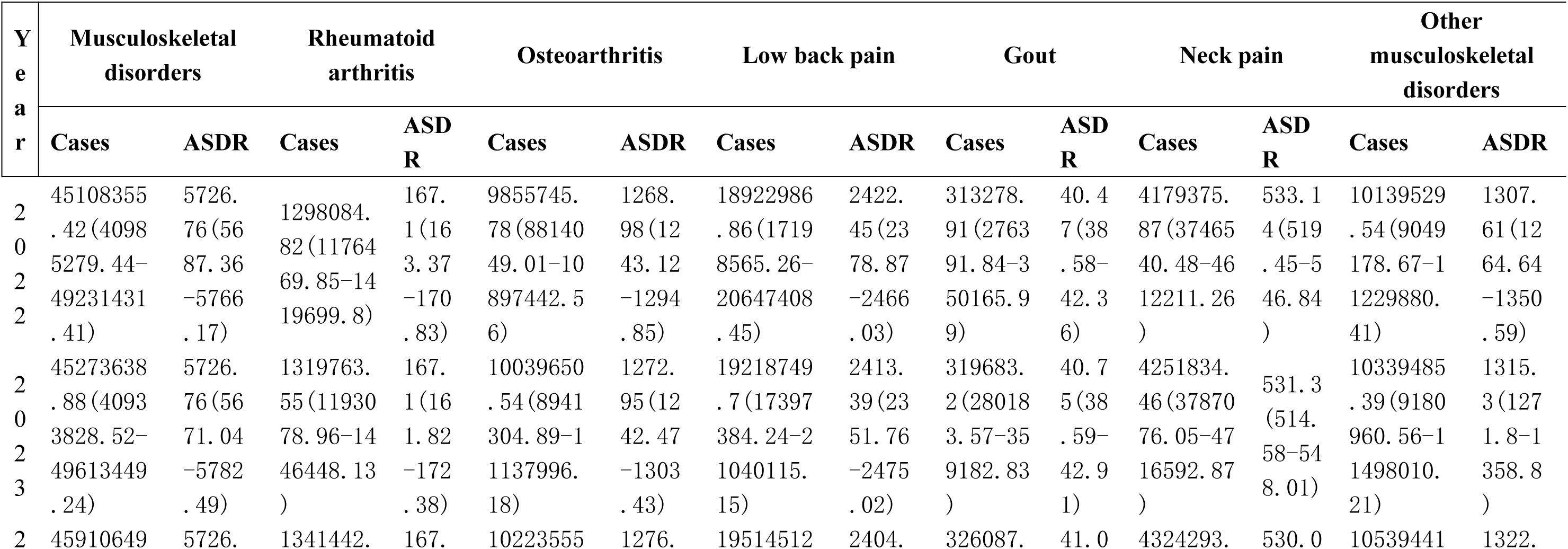
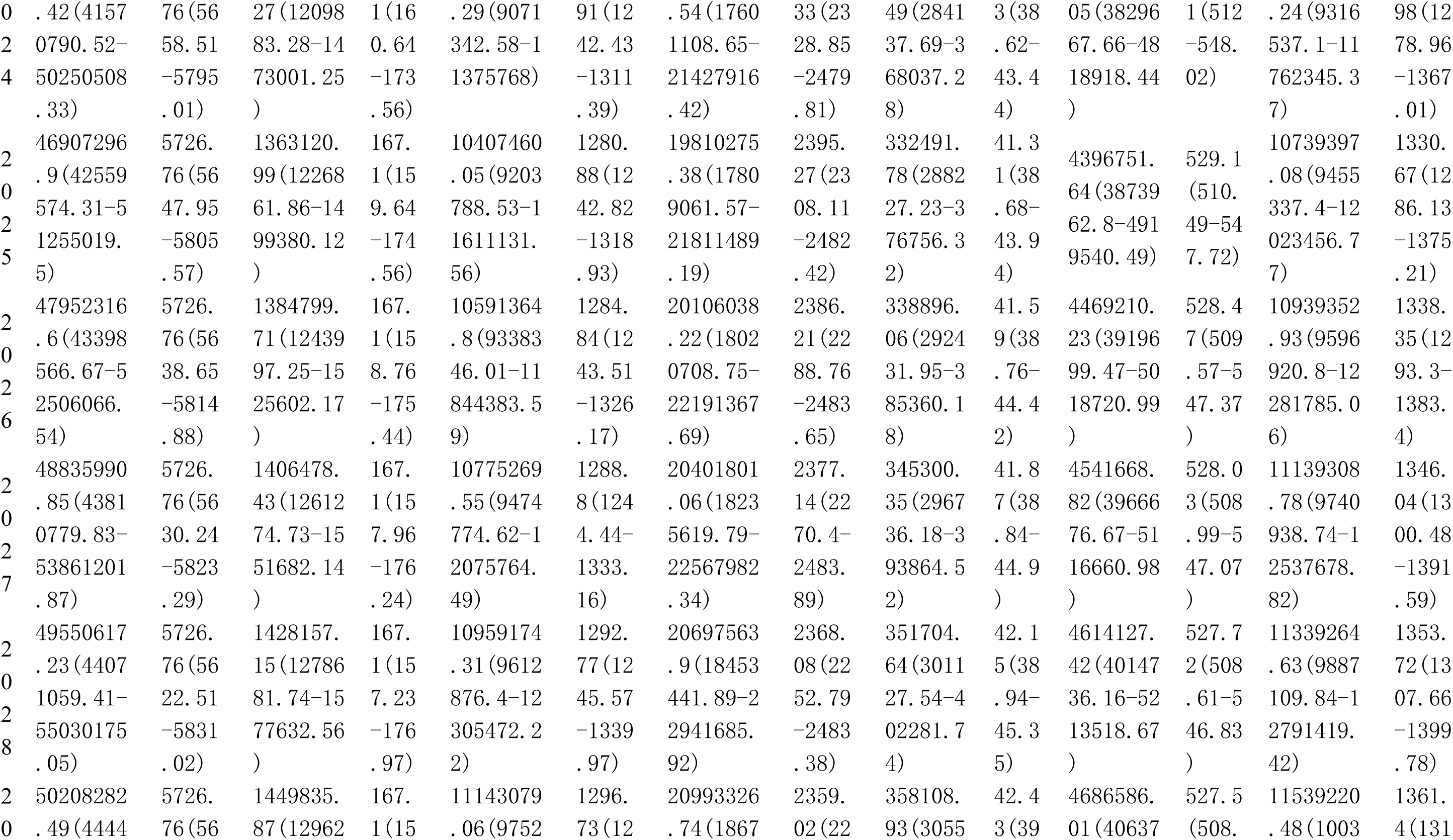
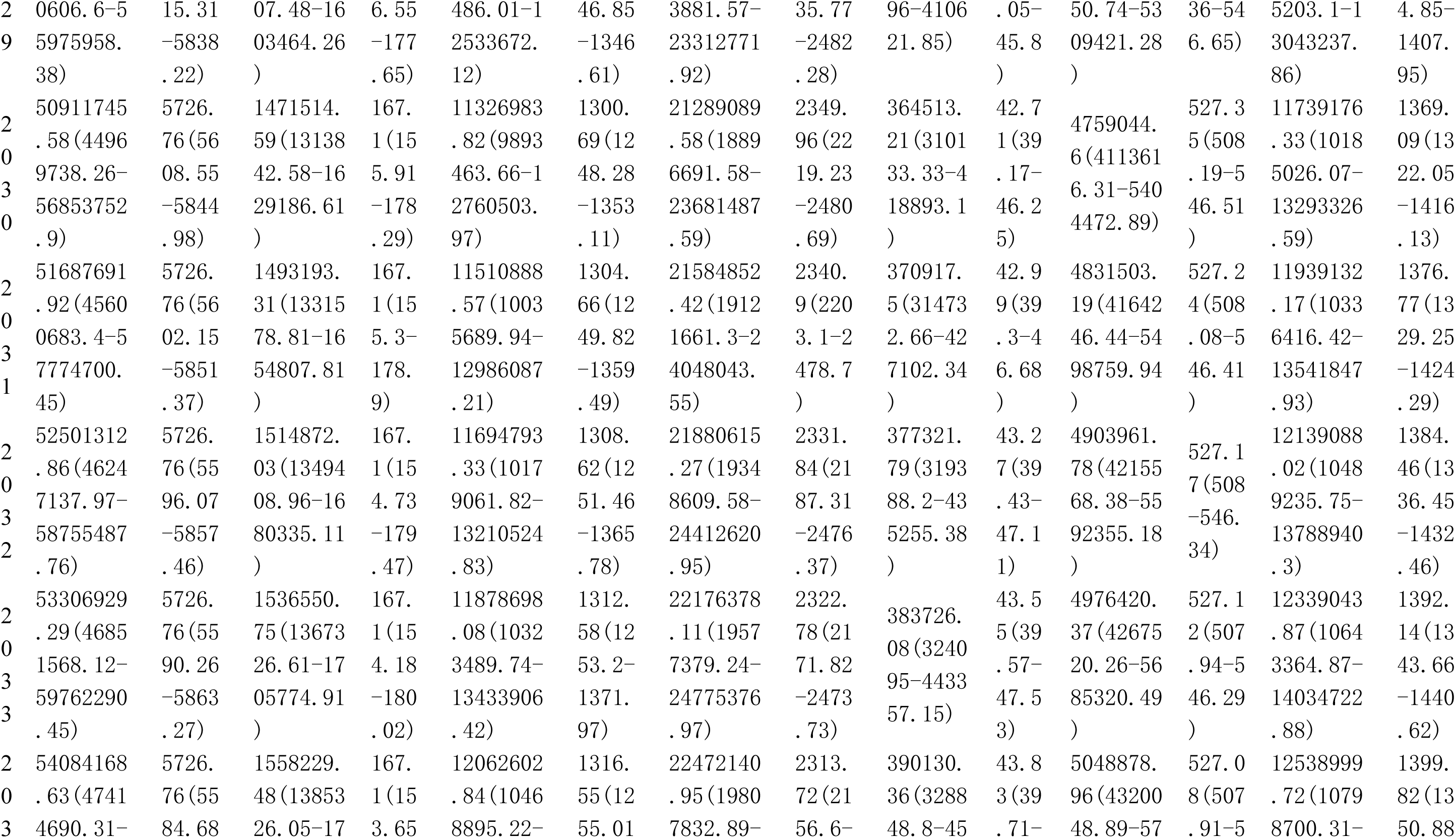
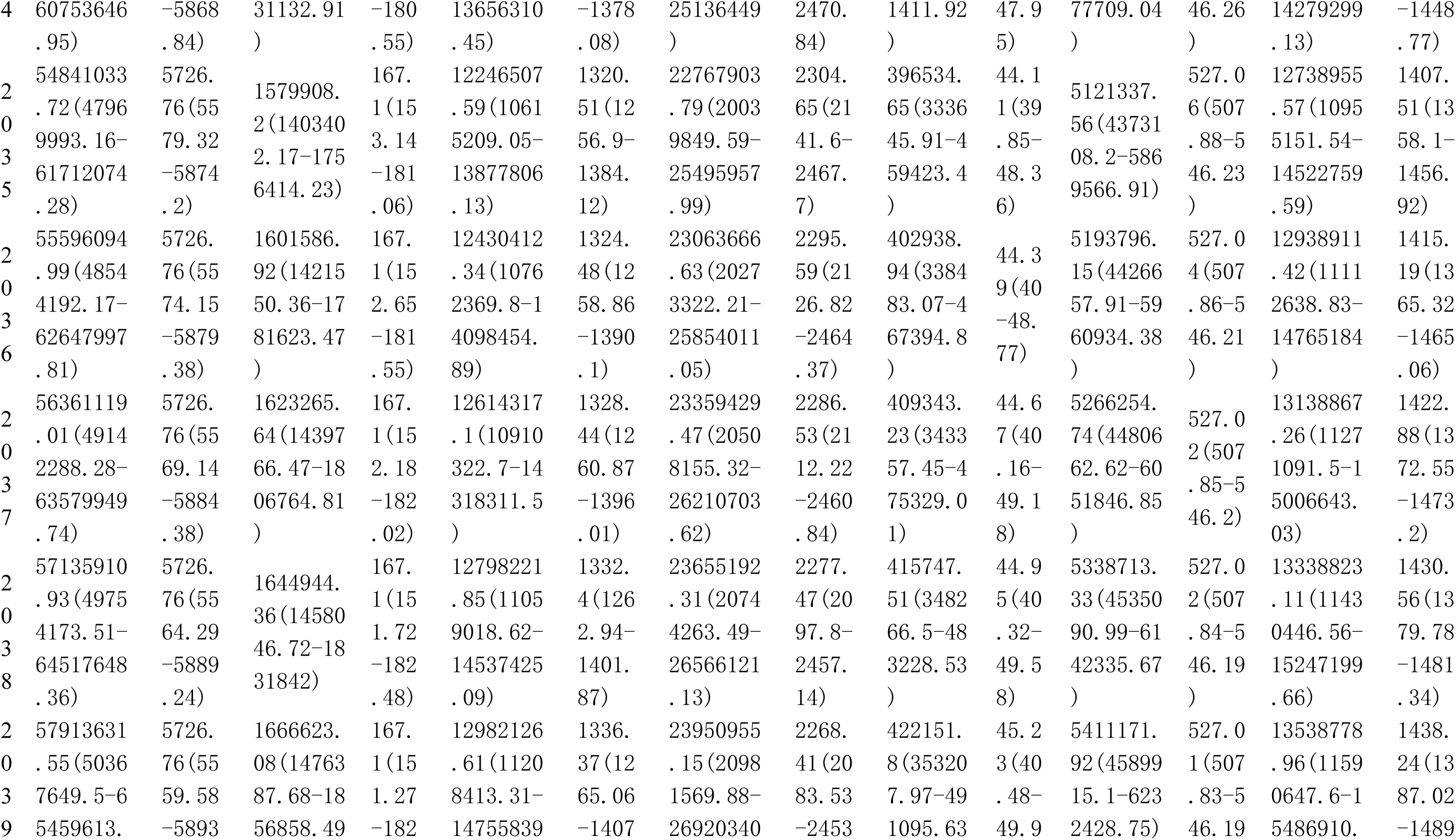
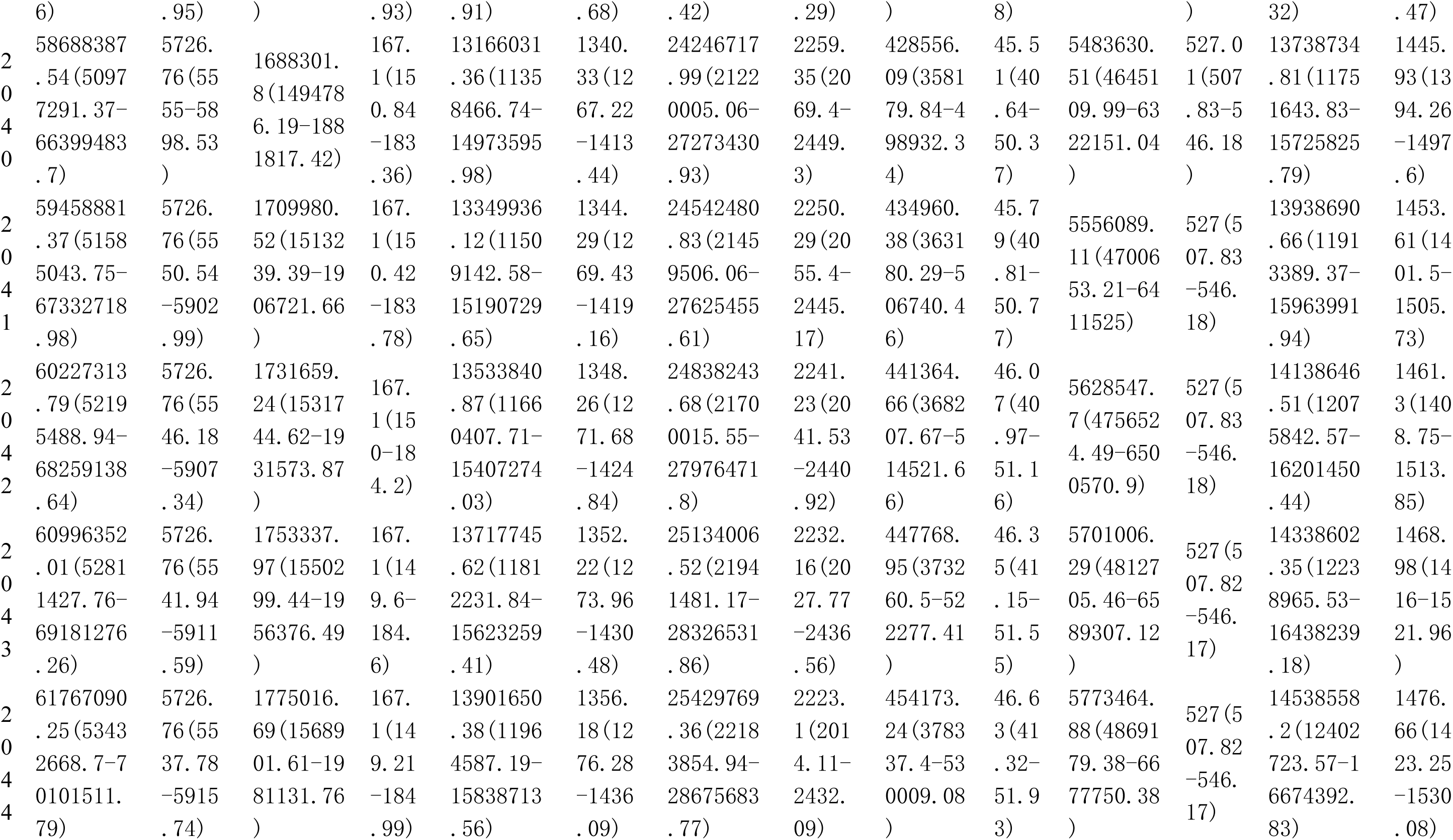
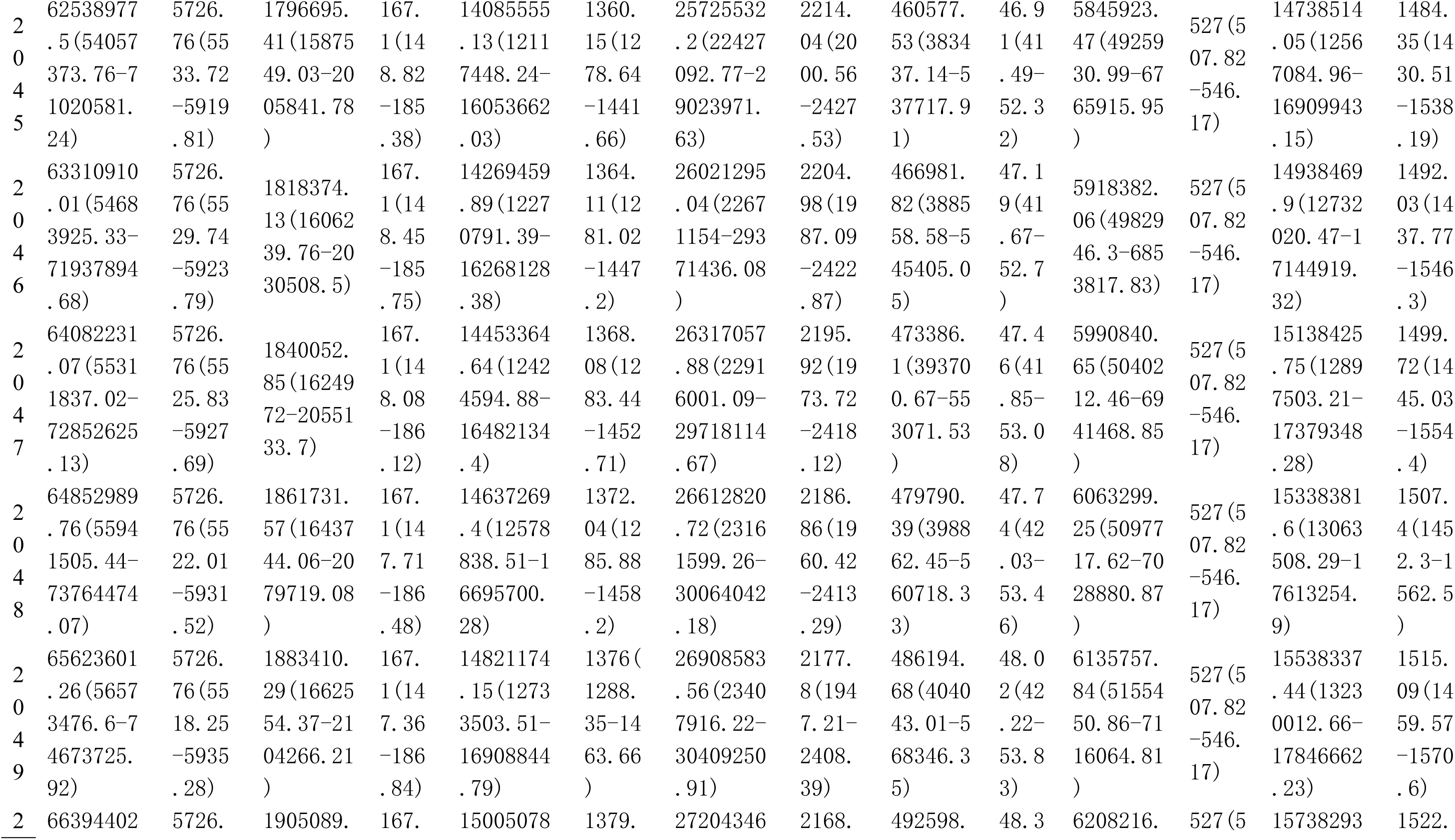
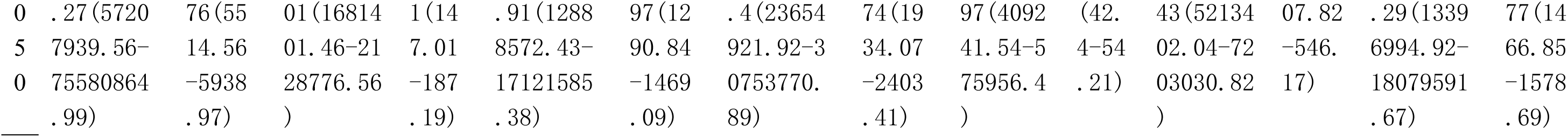
Predictive analysis of DALYs among postmenopausal women from 2022 to 2050.

**Table S5.**
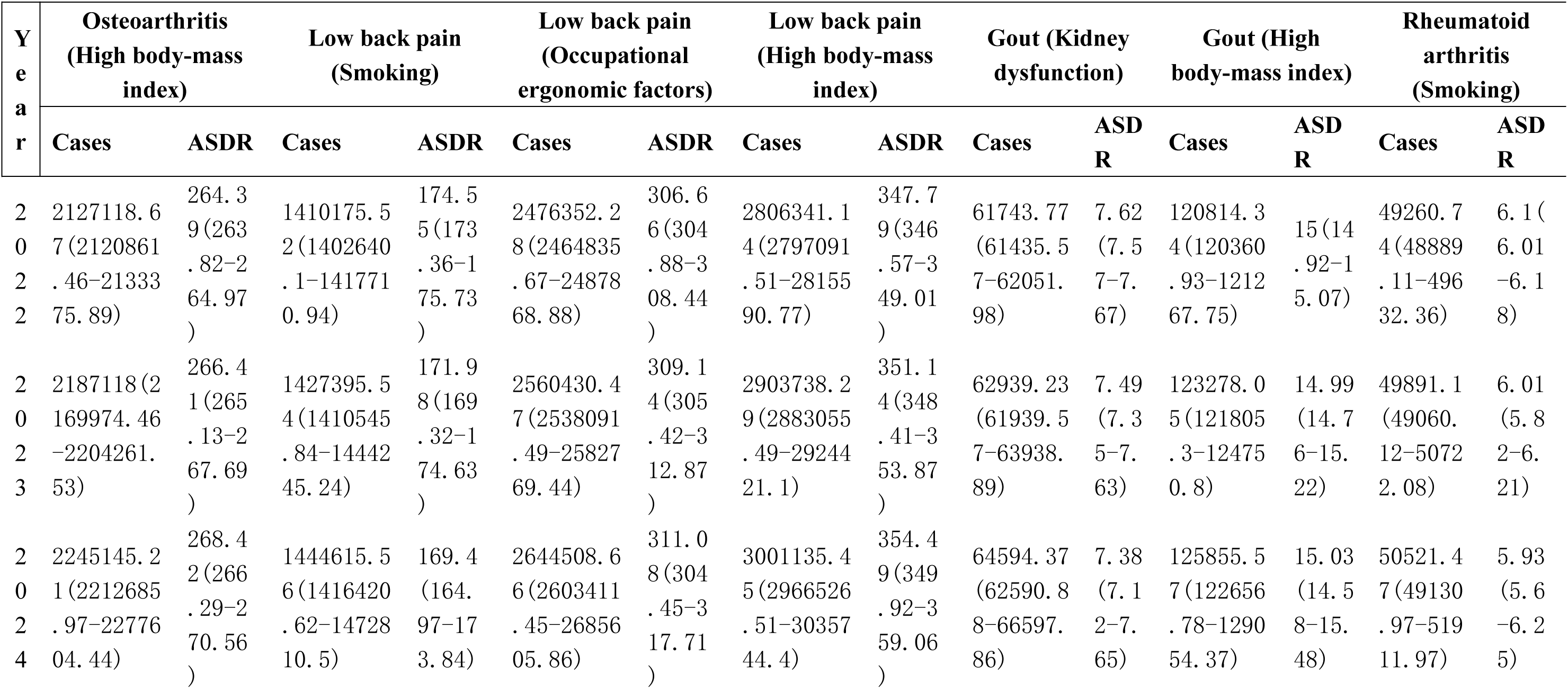
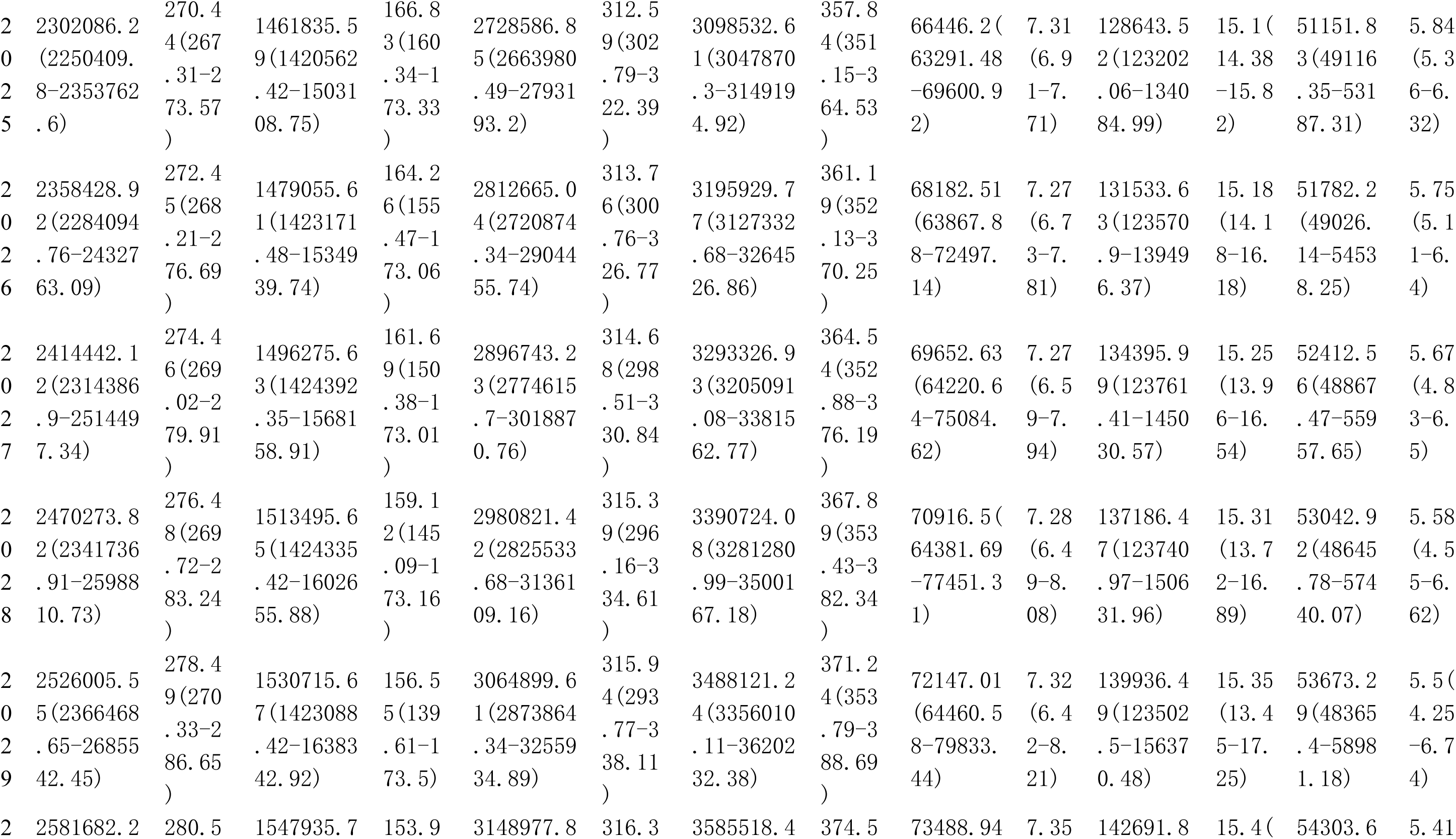
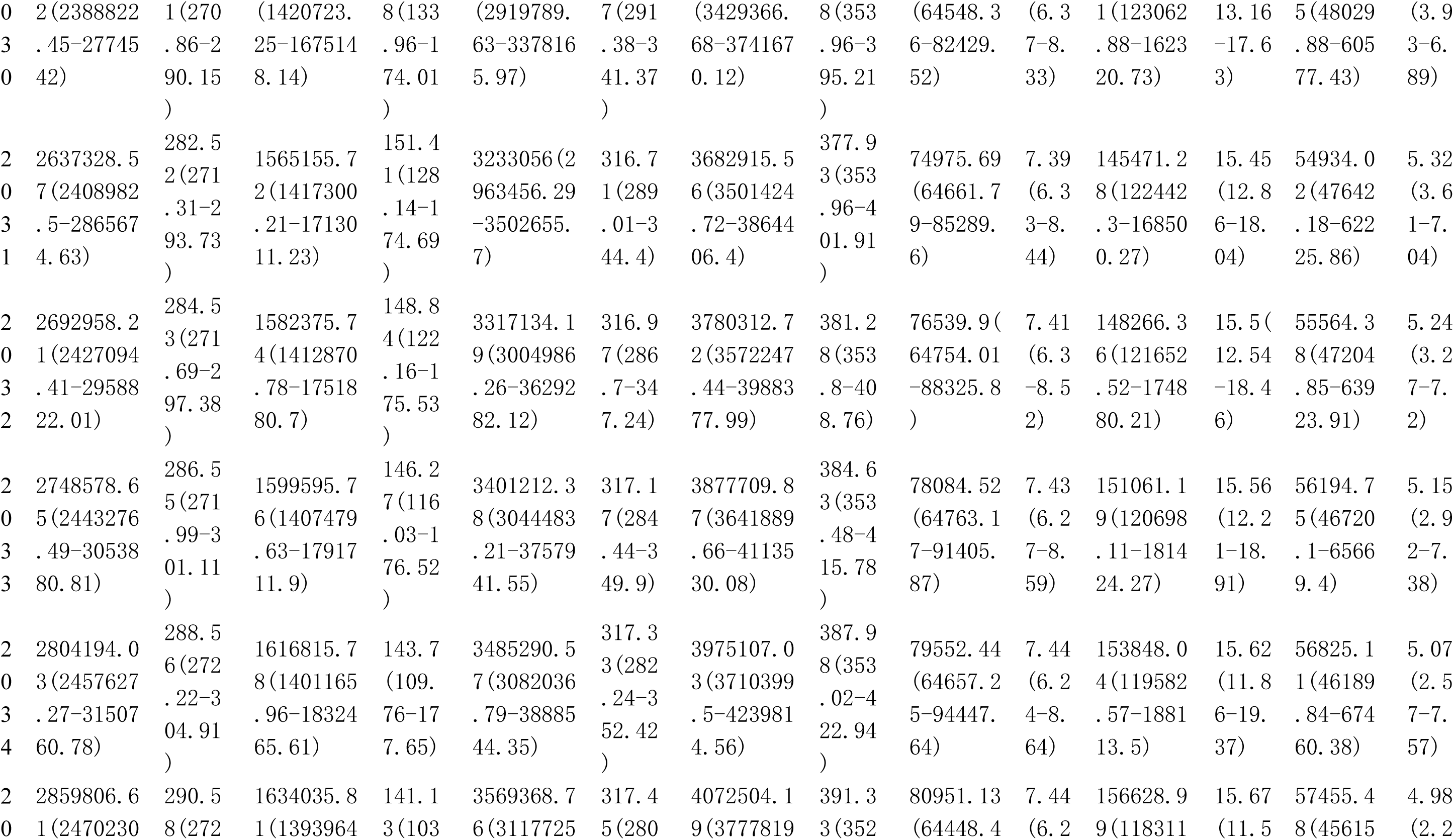
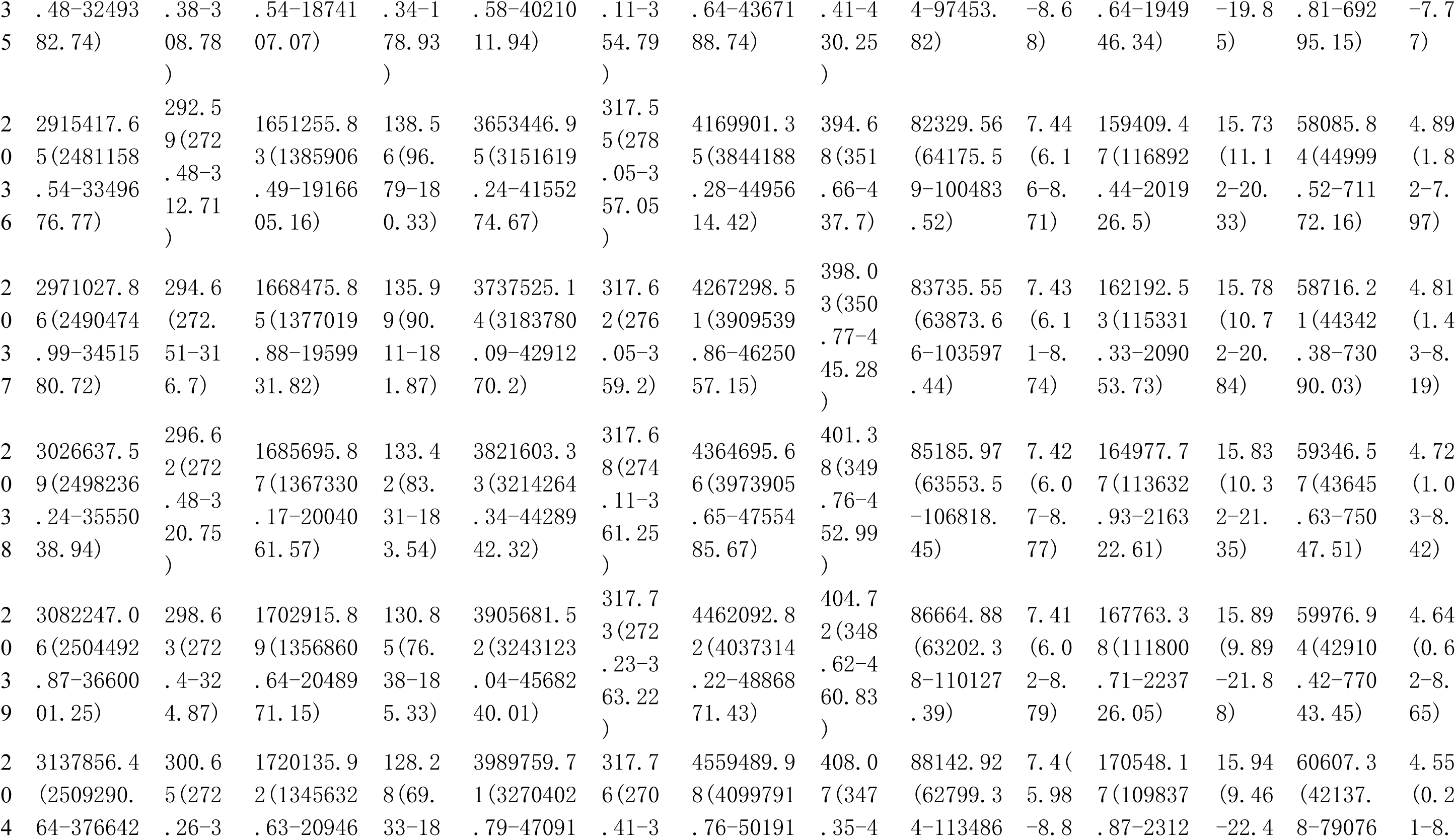
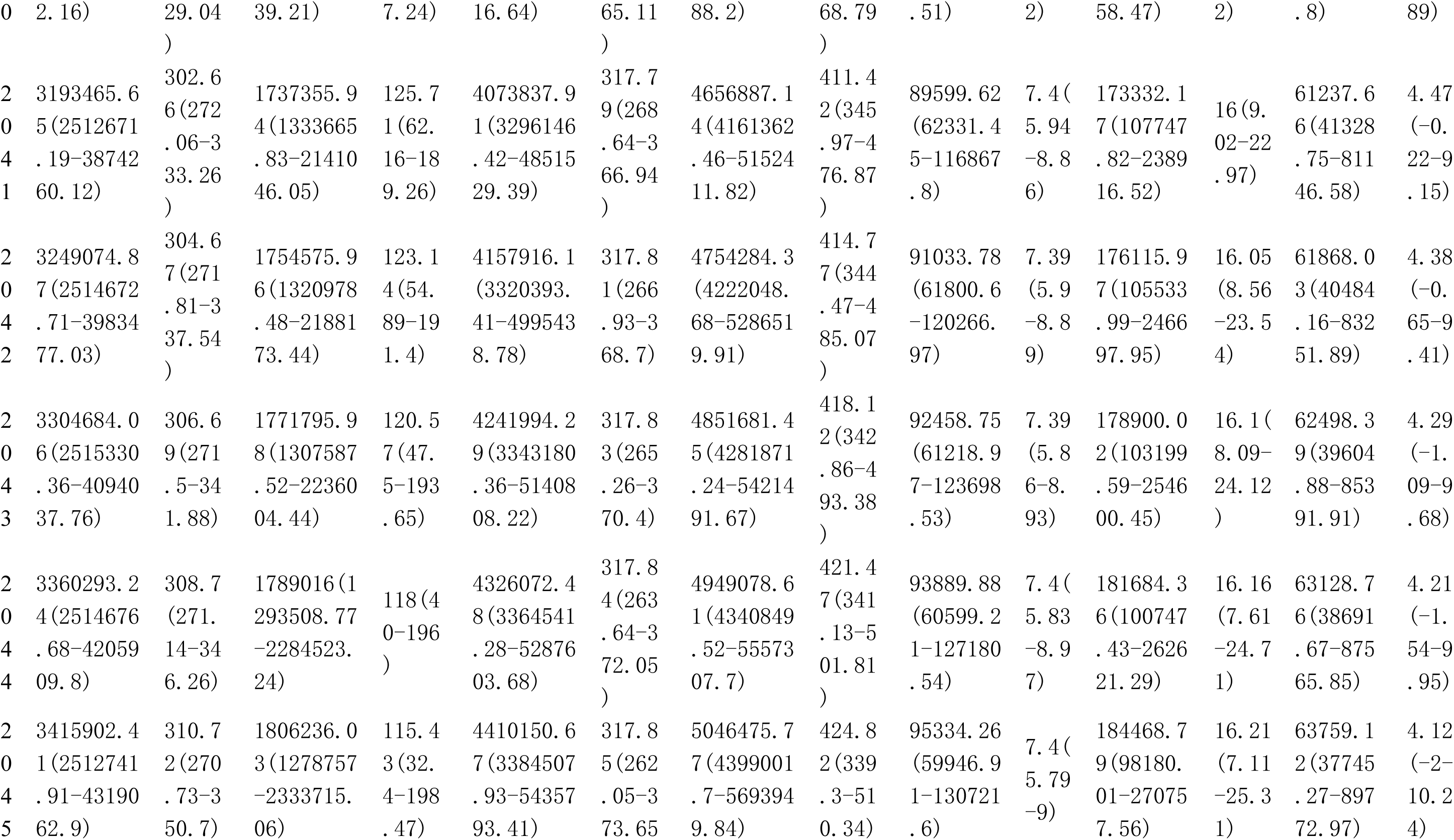
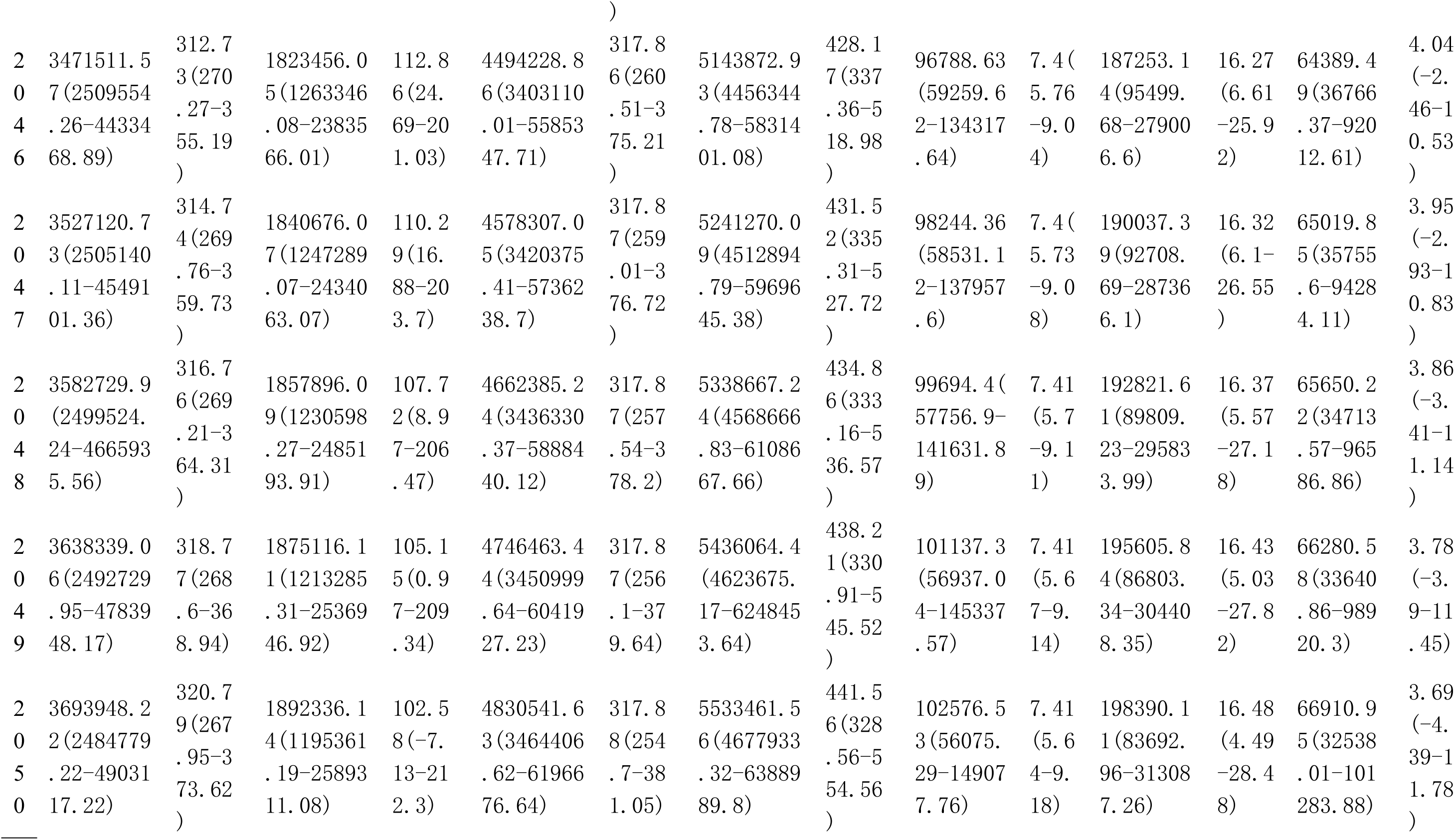
Predictive analysis of DALYs Attributable to Associated Risk Factors among postmenopausal women from 2022 to 2050.

**Table S6.**
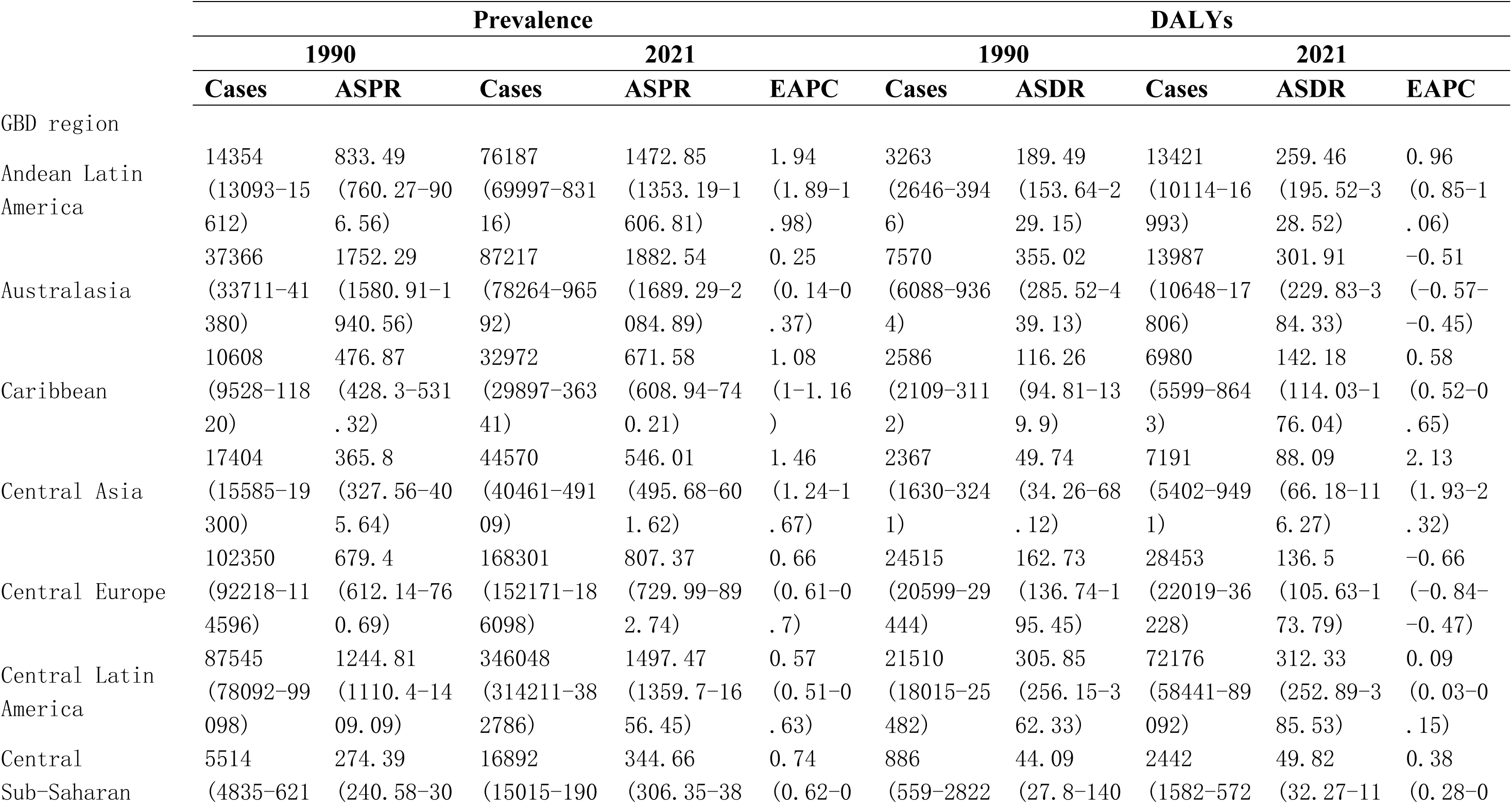
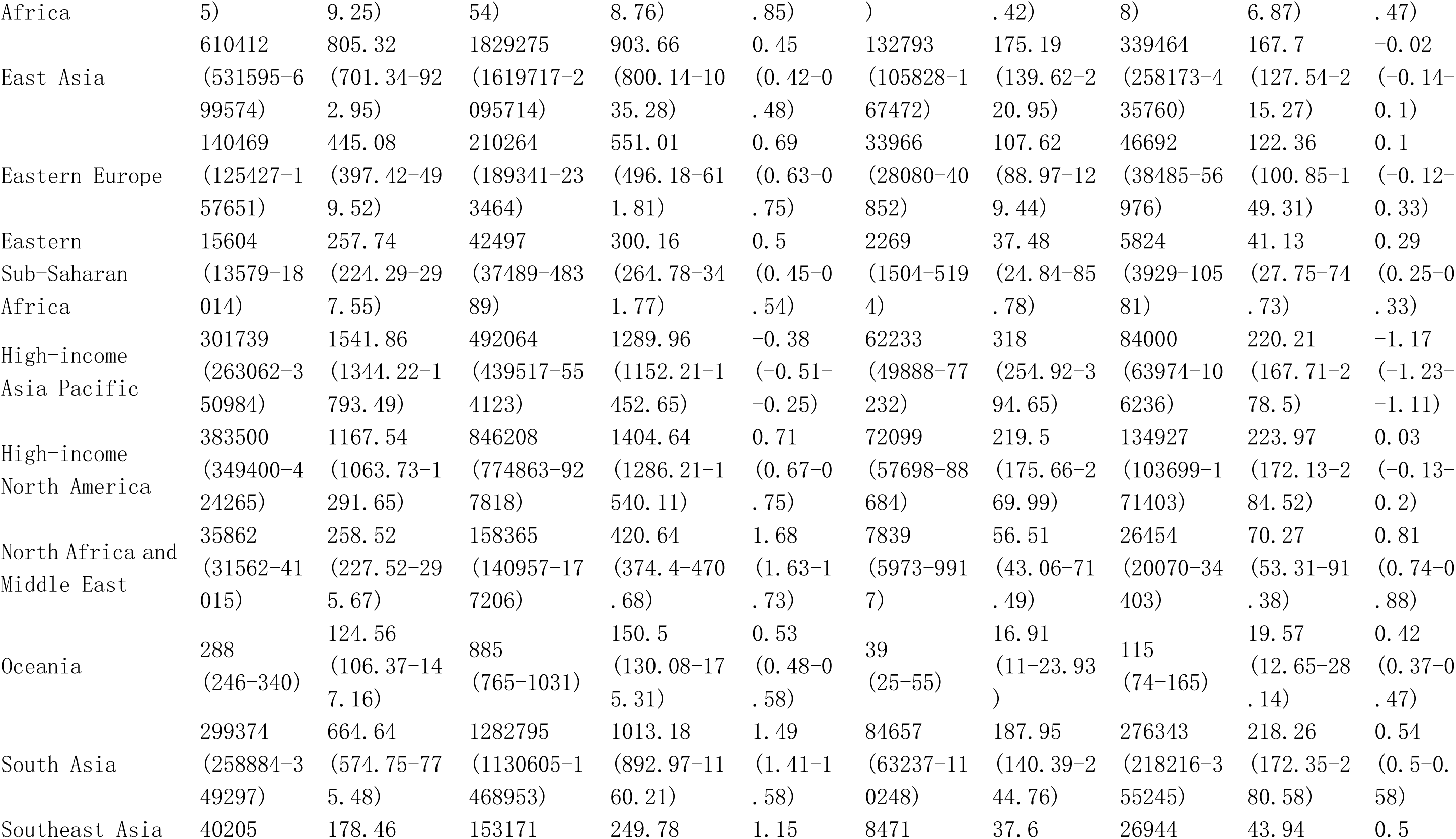
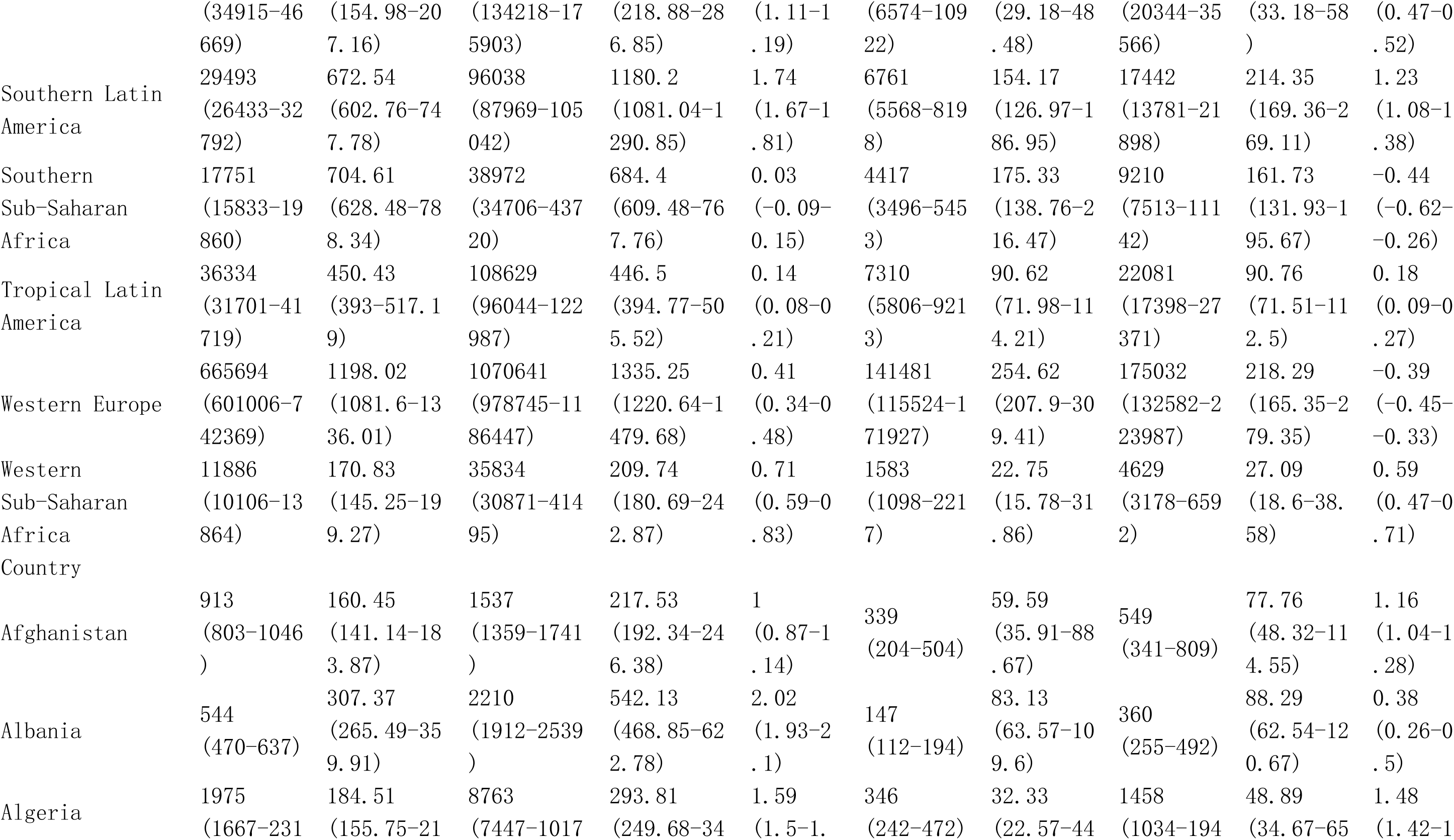
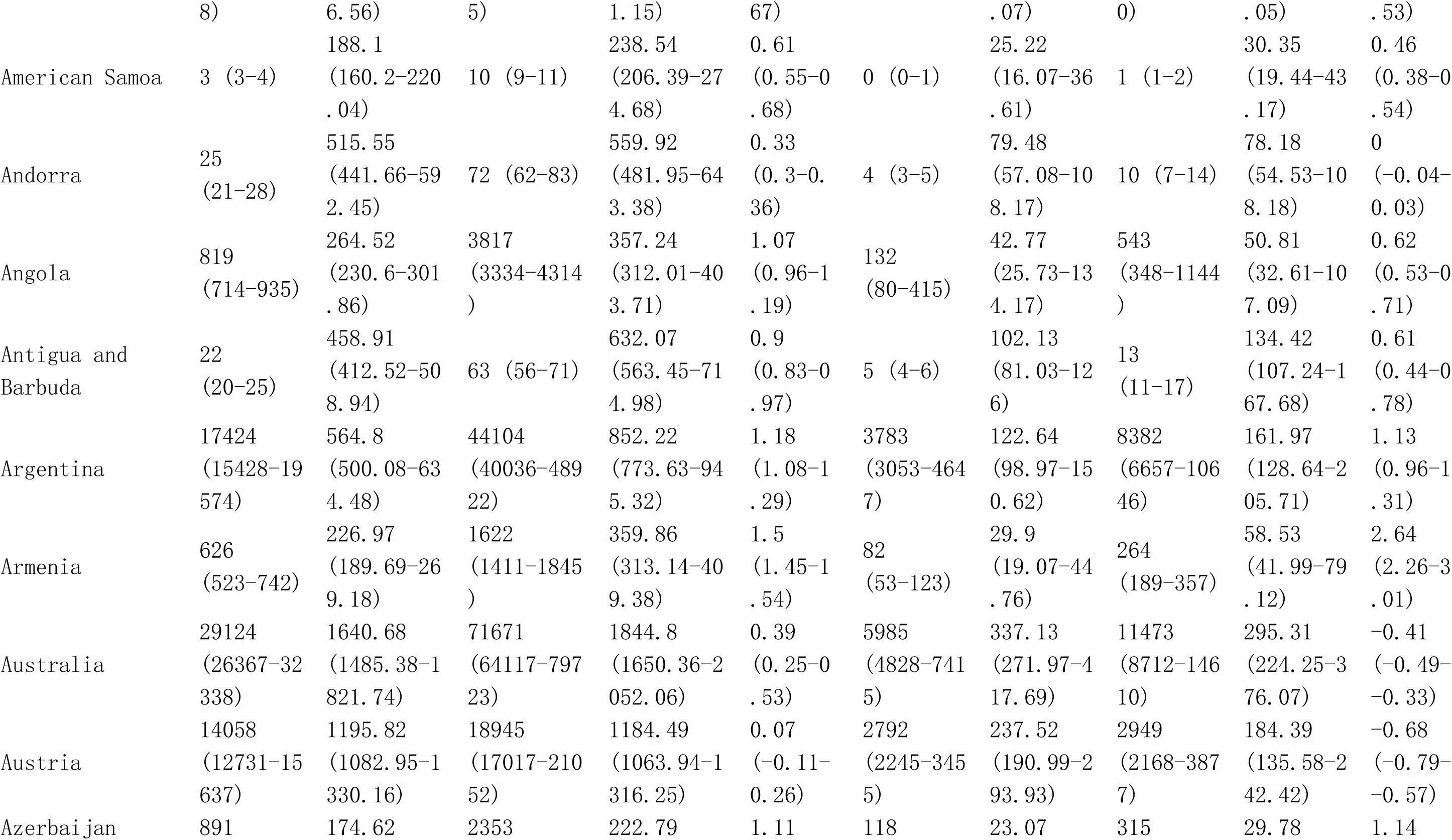
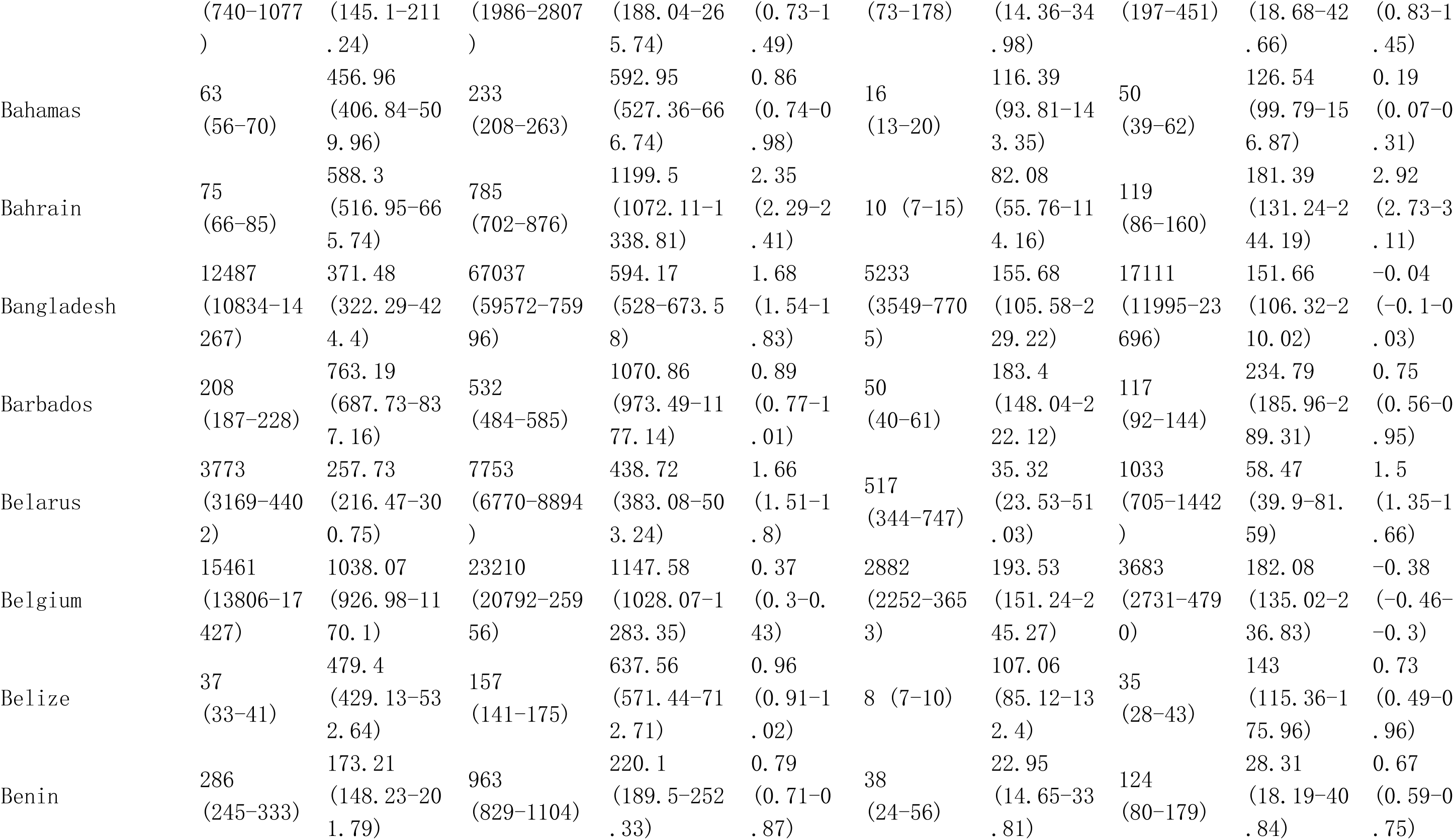
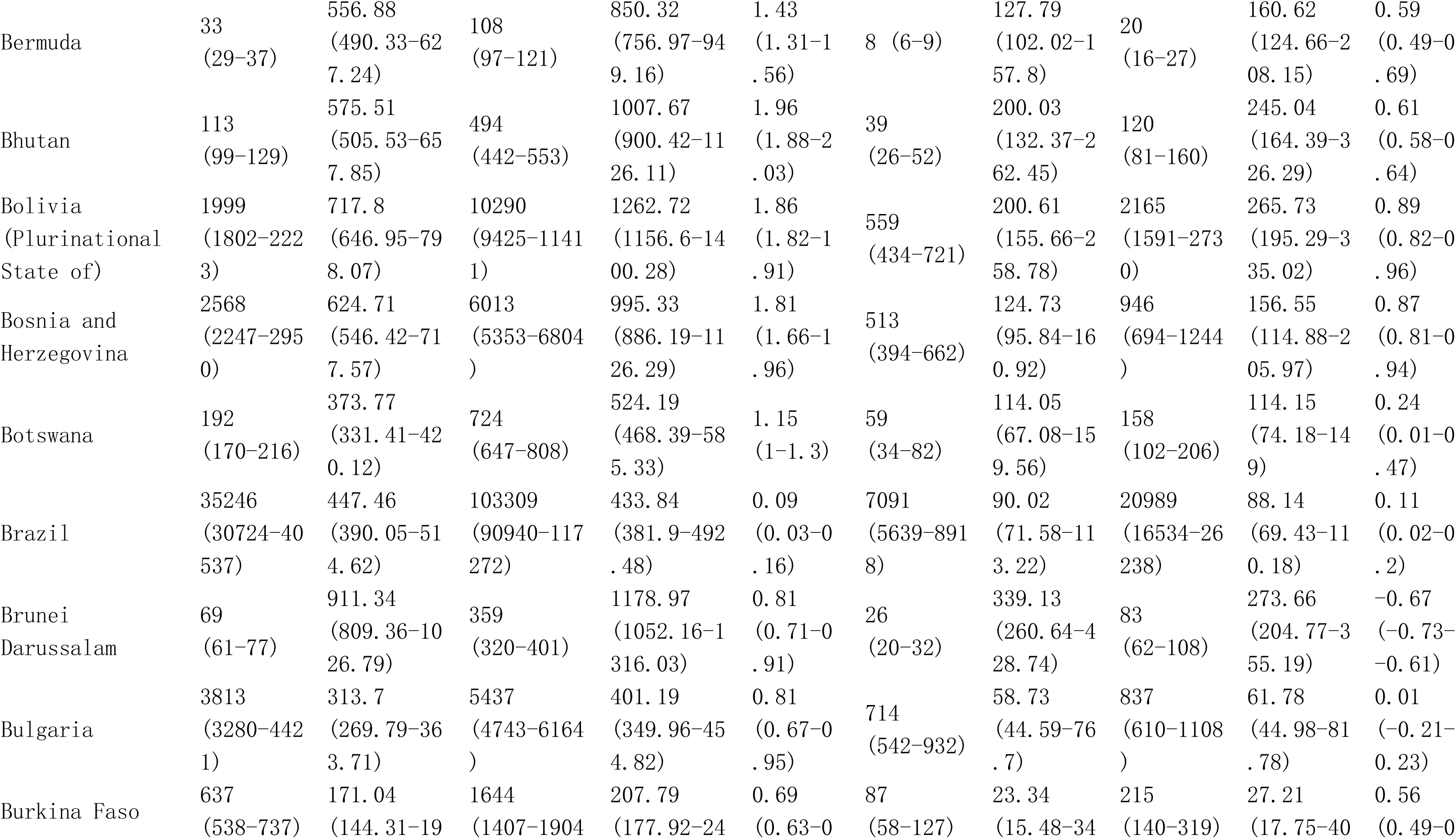
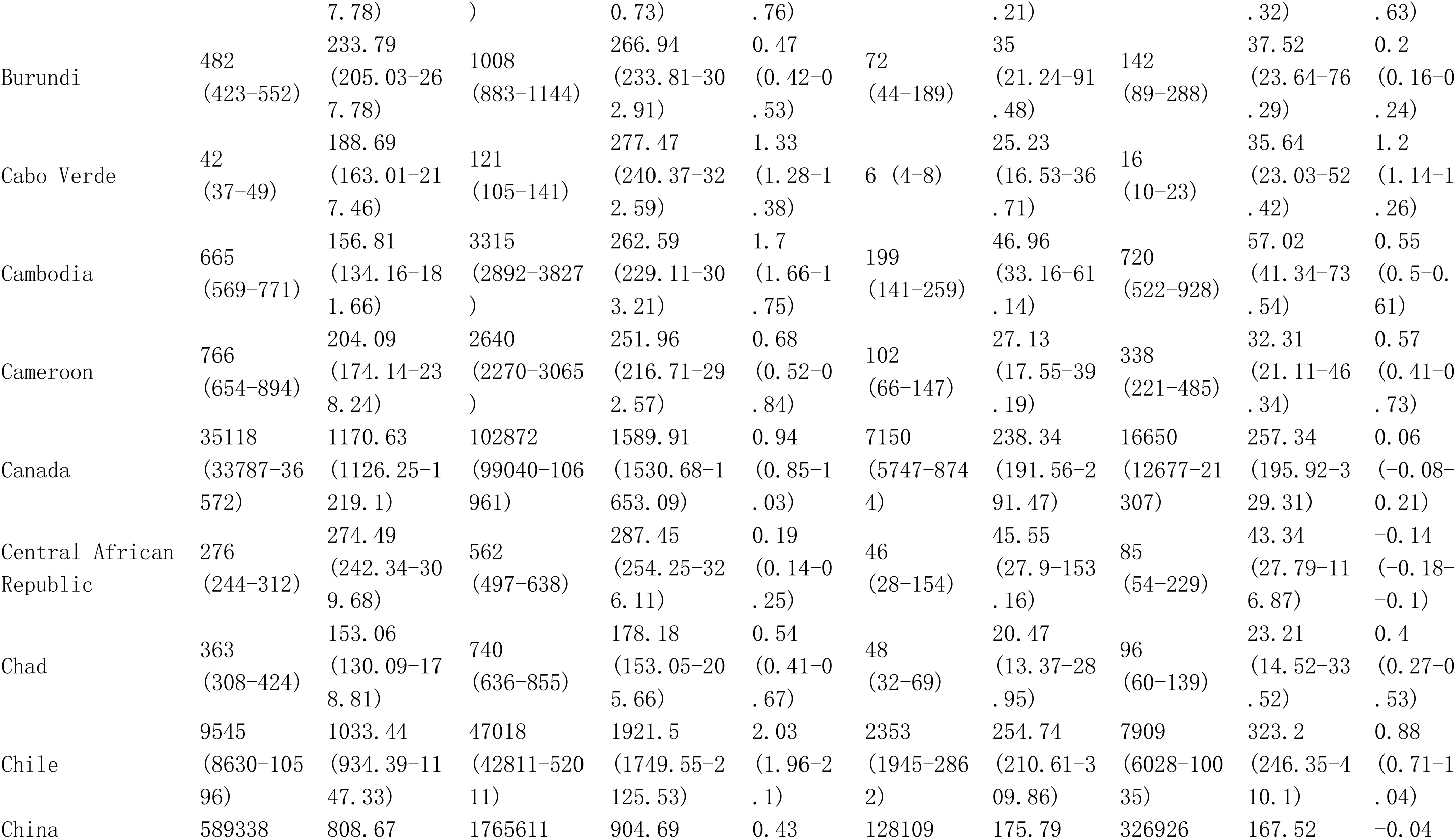
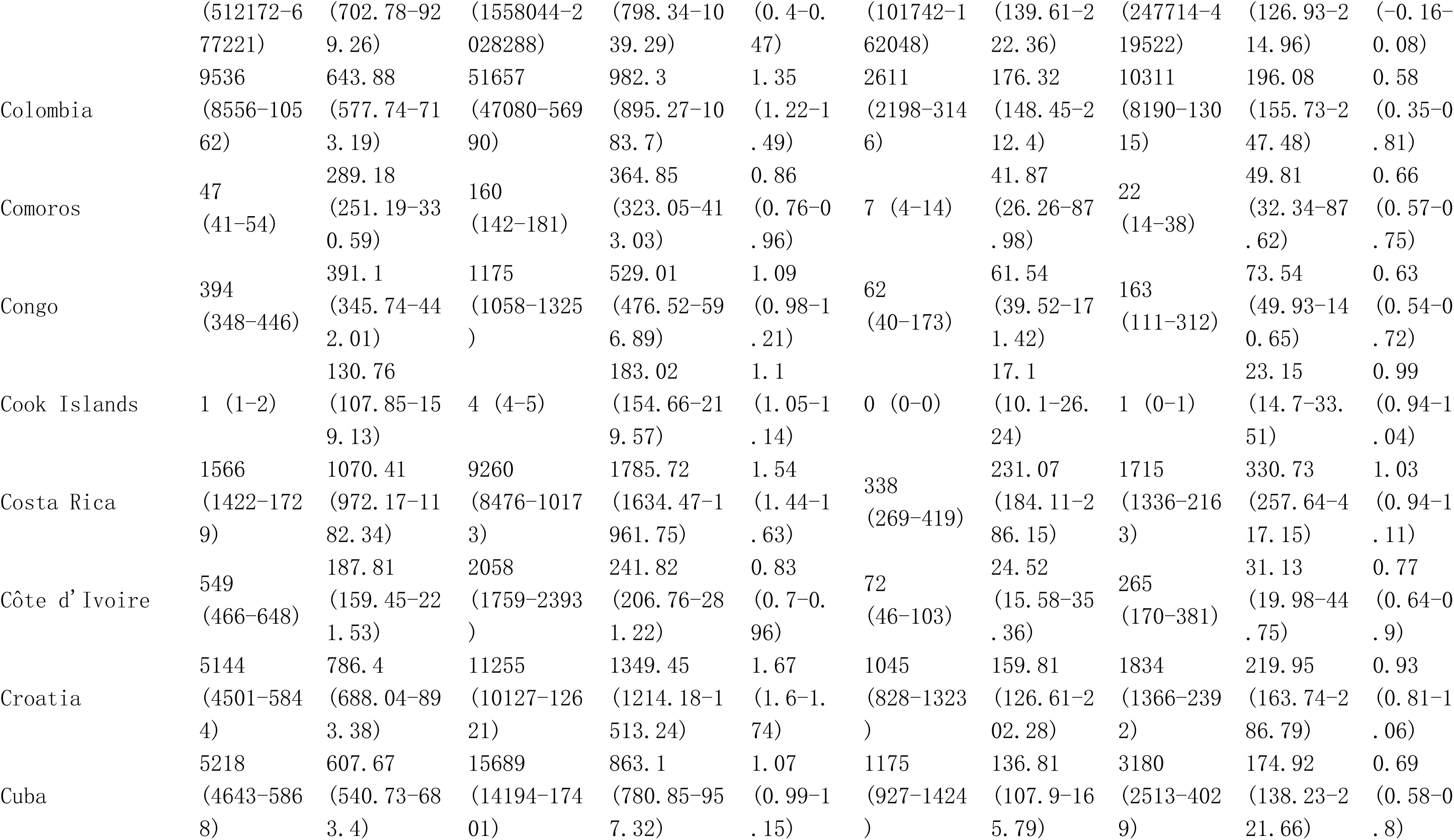
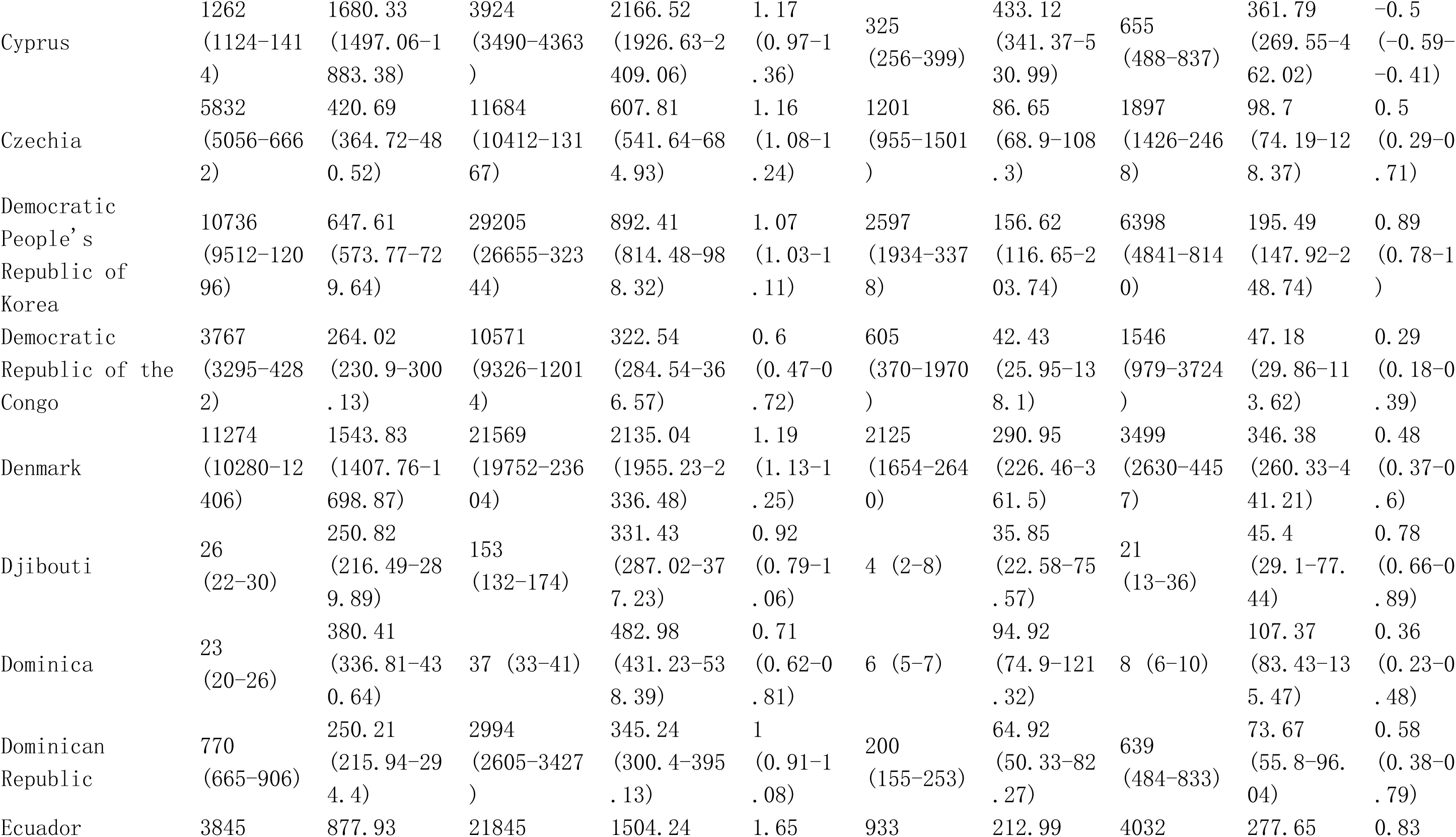
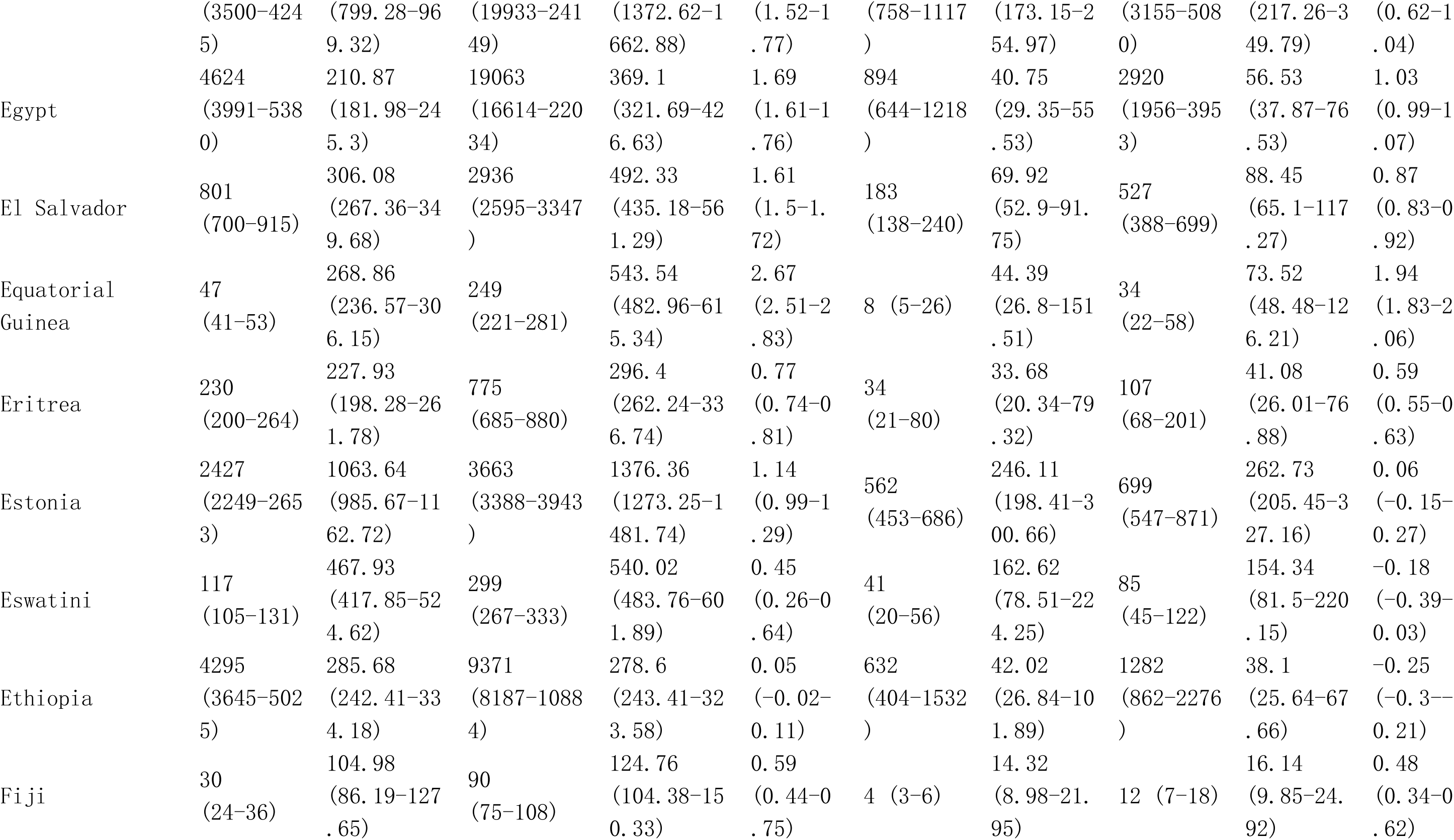
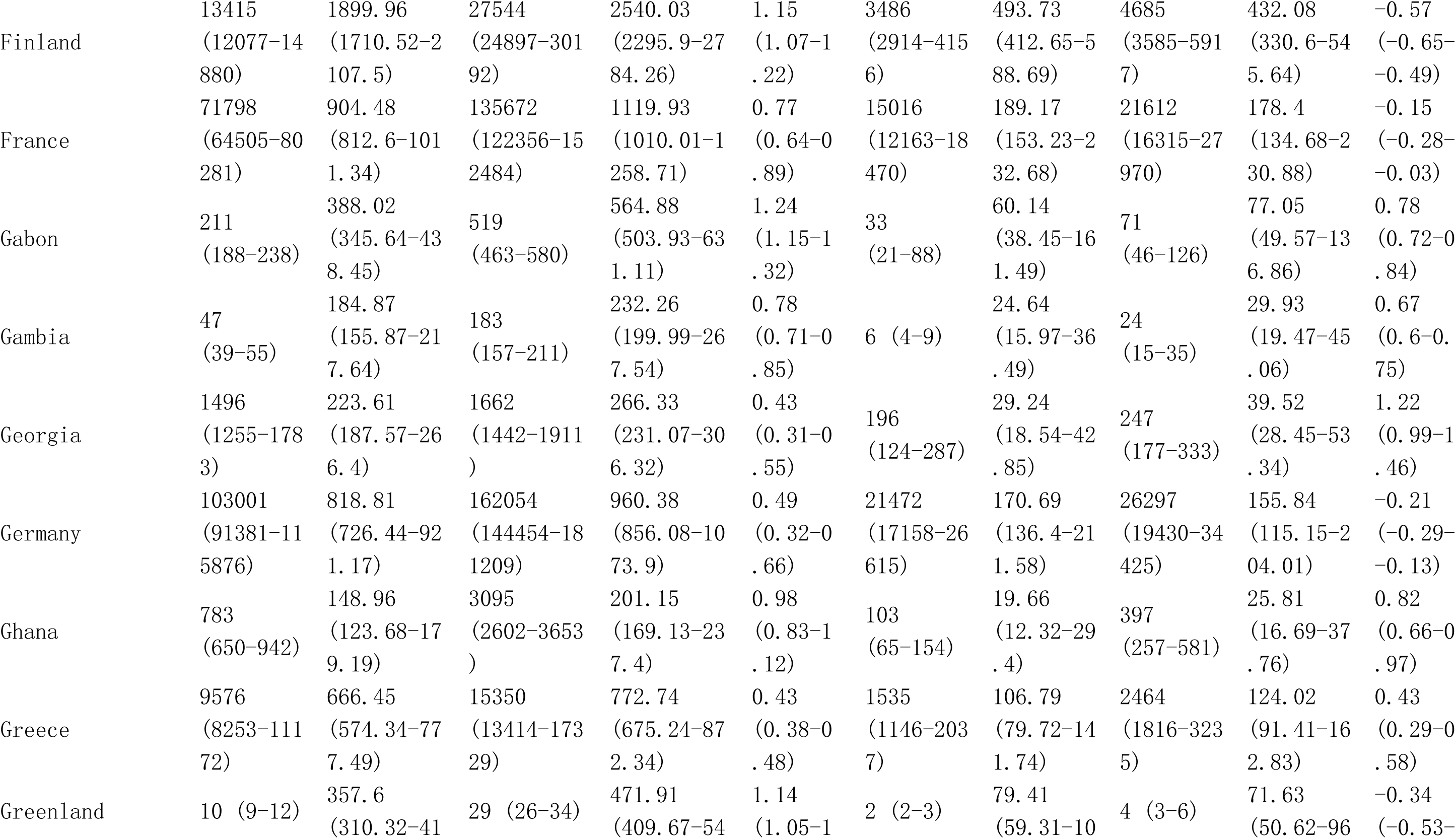
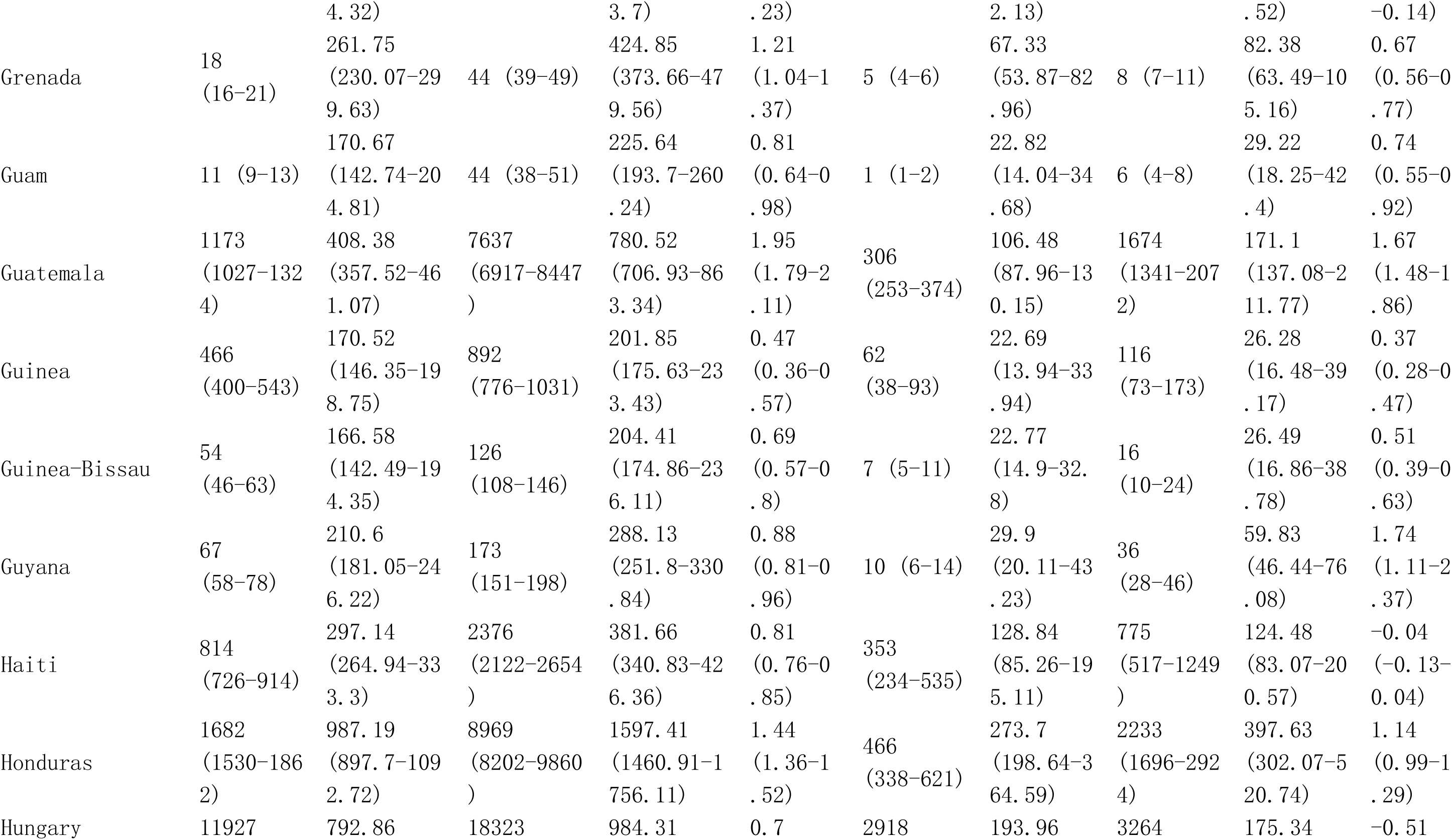
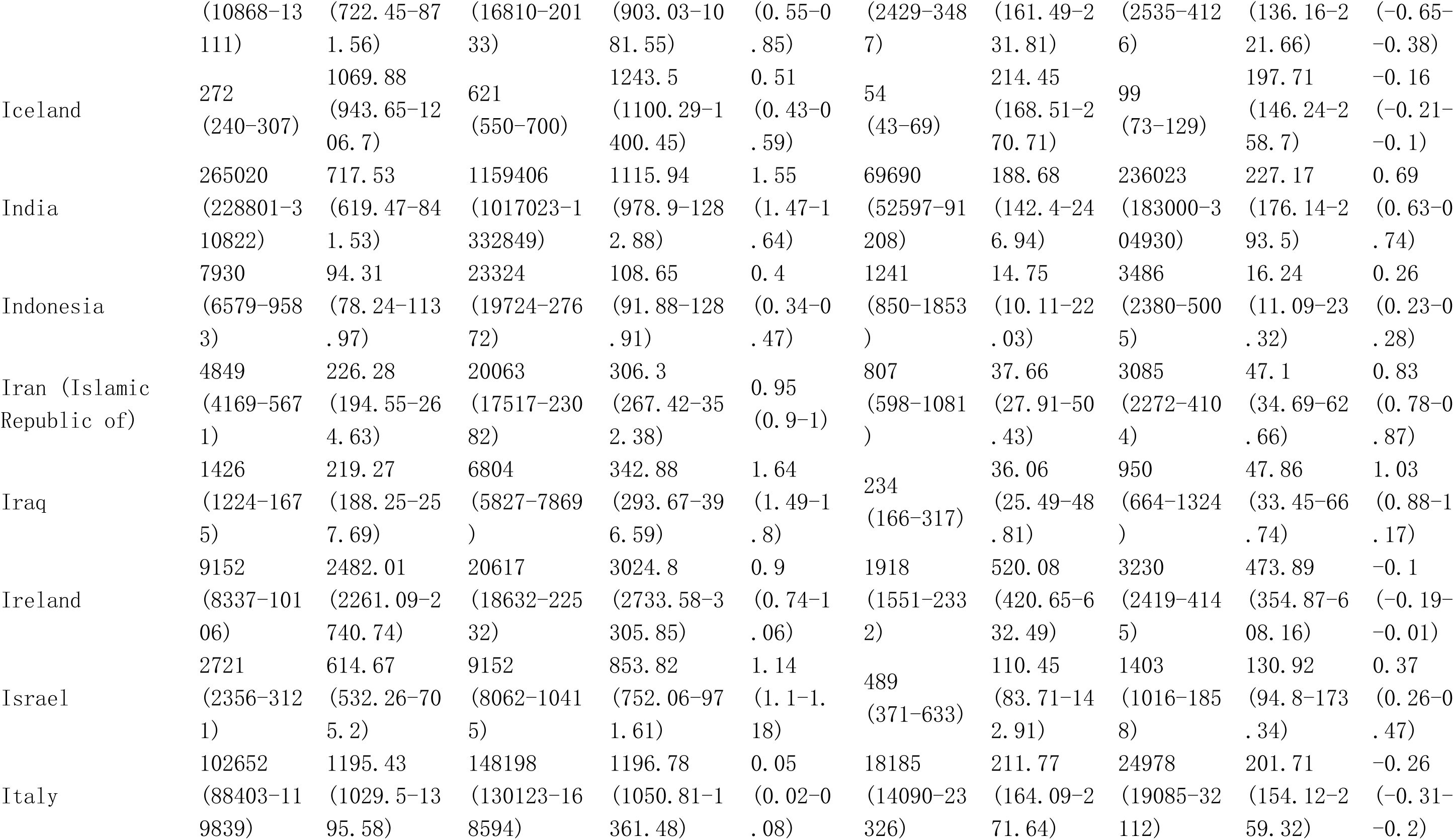
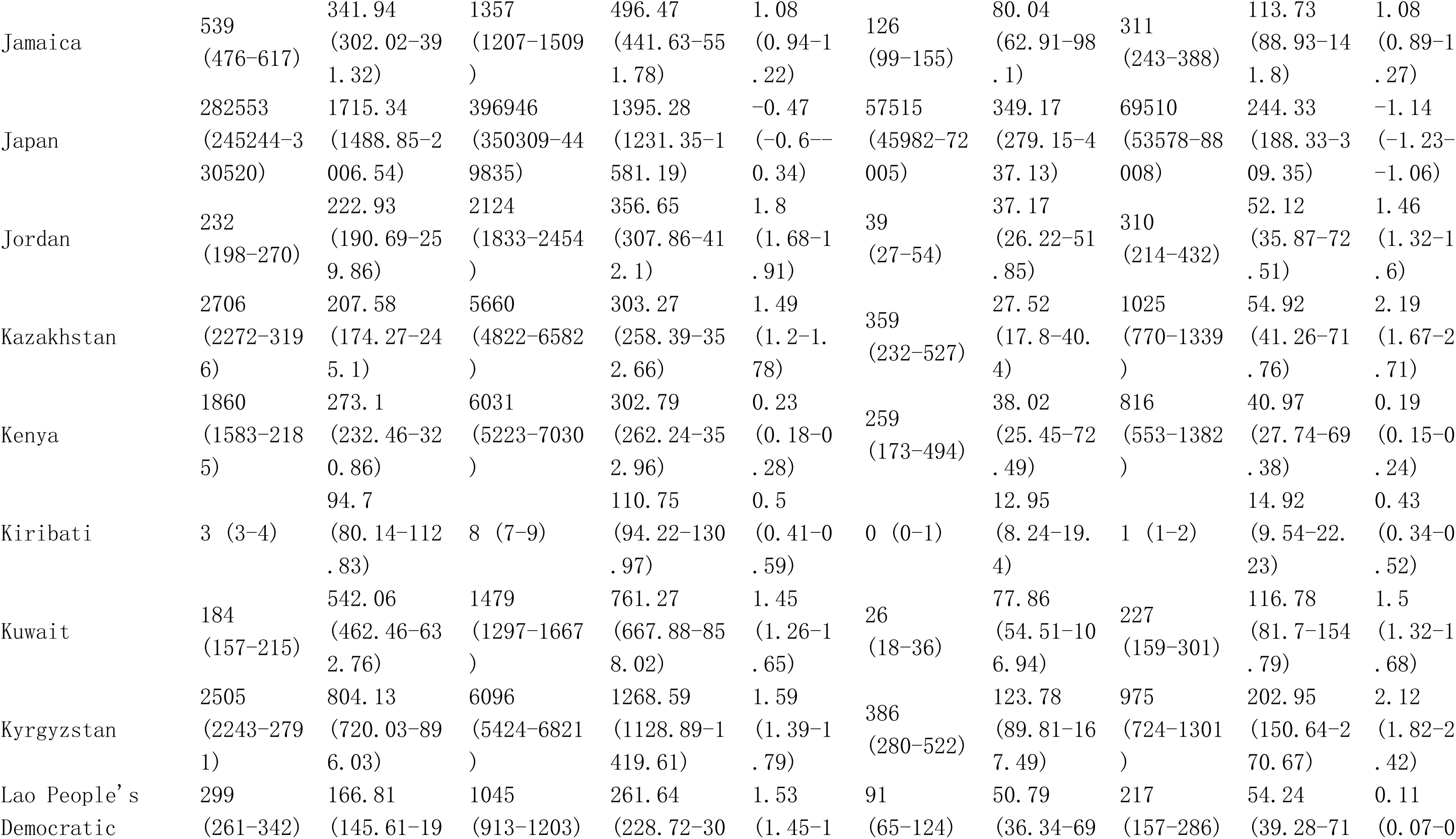
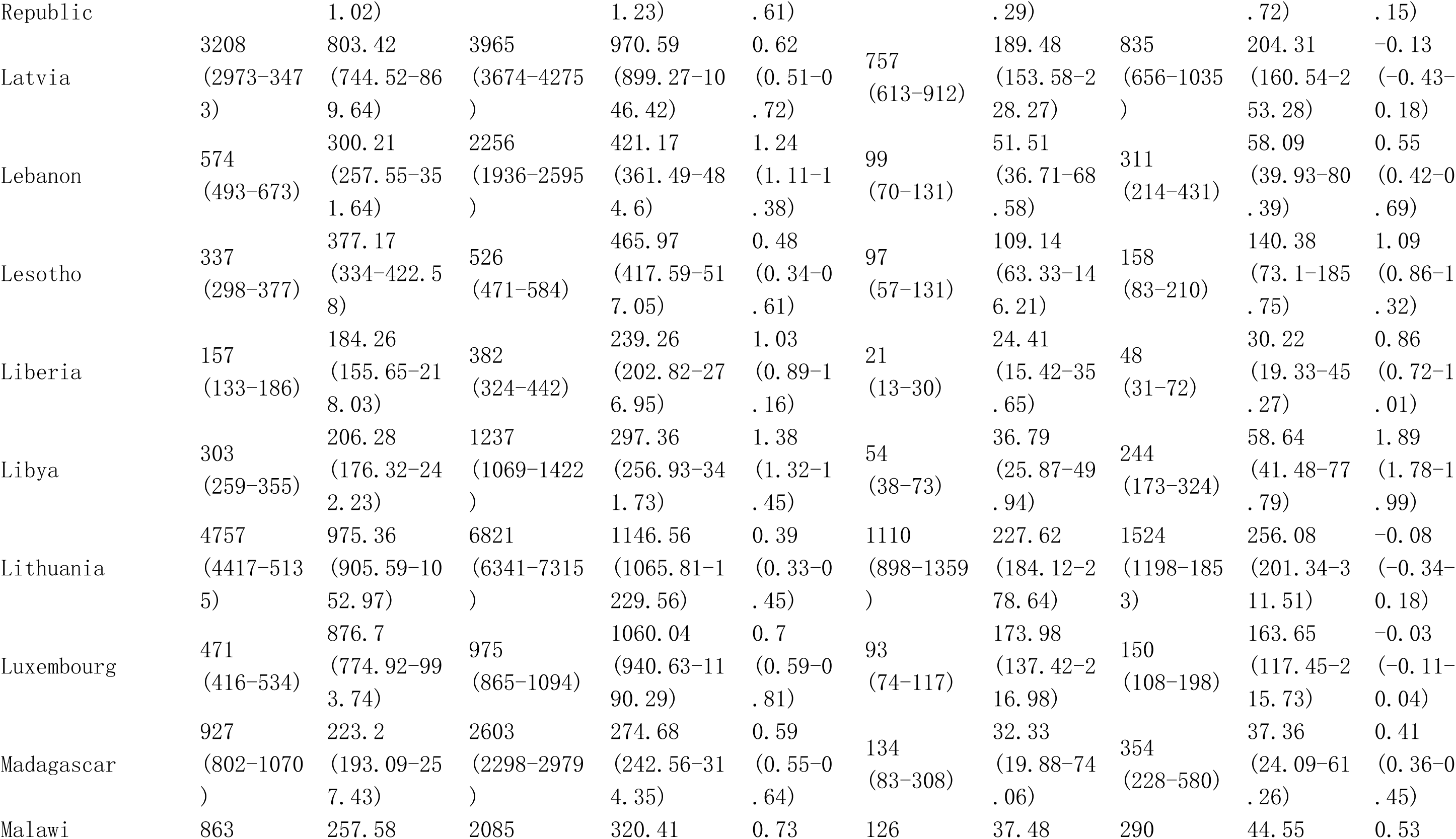
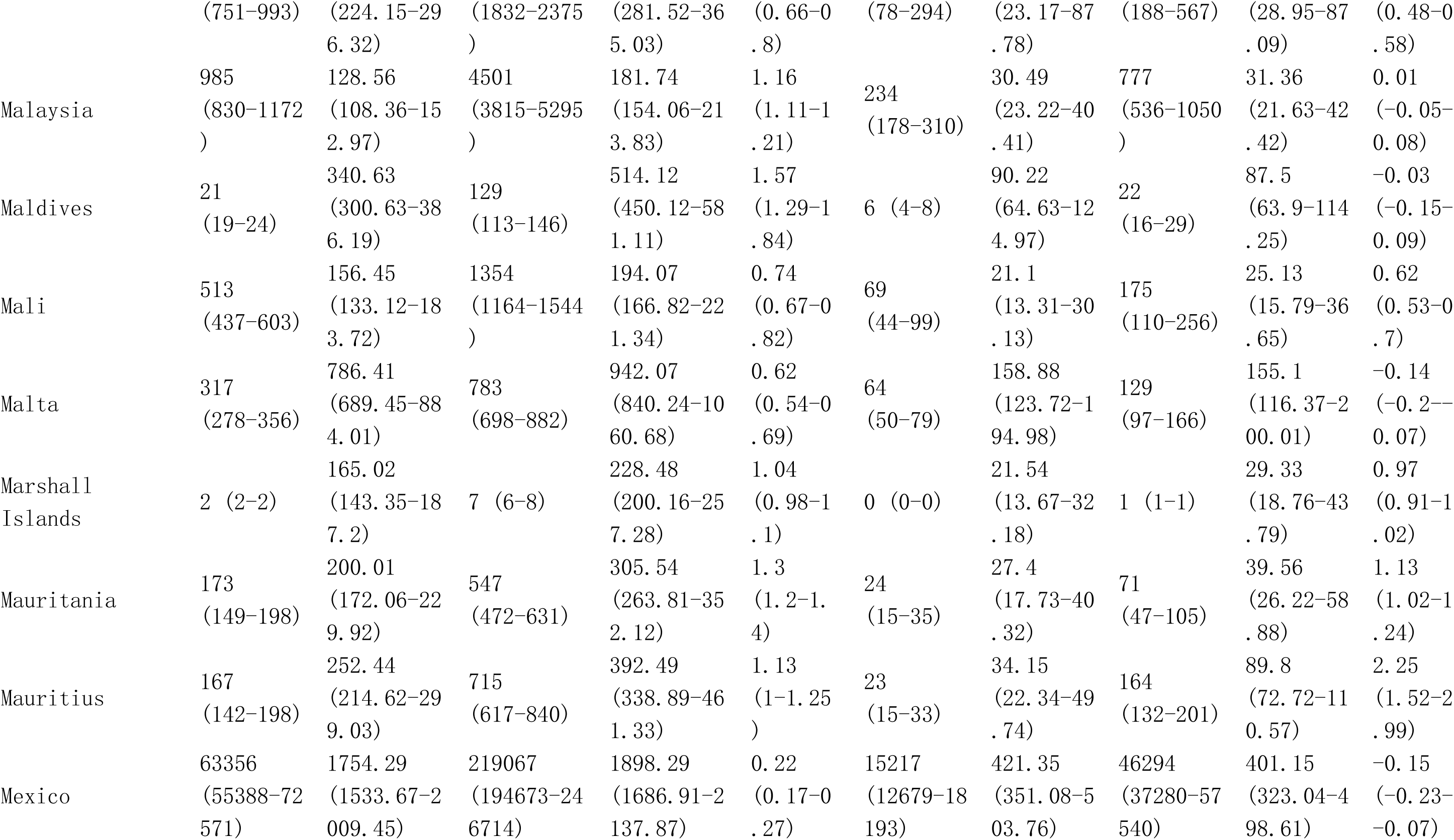
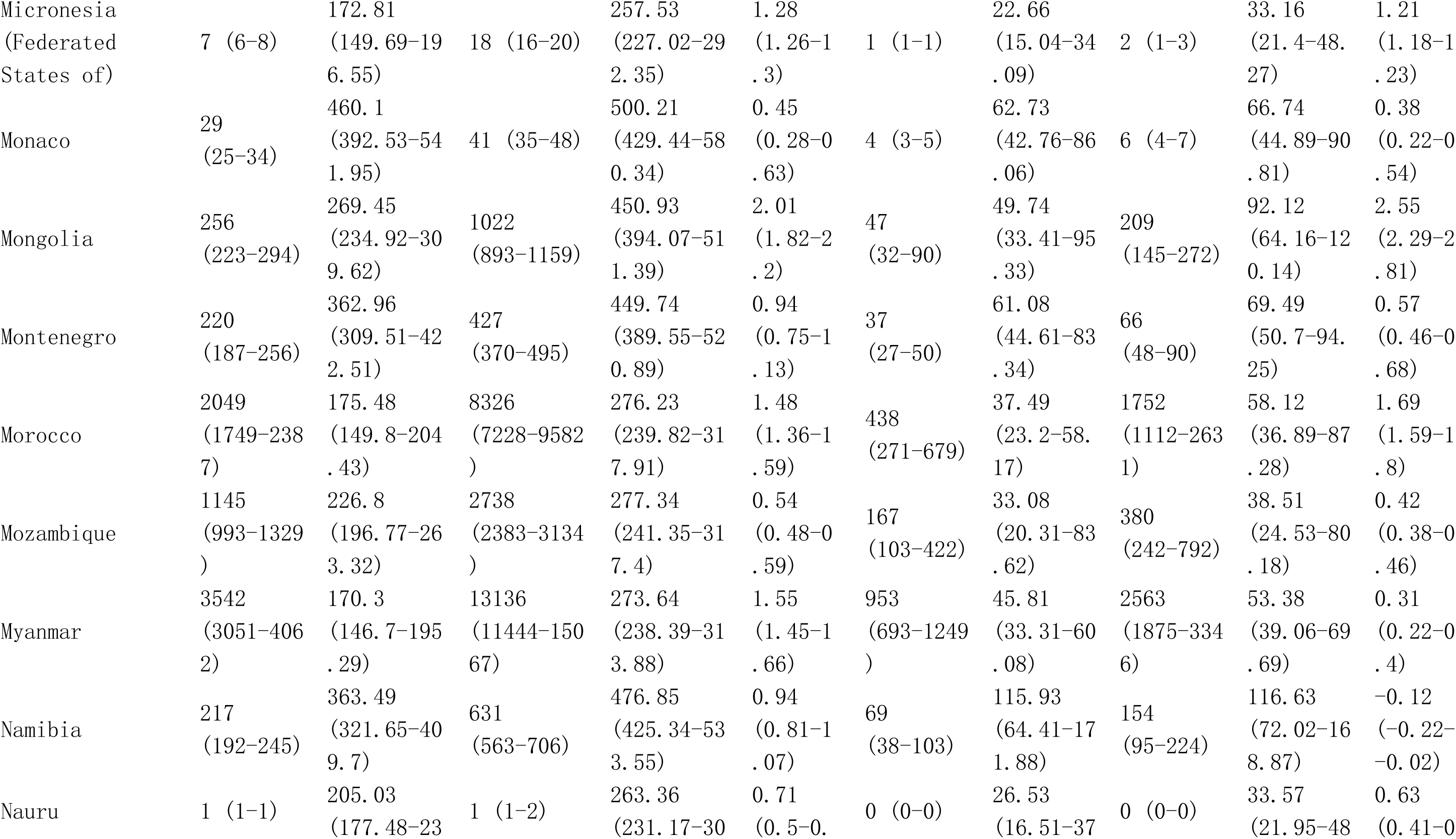
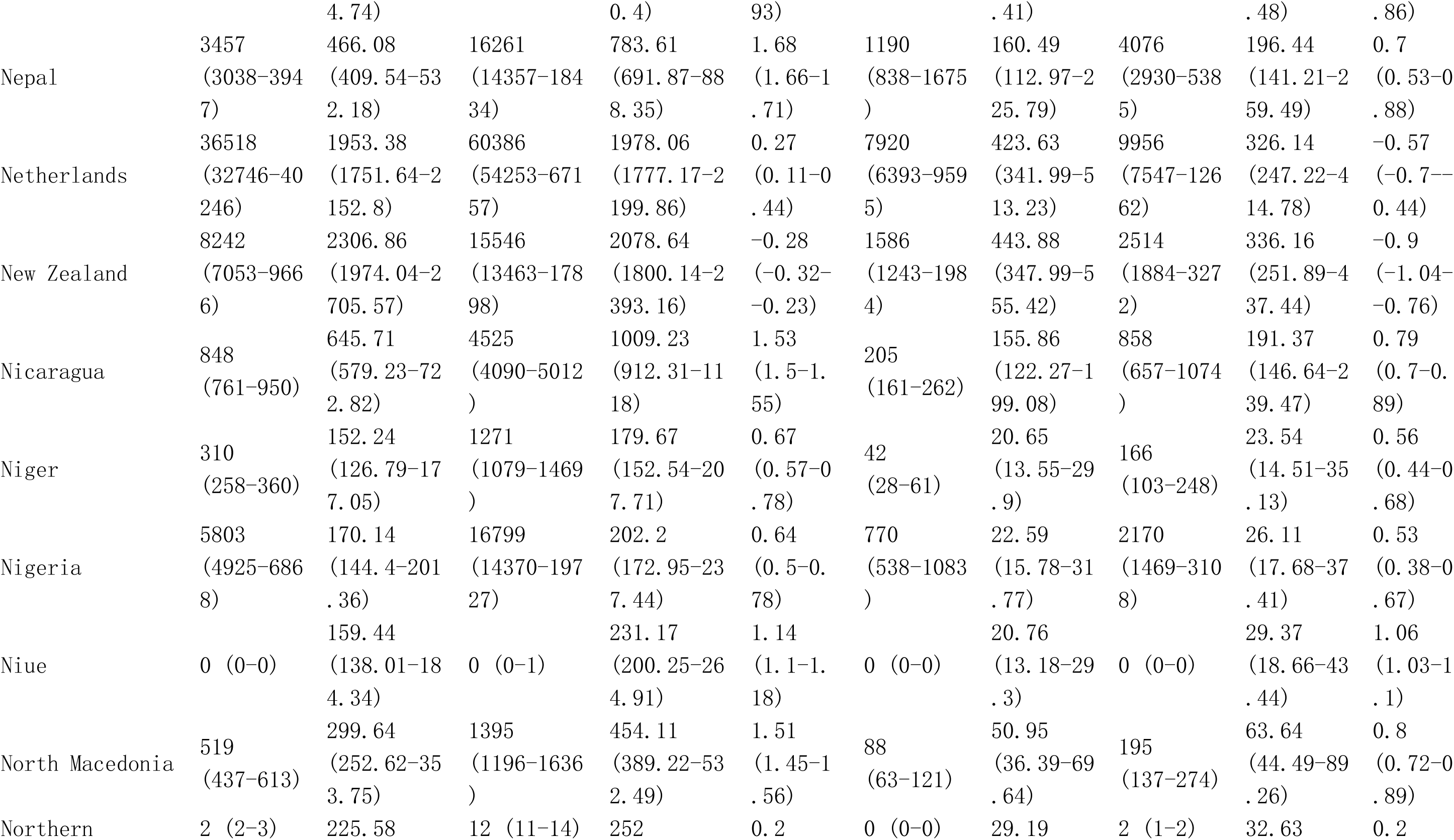
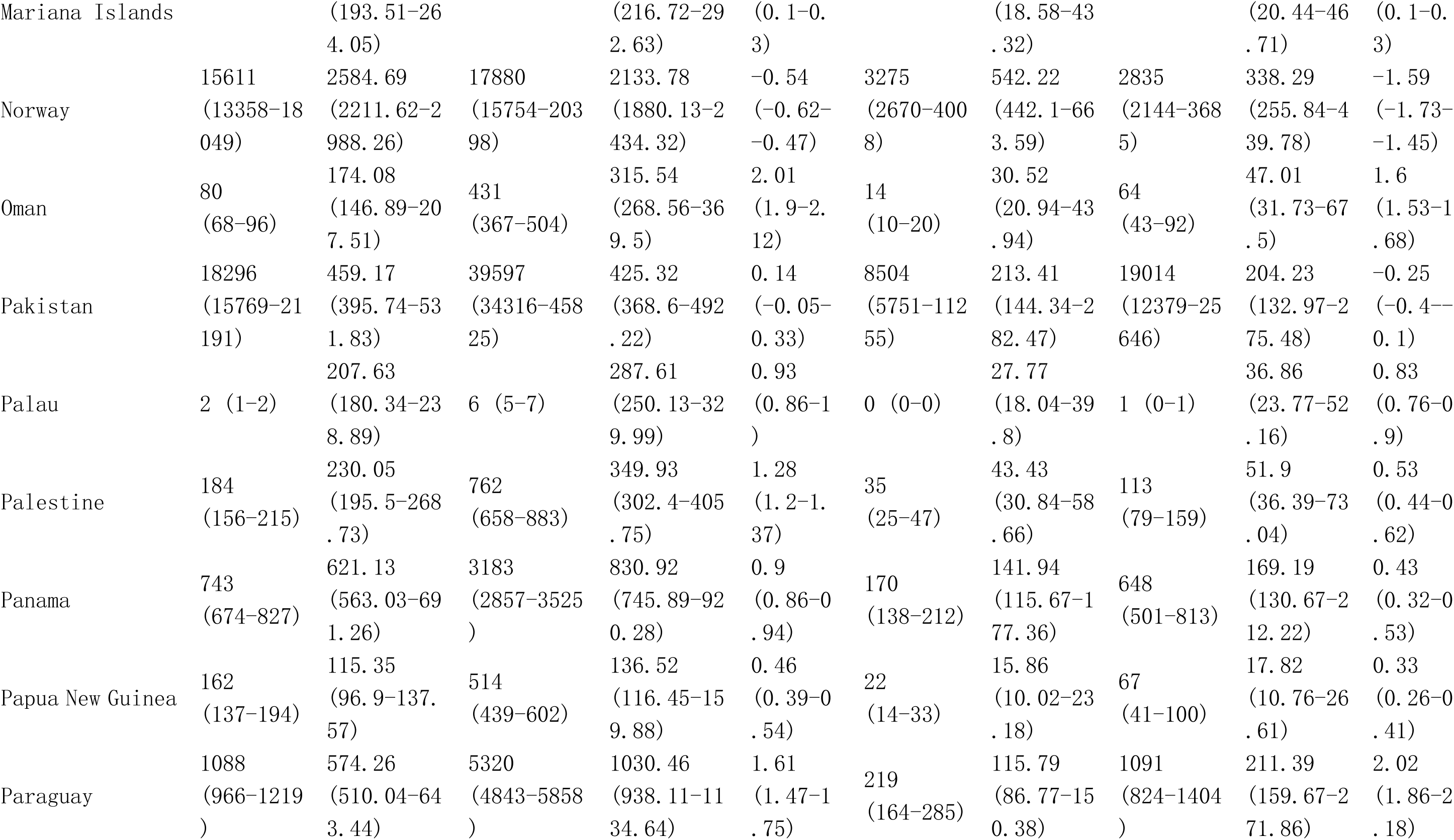
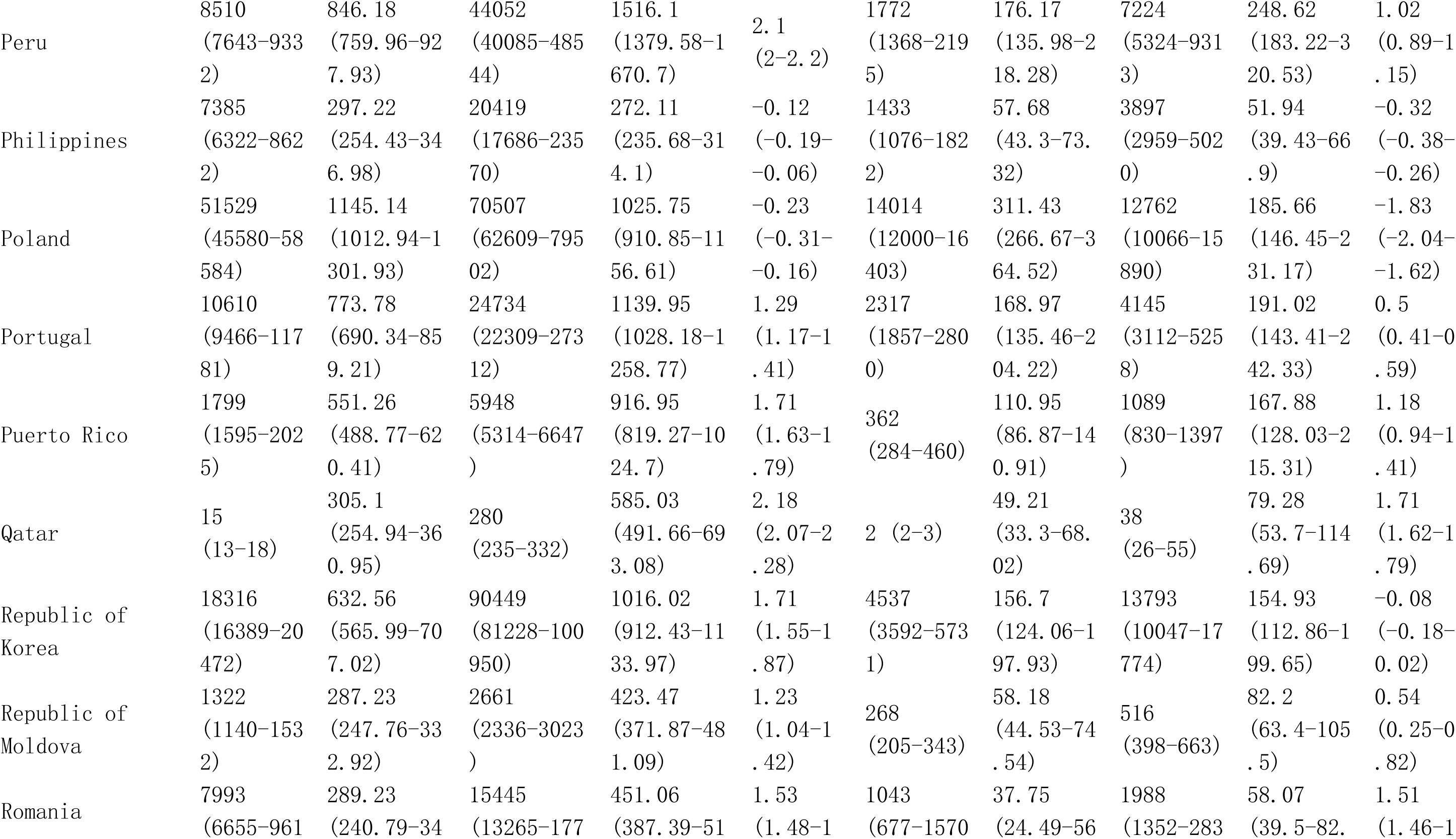
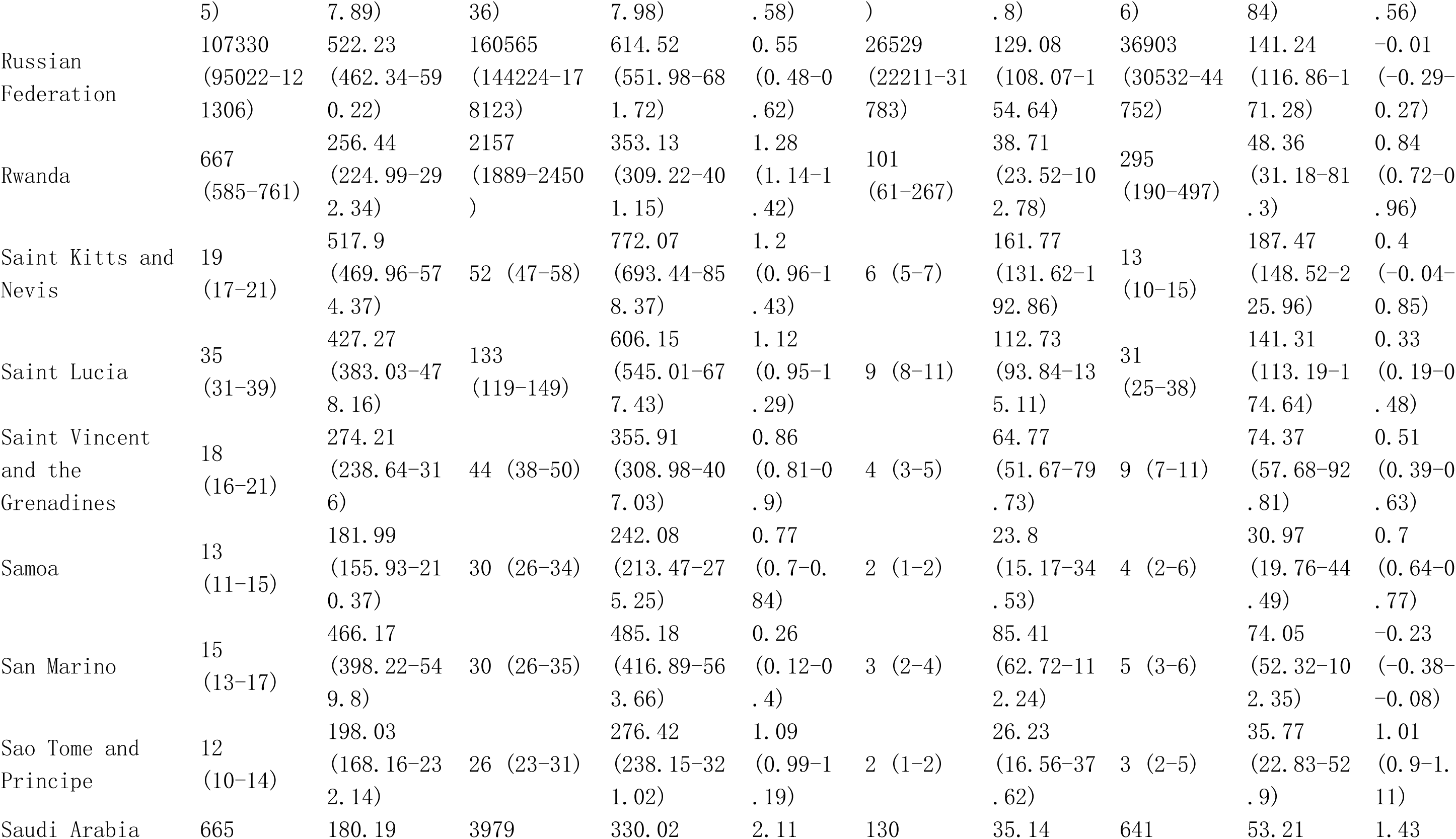
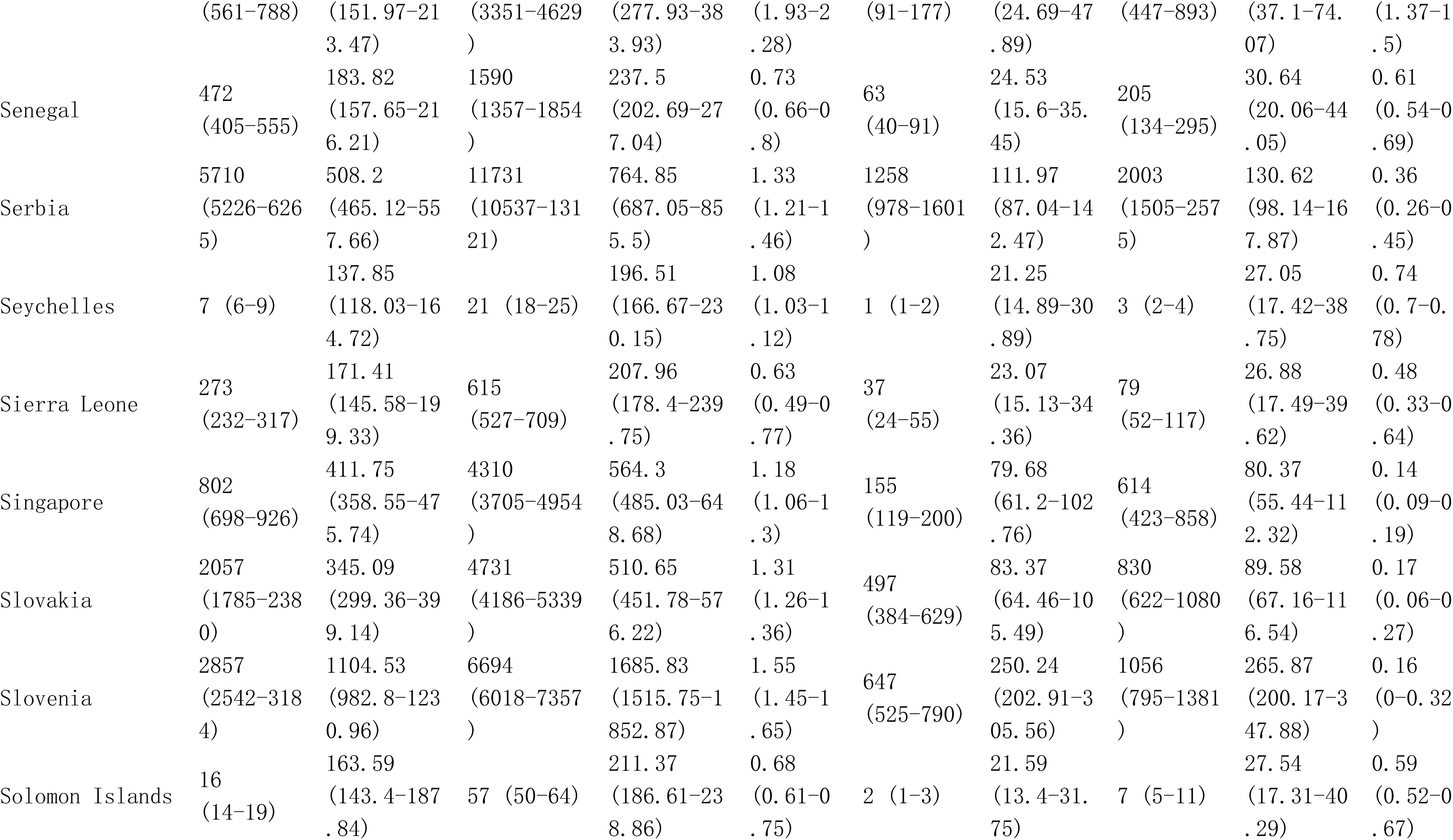
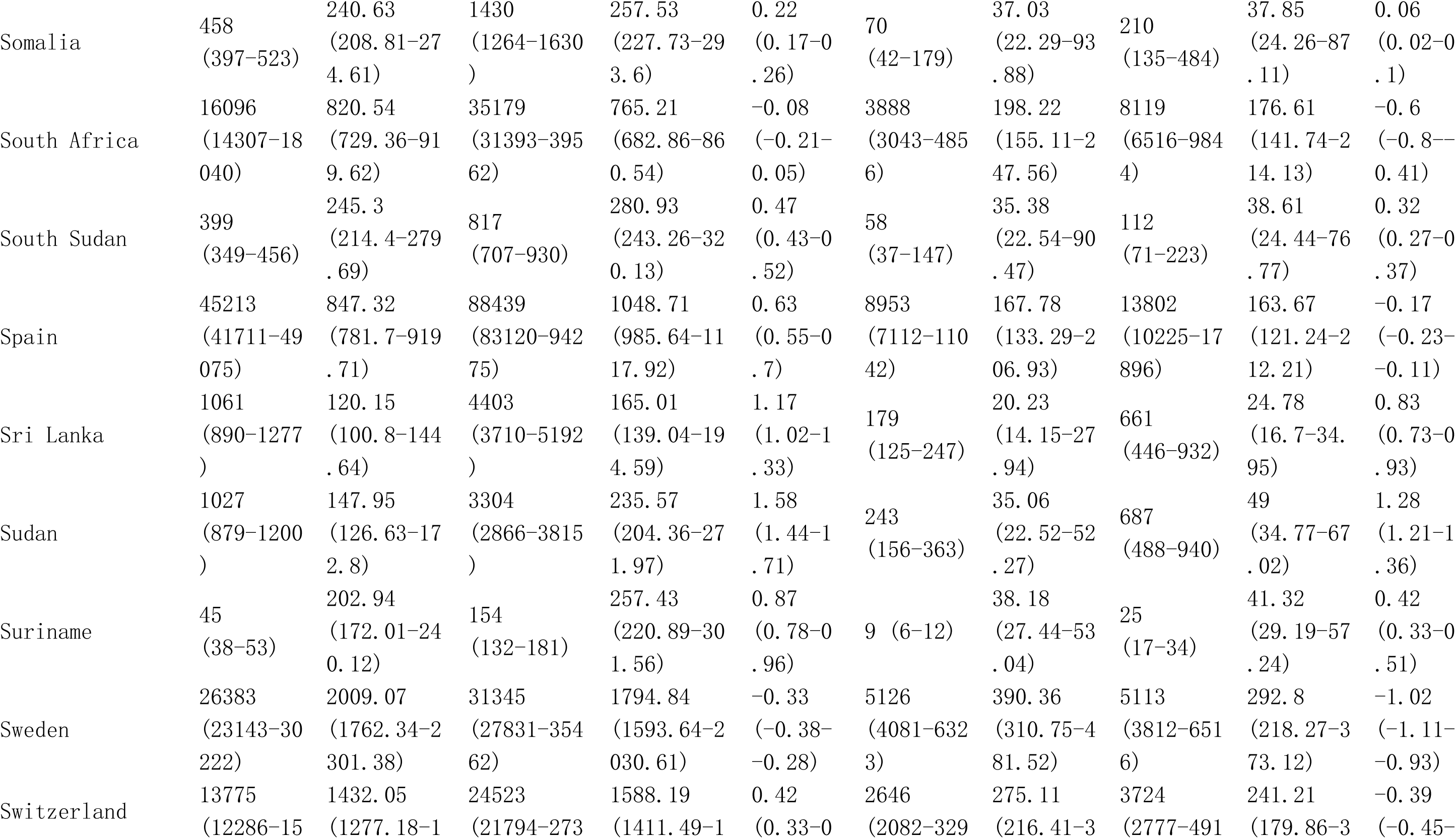
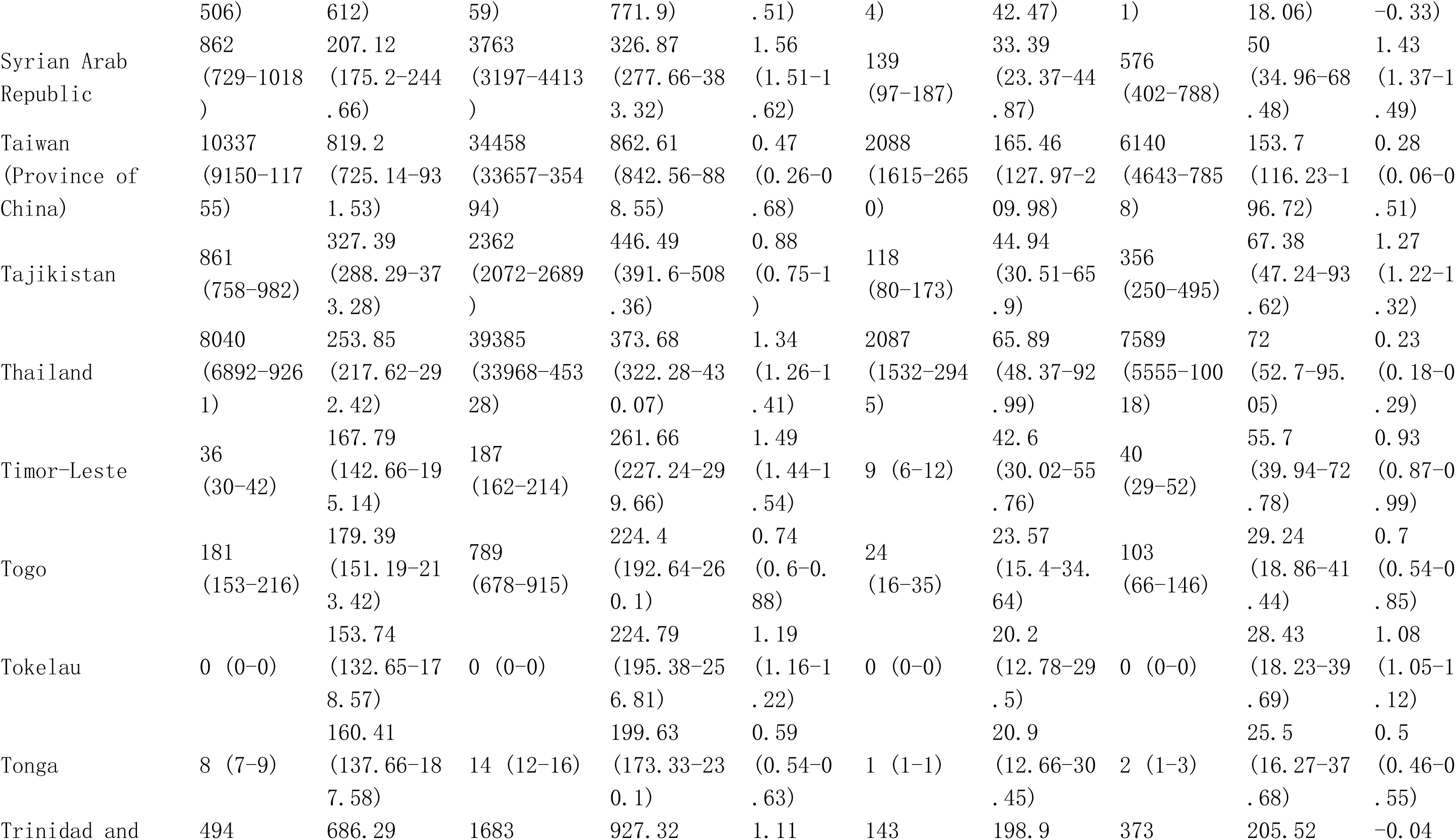
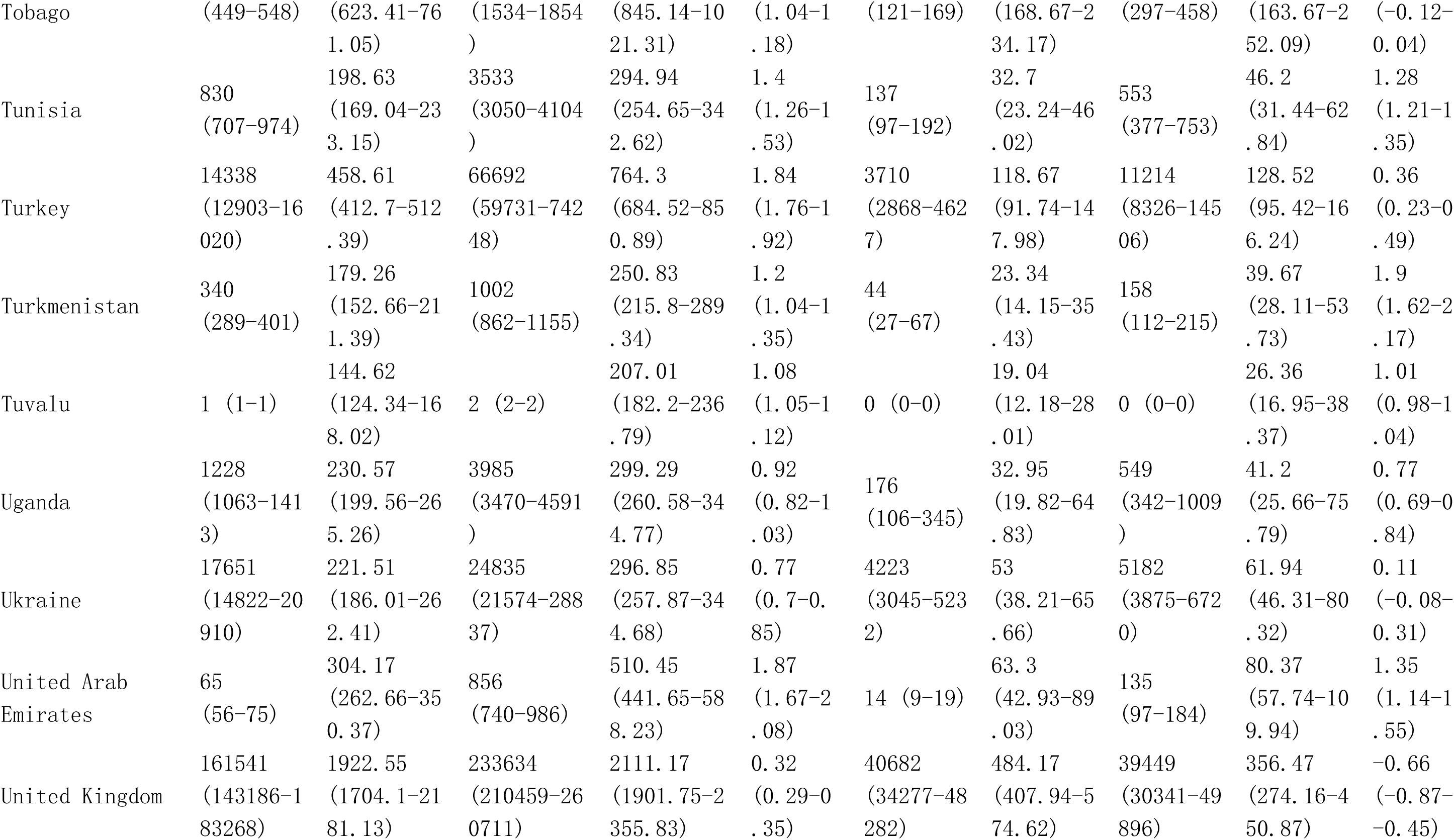
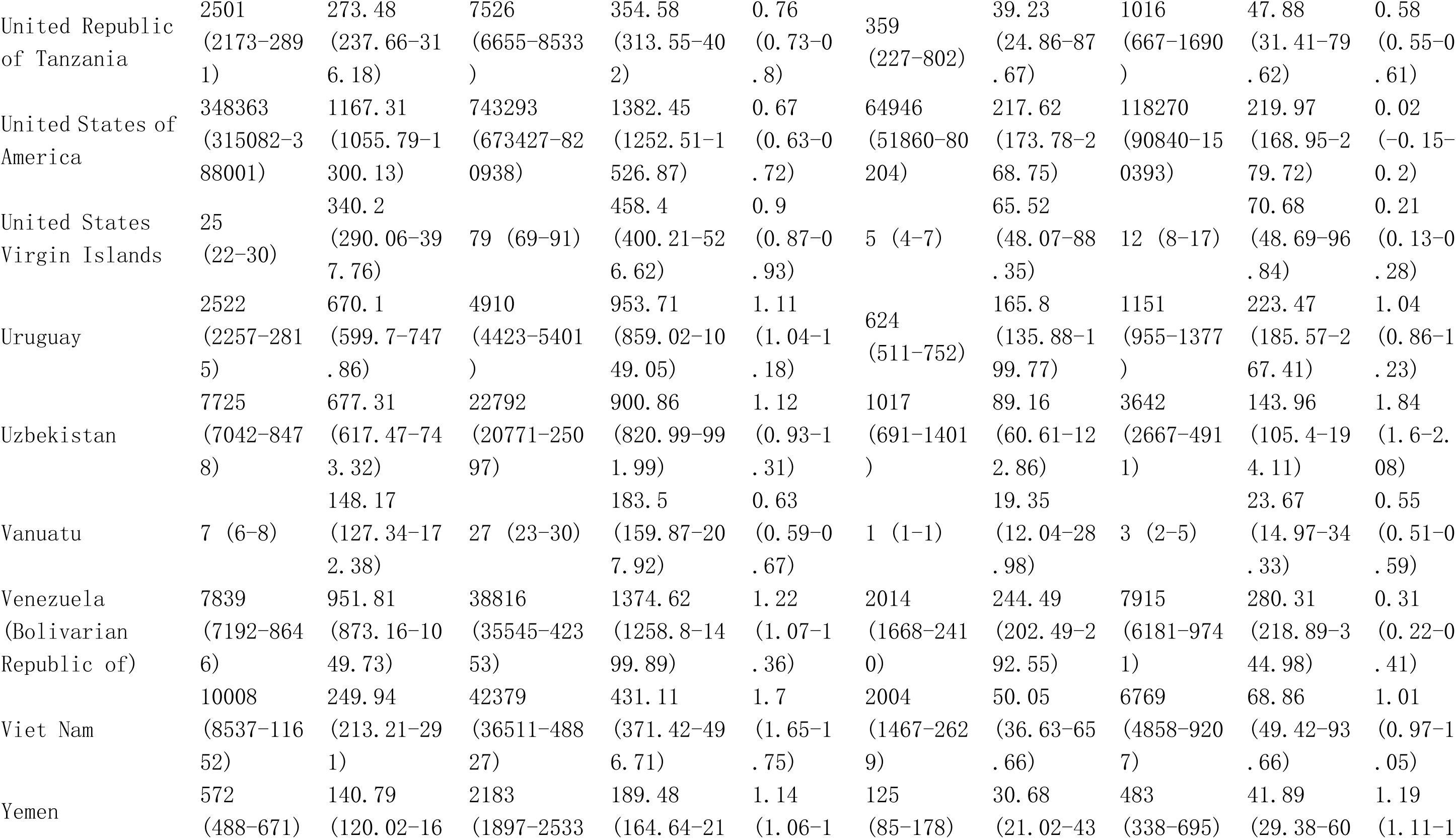
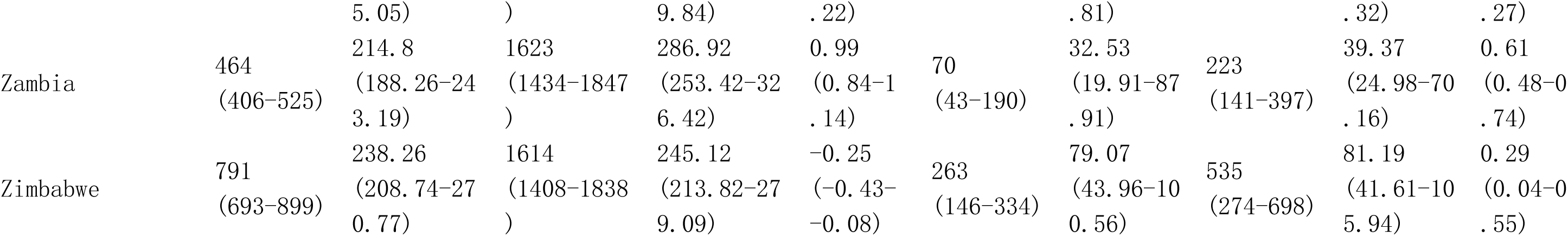
Global burden in Prevalence and DALYs of Rheumatoid Arthritis among postmenopausal women from 1990 to 2021 by 21 GBD geographical regions, and 204 countries and territories.

**Table S7.**
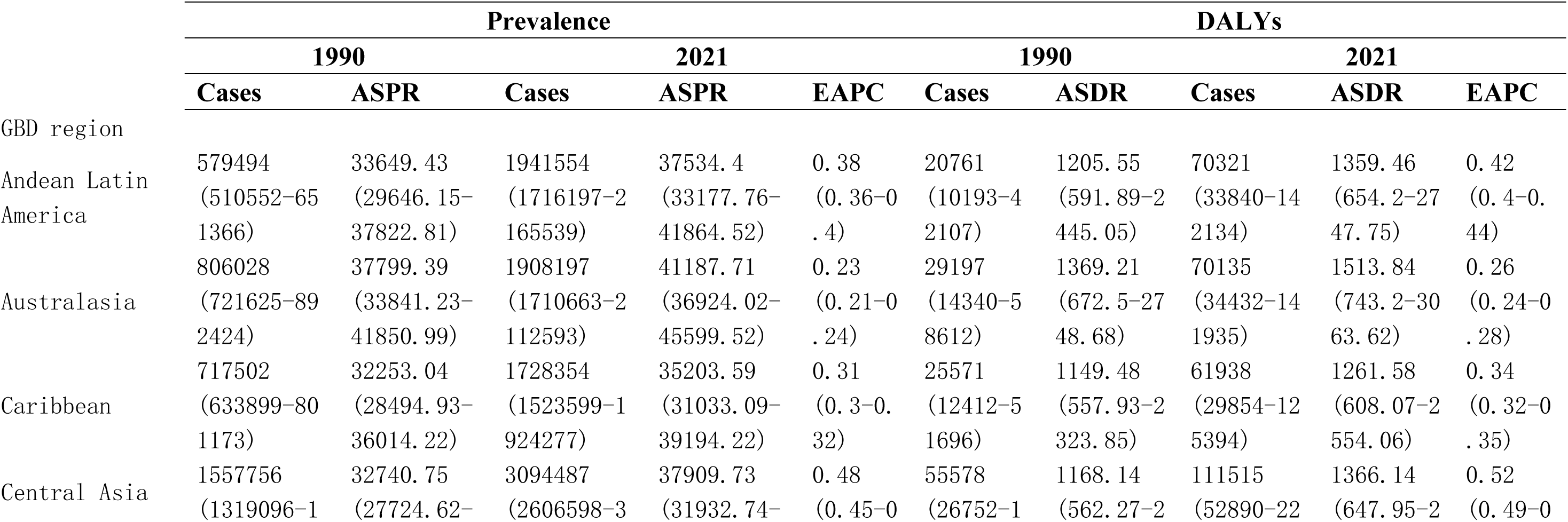
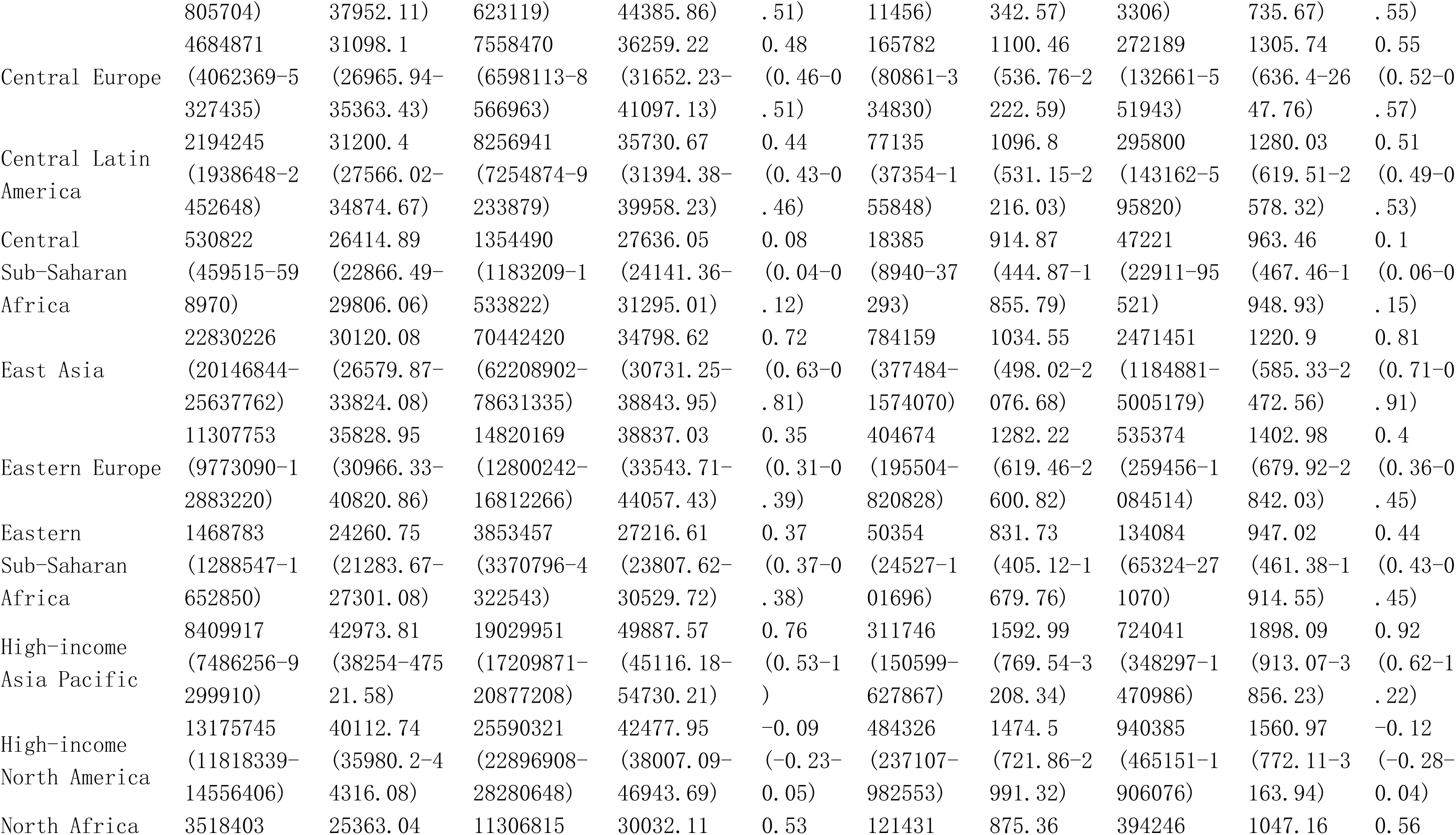
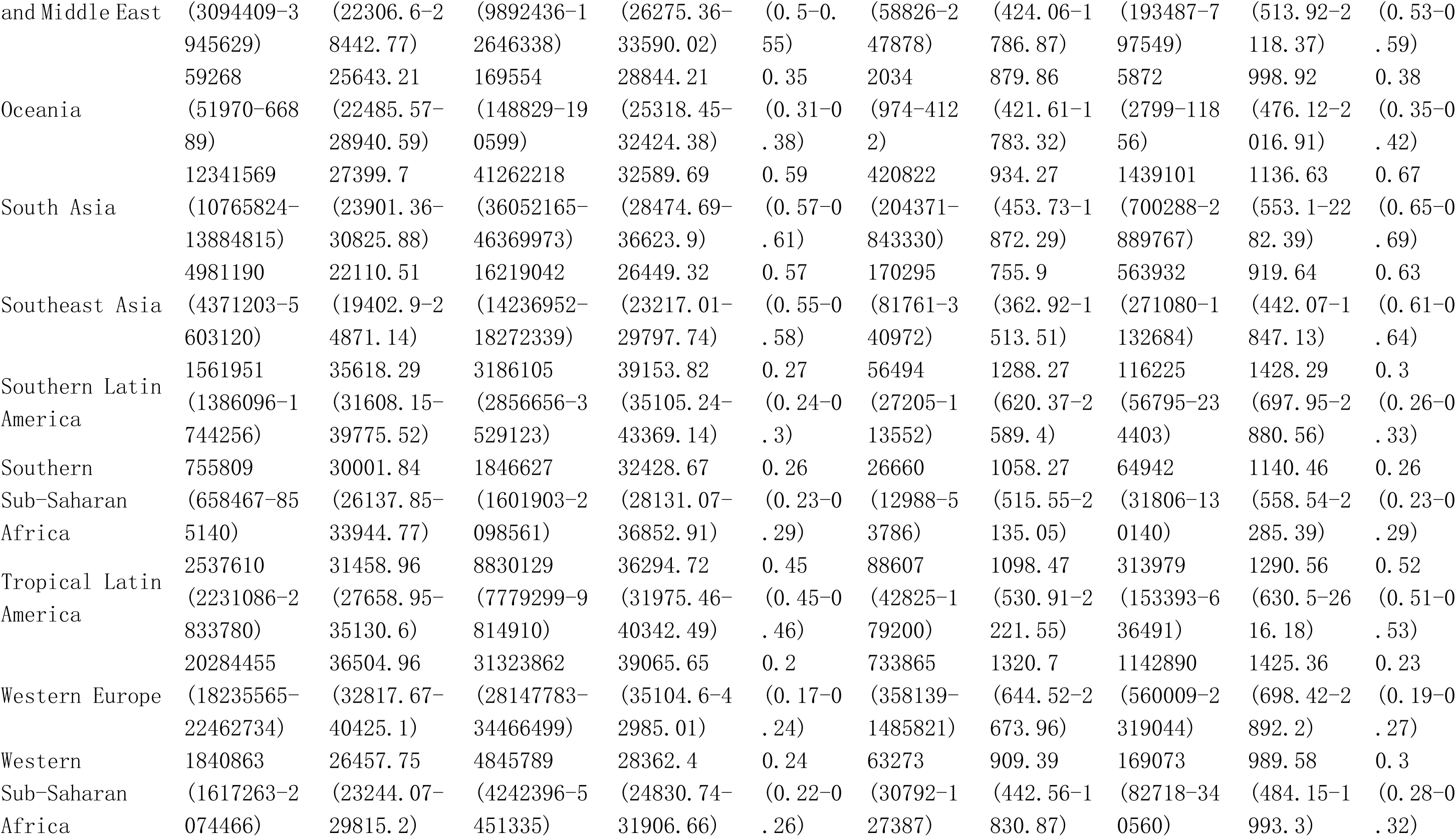
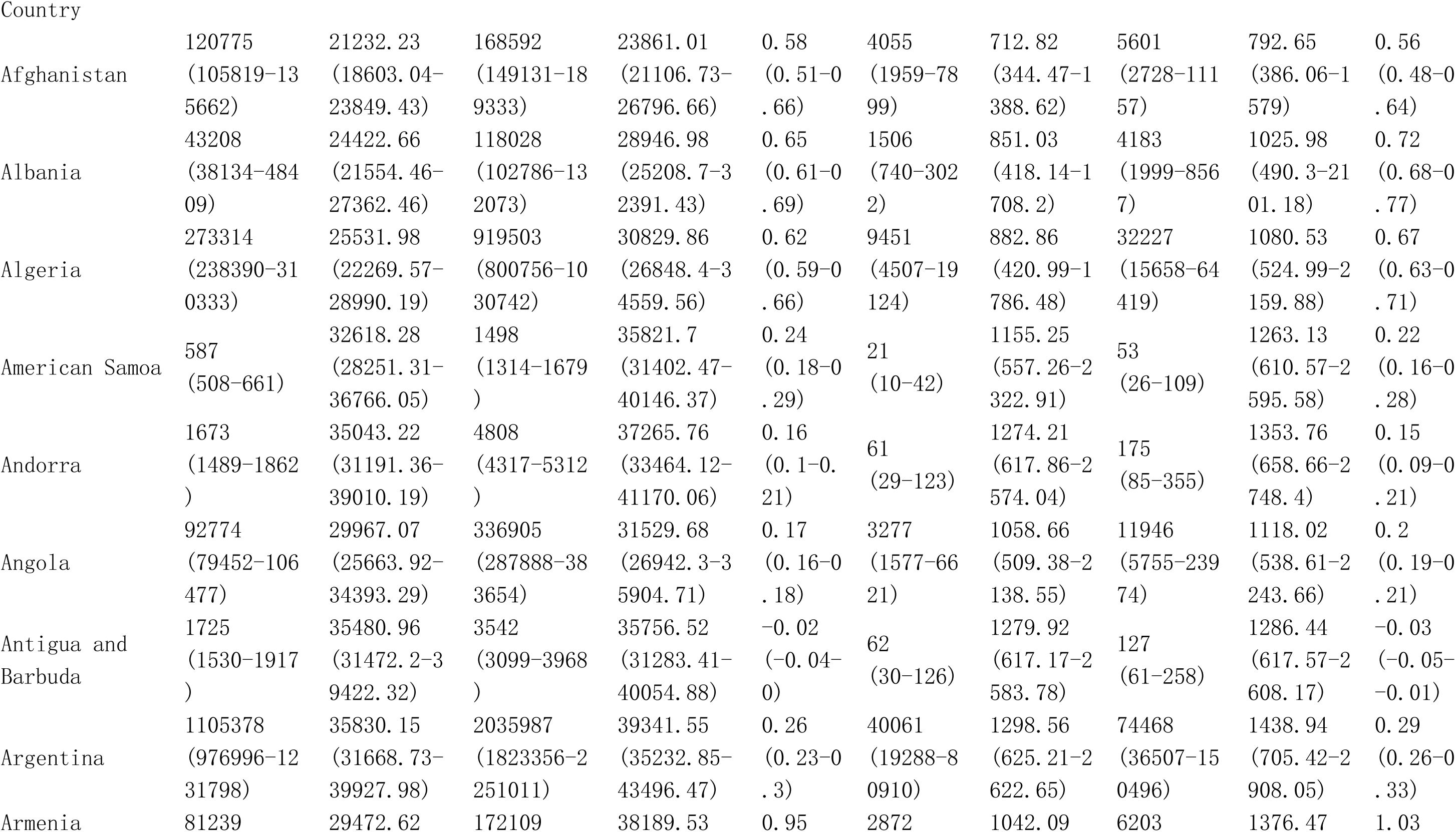
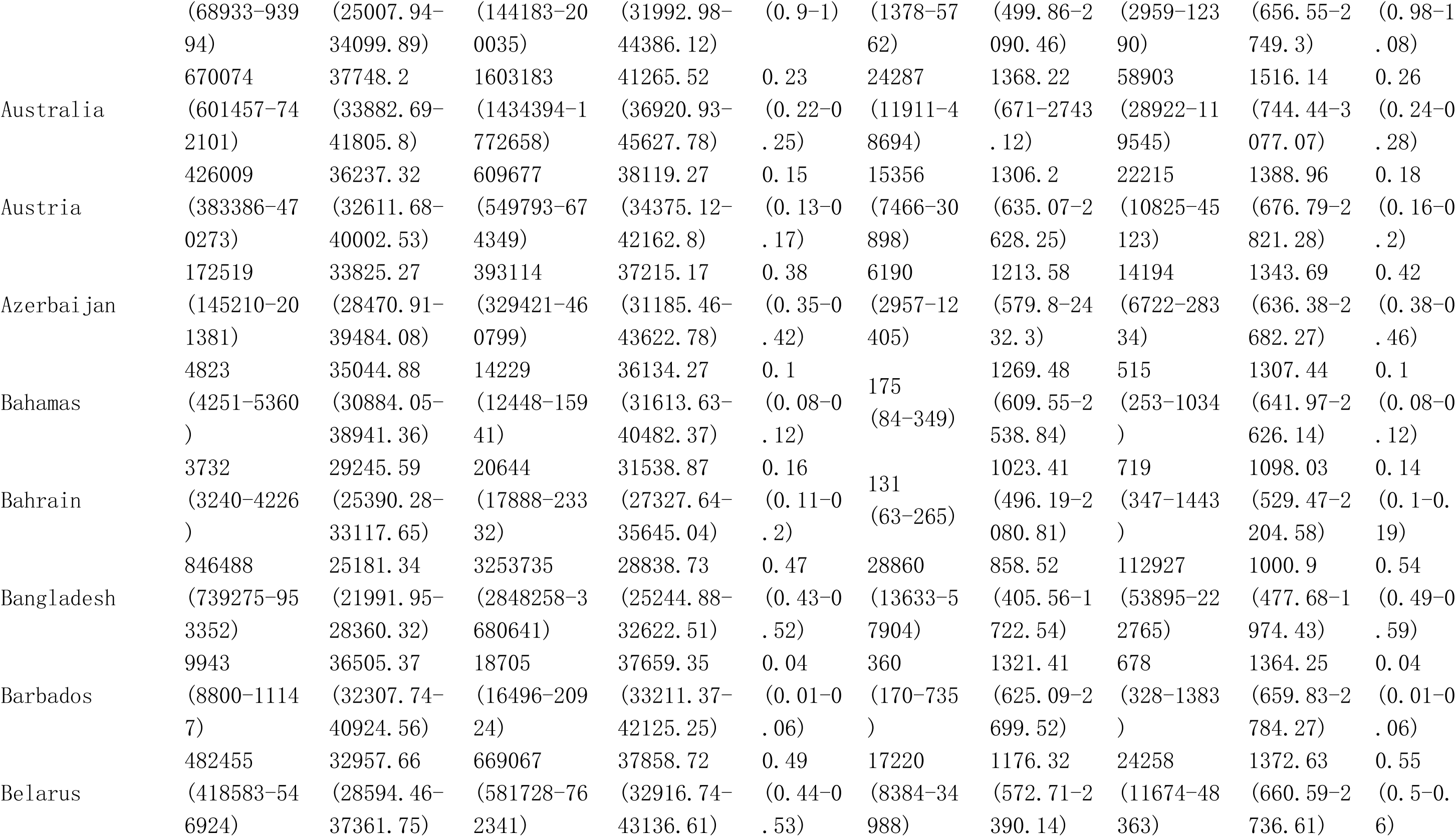
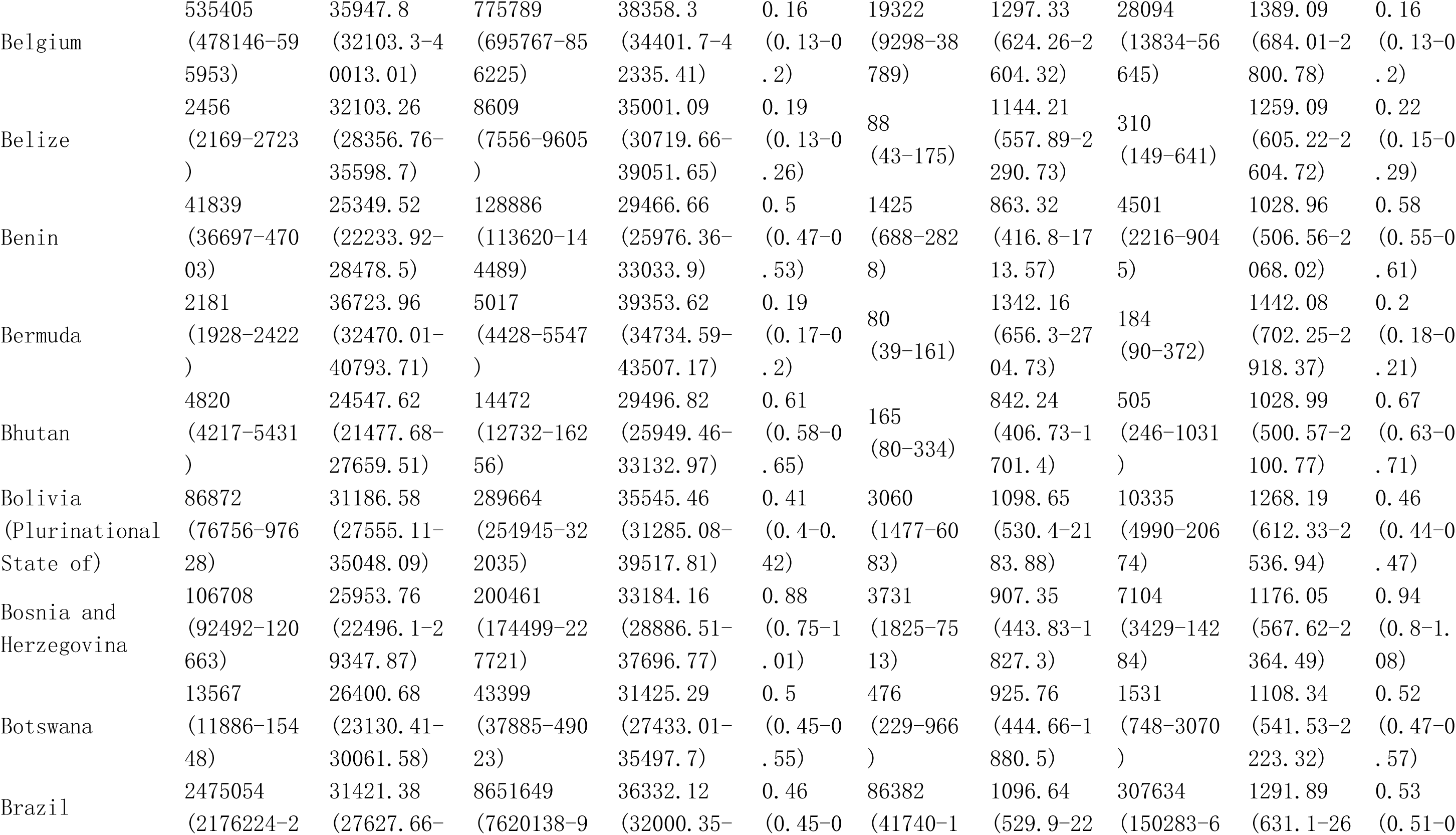
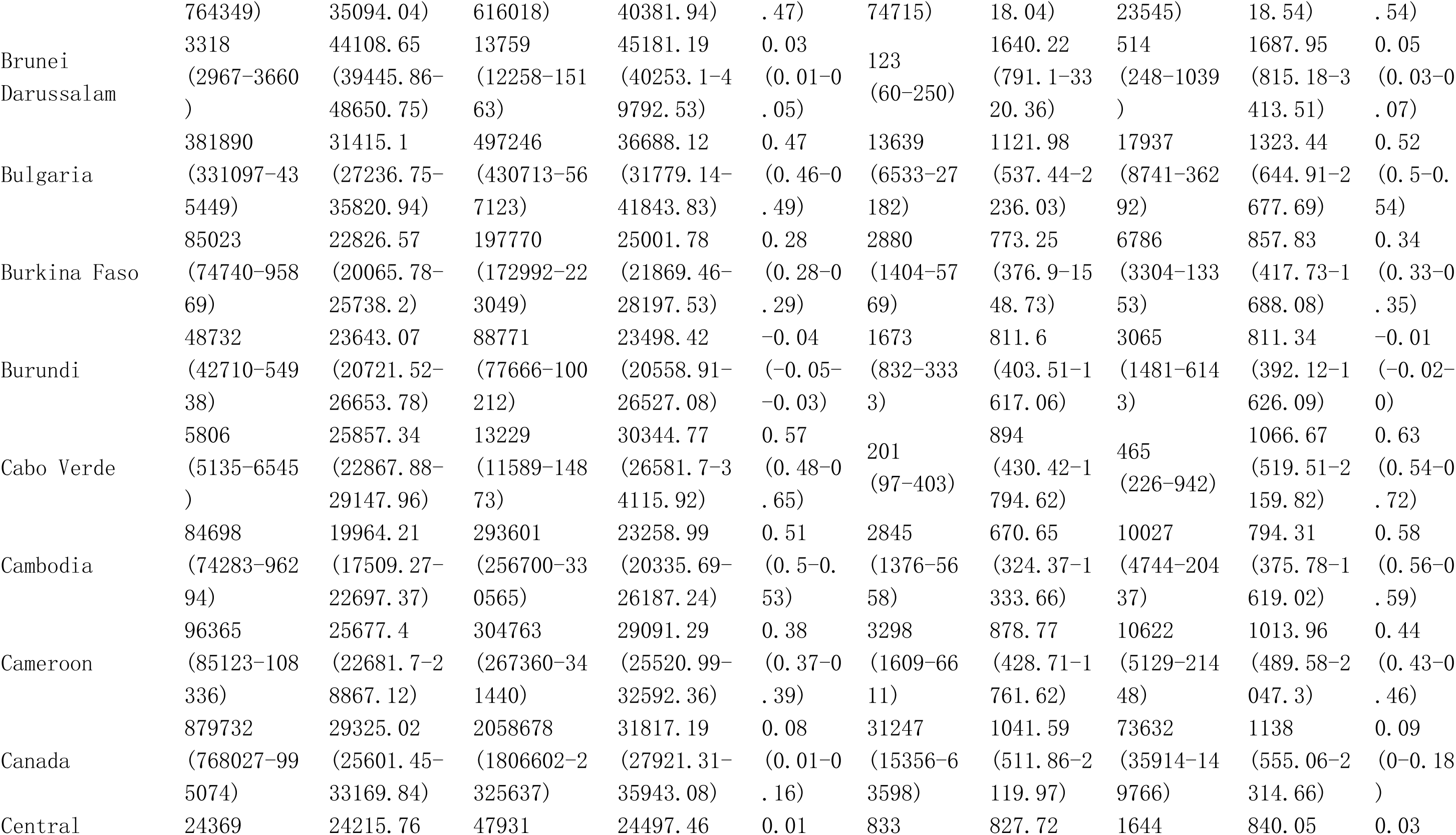
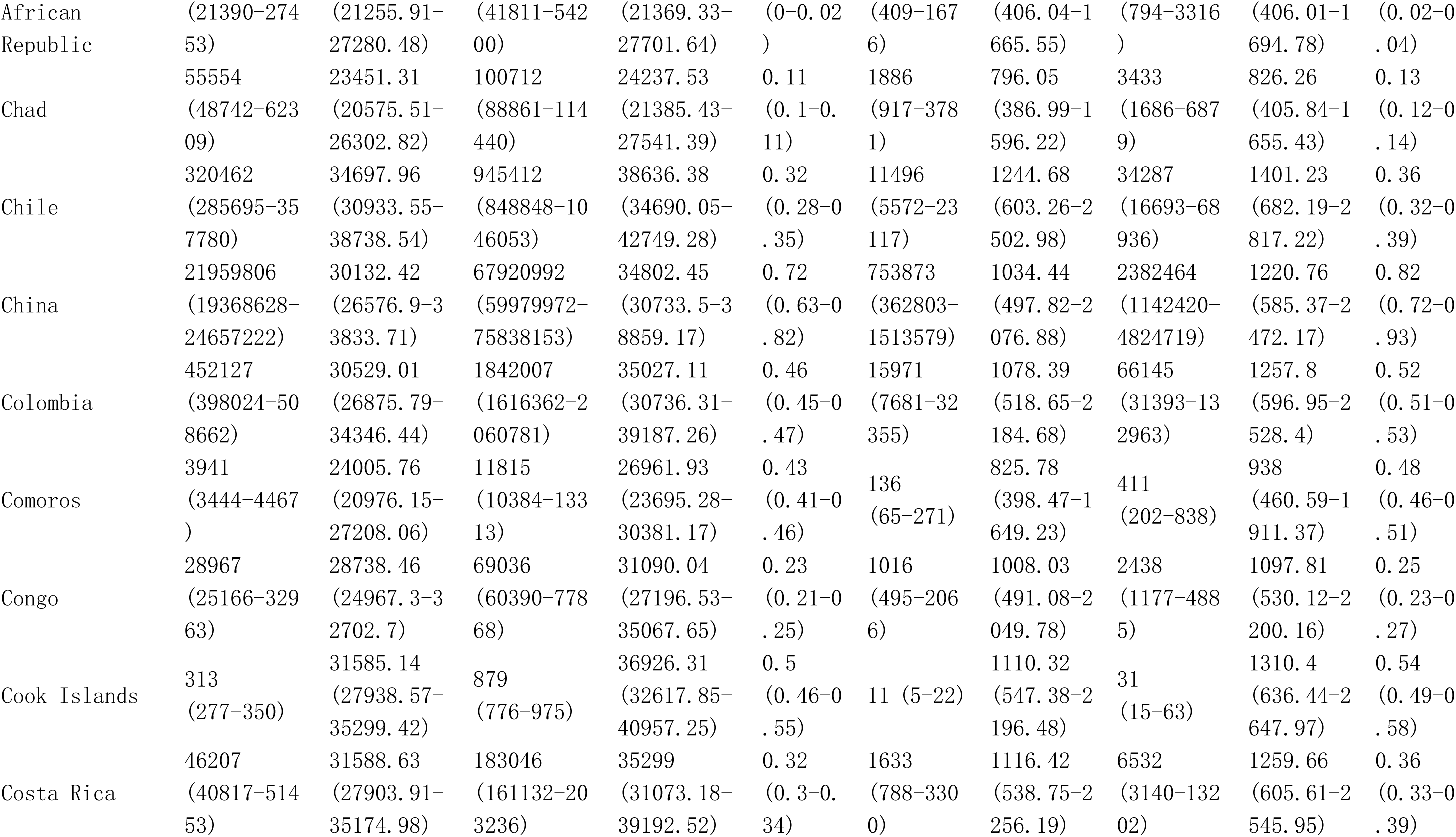
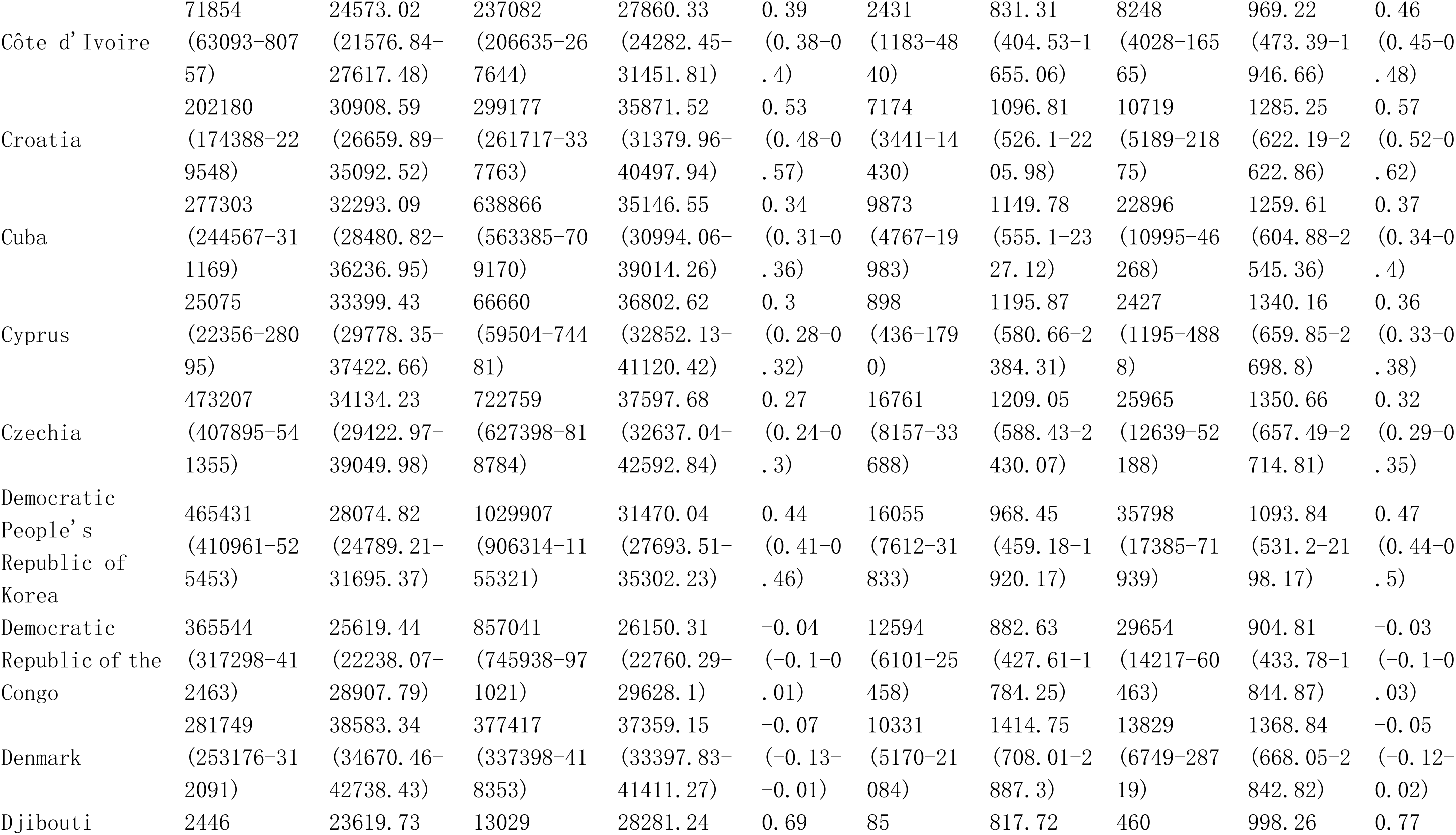
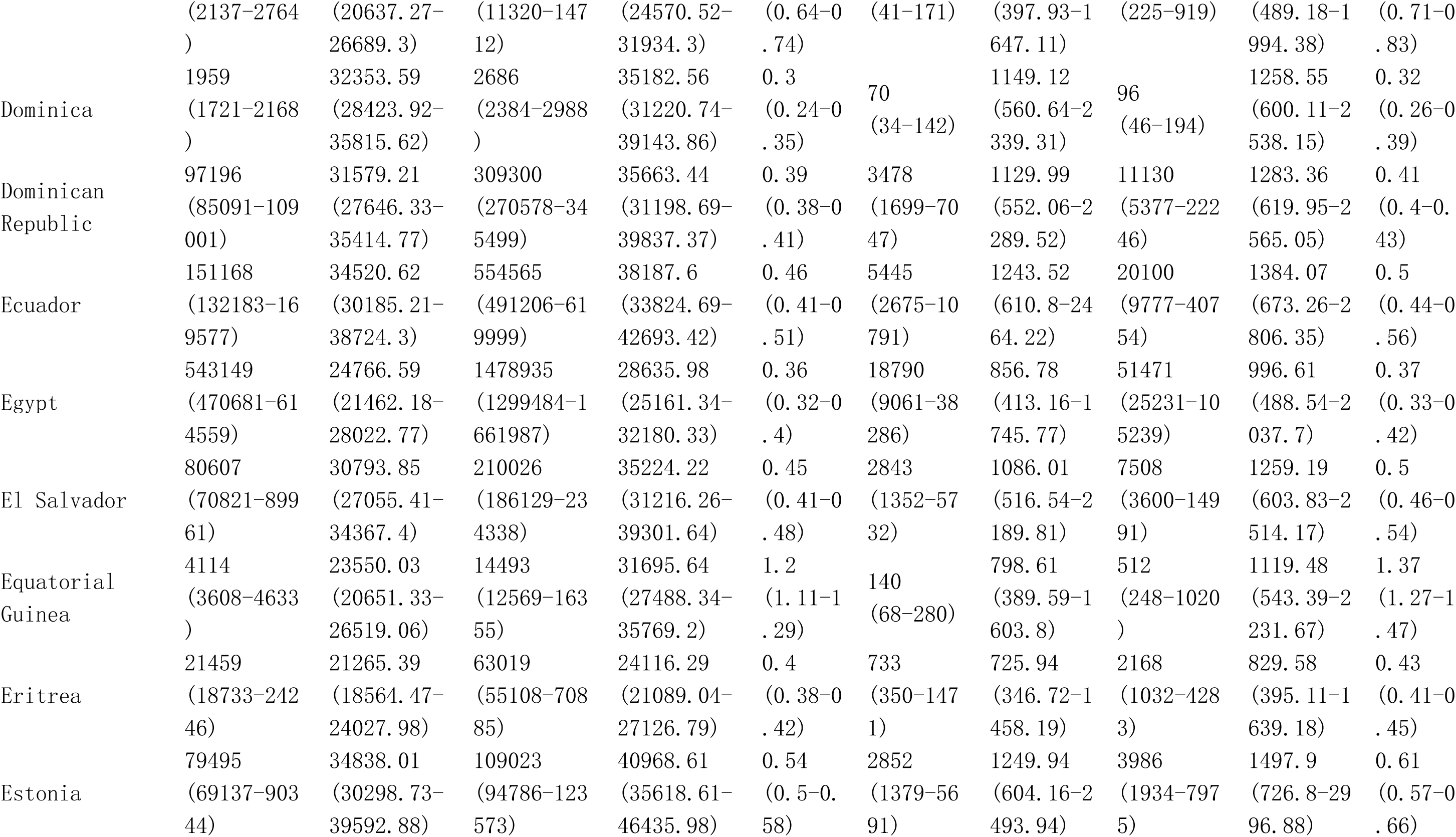
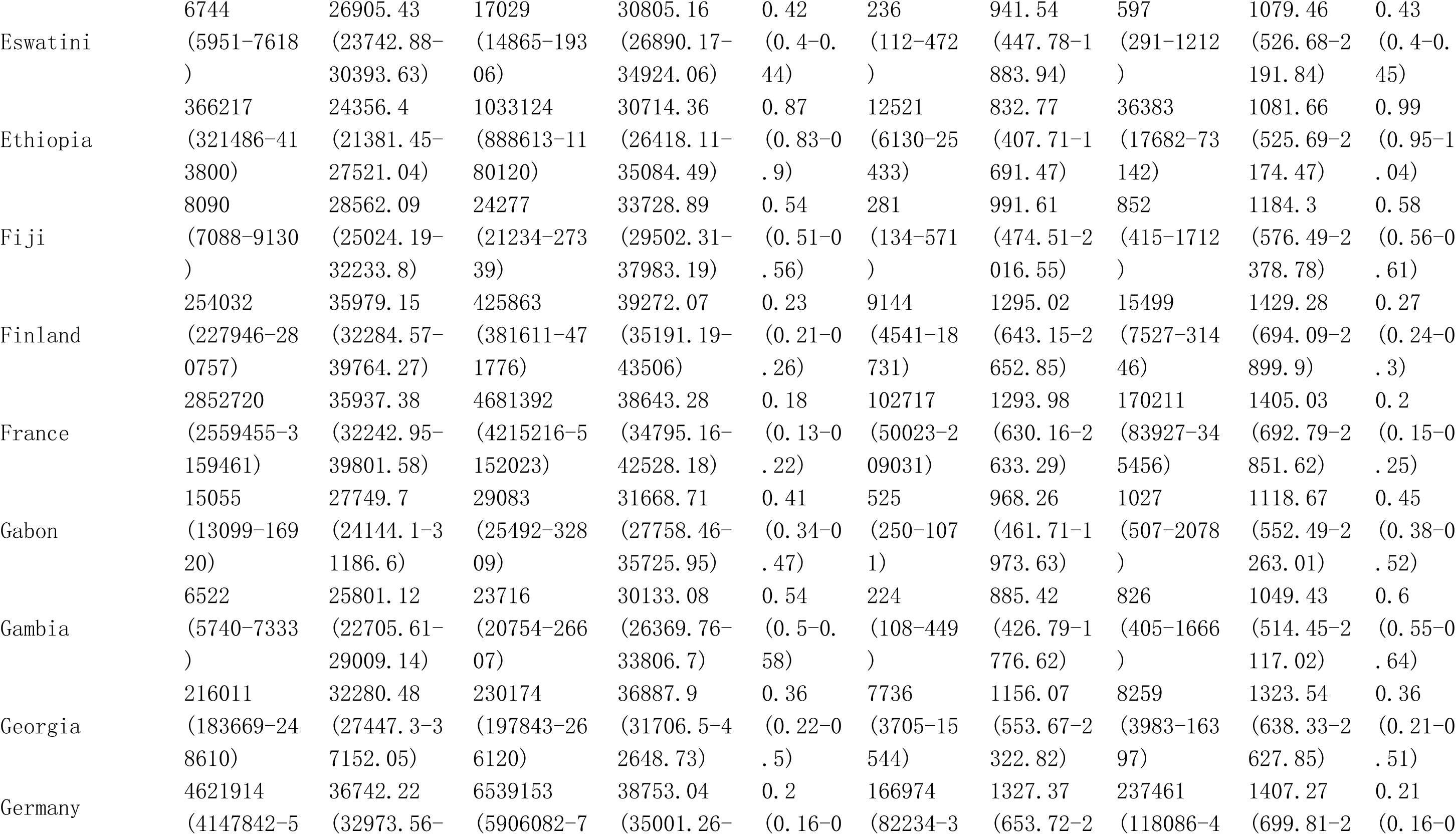
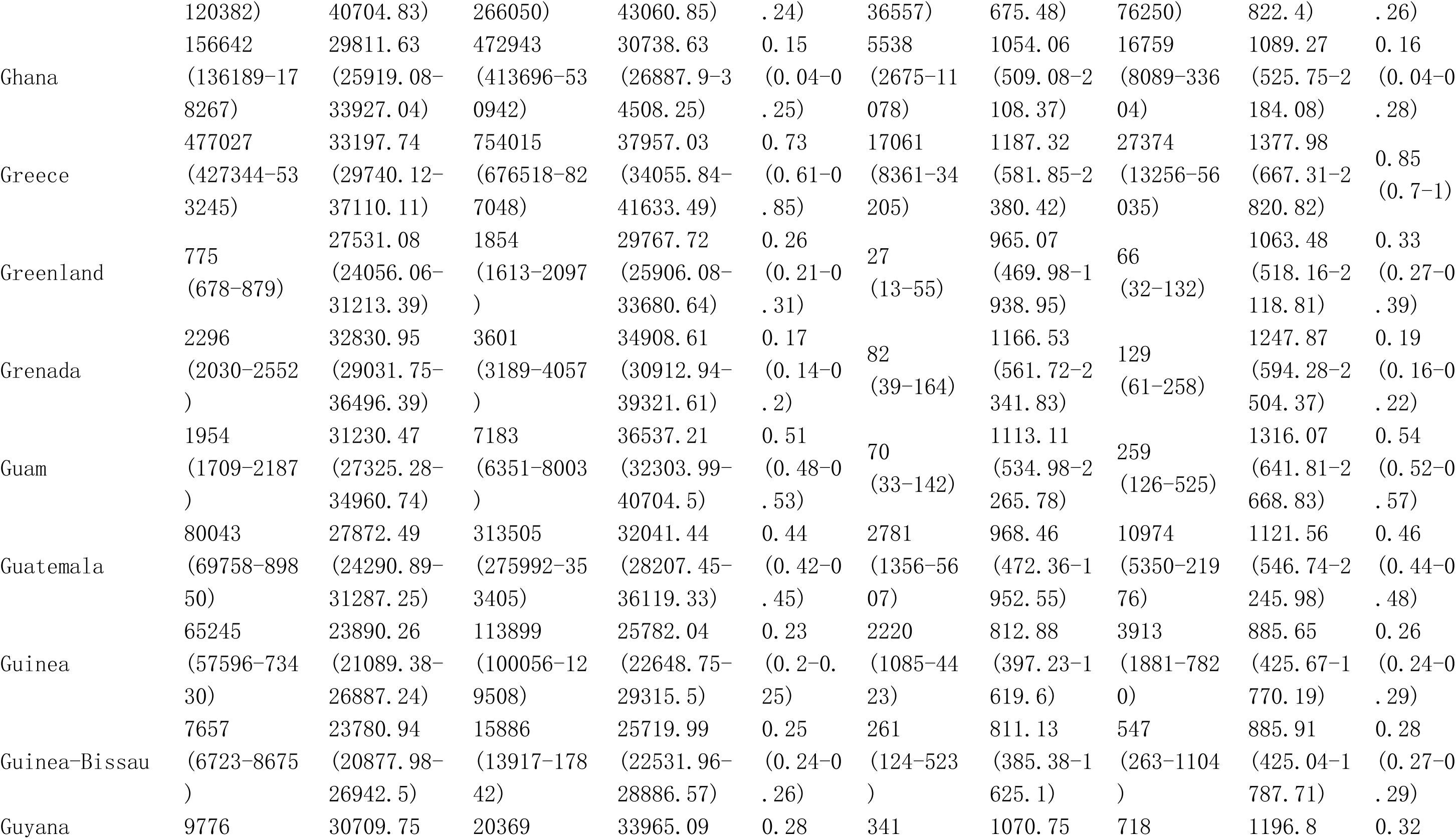
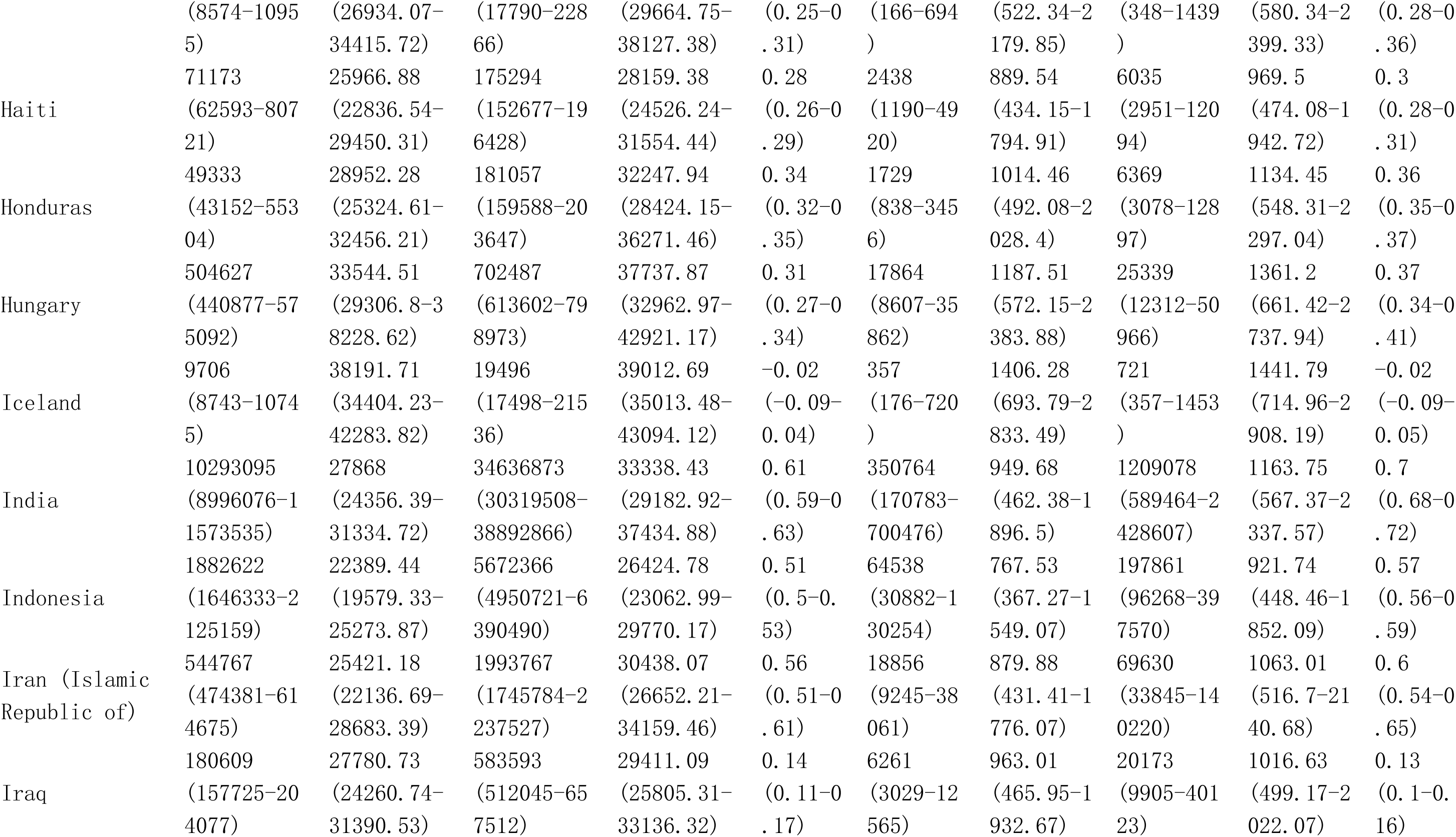
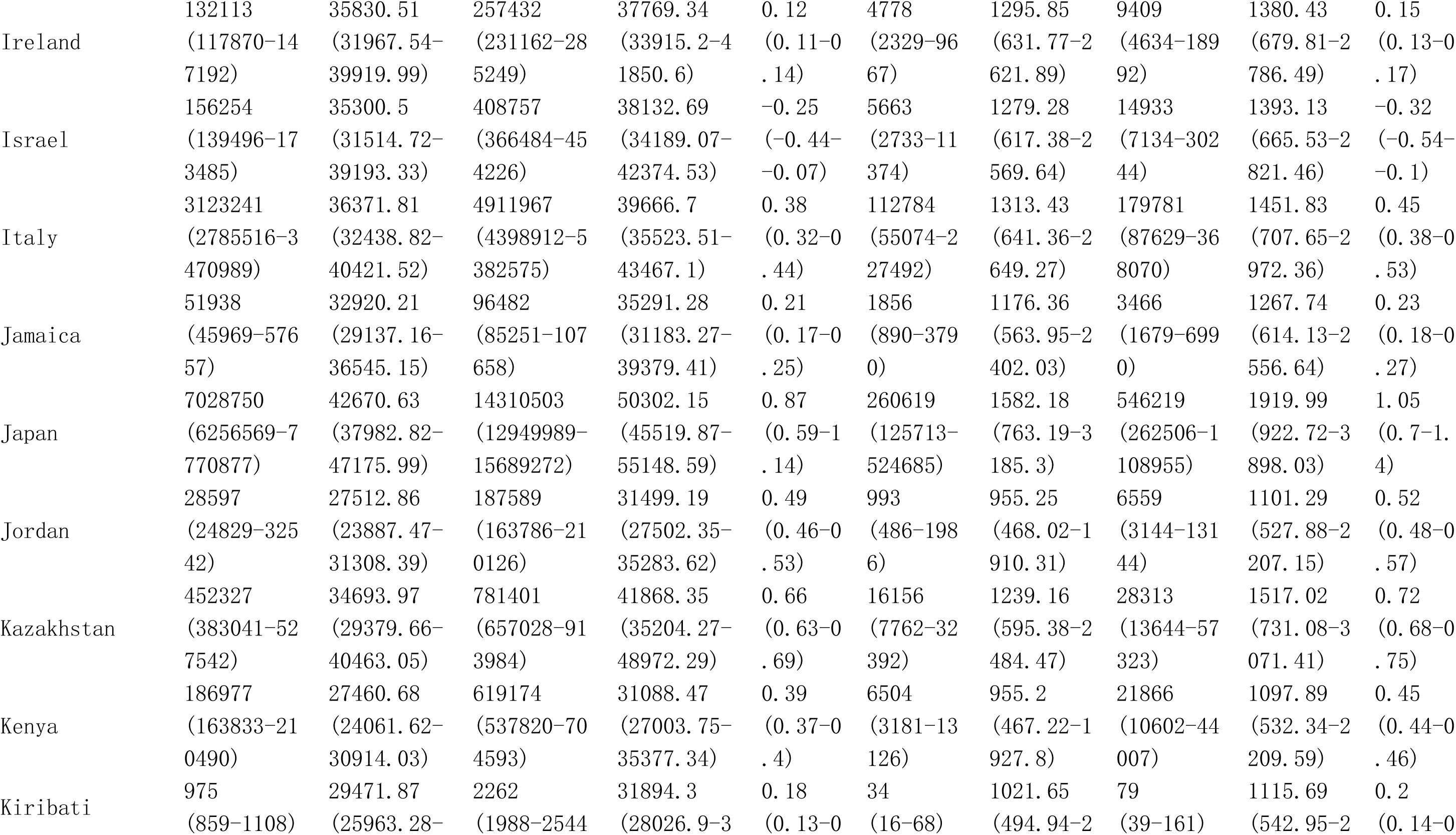
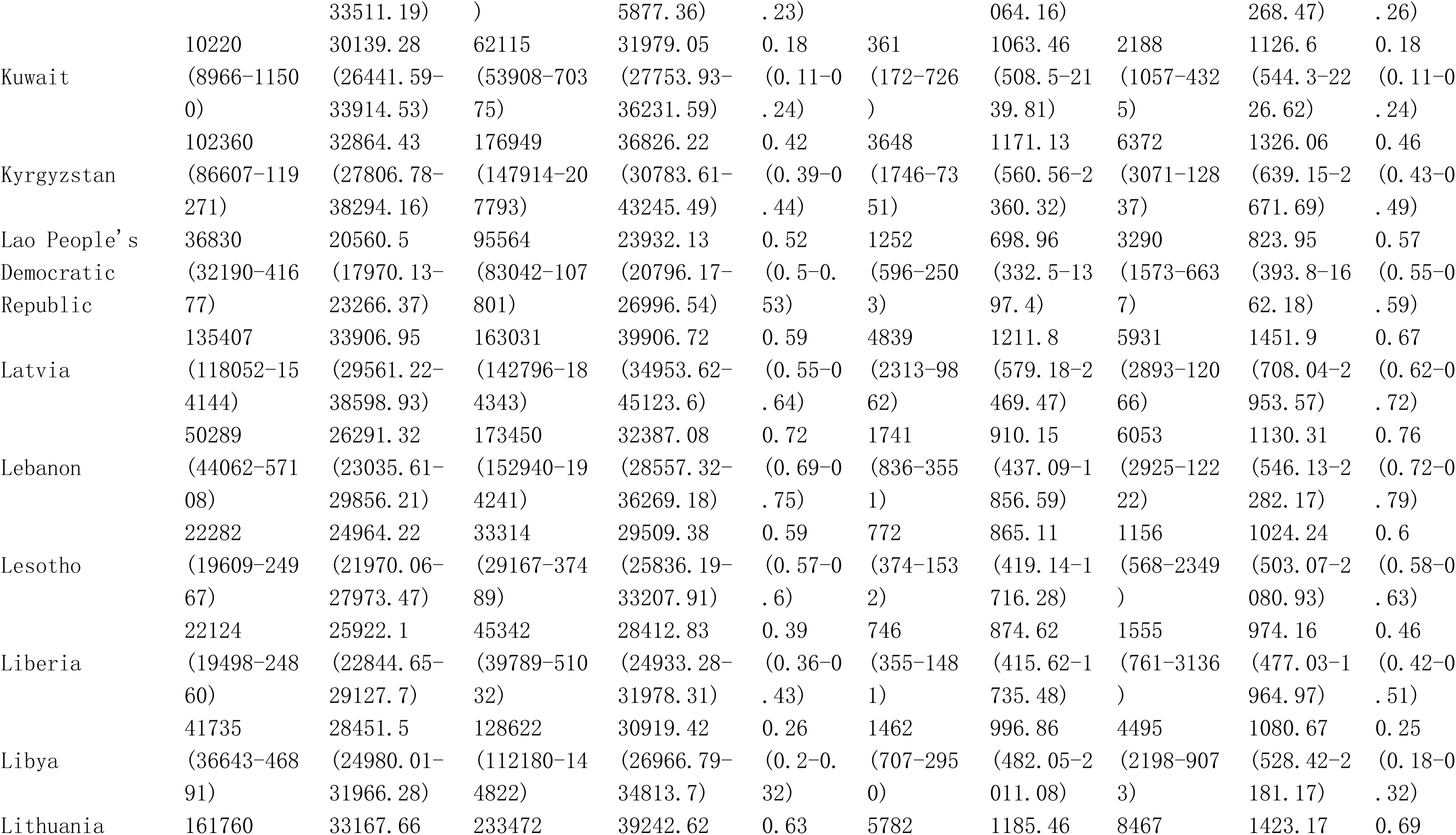
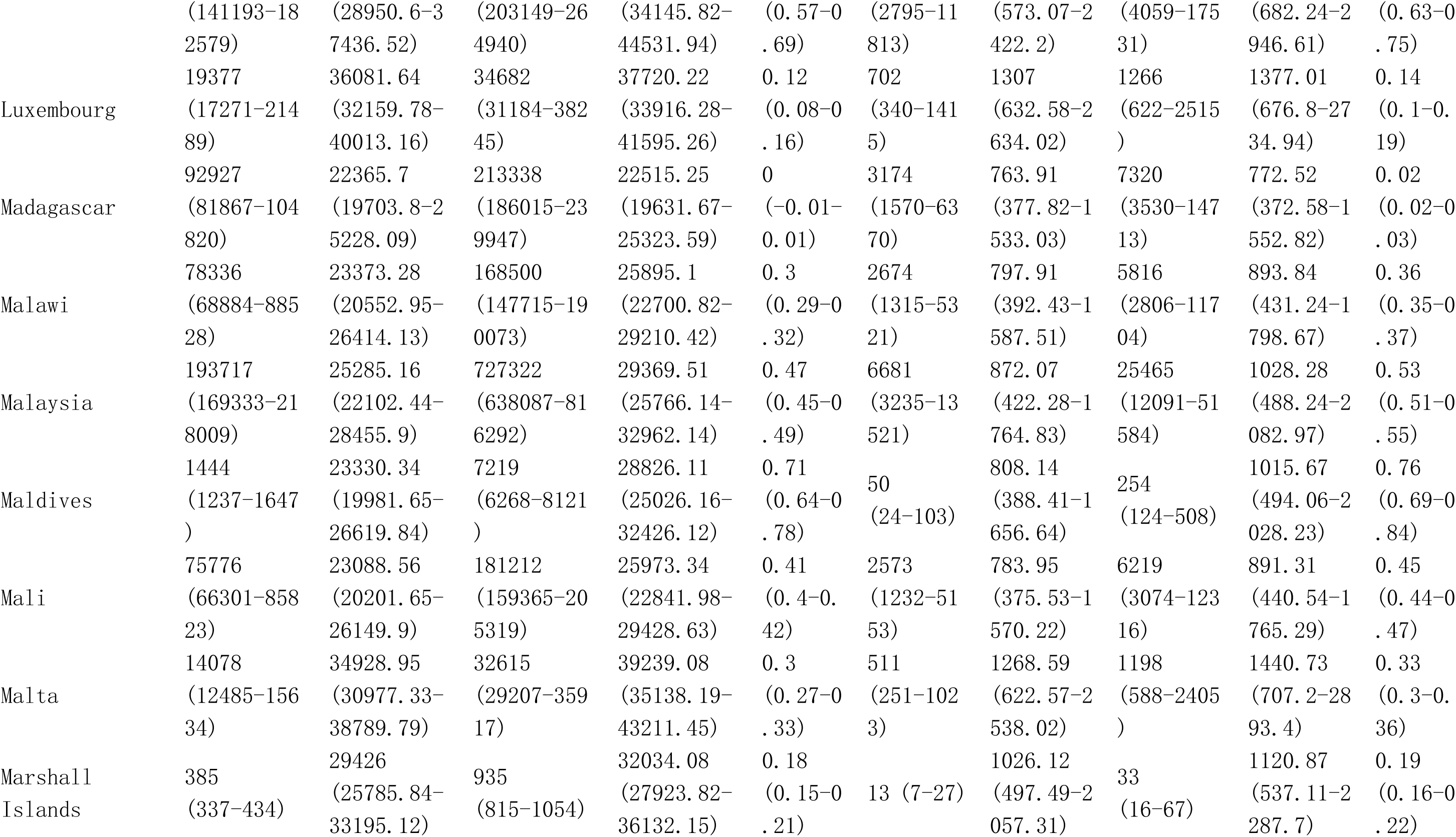
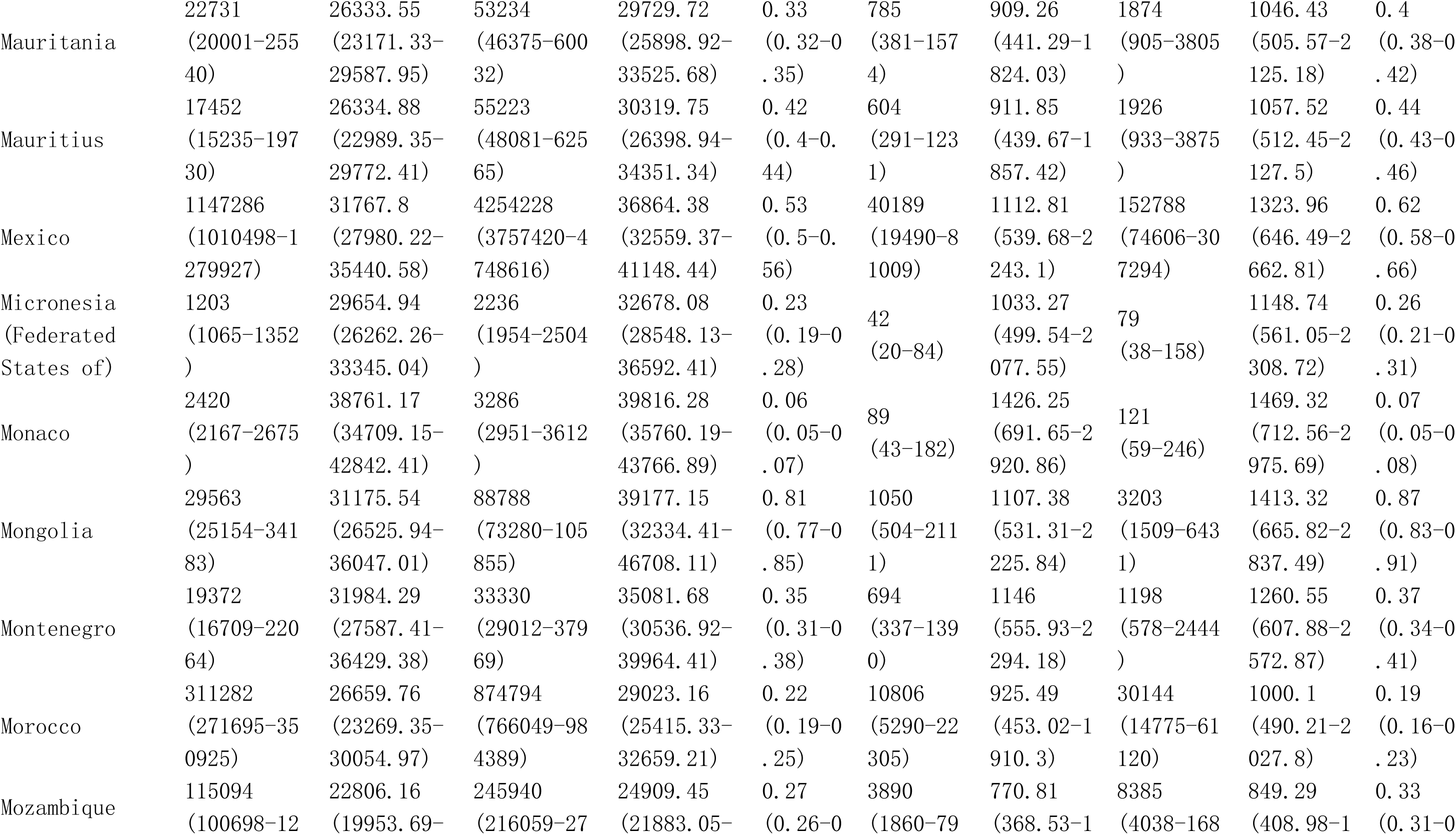
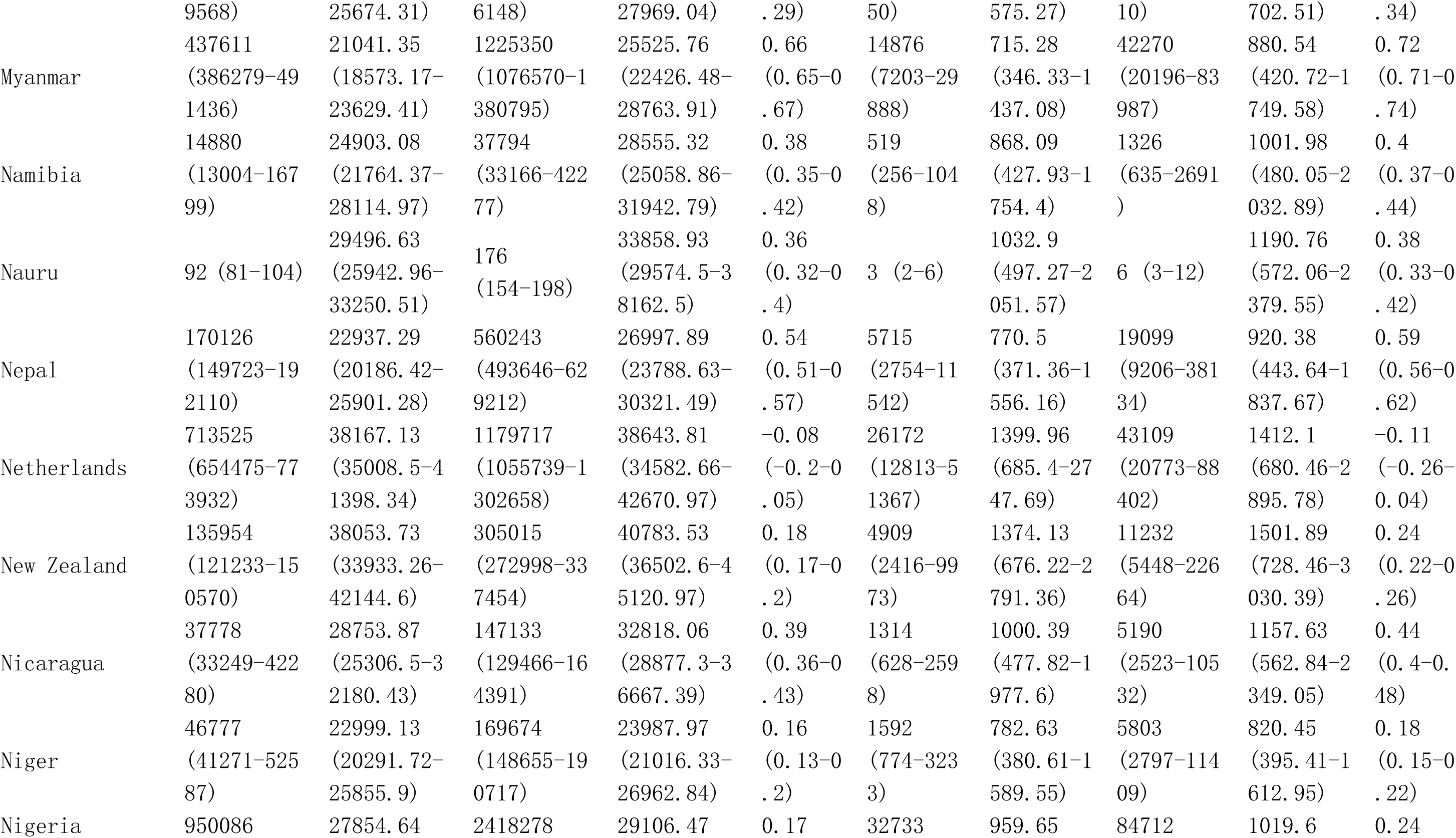
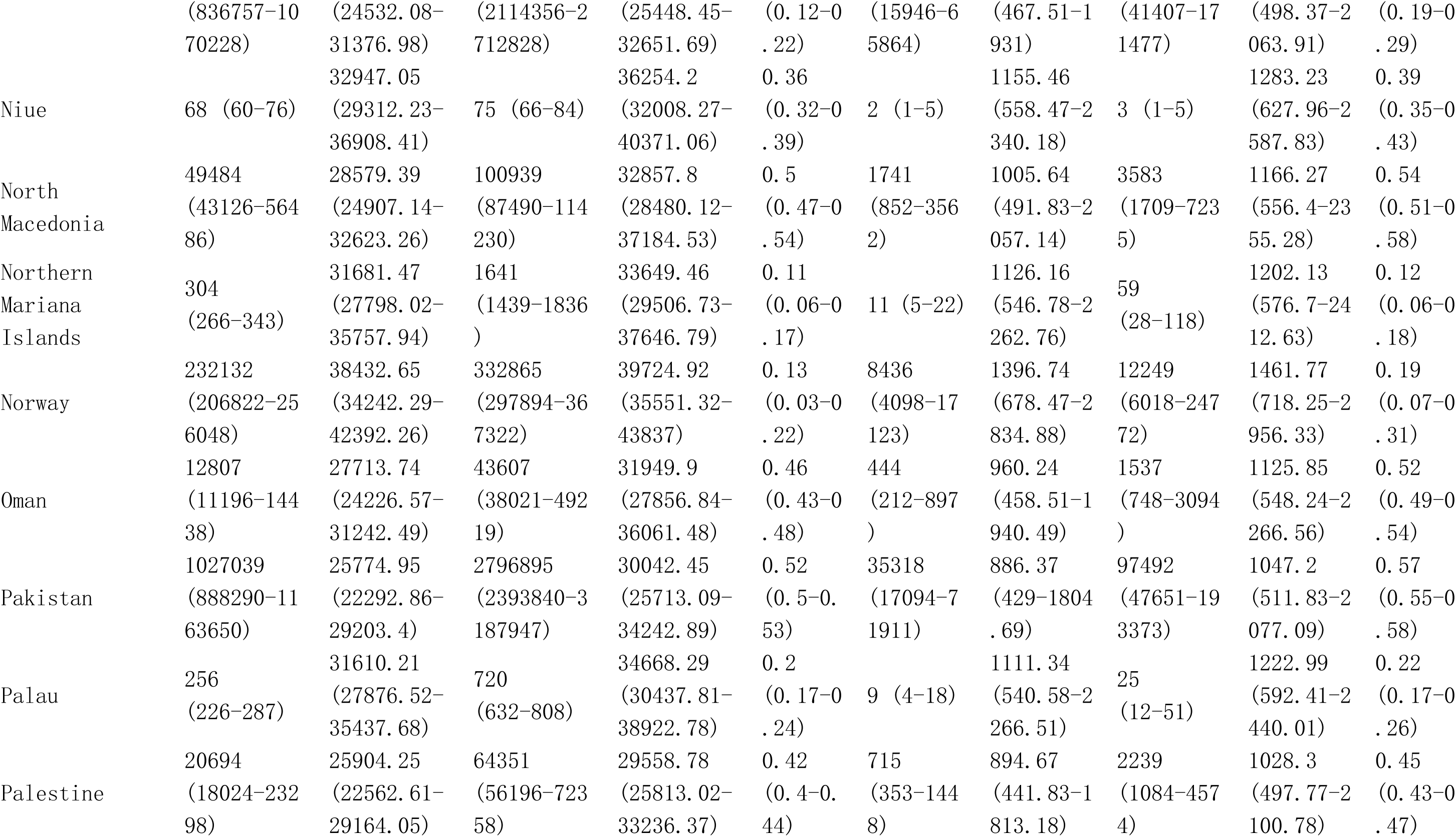
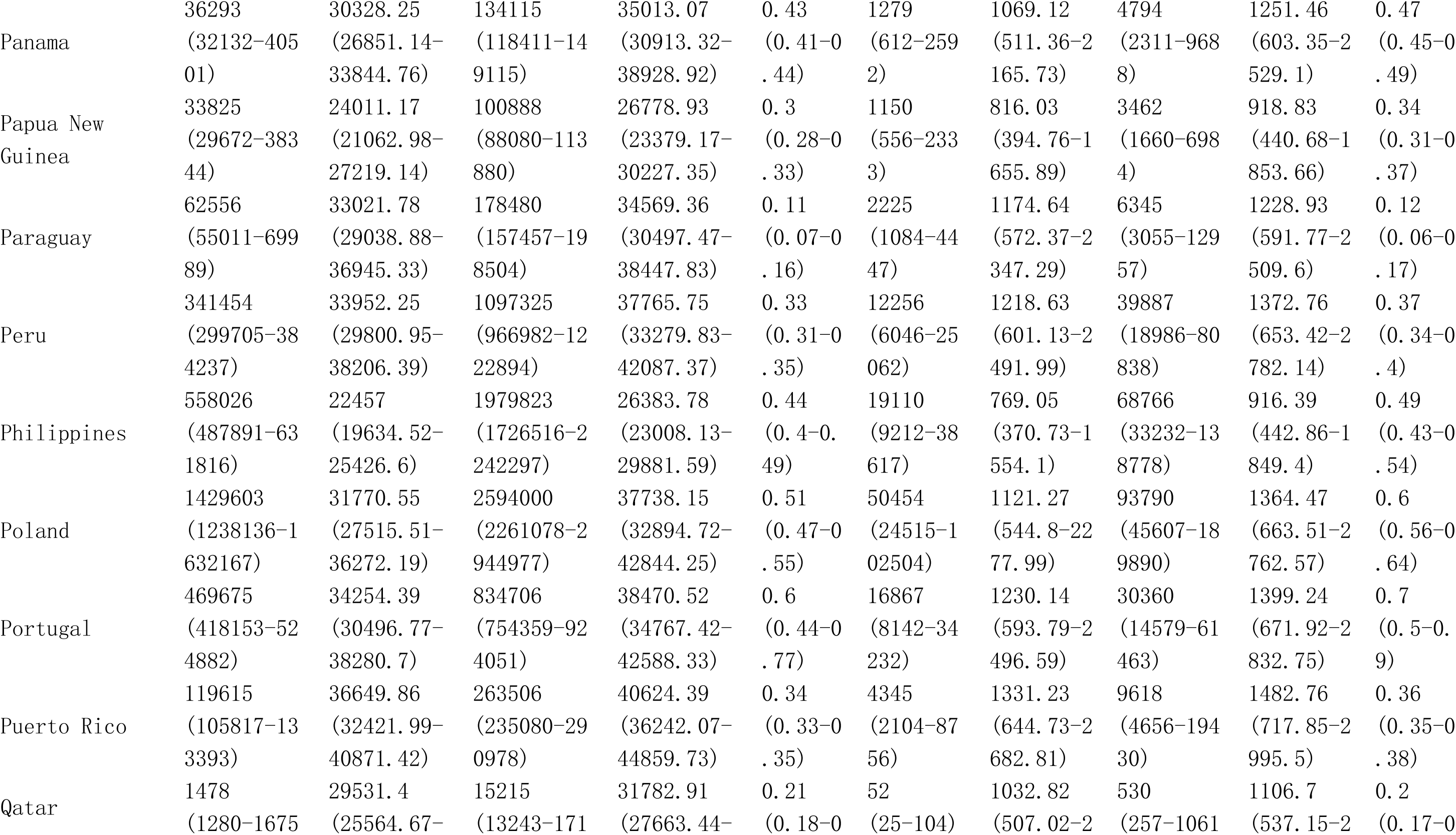
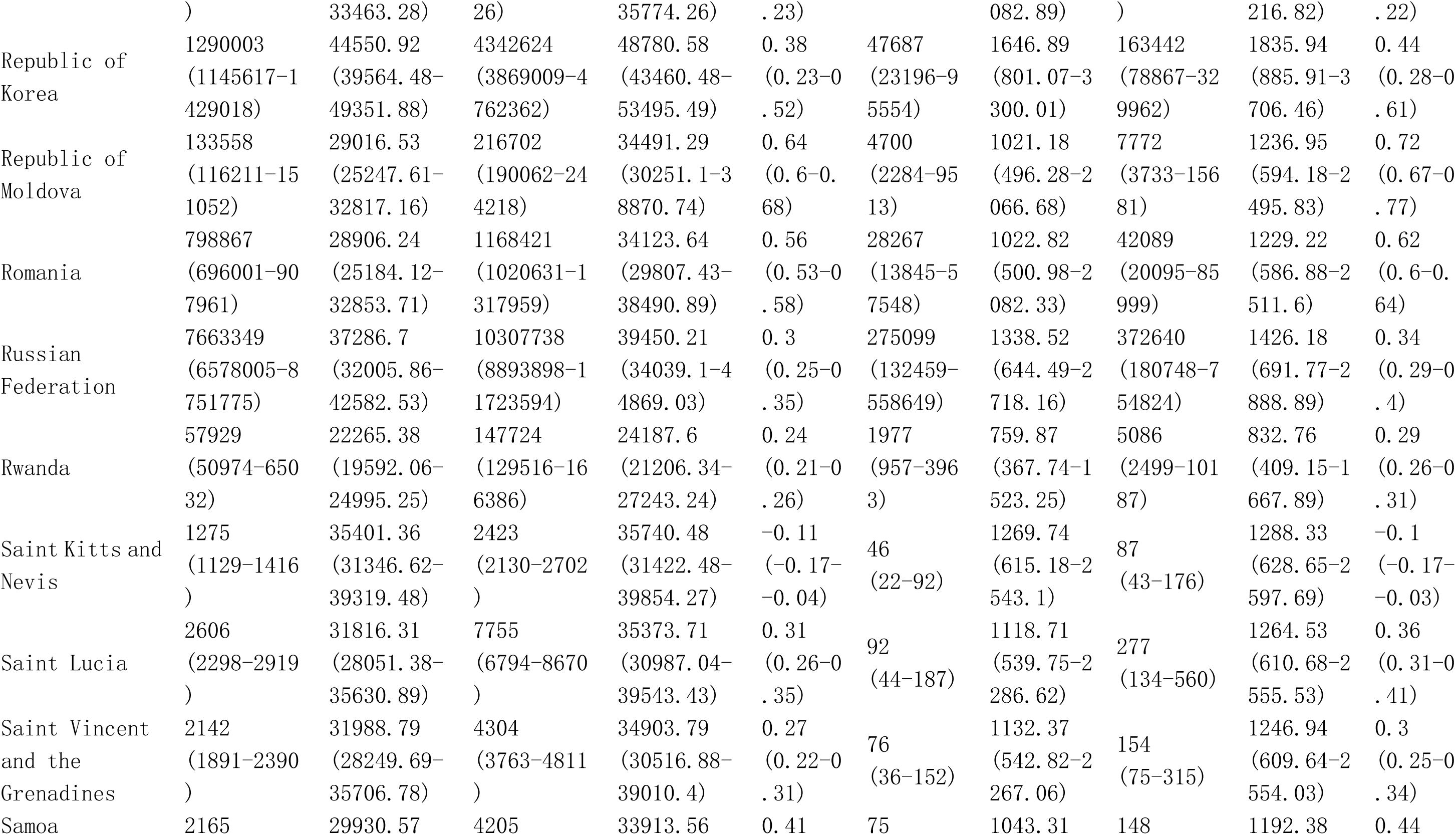
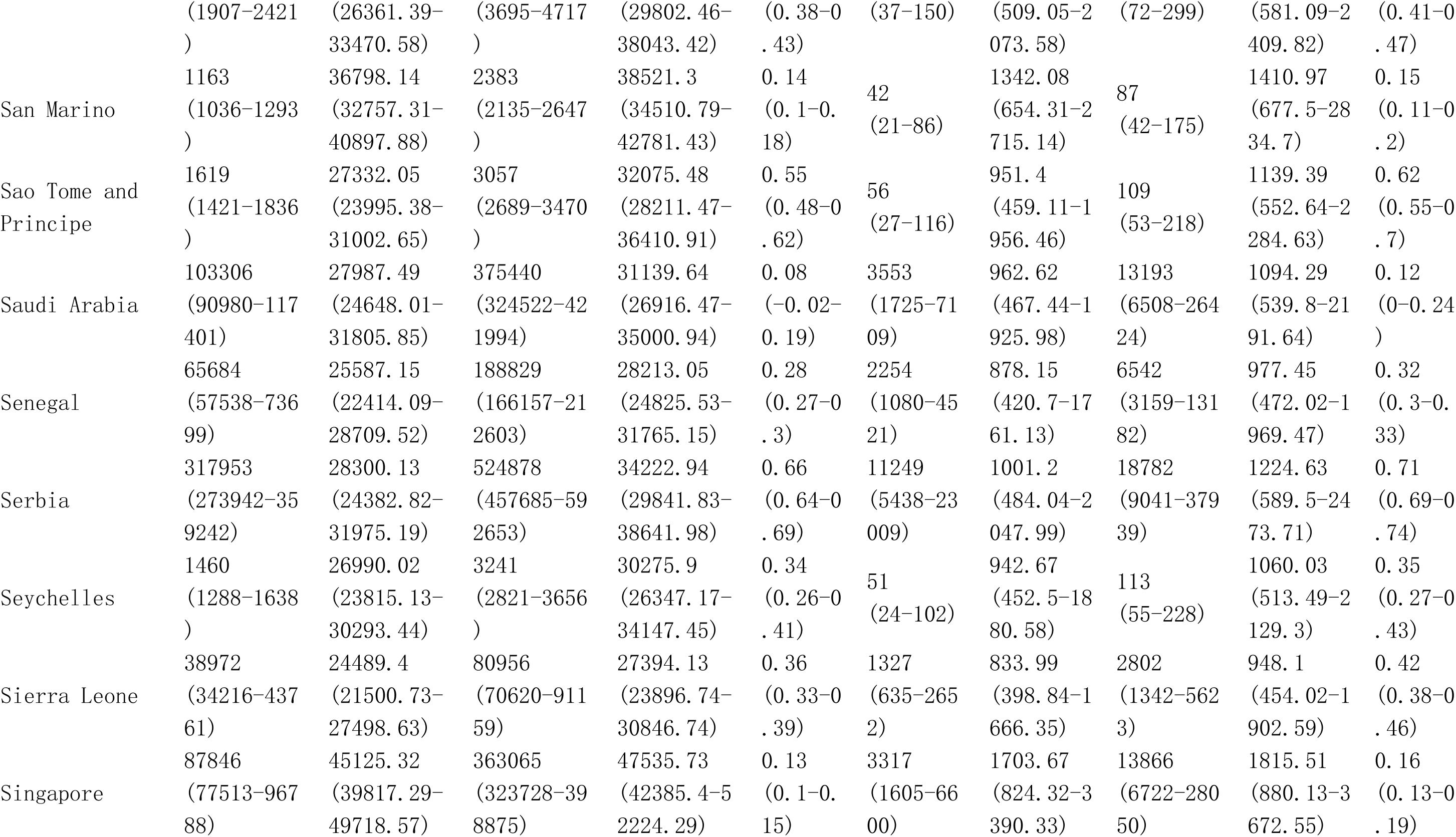
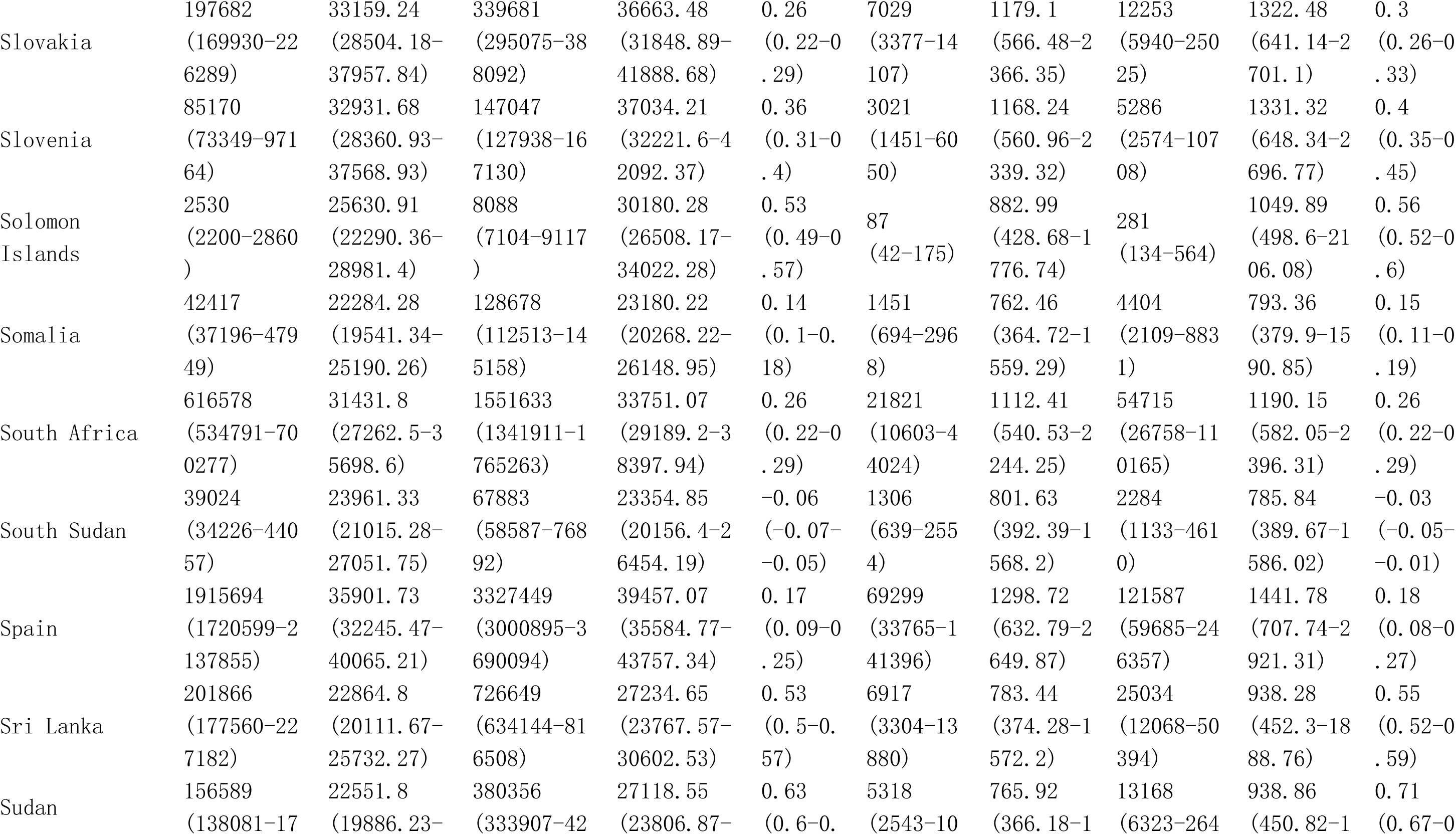
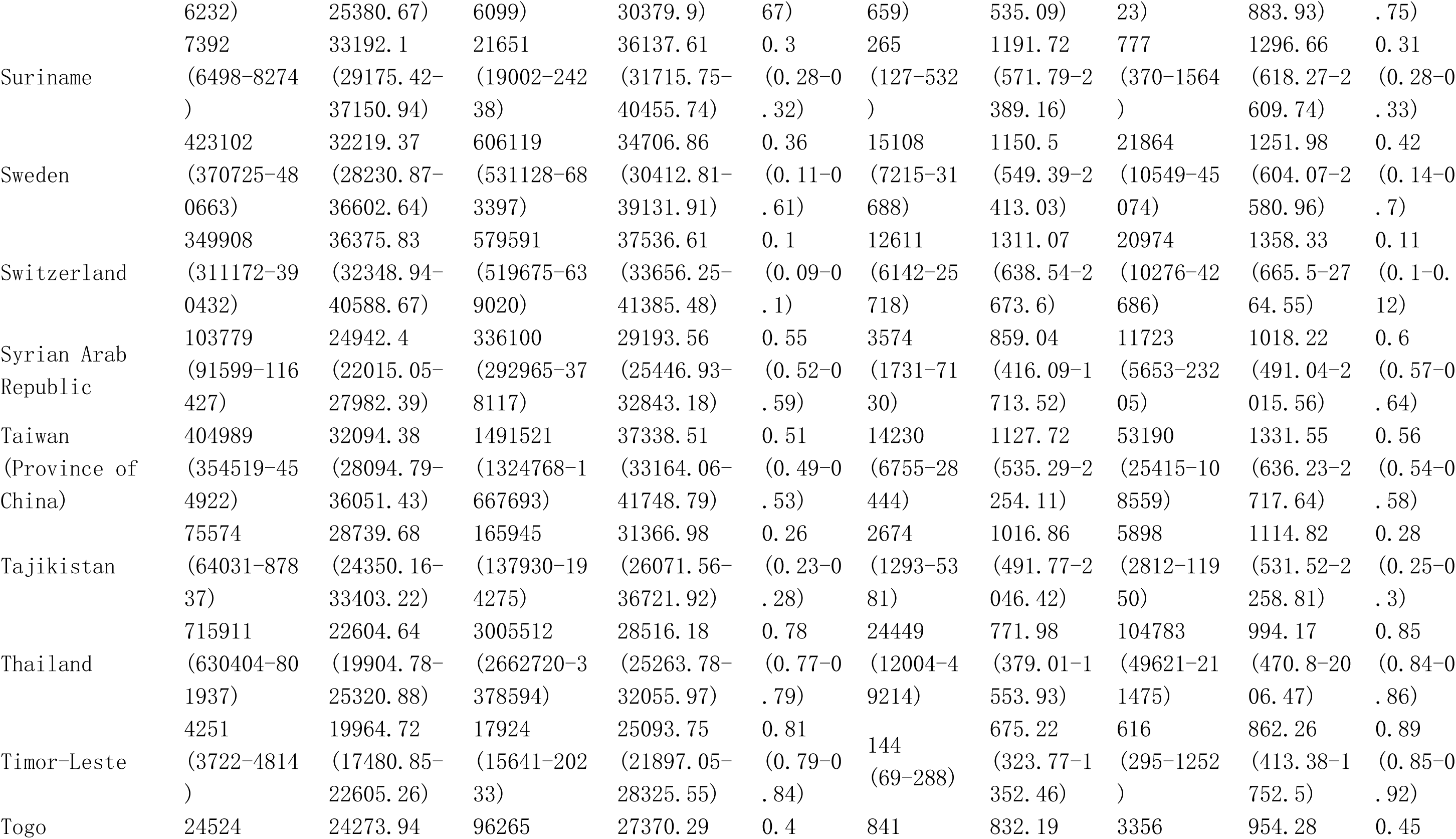
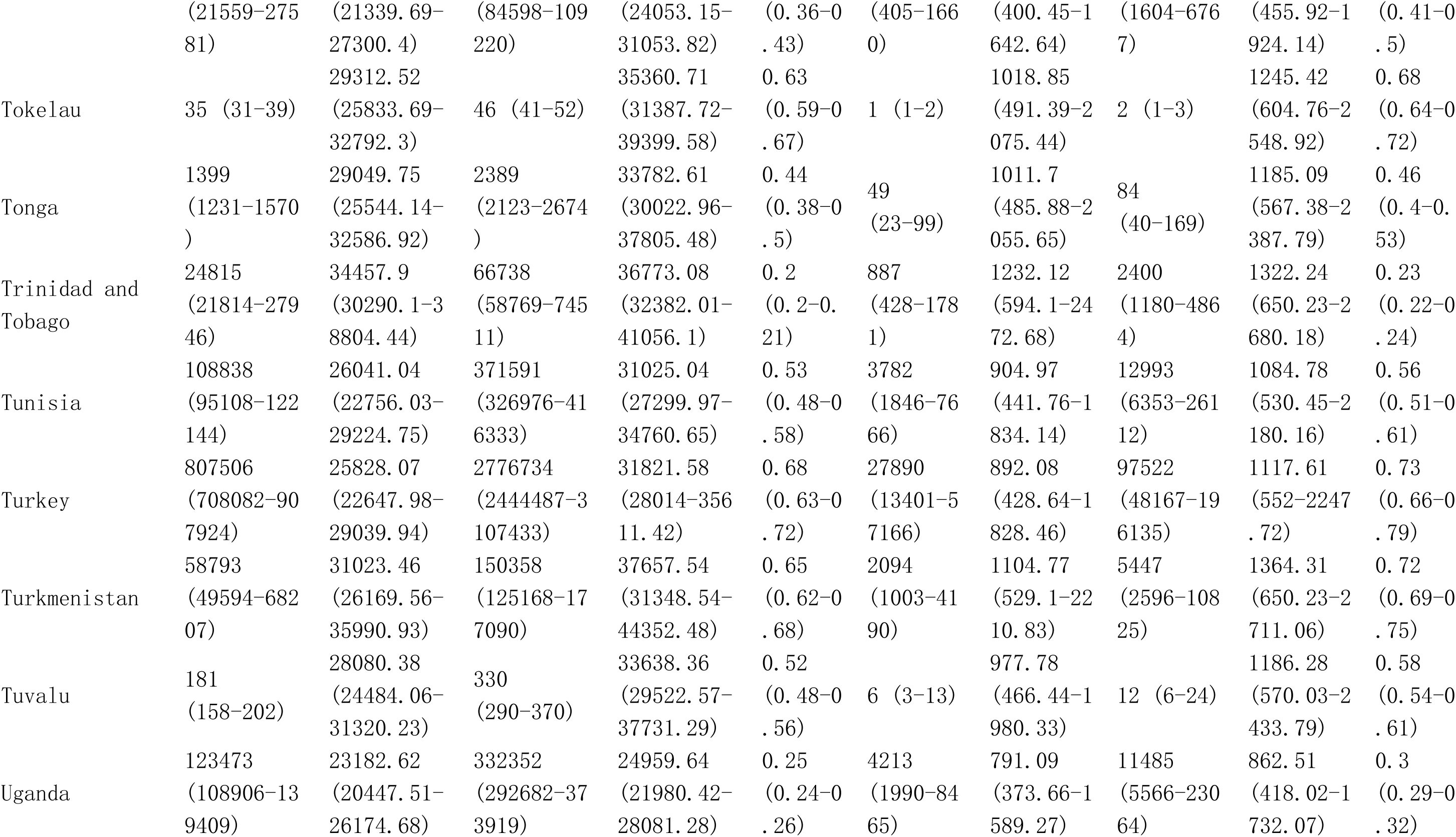
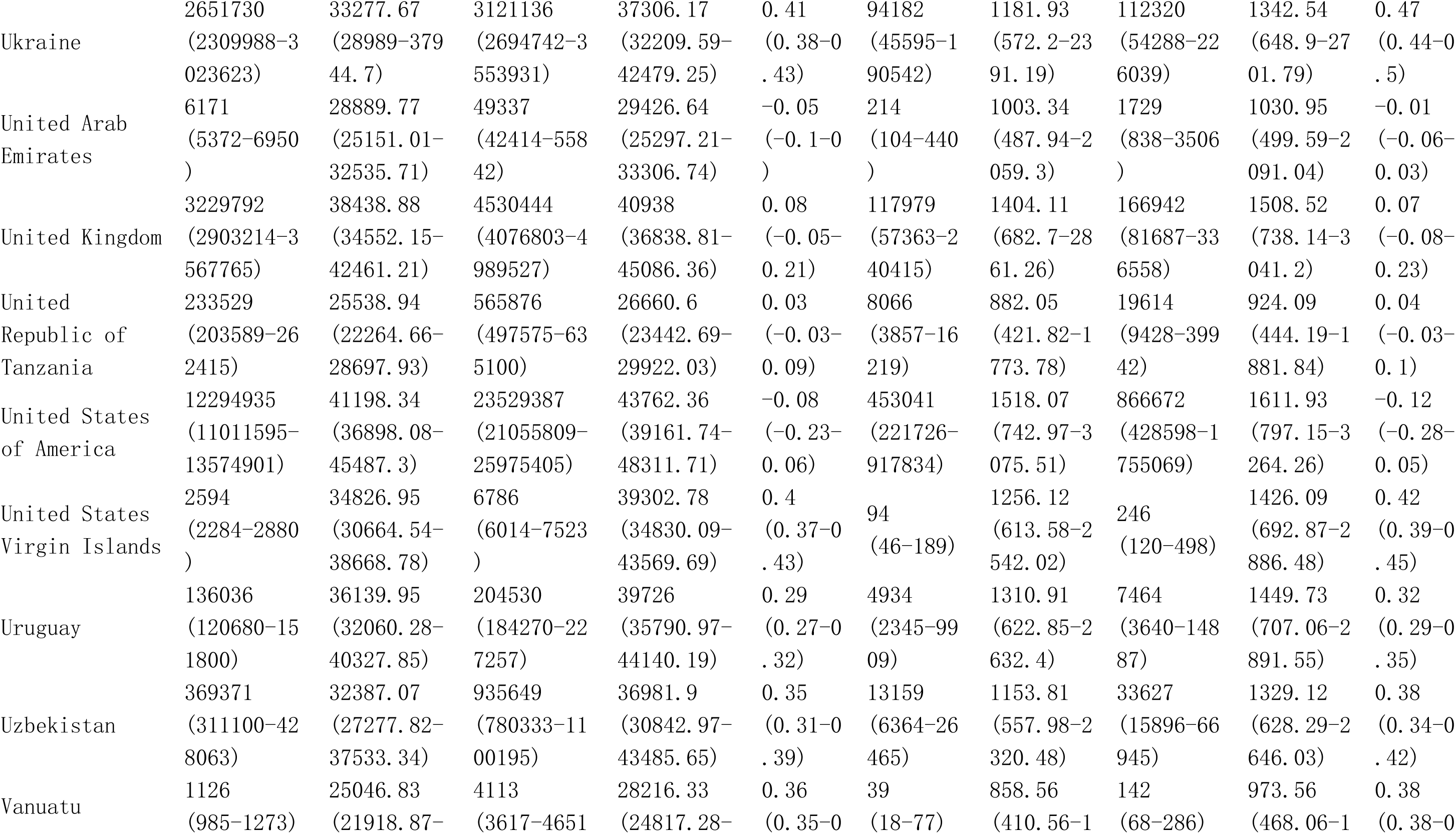
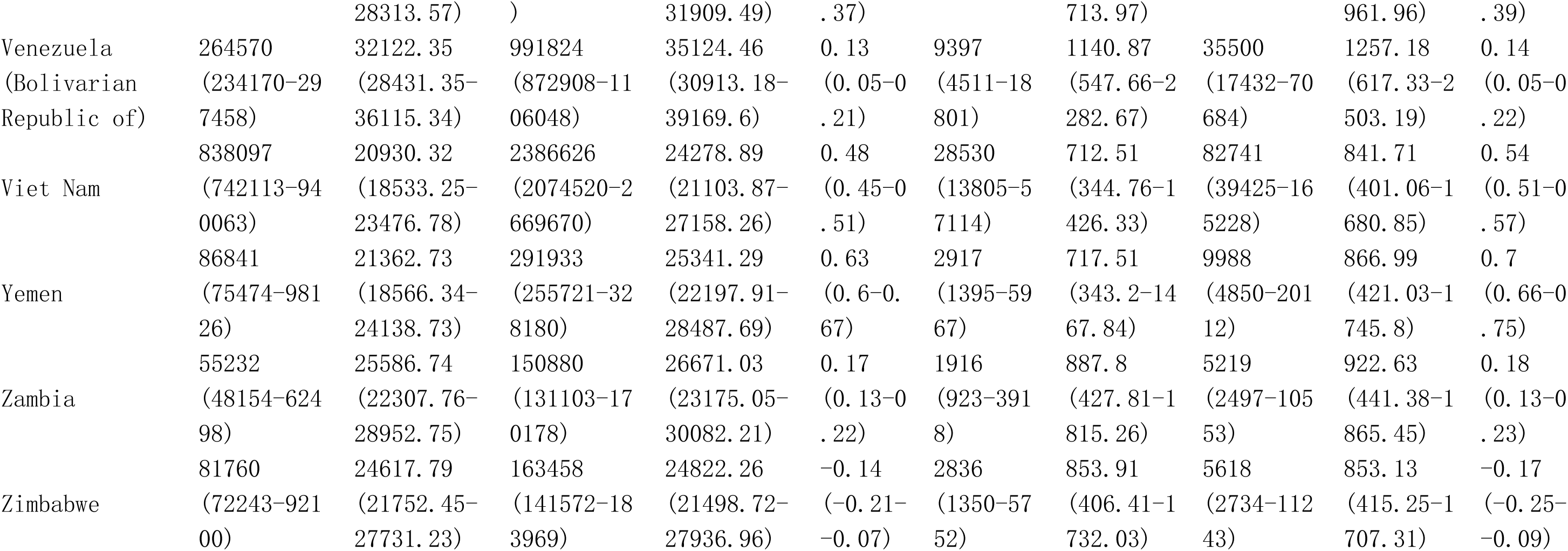
Global burden in Prevalence and DALYs of Osteoarthritis among postmenopausal women from 1990 to 2021 by 21 GBD geographical regions, and 204 countries and territories.

**Table S8.**
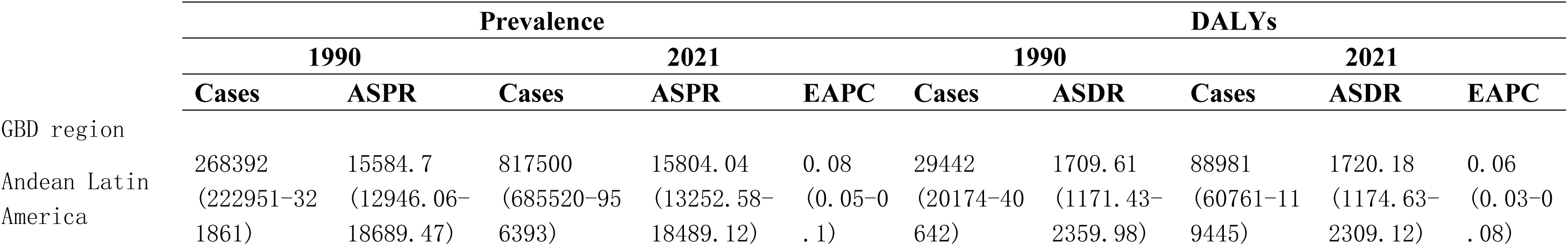
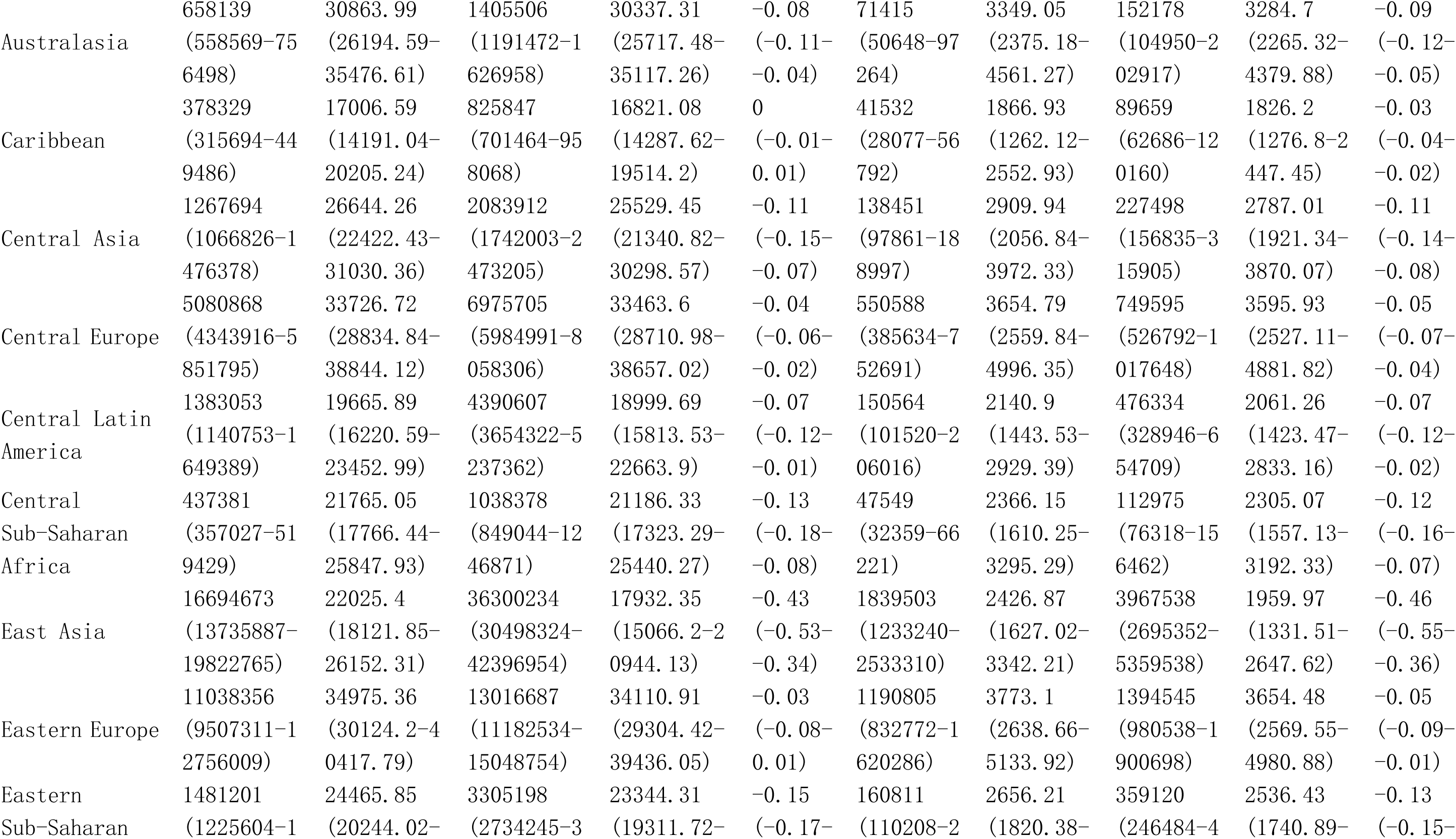
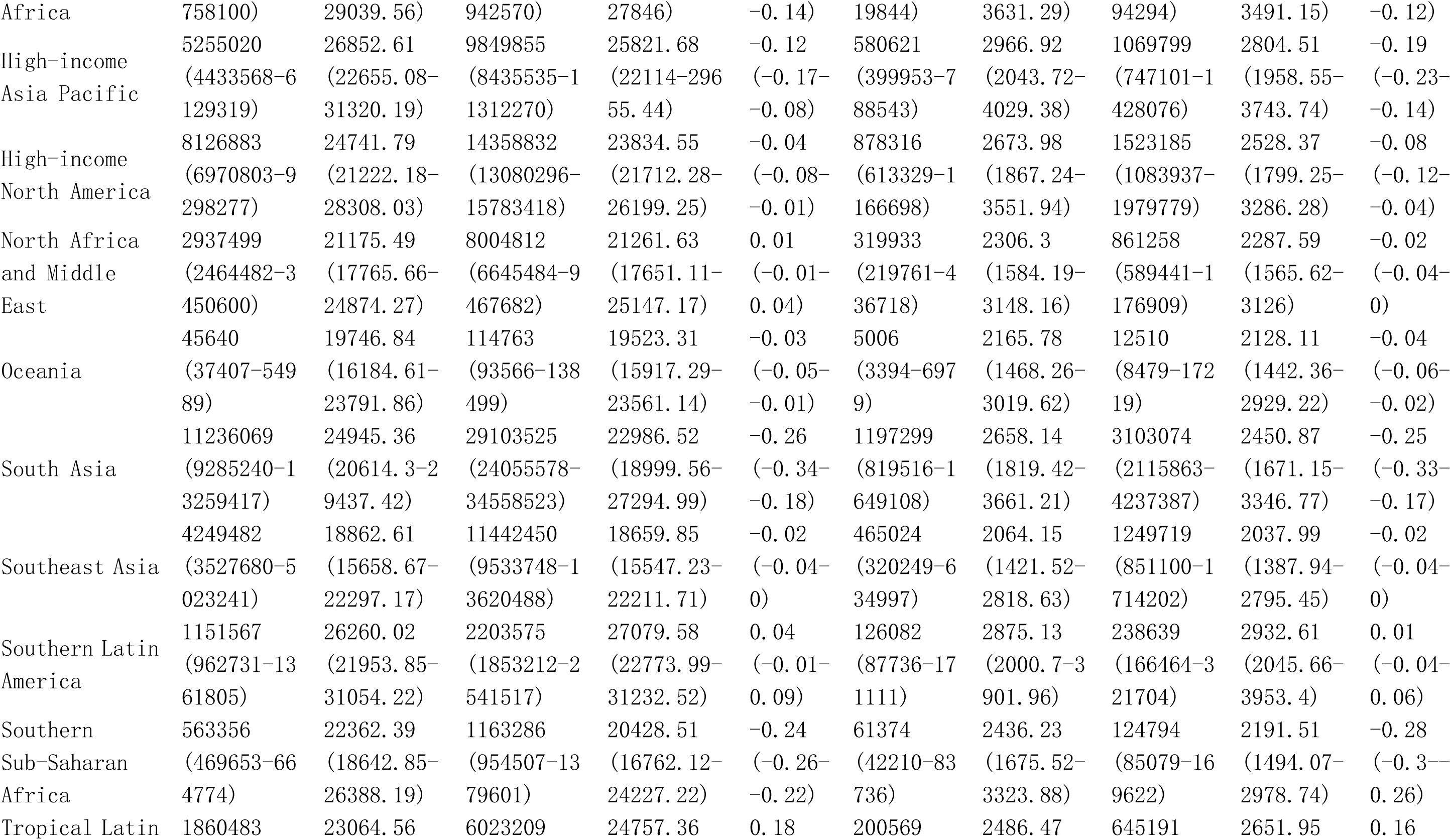
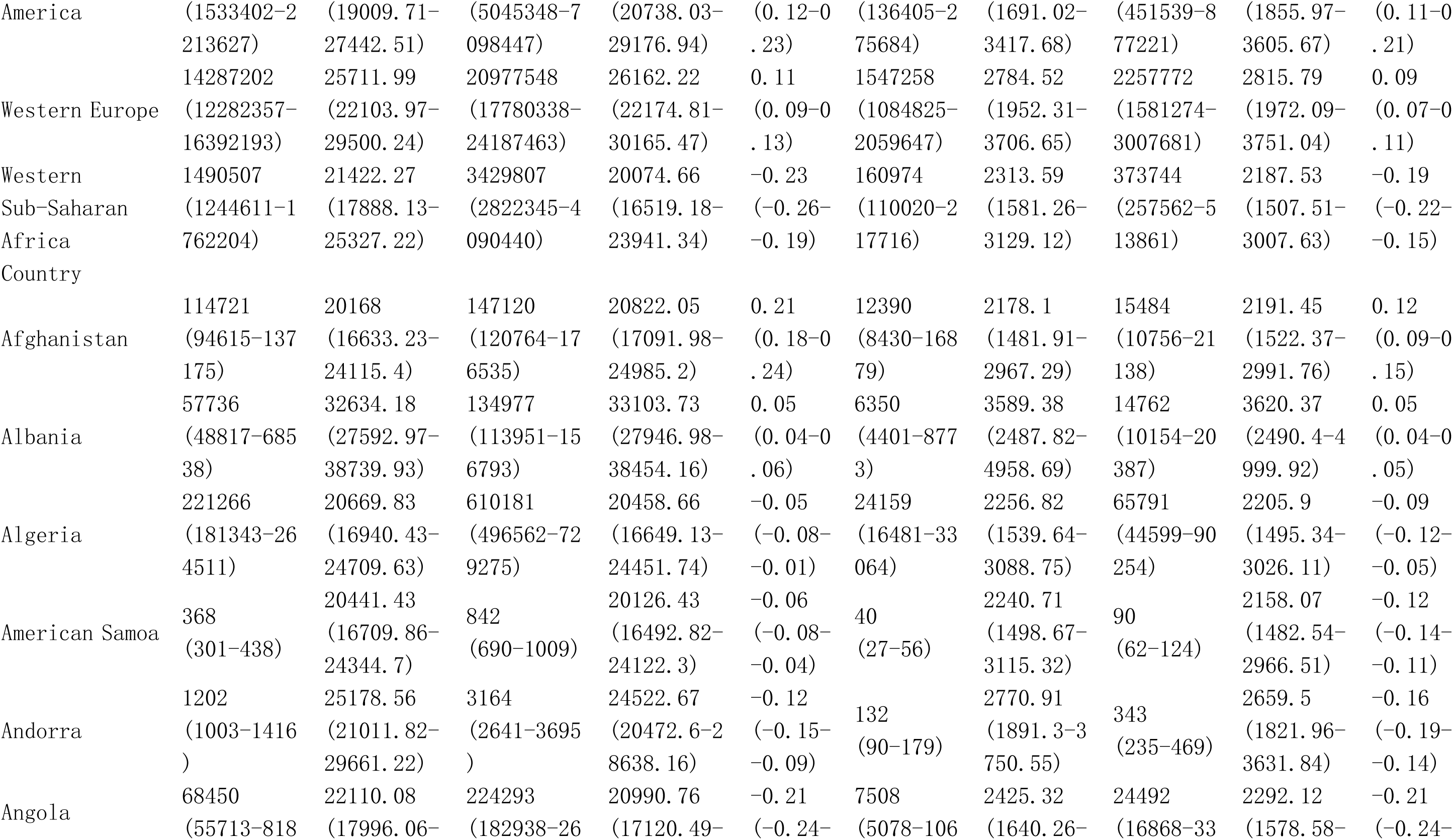
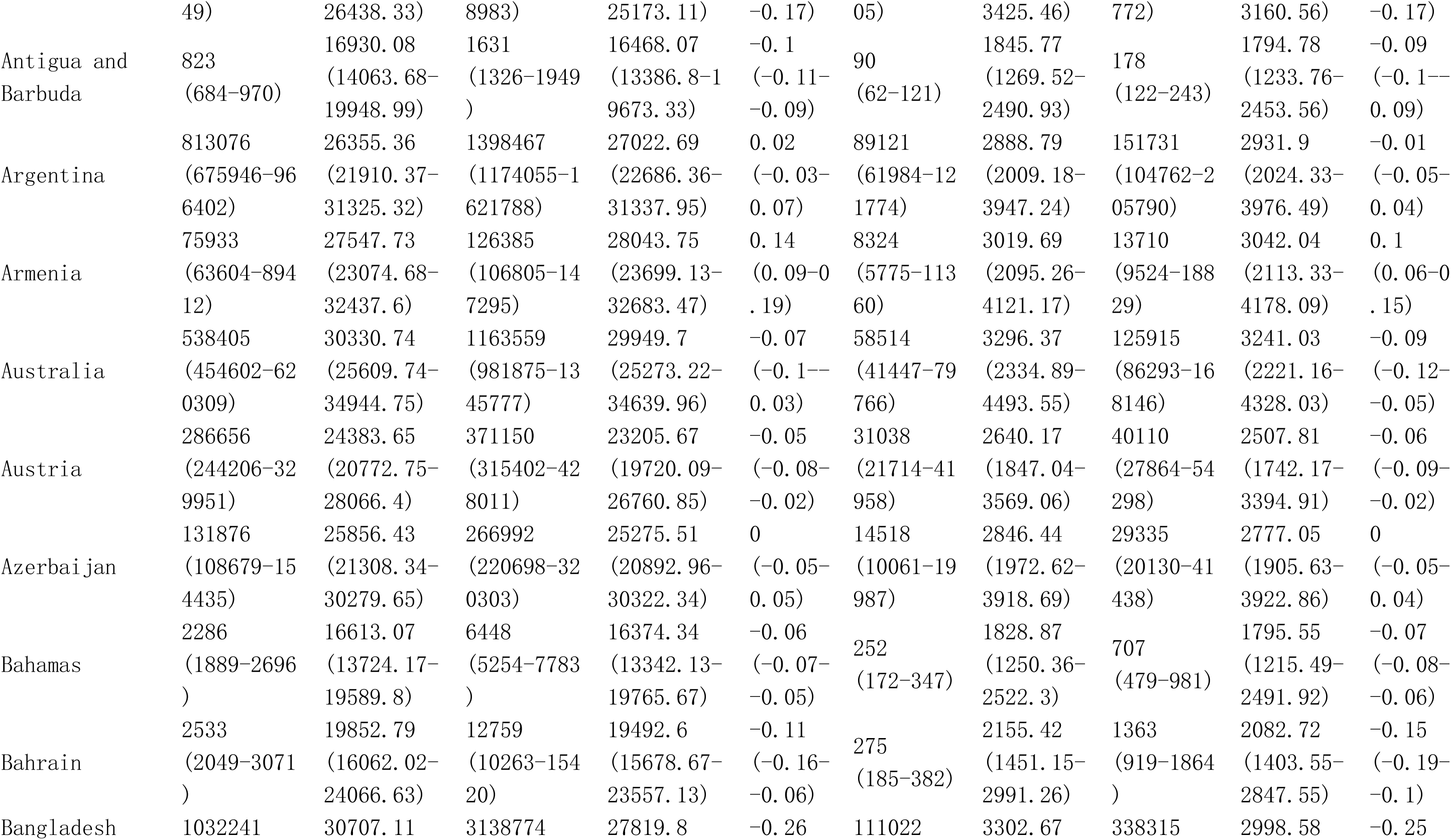
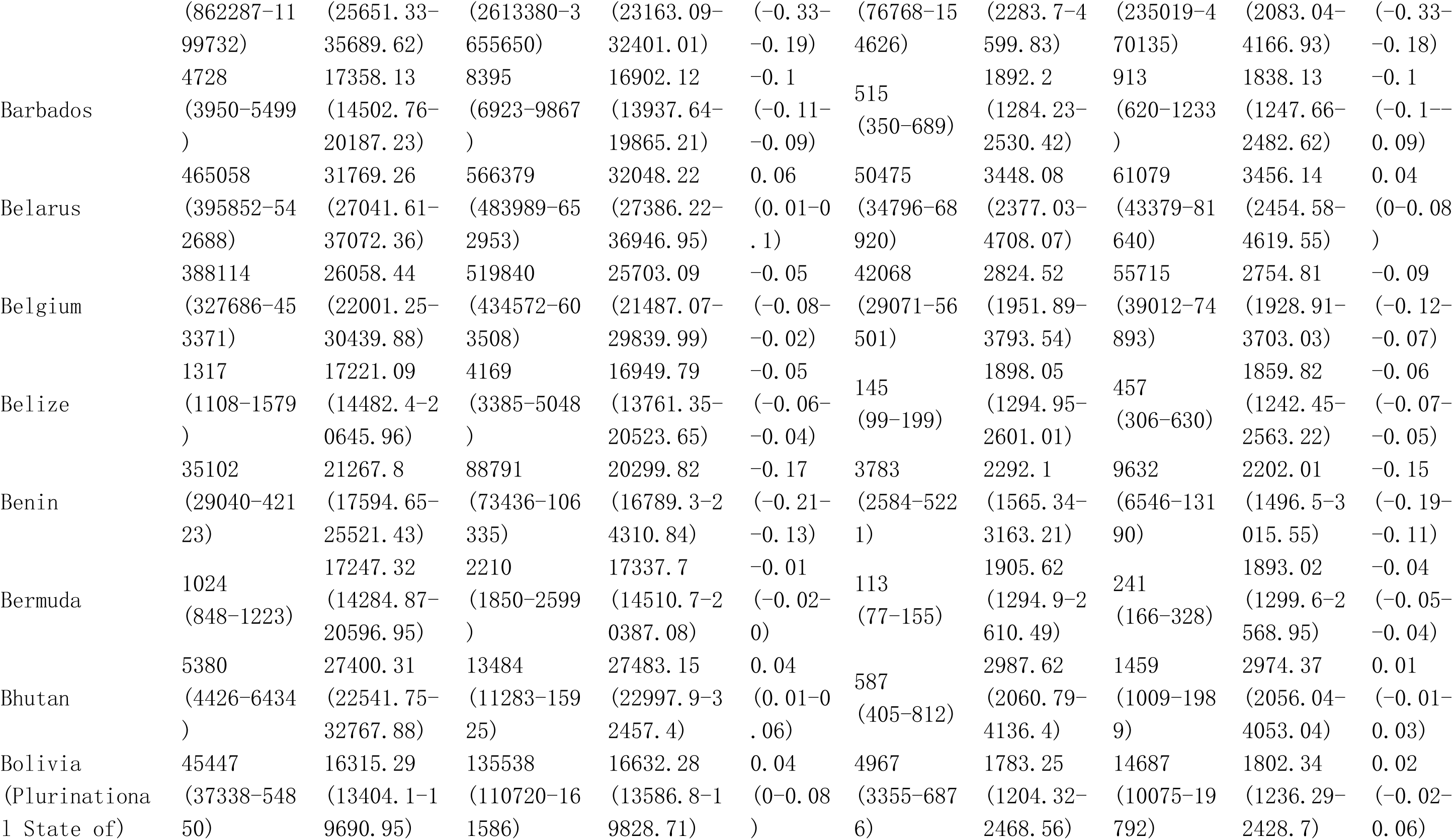
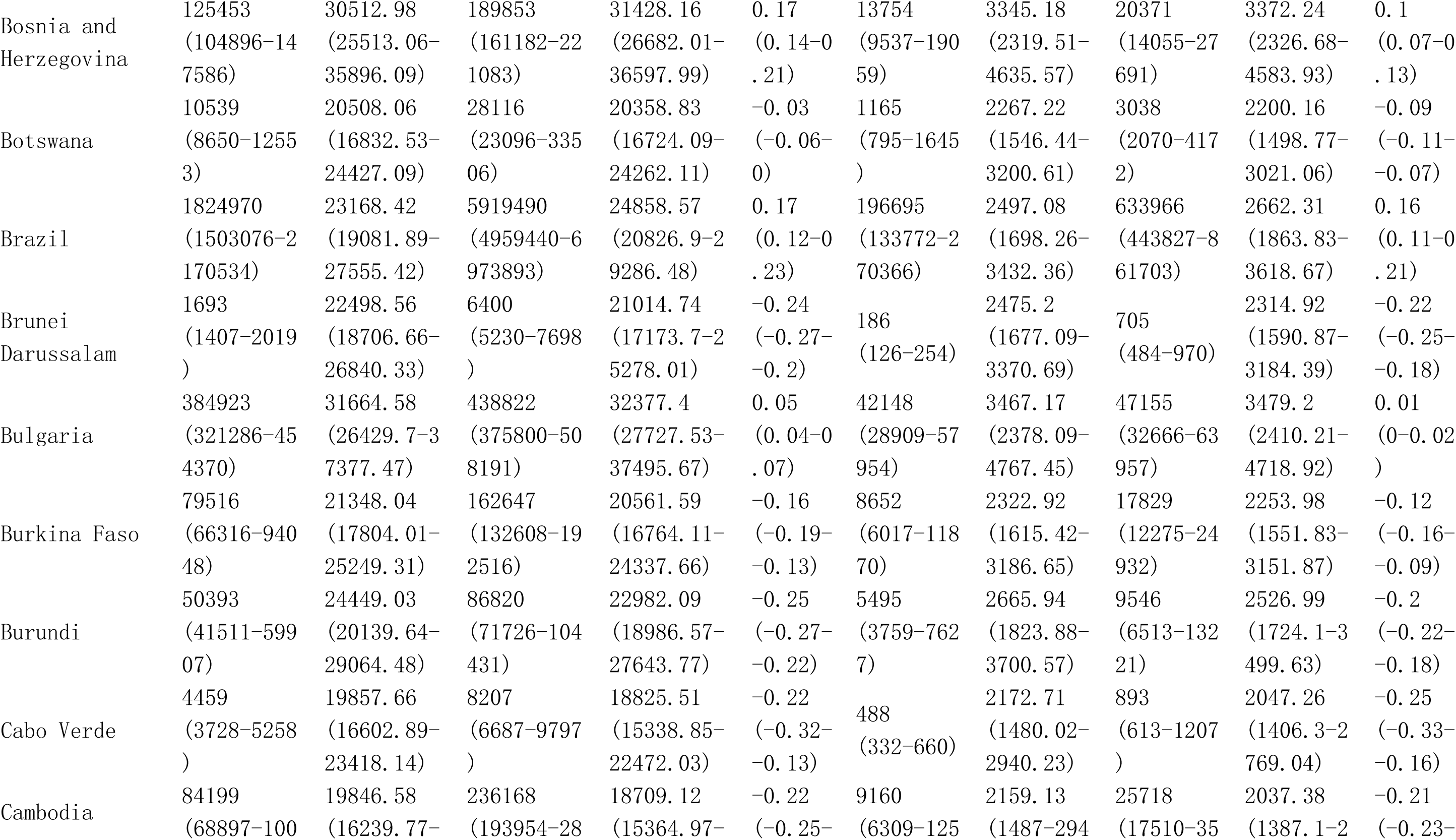
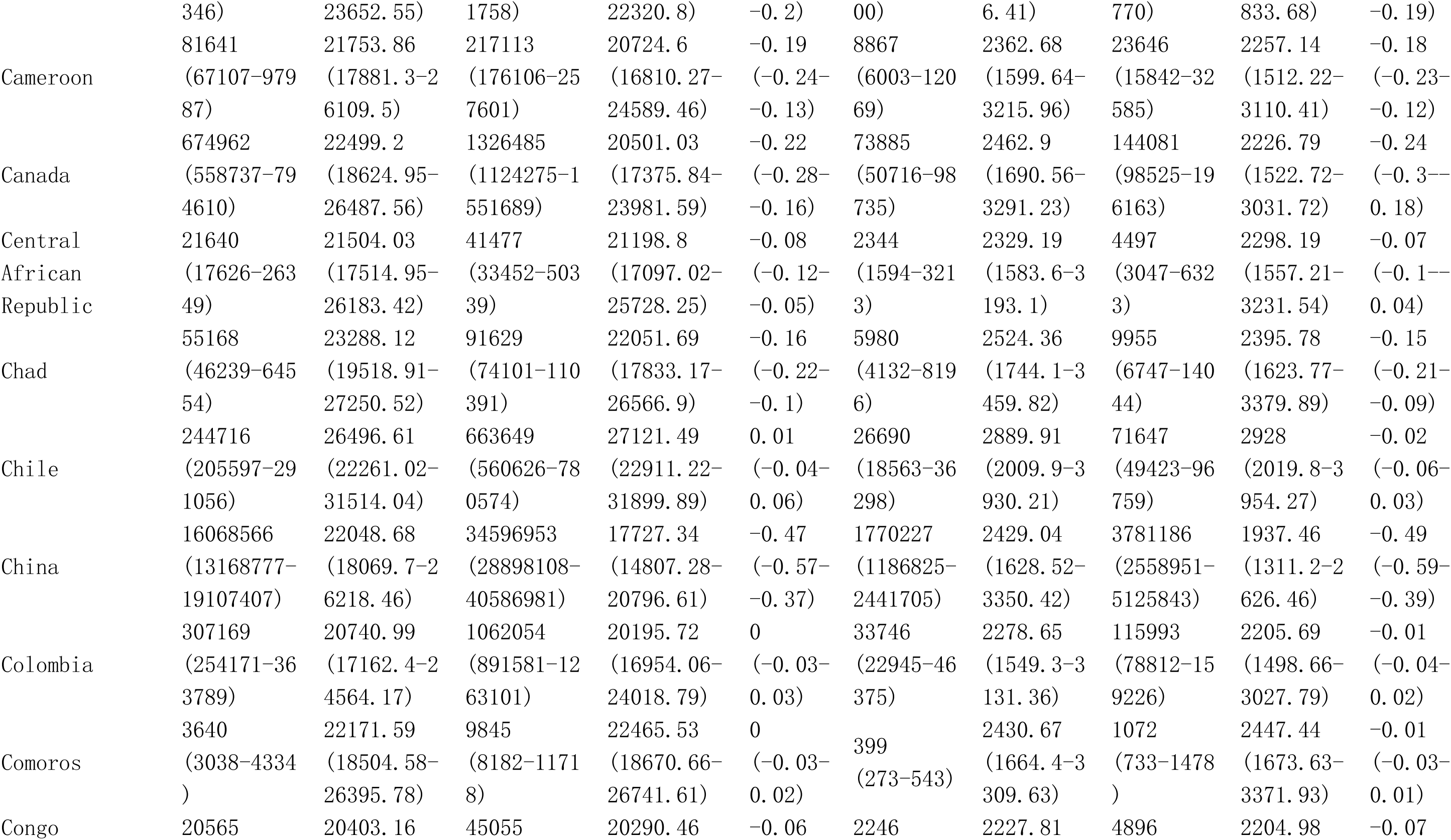
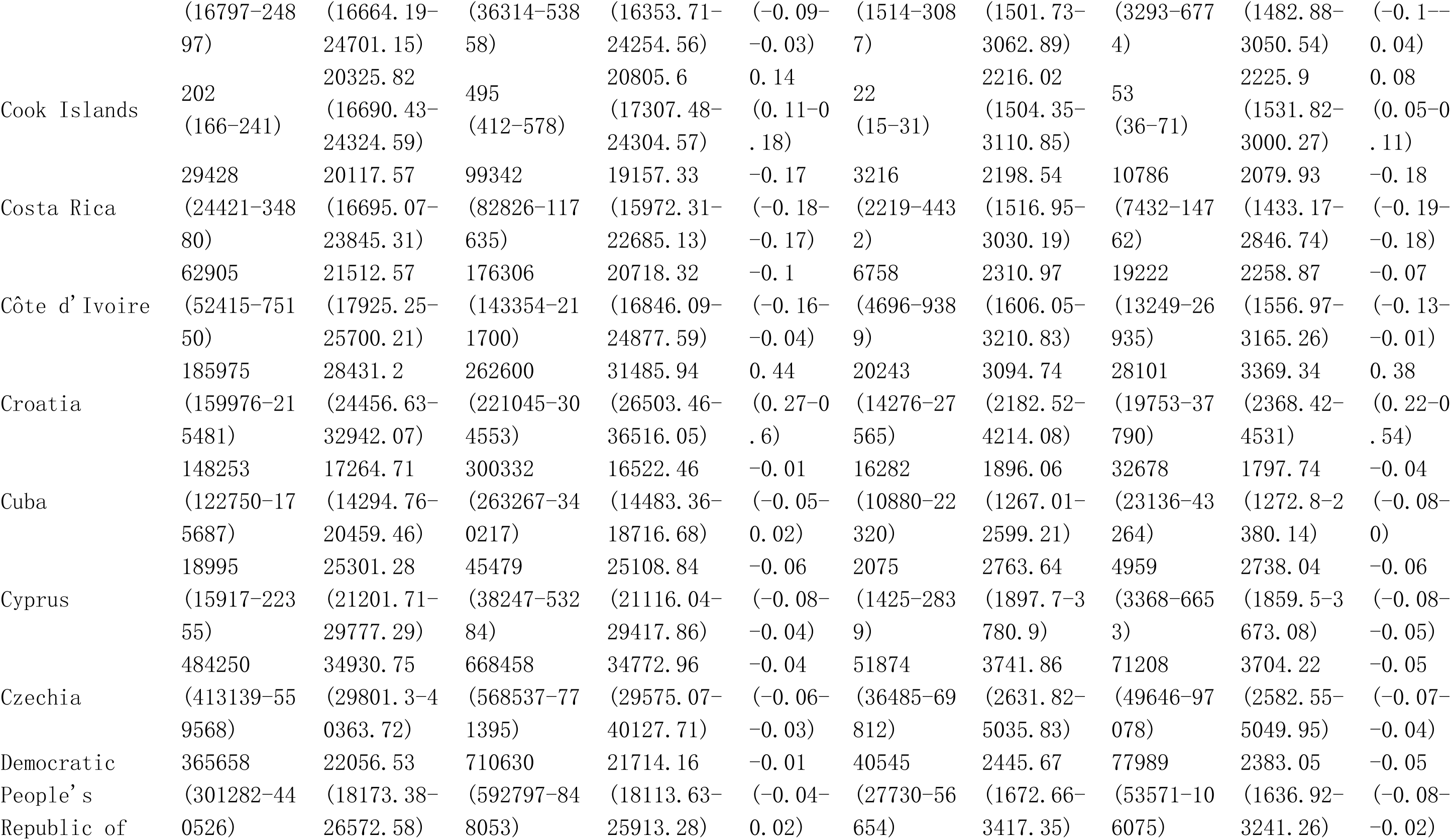
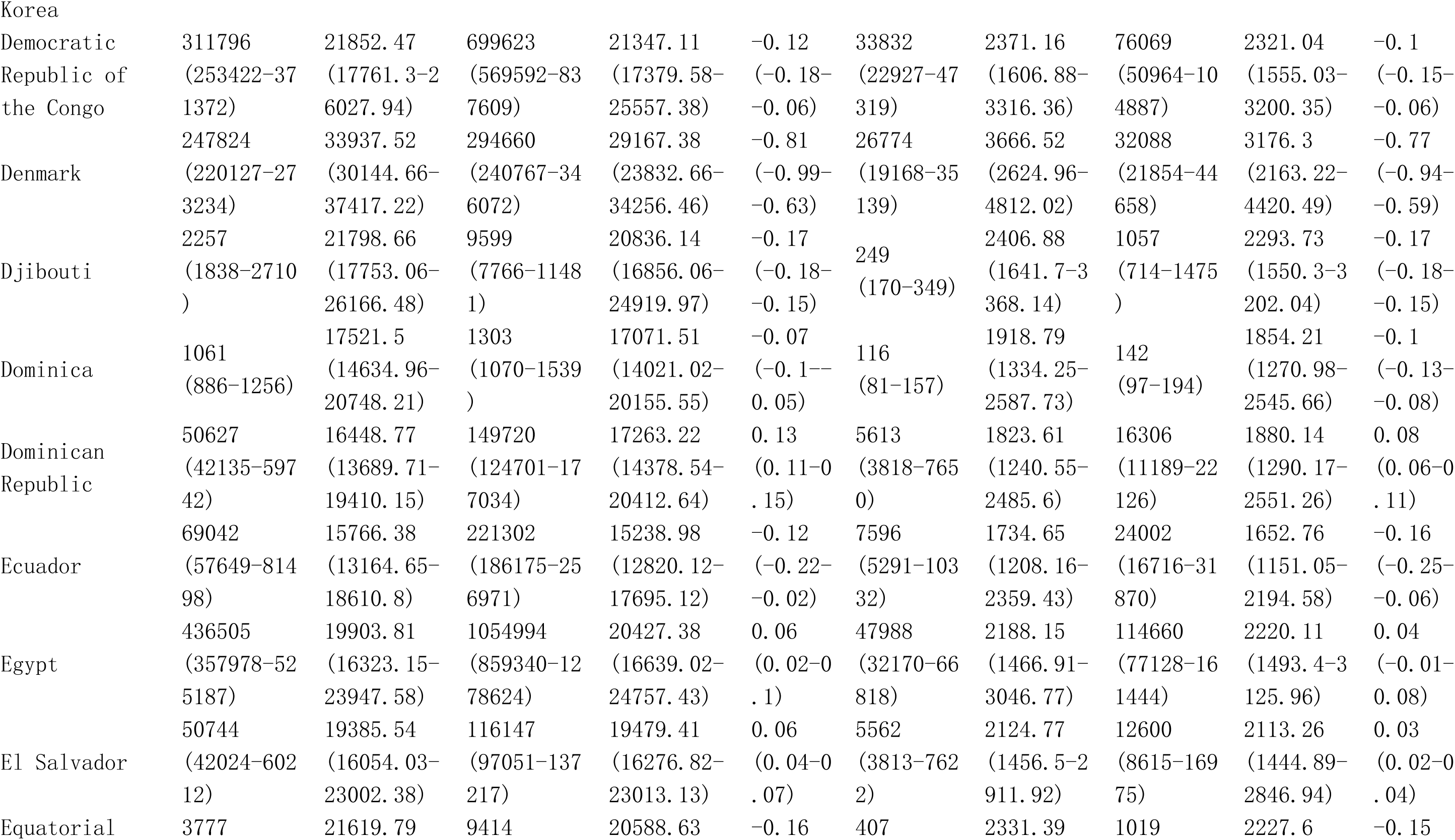
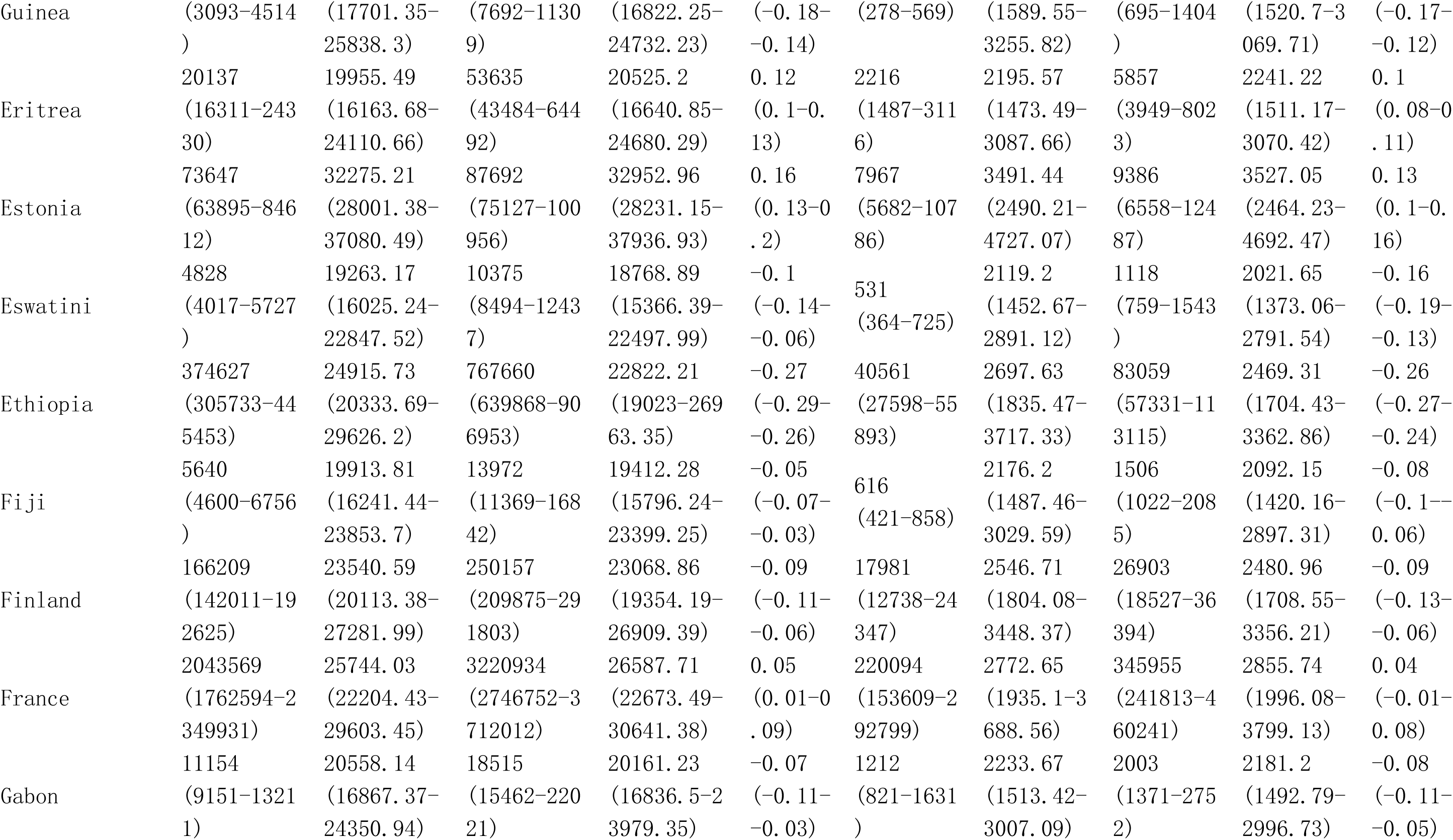
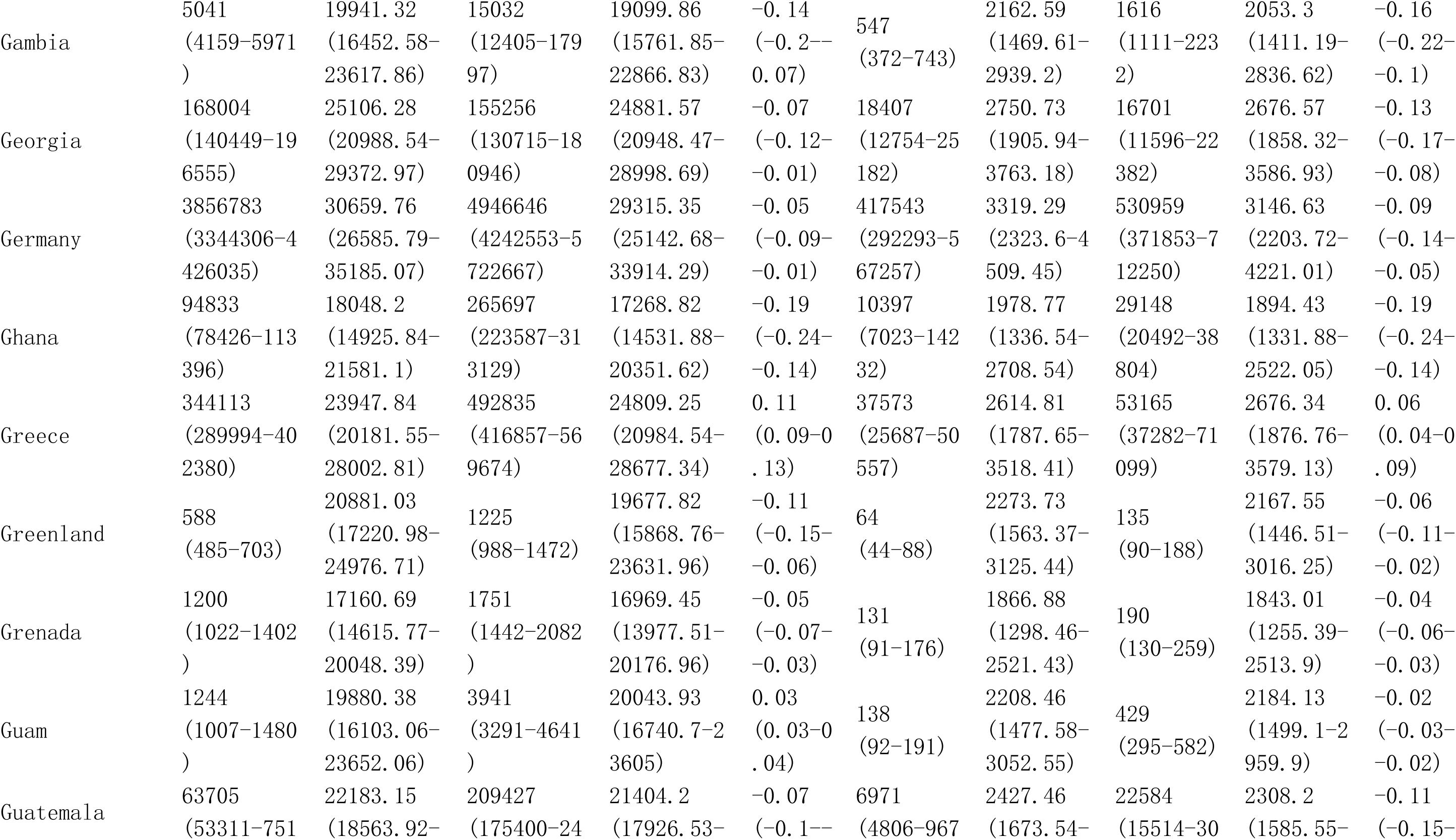
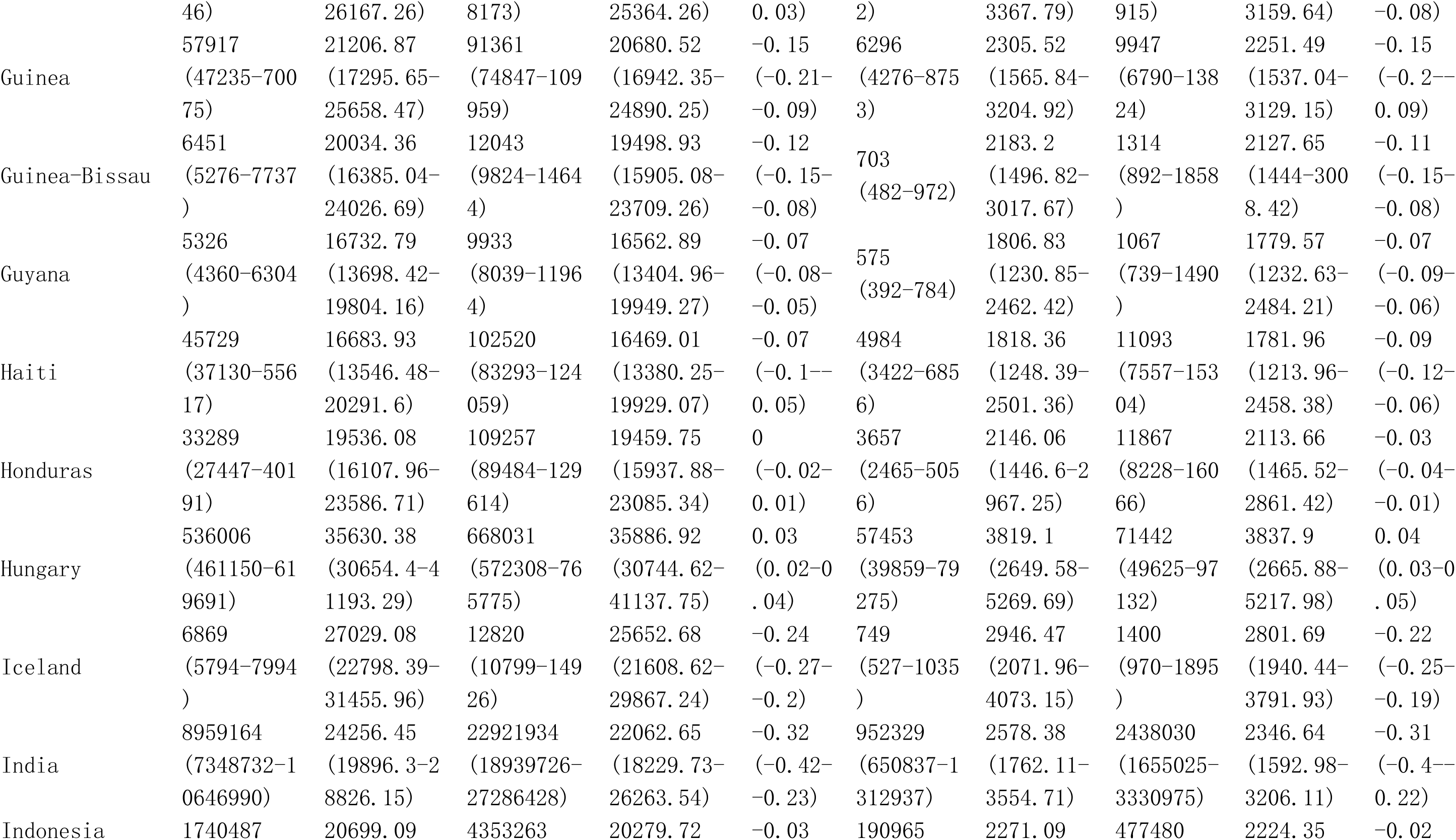
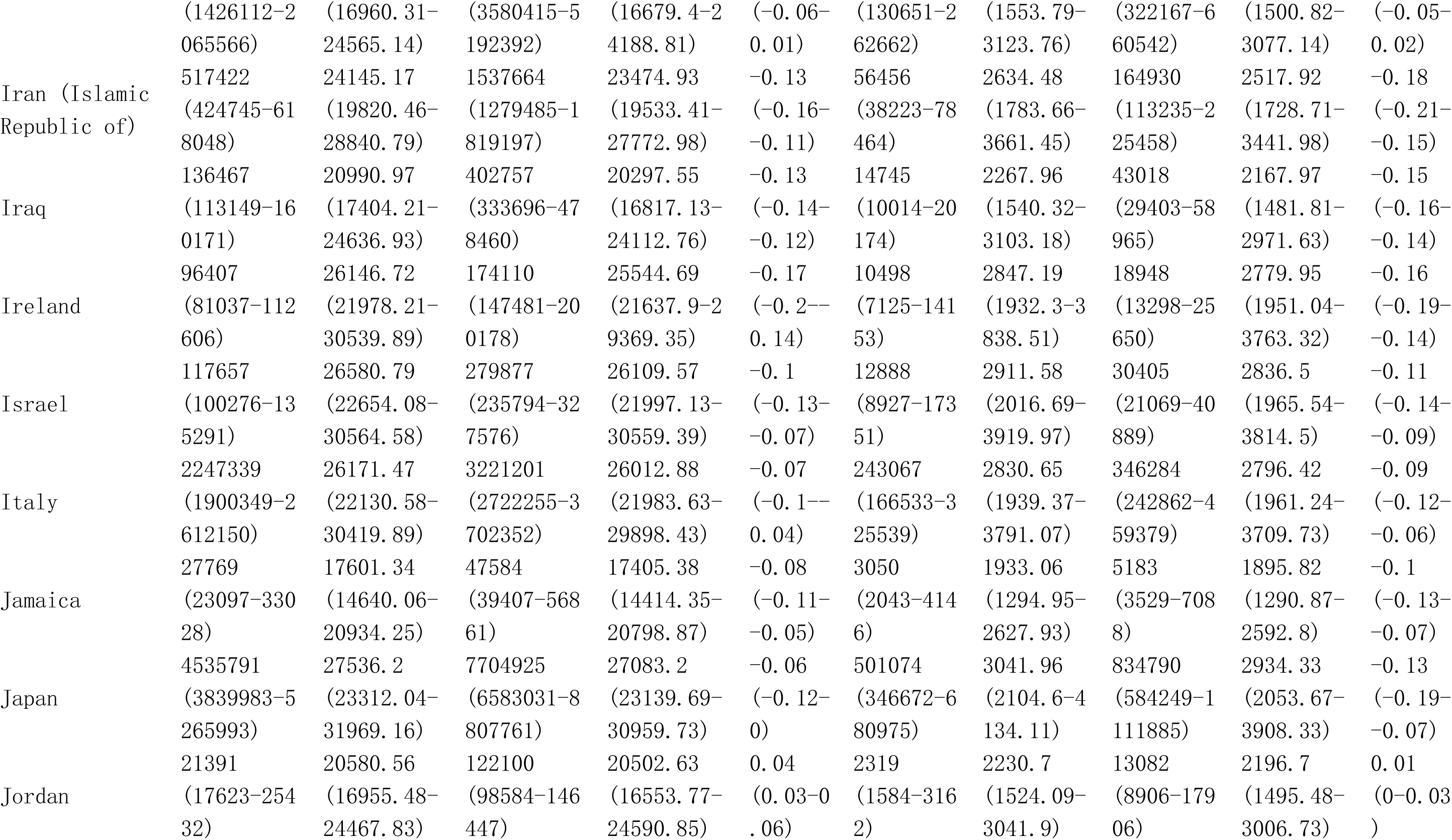
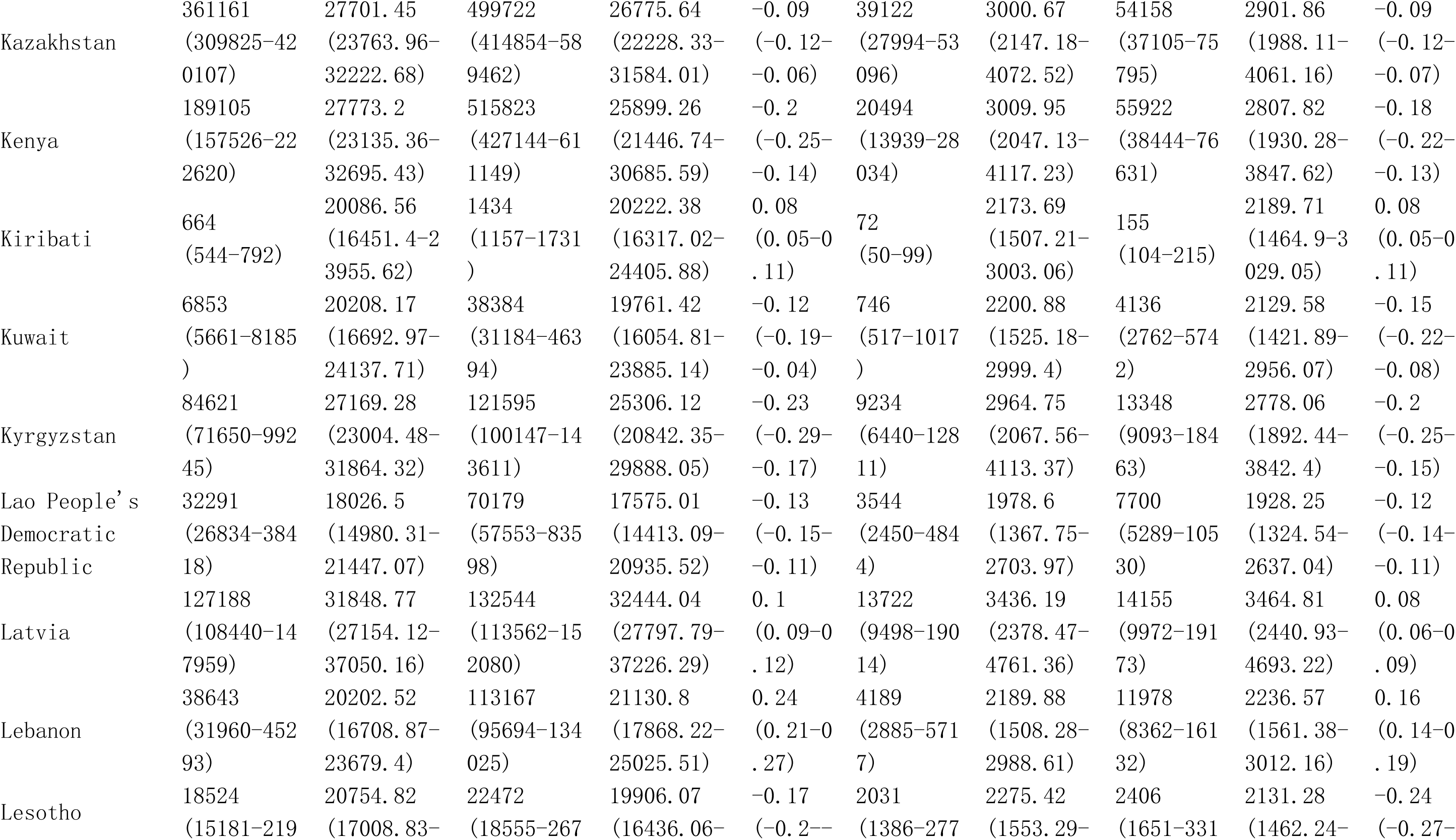
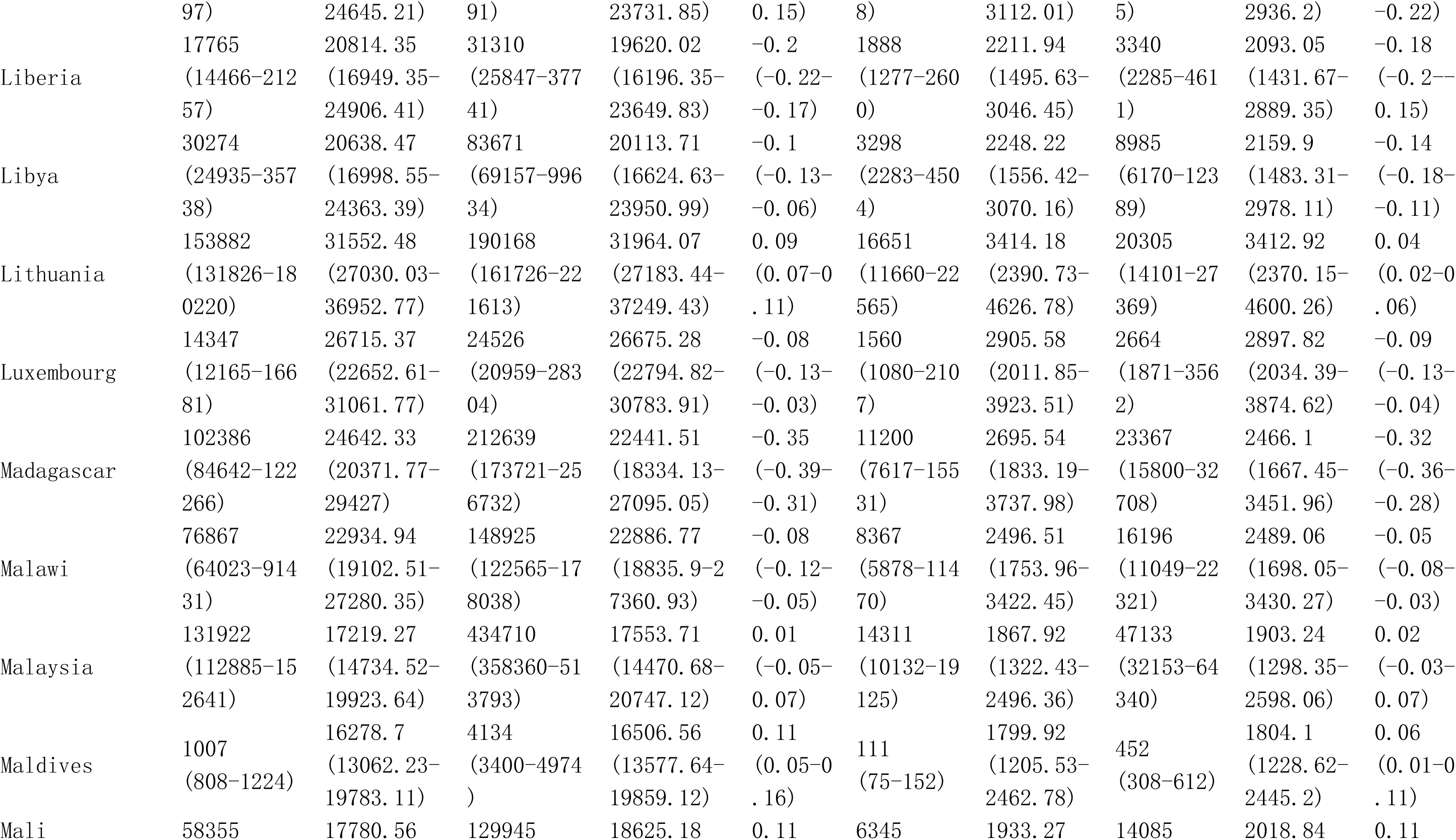
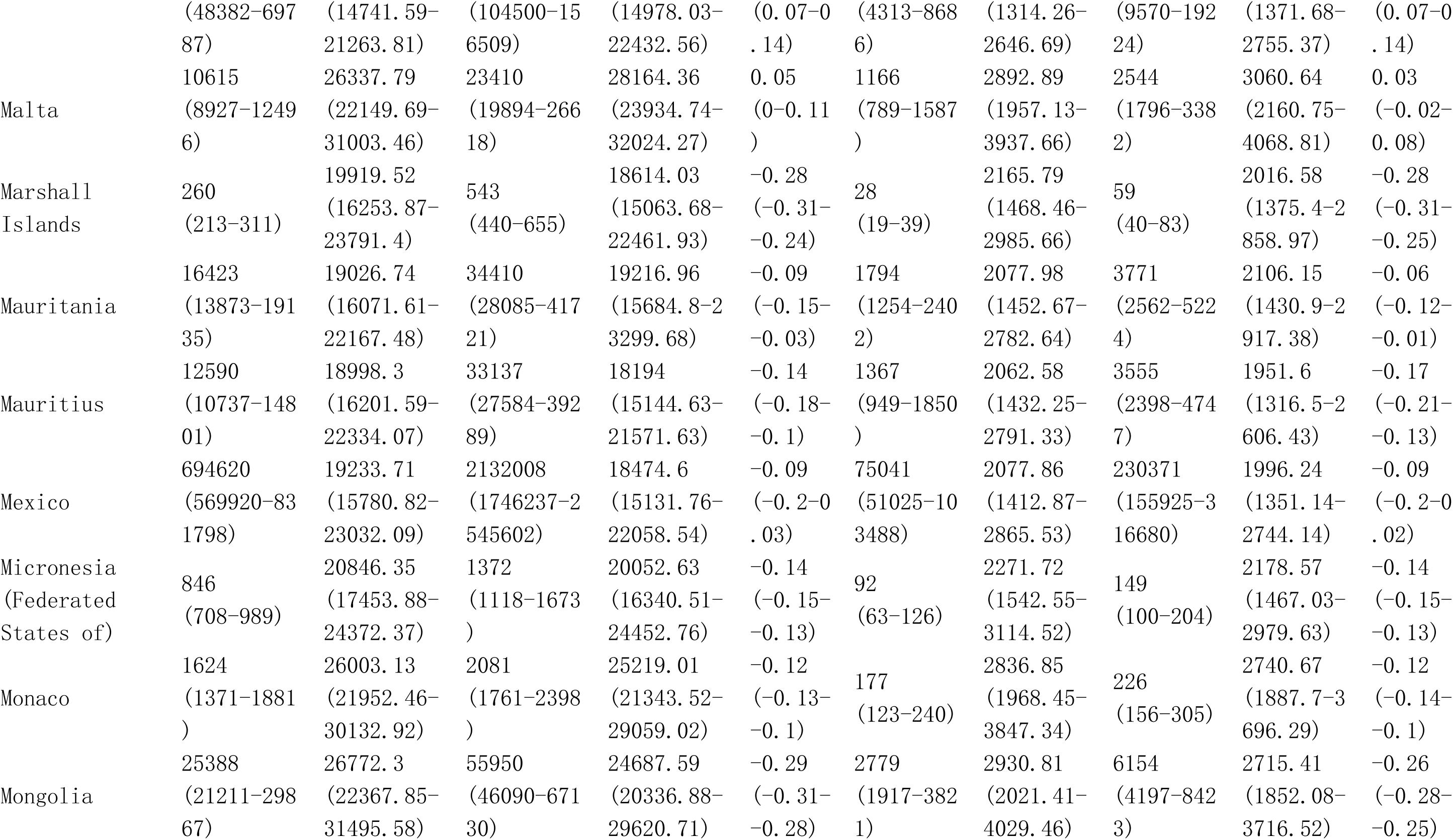
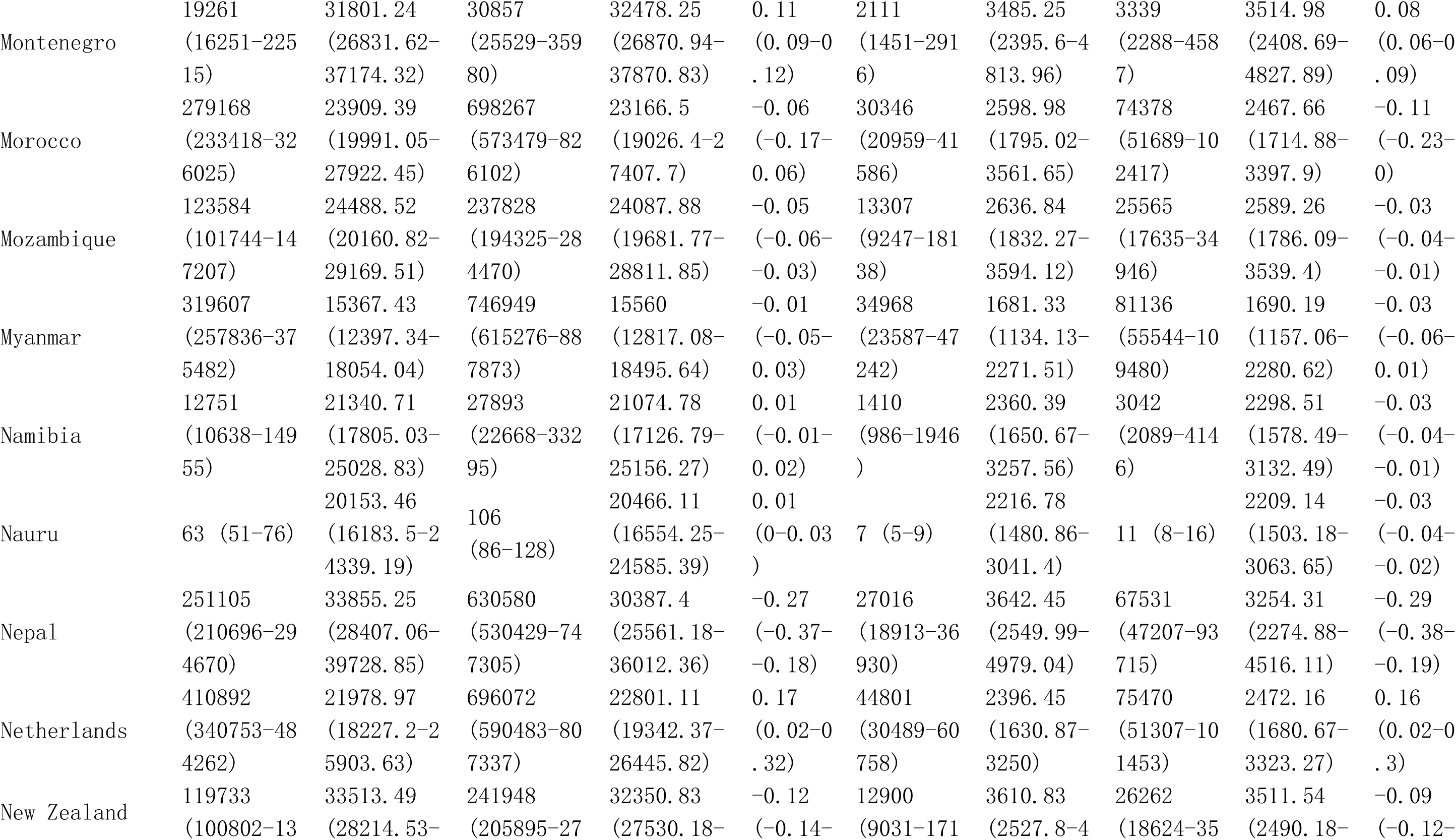
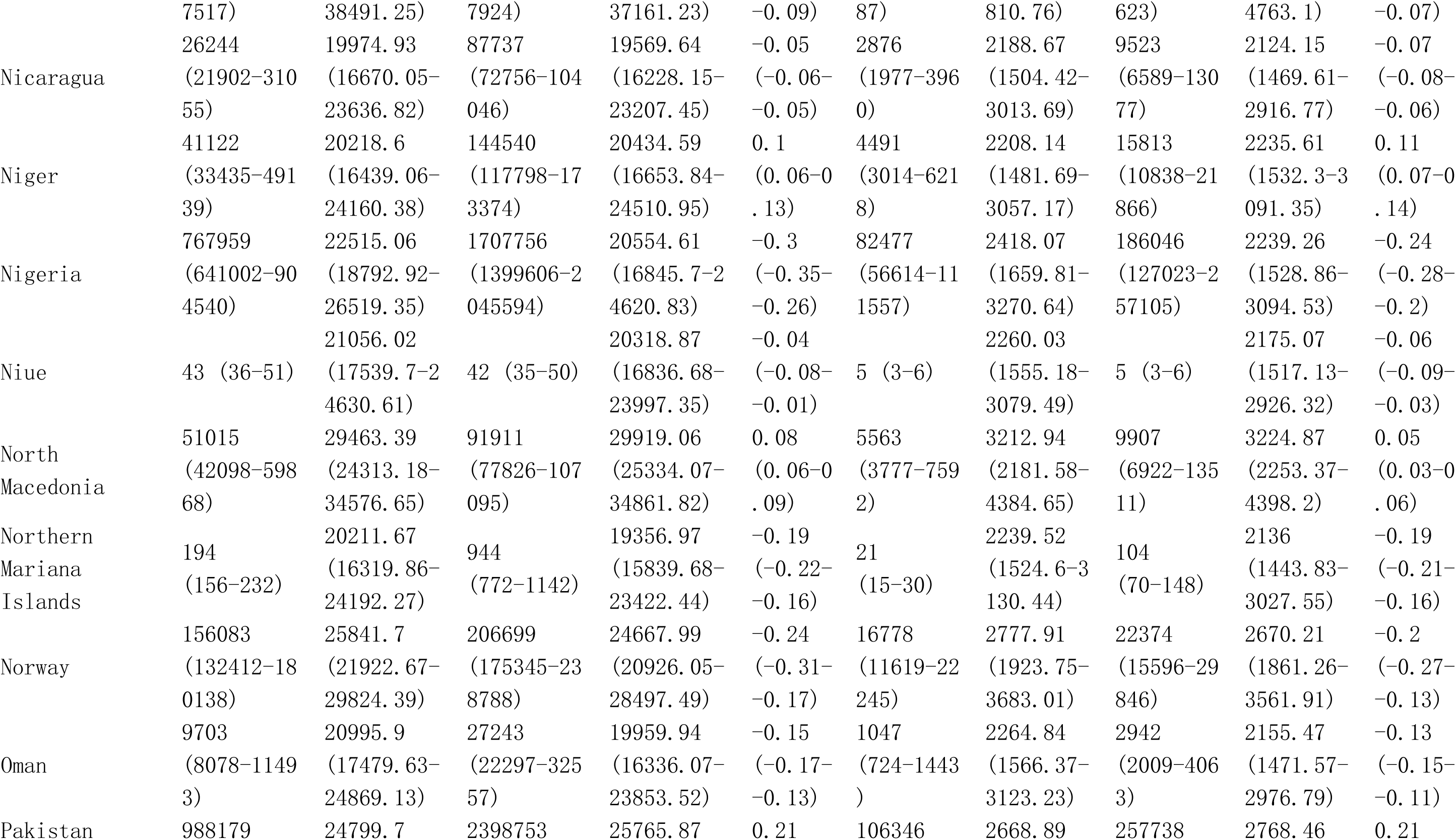
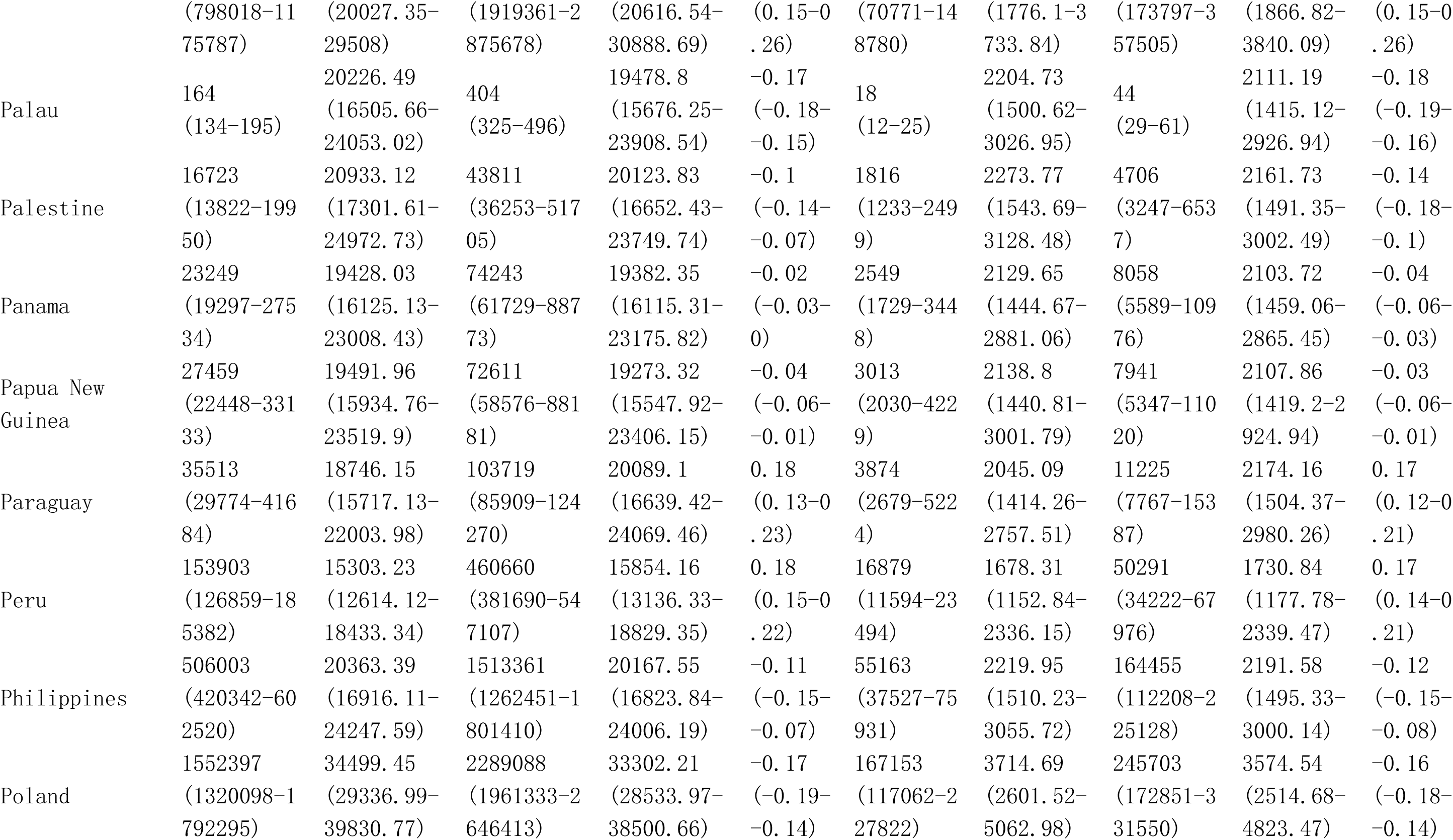
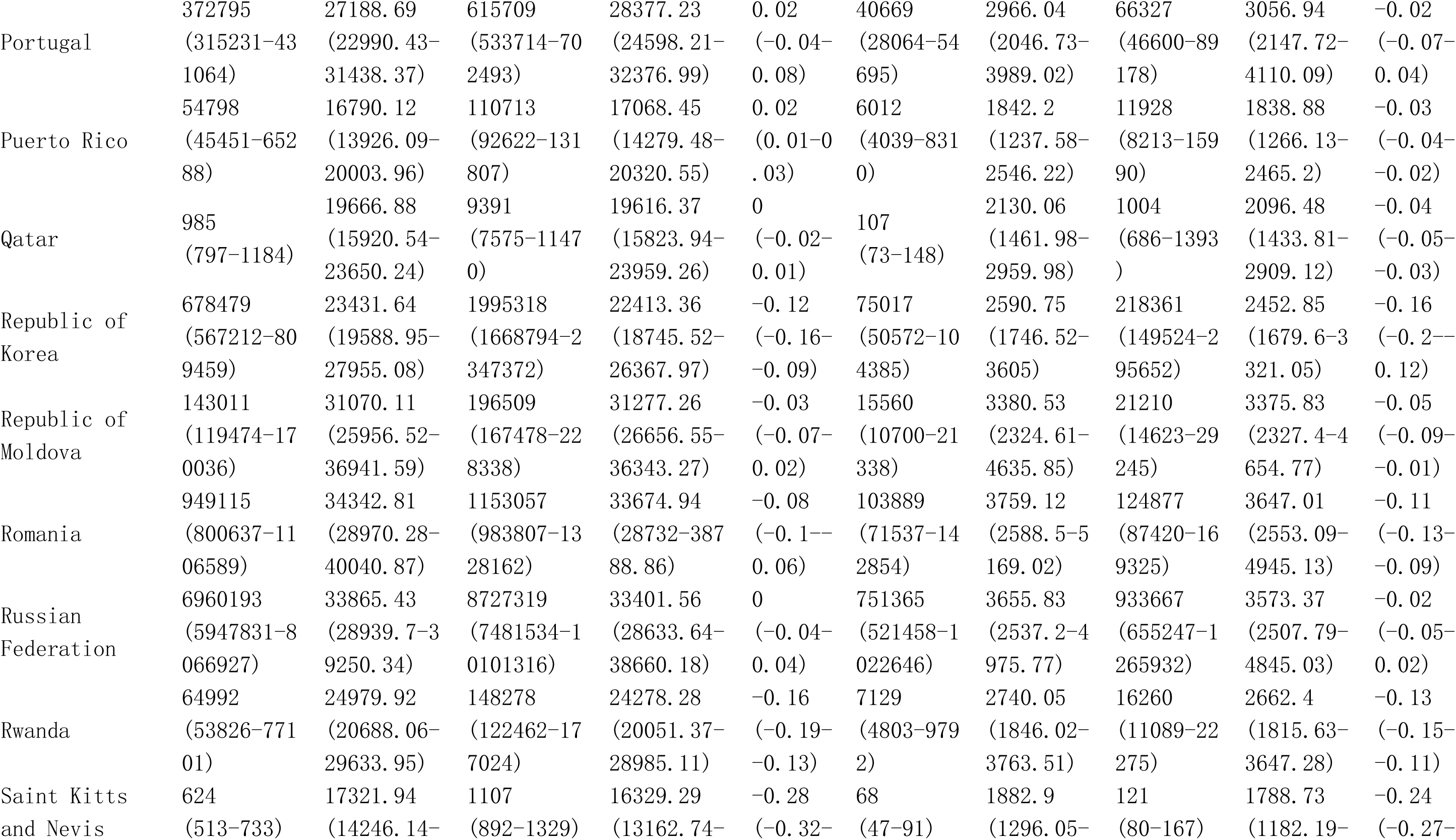
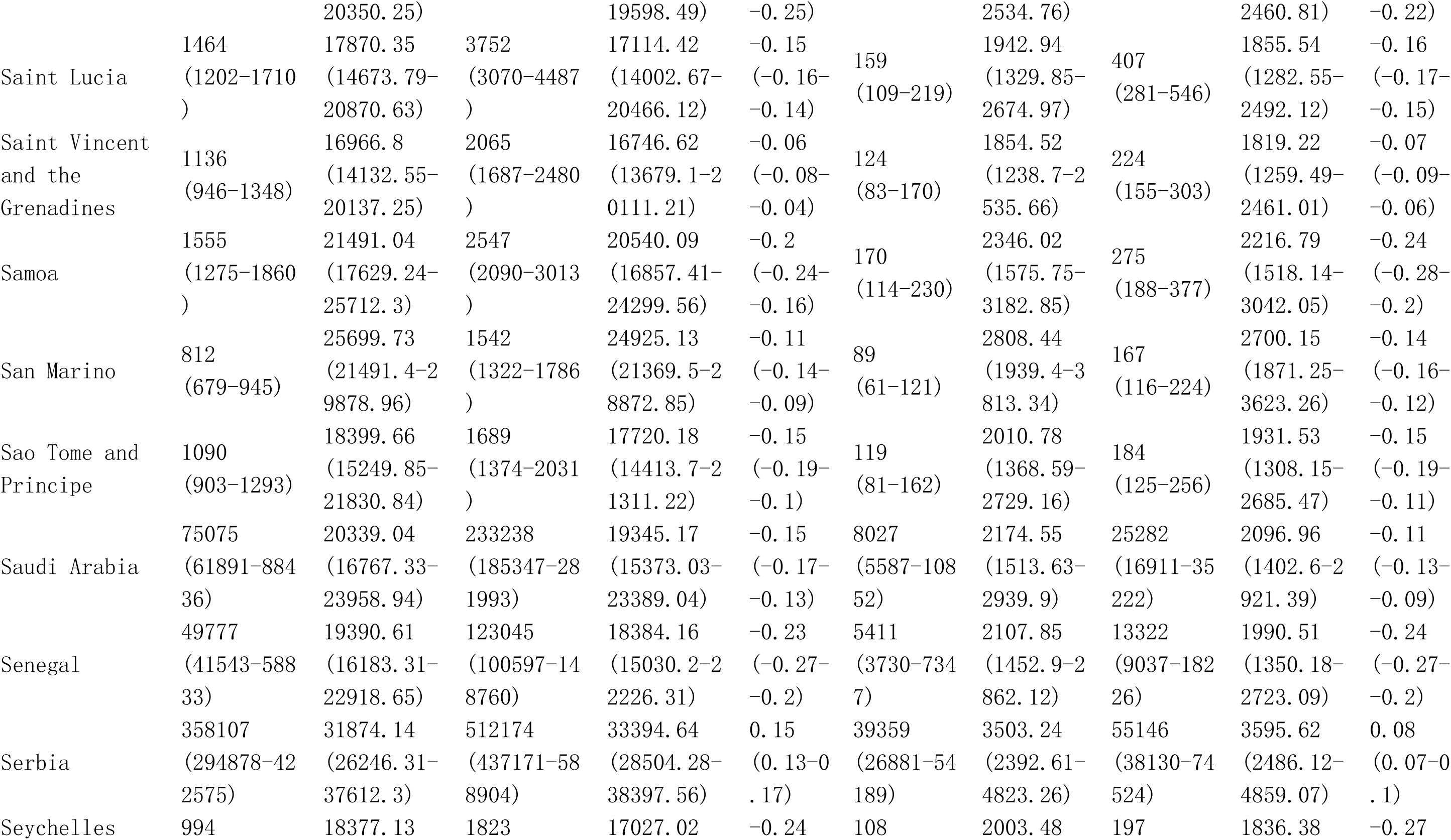
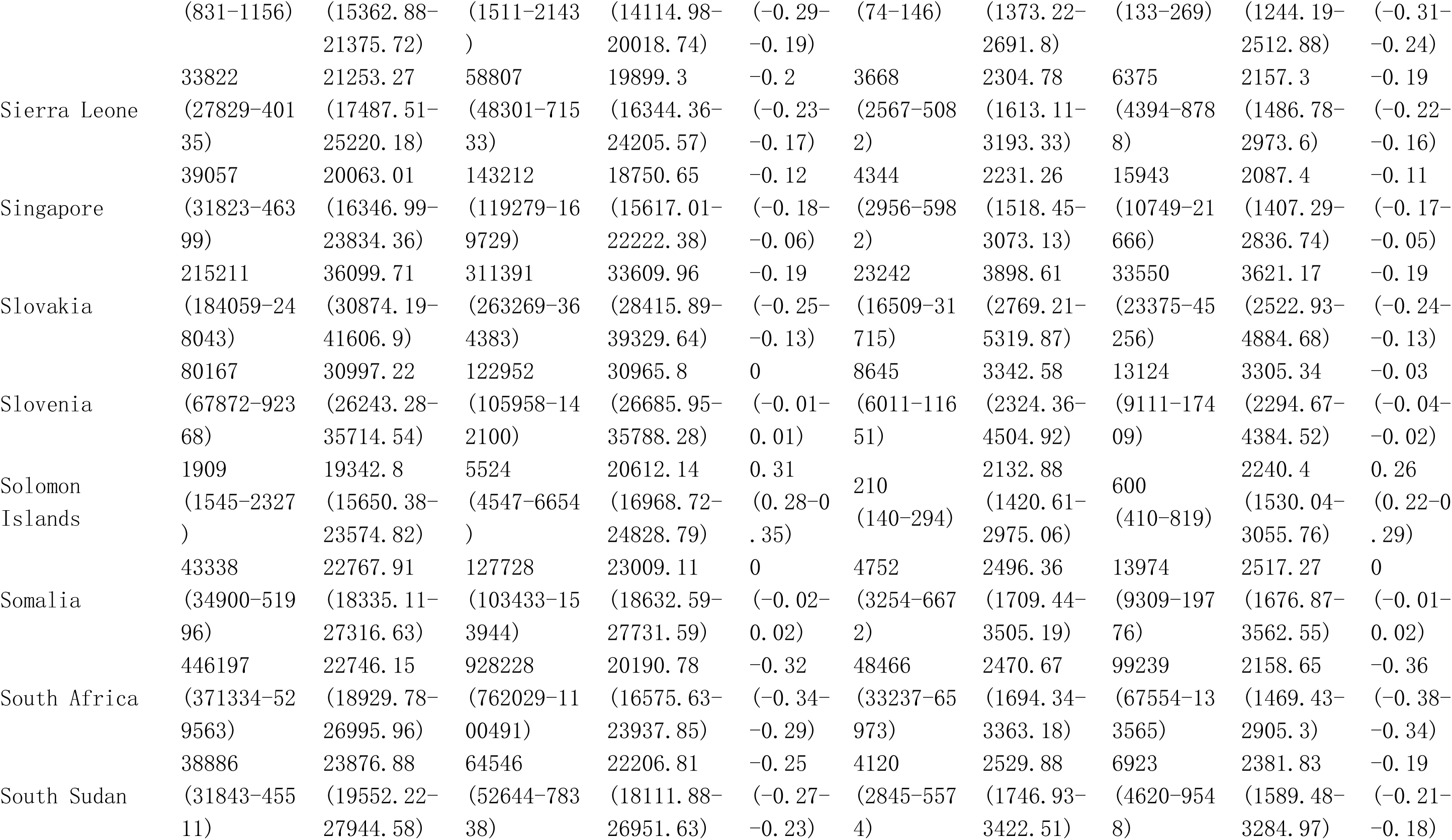
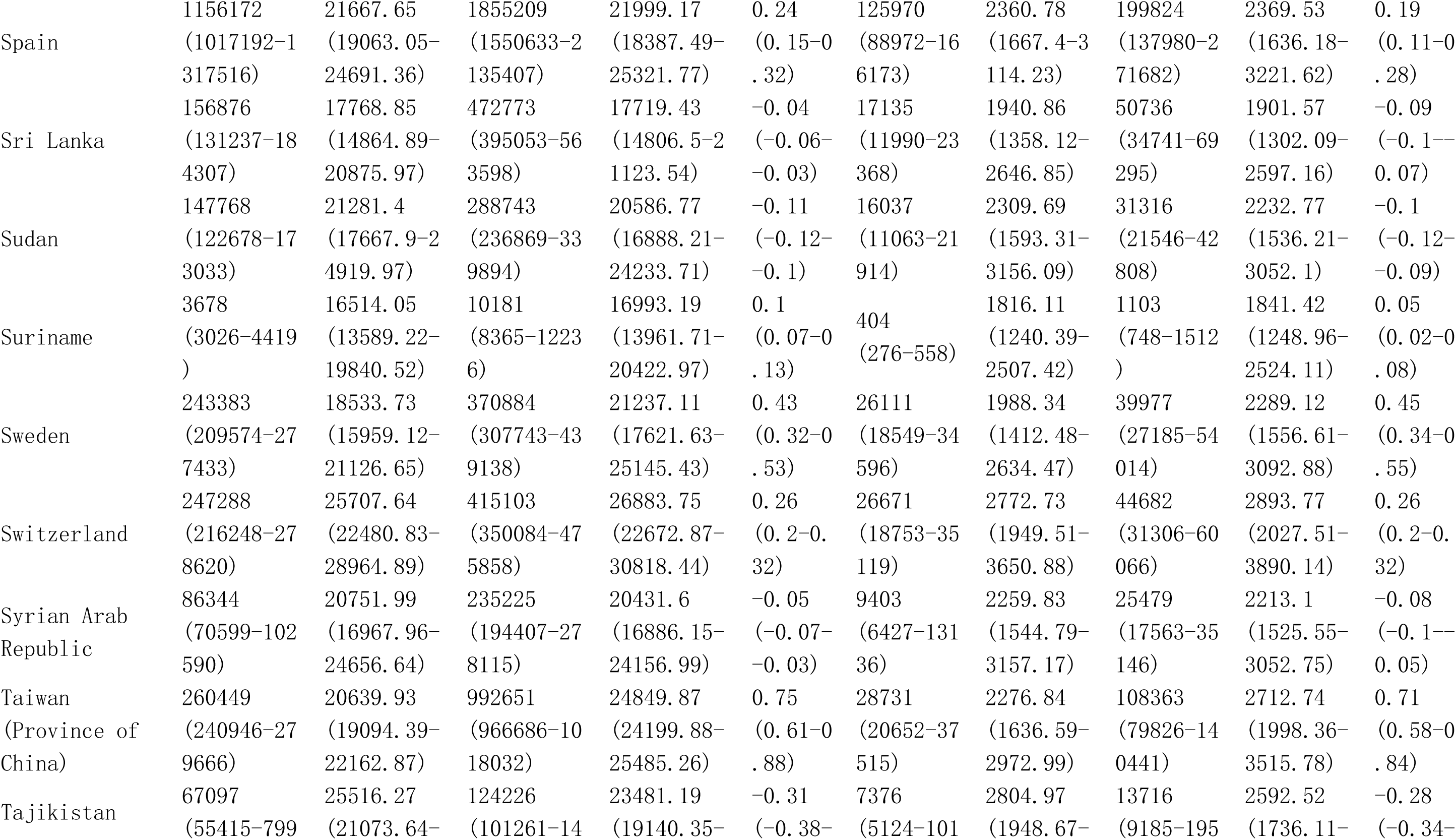
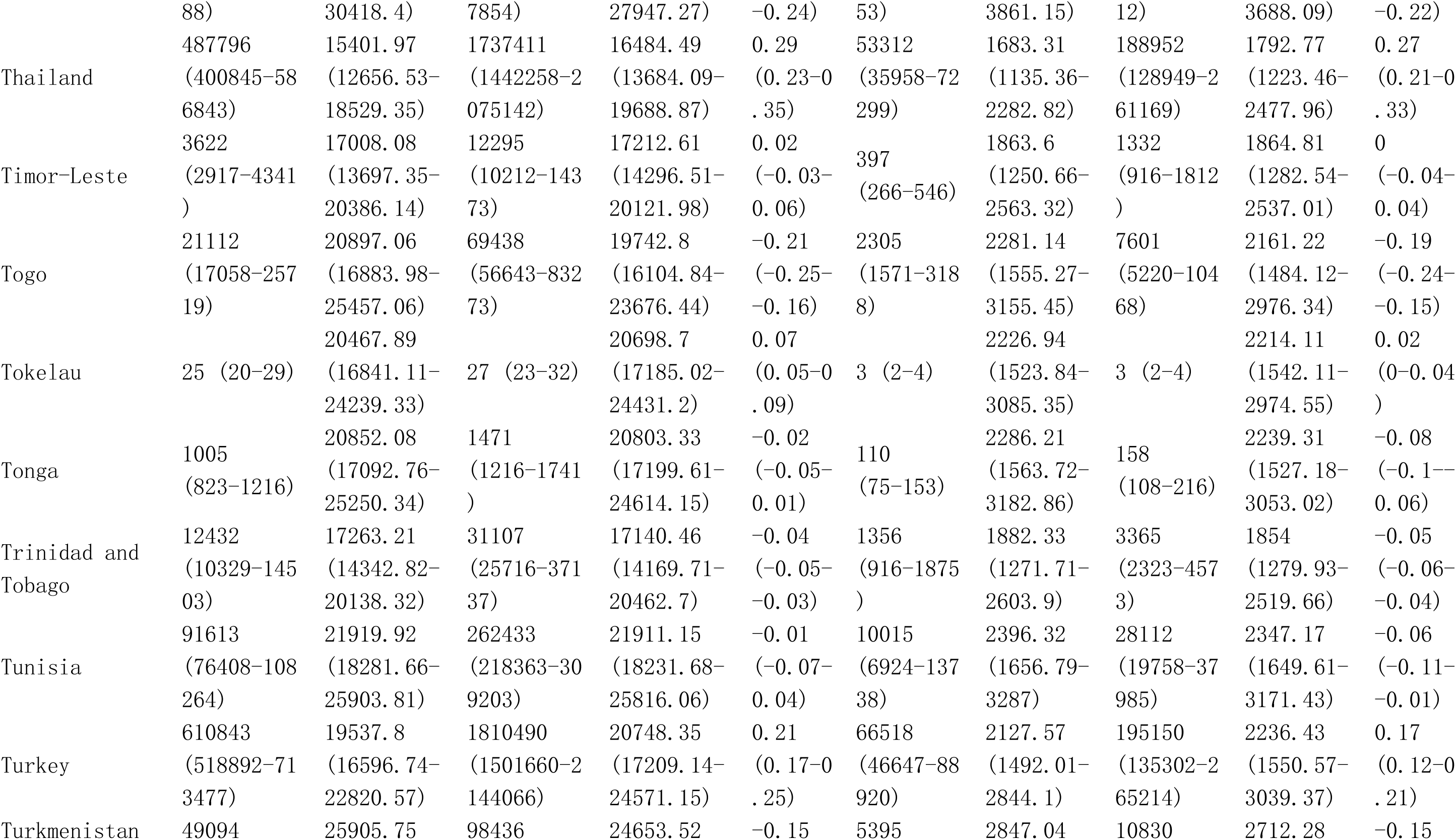
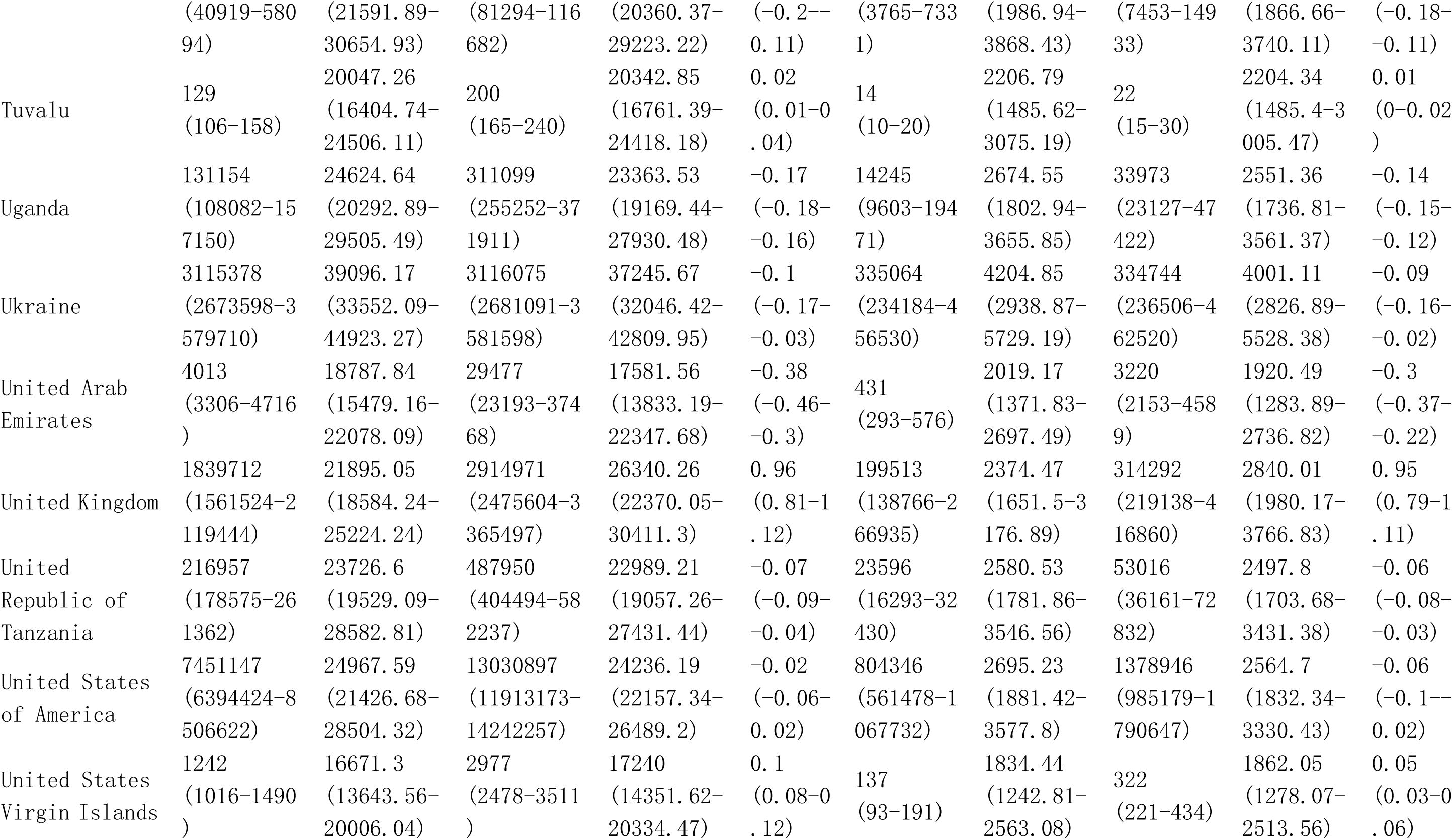
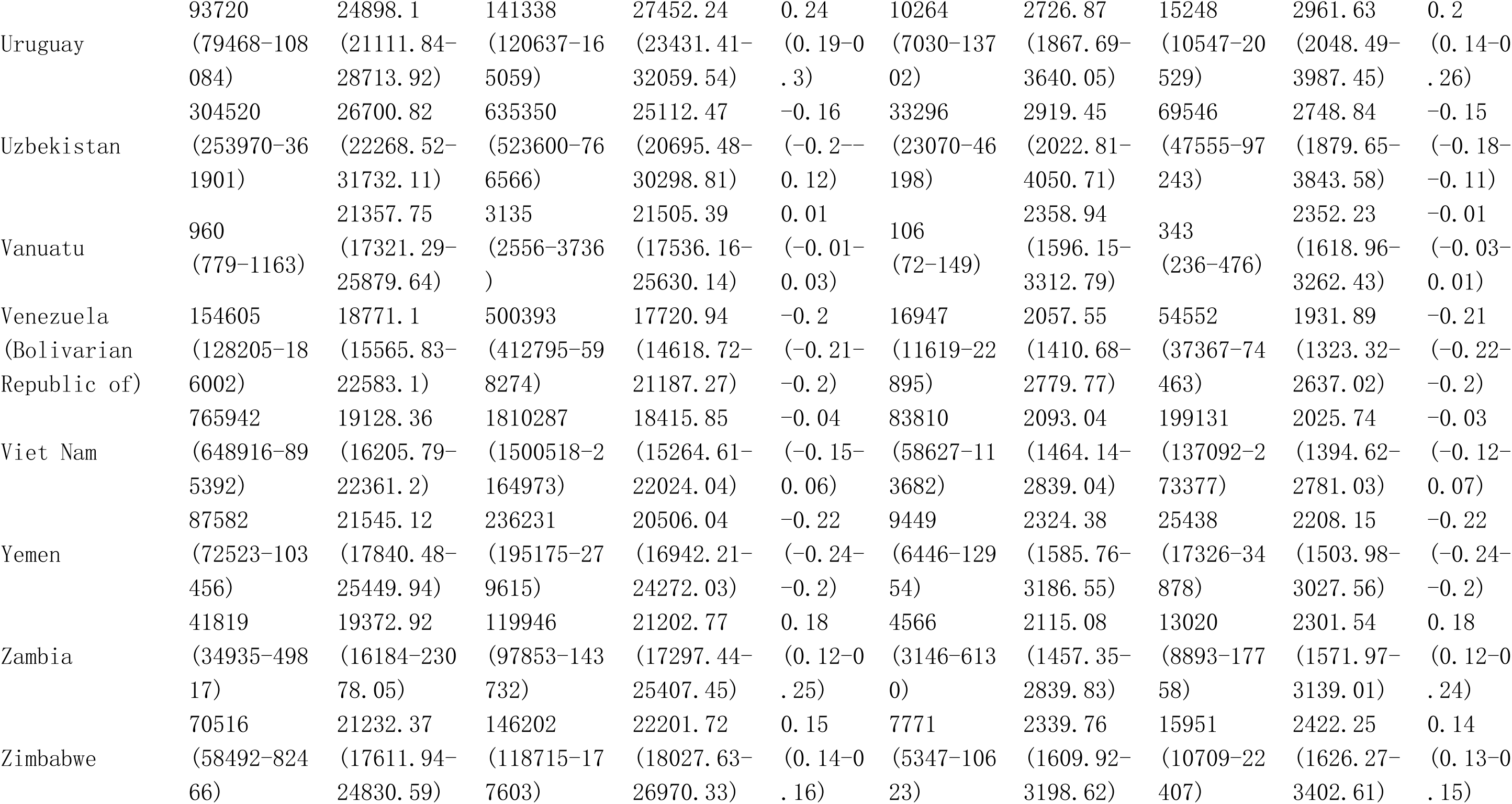
Global burden in Prevalence and DALYs of Low back pain among postmenopausal women from 1990 to 2021 by 21 GBD geographical regions, and 204 countries and territories.

**Table S9.**
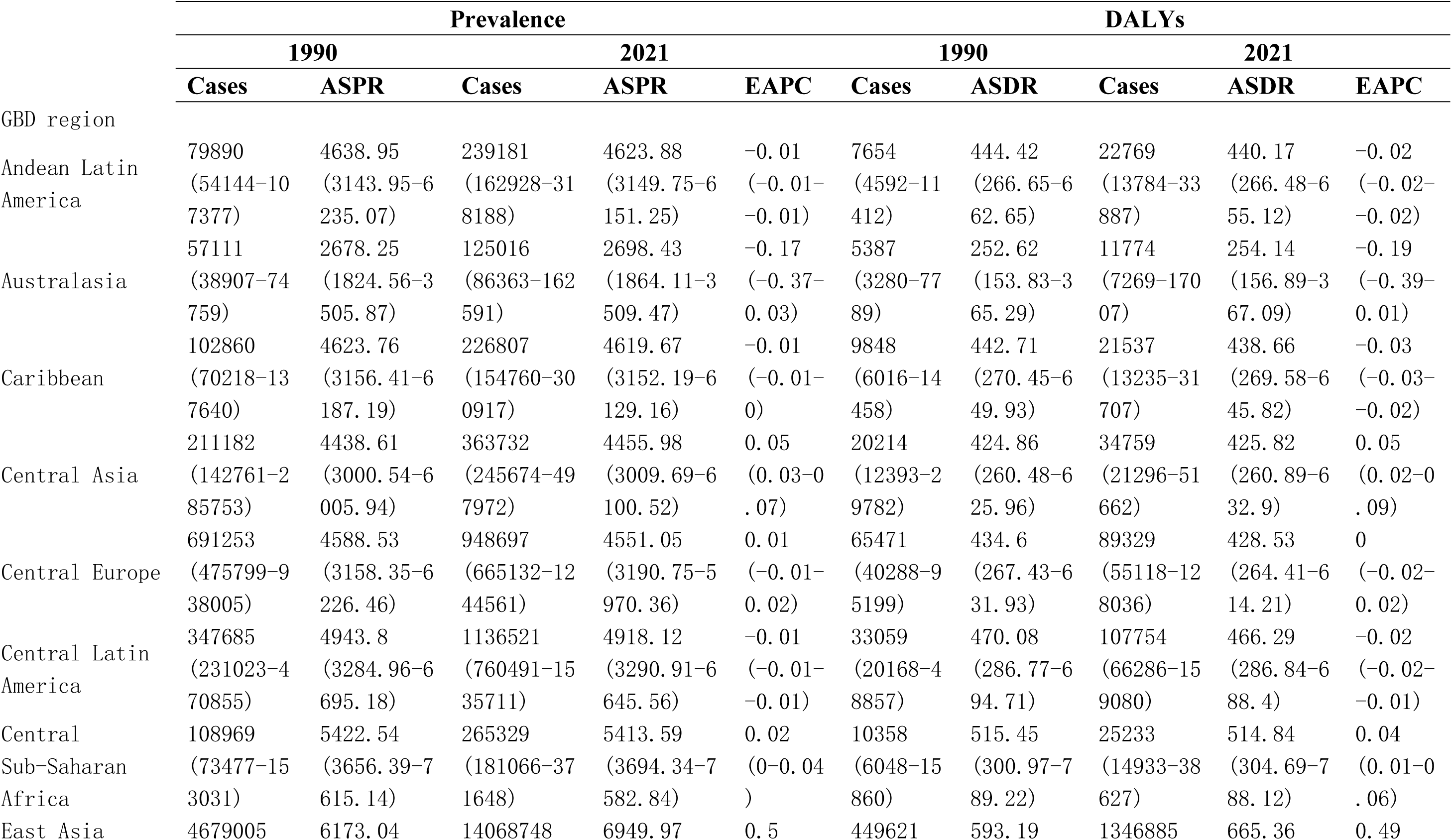
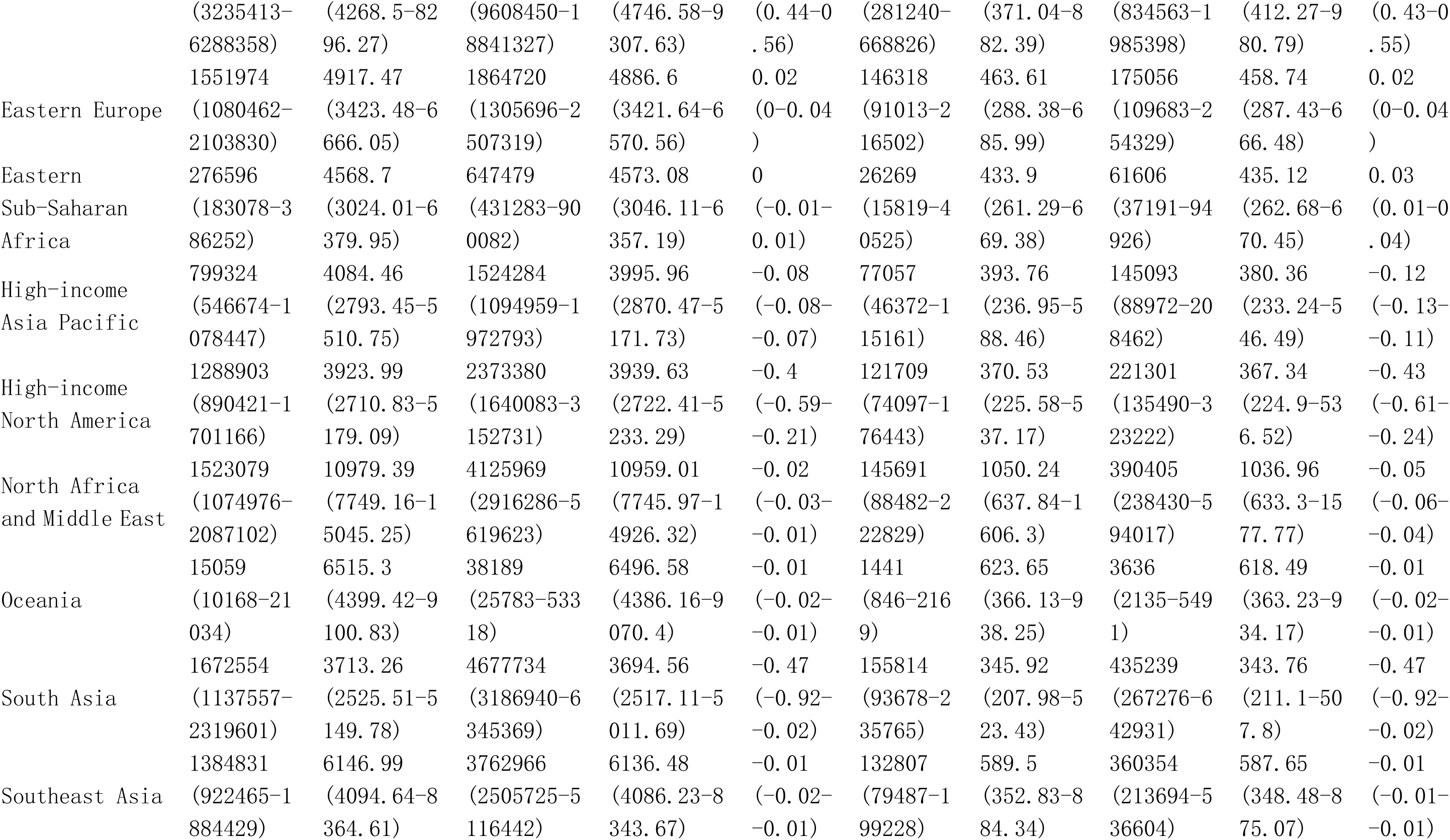
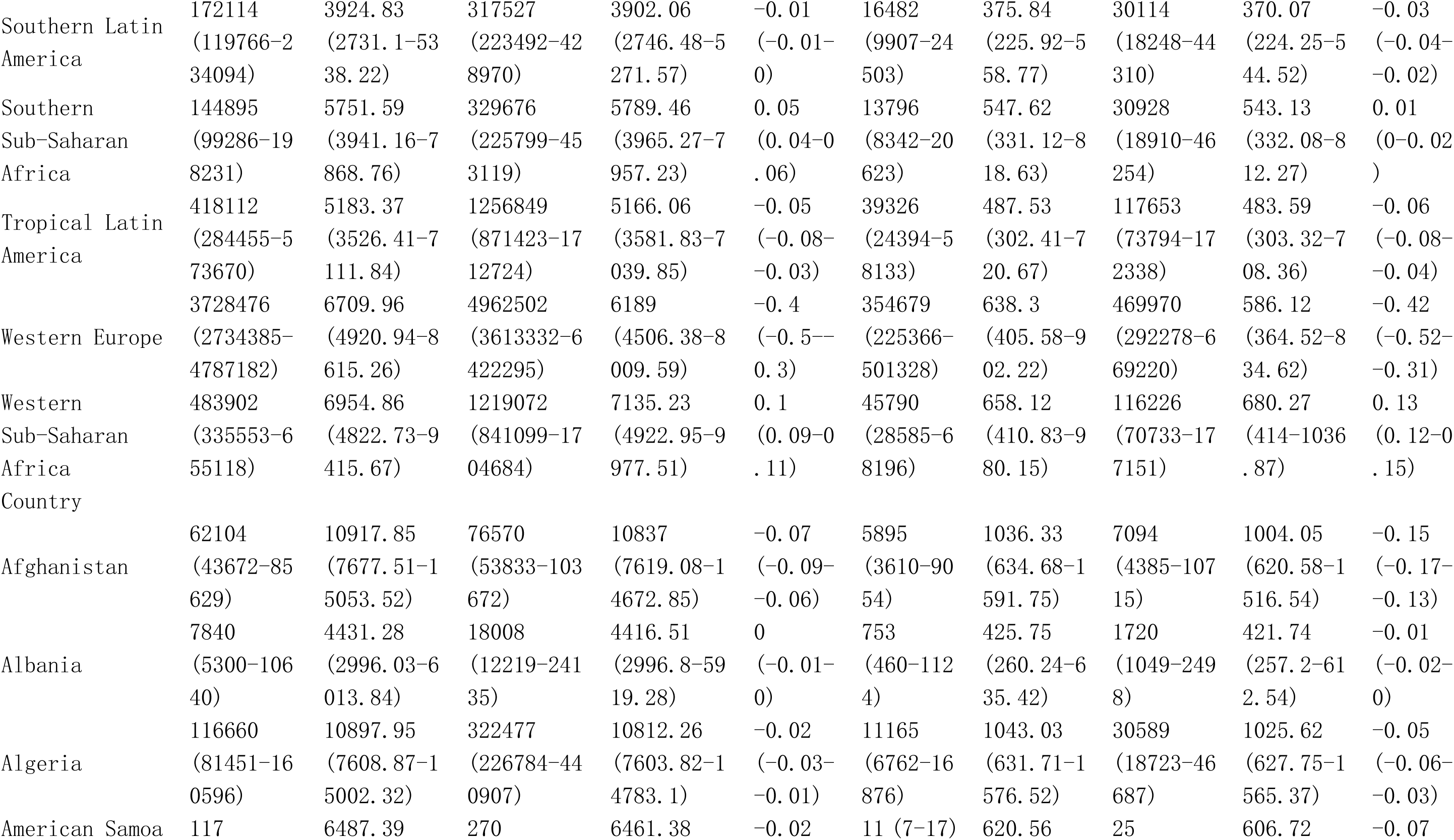
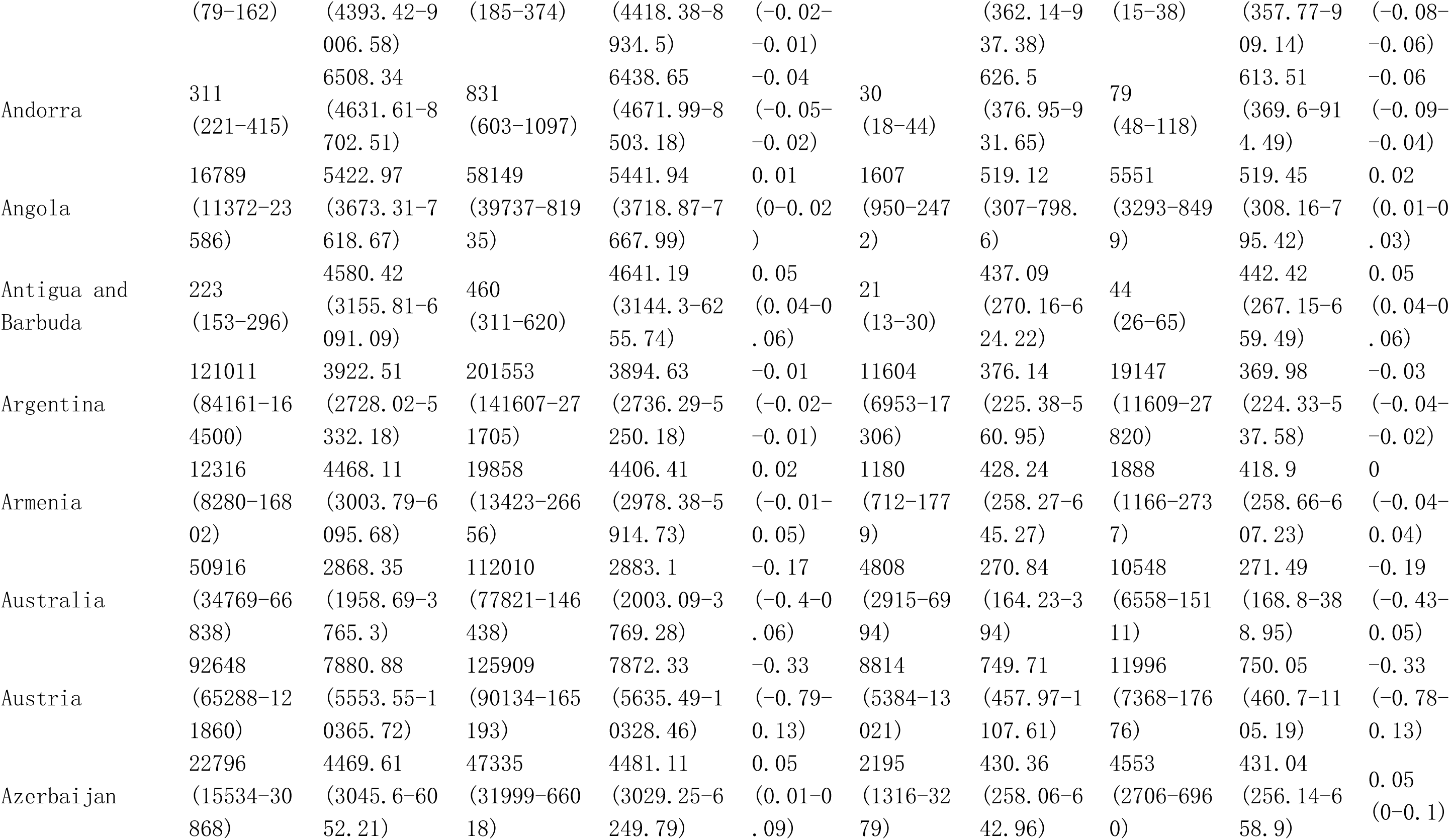
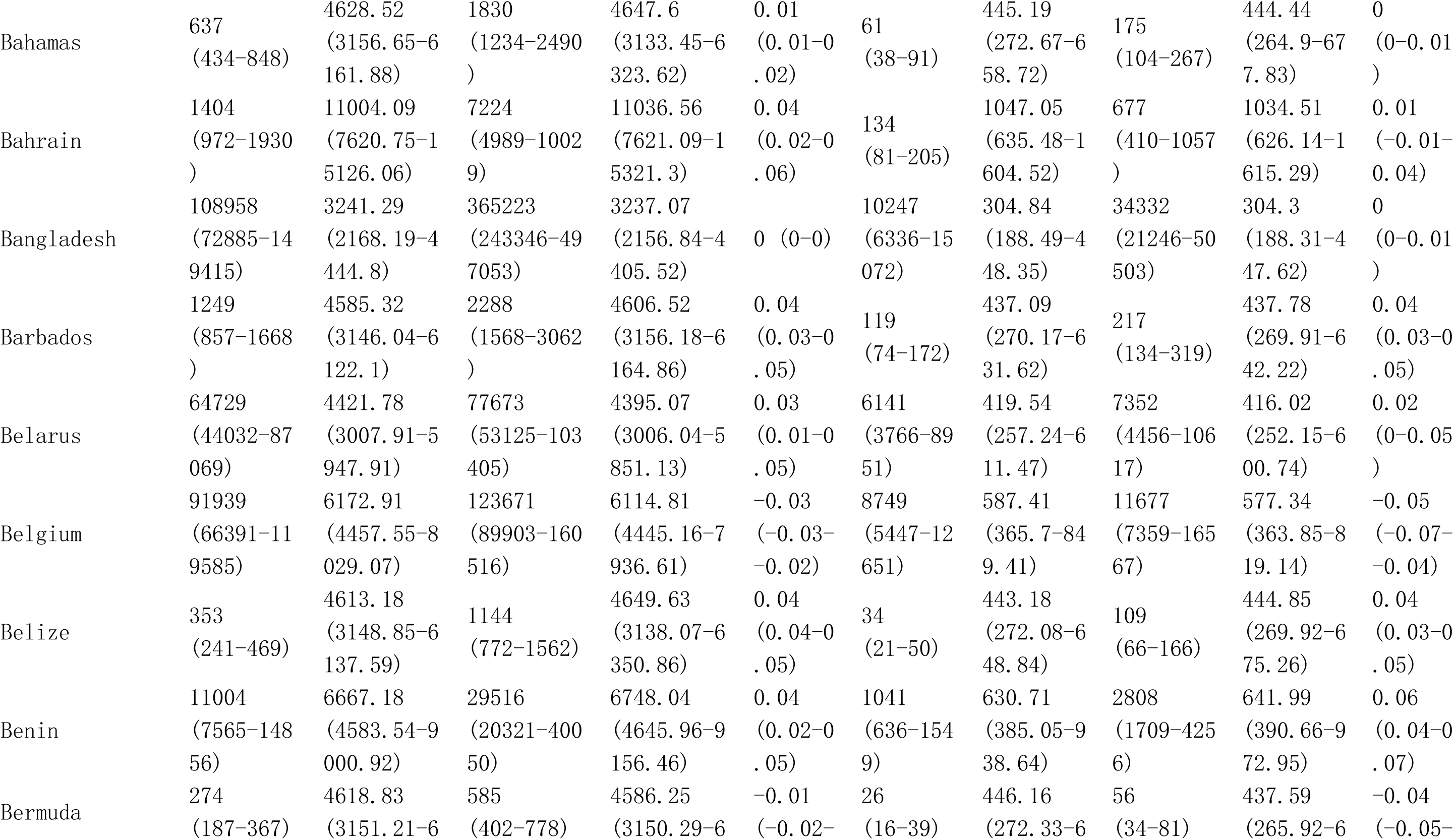
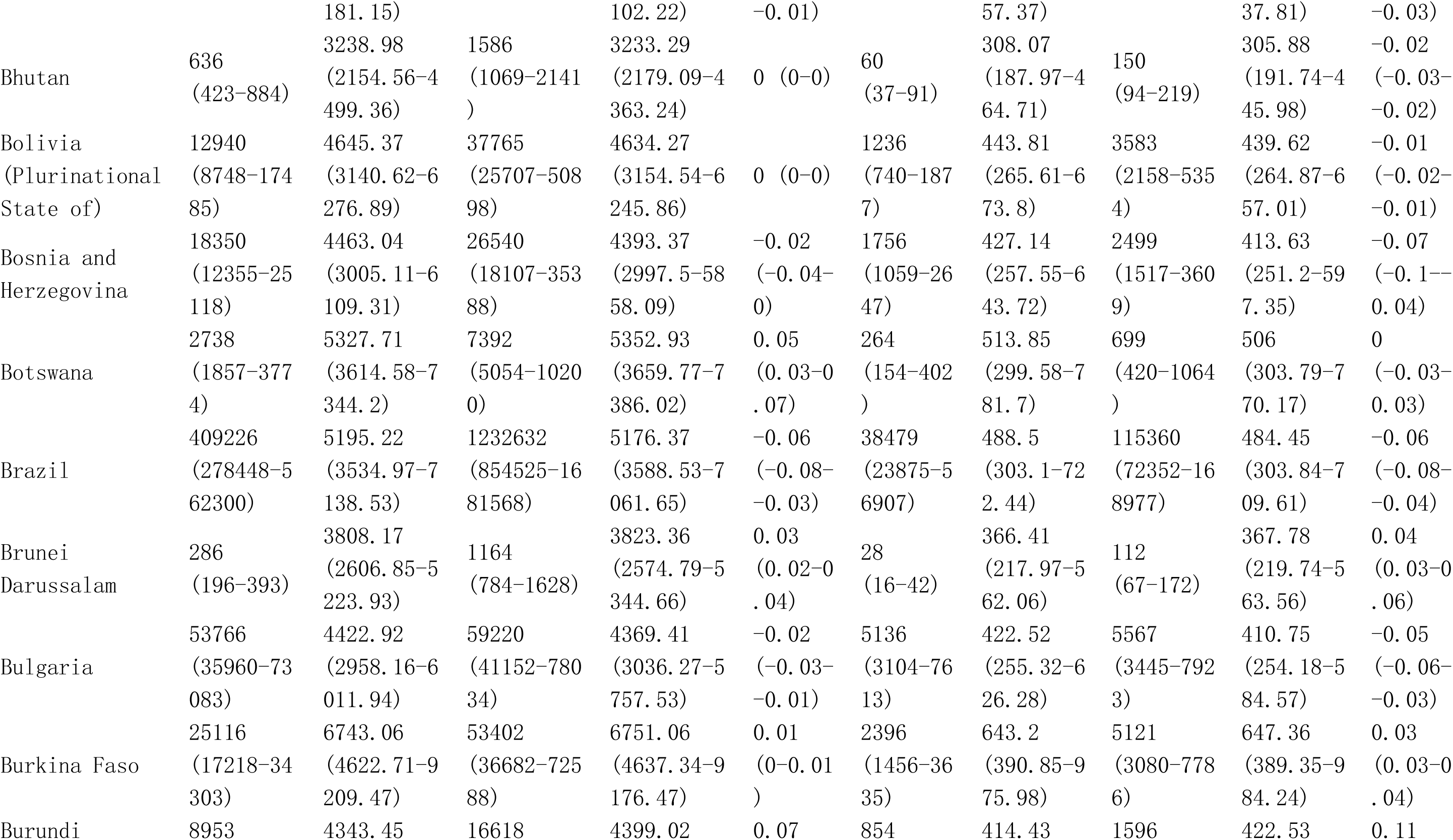
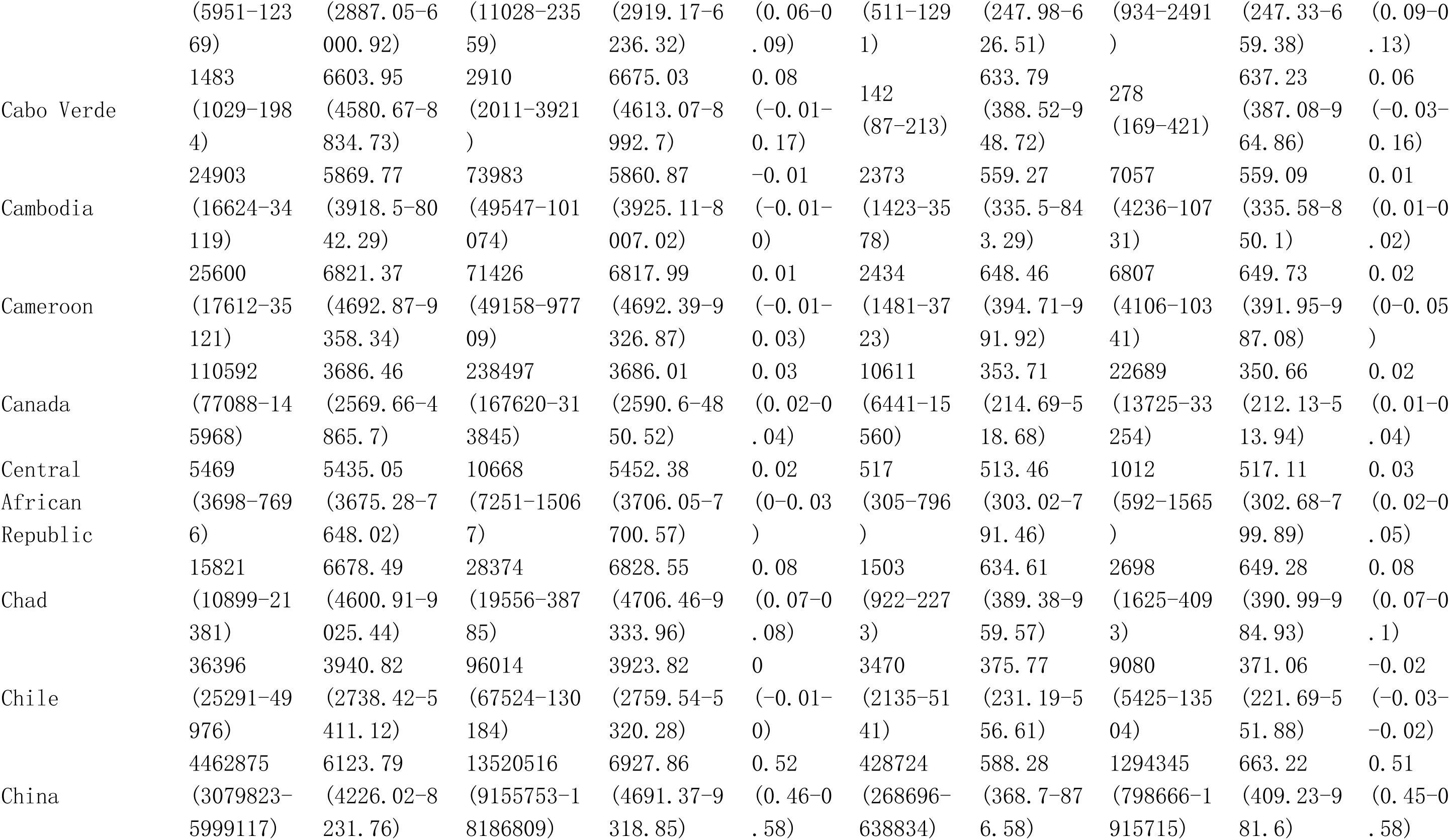
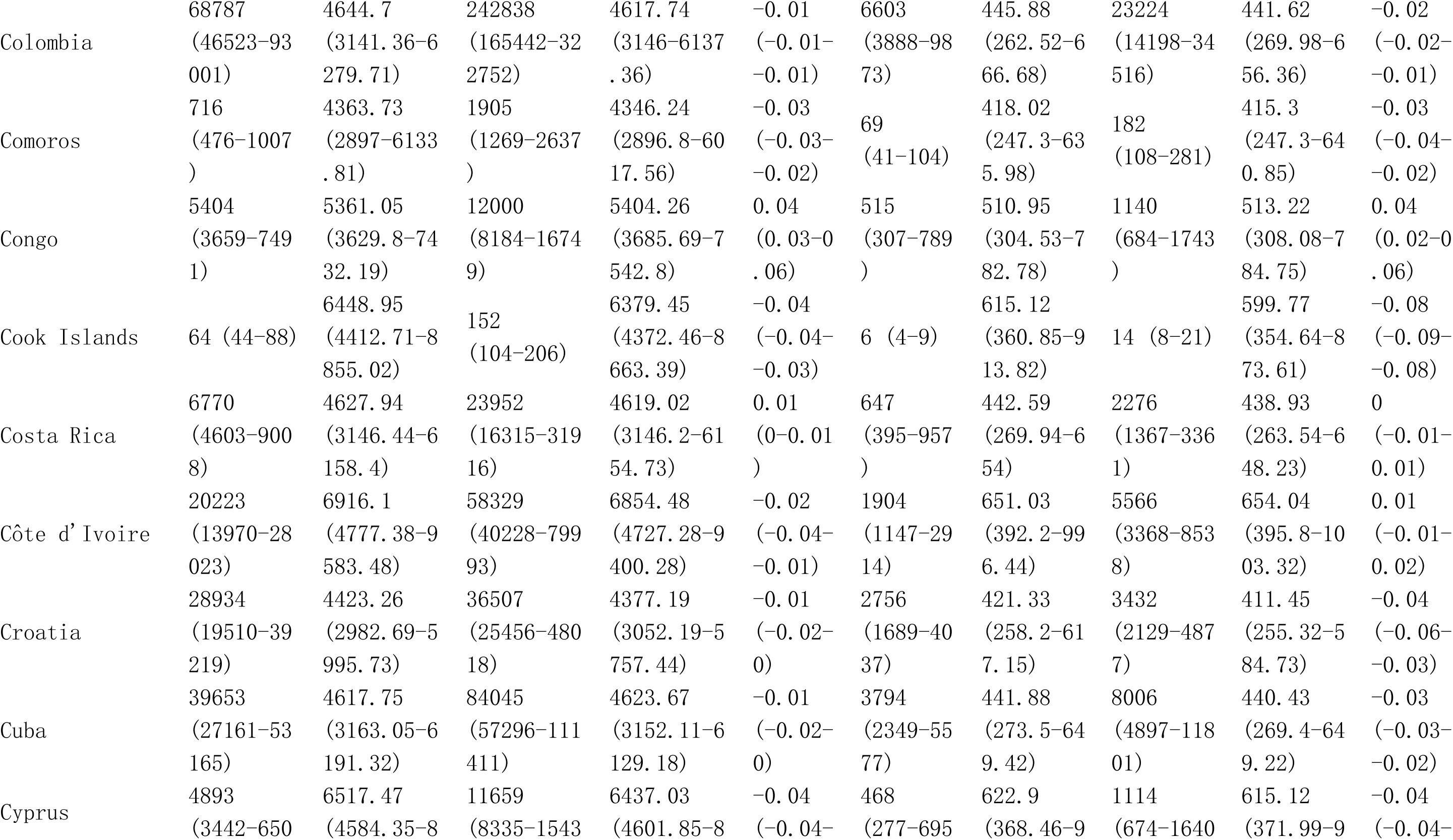
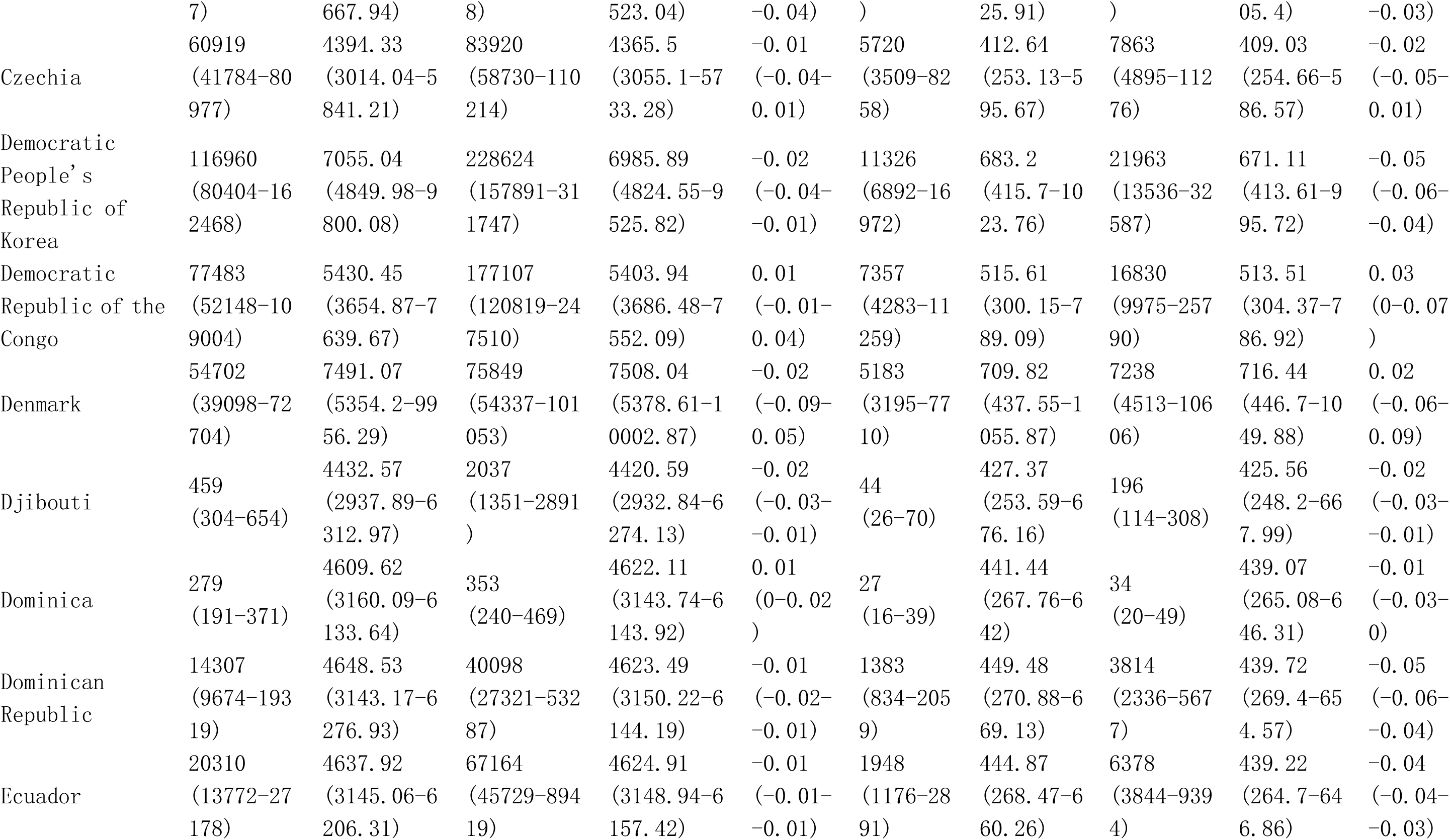
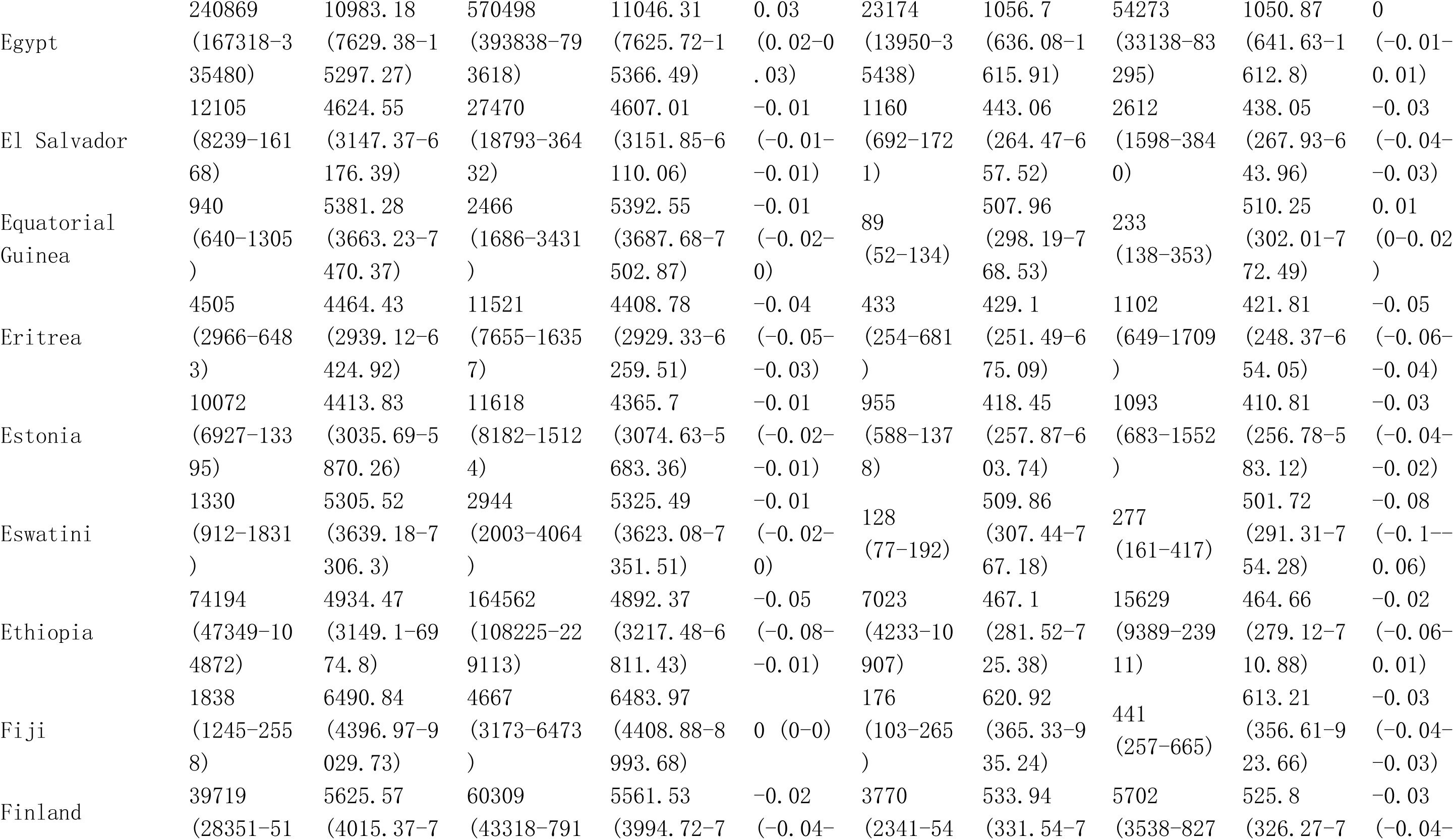
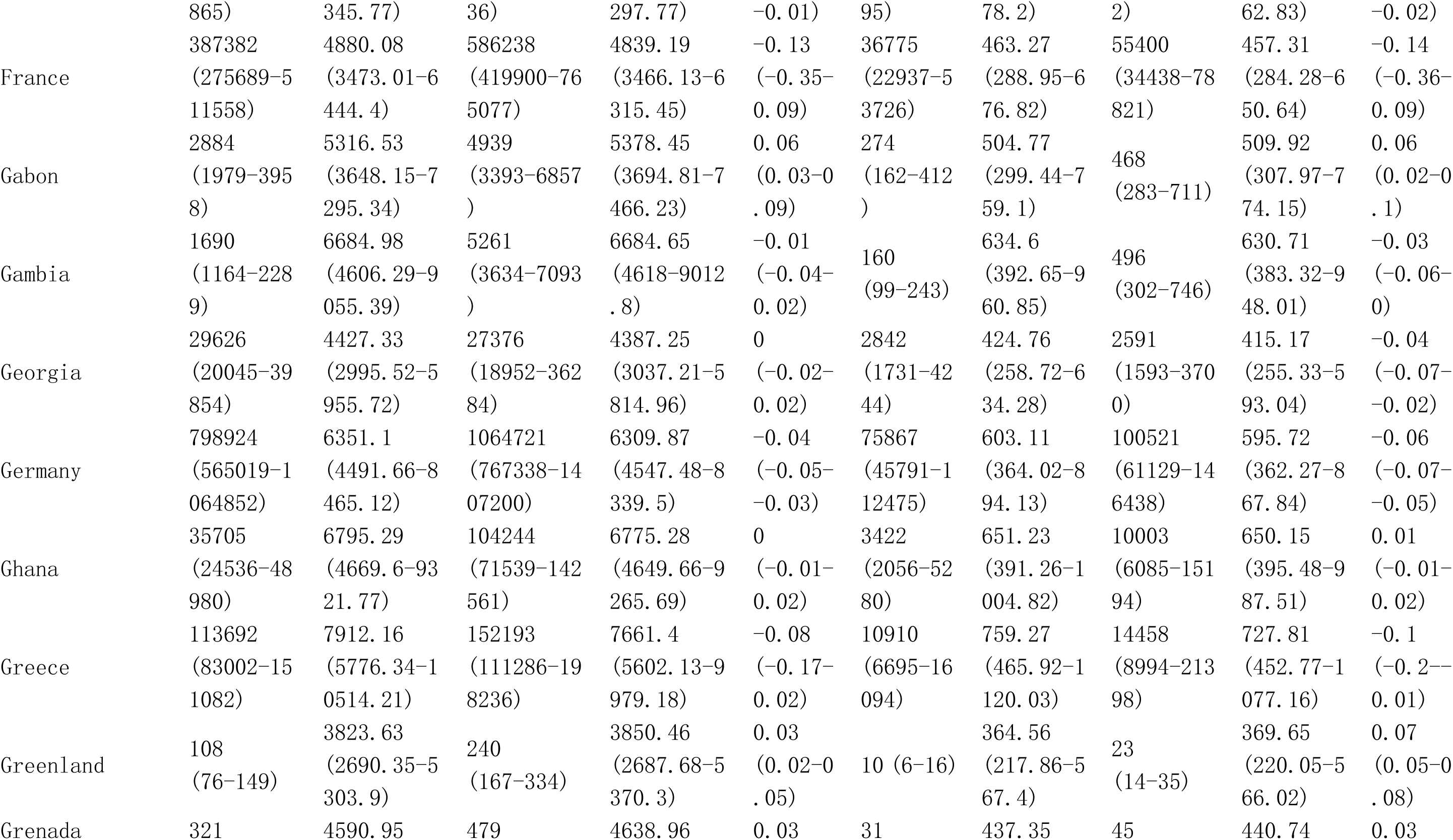
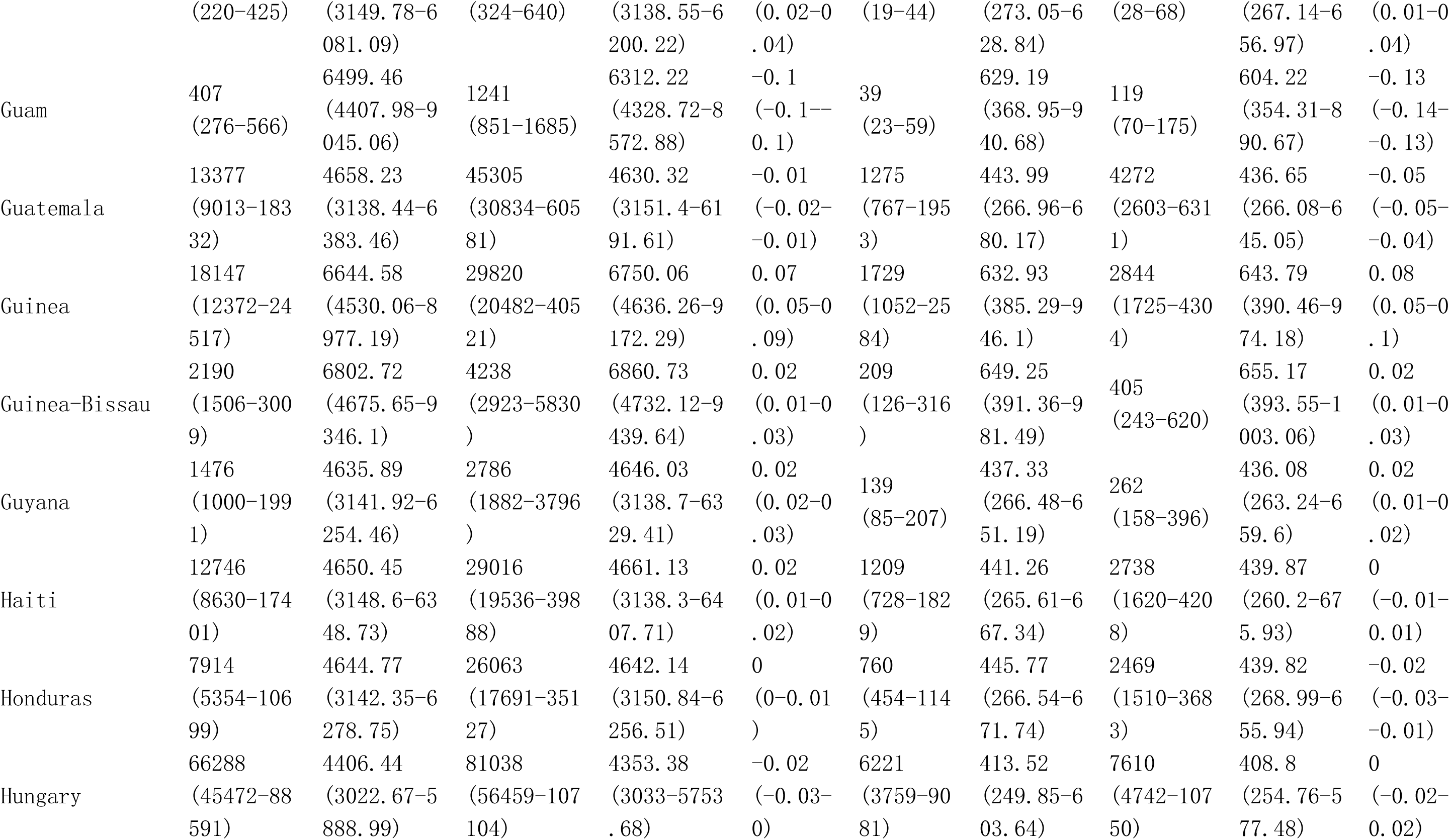
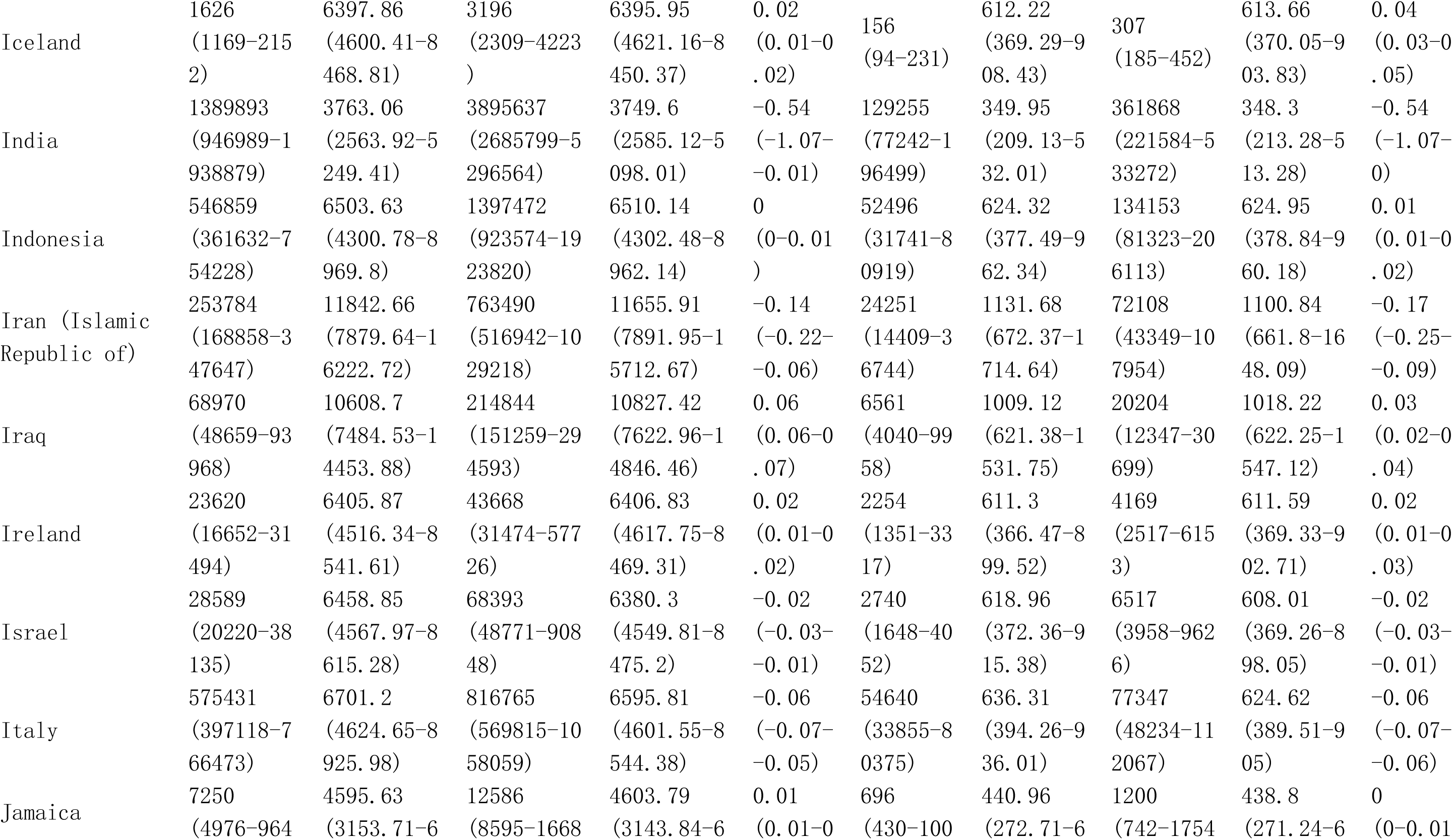
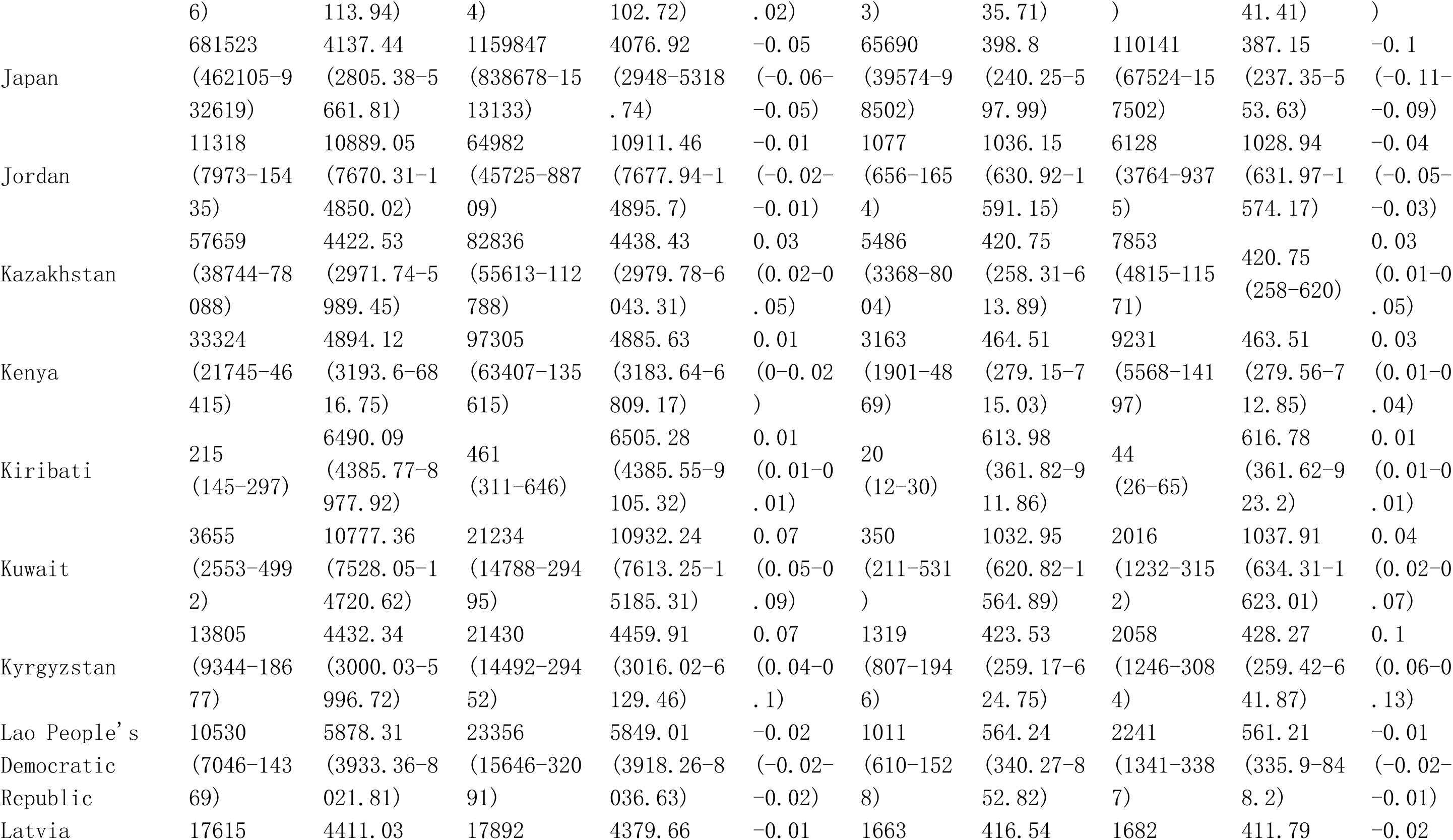
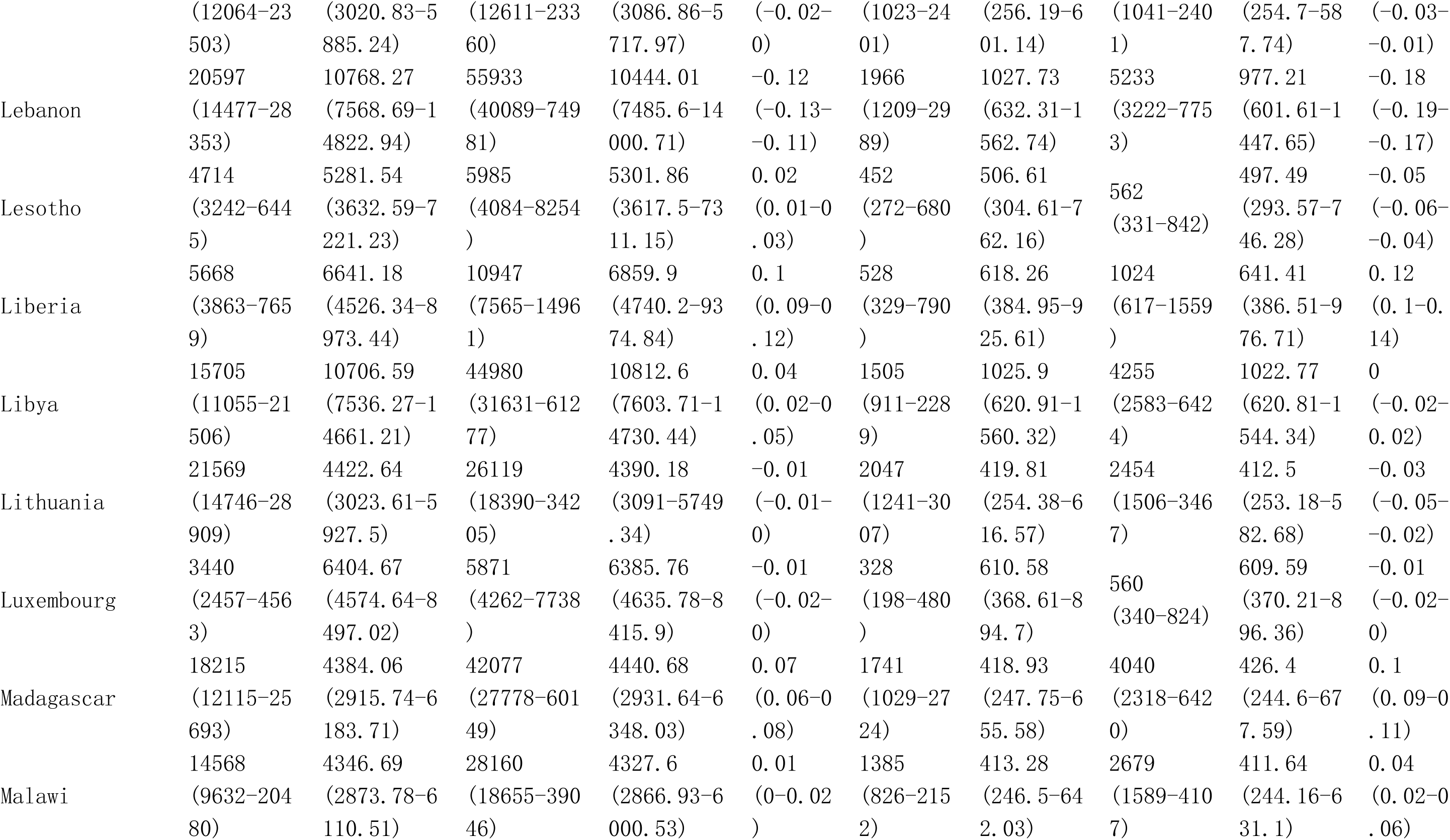
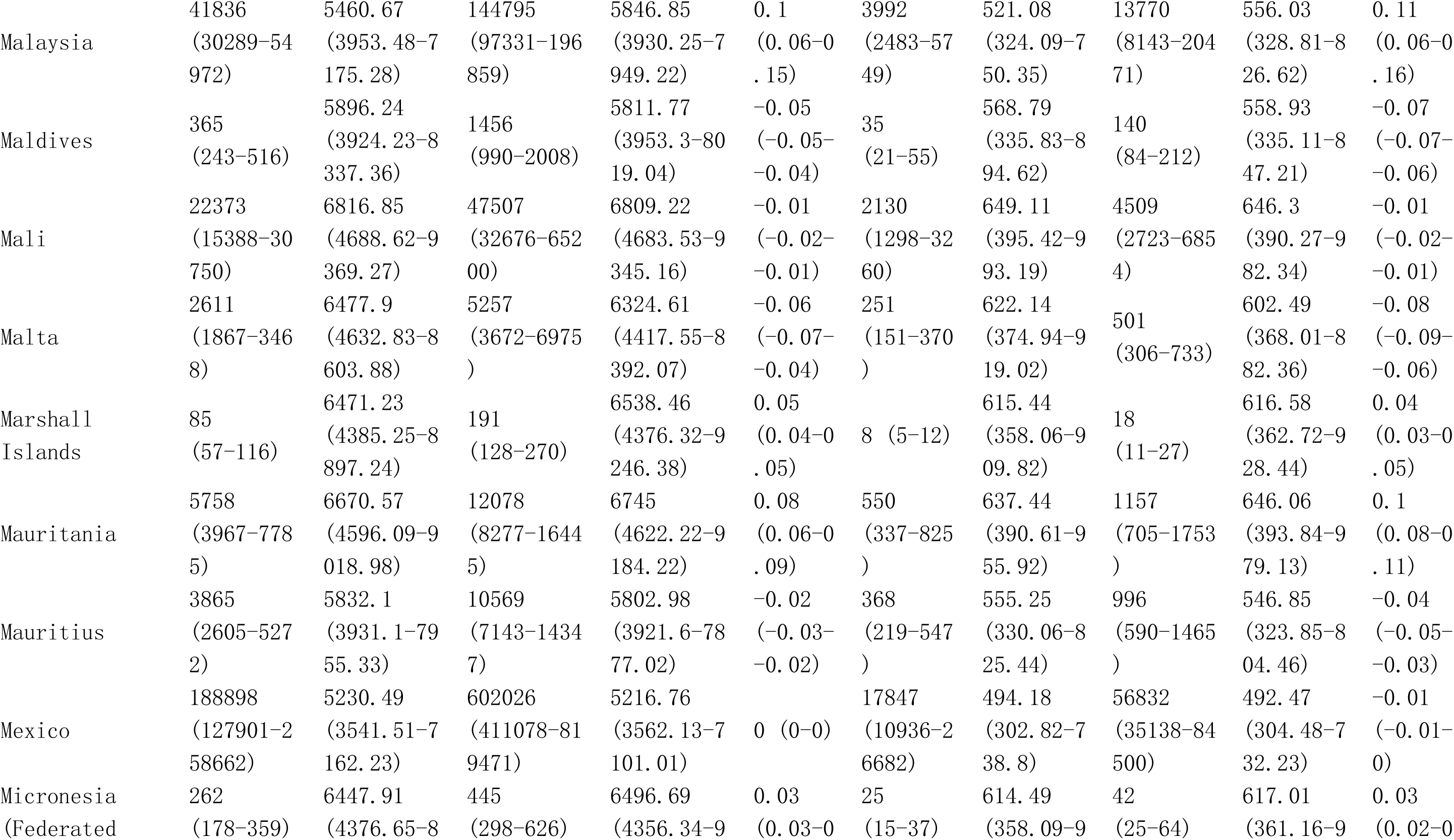
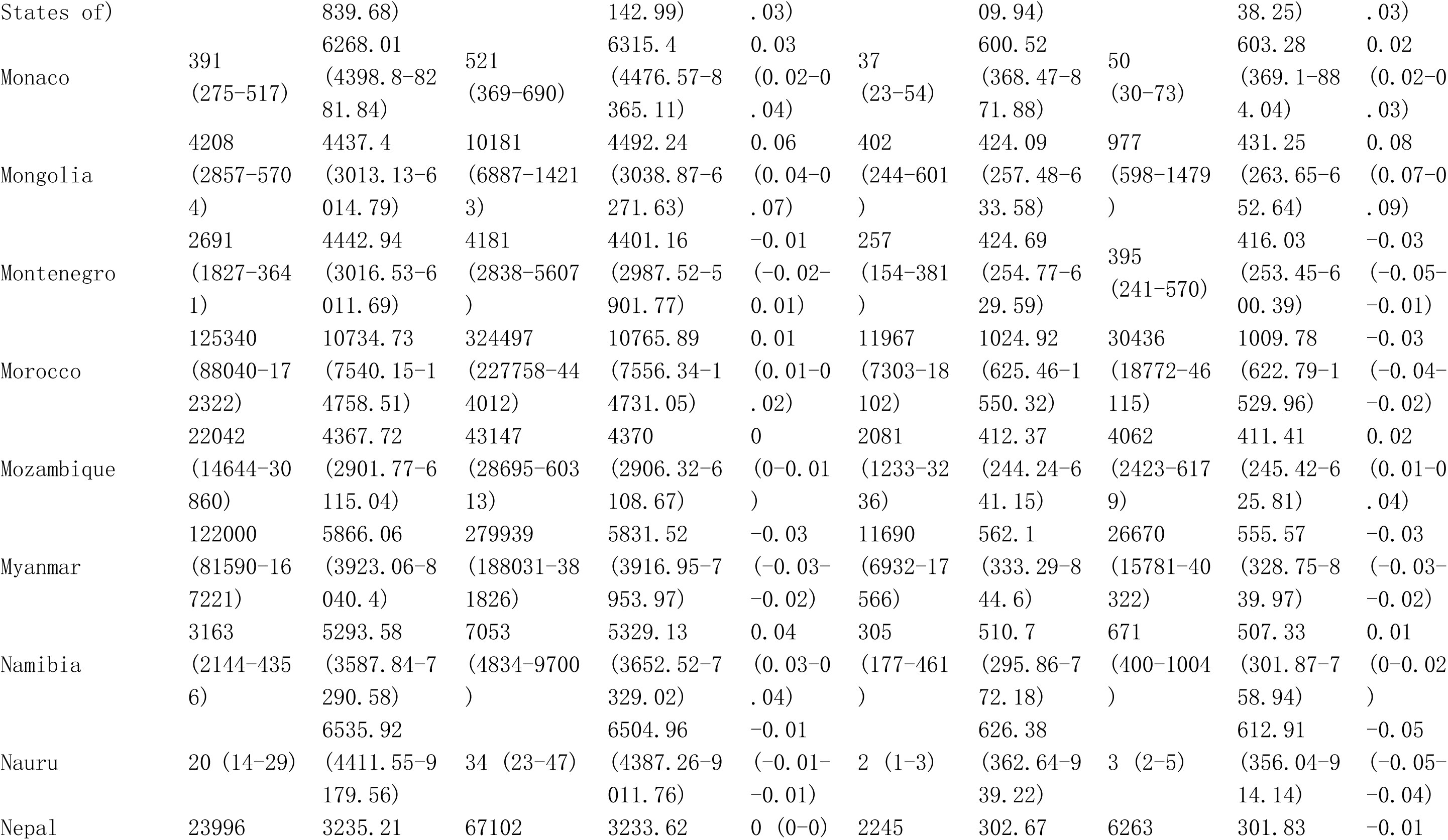
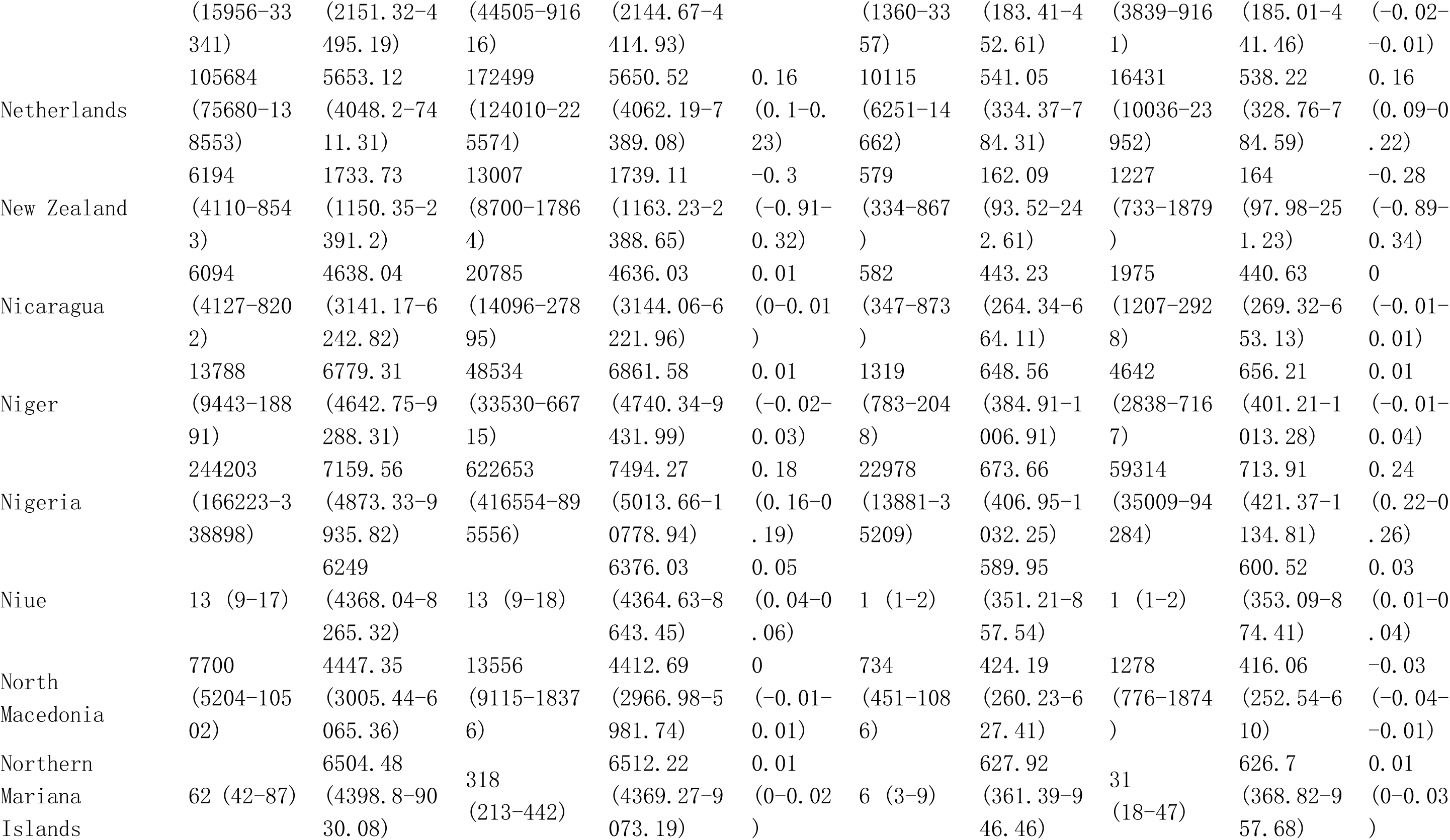
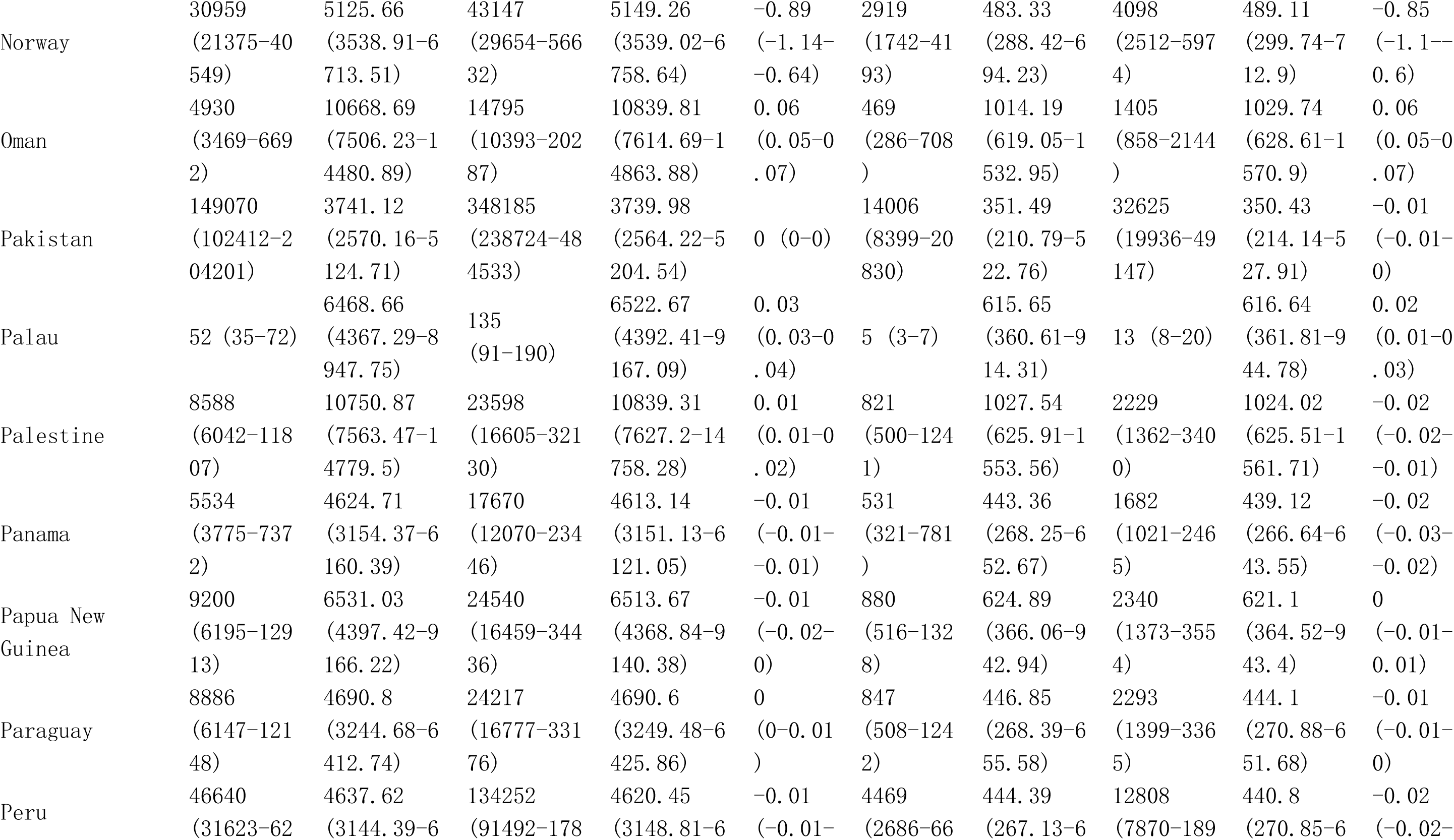
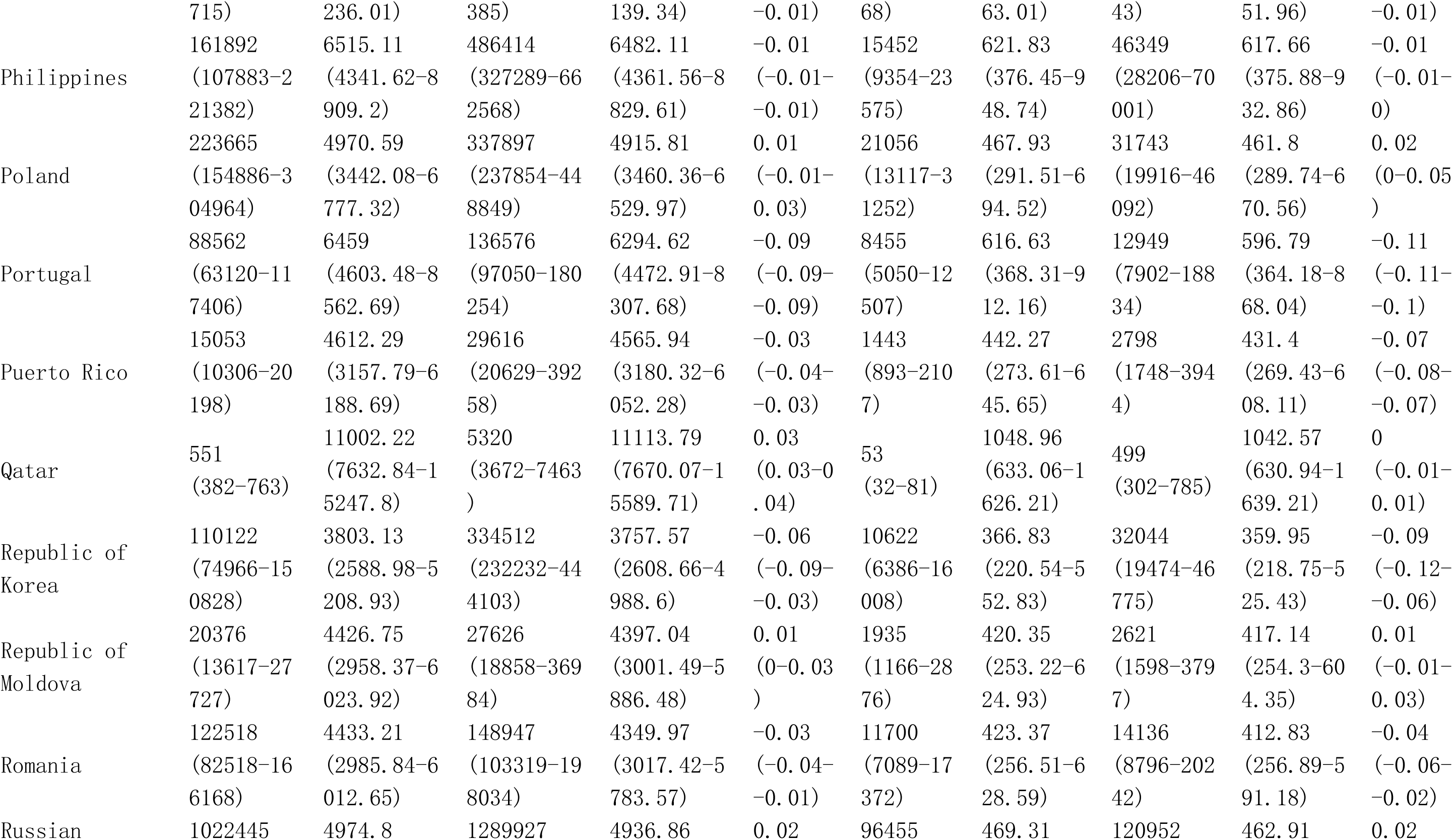
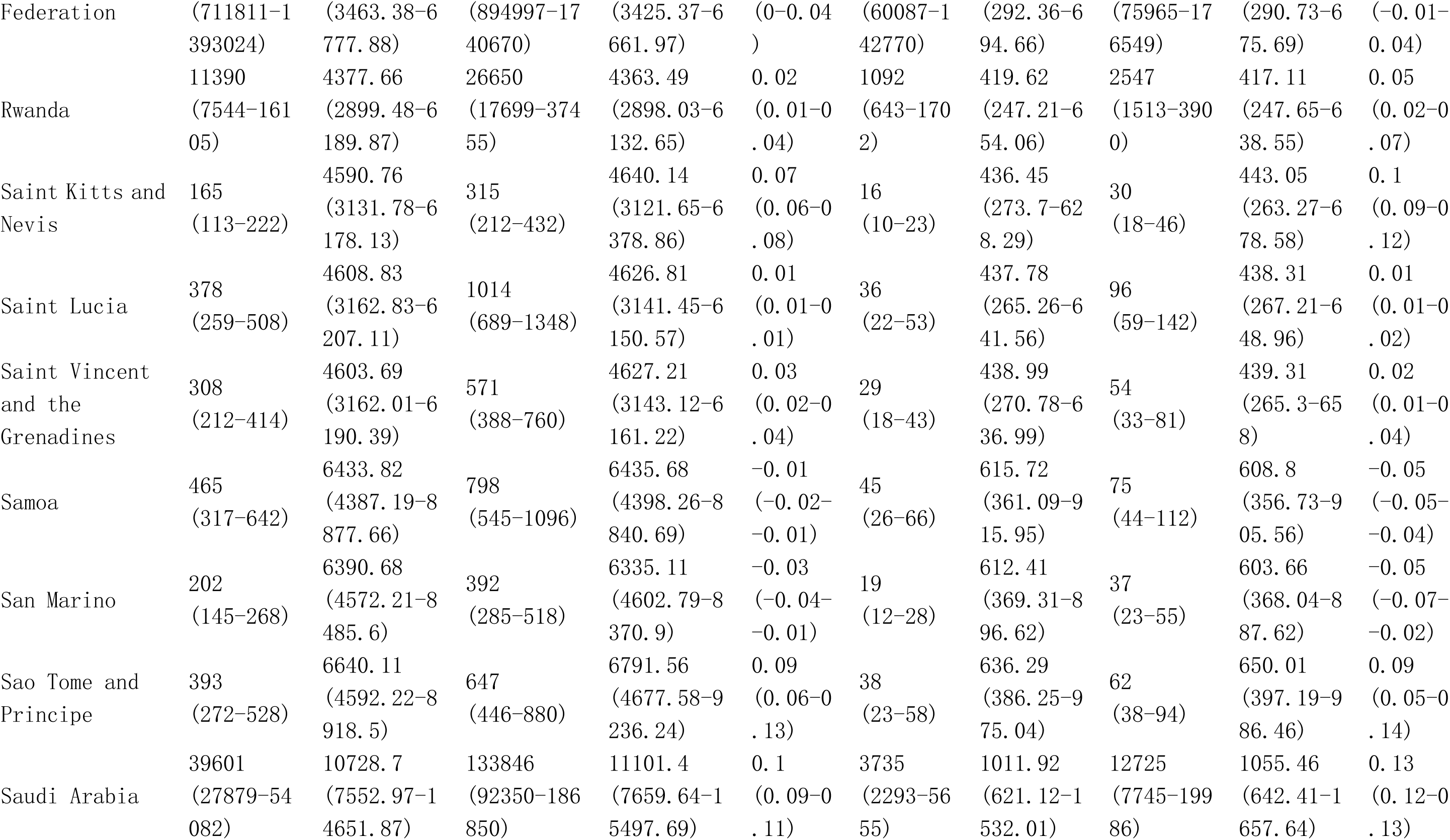
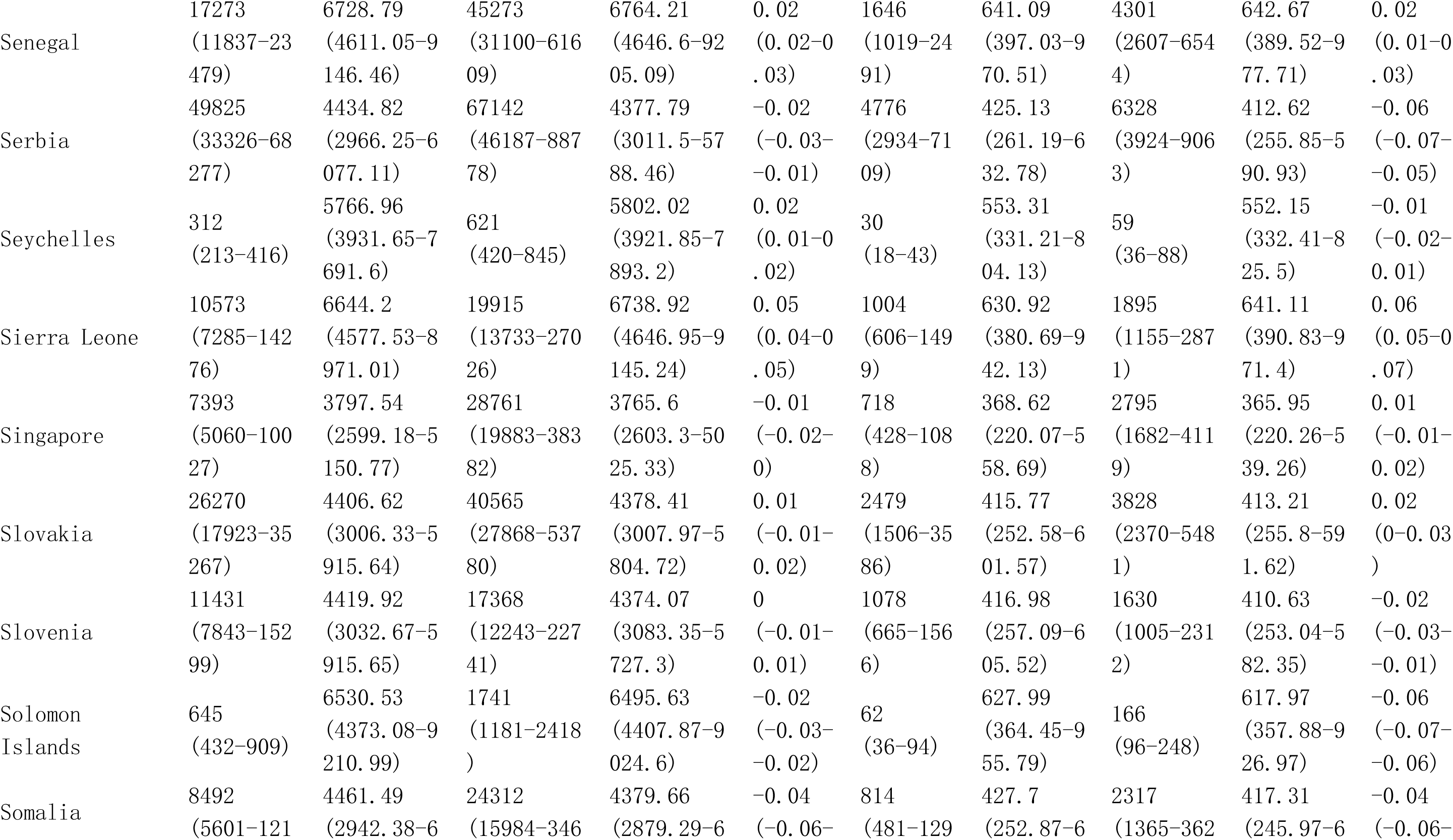
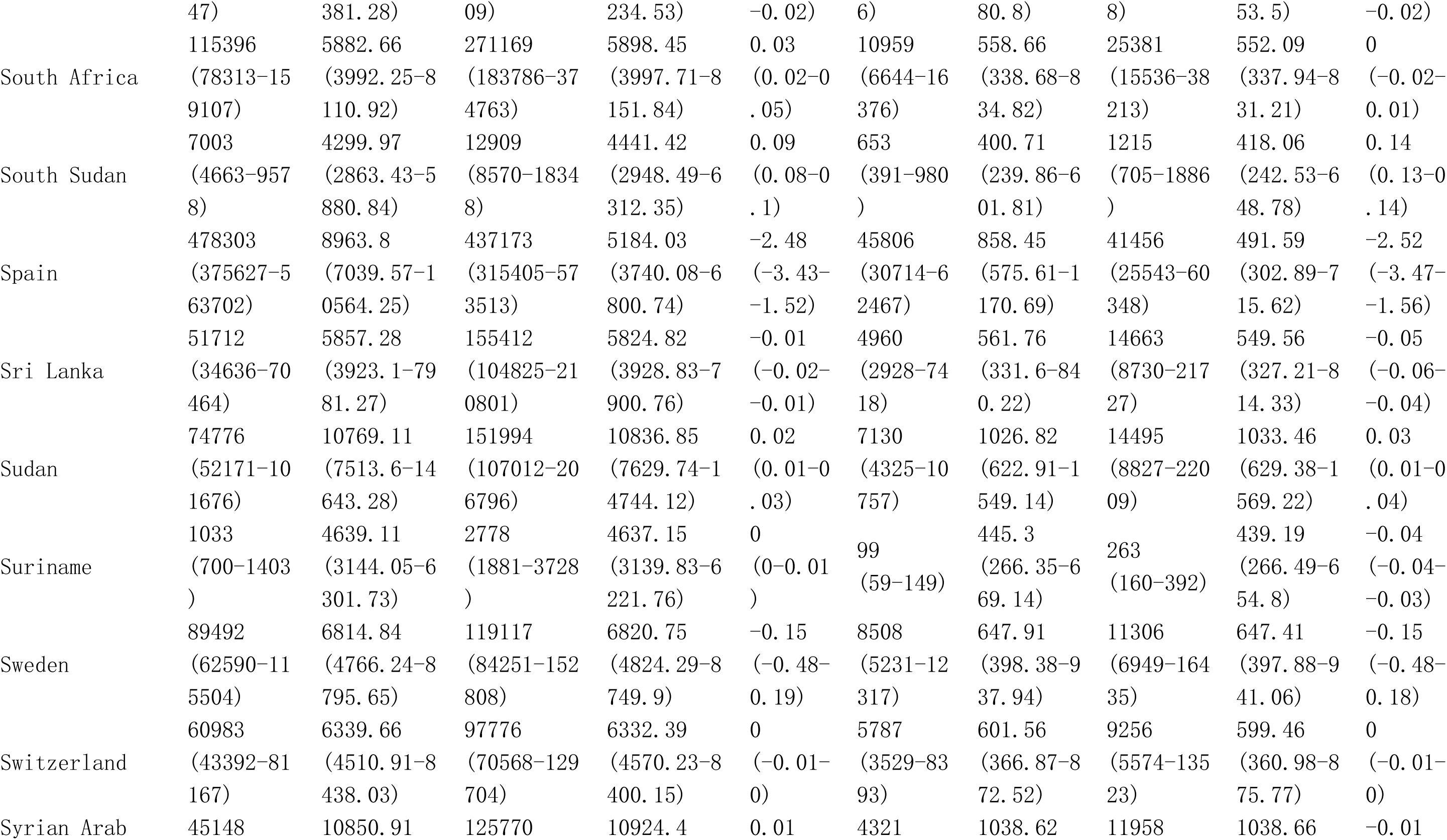
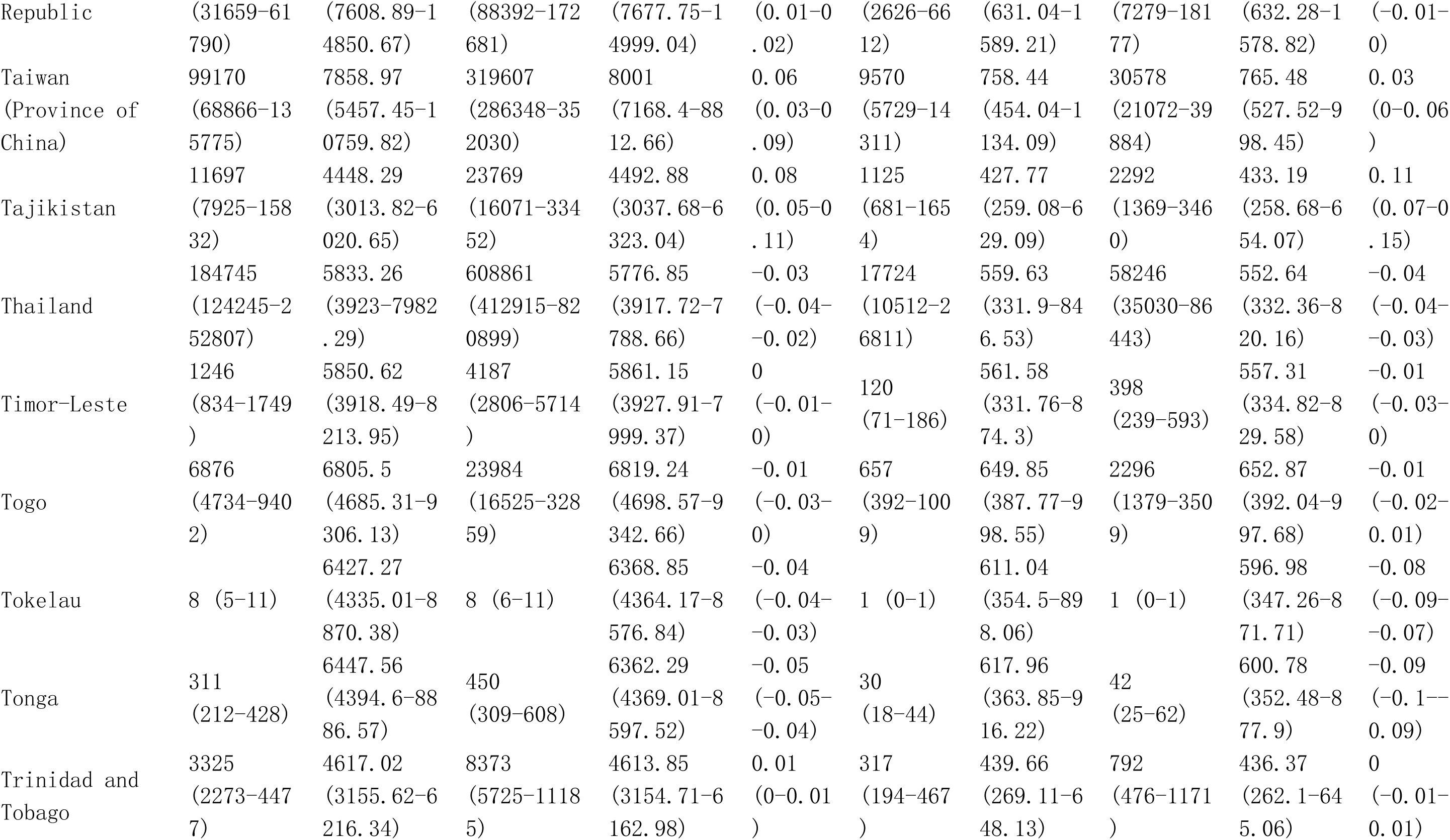
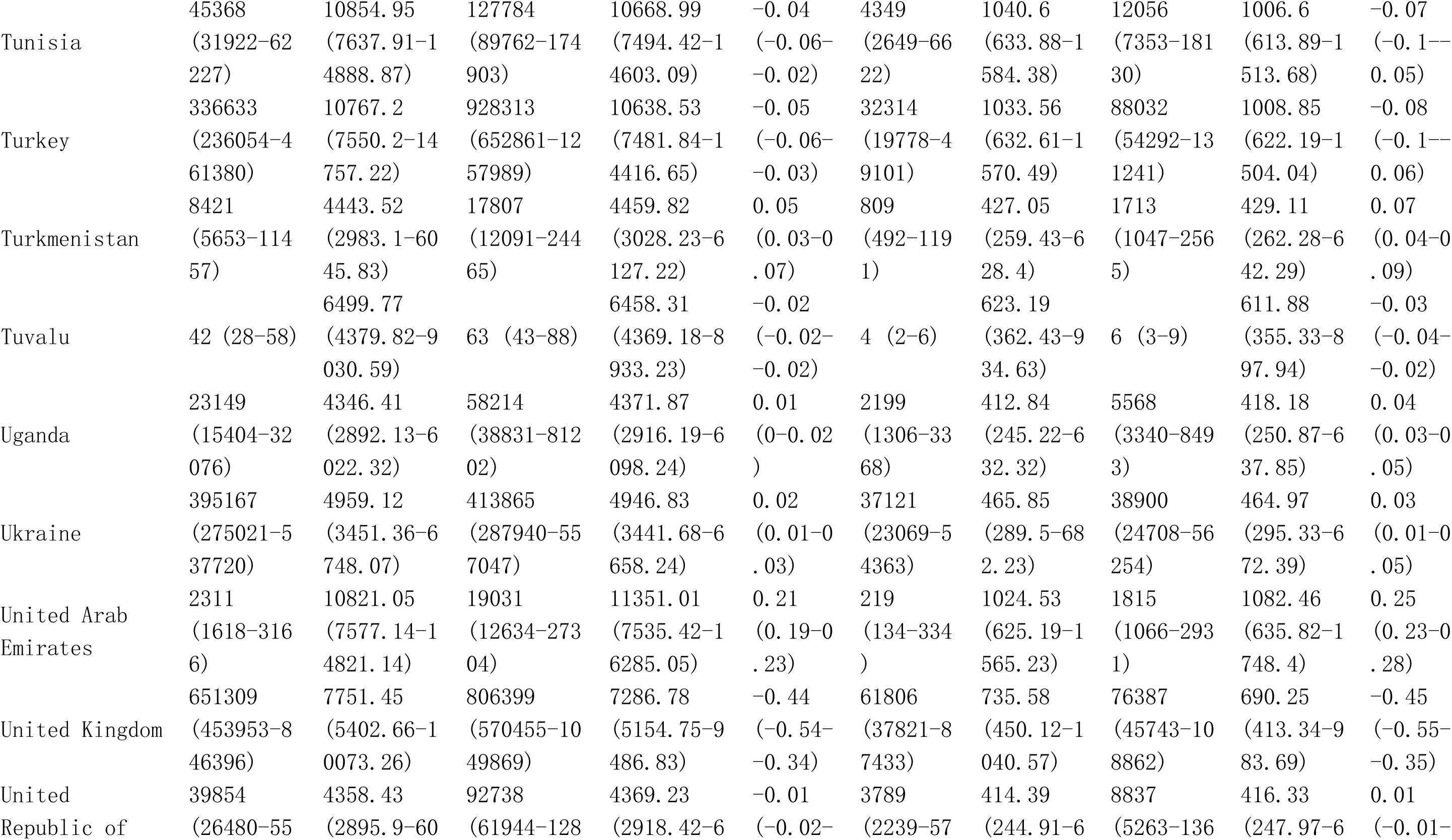
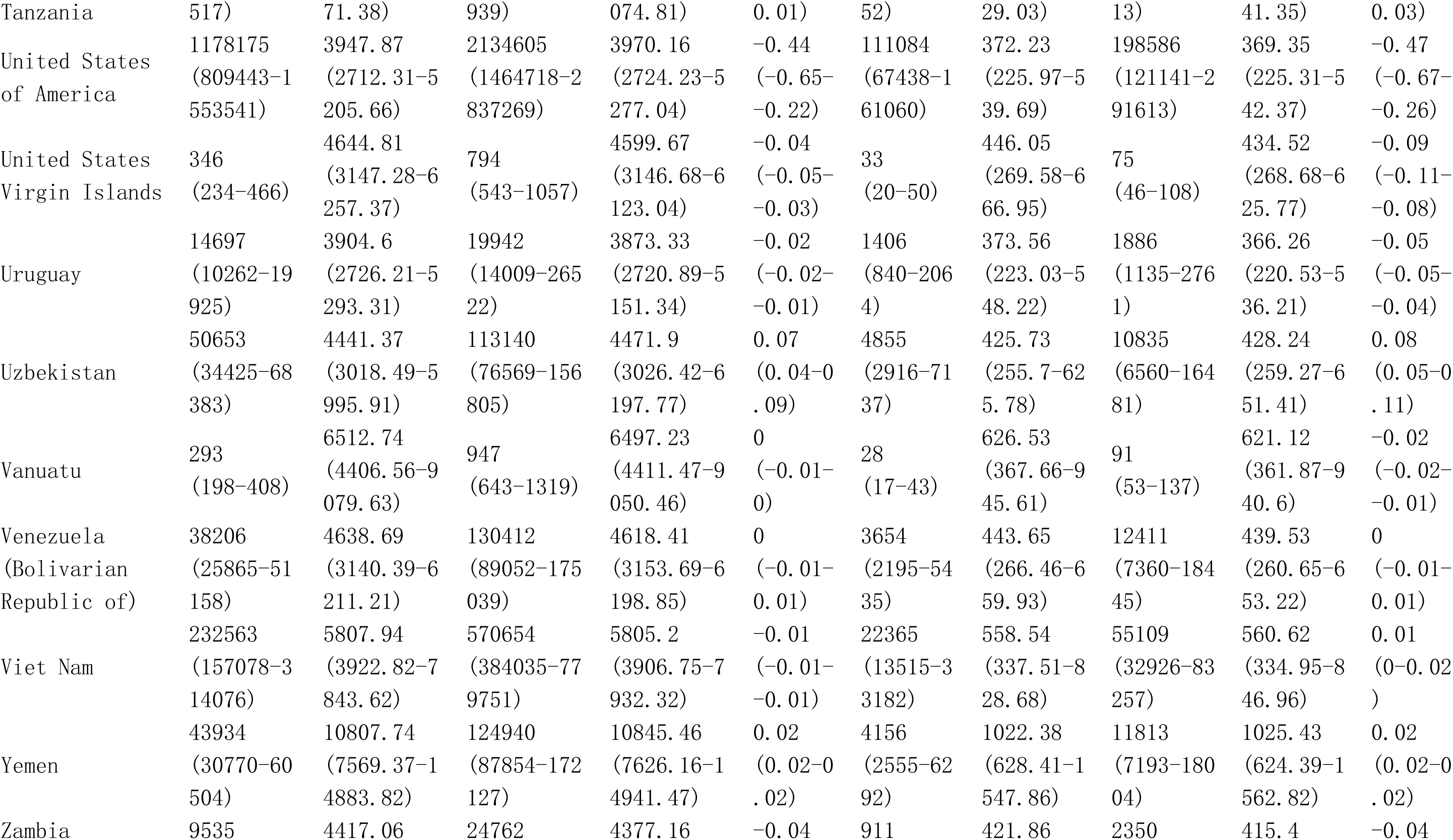
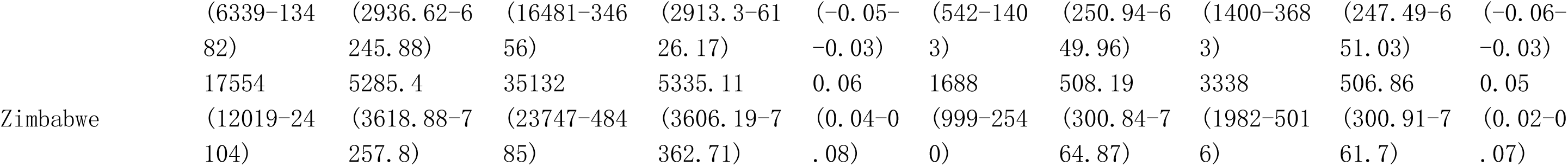
Global burden in Prevalence and DALYs of Neck pain among postmenopausal women from 1990 to 2021 by 21 GBD geographical regions, and 204 countries and territories.

**Table S10.**
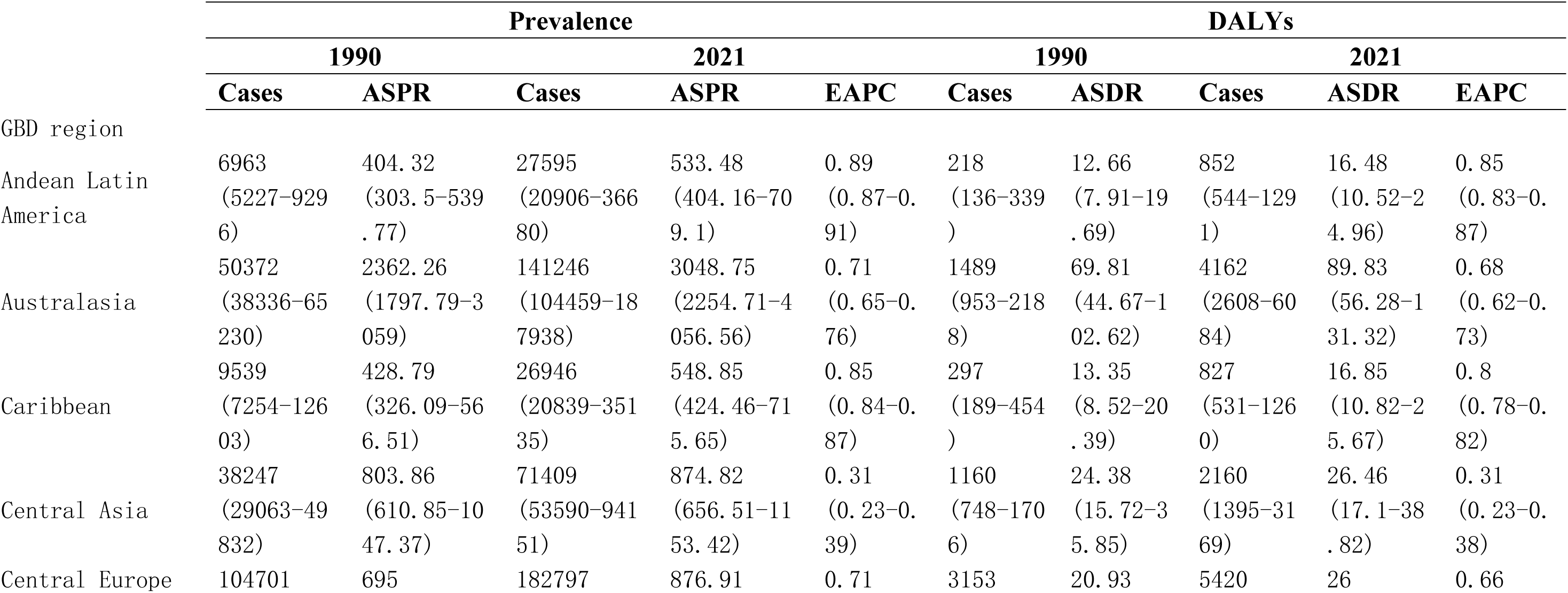
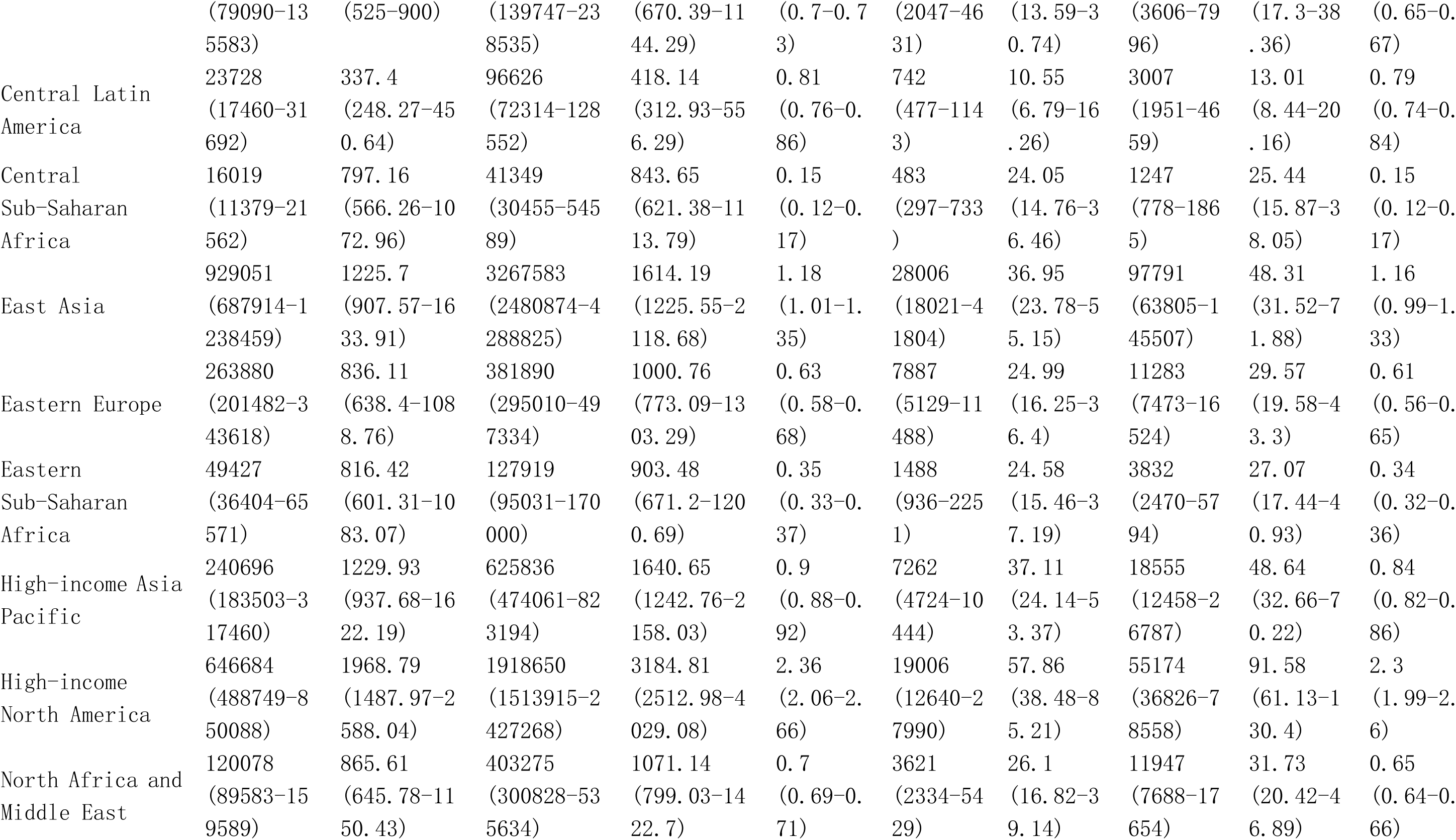
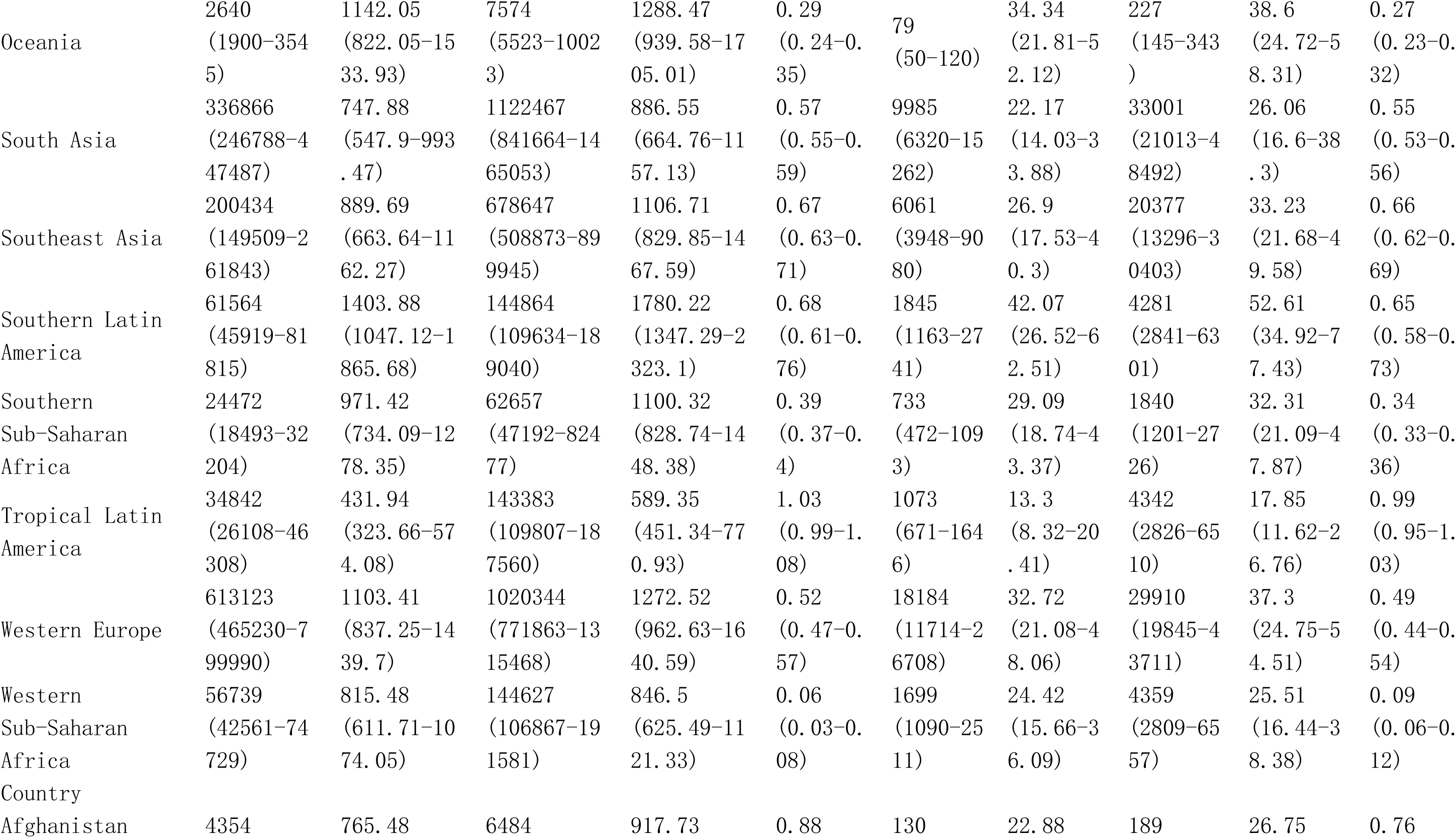
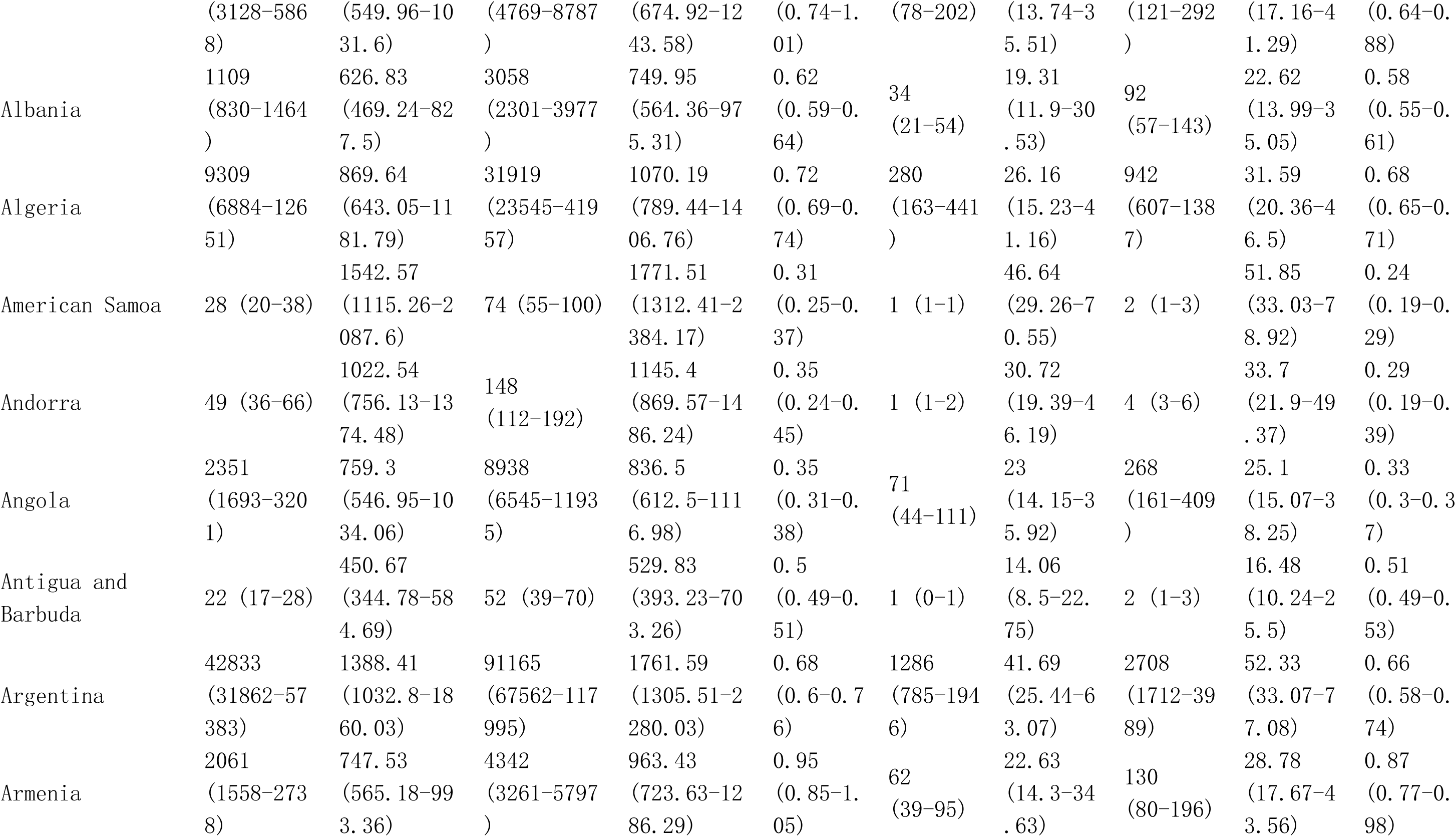
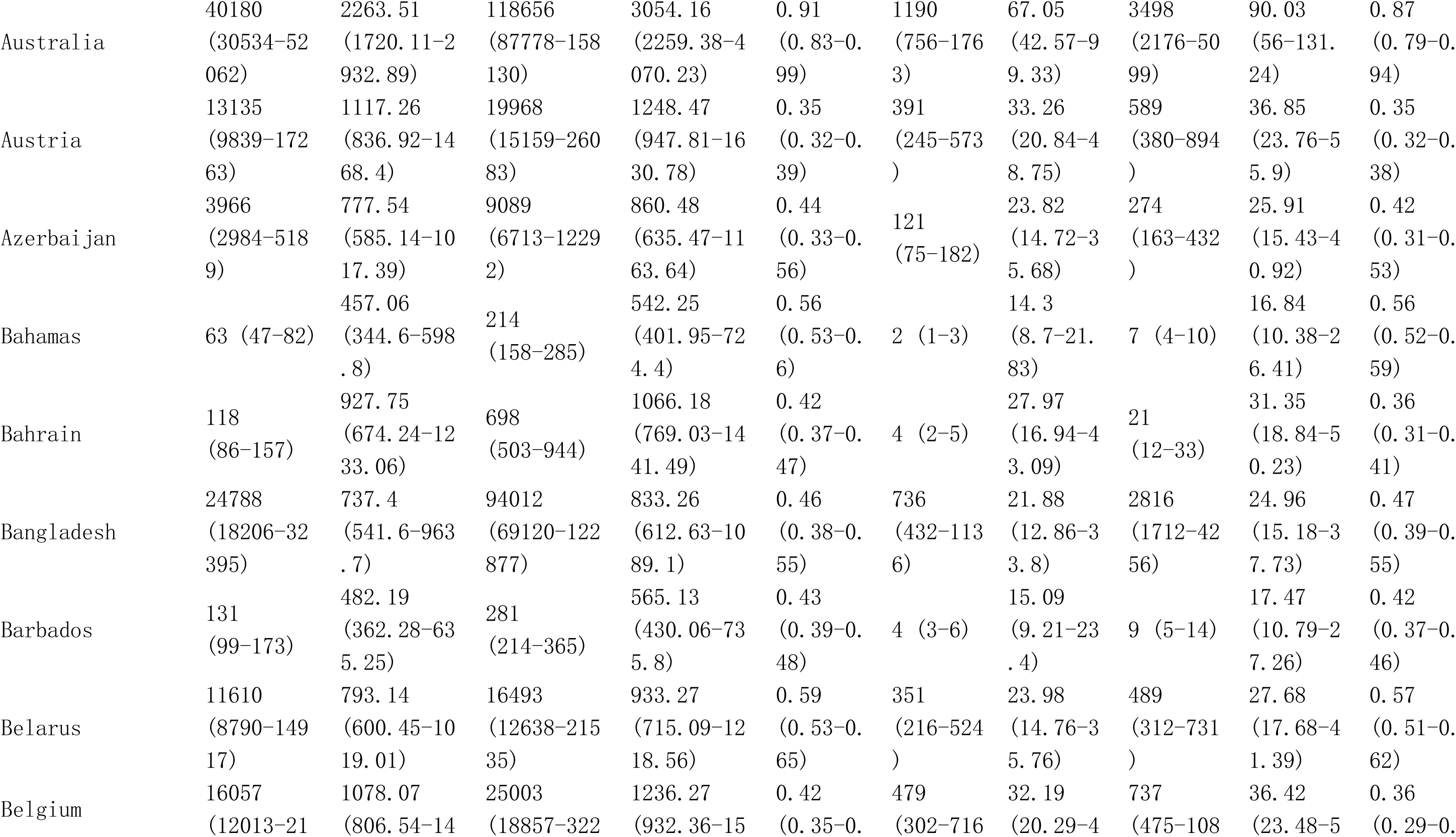
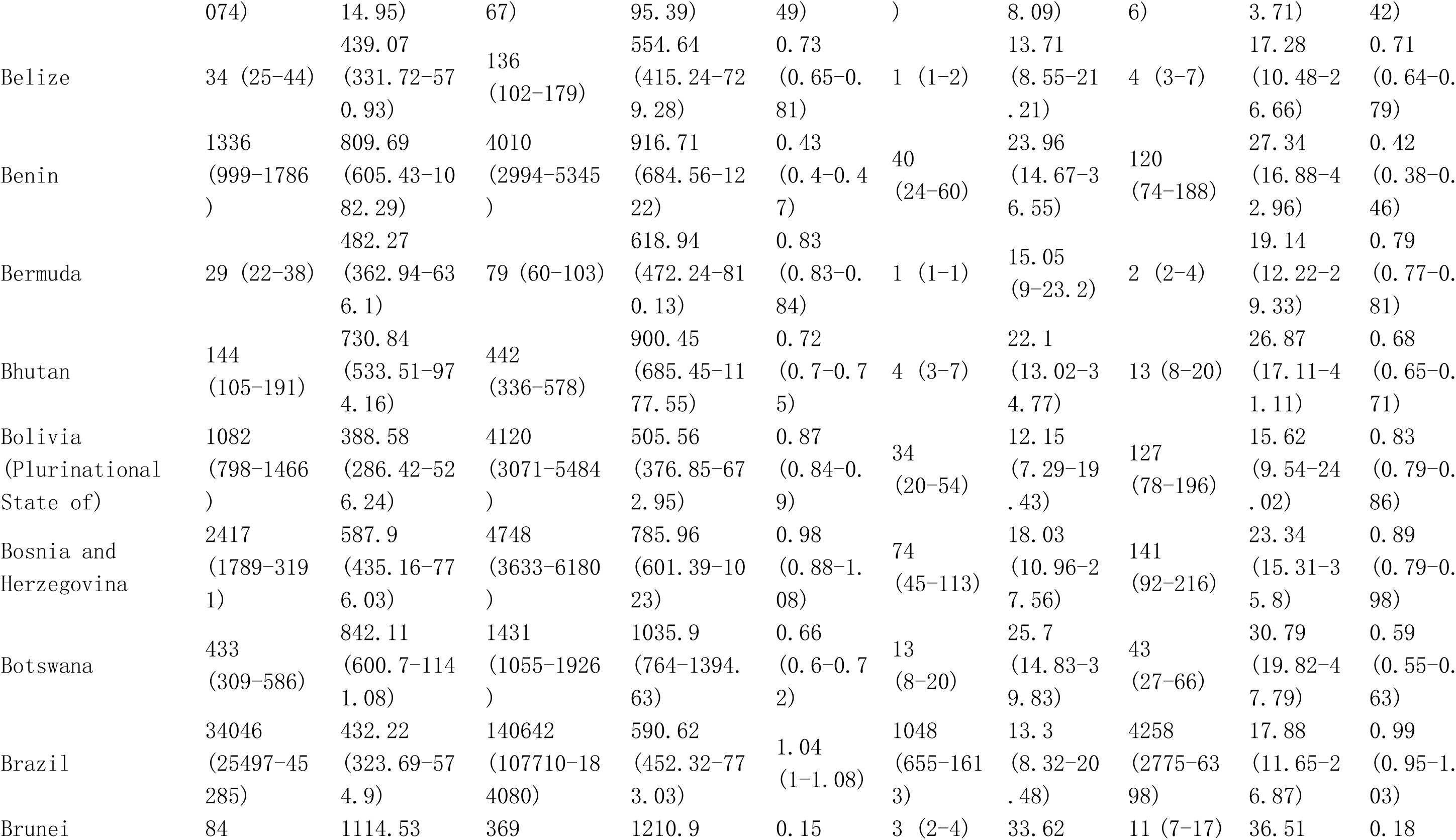
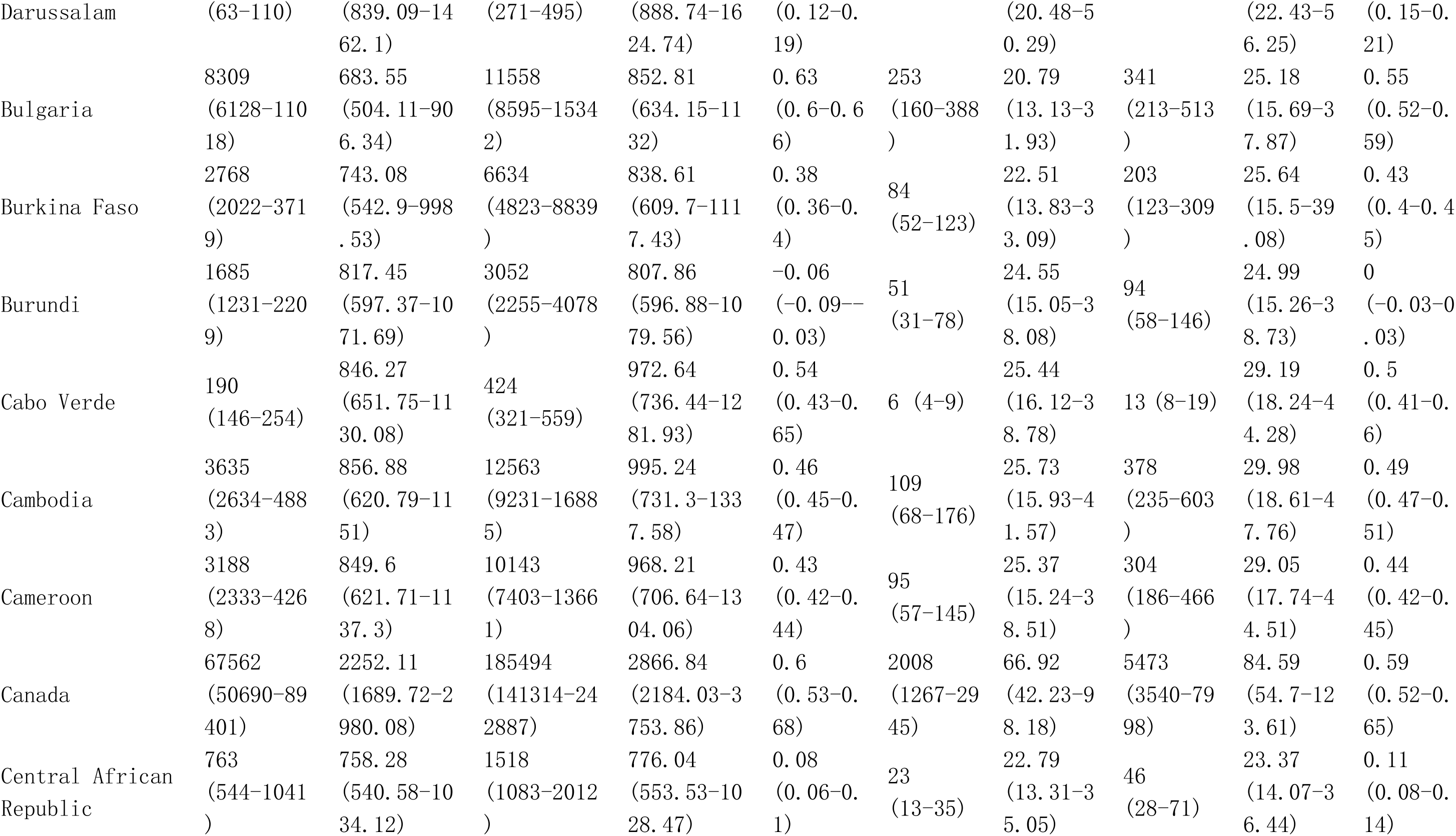
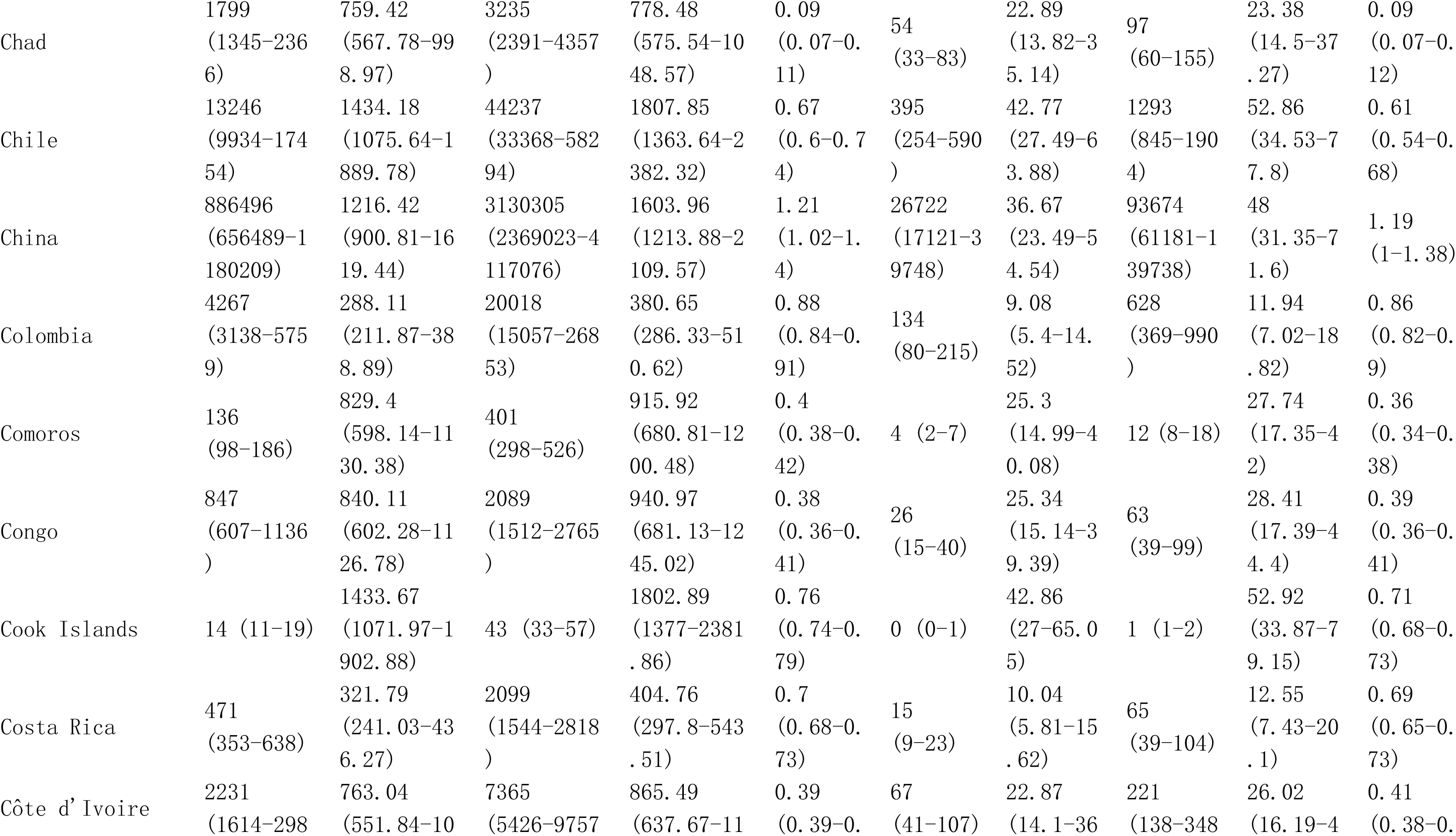
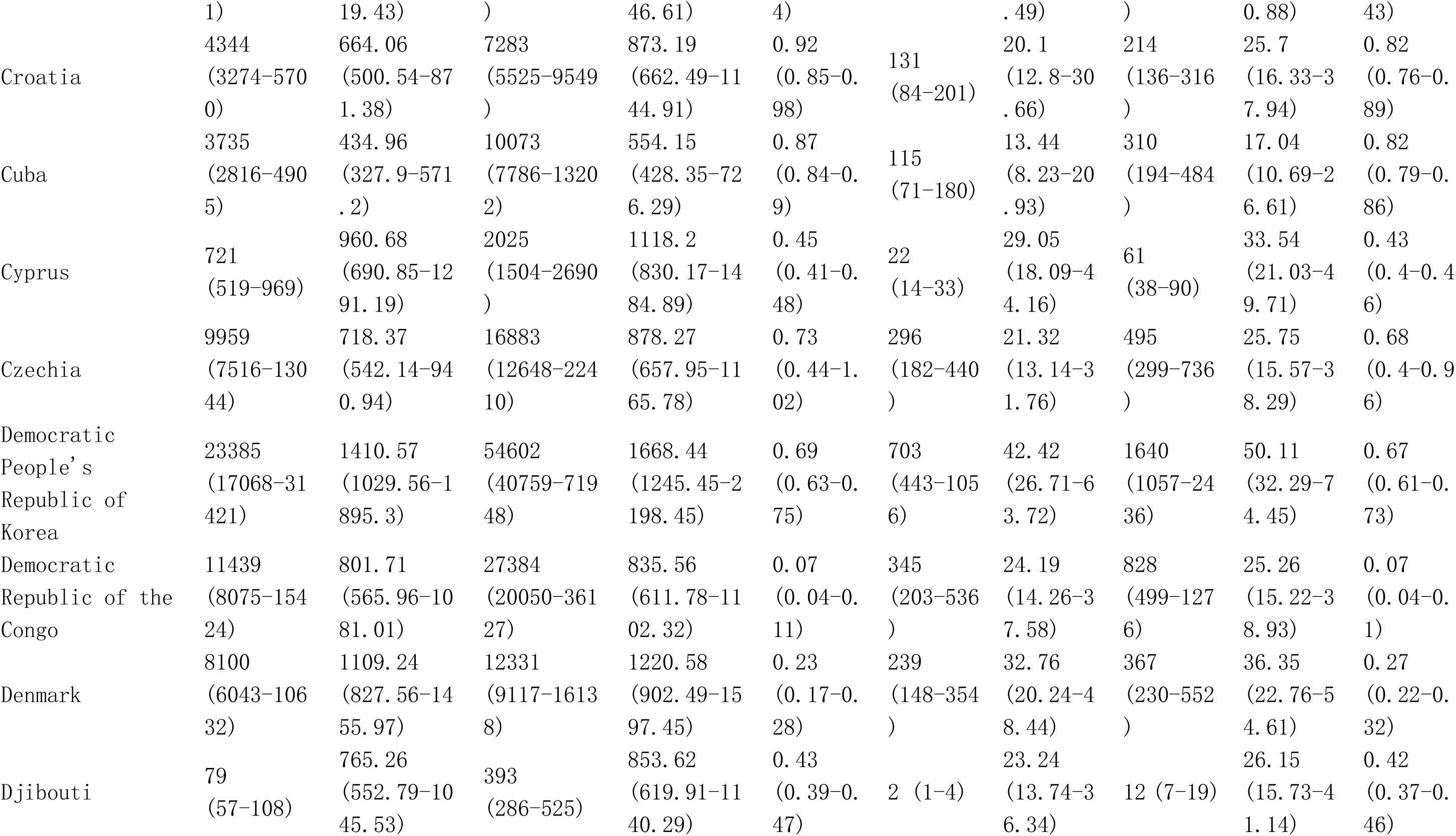
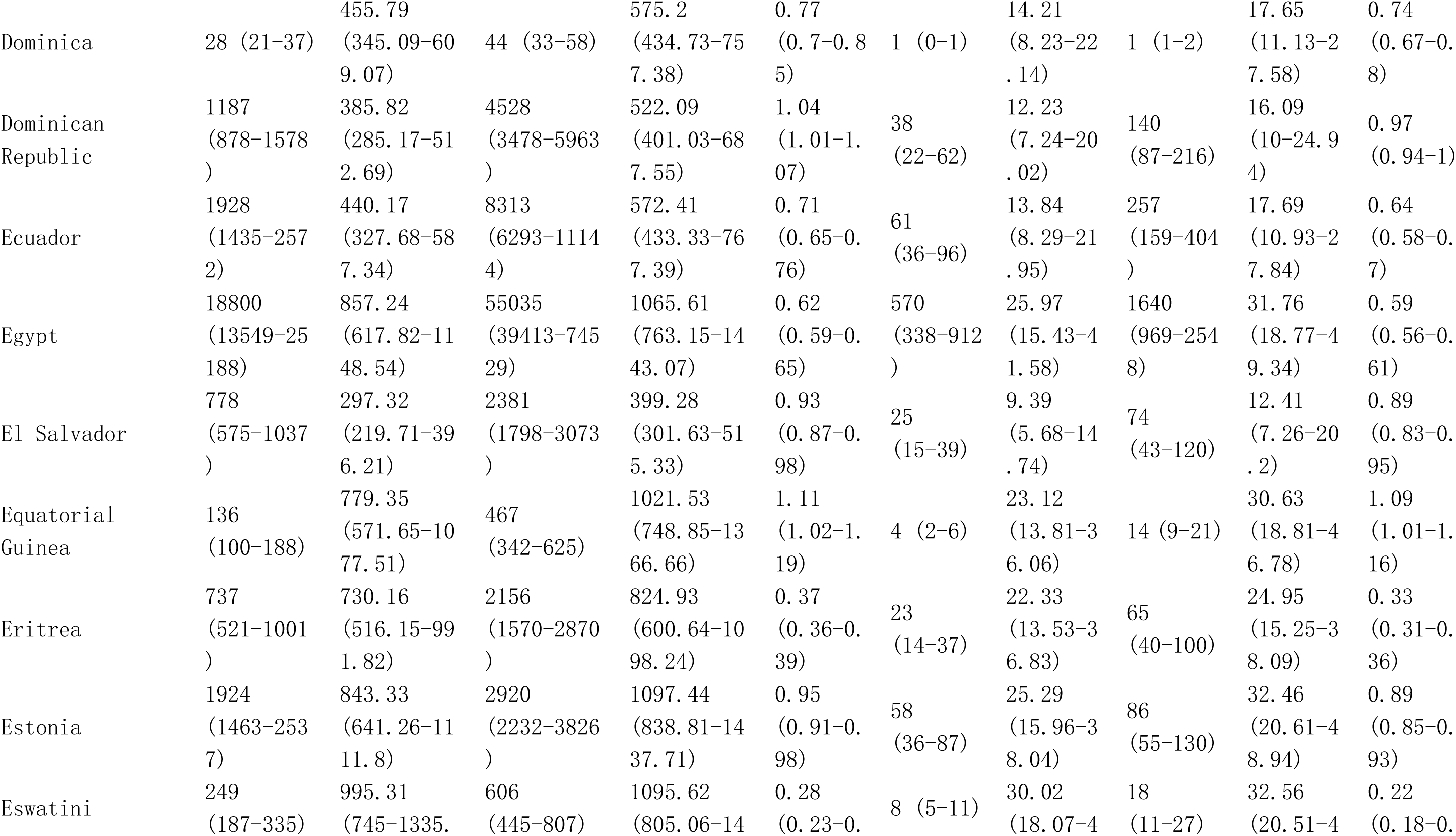
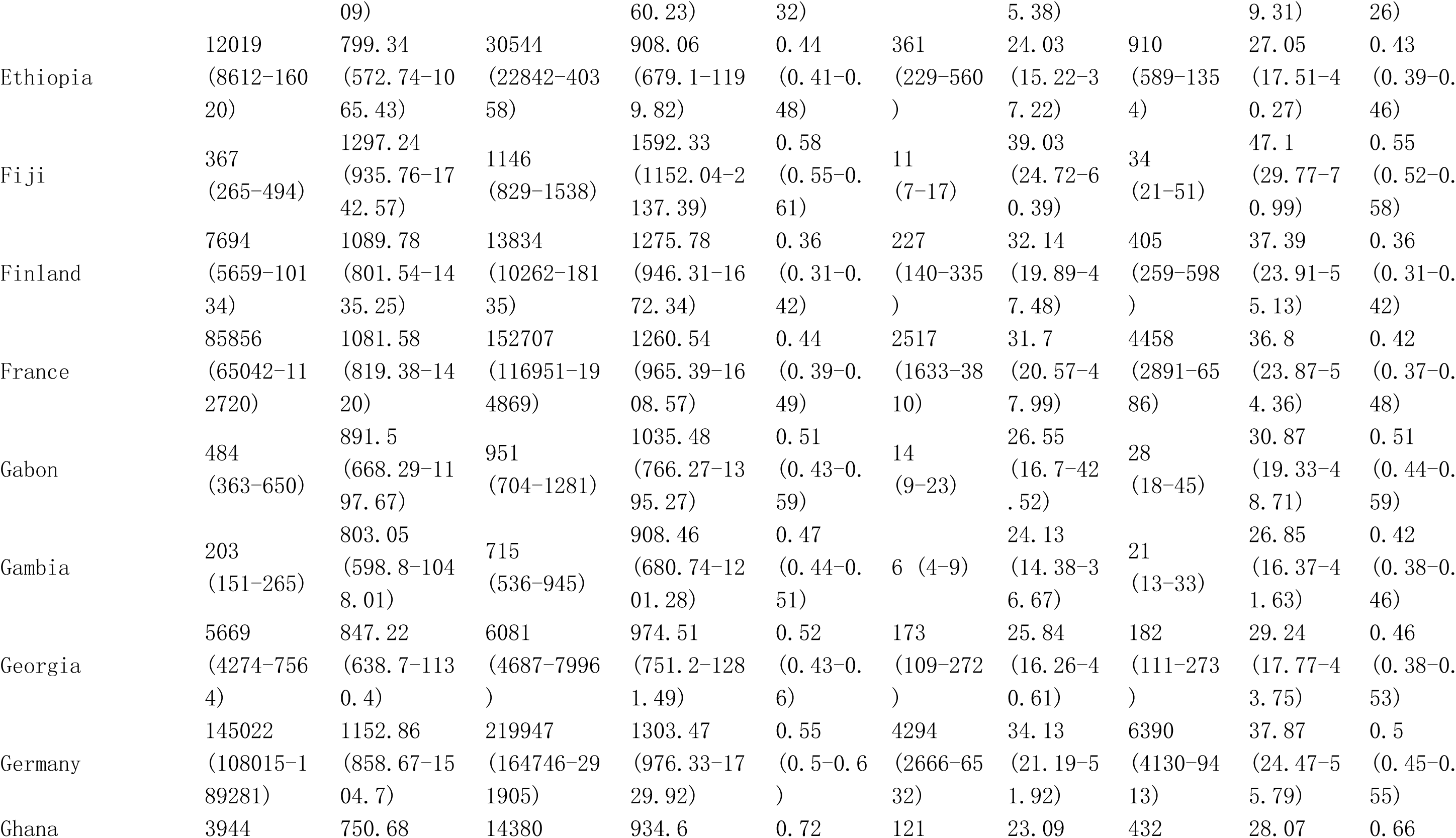
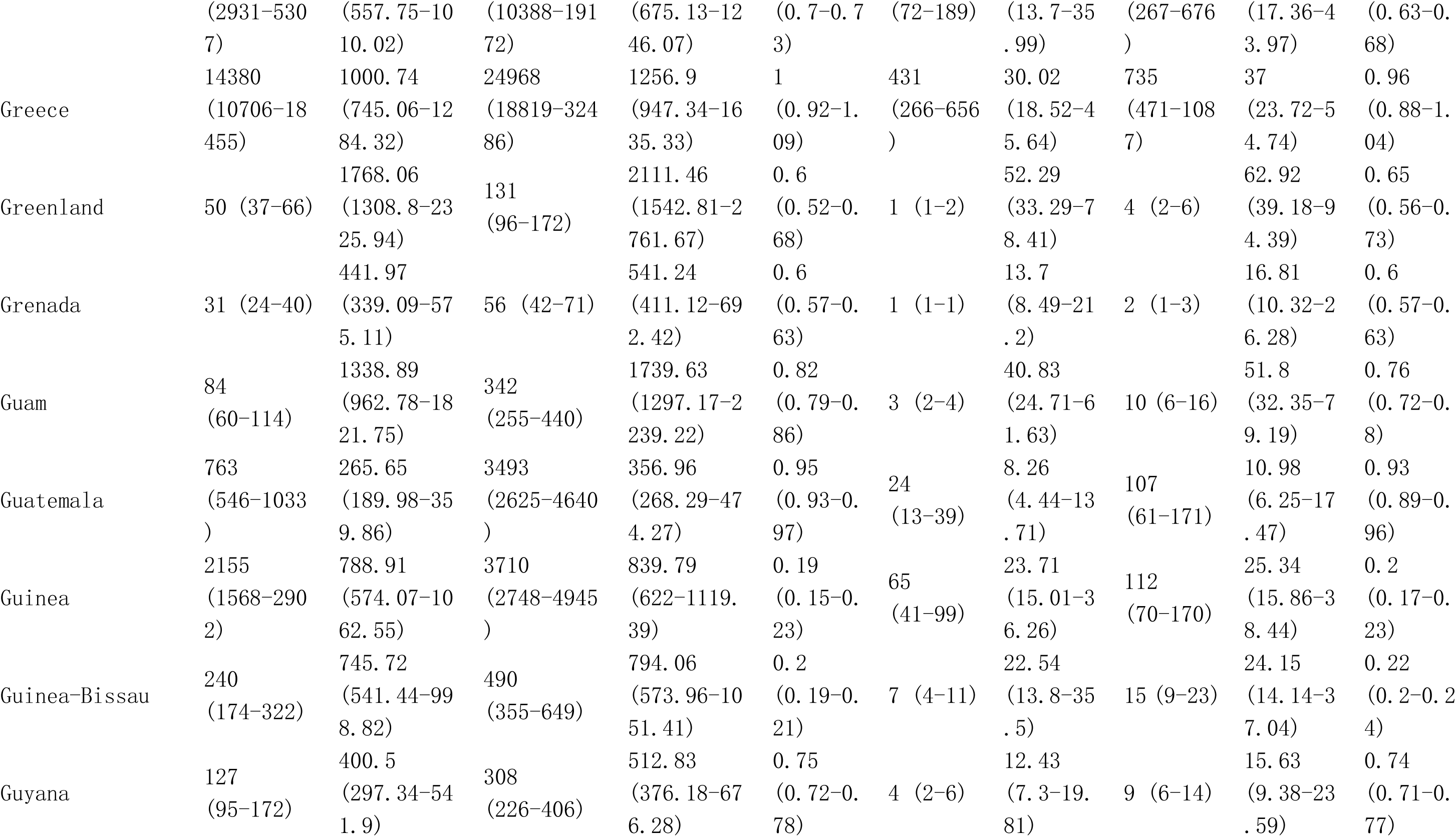
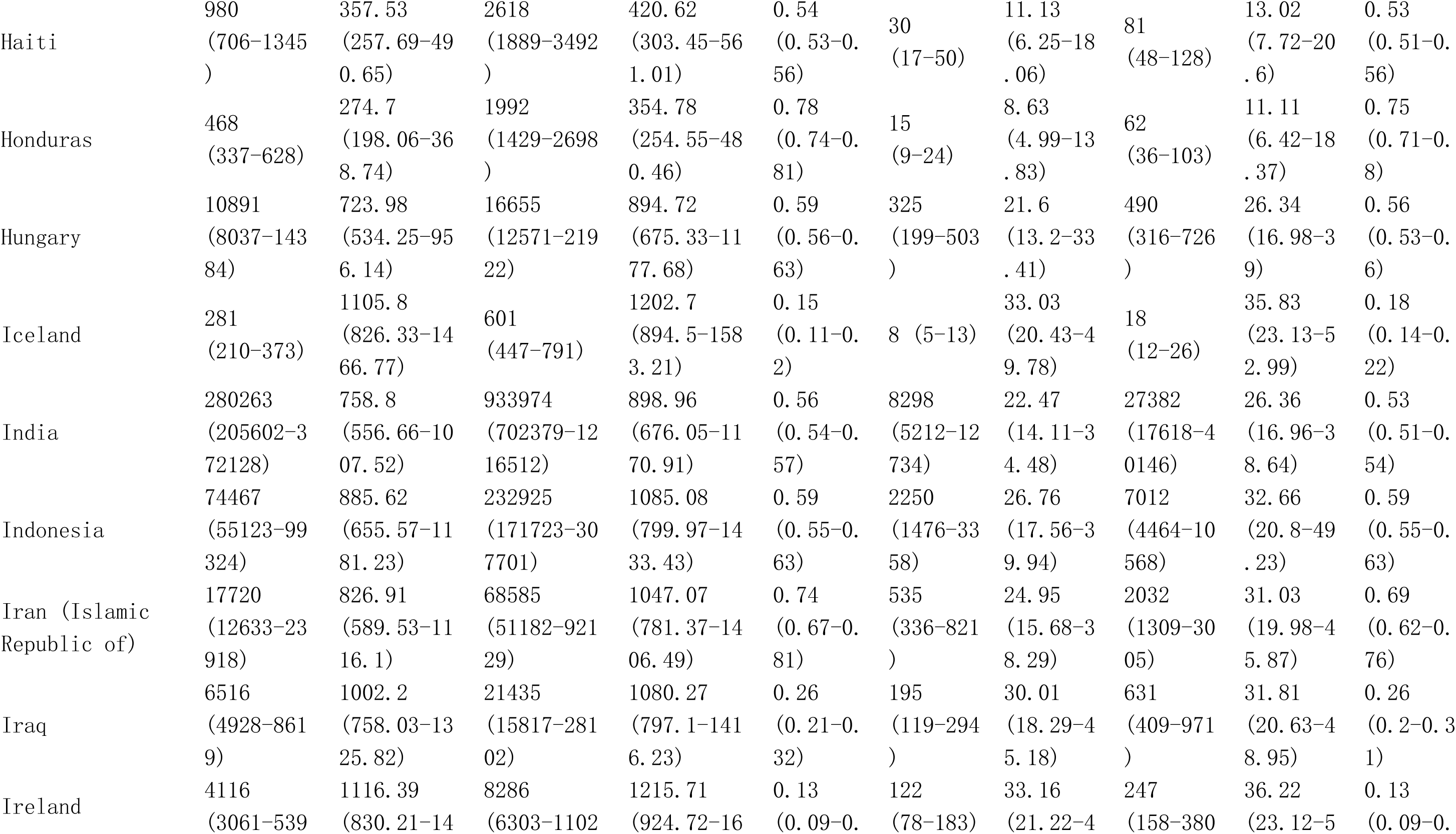
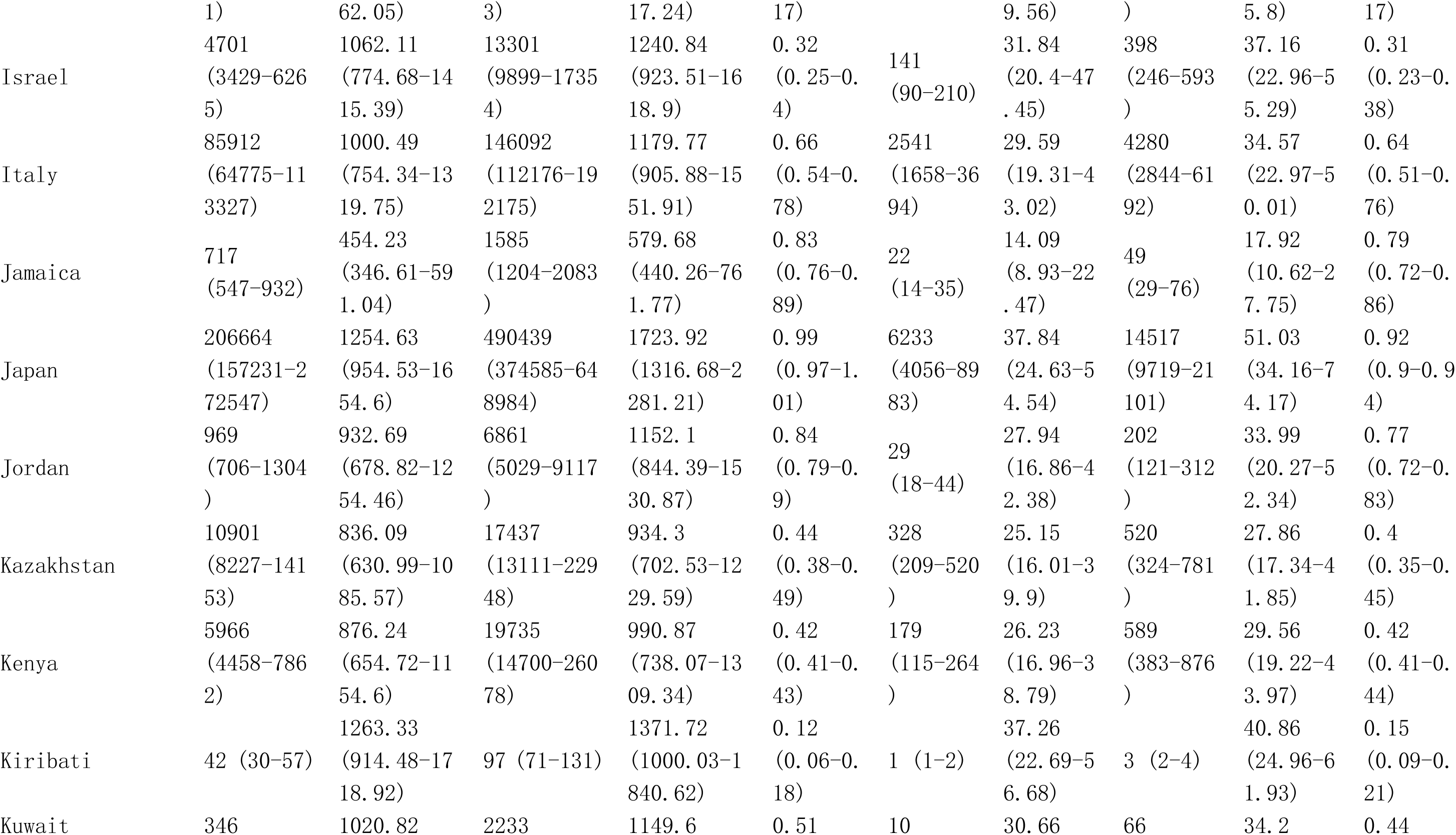
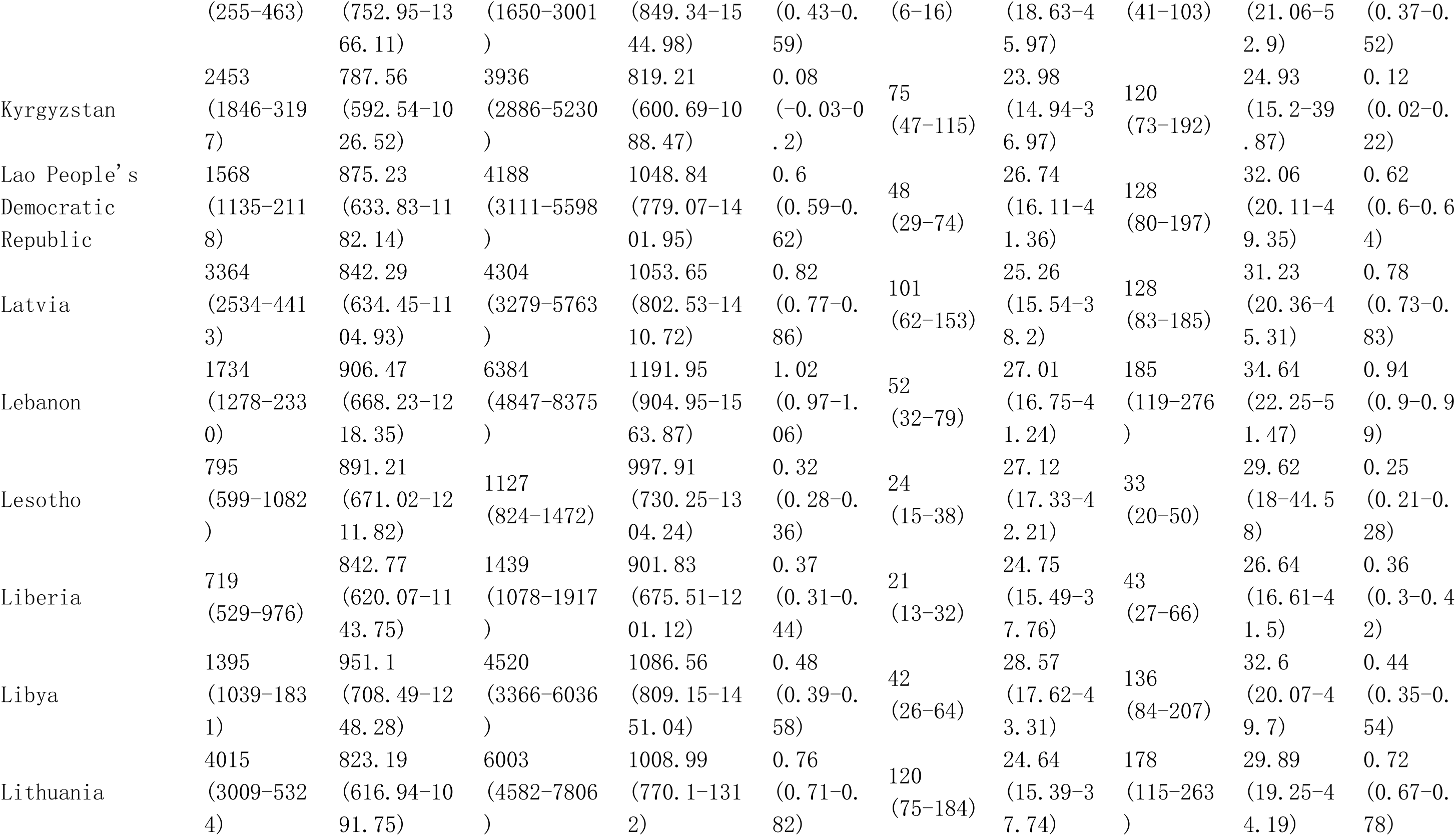
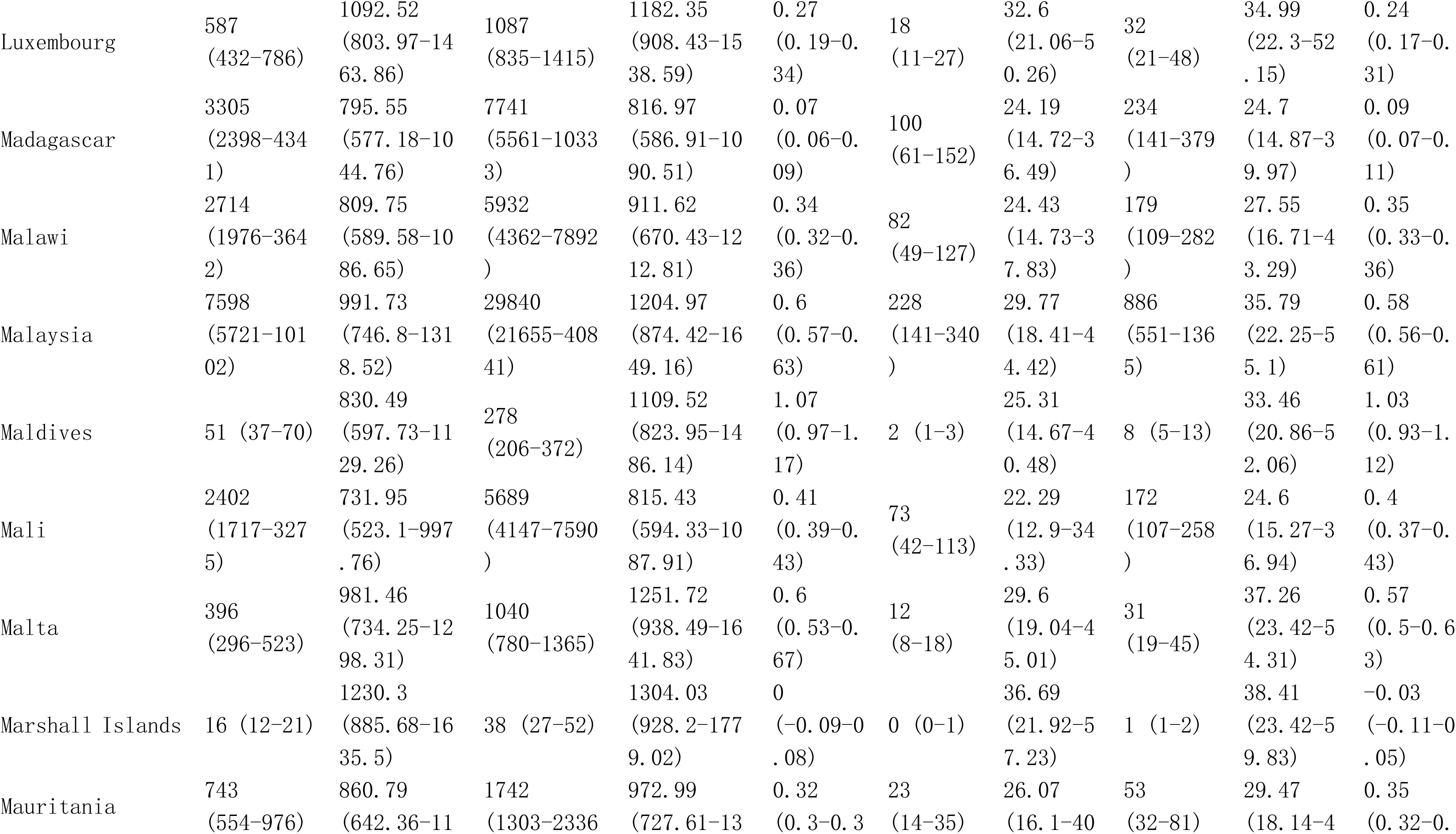
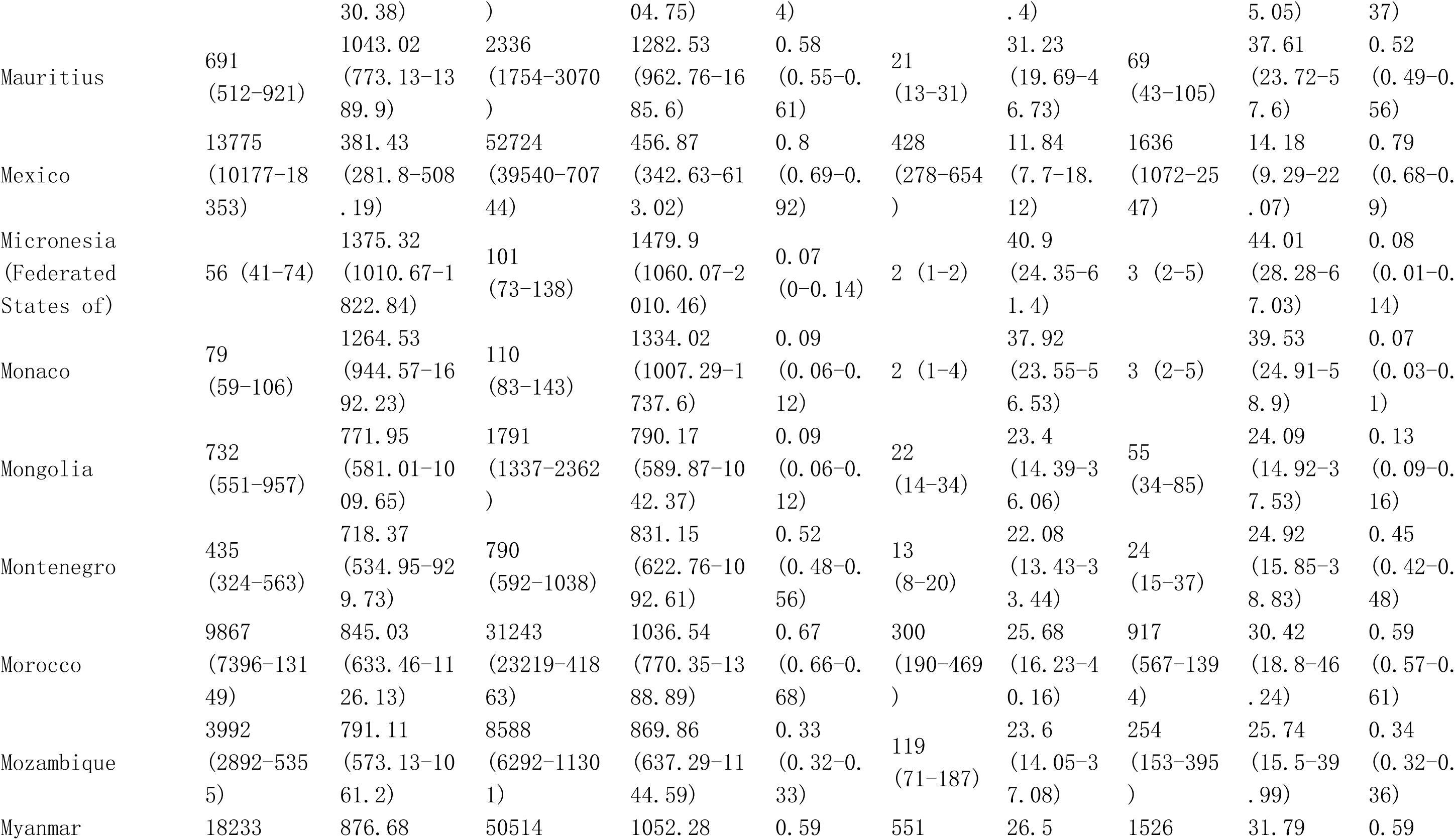
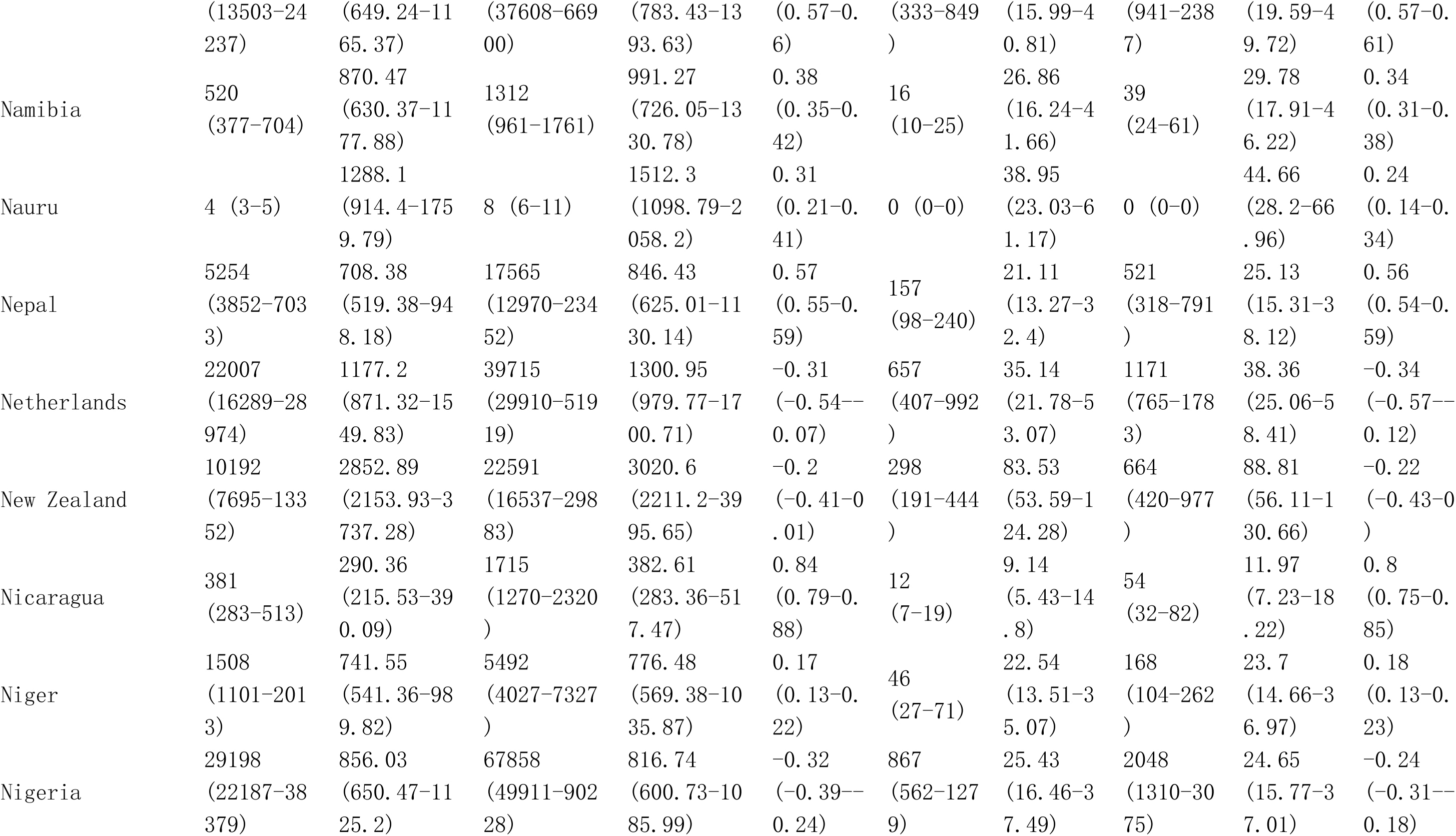
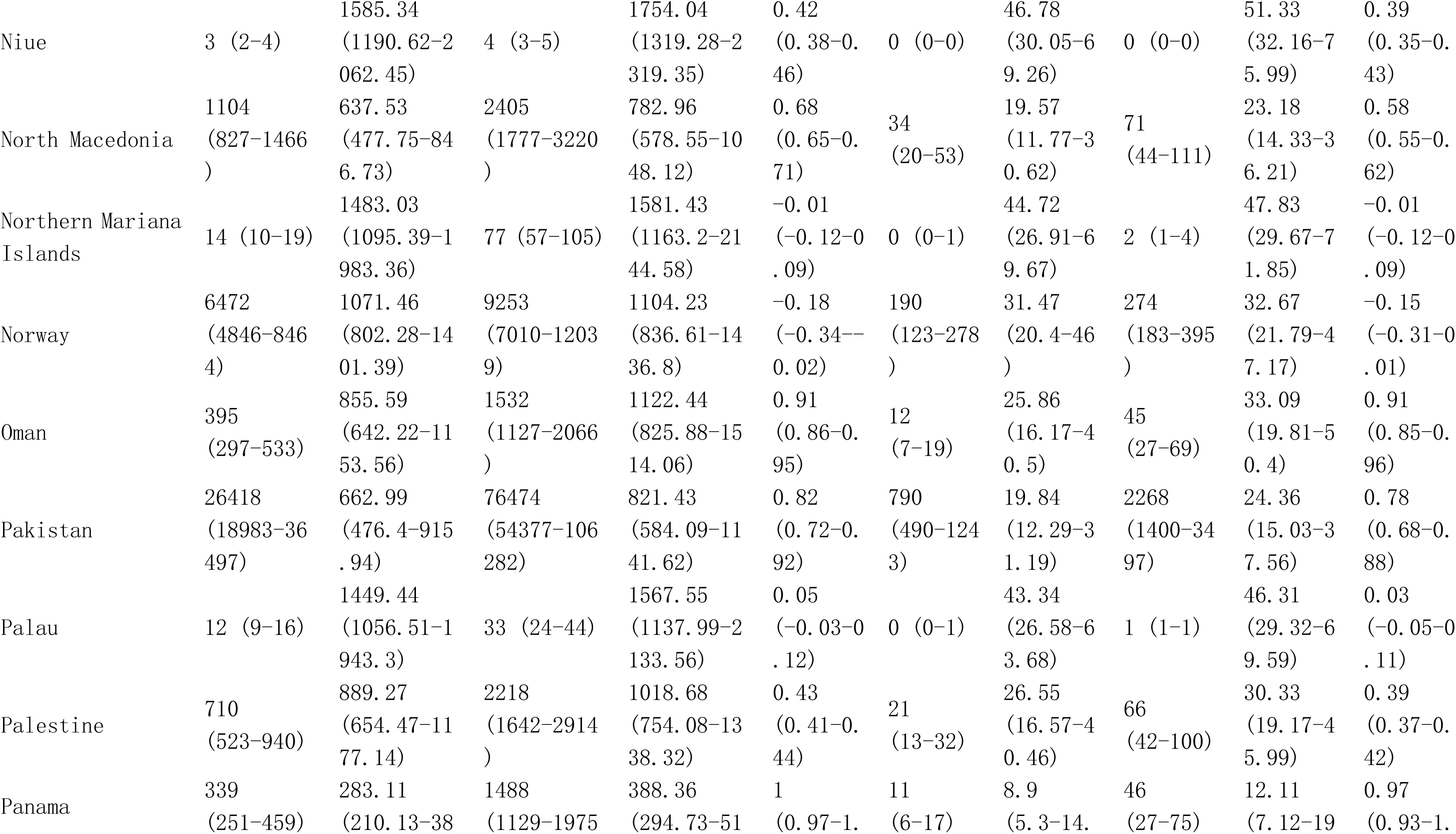
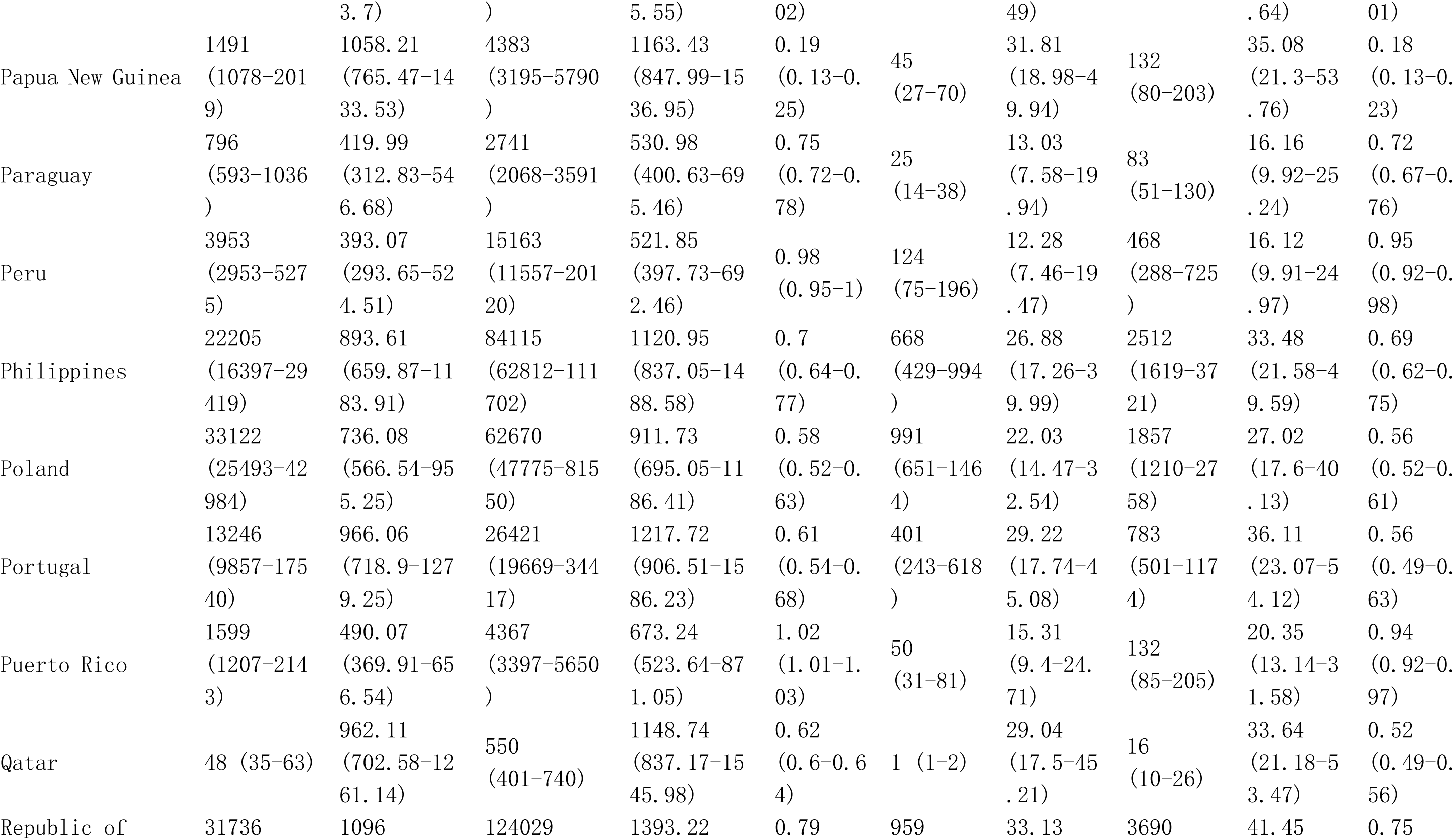
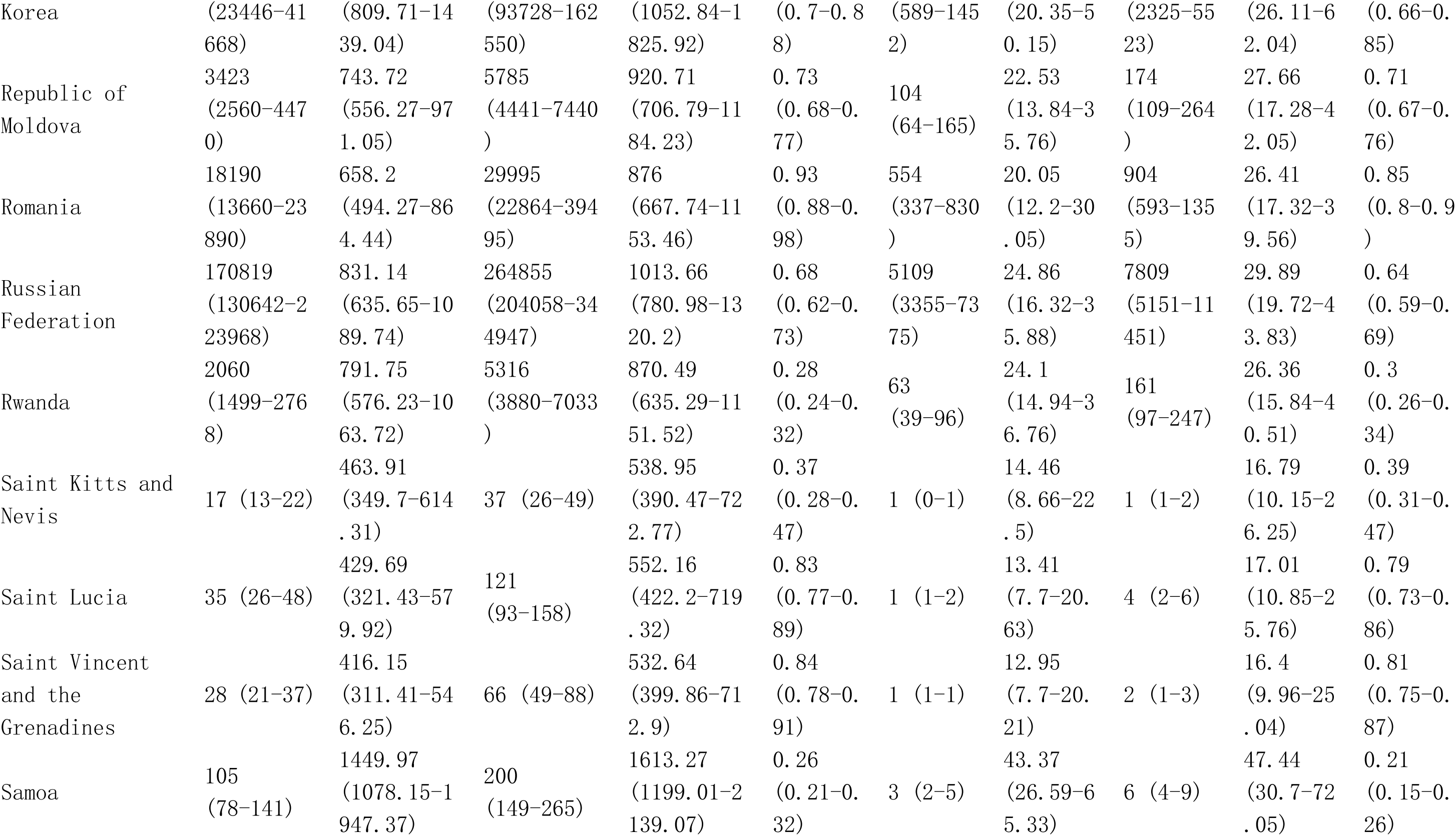
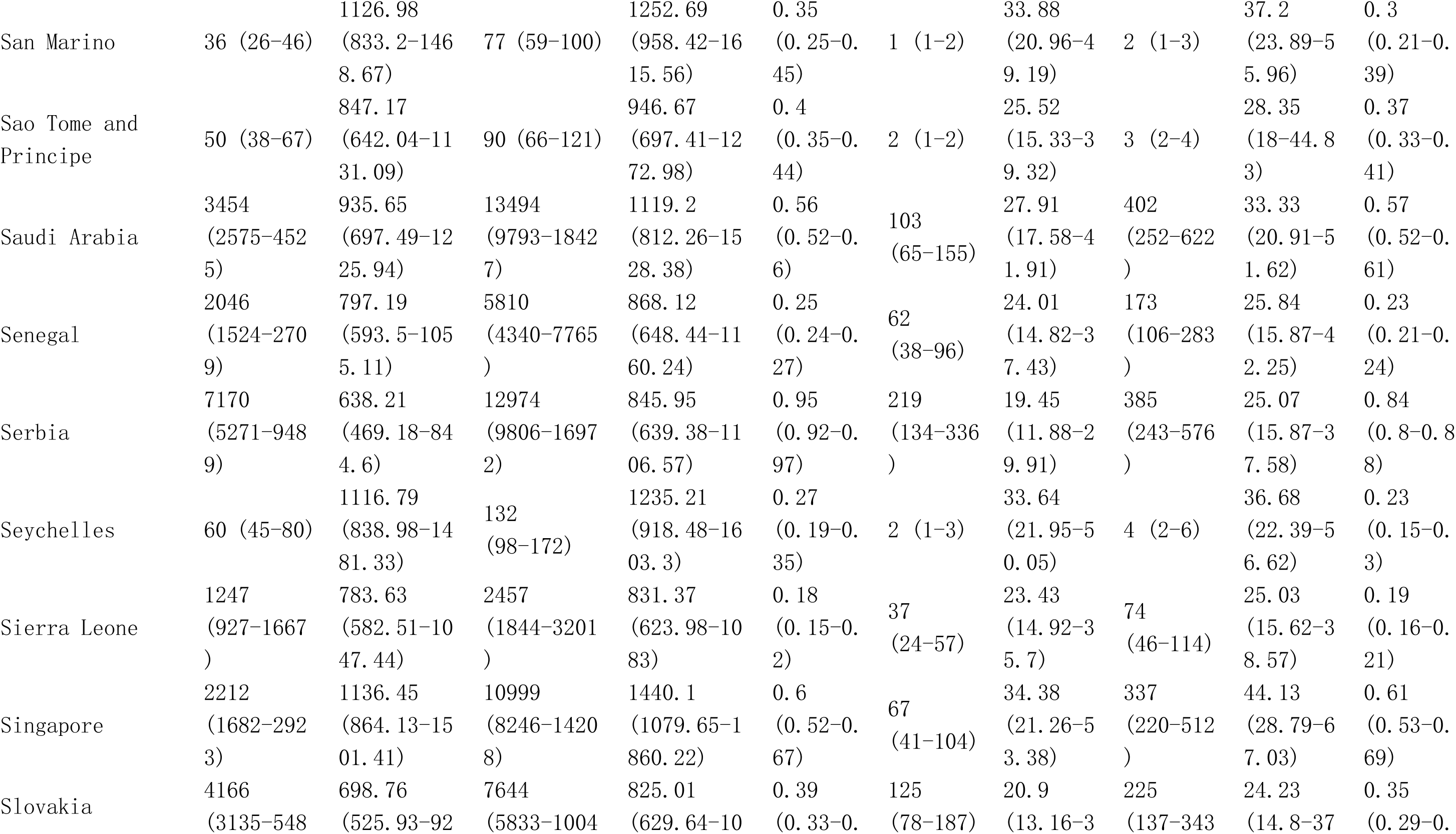
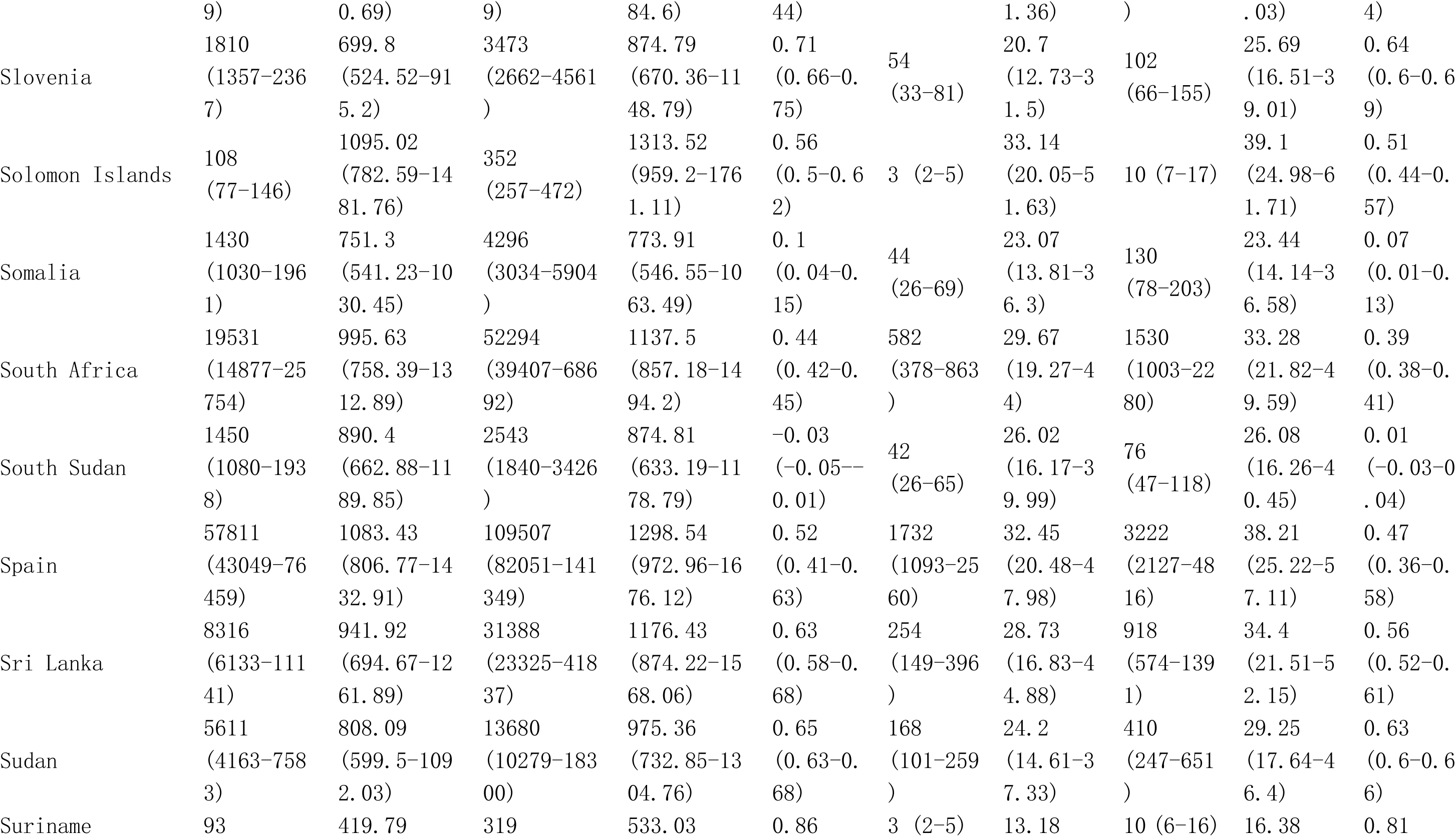
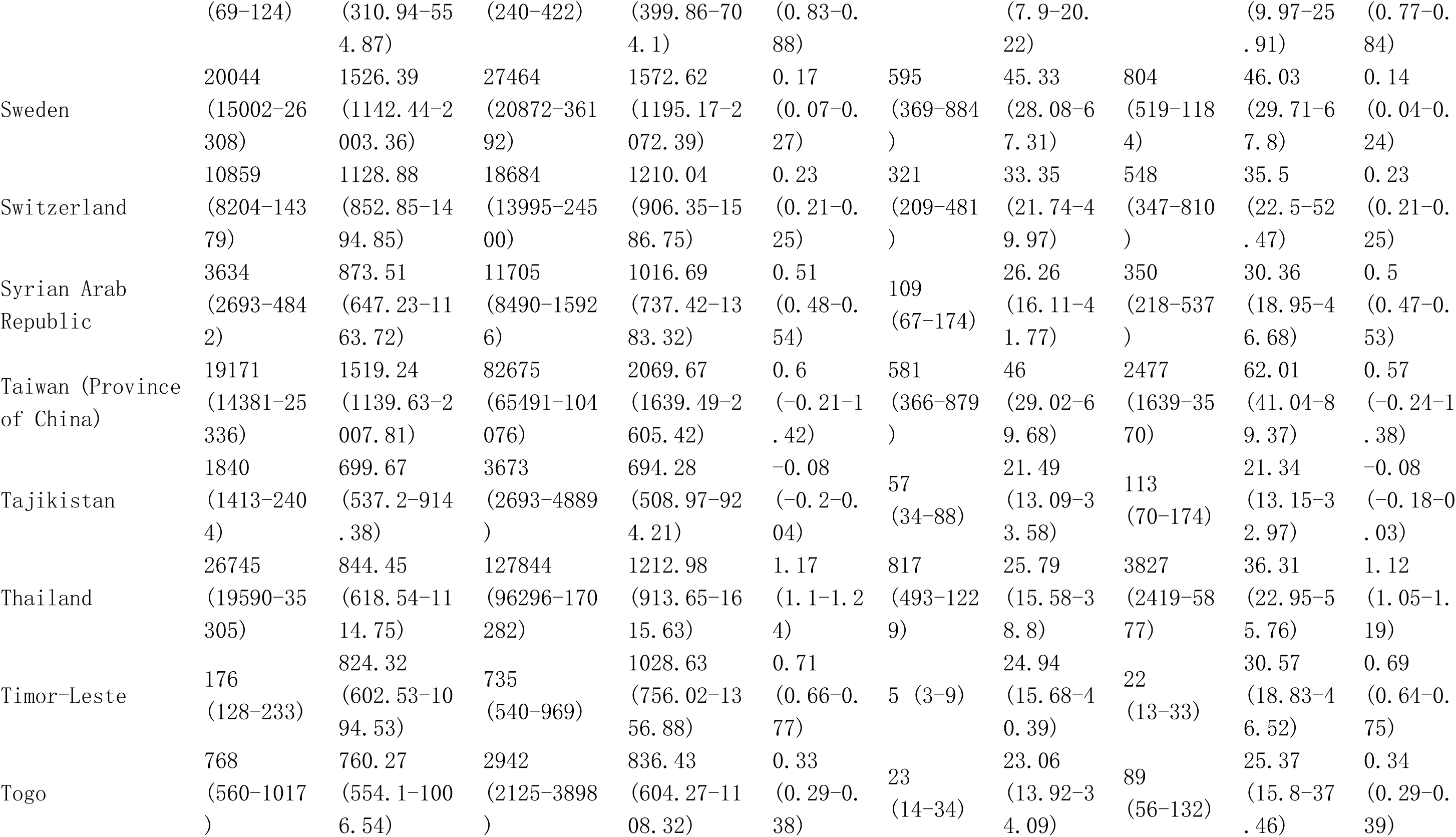
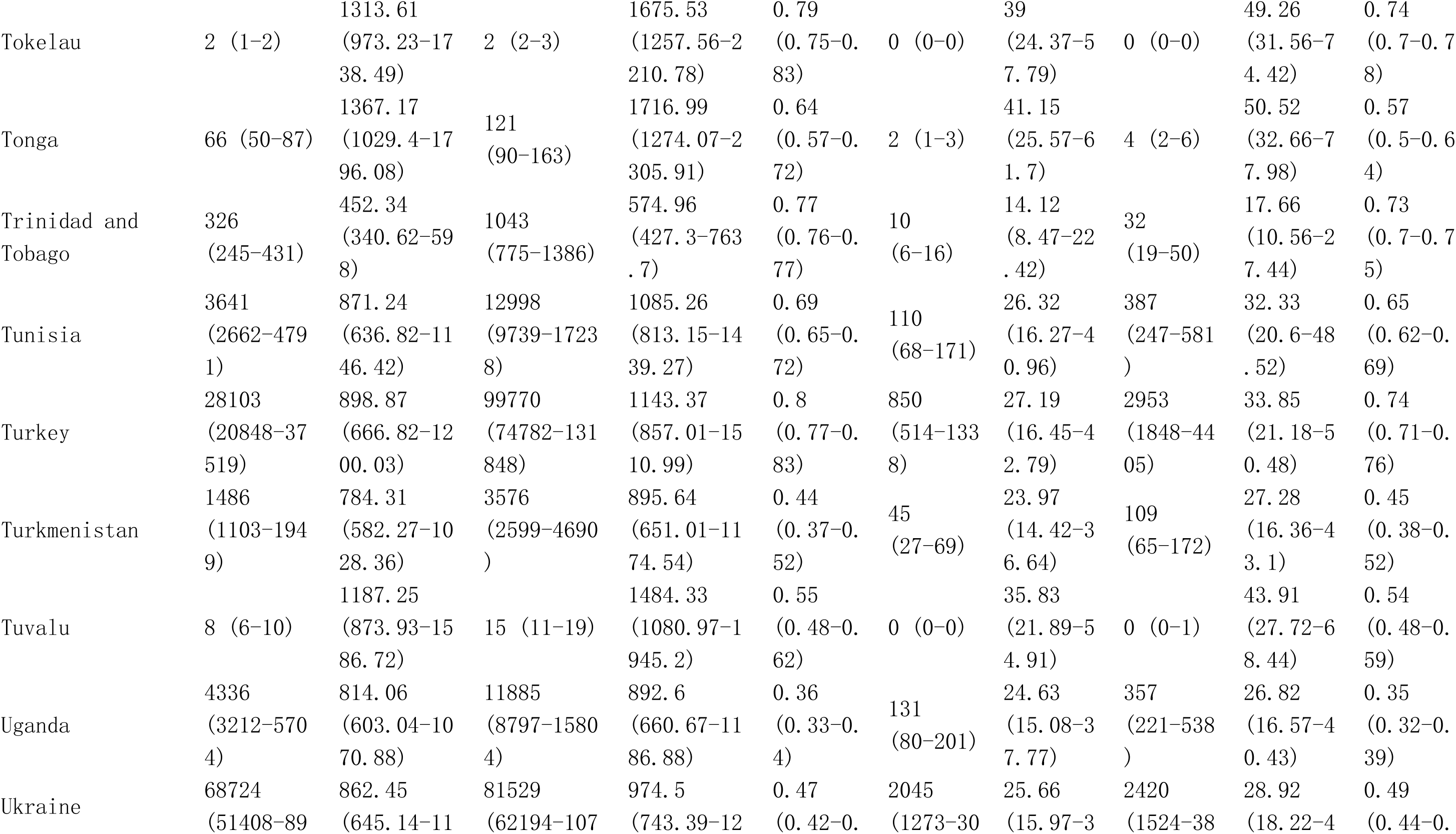
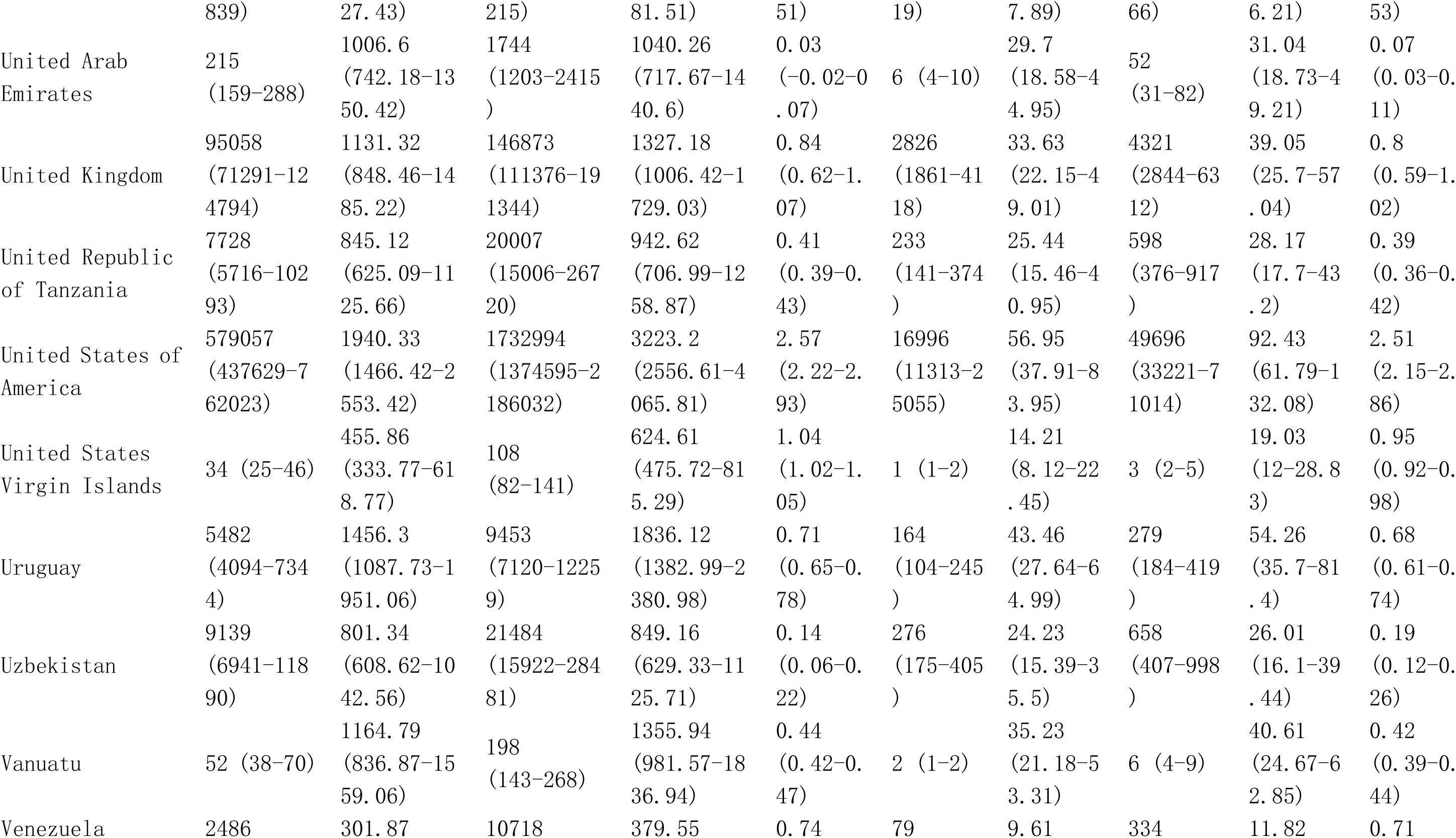
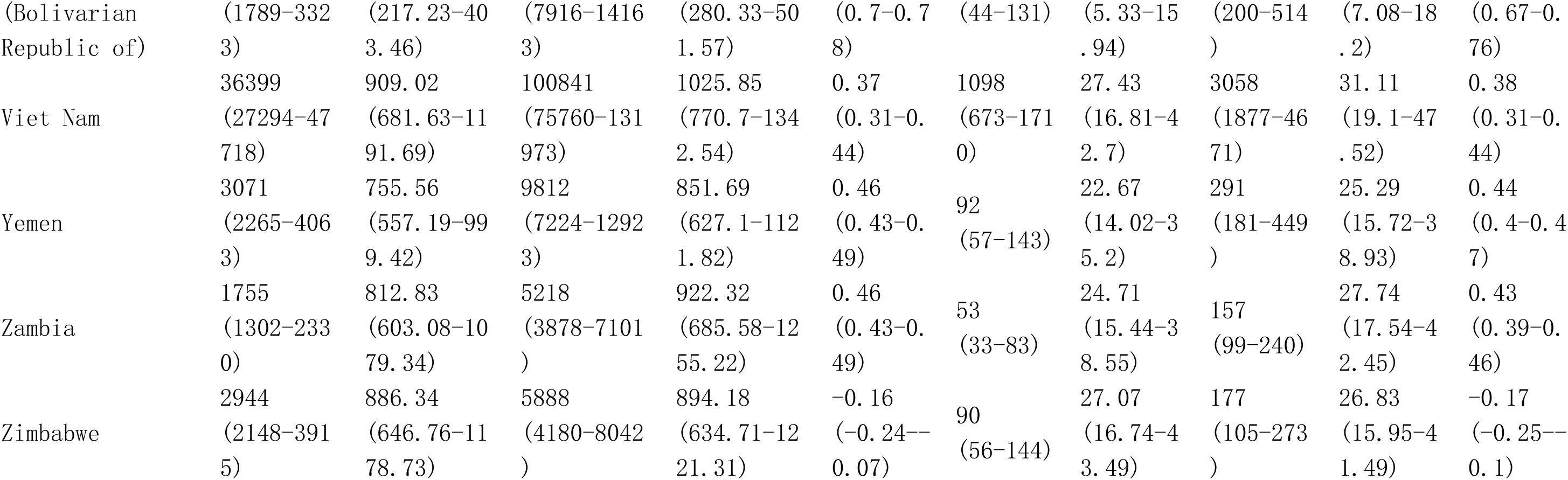
Global burden in Prevalence and DALYs of Gout among postmenopausal women from 1990 to 2021 by 21 GBD geographical regions, and 204 countries and territories.

**Table S11.**
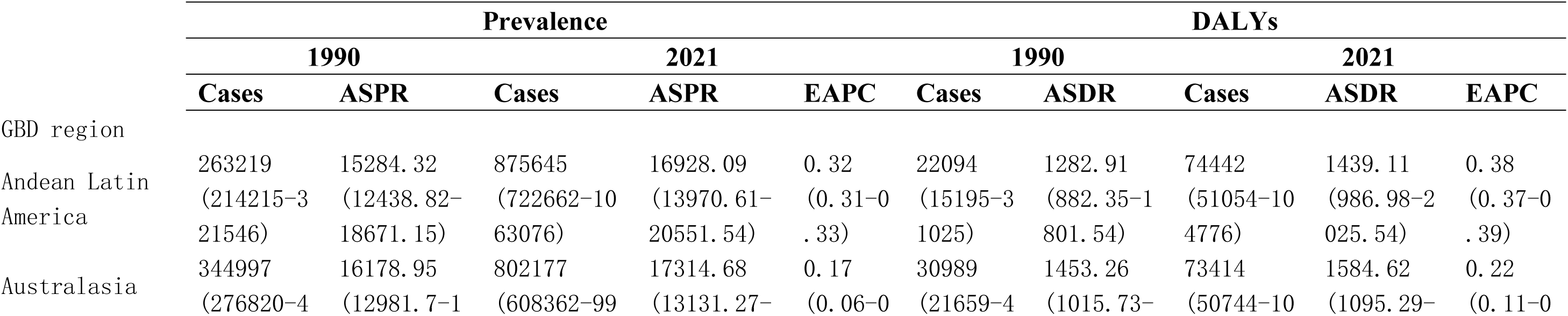
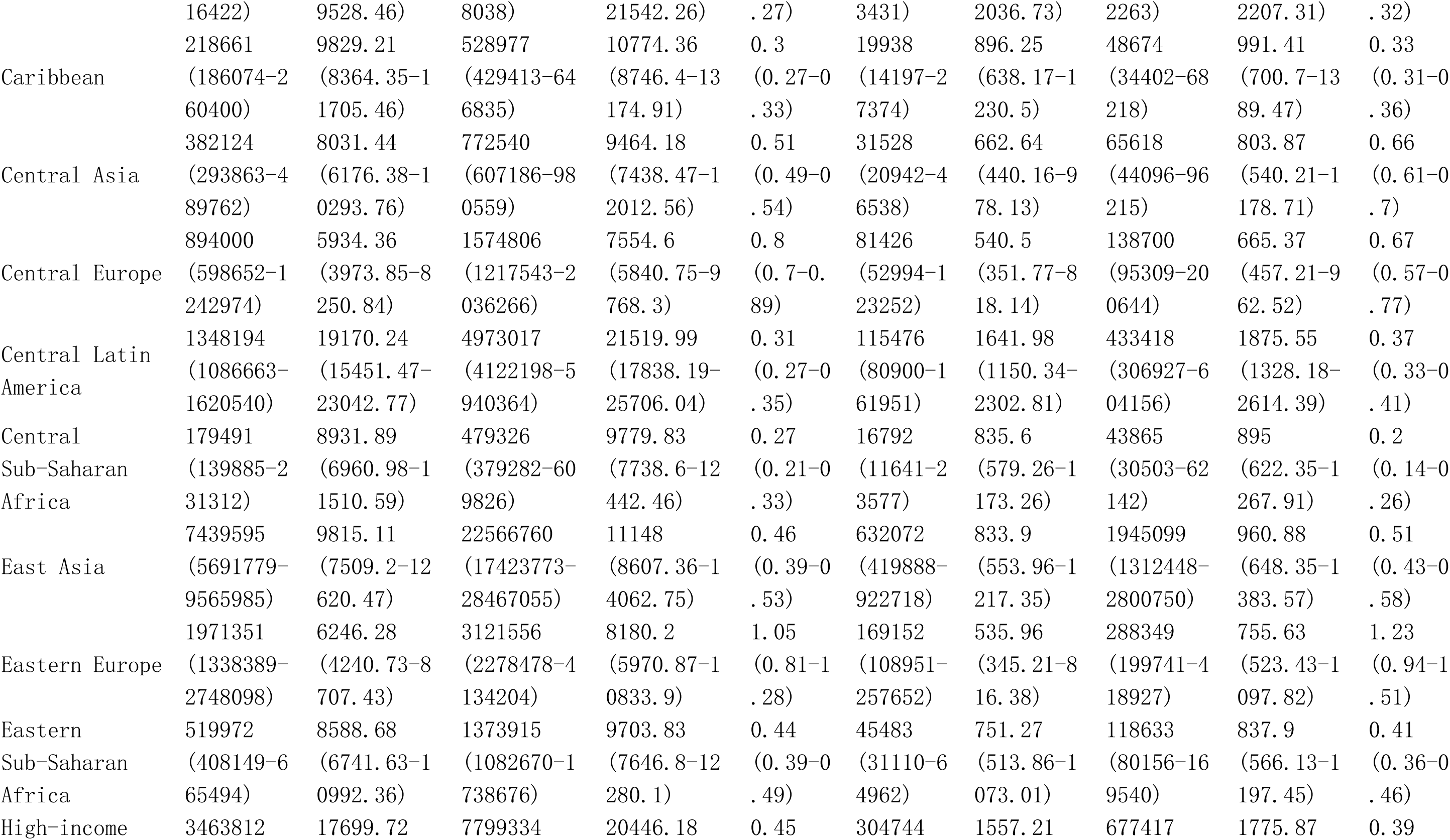
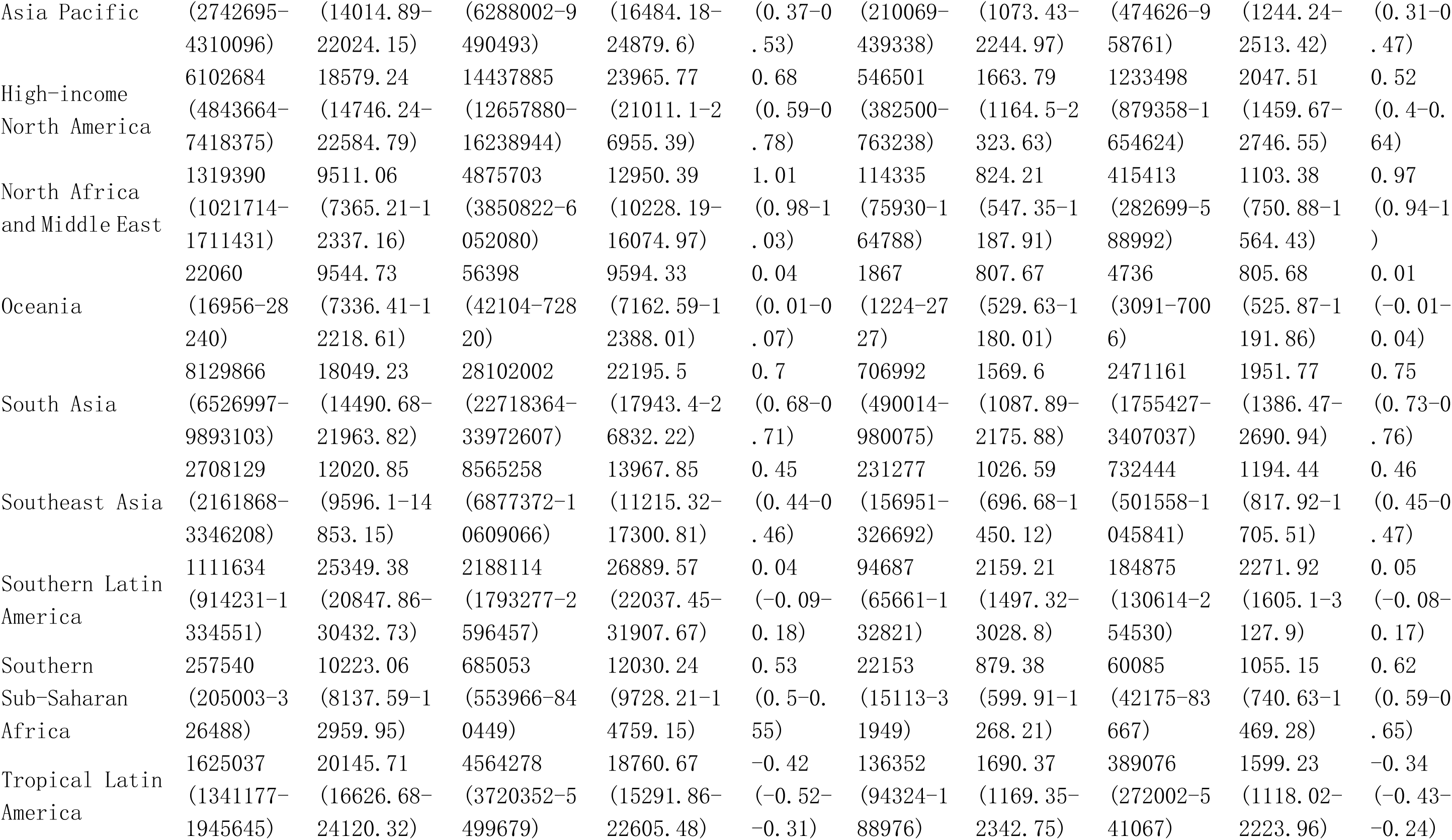
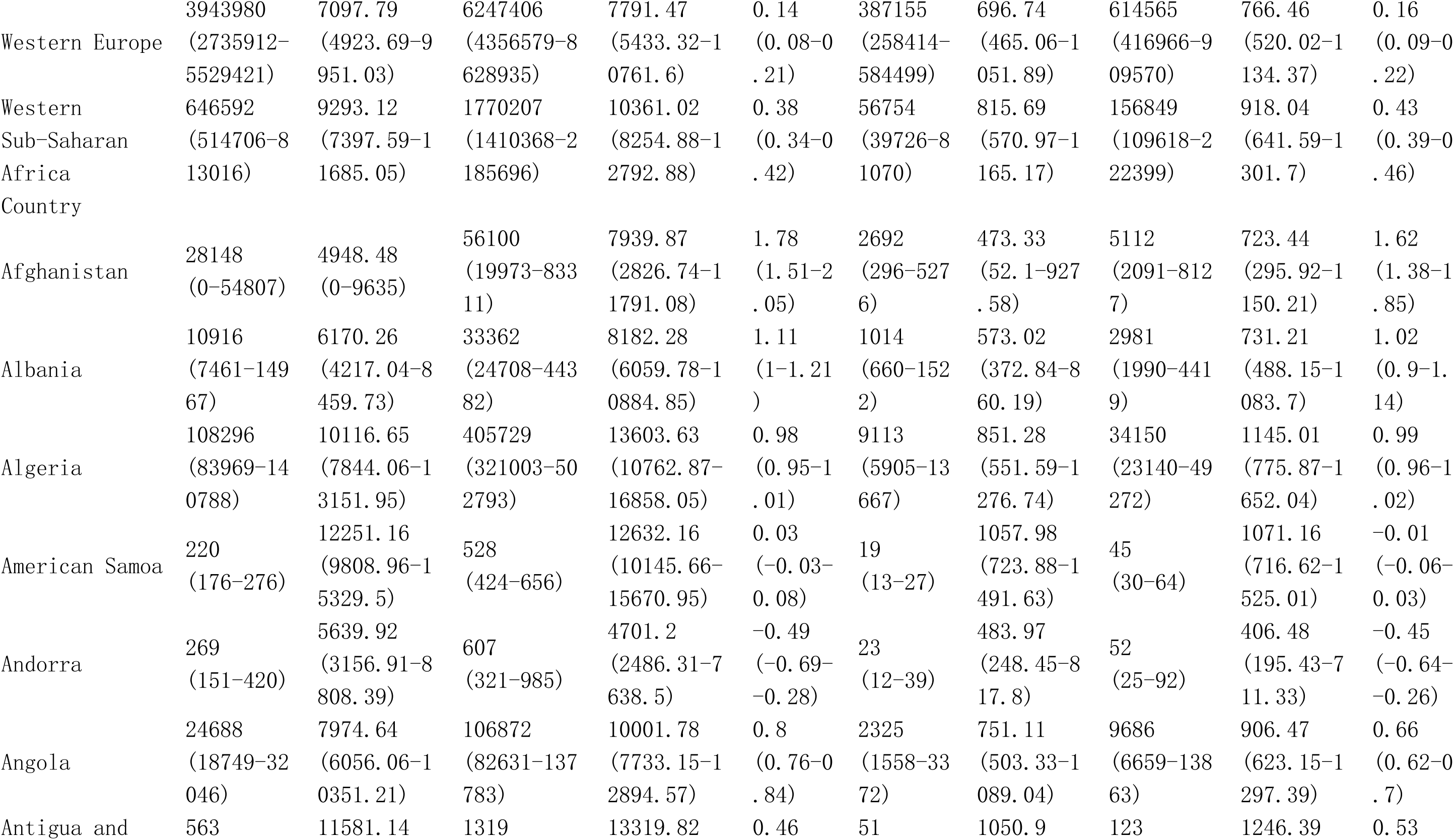
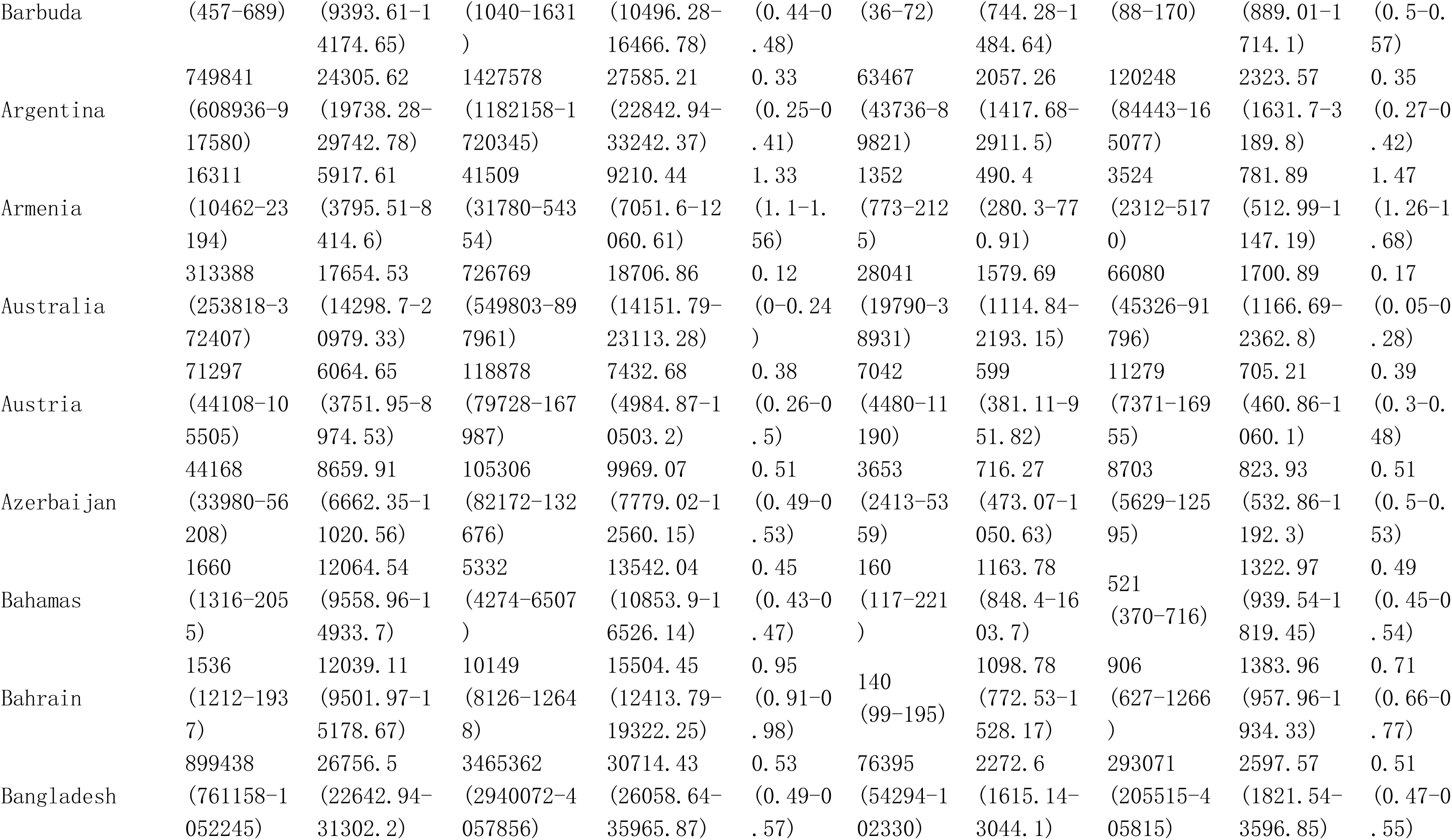
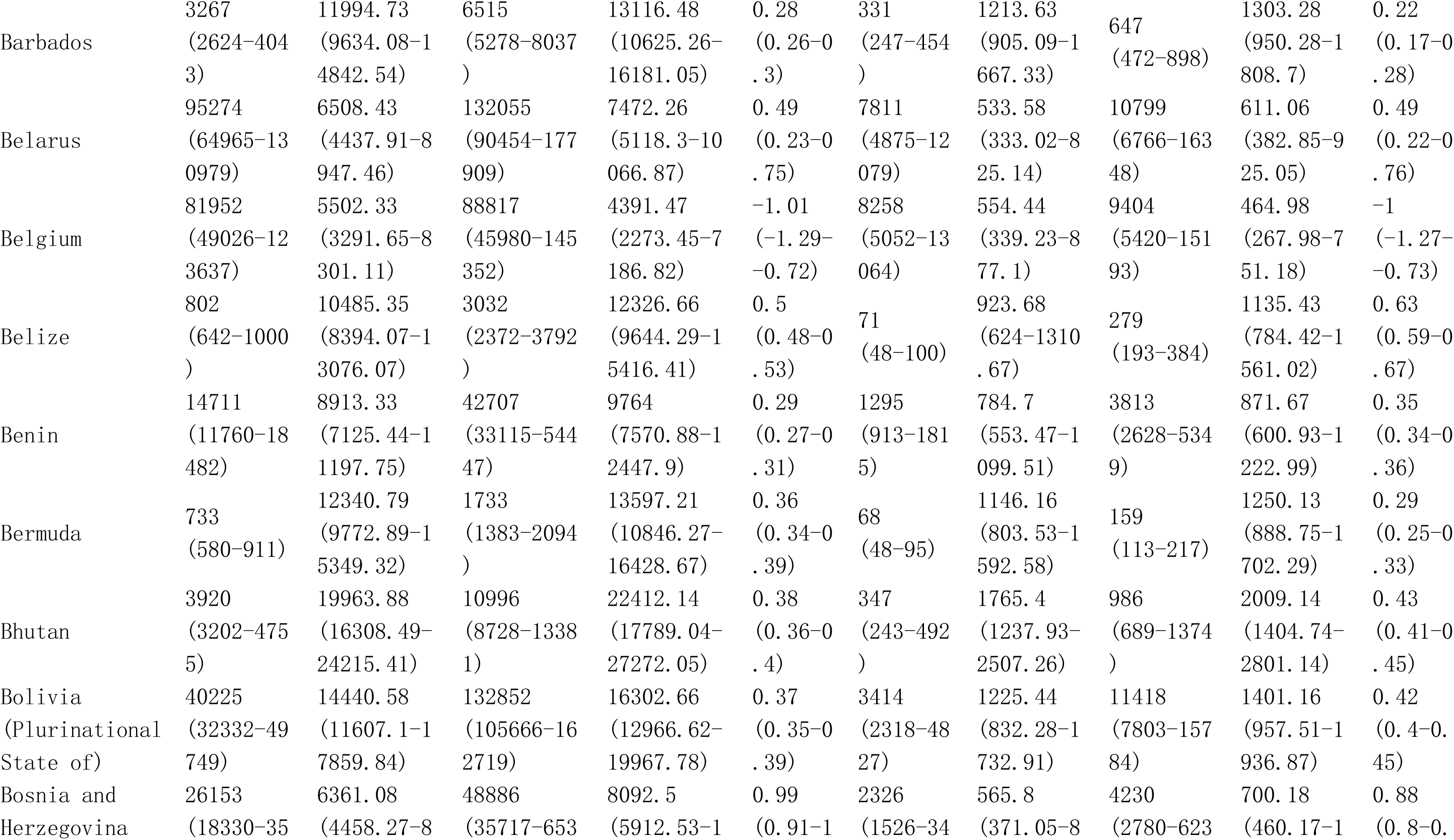
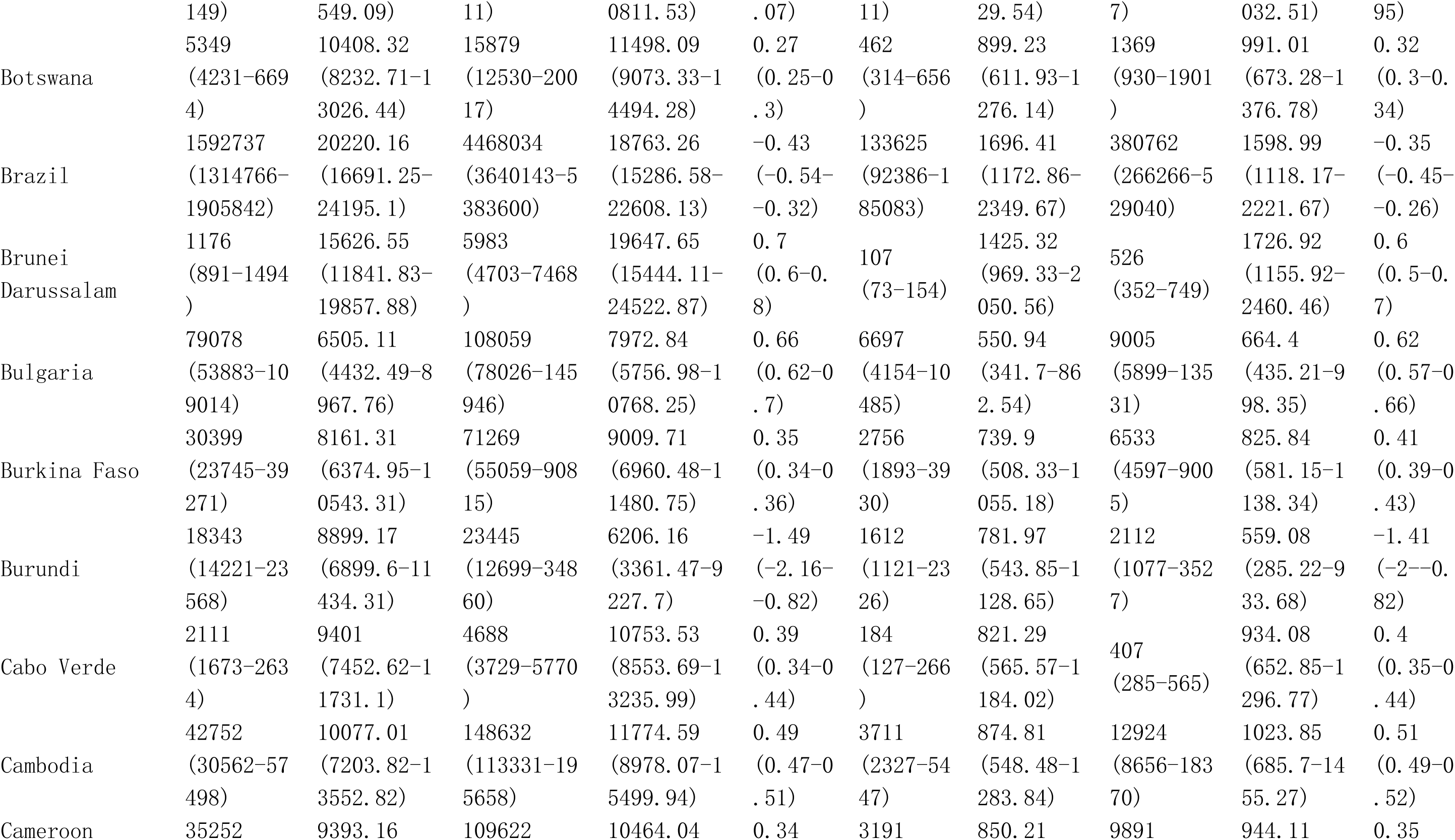
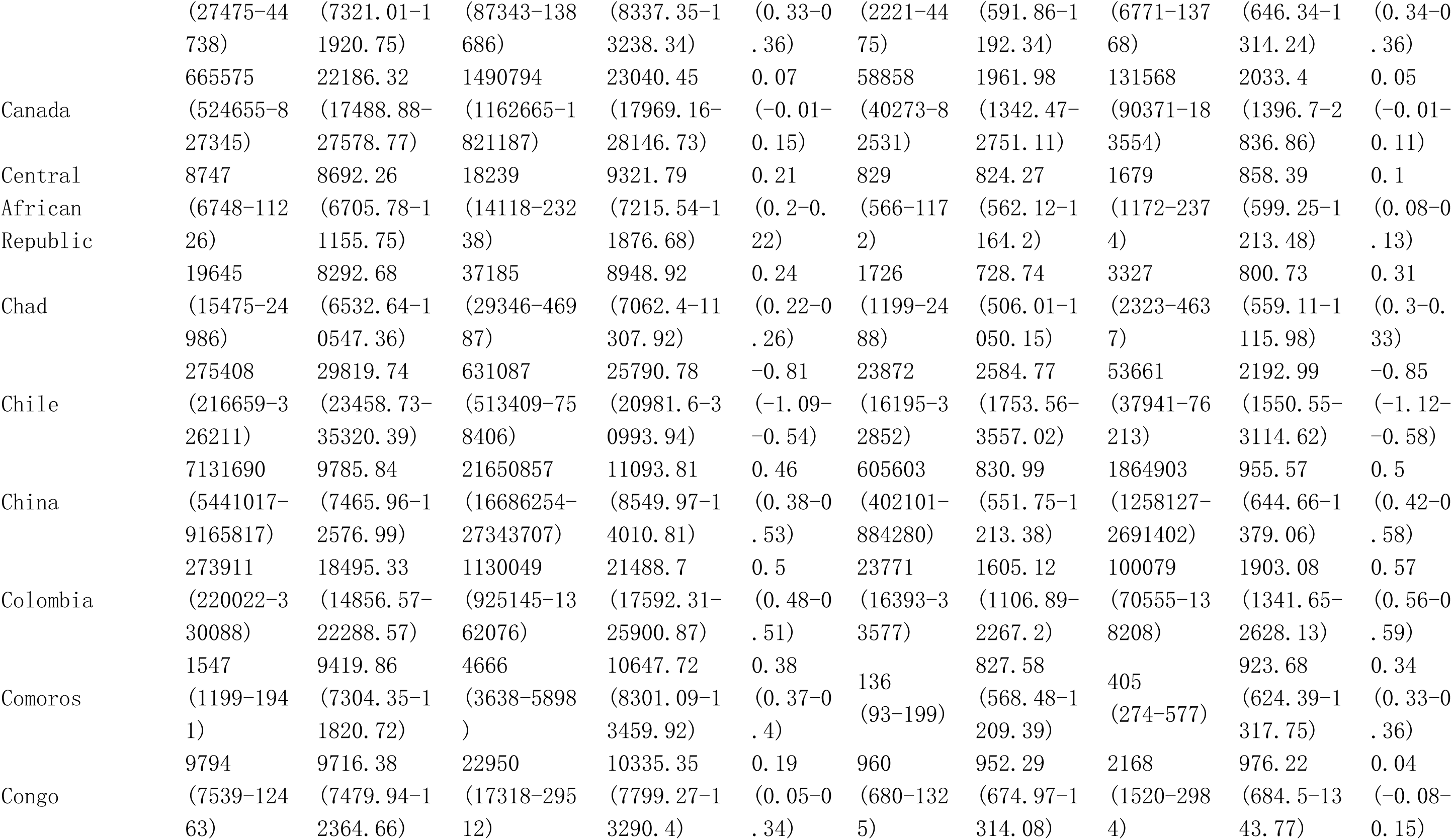
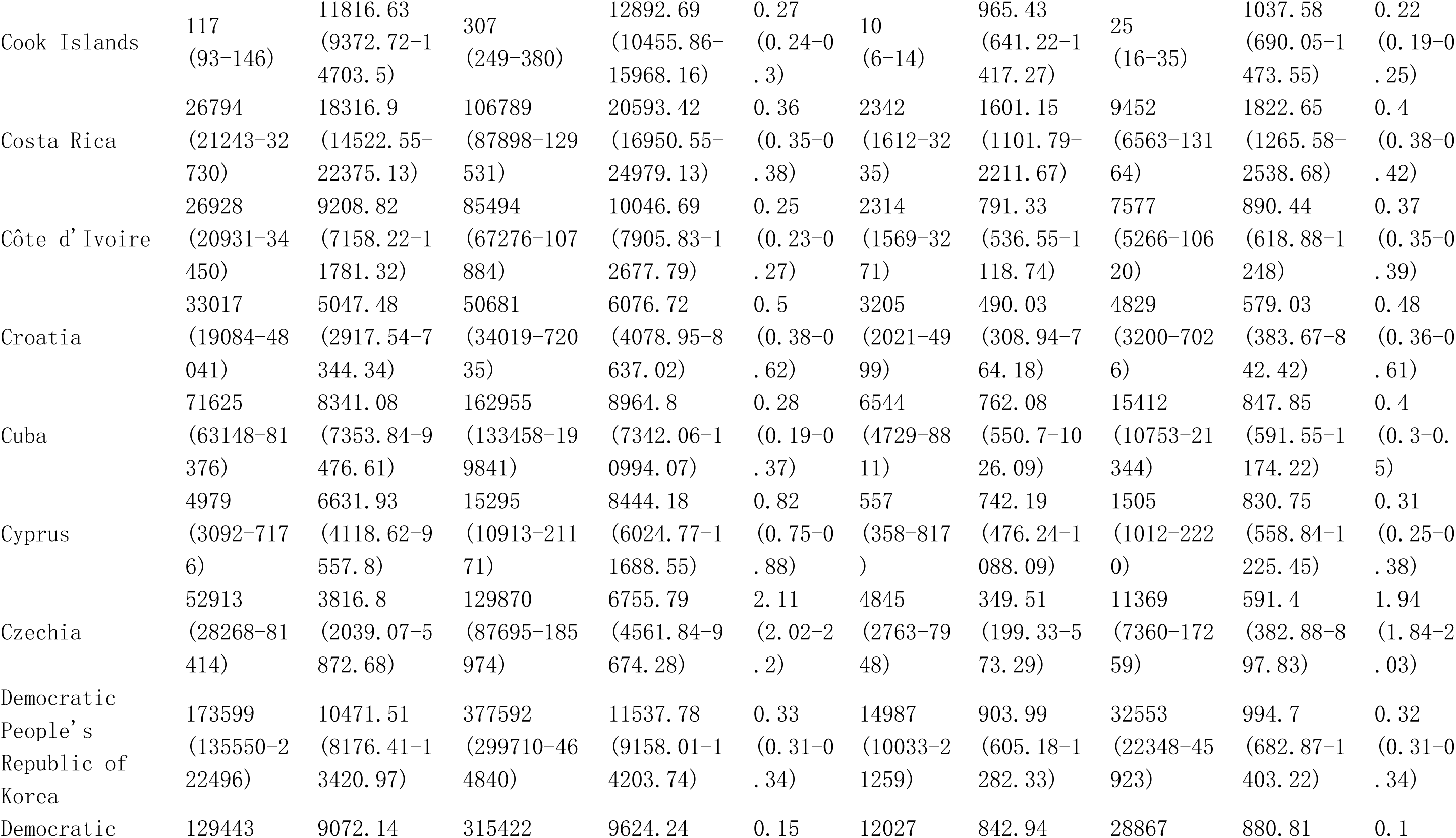
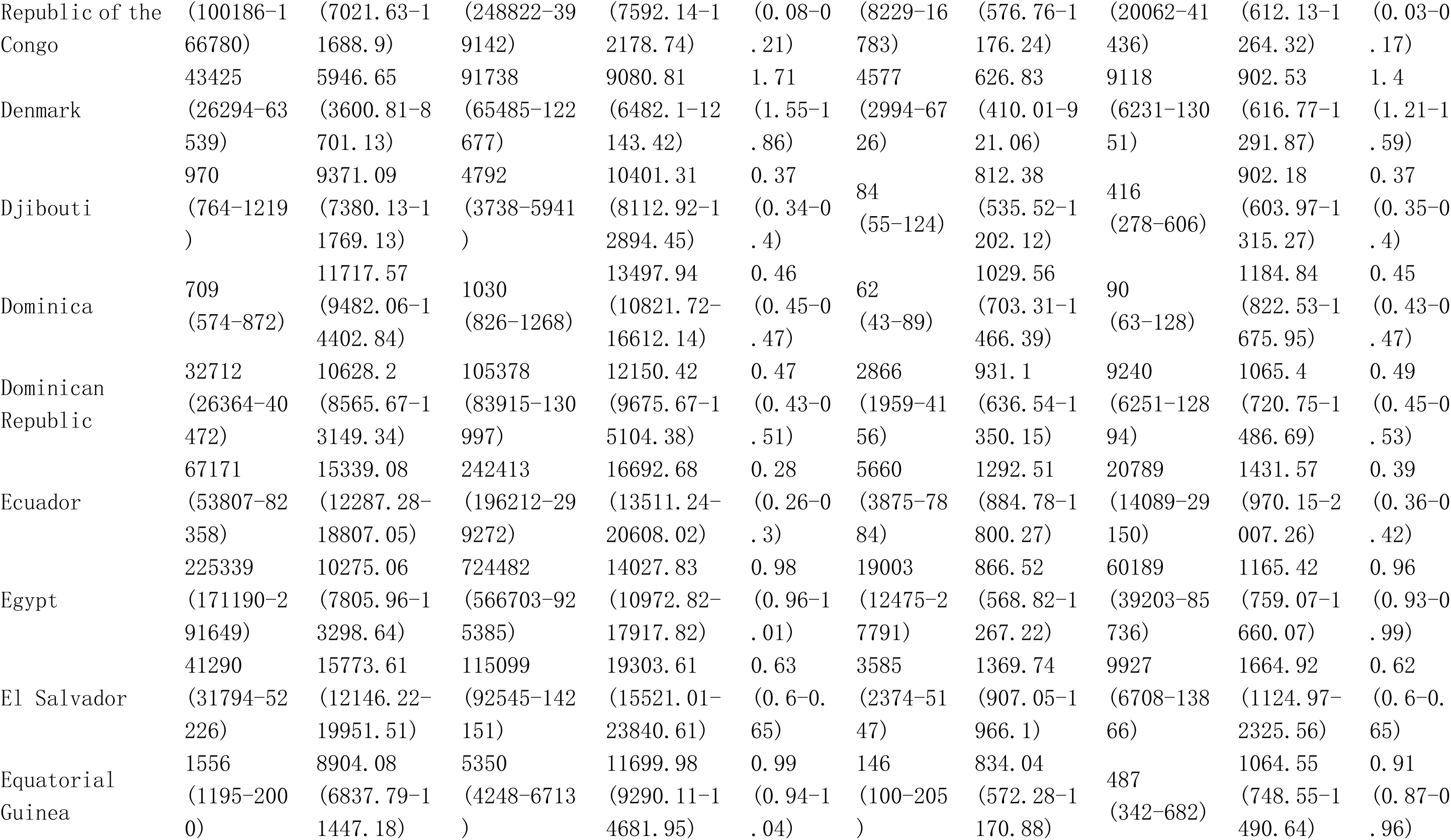
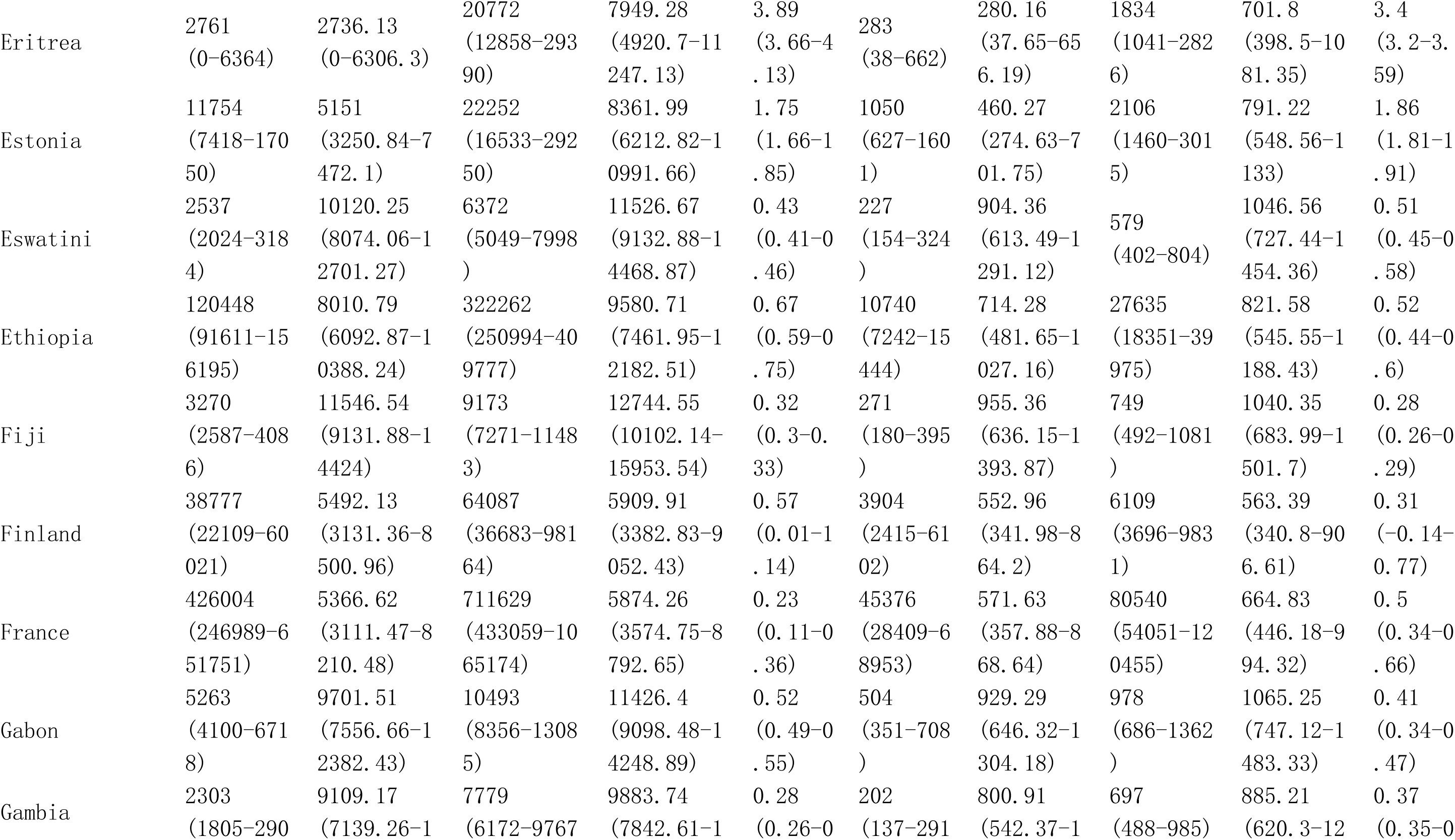
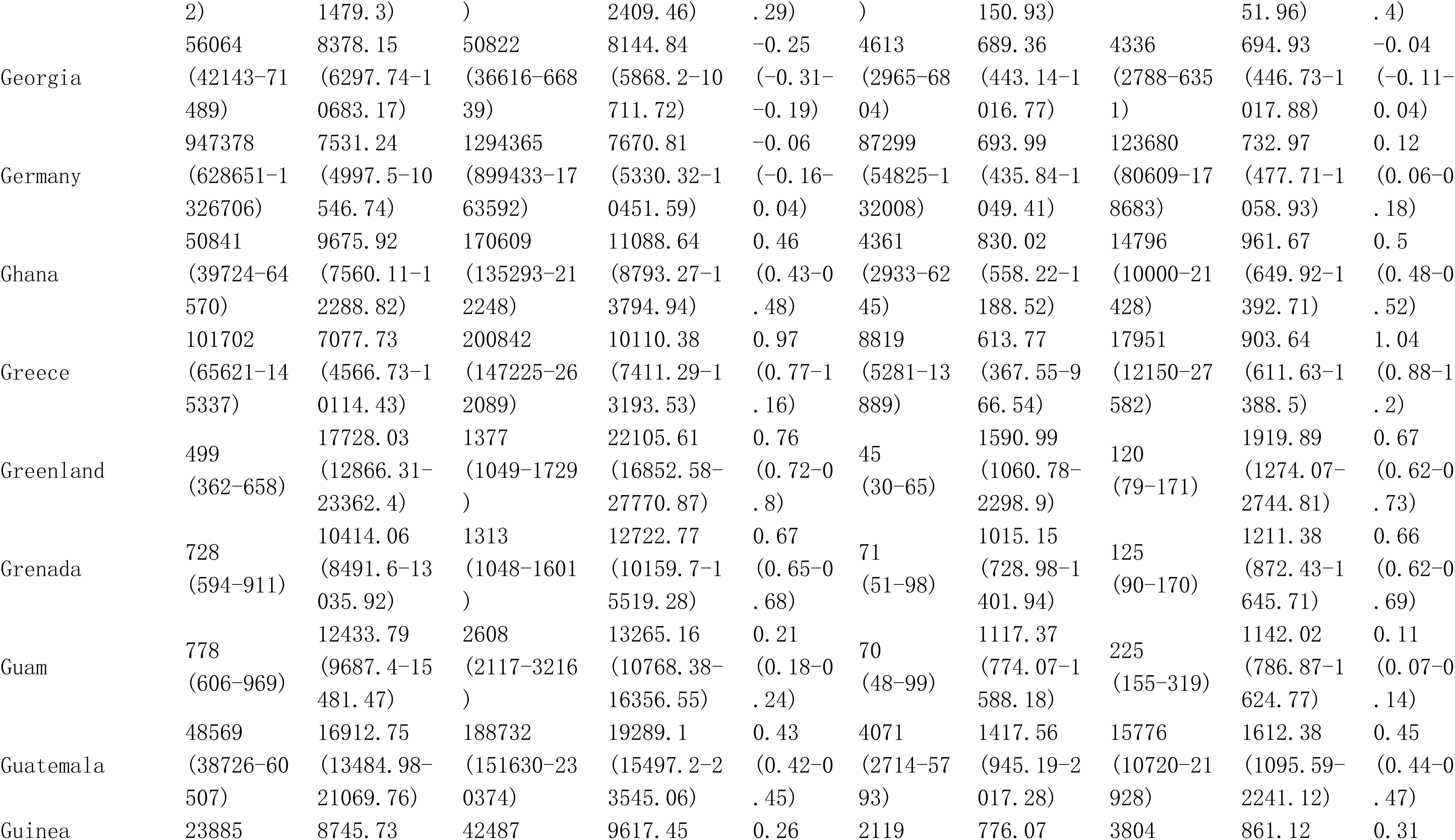
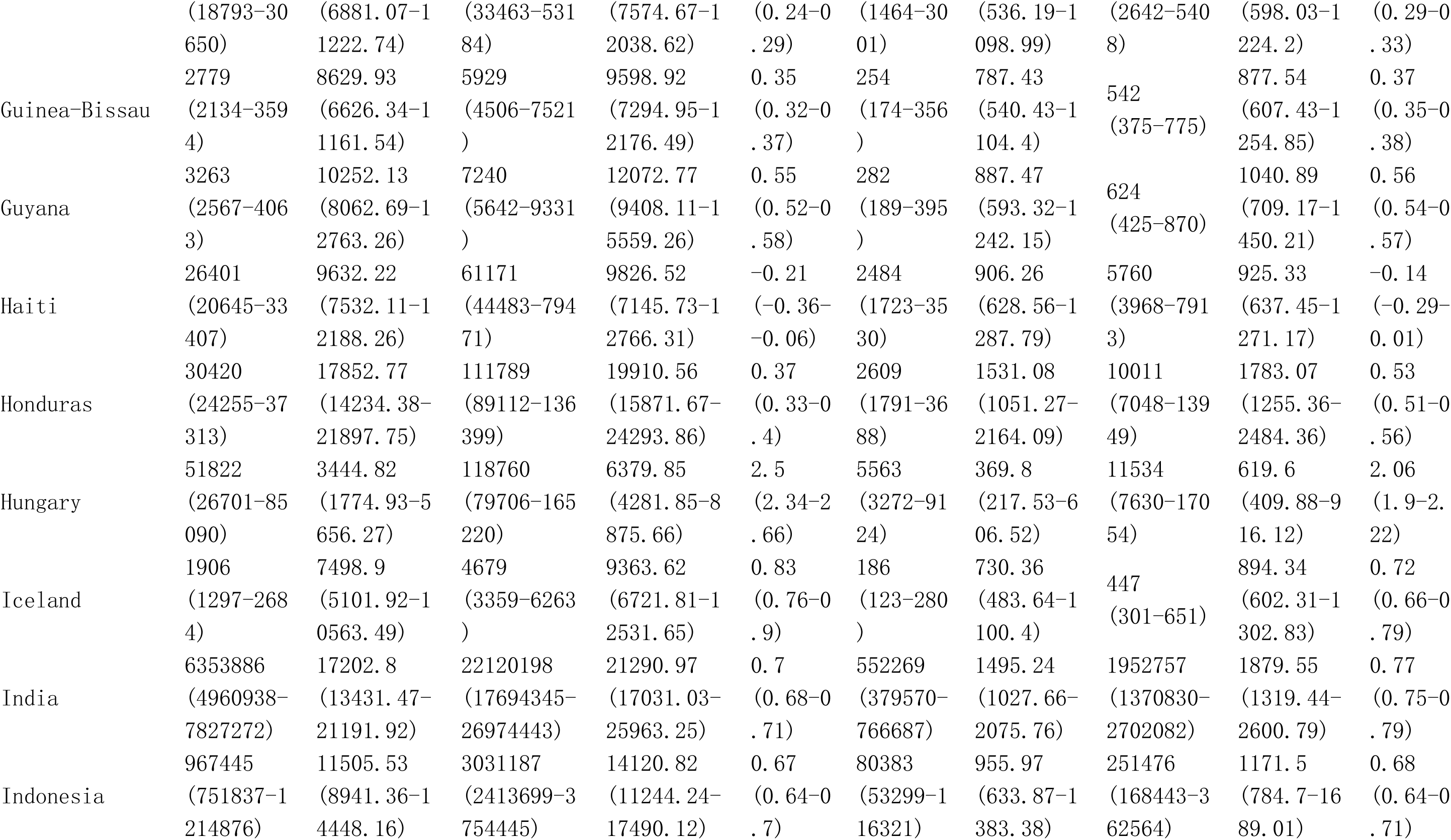
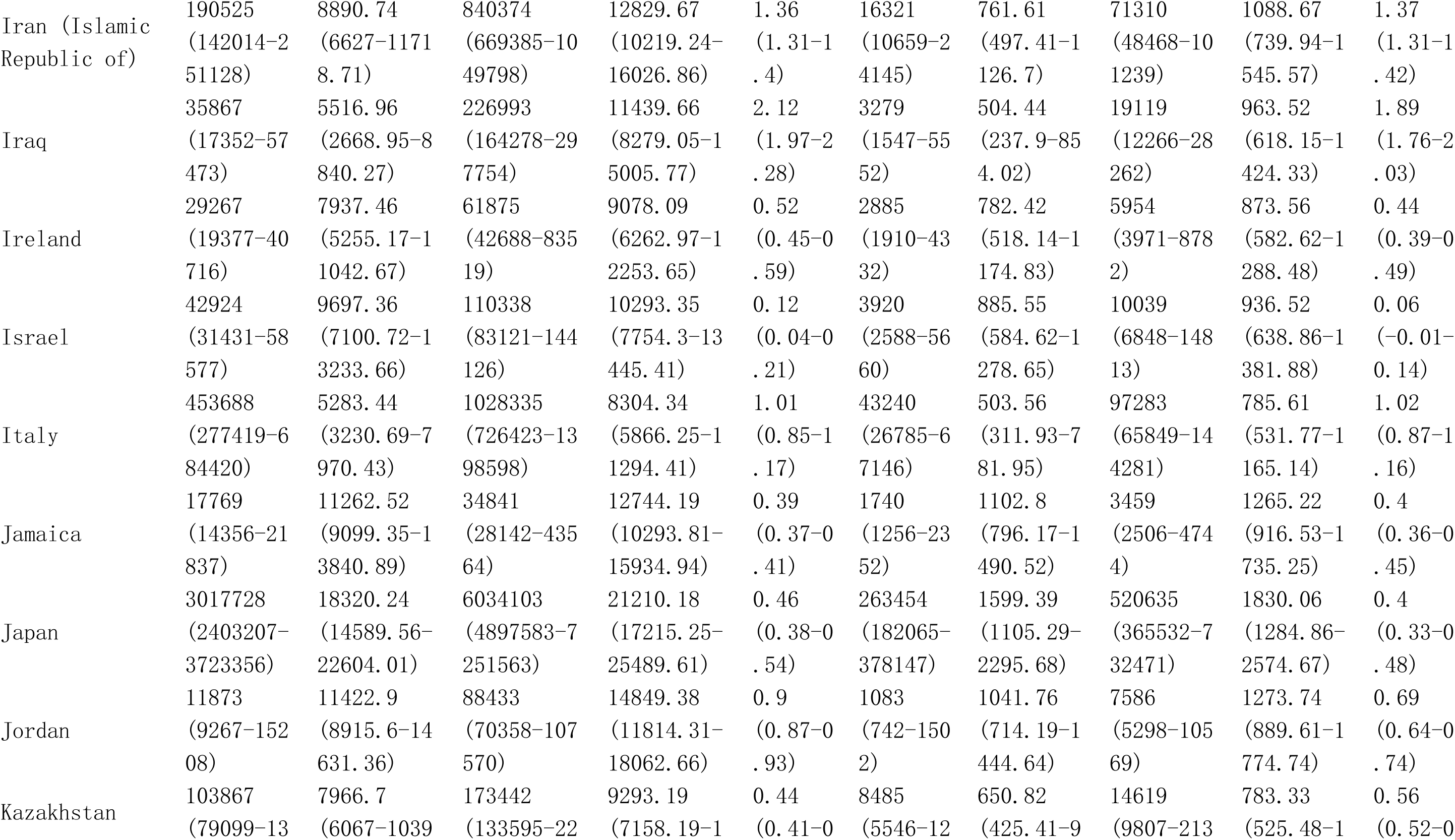
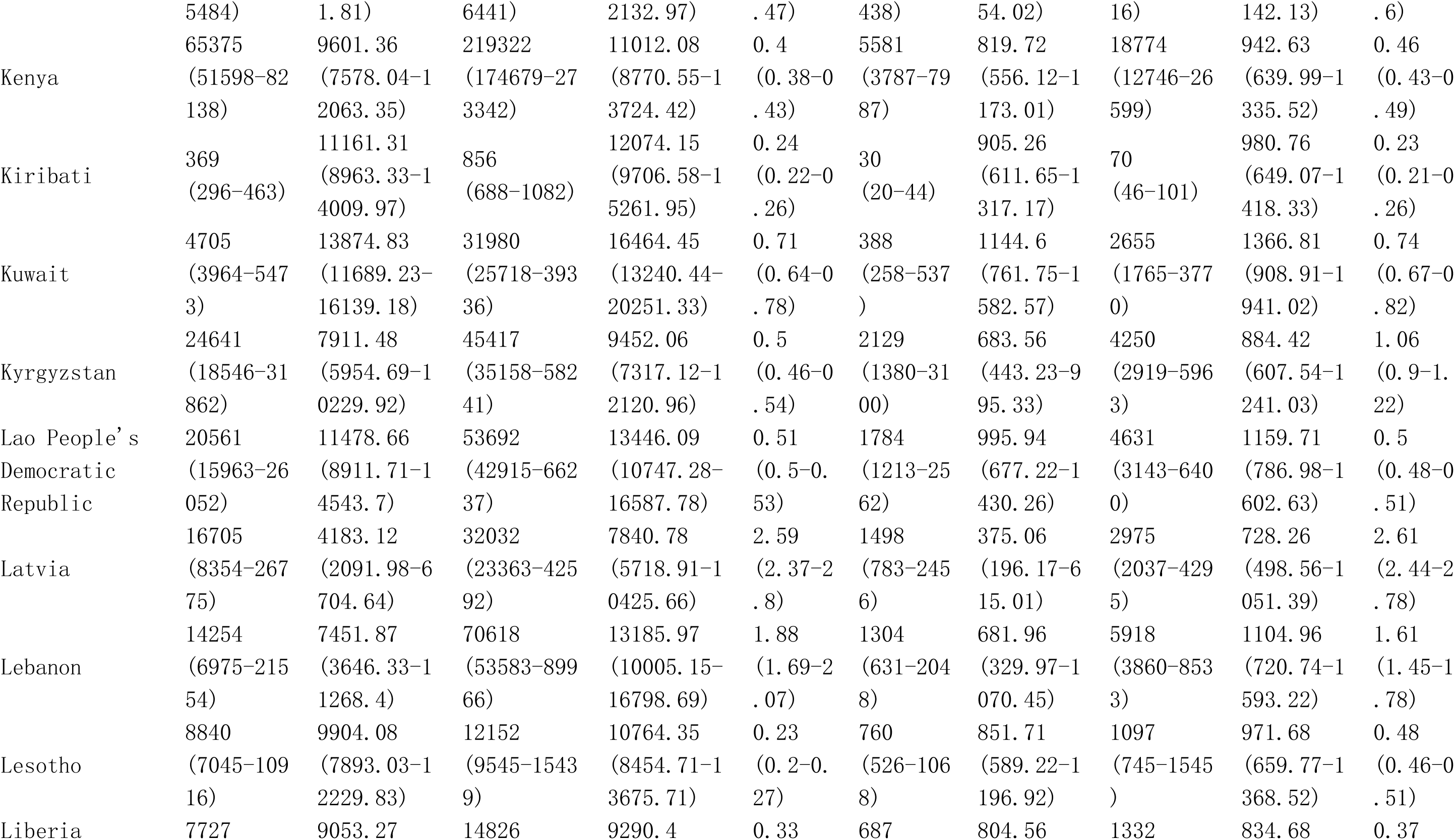
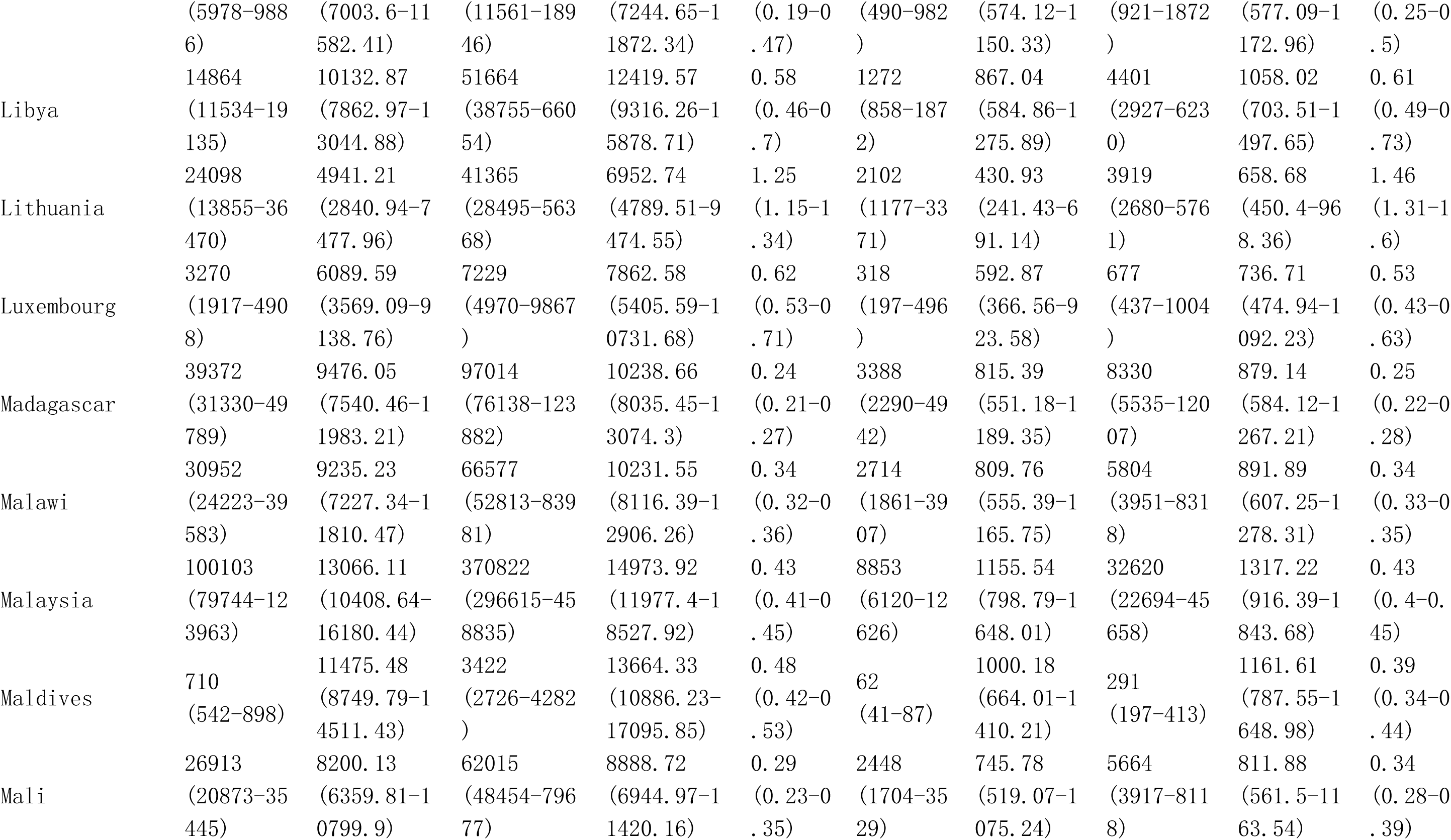
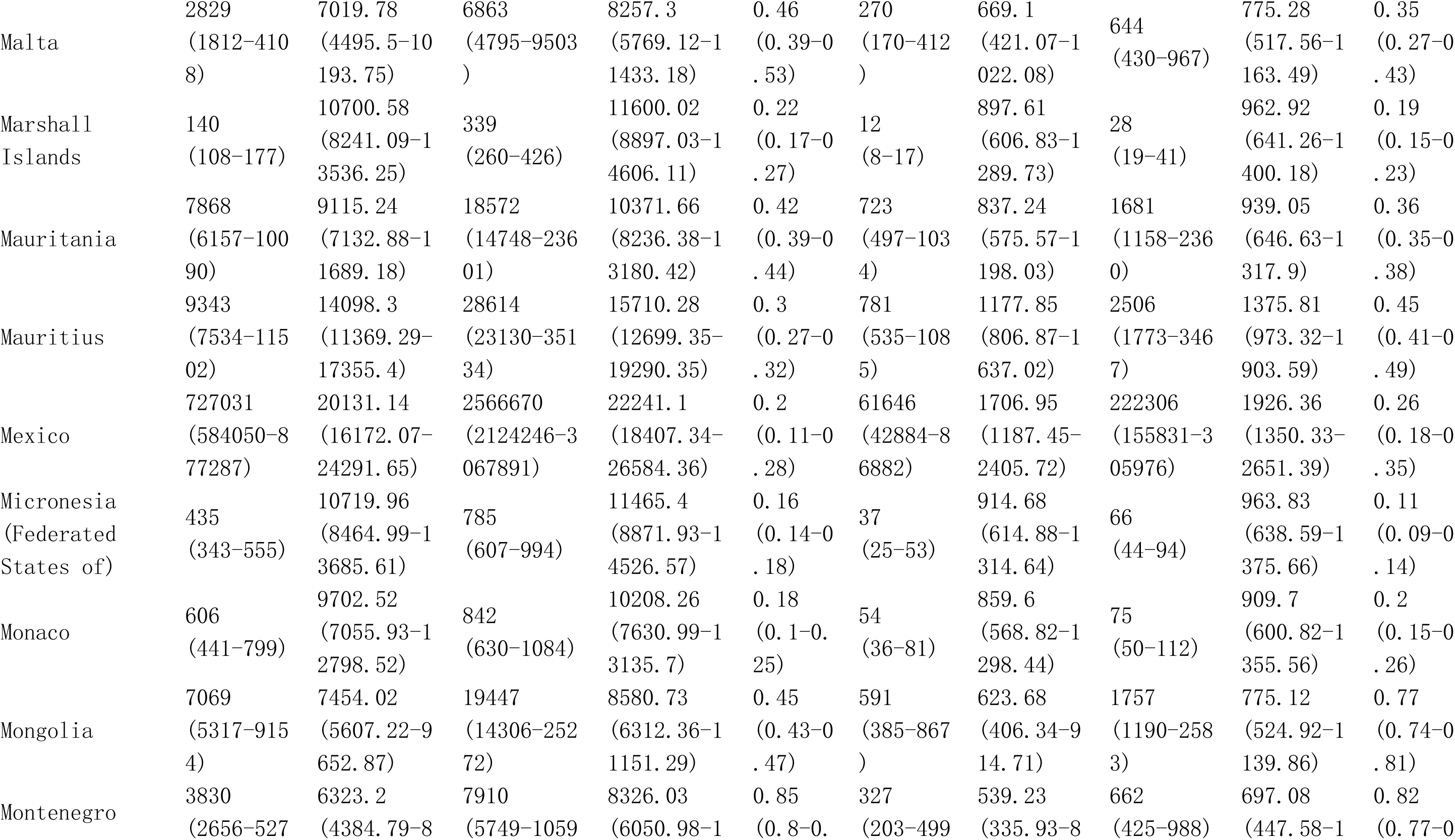
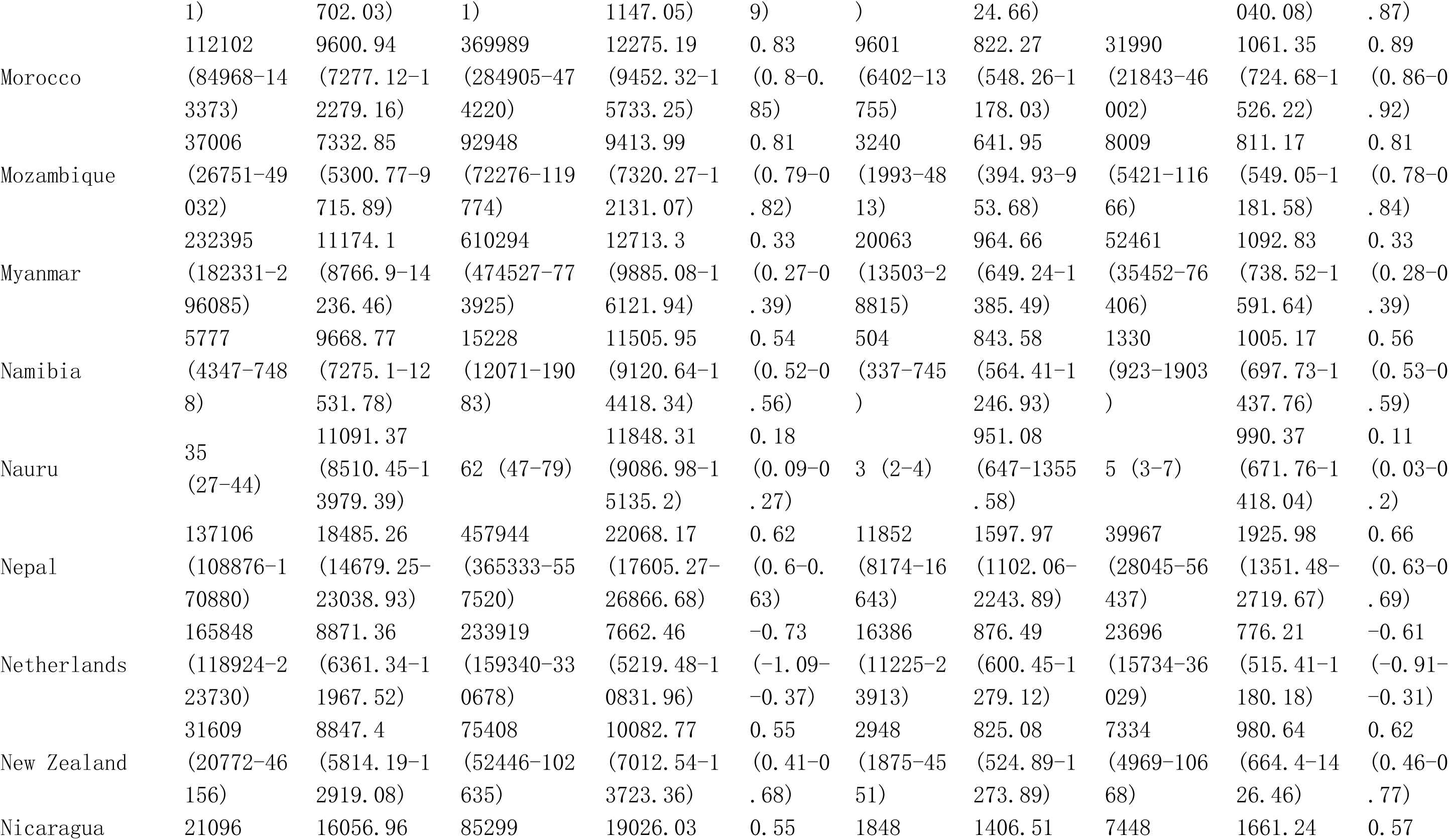
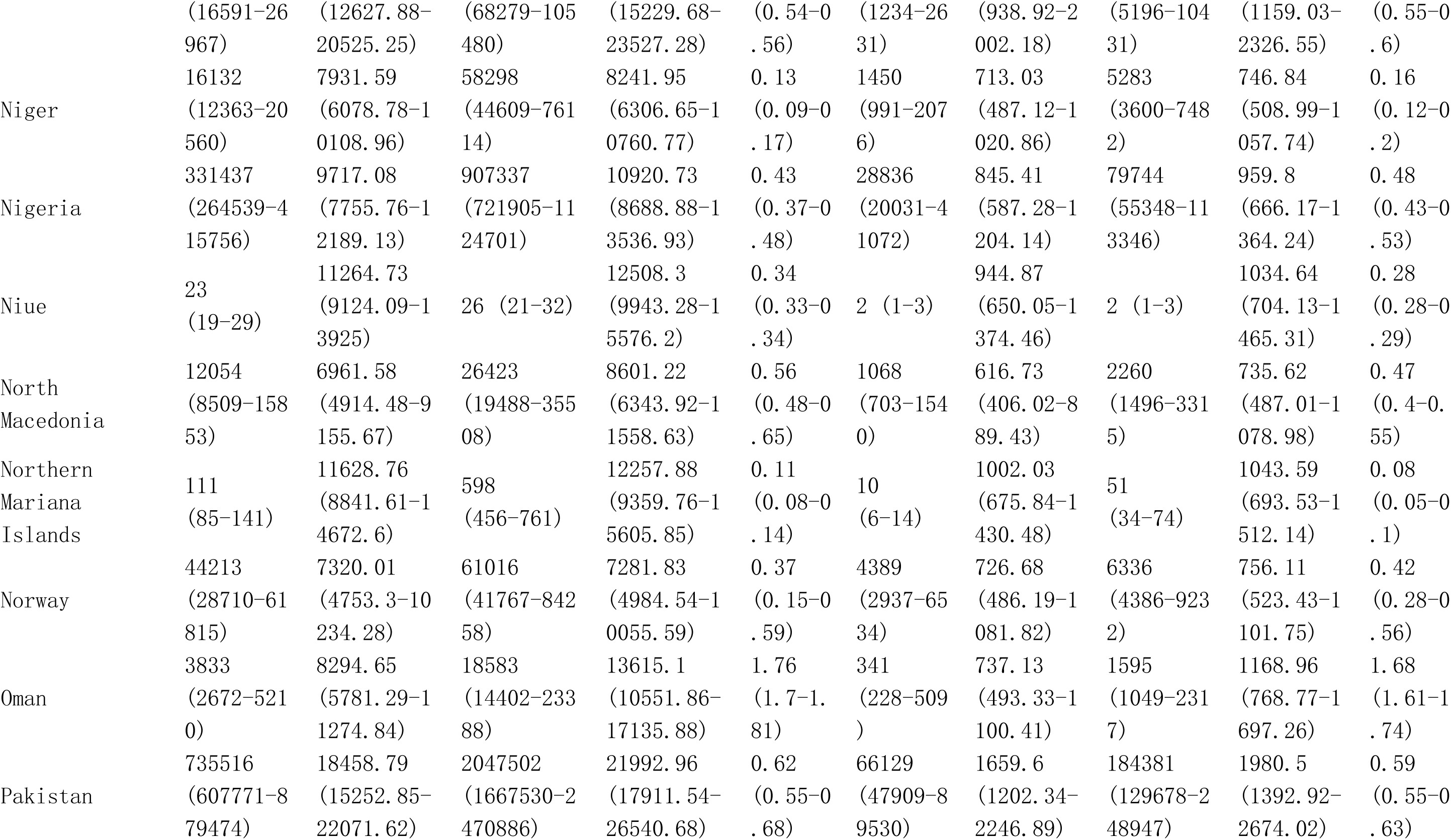
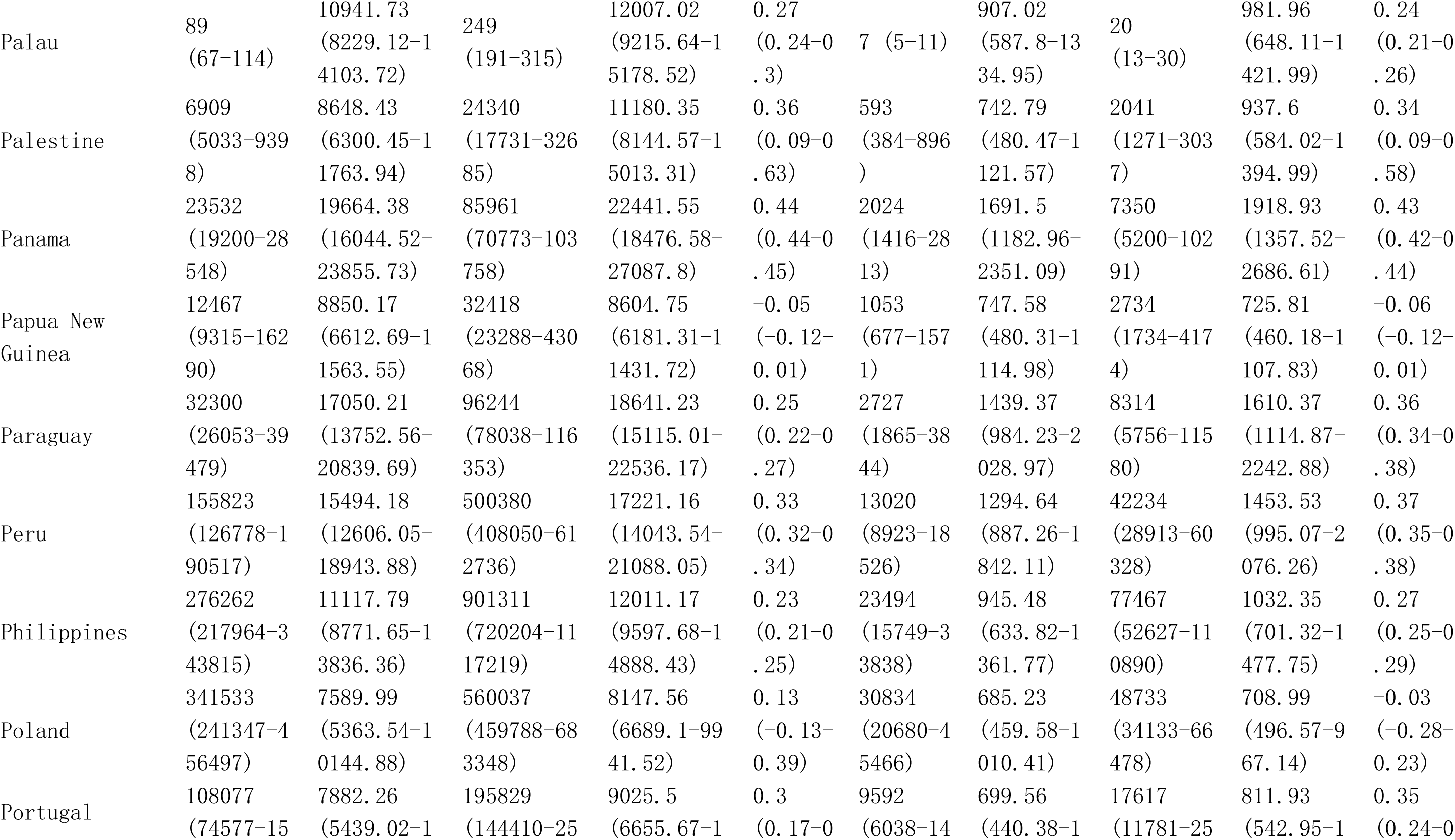
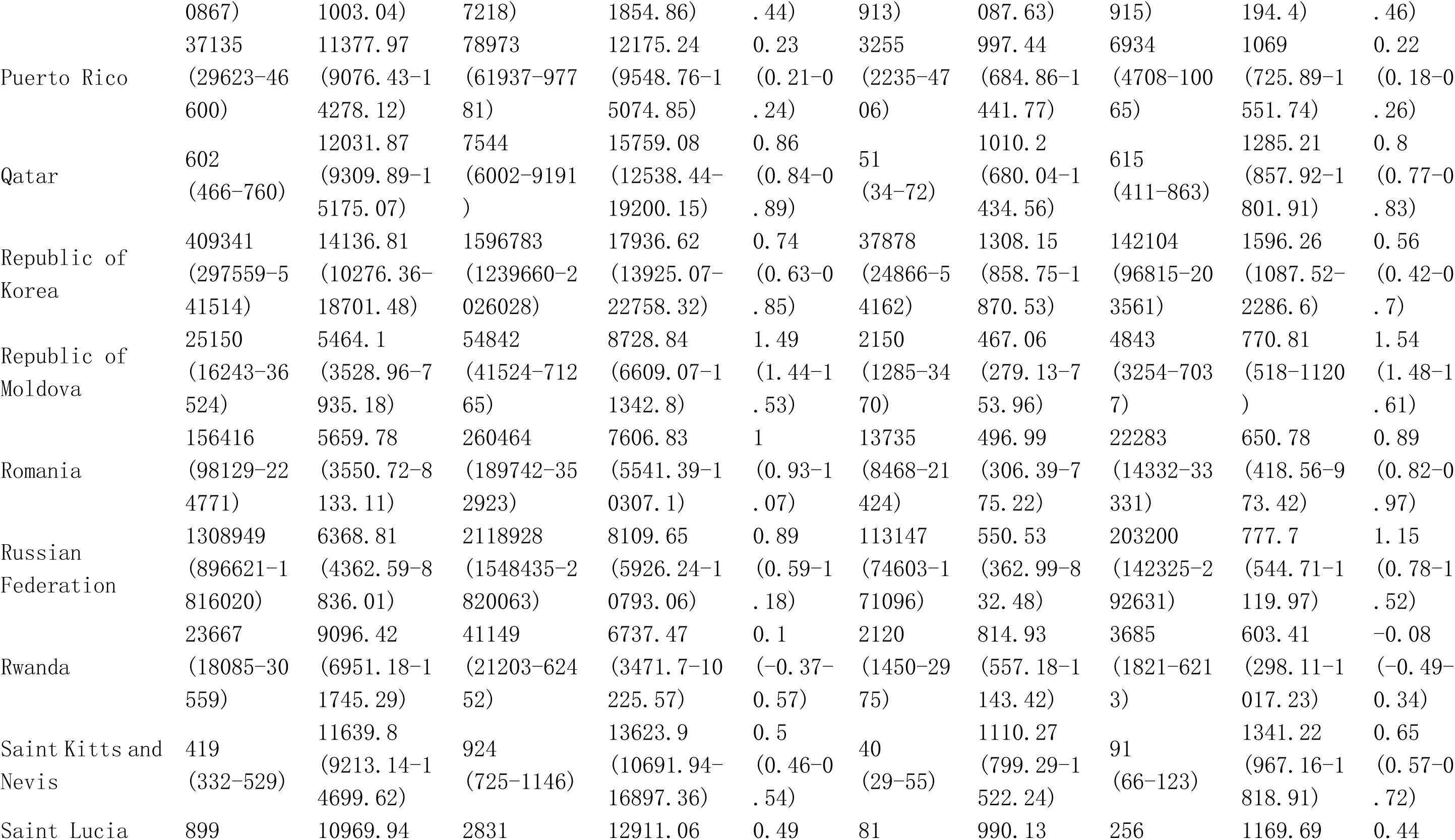
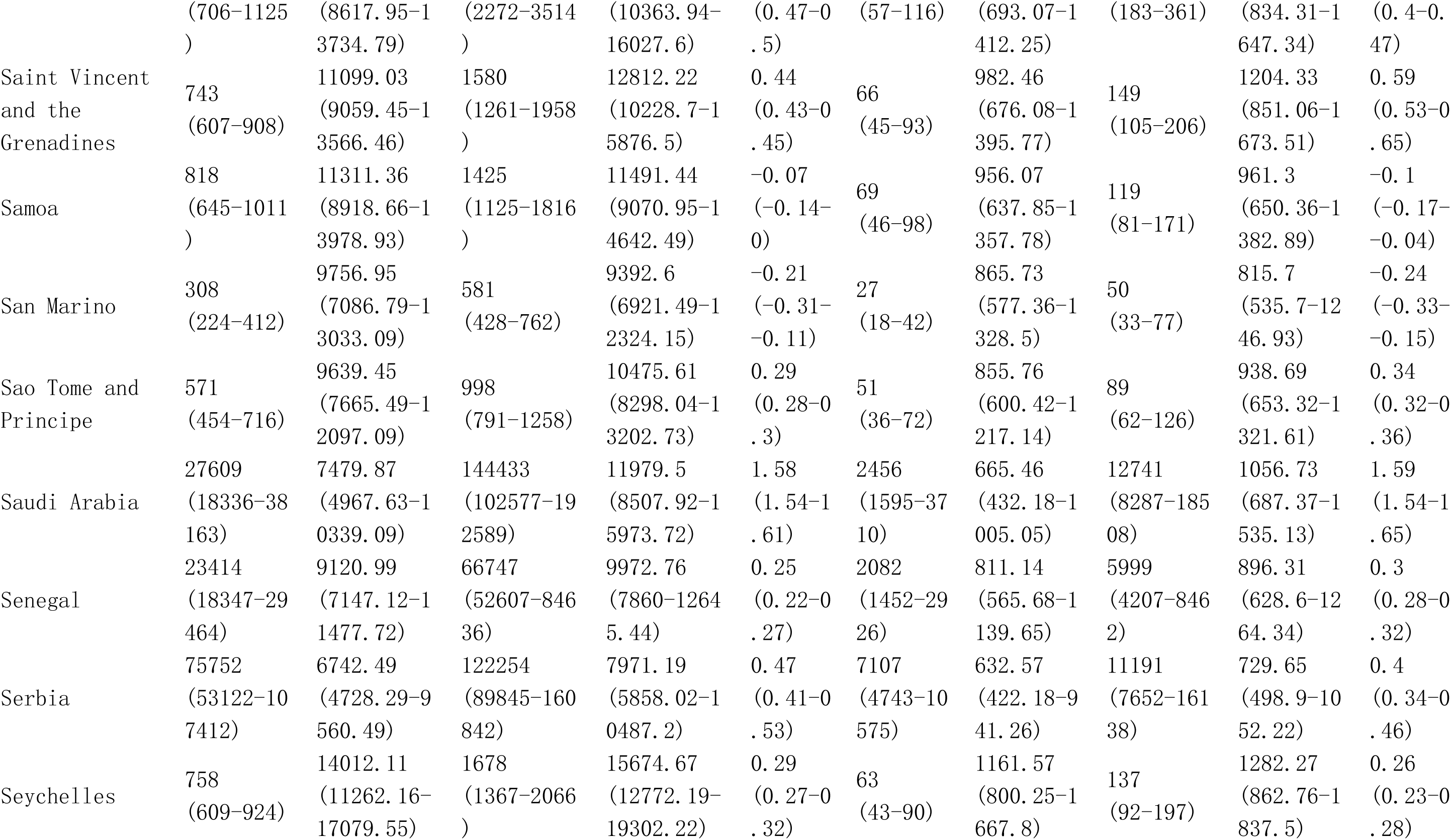
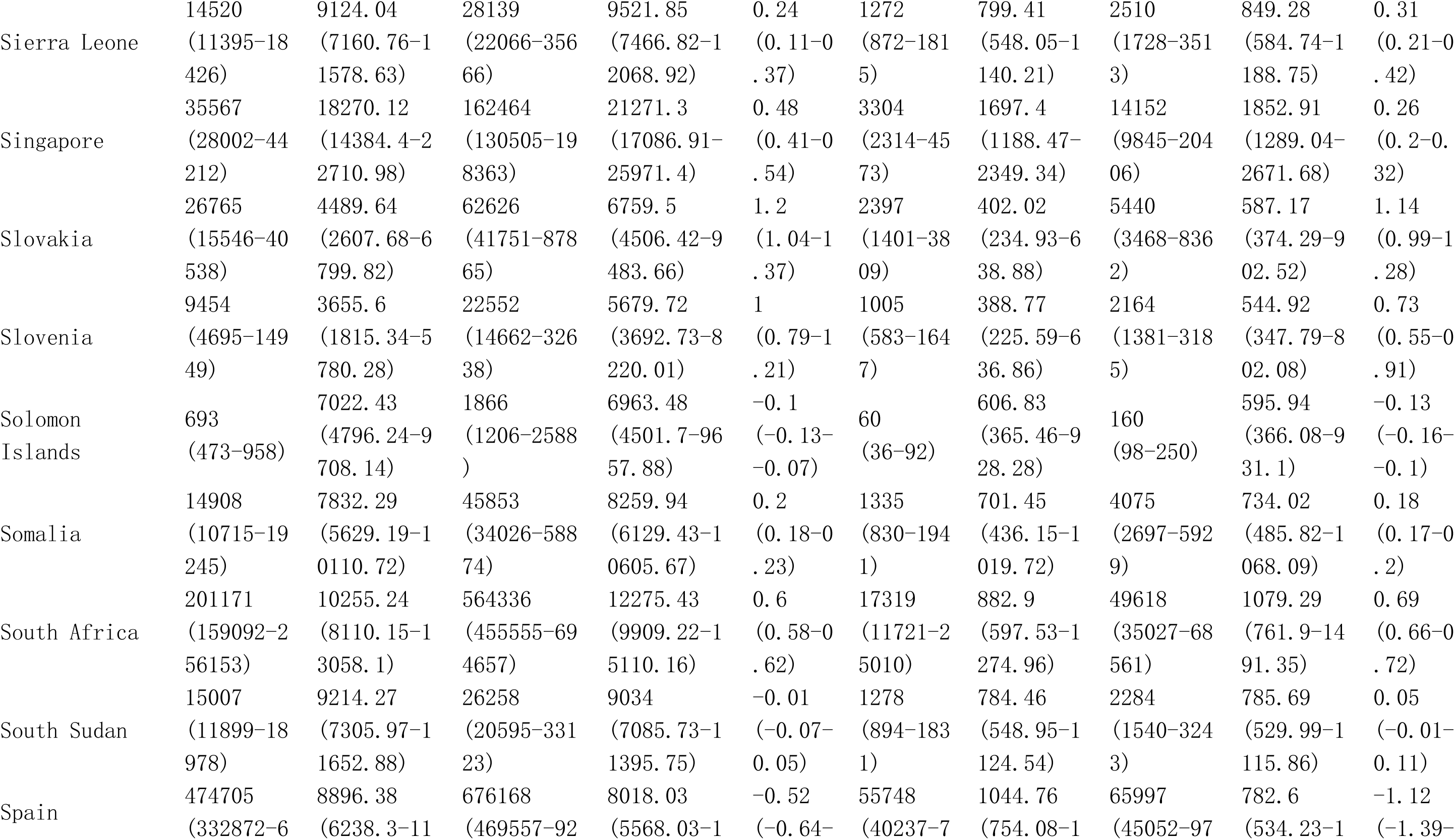
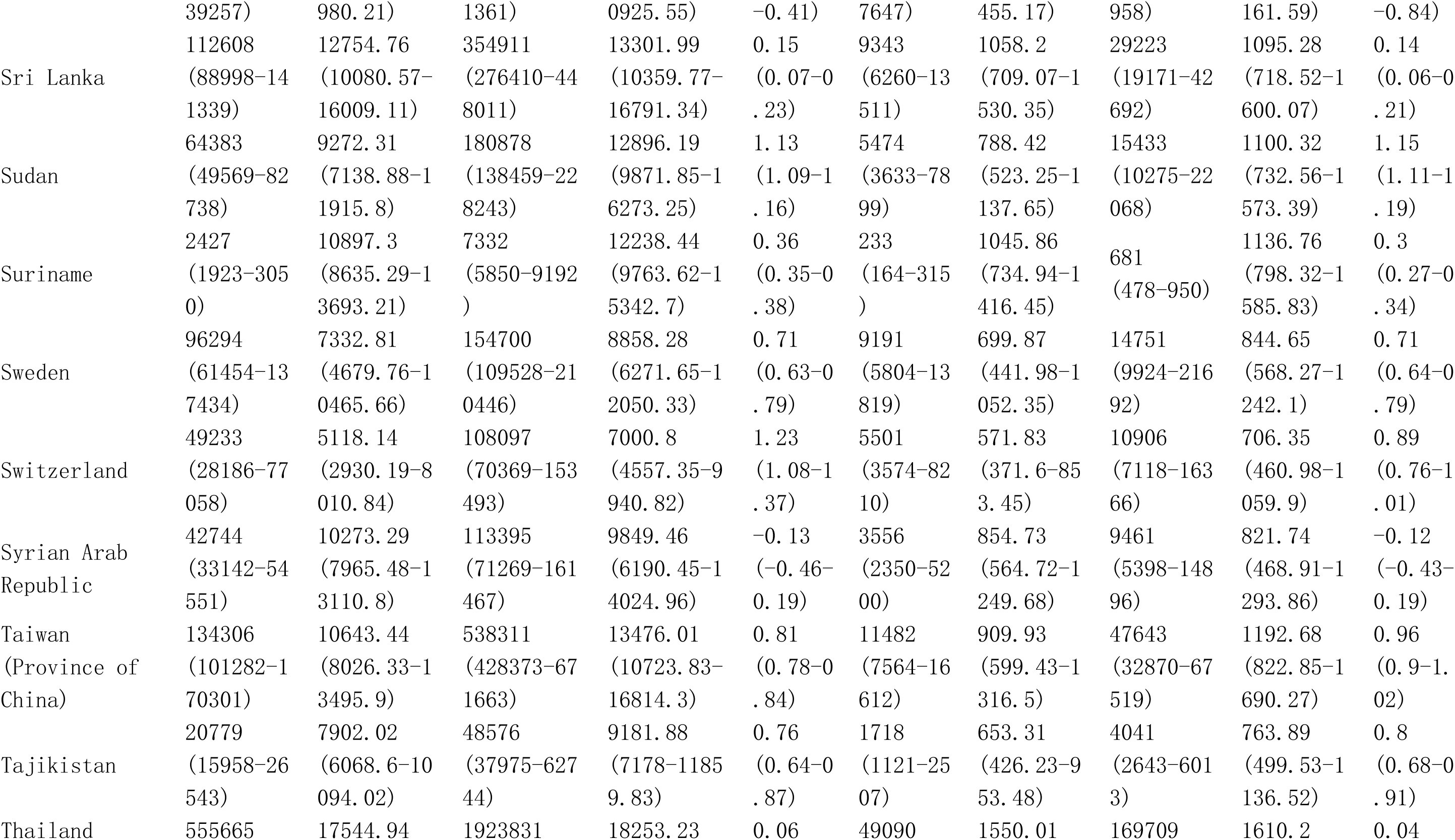
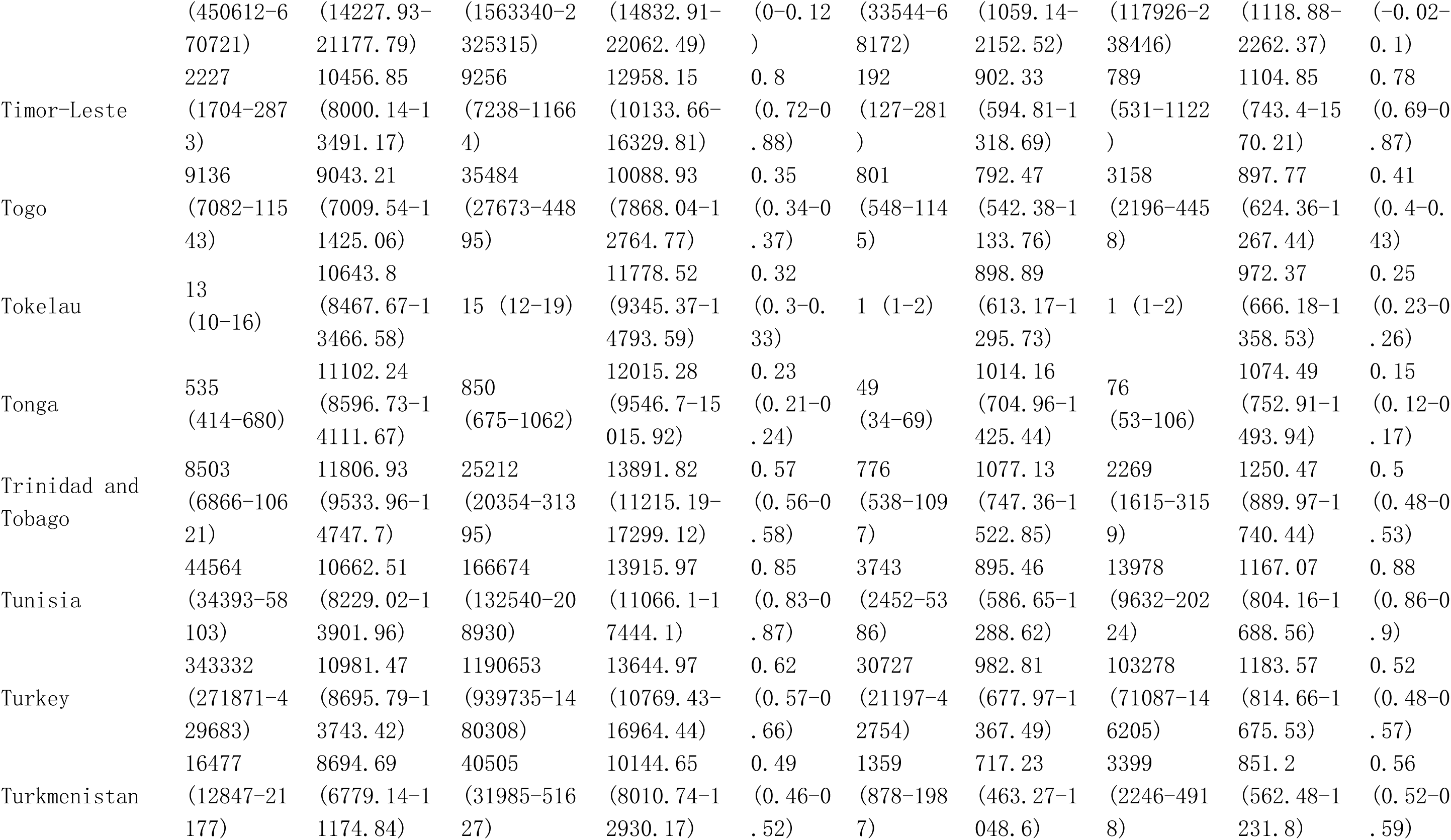
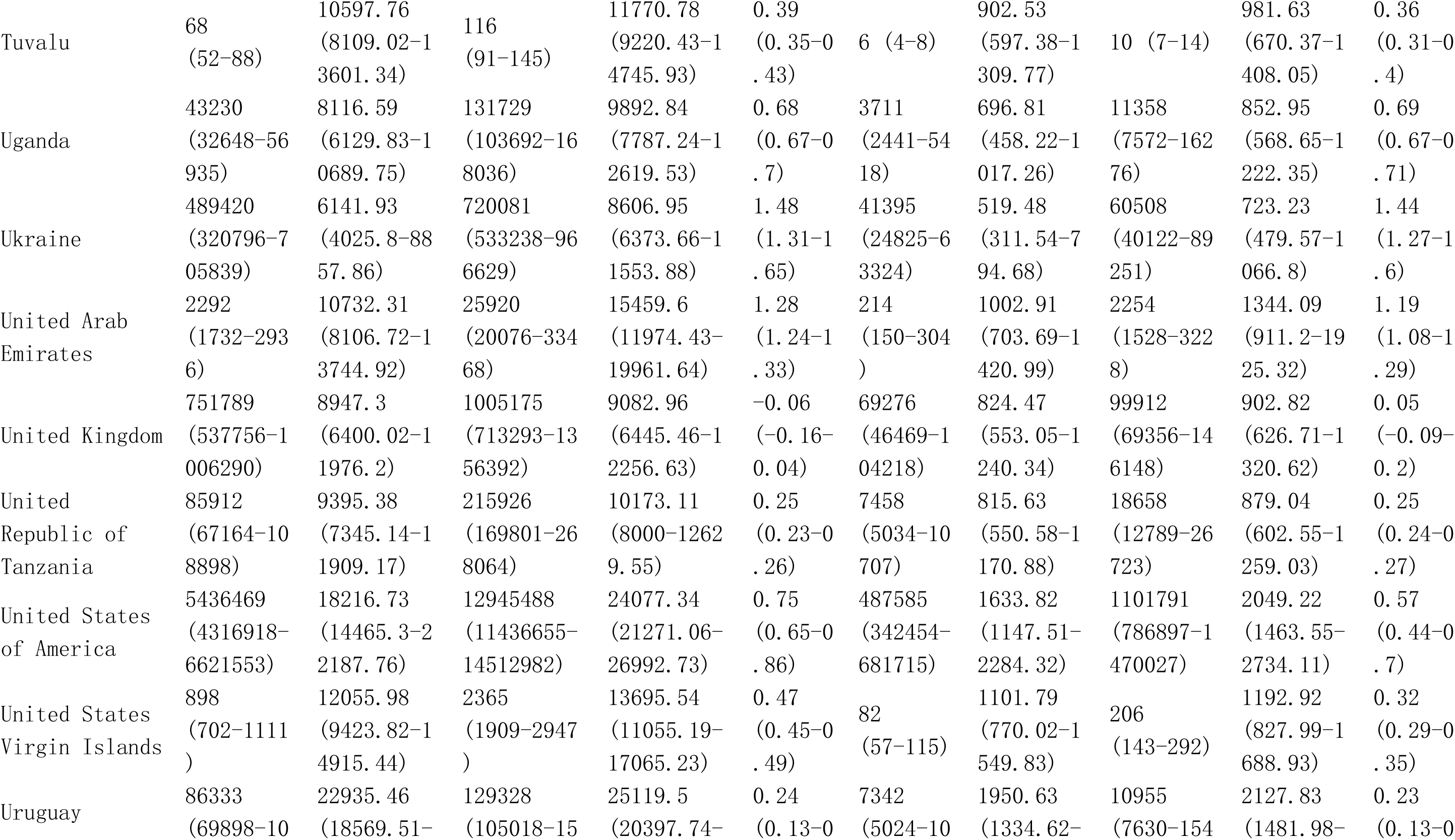
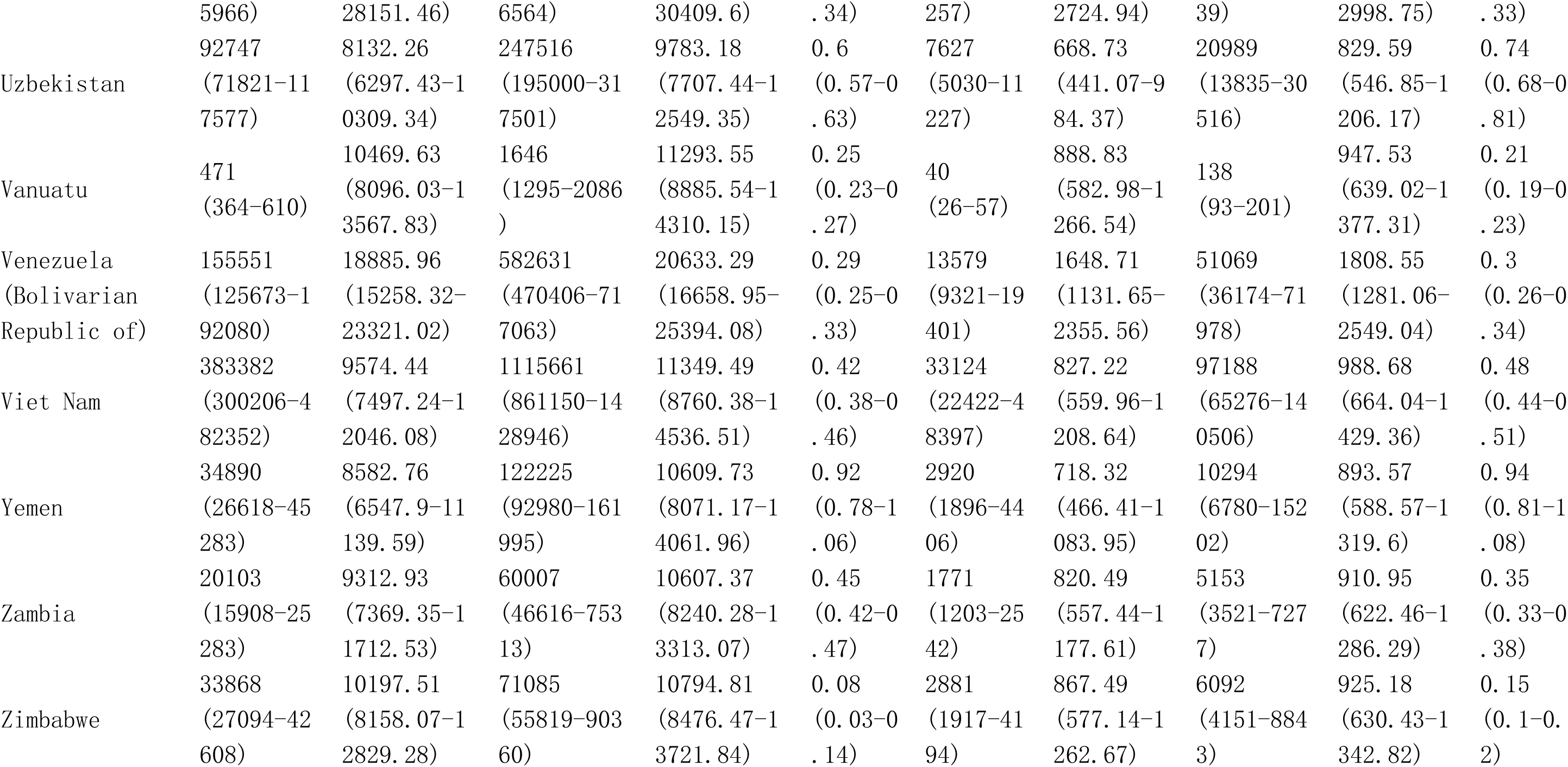
Global burden in Prevalence and DALYs of Other musculoskeletal disorders among postmenopausal women from 1990 to 2021 by 21 GBD geographical regions, and 204 countries and territories.

